# Replicating Health-Economic Simulation Models for Alzheimer’s Disease Using Artificial Intelligence

**DOI:** 10.1101/2025.10.08.25337408

**Authors:** Florence Gilson, Sander Osstyn, Ron Handels

**Affiliations:** Alzheimer Centre Limburg, Faculty of Health Medicine and Life Sciences, Mental Health and Neuroscience Research Institute, Department of Psychiatry and Neuropsychology, Maastricht University. P.O. box 616, 6200MD, Maastricht, The Netherlands; Division of Neurogeriatrics, Department of Neurobiology, Care Sciences and Society, Karolinska Institutet. 171 64 Solna, Sweden

**Keywords:** Health Economics, Health Technology Assessment, Alzheimer’s Disease, Artificial Intelligence, Model Replication, Engineering Prompting, ChatGPT-4, Transparency

## Abstract

**BACKGROUND:** Transparency and credibility of health-economic simulation models is essential to inform reimbursement decisions. Model replication can support model transparency and credibility. Artificial intelligence (AI), particularly large language models, offers new opportunities to accelerate model replication. This led to the research question: “To what extent can the results of existing health-economic Markov models be replicated by models developed using generative AI for eliciting input parameters and code generation?”

**METHODS:** Replication was performed in three steps. First, a chain-of-thought prompting strategy in ChatGPT-4 was developed to replicate in R an open-source model co-developed by one of the authors and with publicly available code. Second, it was applied to replicate a model co-developed by one of the authors but without publicly available code. Third, it was applied to a model without the involvement of the authors and without publicly available code. A mixed-methods approach was employed in terms of qualitatively addressing the face validity of the prompt development and refinement and quantitatively assessing deviations between AI-generated and original model predictions.

**RESULTS:** The first model required approximately one month to replicate, while adaptations to the second and third models took approximately two weeks each. Across the three models and 45 replications (15 per model), the average absolute relative deviations between ChatGPT-4 generated model predictions and published results were: ≤14% for quality-adjusted life years and costs in the first model, ≤7% in the second model, and ≤28% in the third model.

**CONCLUSIONS:** Our approach could support more time-efficient model replication for reimbursement decision-makers, researchers or pharmaceutical companies. This could contribute to transparency and credibility of health-economic models.

## INTRODUCTION

Health Technology Assessment (HTA) increasingly relies on health-economic models, particularly Markov models, to inform policy and reimbursement decisions. These models are essential for HTA bodies to assess the long-term costs and health consequences of new health- care interventions to ensure limited healthcare budgets are allocated efficiently. Given the crucial role of such models it is essential that they are transparent and therewith forming a basis for generating valid model results [^1^]. In addition to using reporting guidelines like the CHEERS checklist [^2^], a practical and robust way to assess the transparency of models is by replication of the results. This process can reveal missing information or insufficient data reporting [^3,4^].

However, a recent meta-research [^5^] highlighted a major gap in practice with nearly 44% of published health-economic evaluations, of which approximately 75% are model-based, not providing enough information for independent replication, emphasizing the urgency of the issue. Fully documenting a model’s details on structure, assumptions and inputs, as well as providing well-annotated code that complies with reporting standards demands considerable time and effort. This may discourage authors from sharing their full work as it is typically not required for publication or for reimbursement [^2,6^].

The increasing use of generative artificial intelligence (AI), particularly large language models such as ChatGPT, offers new opportunities to automate repetitive and resource-intensive tasks. Recent studies have started exploring how generative AI can support various tasks that could be relevant to health-economic model replication. For instance, it has been shown to reduce the time required for extracting and screening data from scientific publications in the context of systematic reviews [^7,8^] and in the replication of four network meta-analyses [^9^]. The latter study also reported promising results in ChatGPT’s ability to generate executable R scripts for replicating each network meta-analysis end-to-end. Reason et al. (2024) [^10^] replicated two published partitioned survival models using structured prompts with GPT-4 and achieved 93% error-free replication across 15 trials. Chhatwal et al. (2024) [^11^] demonstrated the feasibility of developing a de novo Markov-based model for hepatitis C using GPT-4. In their approach, GPT-4 was used to search PubMed for relevant published health-economic models, while ValueGen.AI, a GPT-4-based platform, supported the estimation of model parameters. The model was constructed and executed in R using the “Heemod” package. In another study Chhatwal et al. (2024) [^12^] used ValueGen.AI to replicate a ”simple” published Markov-based health-economic model by extracting the model structure and parameters and implementing the model in R using “Heemod” package. In another study Chhatwal et al. (2025) [^13^] replicated a published report on a cost-effectiveness modeling study on Alzheimer’s treatment, using Python facilitated large language model interactions to build the model using “Heemod” R package.

Given the scarcity of transparent, open-source models and the current knowledge gap in the use of generative AI to replicate closed-source models we developed the following research question: ”To what extent can the results of existing health-economic models be replicated by models developed using generative AI for eliciting input parameters and code generation?”

## METHODS

A three-stage replication workflow was applied to existing health-economic models in the field of Alzheimer’s disease. In the first stage, a chain-of-thought prompting strategy in ChatGPT-4 was developed to replicate in R an open-source model co-developed by one of the authors [RH] and with publicly available code. In the second stage, it was applied to replicate a model also co-developed by one of the authors [RH] but without publicly available code. In the third stage, it was applied to a model developed independently of the authors and without publicly available code. A mixed-methods approach was employed in terms of qualitatively addressing the face validity of the prompt development and refinement and quantitatively assessing deviations between AI-generated and original model predictions.

All prompts, scripts, and decision points are documented in detail and generated data are available in supplementary material. By lack of existing reporting guidelines for our study design we considered the PALISADE checklist (see supplementary material 1), originally designed for machine learning models in health economics [^14^], most appropriate.

### FIRST MODEL SELECTION AND DEVELOPMENT OF PROMPT SEQUENCE

As a starting point, the International Pharmaco-Economic Collaboration on Alzheimer’s Disease (IPECAD) open-source model [^15^] was selected for replication using ChatGPT-4. The IPECAD model is a cohort-based Markov model that reflects disease progression across five health states (mild cognitive impairment, mild, moderate, severe dementia, and death) and two care settings (community and institutional). It operates on annual cycles using transition probabilities from Clinical Dementia Rating (CDR) data, with treatment initiation, discontinuation, and waning effects explicitly modeled. The IPECAD model was developed to support reimbursement decision-makers with an independent modeling framework for early Alzheimer’s disease treatments (e.g., lecanemab) to allow cross-validation of models used in technology appraisal. To this end, the authors first developed the IPECAD model and assessed its internal validity using features from the Institute for Clinical and Economic Review (ICER) model. It was then cross-validated against the Alzheimer’s Disease Archimedes Condition-Event Simulator model. Additional uncertainty scenarios were also explored.

This model was considered ideal for an initial test case, as both its open-access publication and its source code published on GitHub are freely available. The IPECAD publication is intentionally structured with a high level of transparency, clearly detailing the model’s structure, assumptions, and all input parameters. We expected this level of transparency would facilitate the initial development and refinement of the prompt sequence, as the known parameters, outputs, and the original code could be directly compared to the AI-generated output.

A chain-of-thought prompting strategy was used to interact with ChatGPT-4. First, to guide the decomposition of the complete Markov model into small and manageable prompts, we reviewed the IPECAD publication, its supplementary material, and the associated R source code beforehand. We identified key components from the model structure, logic, inputs, and outputs that could be translated into individual prompt objectives. Each of these components were then used to design a corresponding prompt with the ultimate goal that the full prompt sequence would reconstruct the IPECAD model. Second, each prompt was developed alongside a qualitative validation criterion defining what would constitute a correct and complete response. This was based on reproducing the logic of the original code rather than matching its exact syntax, as we expected alternative code could produce the same results. Prompts were refined iteratively, as shown in Figure 1. After submitting each prompt, if the response failed to meet its predefined validation criteria, the prompt was refined and retested until it did. To support this refinement process, in addition to the publication and its source code, insights from ChatGPT-4 itself within the same temporary chat session were also used. These insights were obtained through interaction, such as prompting: ”How can I improve my initial prompt to ensure that you include this variable next time?”. Third, to ensure that each prompt in the sequence would reliably produce the same result for any researcher, once a prompt met its predefined validation criteria, it was tested in five new and temporary chat sessions. Temporary chats were used to ensure ChatGPT-4 did not learn (i.e., remember) from prior prompts. If inconsistencies emerged, the prompt was further refined. Because all prompts were linked in a sequential manner, validating a specific prompt (e.g., prompt 3) required executing the entire sequence from prompt 1 through prompt 3 in each session. Fourth, once the complete sequence of prompts met their respective validation criteria in five temporary chat sessions, the sequence was no longer refined and was tested ten additional times in new temporary chat sessions. These fifteen final trials then served as the basis for evaluating the reproducibility and validity of the AI-generated R code as detailed below.

**Figure 1:**
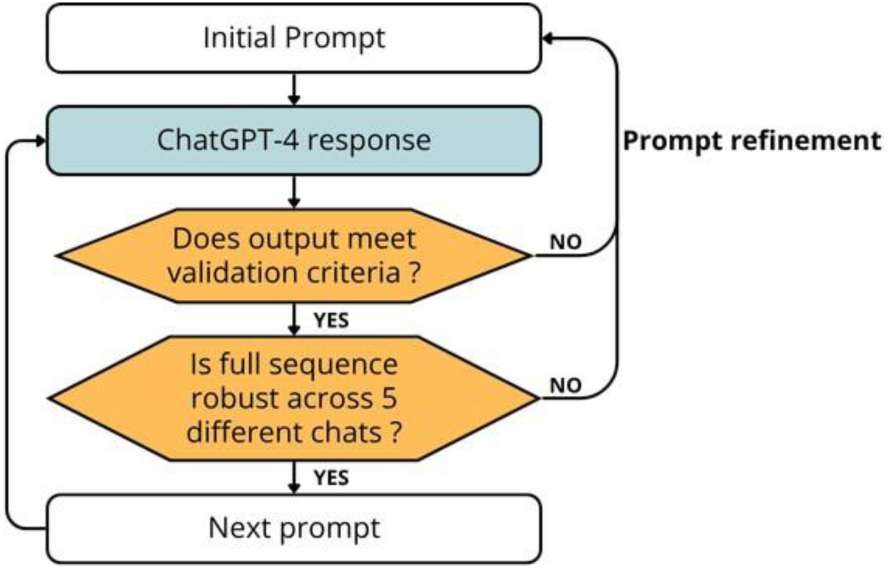
Iterative development and validation process for each prompt in the sequence.

### APPLICATION TO A SECOND MODEL

The earlier developed prompt sequence was applied to a second model developed by Wimo et al. (2020) [^16^]. This model described the natural history and predicted the potential societal costs of Alzheimer’s disease across its disease continuum, as well as assessed the cost effectiveness of a hypothetical disease-modifying treatment (DMT). To this end, the authors used data from the Swedish dementia registry to construct a Markov model simulating a cohort of 100,000 individuals with mild cognitive impairment due to Alzheimer’s disease, starting at age 60 and followed over a 40-year horizon. The intervention scenario assumed a 25% reduction in disease progression.

This model was considered ideal as an initial validation case, as it was co-developed by one of the authors [RH] but did not provide public access to its code or a detailed table of input parameters. This allowed us to test the earlier developed prompt sequence on a less documented case and validate its transferability (i.e., its ability to be adapted and applied to a different model than the one on which the prompt sequence was originally developed).

The earlier developed ChatGPT-4 prompt sequence and validation criteria were adjusted to the second model. First, we reviewed the publication and its supplementary material to identify key differences between the two models. Second, each IPECAD prompt was individually adapted and its validation criterion to reflect these differences. When information was missing in the Wimo et al. (2020) publication, reasonable assumptions were made to maintain logical consistency and completeness. Instead of testing each prompt individually across five temporary chat sessions as done with IPECAD (see Figure 1), the entire adapted sequence was tested as a whole. We reviewed all responses and, when necessary, iteratively refined prompts.

Third, once the full prompt sequence produced correct responses matching all validation criteria across five different temporary chat sessions, it was considered as finalized and no further refinements were made. Fourth, as in the IPECAD replication process, ten additional trials were conducted in temporary chat sessions, and the resulting R scripts and outputs were collected for analysis.

### APPLICATION TO A THIRD MODEL

The further developed prompt sequence was applied to a third model developed by Ross et al. (2022) [^17^]. This model was designed to predict the cost-effectiveness of two anti amyloid monoclonal antibody treatments compared to standard care for early Alzheimer’s disease in the United States. The authors implemented a Markov model in spreadsheet software, using clinical trial data to simulate lifetime health and economic predictions for adults with early-stage AD, from both the United States healthcare sector and societal perspectives. The model used a monthly cycle length over a lifetime horizon and included three treatment strategies: (1) standard care, (2) aducanumab, and (3) donanemab, along with multiple sensitivity analyses and predicted price thresholds for cost-effectiveness.

This model was considered ideal as an initial validation case, as it is entirely independent from the authors and the previous two models. While no public code was available, a full table of input parameters was provided in the model publication. This third replication allowed further testing of the replication workflow’s transferability to an unrelated model.

The replication of this model followed the same steps as for the replication of the previous model by Wimo et al., using the final prompt sequence and validation criteria as a foundation and adjusting them to reflect differences between the two models.

### DATA COLLECTION AND ANALYSIS

A qualitative summary of the prompt development and refinement was generated for all three models (i.e., the initial prompt sequence development using the IPECAD model and the further application to the two other models by Wimo et al. and Ross et al.). Key differences between the three models (IPECAD to Wimo, and Wimo to Ross) were also described. All prompt refinements, prior to meeting validation criteria across five sessions, are provided in supplementary material 2 and final prompt sequences and their validation criteria are presented in supplementary material 3. In addition, the 15 final R scripts, generated from temporary chat sessions and compiled into a single executable R file, are available in supplementary material 4.

For each replicated model, its ChatGPT-4 generated R code was run to obtain quality-adjusted life years (QALYs), costs and a variety of additional outcomes. For the IPECAD model the additional outcome was its life-years. For the model by Wimo et al. the additional outcomes were person-years in each dementia state, total cumulative deaths and deaths. For the model by Ross et al. no additional outcomes were generated. To assess the reproducibility of the original published results by the ChatGPT-4 generated R code, the absolute and relative deviations between the original published model predictions and ChatGPT-4 based model predictions were calculated for each strategy, each metric and each of the fifteen simulations. Relative deviations were expressed as the percentage difference to the original published value. Deviation was summarized by the mean deviations, the minimum and maximum to indicate potential outliers, and the standard deviation and median as an overall measure of spread, with the latter less sensitive to outliers. All formulas used for the descriptive statistics are provided in supplementary material 5.

## RESULTS

### QUALITATIVE RESULTS

Three sets of ChatGPT-4 based prompt sequences were generated, one sequence for the IPECAD model, one sequence for the model by Wimo et al. and one sequence for the model by Ross et al.; all with a variety of refinements (see supplementary material 2).

Overall, no general refinement pattern was identified, as adjustments were relatively specific to each prompt’s objective. However, a key insight was the importance of balancing the level of detail and the prompt’s length. ChatGPT-4 generally performed more reliably when receiving multiple shorter, well-structured prompts rather than a single long one. Larger prompts often led to overlooked instructions or incomplete outputs. Therefore, the approach observed as the most effective was the following: when ChatGPT-4 failed to use, extract or define a specific parameter, that element was explicitly mentioned in the prompt (e.g., ”do not forget to extract mortality probabilities from the lifetable_US_2019_ssa.csv file”). If this adjustment worked, the prompt was retained. If not, the issue was often due to excessive length, and the prompt was then either shortened or split into two. In this context, testing each prompt across several temporary chat sessions proved essential, as even prompts that worked consistently two or three times could fail in subsequent runs. Another key observation was that ChatGPT-4 accurately extracted numeric values from the publication, except when values were embedded within formulas such as a cost calculation. In these cases, it sometimes returned incorrect values. This was resolved by explicitly asking ChatGPT-4 to preserve original formulas. Refinements took up to 39 times for the IPECAD model, up to 13 times for the model by Wimo et al. and up to 11 times for the model by Ross et al. The first prompt sets the context, task instructions, and global instructions for the rest of the prompt sequence. This overview prompt seemed to help reducing the length of subsequent prompts by preventing repeated instructions and ensured consistency across prompts by reminding ChatGPT-4 to reuse the same variable names and avoid redefining parameters already introduced earlier in the sequence.

Specifically for the IPECAD model prompt 5 was split into four sub-prompts (5A–5D) and builds the full transition probability matrix (A), defines transitions from on- and off-treatment states (B and C), and transitions to institutionalized states (D); all relatively specific to the model structure. Validation involved manual comparison with the IPECAD reference code. Moreover, an R script was implemented and computing the absolute difference between each element of the transition array generated by ChatGPT-4 and the reference array from the IPECAD model. Any element with a difference exceeding 10% was flagged. This automated check provided a fast and systematic way to identify problematic transitions and refine the corresponding prompts.

Specifically for the model by Wimo et al. the main differences compared to the IPECAD model were a smaller number of health states, no institutional setting, and no treatment discontinuation or waning effects. Utility input values were only available by health state and five-year age intervals, cost input values were only available in graphical form, and no life table was available. Therefore, these input estimates were manually pre-processed to and fed into a prompt for ChatGPT-4. See supplementary material 6 for details.

Specifically for the model by Ross et al. it included three strategies instead of two, a one-month cycle length, treatment discontinuation, waning effects, explicit modeling of amyloid related imaging abnormality related adverse events, only forward transitions. It included manual preparation of a U.S. life table. See supplementary material 6 for details.

### QUANTITATIVE RESULTS

Reported QALYs and costs predictions of all three models (also referred to as ‘reference values’), as well as the generated QALYs and costs predictions by each of the 15 ChatGPT-4 based replicated model R code are presented in Figure 2. QALYs generated for the IPECAD model tend to slightly overestimate the original values, while most of the other replications across all models and metrics show an underestimation of the reference values. Overall, the tendency for QALYs and costs to be higher under the intervention strategy, compared to no intervention or standard of care, is consistently observed across most replications.

**Figure 2:**
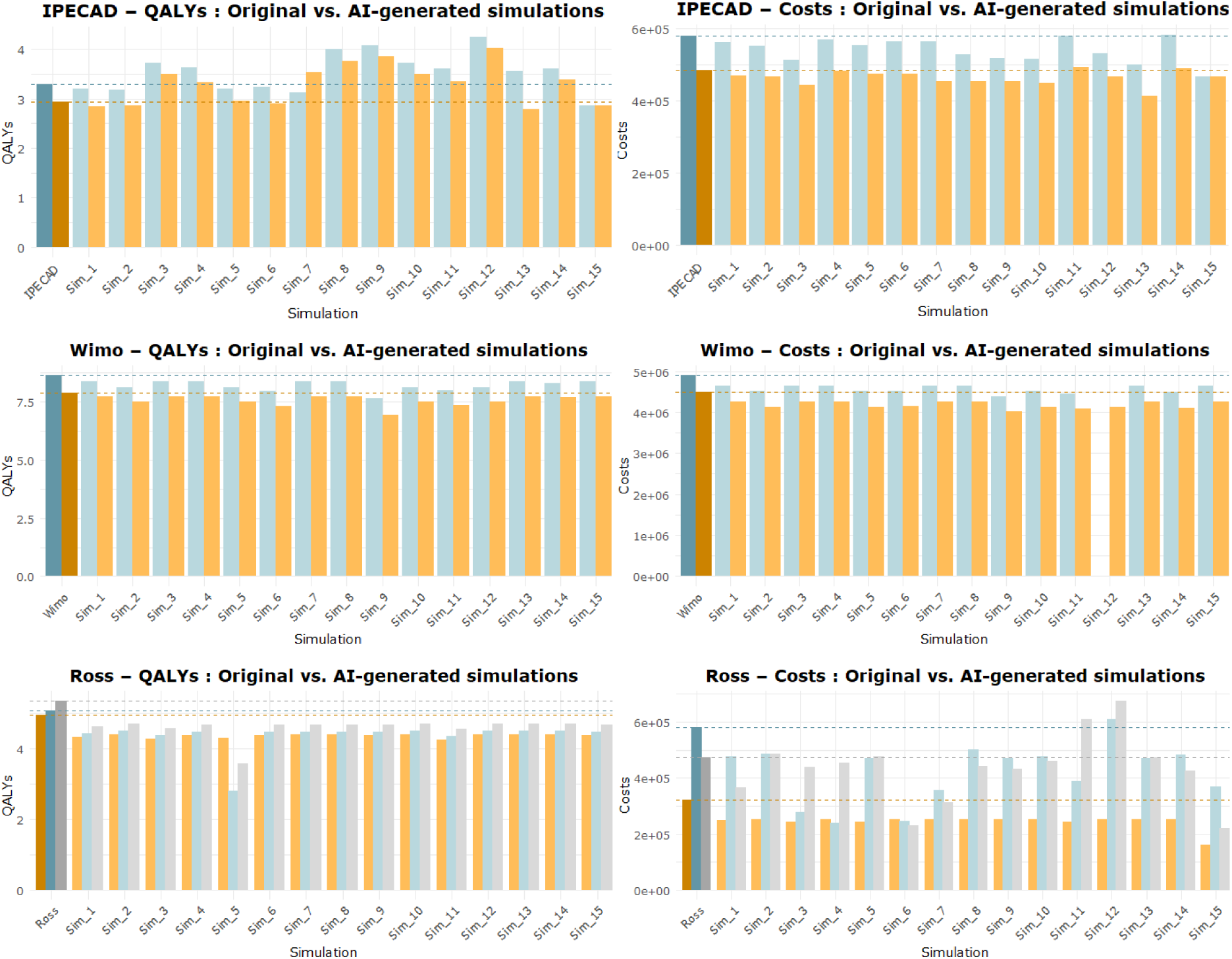
Comparison of originally reported model predictions and ChatGPT-4 generated simulation results across all 15 trials for QALYs (left) and costs (right), by strategy and model. The first darker bars as well as the dashed horizontal lines indicate the original published model values. Strategies are represented as follows: orange = standard of care; blue = intervention; and gray = intervention 2 (only in the model by Ross et al.). Costs are expressed in U.S. dollar (IPECAD and model by Ross et al.) and Swedish Krona (model by Wimo et al.).

Table 3 summarizes statistics calculated for the absolute and relative deviations between the original published model outputs and those generated by ChatGPT-4 across 15 replications for each of the three health-economic models. Overall, IPECAD showed lower deviations for costs than for QALYs, whereas Wimo and Ross displayed the opposite pattern. Across all models, Wimo had the lowest mean deviations, followed by IPECAD, and then Ross. Individual results are presented in supplementary material 7.

**Table 3:**
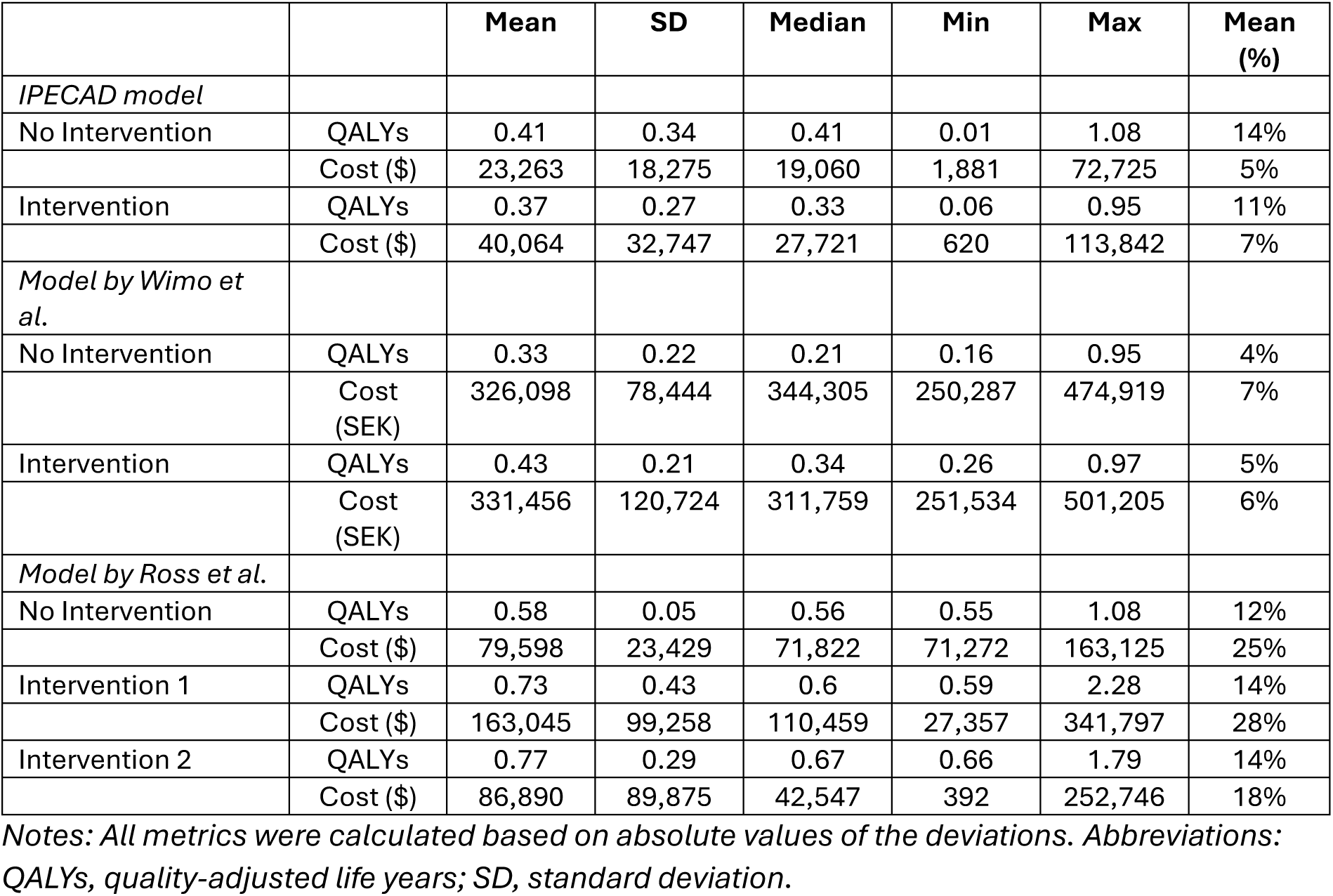
Summary statistics of the 15 absolute deviations between originally published and ChatGPT-4 based replicated model predictions across the three health-economic models. Last column presents relative deviation in terms of deviation relative to the original model predictions.

|  |  | Mean | SD | Median | Min | Max | Mean (%) |
| --- | --- | --- | --- | --- | --- | --- | --- |
| *IPECAD model* |  |  |  |  |  |  |  |
| No Intervention | QALYs | 0.41 | 0.34 | 0.41 | 0.01 | 1.08 | 14% |
| | Cost (\$) | 23,263 | 18,275 | 19,060 | 1,881 | 72,725 | 5% |
| Intervention | QALYs | 0.37 | 0.27 | 0.33 | 0.06 | 0.95 | 11% |
| | Cost (\$) | 40,064 | 32,747 | 27,721 | 620 | 113,842 | 7% |
| *Model by Wimo et al.* |  |  |  |  |  |  |  |
| No Intervention | QALYs | 0.33 | 0.22 | 0.21 | 0.16 | 0.95 | 4% |
|  | Cost (SEK) | 326,098 | 78,444 | 344,305 | 250,287 | 474,919 | 7% |
| Intervention | QALYs | 0.43 | 0.21 | 0.34 | 0.26 | 0.97 | 5% |
|  | Cost (SEK) | 331,456 | 120,724 | 311,759 | 251,534 | 501,205 | 6% |
| *Model by Ross et al.* |  |  |  |  |  |  |  |
| No Intervention | QALYs | 0.58 | 0.05 | 0.56 | 0.55 | 1.08 | 12% |
| | Cost (\$) | 79,598 | 23,429 | 71,822 | 71,272 | 163,125 | 25% |
| Intervention 1 | QALYs | 0.73 | 0.43 | 0.6 | 0.59 | 2.28 | 14% |
| | Cost (\$) | 163,045 | 99,258 | 110,459 | 27,357 | 341,797 | 28% |
| Intervention 2 | QALYs | 0.77 | 0.29 | 0.67 | 0.66 | 1.79 | 14% |
| | Cost (\$) | 86,890 | 89,875 | 42,547 | 392 | 252,746 | 18% |

*Notes: All metrics were calculated based on absolute values of the deviations. Abbreviations: QALYs, quality-adjusted life years; SD, standard deviation.*

## DISCUSSION

A step-by-step chain-of-thought prompting workflow was developed and tested on three published models for Alzheimer’s disease with varying complexity and available details in their publications, allowing evaluation of the method’s robustness and transferability. Results showed deviation between originally published model predictions and replicated model predictions between 4-28%.

ChatGPT-4 generated functional R code across fifteen independent trials (i.e., temporary chat sessions) per model using the same prompt sequence, with a maximum of three human interventions per model. These interventions mainly consisted of correcting recurring errors. For example, for the IPECAD model replications, ChatGPT-4 misspelled the name of a health state (3 times). For the model by Wimo et al., the discounting vector occasionally had an incorrect length (2 times). For the model by Ross et al., the current age variable in the transition probability matrix was not properly bounded between ages 60–99, causing errors in age-specific mortality extraction (3 times). The recurrence of these similar issues for a specific model suggests that further refining the responsible prompts potentially could eliminate manual fixes altogether.

A key insight was that multiple shorter prompts proved more effective than a single long one. Additionally, testing them across several chat sessions was crucial to reveal occasional failures missed in a single trial. The learnings from developing prompt sequence on the first model likely resulted in less modifications required when further developing the prompt sequence on the other two models.

For the IPECAD model, the mean absolute relative deviation from the original published model predictions across all fifteen trials was ≤ 14% for both strategies and outcomes (QALYs and costs). The exact causes of these deviations remain unknown, since not all fifteen AI-generated R scripts were reviewed and compared for this. Nevertheless, a potential explanation could be that the original IPECAD model source code was available and helped refine the prompts, their length was constrained to ensure ChatGPT-4 fully leveraged each detail requested. Additionally, the overall aim was also to keep the prompt sequence relatively short and time-efficient, to allow broader usability for researchers with limited time or expertise. In comparison, the IPECAD model was originally developed as a manual replication of the ICER model [^15, 18^], which reported a deviation of up to 6% in QALYs, up to 7% in life years and up to 27% in costs. Our deviation up to 14% is within that range.

For the model by Wimo et al., the mean absolute relative deviation was ≤ 7% across all strategies and outcomes, which was lower than for IPECAD. Although no source code was available, the model by Wimo et al. was built using similar logic to IPECAD as developed by the same authors and precursor to IPECAD. The model also had a more simple structure with fewer health states (i.e., no separate states for care setting and treatment discontinuation) and no waning effects.

For the model by Ross et al., the mean absolute relative error was higher, at ≤ 28% across strategies and outcomes. This is likely because the model is independent from the other two and included more complexity in terms of treatment discontinuation, amyloid related imaging abnormalities, and diverse cost components.

To compare with other replication efforts, Schwander et al. (2021) [^3^] manually replicated four Markov models for obesity. As experienced modelers with over 20 years in the field, they reported QALY deviations ranging from –4% to 2% (mean absolute of 0.1%) and cost deviations from –4% to 16% (mean absolute of 4%). Compared to the AI-assisted reproductions in this study (mean absolute relative error ≤ 14%, 7% and 28%), we consider Schwander’s results to represent an upper-bound benchmark for manual replication, highlighting that even expert human efforts are subject to deviation. This could be besides other reasons due to limited reporting of model details.

Three conference abstracts report on the use of AI to replicate health-economic models. Chhatwal et al. (2024) [^12^] reproduced a simple HIV/AIDS example from a textbook, reporting errors of 8% in costs, 0.1% in life years, and 2% in the ICER. Chhatwal et al. (2024) [^11^] constructed a de novo health-economic Markov model for hepatitis C treatment and compared their results to expert opinion and published studies. They reported overall face validity and demonstrated the feasibility of their approach but did not report empirical results in their conference abstract. Chhatwal te al. (2025) [^13^] replicated the ICER model (same as replicated by the IPECAD model) using GPT-4-based platform with multi-agent pipelines and Python to facilitate large language model interactions. They reported that despite limitations (general population costs not explicitly reported, and absence of cited references to restrict AI based extraction) their AI-based model replication deviated 5% in incremental QALYs and 14% in incremental costs.

### STRENGTHS AND LIMITATIONS OF THE STUDY

The main strength of this study lies in its high level of transparency. All prompts, validation rules, and refinement steps are fully reported, along with key qualitative and quantitative findings, facilitating future use. Additionally, its approach is simple (using only ChatGPT-4 public interface, subscription-based and R open source software) and required limited time and experience as the results were produced during a 3-month master internship health-economics [FG] under supervision of one experienced modeler [RH].

The study also has several limitations. Expertise on health-economic modeling, R programming skills and the specific Alzheimer’s disease treatment setting is required to correct errors and identify causes of deviation with original model predictions. Also, several validation steps relied on manual inspection, introducing an element of subjectivity that depends on the expertise of the author may have an impact on reliability. Only the final AI-generated model predictions were analyzed and compared, while the actual content of the validated R scripts was not systematically reviewed once all validation criteria were passed. Finally, only the third model by Ross et al. was fully independent from the authors and without their code open source available to our knowledge, which makes the generalizability of our findings mainly rely on the results of replicating this single study.

### RECOMMENDATIONS FOR PRACTICE OR POLICY

Replicating health-economic models is essential to ensure the transparency, quality, and credibility of their results [^6^]. The accessible chain-of-thought workflow proposed here could be valuable for both HTA bodies, pharmaceutical companies, and researchers.

For HTA bodies, our AI-based approach could facilitate reconstruction of company-submitted or existing models to allow cross-comparison of a submitted model to a published model e.g., with a different structure or assumptions.

For pharmaceutical companies, it could accelerate the development of their health-economic model for reimbursement dossiers. For researchers, it could be used to replicate existing models testing their cross-validity or reporting transparency of existing model publications.

Regarding the potential uncertainty around the use of AI in HTA, NICE (United Kingdom) is currently the only HTA agency with a formal position on this topic [^19^] acknowledging its potential benefits in HTA. However, it emphasizes that its clear added value must be justified, that AI should augment but not replace human expertise, and that potential biases and risks must be transparently reported with tools like the PALISADE checklist [^14^]. Other agencies such as ZIN (Netherlands) and ICER (United States) have not yet issued AI-specific guidance [^20, 21^].

### RECOMMENDATIONS FOR FUTURE RESEARCH

Given the reported errors, future research should focus on understanding the causes of deviations between original model predictions and replicated model predictions. This could involve analyzing and comparing the AI-generated scripts by an experienced modeler to determine where they deviate, to identify why some produced better results than others and refine prompts where needed.

A second direction for future research could be to reproduce the entire prompt sequence by an independent research group. This would help mitigate the subjectivity introduced by the validation criteria, which currently rely heavily on the author’s interpretation and technical skills, and could help determine whether the observed deviations can be further reduced.

## CONCLUSION

This study presents a novel chain-of-thought prompting workflow, which uses generative AI to replicate three published health-economic models in the field of Alzheimer’s disease. It provides a first step toward making model replication more accessible for HTA bodies, pharmaceutical companies and researchers, supporting greater transparency and validation in health-economic evaluations.

## Supporting information

final R scripts

## Data Availability

All data produced are available as supplementary material.

## STATEMENTS AND DECLARATIONS

## ACKNOWLEDGMENTS

The authors acknowledge the use of AI tool ChatGPT-4 to replicate health-economic models as detailed in the methods section. The authors acknowledge the use of AI tool ChatGPT-4 to assist in drafting the manuscript. The content has been reviewed and edited by the authors and they take full responsibility for it.

## ETHICS STATEMENT

This study did not involve any human participants or animals. All data used were from publicly available sources and did not contain identifiable personal information.

## AUTHOR CONTRIBUTIONS

Concept and design: FG, RH.

Statistical analysis: FG.

Interpretation of outcomes: FG, RH, SO.

Drafting the manuscript: FG.

Critical revision of the final manuscript: FG, RH, SO.

## DATA AVAILABILITY

Data are included as supplementary material.

## SUPPLEMENTARY MATERIAL 1: PALISADE CHECKLIST

**Table S1.1:** Application of the PALISADE Checklist developed by the ISPOR Task Force.

| Element | Definition | Application to this research |
| --- | --- | --- |
| Purpose | Is the purpose of the algorithm clearly stated at the outset? Is the implementation of the algorithm in a healthcare setting fair and ethical? | The aim to replicate three published health-economic models using ChatGPT-4 (introduction). The application of ChatGPT-4 for model replication is fair and ethical, as it relies exclusively on open-access materials. |
| Appropriateness | Is there a clear justification that the algorithm is acceptable in the context within which it is being applied? | The use of ChatGPT-4 in the context of health-economic model replication is justified by its potential to accelerate the replication process, improve accessibility, and promote greater transparency in research (introduction). |
| Limitations | Have the strengths and limitations, in the context of the purpose, been identified? This should cover both the algorithm and any data used. | Strengths and limitations of using ChatGPT-4 are described (discussion). |
| Implementation | Consideration of access, implementation, and resource issues when implemented in healthcare settings. | All resources used (e.g., ChatGPT-4 and R) are publicly accessible. Implementation details (e.g., session setup, prompt structure, validation protocol, use of temporary chats) are described (methods) and the complete material is available (supplementary material). |
| Sensitivity & Specificity | For classification algorithms, has the model performance and accuracy (specificity and sensitivity) been appropriately evaluated? | Not applicable, as this study does not involve a classification model. |
| Algorithm characteristics | Has the ML mechanism been clearly characterized and described? Is there sufficient transparency for the results to be reproducible? | Prompting sequences, validation criteria, and iteration tracking ensure transparency and reproducibility (methods and results). |
| Data characteristics | Is the selection of data sets justified and are the key characteristics known? This should extend to training sets, test sets and validation sets. | All models replicated rely solely on published articles and publicly available information (i.e., IPECAD model R code). No training/test split is applicable, but the 15-session replication protocol assesses consistency and reproducibility (methods and results). |
| Explainability | Are the outputs of the algorithm clearly understandable by both the healthcare professional and the patient? | All final model outputs (e.g., QALYs, life years, costs, person years) are documented and interpreted (results and discussion). |

## SUPPLEMENTARY MATERIAL 2: PROMPT REFINEMENTS

**Table S2.1:** Model by IPECAD: Overview of prompt 1 and 2 refinement process and rationale for adjustments.

| Trial | Prompt Evolution | Main Limitation | Reason for modification |
| --- | --- | --- | --- |
| 1 | "Please extract all input parameters required to code the Markov model from the IPECAD and its additional material PDF and present them in a structured format suitable for building a Markov model to code in R." | Only table output and Python code provided. | Specify that an R-specific script is required. |
| 2 | "Please extract [...] present them **in a code in R.**" | R script generated, but not downloadable or usable. | Clarify format expectations for the R output. |
| 3 | "Please extract [...] present them **in a ready-to-run R script structure.**" | Missed state names, zero values, and life table. | Clarify full input requirements and attach the *US 2019 life table*. |
| 4 | "Please extract all input parameters required to code the Markov model from the IPECAD publication, its supplemental material **and the 2019 US-specific life table, including model states and without omitting parameters with a zero value.** Present the output in a ready-to-run R script structure." | Incorrect cost values. | Need to find why these values are incorrect. |
| 5-11 | <u>Several cases tested:</u> Prompt emphasized ICER scenario only, no merging with AD-ACE, no simplifications or substitution of data from other scenarios, etc. | Incorrect cost values. | Need to explicitly require preservation of original formulas for costs. |
| 12 | "Please extract all input parameters required to code the IPECAD Markov model from the IPECAD publication and its supplemental material. Include all model states. Do not omit any parameters, even if their value is zero. **When a parameter is computed from a formula, preserve the original formula in the R script rather than just showing the final numeric value.** Present the output as a ready-to-run R script." | Mortality rates not computed from the *US 2019 life table*. | Clarify mortality rate calculation. |
| 13-15 | Several formulations tested to prompt GPT-4 to convert probabilities into mortality rates using the life table, e.g. "Convert probabilities of death from the 2019 US life table into mortality rates..." | Missed some others input parameters. | Need a clear separation between input parameters and mortality rate prompts. |
| 16-20 | **Prompt 1:**<br>"Please extract all input parameters required to code the IPECAD Markov model from the IPECAD publication and its supplemental material. Include all model states and strategies. Do not omit any parameters, even if their value is zero. When a parameter is computed from a formula, preserve the original formula in the R script rather than just showing the final numeric value. Displays the full R script directly in the response (don't put it in a downloadable file)." | Missed the `dampack` package and included unused states by the publication. | Specify expected package (`dampack`) and state names. |
|  | **Prompt 2:**<br>"Please extend the previous R script by adding a fully functional, copy-paste-ready code block that loads the *2019 US life table* from the CSV file named "lifetable_US_2019_ssa.csv" and creates a mortality matrix with the columns "age", "male", "female", and "weighted". Assume the file is in the working directory. The result should be a complete R code block that I can directly append to my existing script." | CSV column name assumptions led to errors and added unnecessary packages. | Specify the required column headers and prohibit package additions.<br>The prompt can be shortened due to the addition of an overview prompt. |
| F<br>I<br>N<br>A<br>L | **Prompt 1:**<br>"Please extract all input parameters required to code the IPECAD Markov model **using the dampack package, based on the content** of the IPECAD publication and its supplemental material. Include all **variables, including descriptive ones (e.g., state names and strategy names) directly as named R objects in the script. Restrict state names to the eleven that are actually used in the publication** ("mcion_c", "mciof_c", "milon_c", "milof_c", "mod_c", "sev_c", "mci_i", "mil_i", "mod_i", "sev_i", "dth"). Do not omit any parameter, even if its value is zero. If a parameter is derived from a formula, preserve the original formula in the R script rather than just showing the final numeric value. Display the full R script directly in the response (do not put it in a downloadable file)."<br>**Prompt 2:**<br>"Please **write a R code** that loads the *2019 US life table* from the CSV file named *"lifetable_US_2019_ssa.csv"*. **The CSV contains the following columns: "age", "male", and "female", which represent age-specific mortality probabilities for males and females, respectively. First, convert these annual mortality probabilities to annual mortality rates. Then, use these columns to compute a weighted averaged of these mortality rates and place them into a new column "weighted". Return a matrix or dataframe with columns: "age", "male", "female", and "weighted", where "male" and "female" contain the converted rates (not the original probabilities). Do not add any other packages. Assume the file is in the working directory.** |  |  |

**Table S2.2:**
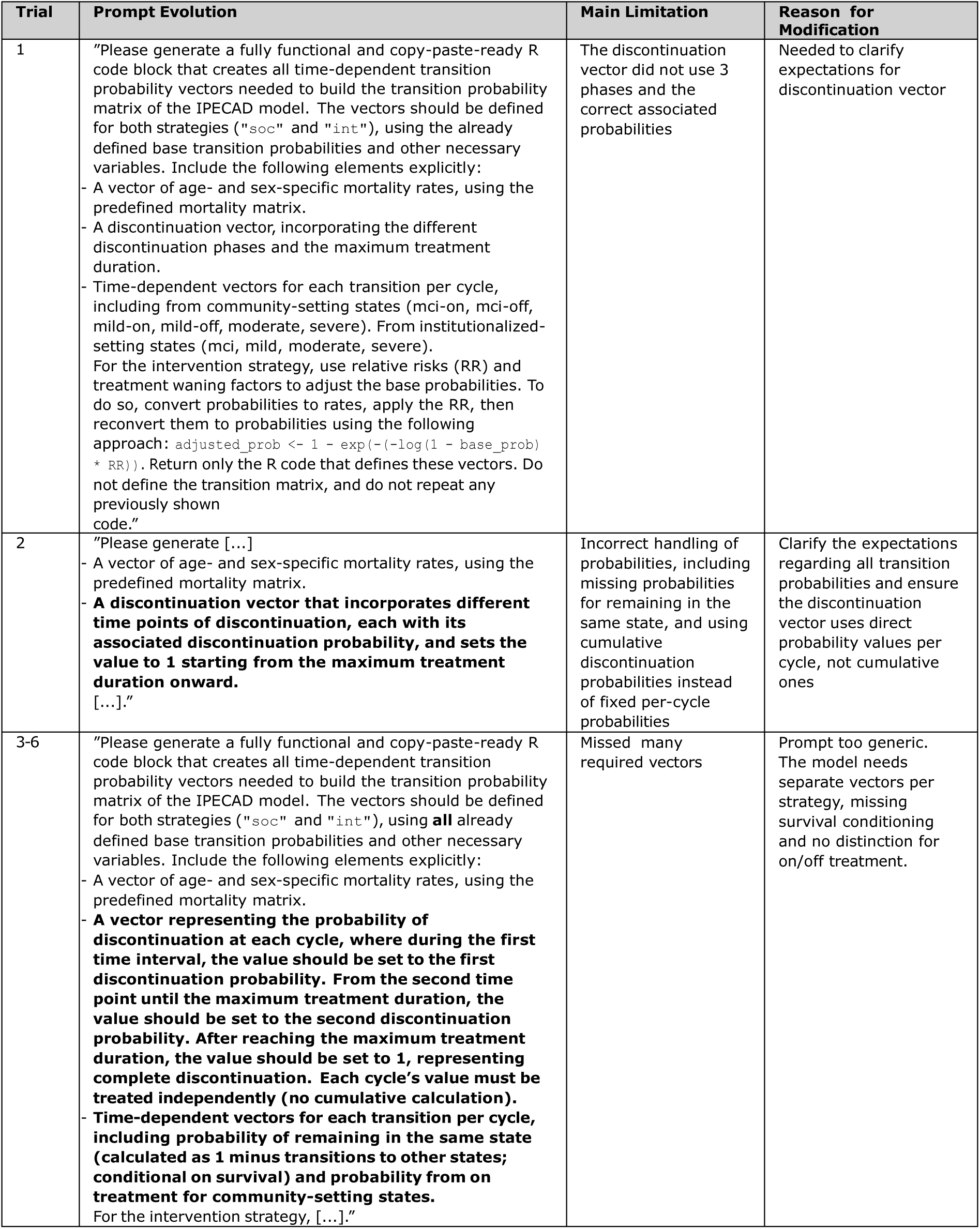
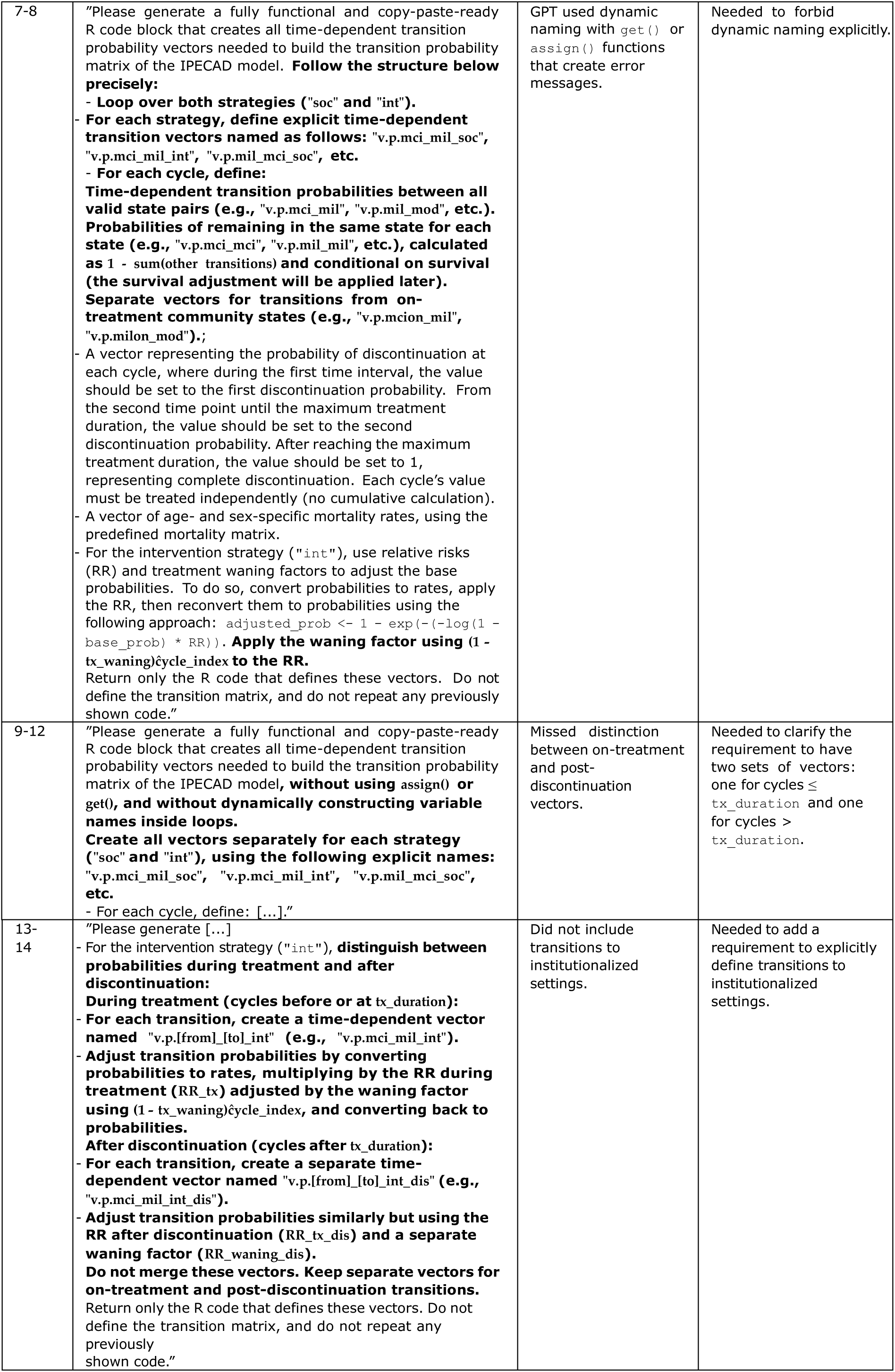
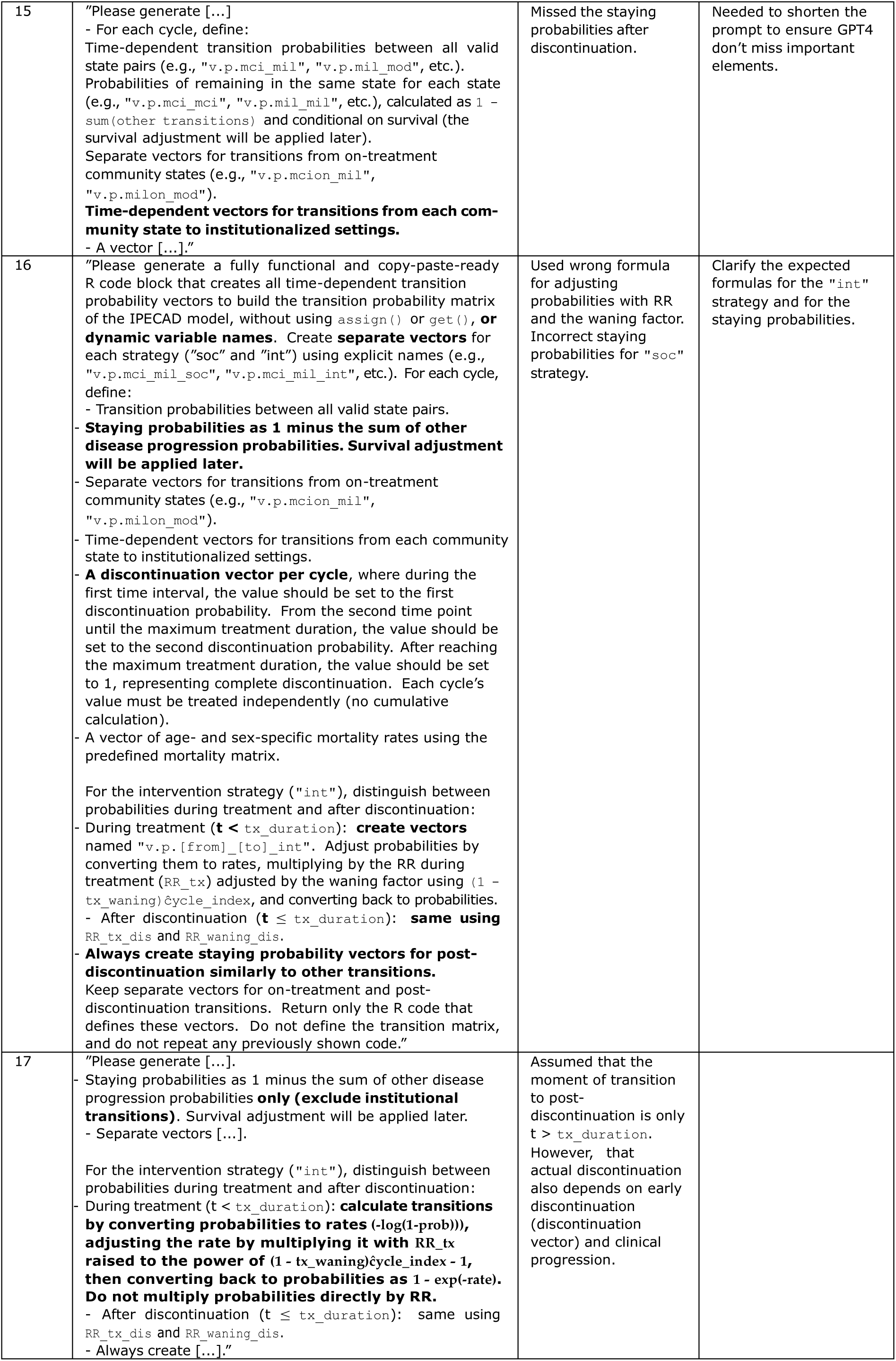
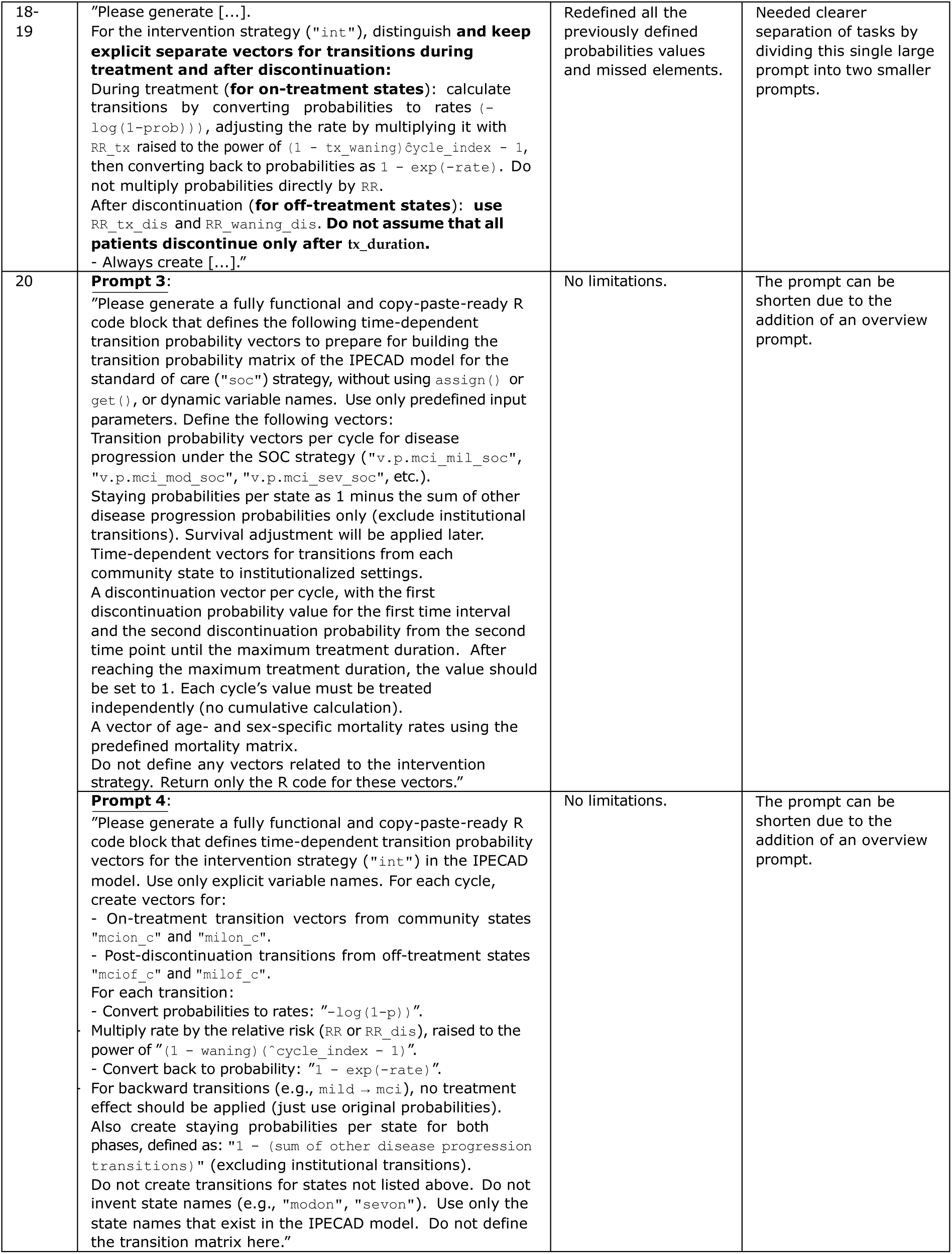
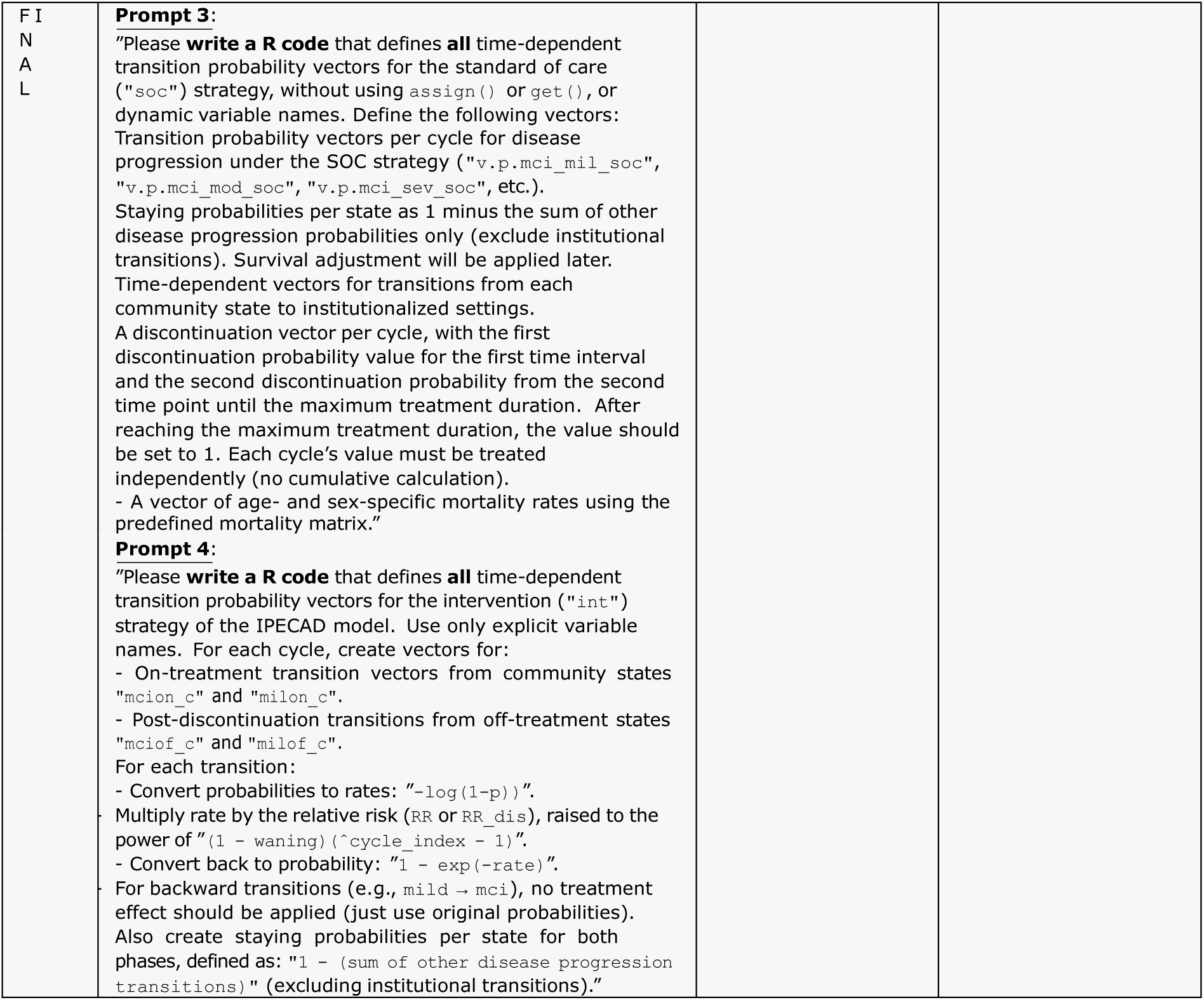
Model by IPECAD: Overview of prompt 3 and 4 refinement process and rationale for adjustments.

**Table S2.3:**
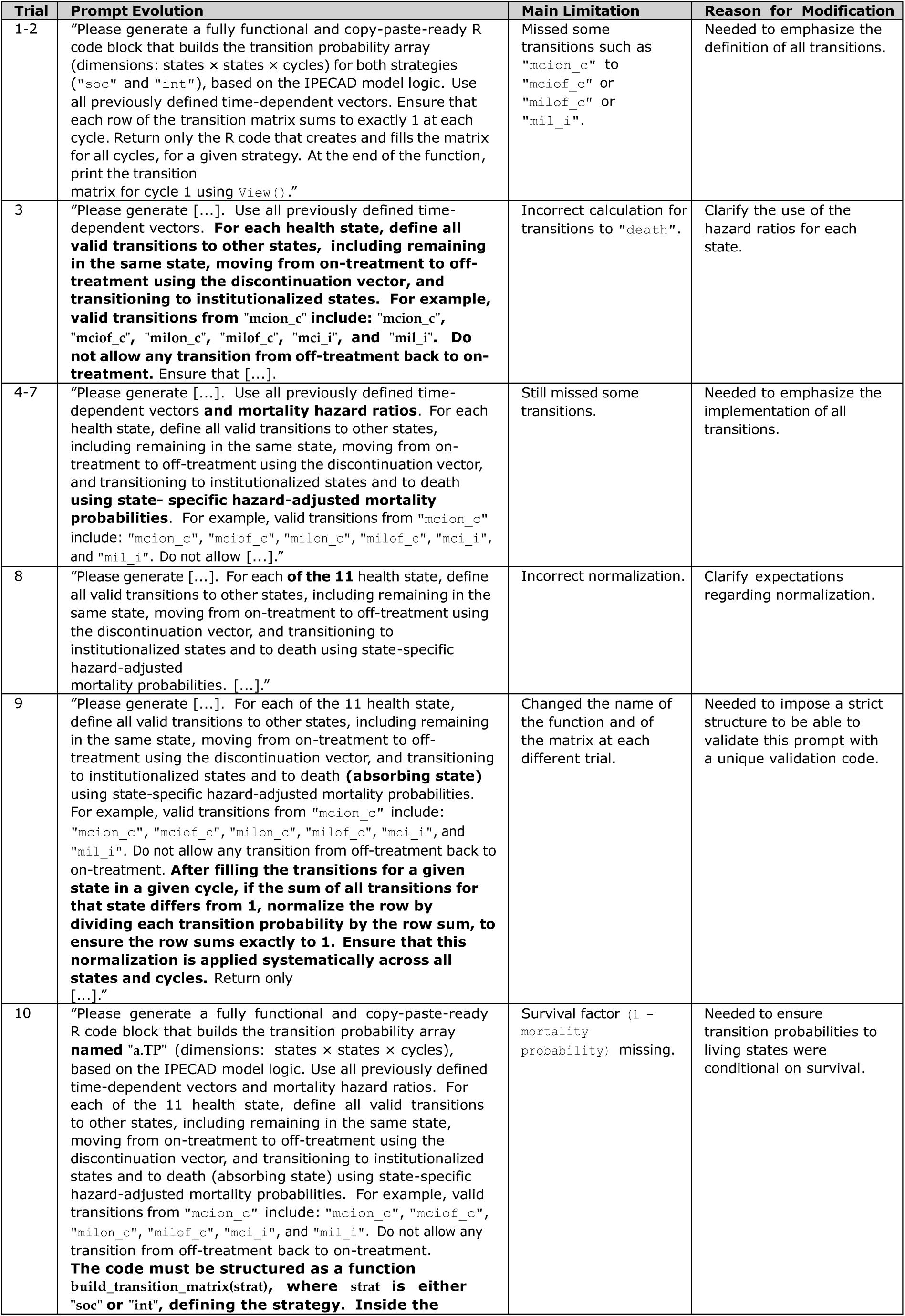
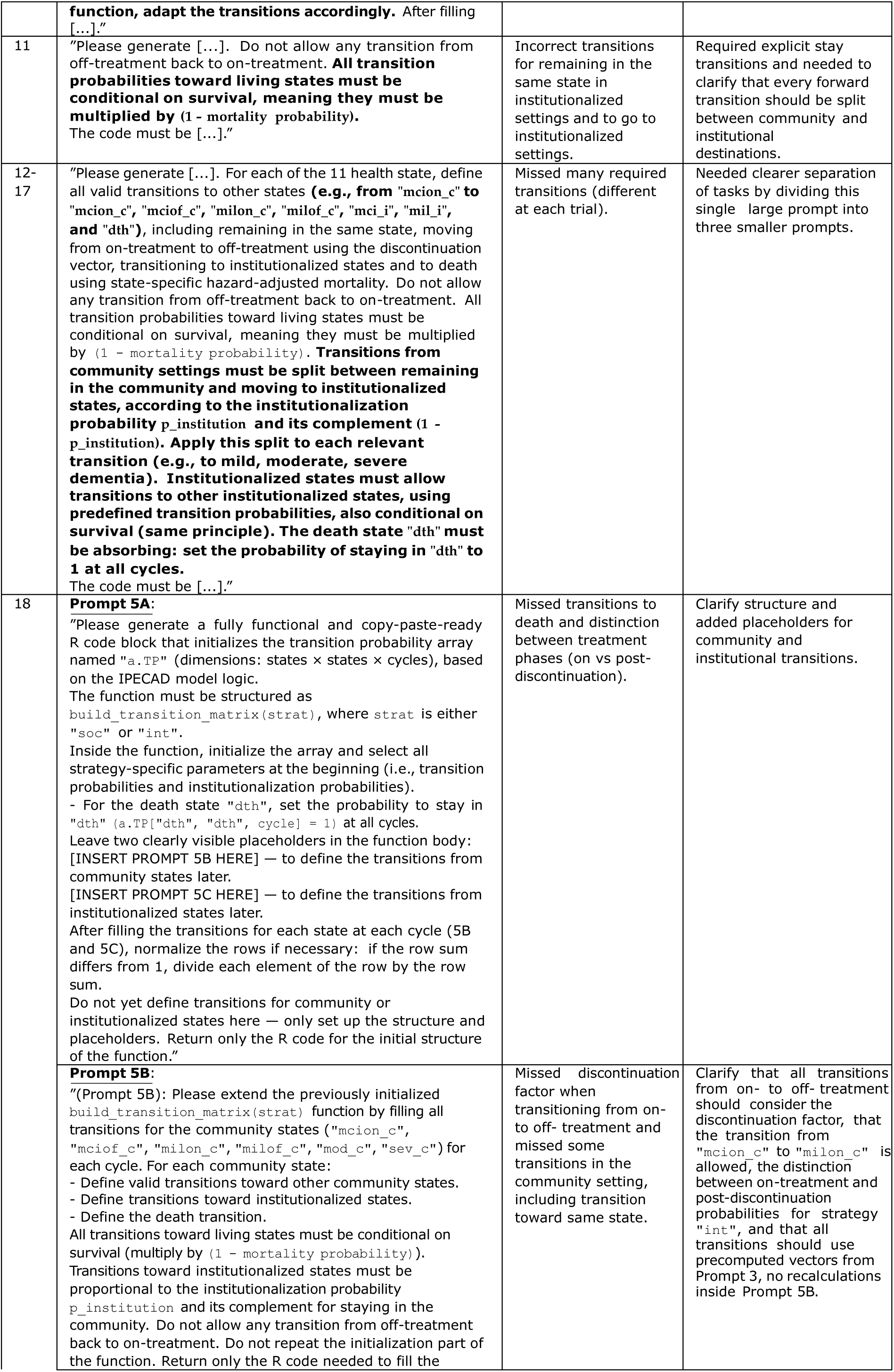
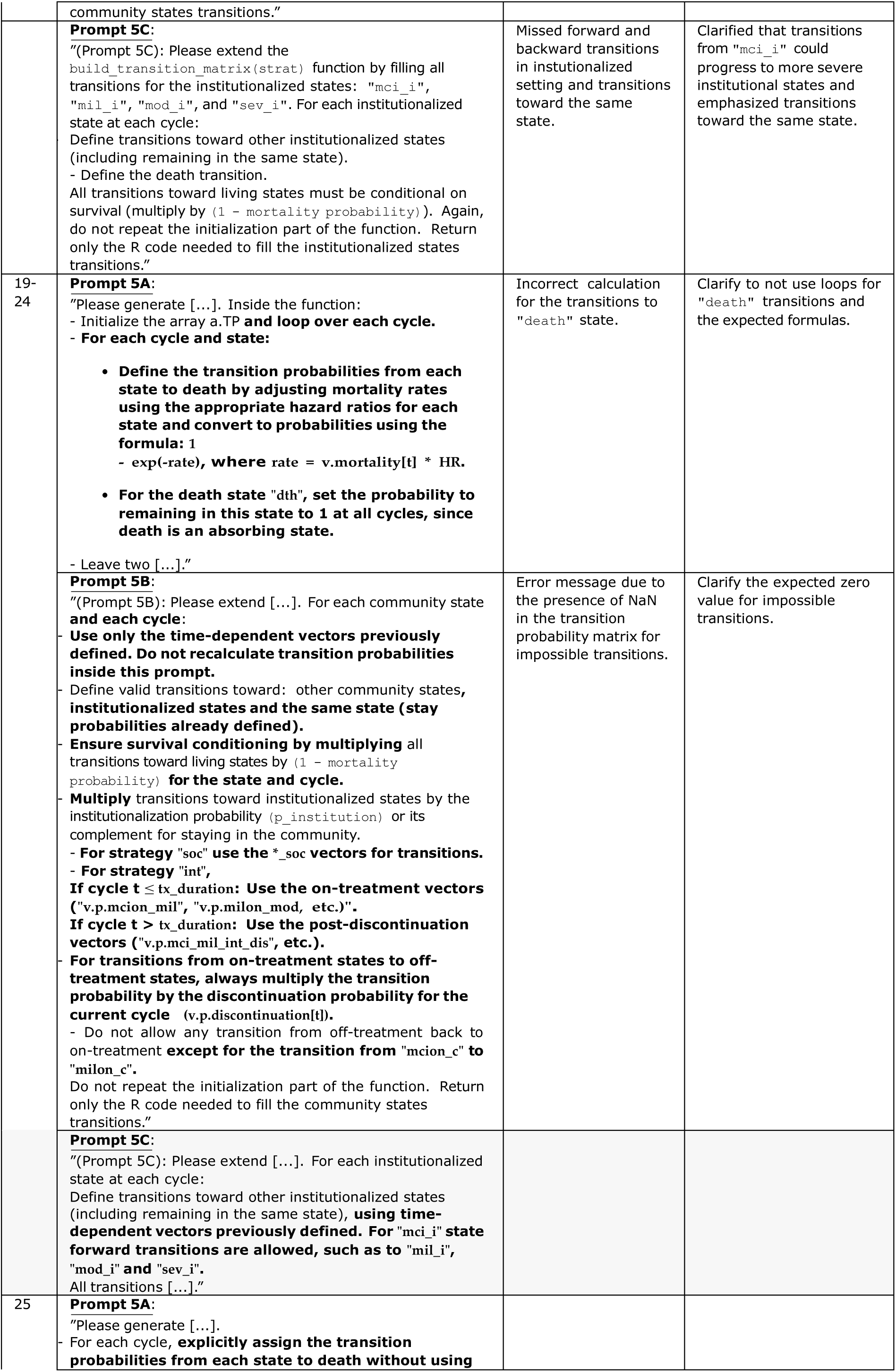
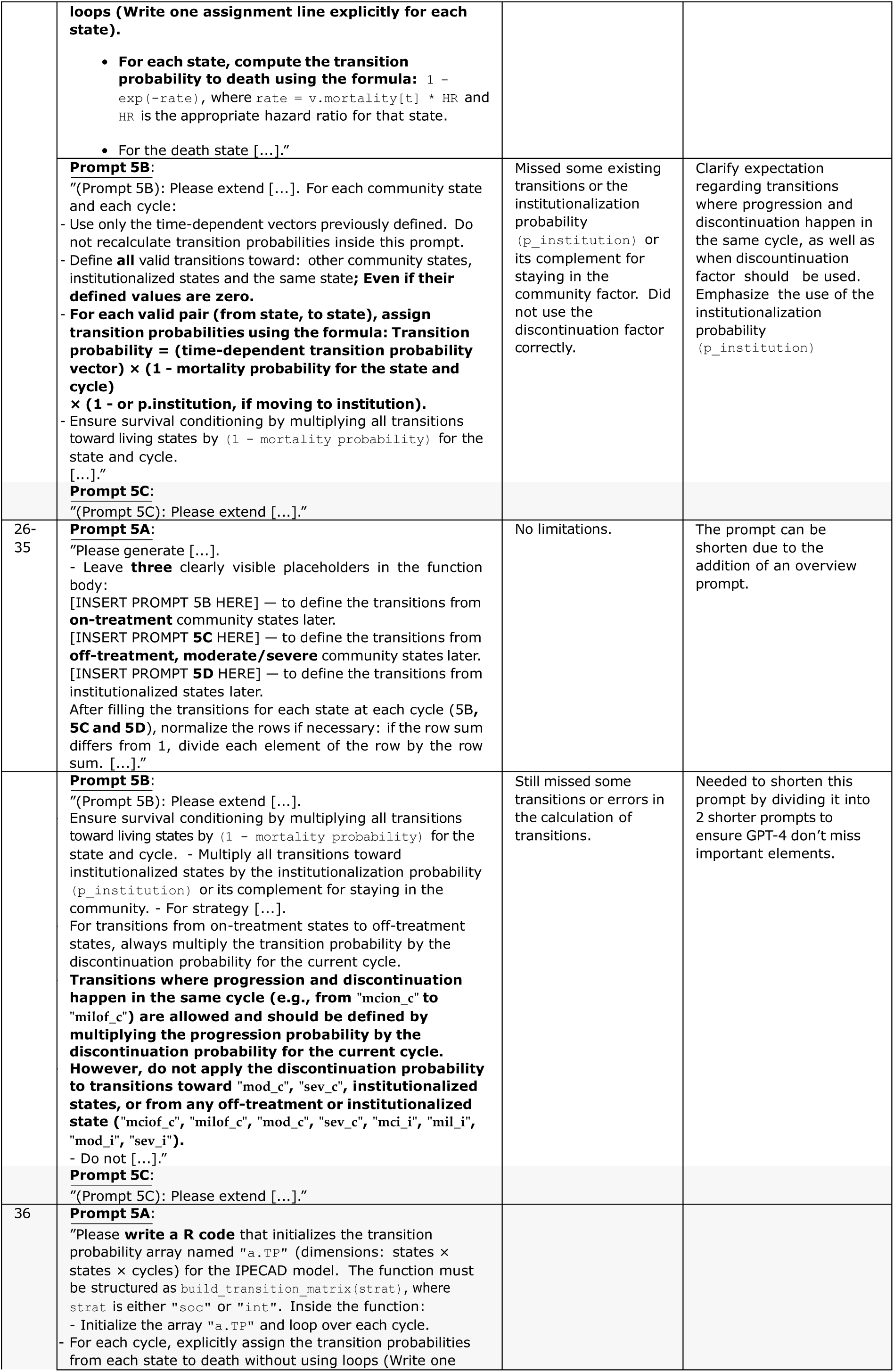
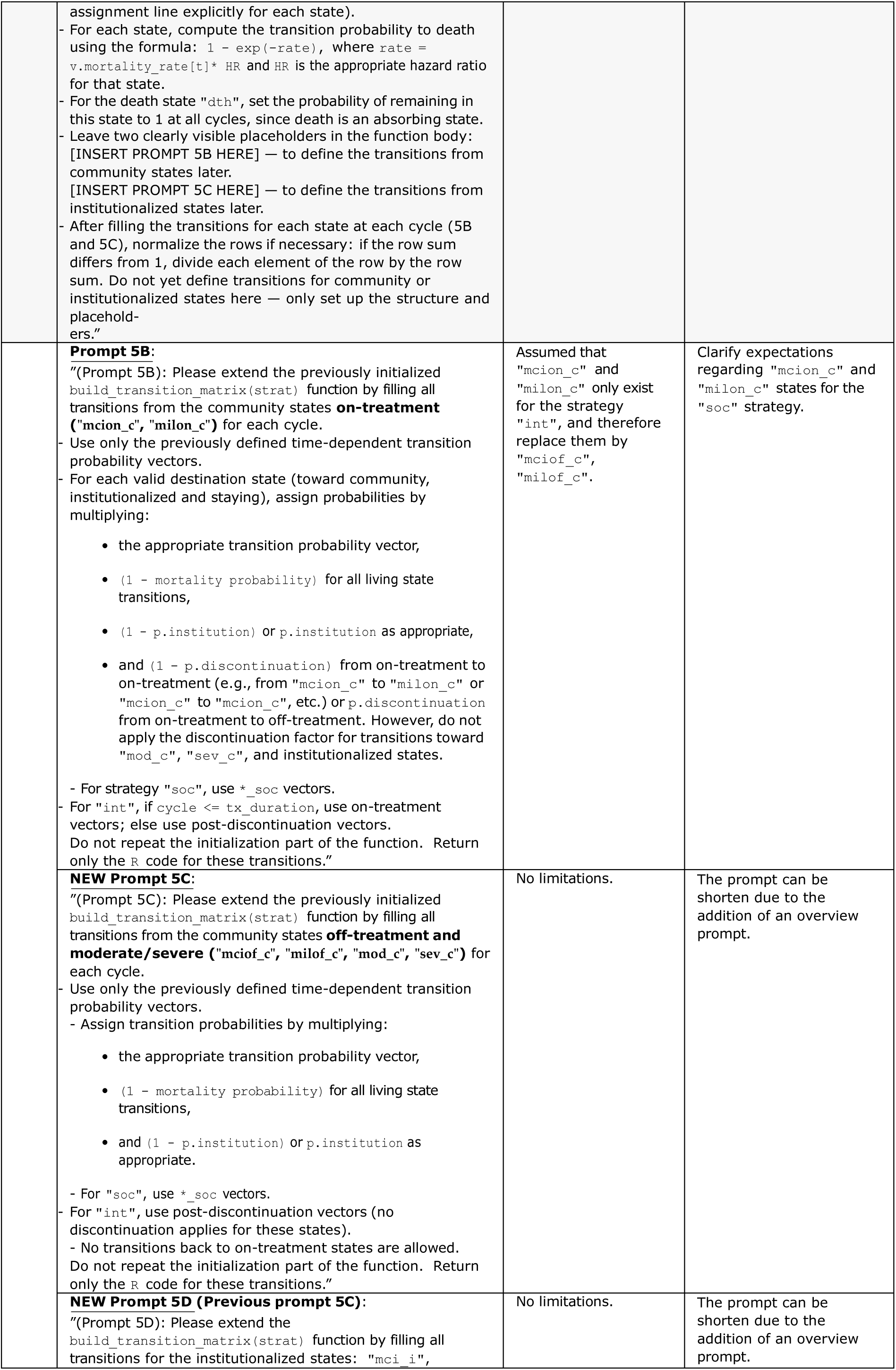
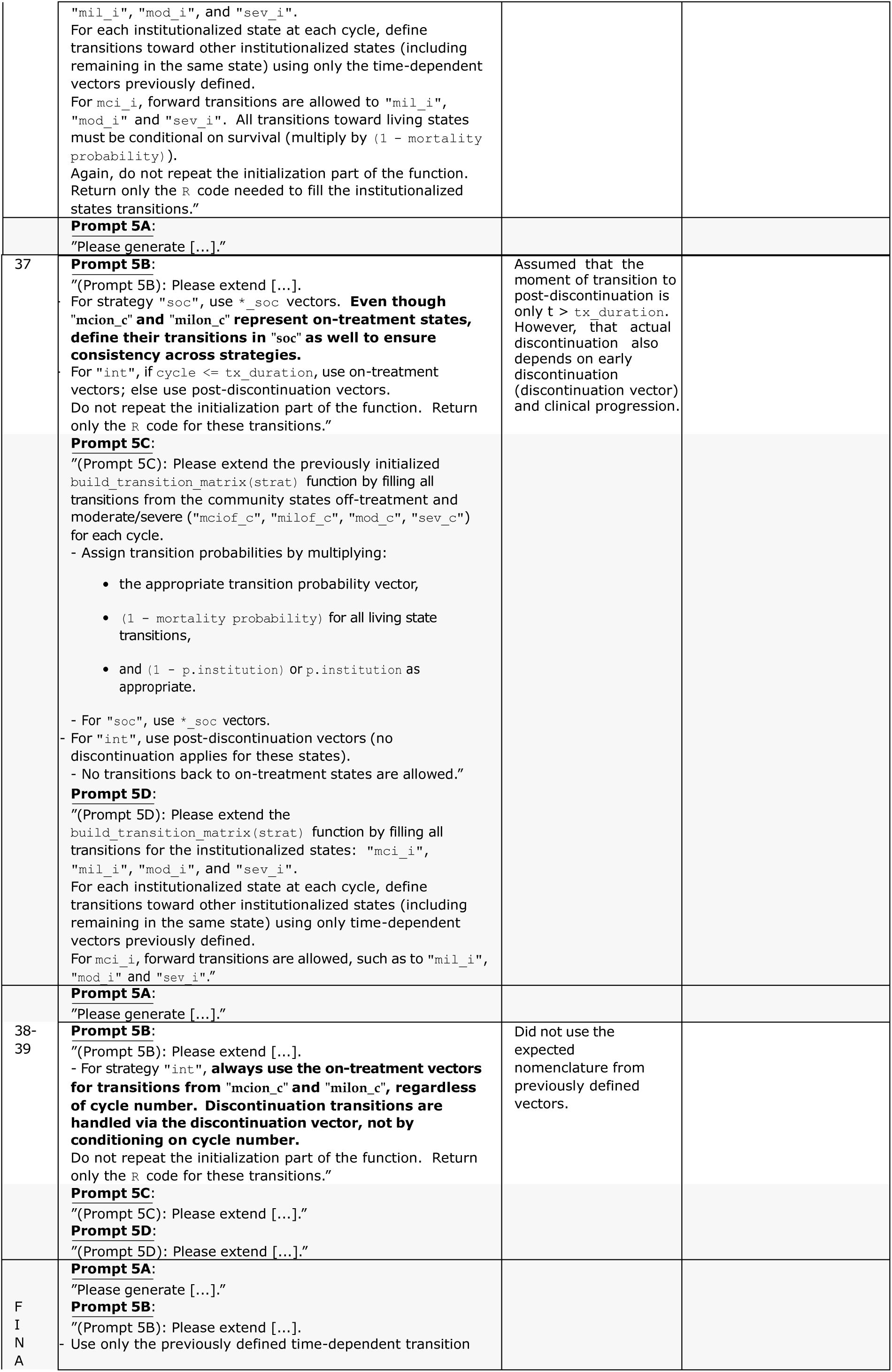
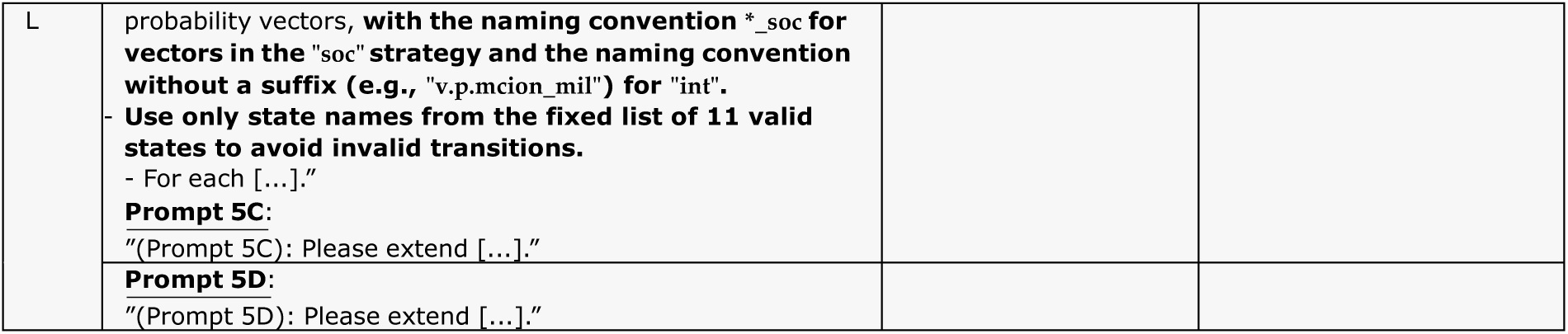
Model by IPECAD: Overview of prompt 5 refinement process and rationale for adjustments.

**Table S2.4:** Model by IPECAD: Overview of prompt 6 refinement process and rationale for adjustments.

| Trial | Prompt Evolution | Main Limitation | Reason for Modification |
| --- | --- | --- | --- |
| 1 | "Please generate a fully functional and copy-paste-ready R code block that initializes and computes the Markov trace matrix and primary outcomes (QUALYs and Costs) for a given strategy ("soc" or "int") over all cycles. Start by setting the initial state distribution. At each cycle, update the state distribution using Markov multiplication with the corresponding cycle's transition probability matrix. Compute total QALYs and total costs by multiplying state occupancy with predefined utility and cost vectors, summing over all cycles. Apply half-cycle correction and discounting to both QALYs and costs. Return only the total (aggregate) QALYs and costs — do not output values per cycle. At the end, display the results using `print()` or a compact summary table." | Half-cycle correction was not aligned with the IPECAD reference (applied starting from cycle 2, final cycle not removed). Additional inputs to cycle 1 (`u.Tx_start` and `c.Tx_start`) were omitted. | Align half-cycle correction and initial cycle adjustments with the published IPECAD methodology. |
| 2 | "Please generate [...]. Compute total QALYs and total costs by multiplying state occupancy with predefined utility and cost vectors, summing over all cycles. **For the intervention strategy, apply recurring treatment costs to on- treatment states every cycle. Apply the initial treatment cost and QUALY gain (`c.Tx_start` and `u.Tx_start`) only once, during the first cycle.** Apply half-cycle correction, **by averaging each outcome with the next cycle, excluding the last cycle.** Apply discounting to both QALYs and costs. Return only the total (aggregate) QALYs and costs **across all cycles.** At the end, display the results using `print()` or a compact summary table." | Order of operations differed from the IPECAD reference. Half-cycle correction applied at the aggregate outcome level rather than cycle-wise. | Needed to clarify the correct sequence of calculations expected. |
| 3 | "Please generate [...]. **Calculates primary outcomes (QALYs and costs) by multiplying state occupancy at each cycle with utility and cost vectors, and storing them in the output matrix. Apply half-cycle correction by averaging each cycle's outcomes with the next cycle's values, then remove the last cycle from all outcome vectors.** For the intervention strategy, apply recurring treatment costs to on-treatment states every cycle. Apply the initial treatment cost and QUALY gain (`c.Tx_start` and `u.Tx_start`) only once, during the first cycle. Apply discounting to both QALYs and costs. **Calculates total outcomes across all cycles, including life-years, total QALYs and total costs.** Return only the total (aggregate) QALYs, costs and LYs across all cycles. At the end, display the results using `print()` or a compact summary table." | Did not display the computed results (QALYs, costs, LYs). | Added output display to clearly report results and allow comparison between strategies. |
| 4 | "Please generate [...]. At the end, **write a script that prints the comparative results clearly.**" | NA caused by implicit indexing without names + incorrect name for costs vectors. | Improve it by explicitly requesting that vectors be constructed with explicit names corresponding to `state_names`, and that any indexing by state name be secured. |
| 5-6 | "Please generate a fully functional and copy-paste-ready R code block that initializes and computes the Markov trace matrix and total outcomes (QUALYs and Costs) for a given strategy ("soc" or "int") over all cycles. **Use predefined state names to define named vectors for state occupancy, utilities, and costs to ensure safe indexing.** Start by [...]. Calculates primary outcomes (QALYs and costs) by multiplying **named** state occupancy [...]" |  | Needed to add the number of the prompt to match with the overview prompt. |
| FINAL | "**(Prompt 6):** Please generate [...]" |  |  |

**Table S2.5:** Model by Wimo et al.: Overview of prompt 1 and 2 refinement process and rationale for adjustments.

| Trial | Prompt Evolution | Main Limitation | Reason for Modification |
| --- | --- | --- | --- |
| 1 | "Please extract all input parameters required to code the IPECAD Markov model using the dampack package, based on the content of the IPECAD publication and its supplemental material. Include all variables, including descriptive ones (e.g., state names and strategy names) directly as named R objects in the script. Restrict state names to the eleven that are actually used in the publication ("mcion_c", "mciof_c", "milon_c", "milof_c", "mod_c", "sev_c", "mci_i", "mil_i", "mod_i", "sev_i", "dth"). Do not omit any parameter, even if its value is zero. If a parameter is derived from a formula, preserve the original formula in the R script rather than just showing the final numeric value. Display the full R script directly in the response (do not put it in a downloadable file)." | IPECAD final prompt. | Needed to be adapted to reflect the Wimo model. |
| 2 | "(Prompt 1): Please extract all input parameters required to code the **health-economic** Markov model **described in the publication by Wimo et al. (2020)** and its supplementary material. Include all variables, including descriptive ones (e.g., state names and strategy names) directly as named R objects in the script. Restrict state names to the 5 used in the publication: "ad_mci", "mild", "moderate", "severe", and "death". Display the full R script directly in the response (do not put it in a downloadable file)." | Used final results costs and QALYs, not the initial costs components and utilities values used as a foundation. Already implemented the transition probability matrix. | Explicitly ask not to use final values from the results section but only those provided in the methods section. Specify not to implement the transition probability matrix yet. |
| 3 | "(Prompt 1): Please extract all input parameters required to code the health-economic Markov model described in the publication by Wimo et al. (2020) and its supplementary material. Include all variables, including descriptive ones (e.g., state names, strategy names), directly as named R objects in the script. **Do not implement the transition probability matrix, just extract the transition probability values.** Restrict state names to the 5 used in the publication: "ad_mci", "mild", "moderate", "severe", and "death". **When defining utility values (QALYs) and cost parameters, use only those described in the Methods section. Do not extract any values from the Results section, as they represent model outcomes, not valid input parameters.** Display the full R script directly in the response (do not put it in a downloadable file)." | Extracted only utilities values for age 60. | Clearly specify where utility and cost components should be extracted from. |
| 4 | "(Prompt 1): Please extract all input parameters required to code the health-economic Markov model described in the publication by Wimo et al. (2020) and its supplementary material. Include all variables, including descriptive ones (e.g., state names, strategy names), directly as named R objects in the script. Do not implement the transition probability matrix. **Instead, extract and define each transition probability as a clearly named R variable (e.g., p.mild_to_moderate).** Restrict state names to the 5 used in the publication: "ad_mci", "mild", "moderate", "severe", and "death". **For QALY utilities, extract all values by age and severity level directly from Supplementary Table 6. For costs, extract only the unit cost parameters from Supplementary Table 5. Do not compute or return average patient costs and QALYs.** Display the full R script directly in the response (do not put it in a downloadable file)." | Missed the hazard ratios, relative risks, and treatment costs. | Explicitly ask to include hazard ratios, relative risks, and treatment costs. |
| 5-9 | "(Prompt 1): Please extract all input parameters required to code the health-economic Markov model described in the publication by Wimo et al. (2020) and its supplementary material. Include all variables, including descriptive ones (e.g., state names, strategy names), directly as named R objects in the script. Do not implement the transition probability matrix. **Instead, extract and define each transition probability as a clearly named R variable (e.g., p.mild_to_moderate).** Restrict state names to the 5 used in the publication: "ad_mci", "mild", "moderate", "severe", and "death". **For QALY utilities, extract all values by age and severity level directly from Supplementary Table 6. For costs, extract only the unit cost parameters from Supplementary Table 5, including the treatment cost for**<br>**the hypothetical DMT.** Do not compute or return average patient costs and QALYs. **Also extract the hazard ratios (HRs) and the two relative risks (RRs) applied in the base case scenario (i.e., for transition from MCI to mild, and from mild to moderate).** Display the full R script directly in the response (do not put it in a downloadable file)." | Final costs were incorrect. | New approach chosen: provide ChatGPT-4 with the extracted fitted GLM equations for costs instead of the initial cost components. However, to keep this prompt relatively short, this will be included in a later prompt. |
| 10 | "(Prompt 1): Please extract all input parameters required to code the health-economic Markov model described in the publication by Wimo et al. (2020) and its supplementary material. Include all variables, including descriptive ones (e.g., state names, strategy names), directly as named R objects in the script. Do not implement the transition probability matrix. Instead, extract and define each transition probability as a clearly named R variable (e.g., `p.mild_to_moderate`). Restrict state names to the 5 used in the publication: "ad_mci", "mild", "moderate", "severe", and "death". For QALY utilities, extract all values by age and severity level directly from Supplementary Table 6. **For costs, extract only the treatment cost for the hypothetical DMT but do not extract unit cost parameters, as annual per-state costs will be derived using fitted equations from Supplementary Figure 2 in a later prompt.** Do not compute or return average patient costs and QALYs. Also extract the hazard ratios (HRs) and the two relative risks (RRs) applied in the base case scenario (i.e., for transition from MCI to mild, and from mild to moderate). Display the full R script directly in the response (do not put it in a downloadable file)." | Missed utilities for age between 60, 65, 70, etc. as these were not provided in the Supplementary Table 6. | Provide ChatGPT-4 with the utilities equations, manually computed in Excel, based on available ages provided in the Supplementary Table 6. |
| FINAL | "(Prompt 1): Please extract all input parameters required to code the health-economic Markov model described in the publication by Wimo et al. (2020) and its supplementary material. Include all variables, including descriptive ones (e.g., state names, strategy names), directly as named R objects in the script. Do not implement the transition probability matrix. Instead, extract and define each transition probability as a clearly named R variable (e.g., `p.mild_to_moderate`). Restrict state names to the 5 used in the publication: "ad_mci", "mild", "moderate", "severe", and "death". **For QALY utilities, define one R function per state based on regression formulas fitted to Supplementary Table 6:**<br>- AD-MCI: $u_{ad\_mci}(age) = -0.002464 \times age + 0.962679$<br>- Mild: $u_{mild}(age) = -0.0025 \times age + 0.8293$<br>- Moderate: $u_{moderate}(age) = -4e-07 \times age^2 - 0.0024 \times age + 0.5828$<br>- Severe: $u_{severe}(age) = -4e-07 \times age^2 - 0.0025 \times age + 0.4337$<br>**Implement each equation as an R function named to be used in outcome calculations. Assume utility = 0 for the death state.** For costs, extract only the treatment cost for the hypothetical DMT but do not extract unit cost parameters, as annual per-state costs will be derived using fitted equations from Supplementary Figure 2 in a later prompt. Do not compute or return average patient costs and QALYs. Also extract the hazard ratios (HRs) and the two relative risks (RRs) applied in the base case scenario (i.e., for transition from MCI to mild, and from mild to moderate). Display the full R script directly in the response (do not put it in a downloadable file)." |  |  |

**Table S2.6:** Model by Wimo et al.: Overview of prompt 2 refinement process and rationale for adjustments.

| Trial | Prompt Evolution | Main Limitation | Reason for Modification |
| --- | --- | --- | --- |
| 1 | "Please write a R code that loads the 2019 US life table from the CSV file named "lifetable_US_2019_ssa.csv". The CSV contains the following columns: "age", "male", and "female", which represent age-specific mortality probabilities for males and females, respectively. First, convert these annual mortality probabilities to annual mortality rates. Then, use these columns to compute a weighted averaged of these mortality rates and place them into a new column "weighted". Return a matrix or dataframe with columns: "age", "male", "female", and "weighted", where "male" and "female" contain the converted rates (not the original probabilities). Do not add any other packages. Assume the file is in the working directory." | IPECAD final prompt. | Needed to be adapted to reflect the Wimo model. |
| 2 | "(Prompt 2): Please write a R code that loads the 2016 Swedish mortality rate table from the CSV file named "mortalityRates_Swedish_2016.csv". The CSV contains the following columns: "age", "male", and "female". The "age" column contains age groups as ranges: "60-64", "65-69", "70-74", "75-79", "80-84", "85-89", and "90+", which represent the mean annual mortality rates for each group and sex. **First, compute a sex-weighted average mortality rate across males and females for each age group. Then, for each disease severity level, apply the following formula to compute the probability of death "1-exp(-age specific annual mortality rate \* HR)". These computed death probabilities will be later inserted into the transition probability matrix. Do not add any other packages. Assume the file is in the working directory."** | Compilation error due to length mismatches, as the 99 different ages were not present, since they were aggregated into groups. | Requested to explicitly include one row per age, regardless of the aggregated age groups in the uploaded file. |
| 3-7 | "(Prompt 2): Please write a R code that loads the 2016 Swedish mortality rate table from the CSV file named "mortalityRates_Swedish_2016.csv". The CSV contains the following columns: "age", "male", and "female". The "age" column contains age groups as ranges: "60-64", "65-69", "70-74", "75-79", "80-84", "85-89", and "90+", which represent the mean annual mortality rates for each group and sex. First, compute a sex-weighted average mortality rate across males and females for each age group. **Then, create a data frame with one row per age (60-99) and with the columns: "age", "rate_avg", "prob_death_ad_mci", "prob_death_mild", "prob_death_moderate", "prob_death_severe". For each age, compute the probability of death for each disease severity state using the formula: "p_death = 1 - exp(-rate \* HR)", using already defined state-specific hazard ratios. Do not add any other packages. Assume the file is in the working directory."** | Did not consider the sex-weighted prevalence. | Manually extract the proportion of female and male for years 2000-2016, compute weights and format as a csv file that will be provided to ChatGPT-4. |
| F<br>I<br>N<br>A<br>L | "(Prompt 2): Please write a R code that loads the 2016 Swedish mortality rate table from the CSV file named "mortalityRates_Swedish_2016.csv". The CSV contains the following columns: "age", "male", and "female". The "age" column contains age groups as ranges: "60-64", "65-69", "70-74", "75-79", "80-84", "85-89", and "90+", which represent the mean annual mortality rates for each group and sex. **Also load the file "weightsMortalityRates_Swedish_2016.csv", which contains columns "age", "weight_male", and "weight_female", representing the actual sex distribution by single year of age from 60 to 99 (40 rows), not by age group.** First, expand the grouped mortality rates to individual ages (60-99), then compute the sex-weighted average mortality rate per age using the corresponding weights. Finally, create a data frame with one row per age (60-99) and the columns: "age", "rate_avg", "prob_death_ad_mci", "prob_death_mild", "prob_death_moderate", "prob_death_severe". For each age, compute the probability of death for each disease severity state using the formula: "p_death = 1 - exp(-rate * HR)", using already defined state-specific hazard ratios. Do not use any additional packages. Assume all files are in the working directory." |  |  |

**Table S2.7:** Model by Wimo et al.: Overview of prompt 3 refinement process and rationale for adjustments.

| Trial | Prompt Evolution | Main Limitation | Reason for Modification |
| --- | --- | --- | --- |
| 1 | "Please write a R code that defines all time-dependent transition probability vectors for the standard of care ("soc") strategy, without using `assign()` or `get()`, or dynamic variable names. Define the following vectors:<br>Transition probability vectors per cycle for disease progression under the SOC strategy ("`v.p.mci_mil_soc`", "`v.p.mci_mod_soc`", "`v.p.mci_sev_soc`", etc.).<br>Staying probabilities per state as 1 minus the sum of other disease progression probabilities only (exclude institutional transitions). Survival adjustment will be applied later.<br>Time-dependent vectors for transitions from each community state to institutionalized settings.<br>A discontinuation vector per cycle, with the first discontinuation probability value for the first time interval and the second discontinuation probability from the second time point until the maximum treatment duration. After reaching the maximum treatment duration, the value should be set to 1. Each cycle's value must be treated independently (no cumulative calculation).<br>- A vector of age- and sex-specific mortality rates using the predefined mortality matrix." | IPECAD final prompt. | Needed to be adapted to reflect the Wimo model. |
| 2 | "(Prompt 3): Please write a R code that defines all time-dependent transition probability vectors for **both the no_intervention and intervention strategy**, without using `assign()` or `get()`, or dynamic variable names.<br>**For the no_intervention strategy, define a transition probability vector for each cycle:**<br>- From "ad_mci" to "mild" and staying in "ad_mci".<br>- From "mild" to "moderate", "severe" and staying in "mild".<br>- From "moderate" to "mild", "severe" and staying in "moderate".<br>- From "severe" to "moderate" and staying in "severe". For the intervention strategy (base-case scenario with hypothetical DMT treatment applied during "ad_mci" and "mild" states only), apply the following formula to each affected transition:<br>- Convert the original probability to a rate: $\text{rate} = -\log(1 - p)$.<br>- Adjust the rate using the appropriate relative risk: $\text{adjusted\_rate} = \text{rate} * \text{RR}$.<br>- Convert the rate back to a probability: $\text{adjusted\_p} = 1 - \exp(-\text{adjusted\_rate})$.<br>Also define the staying probabilities per state for each strategy as: $1 - \text{sum}(\text{other transition probabilities from that state})$. Return only the R code block defining the vectors. Assume that all input probabilities and relative risks have been defined previously." | Defined scalars instead of vectors. | Explicitly requested to use `rep()` function to ensure vectors are returned. |
| FINAL | "(Prompt 3): Please write a R code that defines all time-dependent transition probability vectors for both the no_intervention and intervention strategy, without using `assign()` or `get()`, or dynamic variable names.<br>For the no_intervention strategy, define a transition probability vector for each cycle:<br>- From "ad_mci" to "mild" and staying in "ad_mci".<br>- From "mild" to "moderate", "severe" and staying in "mild".<br>- From "moderate" to "mild", "severe" and staying in "moderate".<br>- From "severe" to "moderate" and staying in "severe".<br>For the intervention strategy (base-case scenario with hypothetical DMT treatment applied during "ad_mci" and "mild" states only), apply the following formula to each affected transition:<br>- Convert the original probability to a rate: $\text{rate} = -\log(1 - p)$.<br>- Adjust the rate using the appropriate relative risk: $\text{adjusted\_rate} = \text{rate} * \text{RR}$.<br>- Convert the rate back to a probability: $\text{adjusted\_p} = 1 - \exp(-\text{adjusted\_rate})$.<br>Also define the staying probabilities per state for each strategy as: $1 - \text{sum}(\text{other transition probabilities from that state})$. Return only the R code block defining the vectors. Assume that all input probabilities and relative risks have<br>been defined previously. **Do not forget to use rep() to make all output vectors of length n_cycles."** |  |  |

**Table S2.8:**
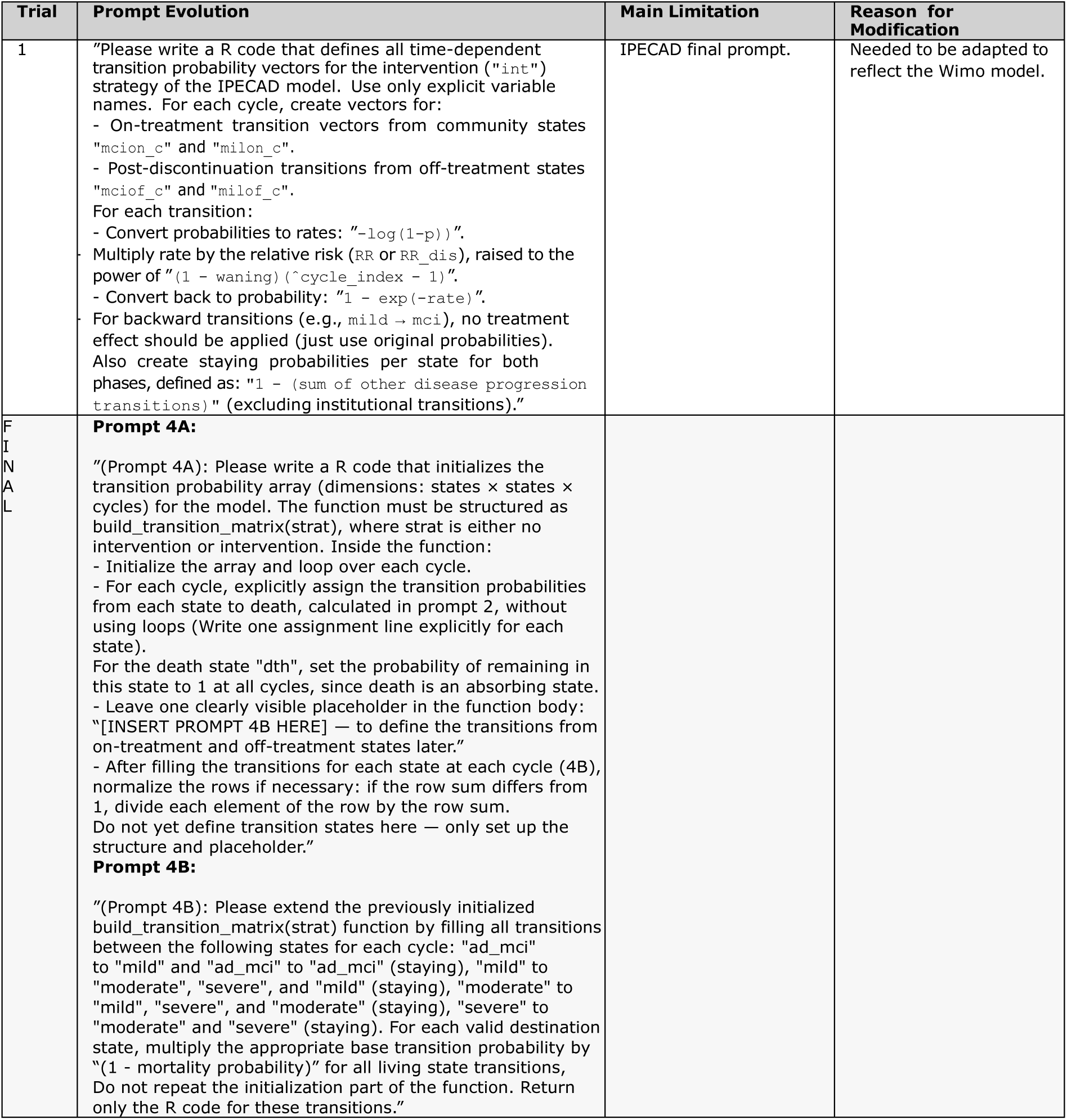
Model by Wimo et al.: Overview of prompt 4 refinement process and rationale for adjustments.

**Table S2.9:**
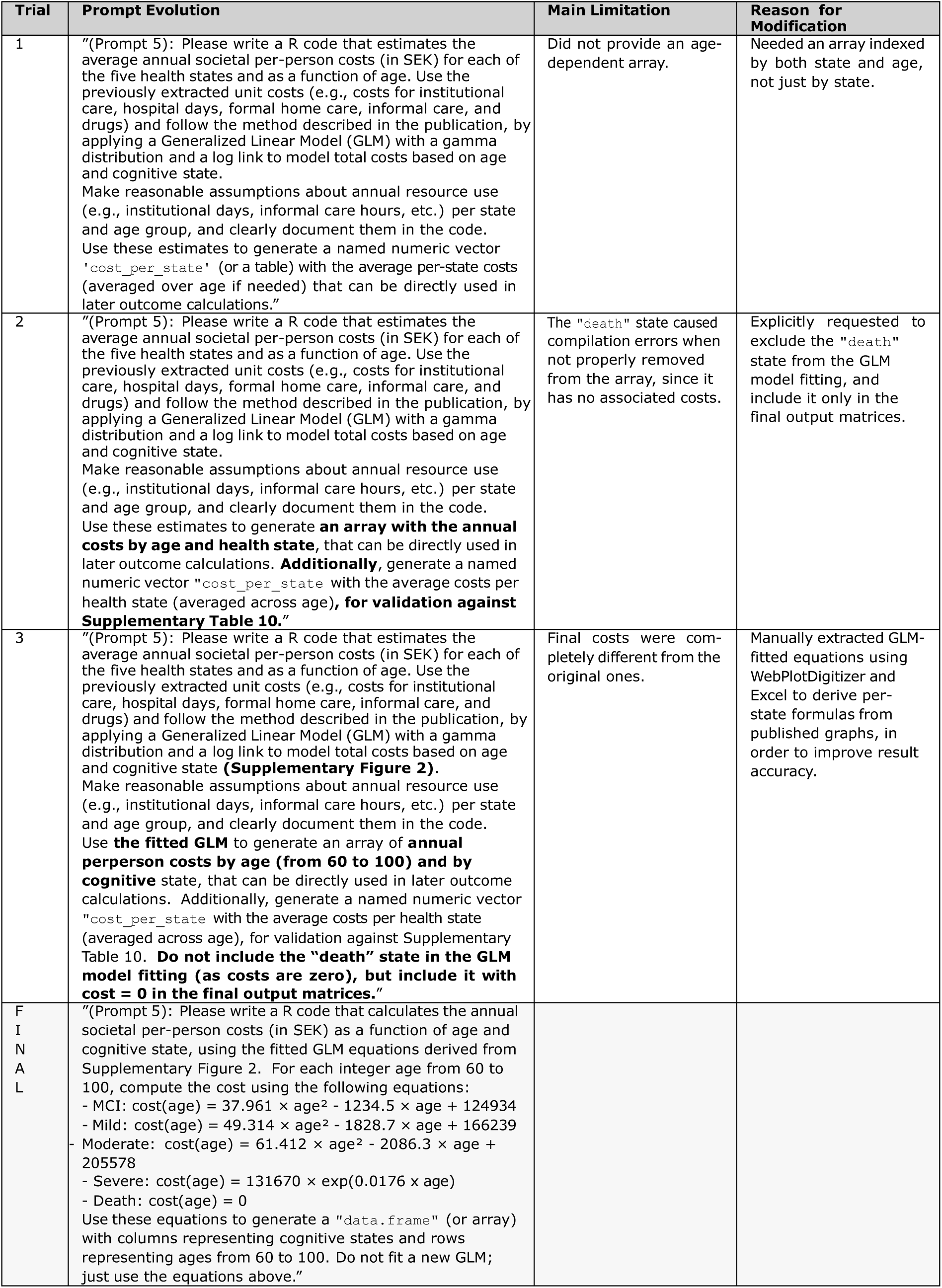
Model by Wimo et al.: Overview of prompt 5 refinement process and rationale for adjustments.

**Table S2.10:** Model by Wimo et al.: Overview of prompt 6 refinement process and rationale for adjustments.

| Trial | Prompt Evolution | Main Limitation | Reason for Modification |
| --- | --- | --- | --- |
| 1 | “(Prompt 6): Please generate a fully functional and copy-paste-ready R code block that initializes and computes the Markov trace matrix and total outcomes (QALYs and Costs) for a given strategy ("soc" or "int") over all cycles. Use predefined state names to define named vectors for state occupancy, utilities, and costs to ensure safe indexing. Start by setting the initial state distribution. At each cycle, update the state distribution using Markov multiplication with the corresponding cycle's transition probability matrix. Calculates primary outcomes (QALYs and costs) by multiplying named state occupancy at each cycle with utility and cost vectors, and storing them in the output matrix. Apply half-cycle correction by averaging each cycle's outcomes with the next cycle's values, then remove the last cycle from all outcome vectors. For the "int" strategy, apply recurring treatment costs to on-treatment states every cycle. Apply the initial treatment cost and QALY gain (c.Tx_start and u.Tx_start) only once, during the first cycle. Apply discounting to both QALYs and costs. Calculates total outcomes across all cycles, including life-years, total QALYs and total costs. Return only the total (aggregate) QALYs, costs and LYs across all cycles. At the end, write a script that prints the comparative results clearly.” | IPECAD final prompt. | Needed to be adapted to reflect the Wimo model. |
| 2 | “(Prompt 6): Please write an R script that runs the Markov model for both strategies ("no_intervention" and "intervention") over all cycles using the 5 predefined states, with the full cohort starting in "ad_mci". At each cycle, update the cohort via Markov multiplication with the corresponding transition matrix, and compute:<br>- Total QALYs by multiplying the cohort state vector by age- and state-specific utility values.<br>- Total costs by multiplying the cohort vector by per-state and age-specific cost values. For the intervention strategy, add the annual DMT cost to the "ad_mci" and "mild" states.<br>- Person-years per state (PYs), storing them for each cycle.<br>Apply half-cycle correction to QALYs, costs, and PYs by averaging each cycle's outcomes with the next cycle's values, then remove the last cycle from all outcome vectors. Apply discounting to both QALYs and costs. At the end, print a summary table in the console showing: total discounted QALYs and costs (in SEK) per strategy, discounted PYs/person per state, incremental Cost-Effectiveness Ratio (ICER), Net Monetary Benefit (NMB) at a WTP threshold of 600,000 SEK.” | More relevant to focus on primary outcomes, such as cumulative clinical outcomes rather than on secondary outcomes (e.g. ICER, NMB). | Refocuses the prompt on primary outcomes, such as cumulative deaths and dementia cases. |
| 3 | “(Prompt 6): Please write an R script that runs the Markov model for both strategies ("no_intervention" and "intervention") over all cycles using the 5 predefined states, with the full cohort starting in "ad_mci". At each cycle, update the cohort via Markov multiplication with the corresponding transition matrix, and compute:<br>- Total QALYs by multiplying the cohort state vector by age- and state-specific utility values.<br>- Total costs by multiplying the cohort vector by per-state and age-specific cost values. For the intervention strategy, add the annual DMT cost to the "ad_mci" and "mild" states.<br>- Person-years per state (PYs), storing them for each cycle.<br>Apply half-cycle correction to QALYs, costs, and PYs by averaging each cycle's outcomes with the next cycle's values, then remove the last cycle from all outcome vectors. Apply discounting to both QALYs and costs. At the end, print a summary table in the console showing: total discounted QALYs and costs (in SEK) per strategy, discounted PYs/person per state, **and the following cumulative values:**<br>- **Total number of deaths,**<br>- **Total number of deaths with dementia,**<br>- **Total number of deaths with AD-MCI,**<br>- **Total number of dementia cases.”** | Returned only the expected results without the R script. | Explicitly requests a complete and copy-paste-ready R script. |
| 4 | “(Prompt 6): Please write **a complete, copy-paste-ready** | QALYs and costs were | Explicitly requests to use |
|  | **R** script that runs the Markov model for both strategies ("no_intervention" and "intervention") over all cycles using the 5 predefined states, with the full cohort starting in "ad_mci". At each cycle, update the cohort via Markov multiplication with the corresponding transition matrix, and compute:<br>- Total QALYs by multiplying the cohort state vector by age- and state-specific utility values.<br>- Total costs by multiplying the cohort vector by per-state and age-specific cost values. For the intervention strategy, add the annual DMT cost to the "ad_mci" and "mild" states.<br>- Person-years per state (PYs), storing them for each cycle.<br>Apply half-cycle correction to QALYs, costs, and PYs by averaging each cycle's outcomes with the next cycle's values, then remove the last cycle from all outcome vectors. Apply discounting to both QALYs and costs. At the end, print a summary table in the console showing: total discounted QALYs and costs (in SEK) per strategy, discounted PYs/person per state, **and the following cumulative values for the total number of deaths, and the total number of deaths with AD-MCI."** | not computed based on previously defined age-specific utility functions. Cumulative deaths were not computed correctly. | previously defined utility and cost functions and ensures correct calculation of deaths using transitions. |
| 5 | “(Prompt 6): Please write a complete, copy-paste-ready R script that runs the Markov model for both strategies ("no_intervention" and "intervention") over all cycles using the 5 predefined states, with the full cohort starting in "ad_mci". At each cycle, update the cohort via Markov multiplication with the corresponding transition matrix, and compute:<br>- Total QALYs by multiplying the cohort state vector **by age-specific utility functions defined for each health state (e.g., u_mild(age)), evaluated for the current cycle's age.**<br>- Total costs by multiplying the cohort vector by per-state and age-specific cost values. For the intervention strategy, add the annual DMT cost to the "ad_mci" and "mild" states.<br>- Person-years per state (PYs), storing them for each cycle.<br>Apply half-cycle correction to QALYs, costs, and PYs by averaging each cycle's outcomes with the next cycle's values, then remove the last cycle from all outcome vectors. Apply discounting to both QALYs and costs. At the end, print a summary table in the console showing: total discounted QALYs and costs (in SEK) per strategy, discounted PYs/person per state, and the following cumulative values for the total number of deaths, and the total number of deaths with AD-MCI." | QALYs and costs were not expressed per person, and death counts were derived from occupancy instead of transitions. | Clarifies that QALYs and costs must be computed per person, and that deaths should be calculated as transitions into "death". |
| 6 | “(Prompt 6): Please write a complete, copy-paste-ready R script that runs the Markov model for both strategies ("no_intervention" and "intervention") over all cycles using the 5 predefined states, with the full cohort starting in "ad_mci". At each cycle, update the cohort via Markov multiplication with the corresponding transition matrix, and compute:<br>- Total QALYs by multiplying the cohort state vector by age-specific utility functions defined for each health state (e.g., u_mild(age)), evaluated for the current cycle's age.<br>- Total costs by multiplying the cohort vector by per-state and age-specific cost values. For the intervention strategy, add the annual DMT cost to the "ad_mci" and "mild" states.<br>- Person-years per state (PYs), storing them for each cycle.<br>Apply half-cycle correction to QALYs, costs, and PYs by averaging each cycle's outcomes with the next cycle's values, then remove the last cycle from all outcome vectors. Apply discounting to both QALYs and costs. At the end, print a summary table in the console showing: total discounted QALYs and costs (in SEK) per strategy **and per person, undiscounted PYs/person per state, the total number of cumulative deaths (based on transitions into "death"), and the total number of deaths with AD-MCI (transitions from "ad_mci" to "death")."** | No fix results structure across trials. | Needed to formalize the name of variables to be able to store the final outcome across the fifteen trials. |
| 7 | “(Prompt 6): Please write a complete, copy-paste-ready R script that runs the Markov model for both strategies ("no_intervention" and "intervention") over all cycles using the 5 predefined states, with the full cohort starting in "ad_mci". At each cycle, update the cohort via Markov multiplication with the corresponding transition matrix, and compute: | Computation error due to vector length mismatch after half-cycle correction. Miscalculated total deaths. | Ensures alignment between vector lengths and refines death computation based on state transitions. |
| | - Total QALYs by multiplying the cohort state vector by age-specific utility functions defined for each health state (e.g., `u_mild(age)`), evaluated for the current cycle's age.<br>- Total costs by multiplying the cohort vector by per-state and age-specific cost values. For the intervention strategy, add the annual DMT cost to the "ad_mci" and "mild" states.<br>- Person-years per state (PYs), storing them for each cycle. Apply half-cycle correction to QALYs, costs, and PYs by averaging each cycle's outcomes with the next cycle's values, then remove the last cycle from all outcome vectors. Apply discounting to both QALYs and costs. At the end, print a summary table in the console showing: total discounted QALYs and costs (in SEK) per strategy and per person, undiscounted PYs/person per state, the total number of cumulative deaths (based on transitions into "death"), and the total number of deaths with AD-MCI (transitions from "ad_mci" to "death"). **Additionally, return the final results in a named list called "results", structured as `results$strategy_name$metric_name`, where `strategy_name` is "no_intervention" or "intervention", and `metric_name` is one of "qalys", "costs", "pys_ad_mci", "pys_mild", "pys_mod", "pys_sev", "deaths_total", "deaths_ad_mci".** | | |
| 8-11 | “(Prompt 6): Please write a complete, copy-paste-ready R script that runs the Markov model for both strategies ("no_intervention" and "intervention") over all cycles using the 5 predefined states, with the full cohort starting in "ad_mci". At each cycle, update the cohort via Markov multiplication with the corresponding transition matrix, and compute:<br>- Total QALYs by multiplying the cohort state vector by age-specific utility functions defined for each health state (e.g., `u_mild(age)`), evaluated for the current cycle's age.<br>- Total costs by multiplying the cohort vector by per-state and age-specific cost values. For the intervention strategy, add the annual DMT cost to the "ad_mci" and "mild" states.<br>- Person-years per state (PYs), storing them for each cycle. Apply half-cycle correction to QALYs, costs, and PYs by averaging each cycle's outcomes with the next, then remove the last cycle from all outcomes **and also from the state trace. Discount only QALYs and costs using a discount factor vector whose length exactly matches the length of the QALY and cost vectors after half-cycle correction.**<br>At the end, print a summary table showing: total discounted QALYs and costs (in SEK) per strategy and per person, undiscounted PYs/person per state, **total deaths (i.e., sum of new deaths = difference in "death" state between cycles), and total cumulative deaths** with AD-MCI (transitions from "ad_mci" to "death"). Additionally, return **all** results in a named list called "results", structured as `results$strategy_name$metric_name`, where `strategy_name` is "no_intervention" or "intervention", and `metric_name` is one of "qalys", "costs", "pys_ad_mci", "pys_mild", "pys_mod", "pys_sev", "deaths_total", "deaths_ad_mci".” | The PYs matrix was not initialized with named columns, and the prompt was becoming too long. | Adds named columns to the PYs matrix and divides the prompt. |
| 12-13 | **Prompt 6A:**<br>“(Prompt 6A): Please write **the structure of an R script that runs a Markov model for both strategies over all cycles using the 5 predefined states, with the full cohort starting in "ad_mci". For each strategy:**<br>- **Initialize a state trace matrix, vectors for QALYs and costs, a PYs matrix with named columns for the four non-death states, and vectors to track total deaths and deaths from AD-MCI.**<br>- **Leave one clearly visible placeholder in the function body: “[INSERT PROMPT 6B HERE] — simulation loop per cycle”.**<br>- **After the loop, apply half-cycle correction to QALYs, costs, and PYs by averaging each cycle's outcomes with the next, then remove the last cycle from all outcomes and also from the state trace.**<br>- **Apply discounting only to QALYs and costs using a discount factor vector matching their corrected lengths.**<br>- Return **all** results in a named list<br>results\$strategy_name\$metric_name, where strategy_name is "no_intervention" or "intervention", and metric_name is one of "qalys" (**discounted, per person**), "final_costs" (**discounted, SEK/person**), "pys_ad_mci", "pys_mild", "pys_mod", "pys_sev", "deaths_total" (**sum of new deaths per cycle**), "deaths_ad_mci" (**cumulative transitions from "ad_mci" to "death"**)."<br>**Prompt 6B:**<br>“(Prompt 6B): Please write the inner simulation loop to be inserted at [INSERT PROMPT 6B HERE] in Prompt 6A. At each cycle, update the cohort via Markov matrix multiplication using the appropriate transition matrix and compute:<br>- The current age<br>- QALYs by multiplying the current state distribution by age-specific utility functions defined for each health state (e.g., `u_mild(age)`), evaluated for the current cycle’s age.<br>- Costs using state- and age-specific cost values. For the intervention strategy, add the annual DMT cost to the "ad_mci" and "mild" states.<br>- Person-years (PYs) per state from the state occupancy vector.<br>- Track total deaths as the difference in the "death" state between the next and current cycle.<br>- Track cumulative deaths from AD-MCI by multiplying the number of individuals in "ad_mci" by the transition probability from "ad_mci" to "death" in the current cycle. " | The final `results` object is inside an R function and therefore not accessible from outside. | Clarifies that the model must be executed outside the function for each strategy and stored in a structured results object. |
| FINAL | **Prompt 6A:**<br>“(Prompt 6A): Please write the structure of an R script that runs a Markov model for both strategies over all cycles using the 5 predefined states, with the full cohort starting in "ad_mci". For each strategy:<br>- Initialize a state trace matrix, vectors for QALYs and costs, a PYs matrix with named columns for the four non-death states, and vectors to track total deaths and cumulative deaths from AD-MCI.<br>- Leave one clearly visible placeholder in the function body: “[INSERT PROMPT 6B HERE] — simulation loop per cycle”.<br>- After the loop, apply half-cycle correction to QALYs, costs, and PYs by averaging each cycle’s outcomes with the next, then remove the last cycle from all outcomes and also from the state trace.<br>- Apply discounting only to QALYs and costs using a discount factor vector matching their corrected lengths.<br>- Return a named list with the final results per strategy, including: "qalys" (discounted, per person), "final_costs" (discounted, SEK/person), "pys_ad_mci", "pys_mild", "pys_mod", "pys_sev", "deaths_total" (sum of new deaths per cycle), "deaths_ad_mci" (cumulative transitions from "ad_mci" to "death").<br>**Outside the function, run the model separately for both strategies and store the outputs in a structure as follows: results\$strategy_name\$metric_name. "**<br>**Prompt 6B:**<br>“(Prompt 6B): Please write the inner simulation loop to be inserted at [INSERT PROMPT 6B HERE] in Prompt 6A. At each cycle, update the cohort via Markov matrix multiplication using the appropriate transition matrix and compute:<br>- The current age<br>- QALYs by multiplying the current state distribution by age-specific utility functions defined for each health state (e.g., `u_mild(age)`), evaluated for the current cycle’s age.<br>- Costs using state- and age-specific cost values. For the intervention strategy, add the annual DMT cost to the "ad_mci" and "mild" states.<br>- Person-years (PYs) per state from the state occupancy vector.<br>- Track total deaths as the difference in the "death" state between the next and current cycle.<br>- Track cumulative deaths from AD-MCI by multiplying the number of individuals in "ad_mci" by the transition probability from "ad_mci" to "death" in the current cycle. " | | |

**Table S2.11:** Model by Ross et al.: Overview of prompt 1 and 2 refinement process and rationale for adjustments.

| Trial | Prompt Evolution | Main Limitation | Reason for Modification |
| --- | --- | --- | --- |
| 1 | “(Prompt 1): Please extract all input parameters required to code the health-economic Markov model described in the publication by Wimo et al. (2020) and its supplementary material. Include all variables, including descriptive ones (e.g., state names, strategy names), directly as named R objects in the script. Do not implement the transition probability matrix. Instead, extract and define each transition probability as a clearly named R variable (e.g., `p.mild_to_moderate`). Restrict state names to the 5 used in the publication: `"ad_mci", "mild", "moderate", "severe", and "death"`. **For QALY utilities, define one R function per state based on regression formulas fitted to Supplementary Table 6:**<br>- **AD-MCI:** $u_{ad\_mci}(age) = -0.002464 \times age + 0.962679$<br>- **Mild:** $u_{mild}(age) = -0.0025 \times age + 0.8293$<br>- **Moderate:** $u_{moderate}(age) = -4e-07 \times age^2 - 0.0024 \times age + 0.5828$<br>- **Severe:** $u_{severe}(age) = -4e-07 \times age^2 - 0.0025 \times age + 0.4337$<br>**Implement each equation as an R function named to be used in outcome calculations. Assume utility = 0 for the death state.** For costs, extract only the treatment cost for the hypothetical DMT but do not extract unit cost parameters, as annual per-state costs will be derived using fitted equations from Supplementary Figure 2 in a later prompt. Do not compute or return average patient costs and QALYs. Also extract the hazard ratios (HRs) and the two relative risks (RRs) applied in the base case scenario (i.e., for transition from MCI to mild, and from mild to moderate). Display the full R script directly in the response (do not put it in a downloadable file).” | Wimo final prompt. | Needed to be adapted to reflect the Ross model. |
| 2 | “(Prompt 1): Please extract all input parameters required to code the health-economic Markov model described in the publication by Ross et al. (2022) and its supplementary material. Include all variables, including descriptive ones (e.g., state names, strategy names), directly as named R objects in the script. Do not implement the transition probability matrix. Instead, extract and define each transition probability as a clearly named R variable (e.g., `p.mild_to_moderate`). Restrict state names to the 5 used in the publication: `"mci", "mild", "moderate", "severe", and "death"`. Simulate a cohort of 1000 individuals by generating a vector of age values drawn from a normal distribution with a mean of 75.2 and standard deviation of 5.5 (as described in the source trial), and store it in a variable called `ages`. Round the ages to the nearest whole number and restrict them to the range 60–99. Display the full R script directly in the response (do not put it in a downloadable file).” | Missed ARIA-related costs. | Explicitly ask to include ARIA-related costs among the cost components. |
| 3 | “(Prompt 1): Please extract all input parameters required to code the health-economic Markov model described in the publication by Ross et al. (2022) and its supplementary material. Include all variables, including descriptive ones (e.g., state names, strategy names), directly as named R objects in the script. Do not implement the transition probability matrix. Instead, extract and define each transition probability as a clearly named R variable (e.g., `p.mild_to_moderate`). Restrict state names to the 5 used in the publication: `"mci", "mild", "moderate", "severe", and "death"`. **Include all costs, including those related to the healthcare and societal sectors, as well as annual drug costs, monthly infusion and monitoring costs, screening costs, and ARIA-related costs.** Simulate a cohort of 1000 individuals by generating a vector of age values drawn from a normal distribution with a mean of 75.2 and standard deviation of 5.5 (as described in the source trial), and store it in a variable called `ages`. Round the ages to the nearest whole number and restrict them to the range 60–99. Display the full R script directly in the response (do not put it in a<br>downloadable file)." | The simulation of the cohort based on a random age distribution led to inconsistencies. | Clearly specify that the cohort should be simulated using a fixed starting age of 75.2 years for all 1,000 individuals. |
| 4 | "(Prompt 1): Please extract all input parameters required to code the health-economic Markov model described in the publication by Ross et al. (2022). Include all variables, including descriptive ones (e.g., state names, strategy names), directly as named R objects in the script. Do not implement the transition probability matrix. Instead, extract and define each transition probability as a clearly named R variable (e.g., `p.mild_to_moderate`). Restrict state names to the 5 used in the publication: "mci", "mild", "moderate", "severe", and "death". Include all costs, including those related to the healthcare and societal sectors, as well as annual drug costs, monthly infusion and monitoring costs, screening costs, and ARIA-related costs. Simulate a cohort of 1000 individuals, **all with a fixed starting age of 75.2 years**. Display the full R script directly in the response (do not put it in a downloadable file)." | The utility values were interpreted as monthly while they were reported annually. | Utility values are divided by 12 to convert annual values into monthly utilities, aligning with the model's time cycle. |
| F<br>I<br>N<br>A<br>L | "(Prompt 1): Please extract all input parameters required to code the health-economic Markov model described in the publication by Ross et al. (2022). Include all variables, including descriptive ones (e.g., state names, strategy names), directly as named R objects in the script. Do not implement the transition probability matrix. Instead, extract and define each transition probability as a clearly named R variable (e.g., `p.mild_to_moderate`). Restrict state names to the 5 used in the publication: "mci", "mild", "moderate", "severe", and "death". Include all costs, including those related to the healthcare and societal sectors, as well as annual drug costs, monthly infusion and monitoring costs, screening costs, and ARIA-related costs. **For utilities, assume the reported values are annual and convert them into monthly utilities by dividing them by 12.** Simulate a cohort of 1000 individuals, all with a fixed starting age of 75.2 years. Display the full R script directly in the response (do not put it in a downloadable file)." |  |  |

**Table S2.12:** Model by Ross et al.: Overview of prompt 2 refinement process and rationale for adjustments.

| Trial | Prompt Evolution | Main Limitation | Reason for Modification |
| --- | --- | --- | --- |
| 1 | “(Prompt 2): Please write a R code that loads the 2016 Swedish mortality rate table from the CSV file named "mortalityRates_Swedish_2016.csv". The CSV contains the following columns: "age", "male", and "female". The "age" column contains age groups as ranges: "60-64", "65-69", "70-74", "75-79", "80-84", "85-89", and "90+", which represent the mean annual mortality rates for each group and sex. **Also load the file "weightsMortalityRates_Swedish_2016.csv", which contains columns "age", "weight_male", and "weight_female", representing the actual sex distribution by single year of age from 60 to 99 (40 rows), not by age group.** First, expand the grouped mortality rates to individual ages (60-99), then compute the sex-weighted average mortality rate per age using the corresponding weights. Finally, create a data frame with one row per age (60-99) and the columns: "age", "rate_avg", "prob_death_ad_mci", "prob_death_mild", "prob_death_moderate", "prob_death_severe". For each age, compute the probability of death for each disease severity state using the formula: "p_death = 1 - exp(-rate * HR)", using already defined state-specific hazard ratios. Do not use any additional packages. Assume all files are in the working directory.” | Wimo final prompt. | Needed to be adapted to reflect the Ross model. |
| 2 | “(Prompt 2): Please write a R code that loads the "mortalityRates_US_2017.csv" located in the working directory. This CSV contains the following columns: "age" (ages from 60 to 99), "male" (male-specific mortality rates), and "female" (female-specific mortality rates). Create a new data frame with one row per age and the following columns: "age", "rate_avg", "prob_death_mci", "prob_death_mild", "prob_death_moderate", "prob_death_severe". For the average mortality rate across sexes ("rate_avg") assumes a 50/50 male-female distribution. Then, for each age, compute the age-specific probability of death for each disease severity state using the formula: "p_death = 1 - exp(-rate_avg * HR)", using already defined state-specific hazard ratios. These computed death probabilities will later be inserted into the transition probability matrix.” | Used annual mortality rates and assumed a 50/50 sex distribution. | The rate is manually converted in month and averaged by sex. |
| F<br>I<br>N<br>A<br>L | “(Prompt 2): Please write a R code that loads the "mortalityRates_US_2017.csv" located in the working directory. This CSV contains the following columns: "age" (ages from 60 to 99), **and "monthly_rate" (age-specific mortality rates).** Create a new data frame with one row per age and the following columns: "age", "prob_death_mci", "prob_death_mild", "prob_death_moderate", "prob_death_severe". For each age, compute the age-specific probability of death for each disease severity state using the formula: "p_death = 1 - exp(-rate_avg * HR)", using already defined state-specific hazard ratios. These computed death probabilities will later be inserted into the transition probability matrix.” |  |  |

**Table S2.13:** Model by Ross et al.: Overview of prompt 3 refinement process and rationale for adjustments.

| Trial | Prompt Evolution | Main Limitation | Reason for Modification |
| --- | --- | --- | --- |
| 1 | “(Prompt 3): Please write a R code that defines all time-dependent transition probability vectors for both the `no_intervention` and `intervention` strategy, without using `assign()` or `get()`, or dynamic variable names.<br>For the `no_intervention` strategy, define a transition probability vector for each cycle:<br>- From “`ad_mci`” to “`mild`” and staying in “`ad_mci`”.<br>- From “`mild`” to “`moderate`”, “`severe`” and staying in “`mild`”.<br>- From “`moderate`” to “`mild`”, “`severe`” and staying in “`moderate`”.<br>- From “`severe`” to “`moderate`” and staying in “`severe`”.<br>For the `intervention` strategy (base-case scenario with hypothetical DMT treatment applied during “`ad_mci`” and “`mild`” states only), apply the following formula to each affected transition:<br>- Convert the original probability to a rate: $\text{rate} = -\log(1 - p)$.<br>- Adjust the rate using the appropriate relative risk: $\text{adjusted\_rate} = \text{rate} * \text{RR}$.<br>- Convert the rate back to a probability: $\text{adjusted\_p} = 1 - \exp(-\text{adjusted\_rate})$.<br>Also define the staying probabilities per state for each strategy as: $1 - \text{sum}(\text{other transition probabilities from that state})$. Return only the R code block defining the vectors. Assume that all input probabilities and relative risks have been defined previously. Do not forget to use `rep()` to make all output vectors of length `n_cycles`.” | Wimo final prompt. | Needed to be adapted to reflect the Ross model. |
| F<br>I<br>N<br>A<br>L | “(Prompt 3): Please write a R code that defines all time-dependent transition probability vectors for both the `no_intervention` and `intervention` strategies (Aducanumab and Donanemab), without using `assign()` or `get()`, or dynamic variable names. For each strategy, define a vector of length `n_cycles` for each transition:<br>- From “`mci`” to “`mild`” and staying in “`mci`”.<br>- From “`mild`” to “`moderate`” and staying in “`mild`”.<br>- From “`moderate`” to “`severe`” and staying in “`moderate`”.<br>- From “`severe`” to staying in “`severe`”.<br>For the `intervention` strategies, apply the intervention effect (hazard ratio) to all progression transitions between disease stage, using the following transformation:<br>- Convert the original probability to a rate: $\text{rate} = -\log(1 - p)$.<br>- Adjust the rate using the appropriate hazard ratio: $\text{adjusted\_rate} = \text{rate} * \text{HR}$.<br>- Convert the rate back to a probability: $\text{adjusted\_p} = 1 - \exp(-\text{adjusted\_rate})$.<br>Define the staying probabilities per state for each strategy as: $1 - \text{sum}(\text{other transition probabilities from that state})$. Return only the R code block defining the vectors. Assume that all input probabilities and HRs have been defined previously. Do not forget to use `rep()` to make all output vectors of length `n_cycles`.” |  |  |

**Table S2.14:** Model by Ross et al.: Overview of prompt 4 refinement process and rationale for adjustments.

| Trial | Prompt Evolution | Main Limitation | Reason for Modification |
| --- | --- | --- | --- |
| 1 | **Prompt 4A:**<br>"(Prompt 4A): Please write a R code that initializes the transition probability array (dimensions: states × states × cycles) for the model. The function must be structured as `build_transition_matrix(strat)`, where `strat` is either no intervention or intervention. Inside the function:<br>- Initialize the array and loop over each cycle.<br>- For each cycle, explicitly assign the transition probabilities from each state to death, calculated in prompt 2, without using loops (Write one assignment line explicitly for each state). For the death state "dth", set the probability of remaining in this state to 1 at all cycles, since death is an absorbing state.<br>- Leave one clearly visible placeholder in the function body: "[INSERT PROMPT 4B HERE] — to define the transitions from on-treatment and off-treatment states later."<br>- After filling the transitions for each state at each cycle (4B), normalize the rows if necessary: if the row sum differs from 1, divide each element of the row by the row sum.<br>Do not yet define transition states here — only set up the structure and placeholder." | Wimo final prompt. | Needed to be adapted to reflect the Ross model. |
|  | **Prompt 4B:**<br>"(Prompt 4B): Please extend the previously initialized `build_transition_matrix(strat)` function by filling all transitions between the following states for each cycle: "ad_mci" to "mild" and "ad_mci" to "ad_mci" (staying), "mild" to "moderate", "severe", and "mild" (staying), "moderate" to "mild", "severe", and "moderate" (staying), "severe" to "moderate" and "severe" (staying). For each valid destination state, multiply the appropriate base transition probability by "(1 - mortality probability)" for all living state transitions. Do not repeat the initialization part of the function. Return only the R code for these transitions." | Wimo final prompt. | Needed to be adapted to reflect the Ross model. |
| 2 | **Prompt 4A:**<br>"(Prompt 4A): Please write a R code that initializes the transition probability array (dimensions: states × states × cycles) for the model. The function must be structured as `build_transition_matrix(strat)`, where `strat` is either no intervention, aducanumab or donanemab. Inside the function:<br>- Initialize the array and loop over each cycle.<br>- For each cycle, explicitly assign the transition probabilities from each state to death, calculated in prompt 2, without using loops (Write one assignment line explicitly for each state). For the death state "dth", set the probability of remaining in this state to 1 at all cycles, since death is an absorbing state.<br>- Leave one clearly visible placeholder in the function body: "[INSERT PROMPT 4B HERE] — define transitions to next disease stages (with or without treatment)."<br>- After filling the transitions for each state at each cycle (4B), normalize the rows if necessary: if the row sum differs from 1, divide each element of the row by the row sum.<br>Do not yet define transition states here — only set up the structure and placeholder." | Computation error<br>current age variable was no restricted to ages 60-99. | Explicitly restricts the age range to 60–99, aligning it with the available mortality data. |
|  | **Prompt 4B:**<br>"(Prompt 4B): Please extend the previously initialized `build transition matrix(strat)` function by filling all transitions between the following states for each cycle: "mci" to "mild" and "mci" (staying), "mild" to "moderate" and "mild" (staying), "moderate" to "severe" and "moderate" (staying), "severe" to "severe" (staying). For each valid destination state, multiply the appropriate base transition probability by "(1 - mortality probability)" for all living state transitions. Do not repeat the initialization part of the function. Return only the R code for these transitions." | Wimo final prompt. | Needed to be adapted to reflect the Ross model. |
| FIN | **Prompt 4A:**<br>"(Prompt 4A): Please write a R code that initializes the transition probability array (dimensions: states × states × cy- |  |  |
| A<br>L | cles) for the model. The function must be structured as build_transition_matrix(strat), where strat is either no intervention, aducanumab or donanemab. Inside the function:<br>- Initialize the array and loop over each cycle.<br>- For each cycle, assign the transition probabilities from each state to death, calculated in prompt 2. **These assignments must be written explicitly per state (no for-loops). Assume that the selected age is between 60 and 99.** For the death state, set the probability of staying in death to 1 at all cycles, since death is an absorbing state.<br>- Leave one clearly visible placeholder in the function body: "[INSERT PROMPT 4B HERE] — define transitions to next disease stages (with or without treatment)."<br>- After filling the transitions for each state at each cycle (4B), normalize the rows if necessary: if the row sum differs from 1, divide each element of the row by the row sum.<br>Do not yet define transition states here — only set up the structure and placeholder."<br>**Prompt 4B:**<br>“(Prompt 4B): Please extend the previously initialized build_transition_matrix(strat) function by filling all transitions between the following states for each cycle: "mci" to "mild" and "mci" (staying), "mild" to "moderate" and "mild" (staying), "moderate" to "severe" and "moderate" (staying), "severe" to "severe" (staying). For each valid destination state, multiply the appropriate base transition probability by "(1 - mortality probability)" for all living state transitions. Do not repeat the initialization part of the function. Return only the R code for these transitions." |  |  |

**Table S2.15:**
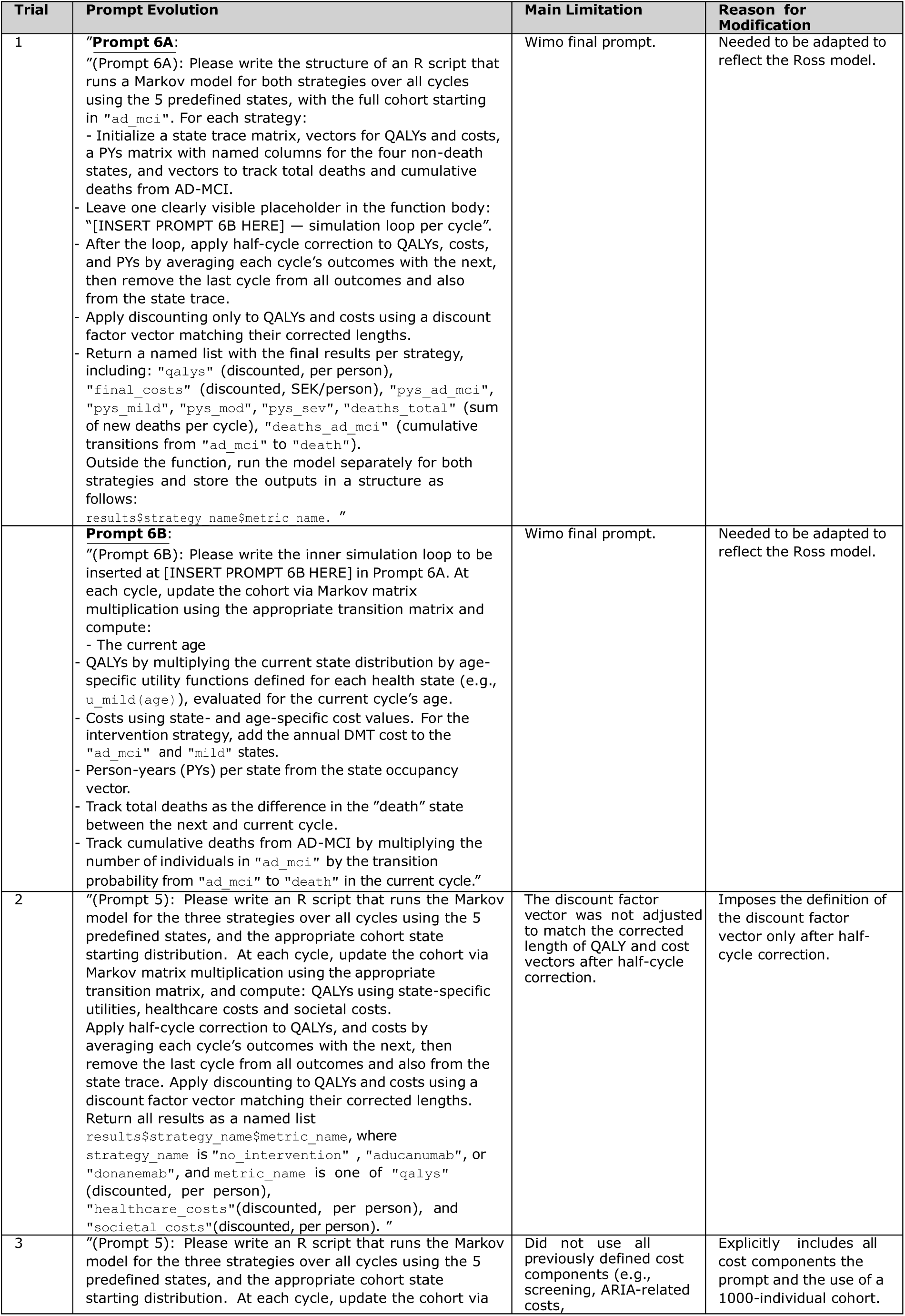
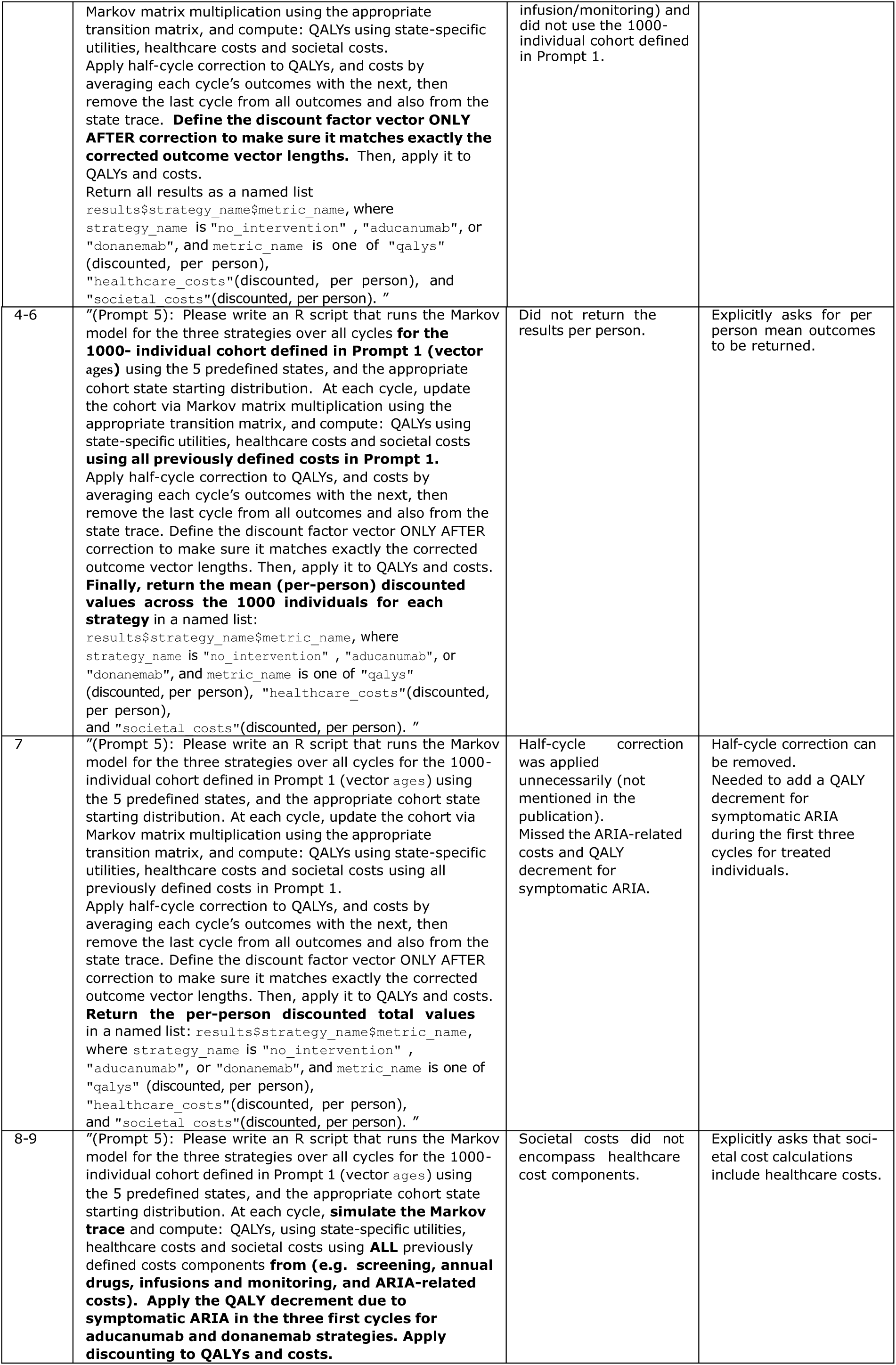
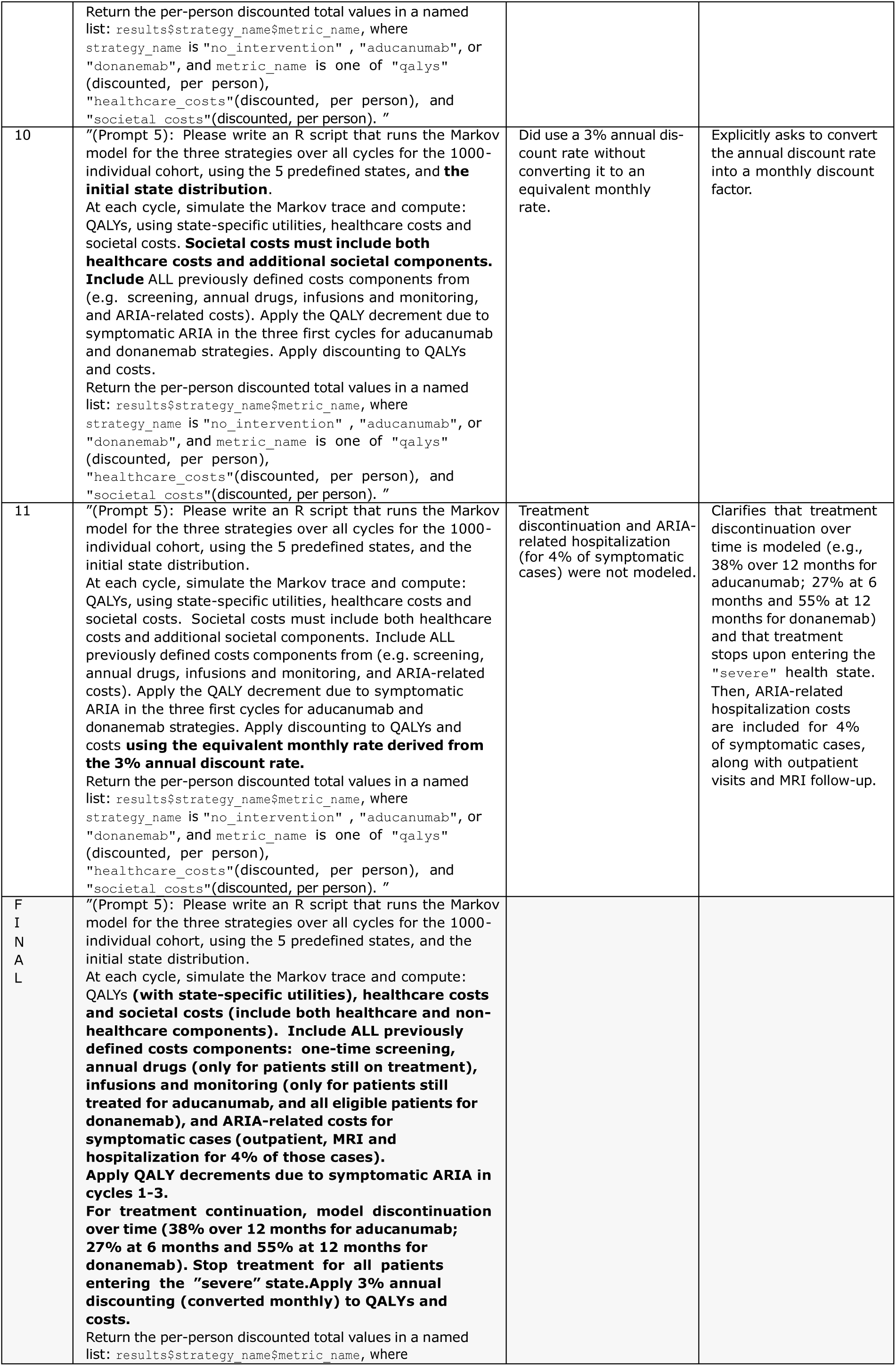
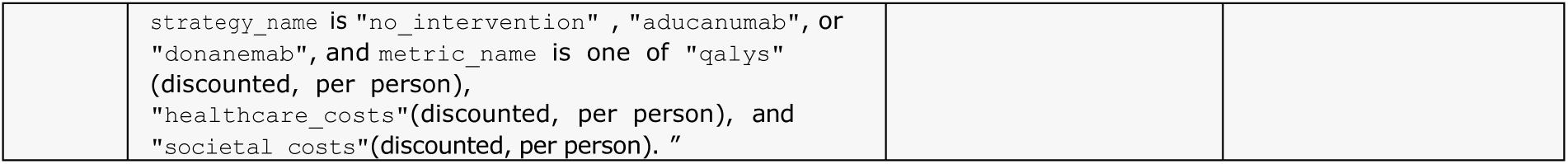
Model by Ross et al.: Overview of prompt 5 refinement process and rationale for adjustments.

## SUPPLEMENTARY MATERIAL 3: FINAL PROMPT SEQUENCE AND VALIDATION CRITERIA

**Figure S3.1:**
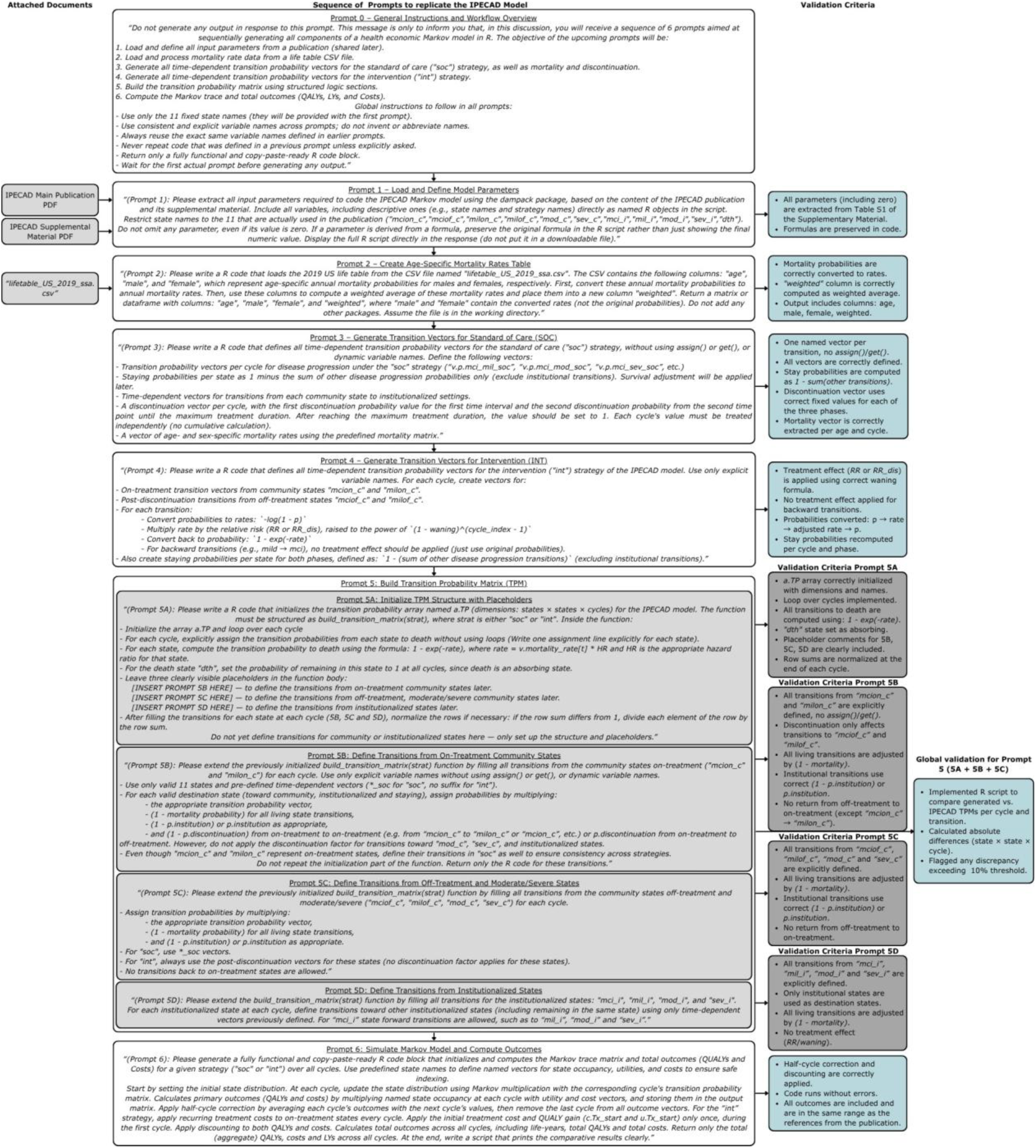
Final sequence of prompts to implement the IPECAD model, with associated files and validation process.

**Figure S3.2:**
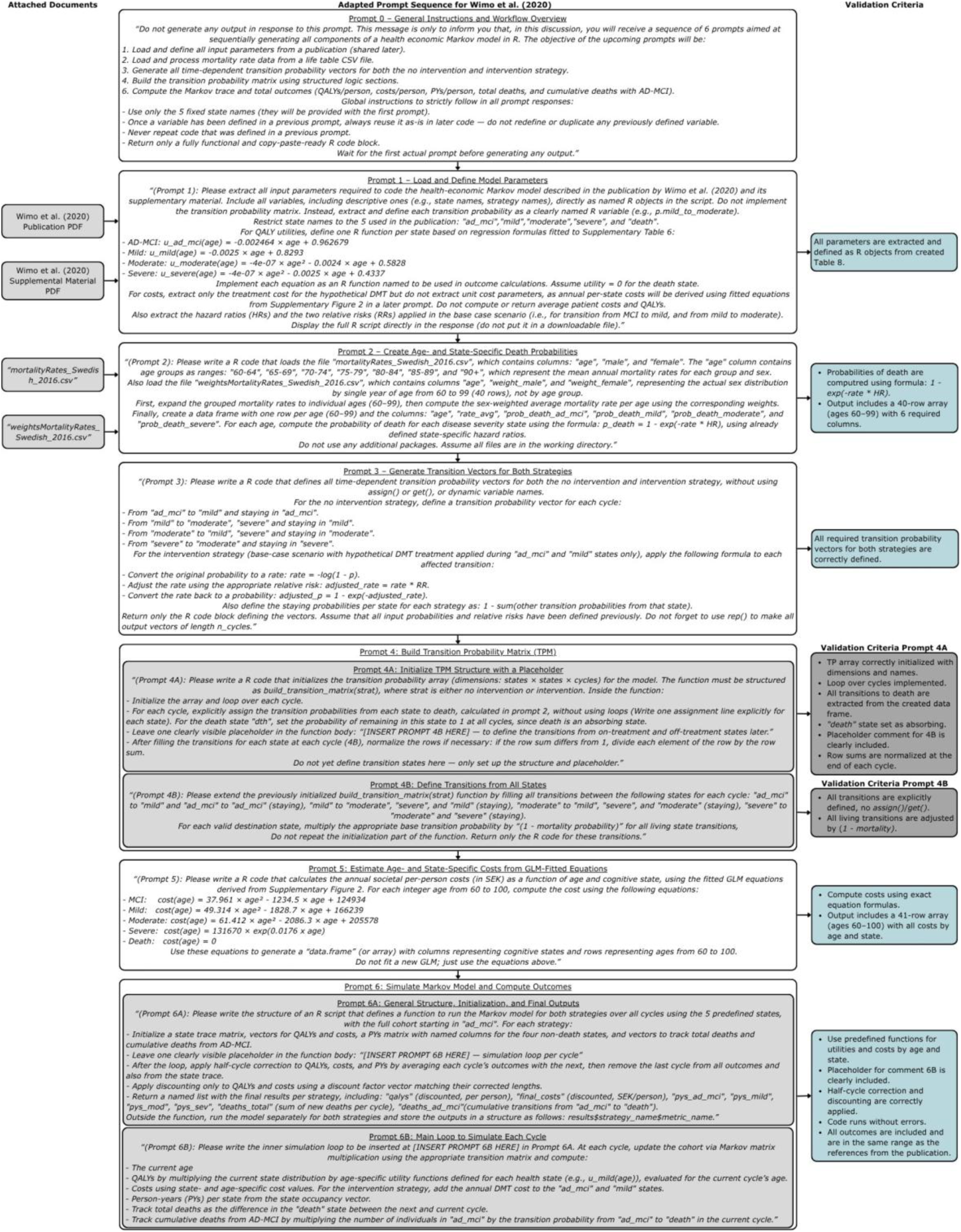
Final sequence of prompts to implement the Wimo et al. (2020) model, with associated files and validation process.

**Figure S3.3:**
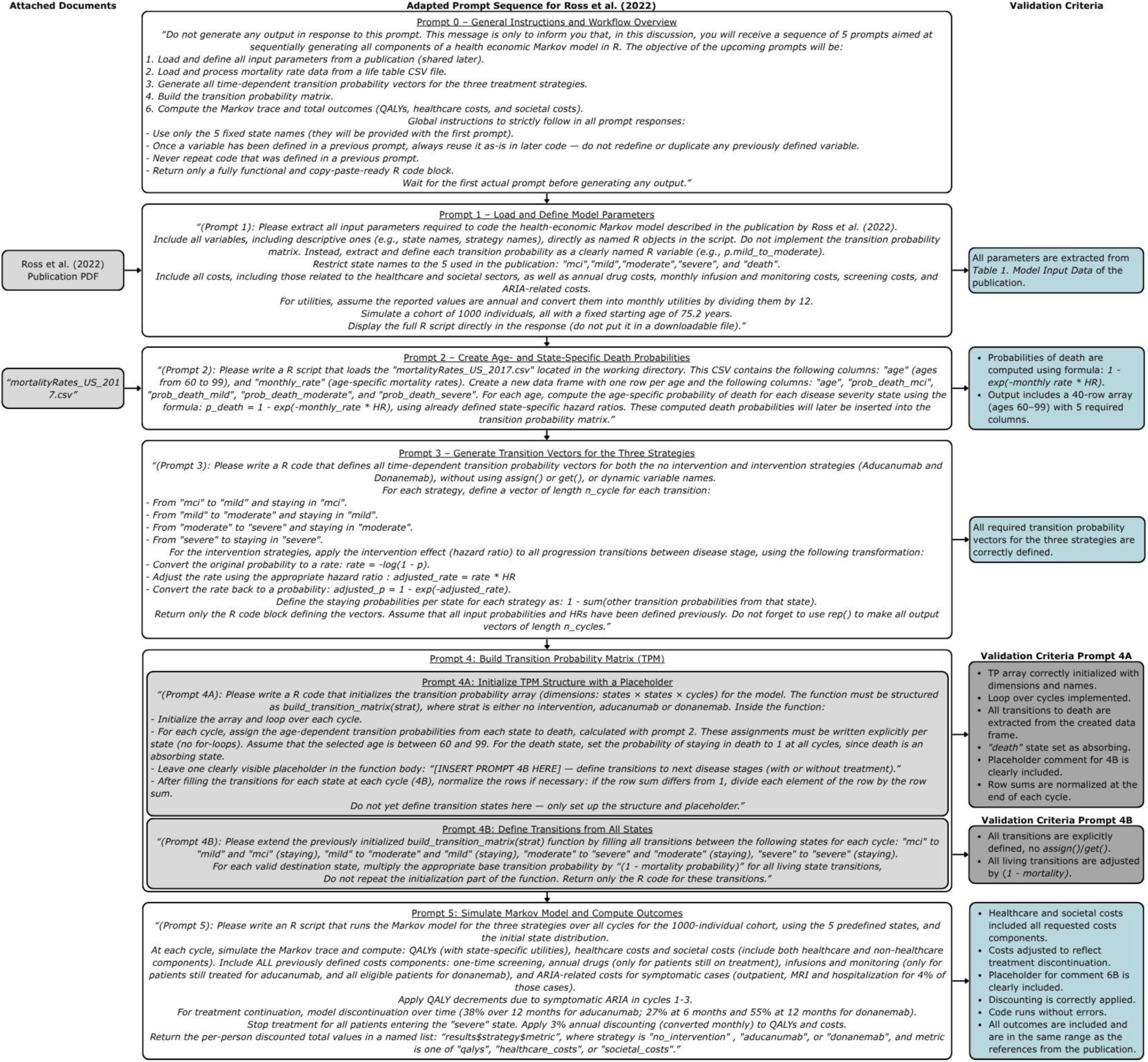
Final sequence of prompts to implement the Ross et al. (2022) model, with associated files and validation process.

## SUPPLEMENTARY MATERIAL 4: 15 FINAL R SCRIPTS

See supplemented zip file.

## SUPPLEMENTARY MATERIAL 5: EQUATIONS

Equations 1 and 2 define the formulas used to quantify the reproducibility of ChatGPT-4 generated model outputs across the fifteen simulations (n = 15).

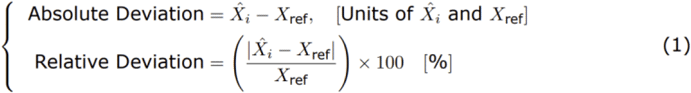

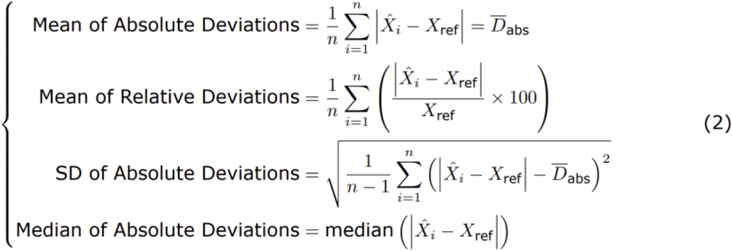

## SUPPLEMENTARY MATERIAL 6: PROMTP ADAPTATIONS

**Table S6.1:** Prompt adaptation and associated validation criteria for replicating the Wimo et al. (2020) model. Text in blue indicates assumptions made during adaptation.

| Wimo Prompt | IPECAD Prompt | Adaptation to Wimo et al. (2020) | Adapted Validation Criteria |
| --- | --- | --- | --- |
| 0 | 0 | Adjusted to reflect the actual aim of each prompt required for Wimo et al. (2020). |  |
| 1 | 1 | - Do not force the use of the *dampack* package.<br>- Use five health states: AD-MCI, Mild, Moderate, Severe, and Death.<br>- No institutionalization states.<br>- No treatment discontinuation or stopping rules.<br>- Utility functions were derived from Supplementary Table 6 (5-year intervals) to estimate utilities by age and health state. | Creation of a table (similar to Table S1 from the IPECAD supplementary material) summarizing the required model input parameters based on the Wimo publication. |
| 2 | 2 | - Use Swedish age- and sex-specific mean mortality rates (2000–2016) from Statistics Sweden.<br>- Sex proportions were extracted from Statistics Sweden (2000–2016) and provided to ChatGPT-4 into a CSV file.<br>- Compute transition probabilities from each health state to death using: $1 - \exp(-\text{age rate} * \text{HR})$. | The code must return a 40-row array (ages 60–99) with columns: sex-weighted rate and state-specific death probabilities. |
| 3 | 3 + 4 | - No discontinuation and treatment phases.<br>- No waning effect.<br>- Intervention only affects transitions from AD-MCI and Mild states. | Verification that all transition probability vectors are correctly defined. |
| 4<br>(4A, 4B) | 5<br>(5A-5D) | - Assumed the Death state is an absorbing state.<br>- No treatment discontinuation or transition toward institution factors in the calculation of transition probability vectors.<br>- Subprompts 5B and 5C were merged into now Prompt 4B.<br>- Subprompt 5D was removed as no institutionalization transitions. | Verification that the transition probability matrix is correctly defined with all transitions. |
| 5 | - | - New prompt created to extract age- and state-specific costs based on previously fitted GLM equations from Supplementary Figure 2. | The code must return a 40-row array (ages 60–99) with columns representing all health states. |
| 6<br>(6A, 6B) | 6 | - Selected final outcomes include: (1) total QALYs/person, (2) total costs (SEK/person), (3) person-years per health state, and (4) cumulative deaths and (5) deaths with AD- MCI. | Implementation of an R script comparing the generated outcomes to those reported in the publication, and computing absolute and relative differences. |

**Table S6.2:** Prompt adaptation and associated validation criteria for replicating the Ross et al. (2022) model. Text in blue indicates assumptions made during adaptation.

| Ross Prompt | Wimo Prompt | Adaptation to Ross et al. (2022) | Adapted Validation Criteria |
| --- | --- | --- | --- |
| 0 | 0 | Adjusted to reflect the actual aim of each prompt required for Ross et al. (2022). |  |
| 1 | 1 | - 3 treatment strategies: standard care, aducanumab, and donanemab.<br>- Assumed a cohort of 1000 individuals.<br>- Baseline age set to 75.2 years.<br>- Use 1-month cycle length.<br>- No backward transitions or multi-step transitions (only forward or stay).<br>- Provide full costing components: screening, drug, infusion/monitoring, and ARIA-related.<br>- Include utility decrement and ARIA-related adverse events.<br>- Provide all utilities by state.<br>- Supplementary Material is not needed for model replication. | Table 1. Model Input Data, provided into the Ross et al. (2022) publication, used to ensure all required parameters were extracted and correct. |
| 2 | 2 | - Use US Centers for Disease Control and Prevention life table.<br>- No distinction made between sexes. | The code must return a 40-row array (ages 60–99) with columns: age and state-specific death probabilities. |
| 3 | 3 | - No backward transitions or multi-step transitions (only forward or stay).<br>- Apply treatment effects (hazard ratios) to all forward transitions.<br>- Use hazard ratio (HR) to reduce the disease progression for the interventions.<br>- Interventions affect all transitions. | Verification that all transition probability vectors are correctly defined. |
| 4<br>(4A, 4B) | 4<br>(4A, 4B) | - Assumed the “death” state is an absorbing state. | Verification that the transition probability matrix is correctly defined with all transitions. |
| 5 | 6<br>(6A, 6B) | - At baseline, 65% of patients had MCI and 35% had mild dementia.<br>- Apply treatment discontinuation patterns for cost calculations. **Aducanumab**: linear tapering to 62% over 12 months. **Donanemab**: stepwise discontinuation at months 6 and 12.<br>- Treatment costs are discontinued upon progression to severe dementia.<br>- No half-cycle correction applied.<br>- Selected final outcomes include: (1) total QALYs/person,<br>(2) total healthcare costs (USD/person), (3) total societal costs (USD/person). | Implementation of an R script comparing the generated outcomes to those reported in the publication, and computing absolute and relative differences. |

## SUPPLEMENTARY MATERIAL 7: INDIVIDUAL RESULTS

**Table S7.1:** Detailed Simulation Results: Values, and Absolute (”Abs”) and Relative (”Rel%”) Differences of Model Predictions Compared to IPECAD Reference.

| QALYs |  |  |  |  |  |  |
| --- | --- | --- | --- | --- | --- | --- |
| Simulation | Standard Of Care |  |  | Intervention |  |  |
|  | Val | Abs | Rel% | Val | Abs | Rel% |
| IPECAD (Reference) | 2.94 | NA | NA | 3.29 | NA | NA |
| 1 | 2.85 | -0.09 | 3 | 3.19 | -0.10 | 3 |
| 2 | 2.86 | -0.08 | 3 | 3.18 | -0.11 | 3 |
| 3 | 3.50 | 0.56 | 19 | 3.73 | 0.44 | 13 |
| 4 | 3.32 | 0.38 | 13 | 3.64 | 0.35 | 11 |
| 5 | 2.95 | 0.01 | 0 | 3.20 | -0.09 | 3 |
| 6 | 2.90 | -0.04 | 1 | 3.23 | -0.06 | 2 |
| 7 | 3.53 | 0.59 | 20 | 3.12 | -0.17 | 5 |
| 8 | 3.77 | 0.83 | 28 | 4.00 | 0.71 | 22 |
| 9 | 3.86 | 0.92 | 31 | 4.08 | 0.79 | 24 |
| 10 | 3.49 | 0.55 | 19 | 3.72 | 0.43 | 13 |
| 11 | 3.35 | 0.41 | 14 | 3.62 | 0.33 | 10 |
| 12 | 4.02 | 1.08 | 37 | 4.24 | 0.95 | 29 |
| 13 | 2.79 | -0.15 | 5 | 3.56 | 0.27 | 8 |
| 14 | 3.38 | 0.44 | 15 | 3.61 | 0.32 | 10 |
| 15 | 2.86 | -0.08 | 3 | 2.86 | -0.43 | 13 |
| Costs (USD) |  |  |  |  |  |  |
| Simulation | Standard Of Care |  |  | Intervention |  |  |
|  | Val | Abs | Rel% | Val | Abs | Rel% |
| IPECAD (Reference) | 484,963 | NA | NA | 580,334 | NA | NA |
| 1 | 470,838 | -14,125 | 3 | 562,842 | -17,492 | 3 |
| 2 | 465,903 | -19,060 | 4 | 552,612 | -27,721 | 5 |
| 3 | 444,403 | -40,560 | 8 | 512,352 | -67,982 | 12 |
| 4 | 483,082 | -1,881 | 0 | 569,672 | -10,662 | 2 |
| 5 | 475,542 | -9,421 | 2 | 555,304 | -25,029 | 4 |
| 6 | 475,729 | -9,234 | 2 | 563,876 | -16,458 | 3 |
| 7 | 454,383 | -30,580 | 6 | 565,084 | -15,250 | 3 |
| 8 | 453,179 | -31,784 | 7 | 528,637 | -51,697 | 9 |
| 9 | 453,333 | -31,630 | 7 | 519,074 | -61,260 | 11 |
| 10 | 448,098 | -36,865 | 8 | 516,120 | -64,213 | 11 |
| 11 | 492,280 | 7,317 | 2 | 580,953 | 619 | 0 |
| 12 | 465,847 | -19,116 | 4 | 532,458 | -47,876 | 8 |
| 13 | 412,238 | -72,725 | 15 | 500,660 | -79,674 | 14 |
| 14 | 490,543 | 5,580 | 1 | 581,521 | 1,187 | 0 |
| 15 | 465,903 | -19,060 | 4 | 466,492 | -113,842 | 20 |
| Life Years |  |  |  |  |  |  |
| Simulation | Standard Of Care |  |  | Intervention |  |  |
|  | Val | Abs | Rel% | Val | Abs | Rel% |
| IPECAD (Reference) | 6.15 | NA | NA | 6.50 | NA | NA |
| 1 | 5.98 | -0.17 | 3 | 6.31 | -0.19 | 3 |
| 2 | 5.98 | -0.17 | 3 | 6.29 | -0.21 | 3 |
| 3 | 6.60 | 0.45 | 7 | 6.83 | 0.33 | 5 |
| 4 | 19.26 | 13.11 | 213 | 19.26 | 12.76 | 196 |
| 5 | 6.15 | 0.00 | 0 | 6.39 | -0.11 | 2 |
| 6 | 6.03 | -0.12 | 2 | 6.34 | -0.16 | 2 |
| 7 | 6.65 | 0.50 | 8 | 6.24 | -0.26 | 4 |
| 8 | 7.01 | 0.86 | 14 | 7.23 | 0.73 | 11 |
| 9 | 6.62 | 0.47 | 8 | 6.84 | 0.34 | 5 |
| 10 | 6.60 | 0.45 | 7 | 6.83 | 0.33 | 5 |
| 11 | 19.55 | 13.40 | 218 | 19.55 | 13.05 | 201 |
| 12 | 6.87 | 0.72 | 12 | 7.09 | 0.59 | 9 |
| 13 | 6.50 | 0.44 | 7 | 6.50 | 0.00 | 0 |
| 14 | 6.22 | 0.07 | 1 | 6.45 | -0.05 | 1 |
| 15 | 5.98 | -0.17 | 3 | 6.80 | 0.52 | 8 |

**Table S7.2:** Detailed Simulation Results: Values, Absolute (”Abs”) and Relative (”Rel%”) Differences Compared to Wimo et al. (2020) Reference.

| QALYs |  |  |  |  |  |  | Costs (SEK) |  |  |  |  |  |
| --- | --- | --- | --- | --- | --- | --- | --- | --- | --- | --- | --- | --- |
| Simulation | No Intervention |  |  | Intervention |  |  | No Intervention |  |  | Intervention |  |  |
|  | Val | Abs | Rel% | Val | Abs | Rel% | Val | Abs | Rel% | Val | Abs | Rel % |
| Wimo (Reference) | 7.90 | NA | NA | 8.63 | NA | NA | 4,503,751 | NA | NA | 4,893,703 | NA | NA |
| 1 | 7.74 | -0.16 | 2 | 8.37 | -0.26 | 3 | 4,253,208 | -250,543 | 6 | 4,642,112 | -251,591 | 5 |
| 2 | 7.51 | -0.39 | 5 | 8.13 | -0.50 | 6 | 4,129,328 | -374,423 | 8 | 4,506,905 | -386,798 | 8 |
| 3 | 7.73 | -0.17 | 2 | 8.37 | -0.26 | 3 | 4,253,464 | -250,287 | 6 | 4,642,169 | -251,534 | 5 |
| 4 | 7.74 | -0.16 | 2 | 8.37 | -0.26 | 3 | 4,253,208 | -250,543 | 6 | 4,642,112 | -251,591 | 5 |
| 5 | 7.51 | -0.39 | 5 | 8.13 | -0.50 | 6 | 4,129,328 | -374,423 | 8 | 4,506,905 | -386,798 | 8 |
| 6 | 7.33 | -0.57 | 7 | 7.97 | -0.66 | 8 | 4,159,446 | -344,305 | 8 | 4,523,351 | -370,352 | 8 |
| 7 | 7.74 | -0.16 | 2 | 8.37 | -0.26 | 3 | 4,253,208 | -250,543 | 6 | 4,642,112 | -251,591 | 5 |
| 8 | 7.73 | -0.17 | 2 | 8.37 | -0.26 | 3 | 4,252,595 | -251,156 | 6 | 4,640,537 | -253,166 | 5 |
| 9 | 6.95 | -0.95 | 12 | 7.66 | -0.97 | 11 | 4,028,832 | -474,919 | 11 | 4,392,498 | -501,205 | 10 |
| 10 | 7.51 | -0.39 | 5 | 8.13 | -0.50 | 6 | 4,129,328 | -374,423 | 8 | 4,506,905 | -386,798 | 8 |
| 11 | 7.36 | -0.54 | 7 | 8.00 | -0.63 | 7 | 4,085,790 | -417,781 | 9 | 4,450,568 | -443,135 | 9 |
| 12 | 7.51 | -0.39 | 5 | 8.13 | -0.50 | 6 | 4,129,328 | -374,423 | 8 | NA | NA | NA |
| 13 | 7.73 | -0.17 | 2 | 8.37 | -0.26 | 3 | 4,253,464 | -250,287 | 6 | 4,642,169 | -251,534 | 5 |
| 14 | 7.69 | -0.21 | 3 | 8.29 | -0.34 | 4 | 4,100,807 | -403,664 | 9 | 4,490,939 | -402,764 | 8 |
| 15 | 7.73 | -0.17 | 2 | 8.37 | -0.26 | 3 | 4,253,464 | -250,287 | 6 | 4,642,169 | -251,534 | 5 |
| Person years/person in AD-MCI |  |  |  |  |  |  | Person years/person in Mild AD |  |  |  |  |  |
| Simulation | No Intervention |  |  | Intervention |  |  | No Intervention |  |  | Intervention |  |  |
|  | Val | Abs | Rel% | Val | Abs | Rel% | Val | Abs | Rel% | Val | Abs | Rel% |
| Wimo (Reference) | 4.20 | NA | NA | 5.64 | NA | NA | 4.97 | NA | NA | 8.83 | NA | NA |
| 1 | 4.17 | -0.03 | 1 | 5.42 | -0.22 | 4 | 4.75 | -0.22 | 5 | 5.42 | -0.41 | 7 |
| 2 | 4.17 | -0.03 | 1 | 5.42 | -0.22 | 4 | 4.75 | -0.22 | 5 | 5.42 | -0.41 | 7 |
| 3 | 4.16 | -0.04 | 1 | 5.41 | -0.23 | 4 | 4.75 | -0.22 | 4 | 5.42 | -0.41 | 7 |
| 4 | 4.17 | -0.03 | 1 | 5.42 | -0.22 | 4 | 4.75 | -0.22 | 5 | 5.42 | -0.41 | 7 |
| 5 | 4.17 | -0.03 | 1 | 5.42 | -0.22 | 4 | 4.75 | -0.22 | 5 | 5.42 | -0.41 | 7 |
| 6 | 3.67 | -0.53 | 13 | 4.92 | -0.72 | 13 | 4.75 | -0.22 | 5 | 5.42 | -0.41 | 7 |
| 7 | 4.17 | -0.03 | 1 | 5.42 | -0.22 | 4 | 4.75 | -0.22 | 5 | 5.42 | -0.41 | 7 |
| 8 | 4.16 | -0.04 | 1 | 5.41 | -0.23 | 4 | 4.75 | -0.22 | 5 | 5.42 | -0.41 | 7 |
| 9 | 4.17 | -0.03 | 1 | 5.42 | -0.22 | 4 | 3.10 | 1.87 | 38 | 3.75 | -2.08 | 36 |
| 10 | 4.17 | -0.03 | 1 | 5.42 | -0.22 | 4 | 4.75 | -0.22 | 5 | 5.42 | -0.41 | 7 |
| 11 | 3.66 | -0.54 | 13 | 4.91 | -0.73 | 13 | 4.75 | -0.22 | 4 | 5.42 | -0.41 | 7 |
| 12 | 4.17 | -0.03 | 1 | 5.42 | -0.22 | 4 | 4.75 | -0.22 | 5 | 5.42 | -0.41 | 7 |
| 13 | 4.16 | -0.04 | 1 | 5.41 | -0.23 | 4 | 4.75 | -0.22 | 4 | 5.42 | -0.41 | 7 |
| 14 | 4.67 | 0.47 | 11 | 5.92 | -0.28 | 5 | 4.75 | -0.22 | 5 | 5.42 | -0.41 | 7 |
| 15 | 4.16 | -0.04 | 1 | 5.41 | -0.23 | 4 | 4.75 | -0.22 | 4 | 5.42 | -0.41 | 7 |
| Person years/person in Moderate AD |  |  |  |  |  |  | Person years /person in Severe AD |  |  |  |  |  |
| Simulation | No Intervention |  |  | Intervention |  |  | No Intervention |  |  | Intervention |  |  |
|  | Val | Abs | Rel% | Val | Abs | Rel% | Val | Abs | Rel% | Val | Abs | Rel% |
| Wimo (Reference) | 7.31 | NA | NA | 6.14 | NA | NA | 2.50 | NA | NA | 2.02 | NA | NA |
| 1 | 6.74 | -0.57 | 8 | 5.77 | -0.37 | 6 | 2.23 | -0.27 | 11 | 1.85 | -0.17 | 8 |
| 2 | 6.74 | -0.57 | 8 | 5.77 | -0.37 | 6 | 2.23 | -0.27 | 11 | 1.85 | -0.17 | 8 |
| 3 | 6.75 | -0.56 | 8 | 5.77 | -0.37 | 6 | 2.23 | -0.27 | 11 | 1.85 | -0.17 | 8 |
| 4 | 6.74 | -0.57 | 8 | 5.77 | -0.37 | 6 | 2.23 | -0.27 | 11 | 1.85 | -0.17 | 8 |
| 5 | 6.74 | -0.57 | 8 | 5.77 | -0.37 | 6 | 2.23 | -0.27 | 11 | 1.85 | -0.17 | 8 |
| 6 | 6.74 | -0.57 | 8 | 5.77 | -0.37 | 6 | 2.23 | -0.27 | 11 | 1.85 | -0.17 | 8 |
| 7 | 6.74 | -0.57 | 8 | 5.77 | -0.37 | 6 | 2.23 | -0.27 | 11 | 1.85 | -0.17 | 8 |
| 8 | 6.74 | -0.57 | 8 | 5.77 | -0.37 | 6 | 2.23 | -0.27 | 11 | 1.85 | -0.17 | 8 |
| 9 | 5.16 | -2.15 | 29 | 4.63 | -1.51 | 25 | 3.73 | 1.22 | 49 | 3.11 | 1.09 | 54 |
| 10 | 6.74 | -0.57 | 8 | 5.77 | -0.37 | 6 | 2.23 | -0.27 | 11 | 1.85 | -0.17 | 8 |
| 11 | 6.75 | -0.56 | 8 | 5.77 | -0.37 | 6 | 2.23 | -0.27 | 11 | 1.85 | -0.17 | 8 |
| 12 | 6.74 | -0.57 | 8 | 5.77 | -0.37 | 6 | 2.23 | -0.27 | 11 | 1.85 | -0.17 | 8 |
| 13 | 6.75 | -0.56 | 8 | 5.77 | -0.37 | 6 | 2.23 | -0.27 | 11 | 1.85 | -0.17 | 8 |
| 14 | 6.74 | -0.57 | 8 | 5.77 | -0.37 | 6 | 2.23 | -0.27 | 11 | 1.85 | -0.17 | 8 |
| 15 | 6.75 | -0.56 | 8 | 5.77 | -0.37 | 6 | 2.23 | -0.27 | 11 | 1.85 | -0.17 | 8 |
| Cumulative Deaths |  |  |  |  |  |  | Cumulative Deaths With AD-MCI |  |  |  |  |  |
| Simulation | No Intervention |  |  | Intervention |  |  | No Intervention |  |  | Intervention |  |  |
|  | Val | Abs | Rel% | Val | Abs | Rel% | Val | Abs | Rel% | Val | Abs | Rel% |
| Wimo (Reference) | 99,987.00 | NA | NA | 99,969.00 | NA | NA | 4,030.00 | NA | NA | 6,473.00 | NA | NA |
| 1 | 99,978.24 | -8.76 | 0 | 99,957.41 | -11.59 | 0 | 4,754.10 | 724.10 | 18 | 7,130.39 | 657.39 | 10 |
| 2 | 99,963.88 | -23.12 | 0 | 99,932.66 | -36.34 | 0 | 4,754.02 | 724.02 | 18 | 7,129.61 | 656.61 | 10 |
| 3 | 99,978.26 | -8.74 | 0 | 99,957.50 | -11.50 | 0 | 4,743.68 | 713.68 | 18 | 7,113.40 | 640.40 | 10 |
| 4 | 99,978.24 | -8.76 | 0 | 99,957.41 | -11.59 | 0 | 4,754.10 | 724.10 | 18 | 7,130.39 | 657.39 | 10 |
| 5 | 99,978.24 | -8.76 | 0 | 99,957.41 | -11.59 | 0 | 4,754.10 | 724.10 | 18 | 7,130.39 | 657.39 | 10 |
| 6 | 99,963.88 | -23.12 | 0 | 99,932.66 | -36.34 | 0 | 4,754.02 | 724.02 | 19 | 7,129.61 | 656.61 | 10 |
| 7 | 99,224.76 | -762.24 | 1 | 99,203.94 | -765.06 | 1 | 4,000.62 | -29.38 | 1 | 6,376.92 | -96.08 | 1 |
| 8 | 99,964.30 | -22.70 | 0 | 99,935.44 | -33.56 | 0 | 4,774.83 | 744.83 | 18 | 7,173.73 | 700.73 | 11 |
| 9 | 99,997.29 | 10.29 | 0 | 99,984.65 | 15.65 | 0 | 4,754.02 | 724.02 | 18 | 7,129.61 | 656.61 | 10 |
| 10 | 99,224.76 | -762.24 | 1 | 99,203.94 | -765.06 | 1 | 4,000.62 | -29.38 | 1 | 6,376.92 | -96.08 | 1 |
| 11 | 99,963.92 | -23.08 | 0 | 99,932.79 | -36.21 | 0 | 4,743.60 | 713.60 | 18 | 7,112.63 | 639.63 | 10 |
| 12 | 99,978.24 | -8.76 | 0 | 99,957.41 | -11.59 | 0 | 4,754.10 | 724.10 | 18 | 7,130.39 | 657.39 | 10 |
| 13 | 99,963.92 | -23.08 | 0 | 99,932.79 | -36.21 | 0 | 4,743.60 | 713.60 | 18 | 7,112.63 | 639.63 | 10 |
| 14 | 99,978.24 | -8.76 | 0 | 99,957.41 | -11.59 | 0 | 4,754.10 | 724.10 | 18 | 7,130.39 | 657.39 | 10 |
| 15 | 99,963.92 | -23.08 | 0 | 99,932.79 | -36.21 | 0 | 4,743.60 | 713.60 | 18 | 7,112.63 | 639.63 | 10 |

**Table S7.3:**
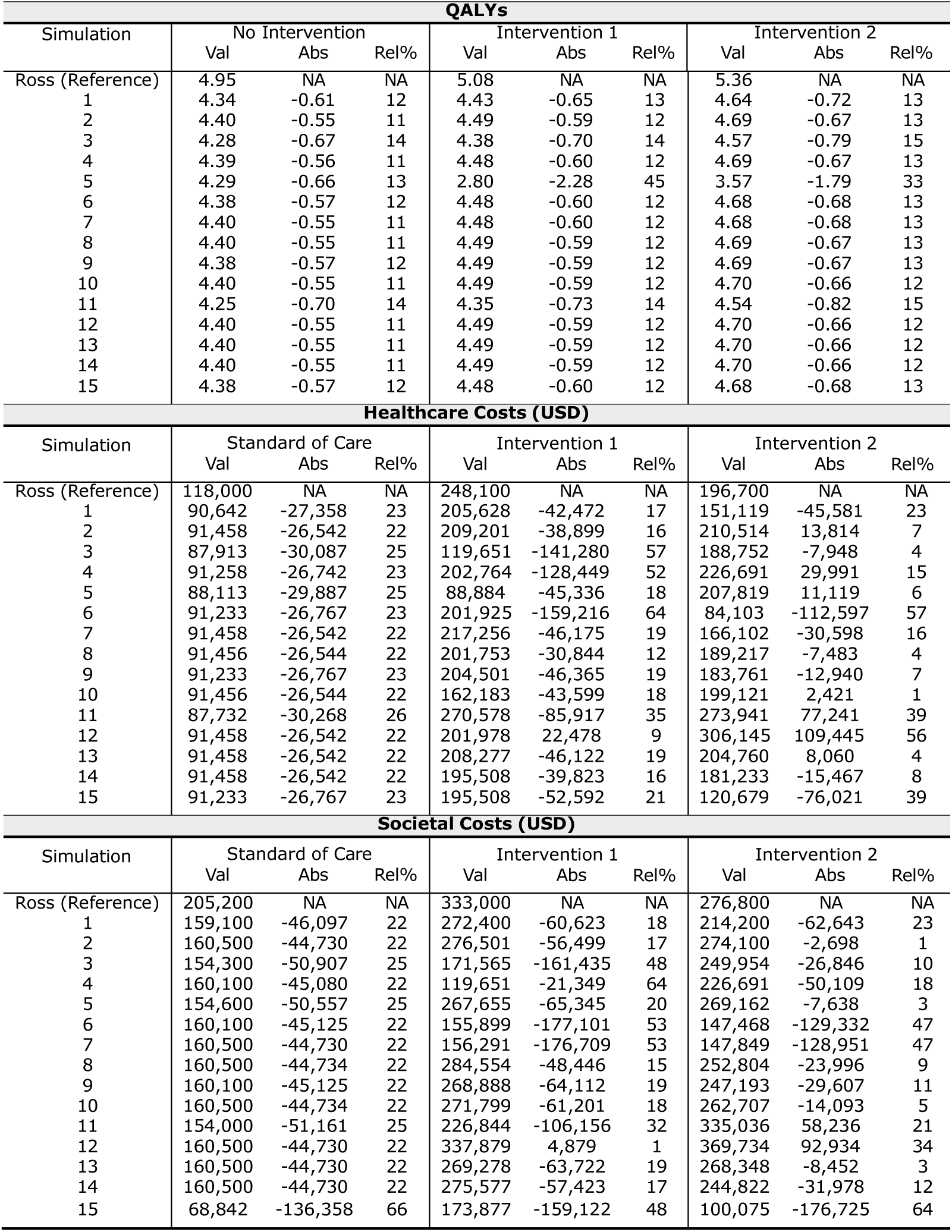
Detailed Simulation Results: Values, Absolute (”Abs”) and Relative (”Rel%”) Differences Compared to Ross et al. (2022) Reference.

