## Supplementary material for "Replicating Health-Economic Simulation Models for Alzheimer’s Disease Using Artificial Intelligence": final R scripts: o2.R

```
# Load required library
library(dampack)

# Define state names
state_names <- c("mcion_c", "mciof_c", "milon_c", "milof_c", "mod_c", "sev_c",
                 "mci_i", "mil_i", "mod_i", "sev_i", "dth")

# Define strategy names
strategy_names <- c("soc", "int")

# Demographic parameters
age_start <- 71
sex_proportion_female <- 0.52  # "weighted" sex for life table
sex_proportion_male <- 1 - sex_proportion_female

# Simulation settings
n_cycles <- 29
discount_EFFECT <- 0.03
discount_QALY <- 0.03
discount_COST <- 0.03
half_cycle_correction <- TRUE
wtp <- 100000

# Initial distribution
p_starting_state_mci <- 0.55
p_starting_state_mil <- 1 - p_starting_state_mci

# Transition probabilities for disease progression
p_mci_mil <- 0.23
p_mci_mod <- 0
p_mci_sev <- 0
p_mil_mci <- 0.03
p_mil_mod <- 0.35
p_mil_sev <- 0.04
p_mod_mil <- 0.03
p_mod_sev <- 0.42
p_sev_mil <- 0
p_sev_mod <- 0.02

# Transition probabilities to institution care
p_mci_i <- 0.024
p_mil_i <- 0.038
p_mod_i <- 0.110
p_sev_i <- 0.259

# Mortality hazard ratios
hr_mort_mci <- 1.82
hr_mort_mil <- 2.92
hr_mort_mod <- 3.85
hr_mort_sev <- 9.52

# Treatment effect (relative risk) on transitions (on-treatment)
rr_tx_mci_mil <- 0.69
rr_tx_mci_mod <- 1
rr_tx_mci_sev <- 1
rr_tx_mil_mod <- 0.69
rr_tx_mil_sev <- 0.69

# Treatment effect after discontinuation (off-treatment)
rr_tx_mci_mil_dis <- 1
rr_tx_mci_mod_dis <- 1
rr_tx_mci_sev_dis <- 1
rr_tx_mil_mod_dis <- 1
rr_tx_mil_sev_dis <- 1

# Treatment discontinuation
p_discontinuation1 <- 0.069
p_discontinuation2 <- 0
discontinuation2_begin <- 2
tx_duration <- 29

# Treatment waning (relative reduction in effect per cycle)
tx_waning <- 0
tx_waning_dis <- 0

# Utilities: patient (community)
u_mci_pt <- 0.851 - 0.17
u_mil_pt <- 0.851 - 0.22
u_mod_pt <- 0.851 - 0.36
u_sev_pt <- 0.851 - 0.53

# Utilities: patient (institution)
u_mci_pt_i <- 0.851 - 0.17
u_mil_pt_i <- 0.851 - 0.19
u_mod_pt_i <- 0.851 - 0.42
u_sev_pt_i <- 0.851 - 0.59

# Utilities: caregiver (community)
u_mci_ic <- -0.03
u_mil_ic <- -0.05
u_mod_ic <- -0.08
u_sev_ic <- -0.10

# Utilities: caregiver (institution)
u_mci_ic_i <- -0.03
u_mil_ic_i <- -0.05
u_mod_ic_i <- -0.08
u_sev_ic_i <- -0.10

# Additional disutility for ARIA (treatment-related)
u_Tx_start <- -0.14 * (12 / 52) * 0.035

# Treatment cost
c_Tx <- 26500 + (52 / 2) * 78.35

# Additional first-year monitoring/adverse event costs
c_Tx_start <- 261.10 * 4 + 261.10 * 3 * 0.215

# Cost functions (healthcare, social, informal) use original formulas where possible
# Healthcare costs (community)
c_mci_hc <- 6042 * 1.12 + 460
c_mil_hc <- 6042 * 1.56 + 965 + 0.21 * 365 * 0.333
c_mod_hc <- 6042 * 1.93 + 1544 + 0.66 * 365 * 0.333
c_sev_hc <- 6042 * 1.93 + 1930

# Healthcare costs (institution)
c_mci_i_hc <- c_mci_hc
c_mil_i_hc <- c_mil_hc
c_mod_i_hc <- c_mod_hc
c_sev_i_hc <- c_sev_hc

# Social care costs (community): set to zero
c_mci_sc <- 0
c_mil_sc <- 0
c_mod_sc <- 0
c_sev_sc <- 0

# Social care costs (institution)
c_mci_i_sc <- 7394 * 12
c_mil_i_sc <- 7394 * 12
c_mod_i_sc <- 7394 * 12
c_sev_i_sc <- 7394 * 12

# Informal care costs (community)
c_mci_ic <- 69 * 12 * 32.46 + 0.204 * 0.049 * 20 * 52 * 32.46
c_mil_ic <- 113 * 12 * 32.46 + 0.112 * 0.086 * 20 * 52 * 32.46
c_mod_ic <- 169 * 12 * 32.46
c_sev_ic <- 298 * 12 * 32.46

# Informal care costs (institution)
c_mci_i_ic <- 69 * 12 * 32.46 * 0.44 + 0.204 * 0.049 * 20 * 52 * 32.46
c_mil_i_ic <- 113 * 12 * 32.46 * 0.44 + 0.112 * 0.086 * 20 * 52 * 32.46
c_mod_i_ic <- 169 * 12 * 32.46 * 0.44
c_sev_i_ic <- 298 * 12 * 32.46 * 0.44


# Load the life table CSV file
life_table <- read.csv("lifetable_US_2019_ssa.csv")

# Convert annual mortality probabilities to annual mortality rates
# Using the standard formula: rate = -log(1 - probability)
life_table$male <- -log(1 - life_table$male)
life_table$female <- -log(1 - life_table$female)

# Weighted average based on sex distribution in starting population (52% female, 48% male)
prop_female <- 0.52
prop_male <- 1 - prop_female

# Compute the weighted mortality rate
life_table$weighted <- prop_male * life_table$male + prop_female * life_table$female

# Return the updated data frame with selected columns
life_table <- life_table[, c("age", "male", "female", "weighted")]

# Display the resulting data frame
life_table


# Define the number of cycles
n_cycles <- 29

# Create time-dependent transition probability vectors for disease progression (SOC strategy)
v.p.mci_mil_soc <- rep(p_mci_mil, n_cycles)
v.p.mci_mod_soc <- rep(p_mci_mod, n_cycles)
v.p.mci_sev_soc <- rep(p_mci_sev, n_cycles)

v.p.mil_mci_soc <- rep(p_mil_mci, n_cycles)
v.p.mil_mod_soc <- rep(p_mil_mod, n_cycles)
v.p.mil_sev_soc <- rep(p_mil_sev, n_cycles)

v.p.mod_mil_soc <- rep(p_mod_mil, n_cycles)
v.p.mod_sev_soc <- rep(p_mod_sev, n_cycles)

v.p.sev_mil_soc <- rep(p_sev_mil, n_cycles)
v.p.sev_mod_soc <- rep(p_sev_mod, n_cycles)

# Staying probabilities for each disease state (excluding institutional and mortality transitions)
v.p.mci_stay_soc <- 1 - (v.p.mci_mil_soc + v.p.mci_mod_soc + v.p.mci_sev_soc)
v.p.mil_stay_soc <- 1 - (v.p.mil_mci_soc + v.p.mil_mod_soc + v.p.mil_sev_soc)
v.p.mod_stay_soc <- 1 - (v.p.mod_mil_soc + v.p.mod_sev_soc)
v.p.sev_stay_soc <- 1 - (v.p.sev_mil_soc + v.p.sev_mod_soc)

# Time-dependent vectors for transitions to institutional care
v.p.mci_i <- rep(p_mci_i, n_cycles)
v.p.mil_i <- rep(p_mil_i, n_cycles)
v.p.mod_i <- rep(p_mod_i, n_cycles)
v.p.sev_i <- rep(p_sev_i, n_cycles)

# Discontinuation vector per cycle
v.p_discontinuation <- numeric(n_cycles)
v.p_discontinuation[1] <- p_discontinuation1
if (n_cycles >= discontinuation2_begin) {
  v.p_discontinuation[2:(tx_duration - 1)] <- p_discontinuation2
  if (tx_duration <= n_cycles) {
    v.p_discontinuation[tx_duration:n_cycles] <- 1
  }
}

# Vector of age- and sex-specific mortality rates using the predefined mortality matrix
# Assuming 'life_table' data frame is already loaded from Prompt 2

# Create age vector from starting age across the number of cycles
v.age <- age_start + 0:(n_cycles - 1)

# Extract corresponding weighted mortality rates from the life table
v.mort_soc <- sapply(v.age, function(a) {
  rate <- life_table$weighted[life_table$age == a]
  if (length(rate) == 0) return(NA) else return(rate)
})


# Initialize vectors for intervention strategy (INT)
v.p.mci_mil_int <- numeric(n_cycles)
v.p.mci_mod_int <- numeric(n_cycles)
v.p.mci_sev_int <- numeric(n_cycles)
v.p.mil_mci_int <- rep(p_mil_mci, n_cycles)  # Backward transition - no treatment effect
v.p.mil_mod_int <- numeric(n_cycles)
v.p.mil_sev_int <- numeric(n_cycles)

v.p.mci_mil_int_dis <- numeric(n_cycles)
v.p.mci_mod_int_dis <- numeric(n_cycles)
v.p.mci_sev_int_dis <- numeric(n_cycles)
v.p.mil_mci_int_dis <- rep(p_mil_mci, n_cycles)  # Backward transition - no treatment effect
v.p.mil_mod_int_dis <- numeric(n_cycles)
v.p.mil_sev_int_dis <- numeric(n_cycles)

# Fill in on-treatment transition vectors
for (i in 1:n_cycles) {
  # Waning factor
  waning_factor <- (1 - tx_waning)^(i - 1)
  
  # MCI on-treatment
  rate_mci_mil <- -log(1 - p_mci_mil)
  rate_mci_mil_tx <- rate_mci_mil * rr_tx_mci_mil^waning_factor
  v.p.mci_mil_int[i] <- 1 - exp(-rate_mci_mil_tx)
  
  rate_mci_mod <- -log(1 - p_mci_mod)
  rate_mci_mod_tx <- rate_mci_mod * rr_tx_mci_mod^waning_factor
  v.p.mci_mod_int[i] <- 1 - exp(-rate_mci_mod_tx)
  
  rate_mci_sev <- -log(1 - p_mci_sev)
  rate_mci_sev_tx <- rate_mci_sev * rr_tx_mci_sev^waning_factor
  v.p.mci_sev_int[i] <- 1 - exp(-rate_mci_sev_tx)
  
  # Mild on-treatment
  rate_mil_mod <- -log(1 - p_mil_mod)
  rate_mil_mod_tx <- rate_mil_mod * rr_tx_mil_mod^waning_factor
  v.p.mil_mod_int[i] <- 1 - exp(-rate_mil_mod_tx)
  
  rate_mil_sev <- -log(1 - p_mil_sev)
  rate_mil_sev_tx <- rate_mil_sev * rr_tx_mil_sev^waning_factor
  v.p.mil_sev_int[i] <- 1 - exp(-rate_mil_sev_tx)
}

# Fill in off-treatment (post-discontinuation) vectors
for (i in 1:n_cycles) {
  # Waning factor after discontinuation
  waning_factor_dis <- (1 - tx_waning_dis)^(i - 1)
  
  # MCI off-treatment
  rate_mci_mil <- -log(1 - p_mci_mil)
  rate_mci_mil_tx_dis <- rate_mci_mil * rr_tx_mci_mil_dis^waning_factor_dis
  v.p.mci_mil_int_dis[i] <- 1 - exp(-rate_mci_mil_tx_dis)
  
  rate_mci_mod <- -log(1 - p_mci_mod)
  rate_mci_mod_tx_dis <- rate_mci_mod * rr_tx_mci_mod_dis^waning_factor_dis
  v.p.mci_mod_int_dis[i] <- 1 - exp(-rate_mci_mod_tx_dis)
  
  rate_mci_sev <- -log(1 - p_mci_sev)
  rate_mci_sev_tx_dis <- rate_mci_sev * rr_tx_mci_sev_dis^waning_factor_dis
  v.p.mci_sev_int_dis[i] <- 1 - exp(-rate_mci_sev_tx_dis)
  
  # Mild off-treatment
  rate_mil_mod <- -log(1 - p_mil_mod)
  rate_mil_mod_tx_dis <- rate_mil_mod * rr_tx_mil_mod_dis^waning_factor_dis
  v.p.mil_mod_int_dis[i] <- 1 - exp(-rate_mil_mod_tx_dis)
  
  rate_mil_sev <- -log(1 - p_mil_sev)
  rate_mil_sev_tx_dis <- rate_mil_sev * rr_tx_mil_sev_dis^waning_factor_dis
  v.p.mil_sev_int_dis[i] <- 1 - exp(-rate_mil_sev_tx_dis)
}

# Staying probabilities (on-treatment)
v.p.mci_stay_int <- 1 - (v.p.mci_mil_int + v.p.mci_mod_int + v.p.mci_sev_int)
v.p.mil_stay_int <- 1 - (v.p.mil_mci_int + v.p.mil_mod_int + v.p.mil_sev_int)

# Staying probabilities (off-treatment)
v.p.mci_stay_int_dis <- 1 - (v.p.mci_mil_int_dis + v.p.mci_mod_int_dis + v.p.mci_sev_int_dis)
v.p.mil_stay_int_dis <- 1 - (v.p.mil_mci_int_dis + v.p.mil_mod_int_dis + v.p.mil_sev_int_dis)


# Function to build the transition probability matrix array for a given strategy
build_transition_matrix <- function(strat) {
  
  # Initialize 3D transition probability array
  a.TP <- array(0, dim = c(length(state_names), length(state_names), n_cycles),
                dimnames = list(state_names, state_names, NULL))
  
  # Loop over each cycle
  for (t in 1:n_cycles) {
    
    # Compute mortality rates for each state based on general mortality and hazard ratios
    mort_mci <- 1 - exp(-v.mort_soc[t] * hr_mort_mci)
    mort_mil <- 1 - exp(-v.mort_soc[t] * hr_mort_mil)
    mort_mod <- 1 - exp(-v.mort_soc[t] * hr_mort_mod)
    mort_sev <- 1 - exp(-v.mort_soc[t] * hr_mort_sev)
    
    # Assign mortality to death from each state (one line per state)
    a.TP["mcion_c", "dth", t]   <- mort_mci
    a.TP["mciof_c", "dth", t]   <- mort_mci
    a.TP["milon_c", "dth", t]   <- mort_mil
    a.TP["milof_c", "dth", t]   <- mort_mil
    a.TP["mod_c", "dth", t]     <- mort_mod
    a.TP["sev_c", "dth", t]     <- mort_sev
    a.TP["mci_i", "dth", t]     <- mort_mci
    a.TP["mil_i", "dth", t]     <- mort_mil
    a.TP["mod_i", "dth", t]     <- mort_mod
    a.TP["sev_i", "dth", t]     <- mort_sev
    a.TP["dth", "dth", t]       <- 1  # Absorbing state
    
    # Transitions from "mcion_c" (MCI on-treatment, community)
    if (strat == "soc") {
      p_mci_mil <- v.p.mci_mil_soc[t]
      p_mci_mod <- v.p.mci_mod_soc[t]
      p_mci_sev <- v.p.mci_sev_soc[t]
      p_mci_stay <- v.p.mci_stay_soc[t]
    } else {
      p_mci_mil <- v.p.mci_mil_int[t]
      p_mci_mod <- v.p.mci_mod_int[t]
      p_mci_sev <- v.p.mci_sev_int[t]
      p_mci_stay <- v.p.mci_stay_int[t]
    }
    
    # Transition probabilities (base terms)
    surv <- 1 - a.TP["mcion_c", "dth", t]
    disc <- 1 - v.p_discontinuation[t]
    inst <- v.p.mci_i[t]
    comm <- 1 - inst
    
    # To "mcion_c" (stay in MCI on-treatment, community)
    a.TP["mcion_c", "mcion_c", t] <- p_mci_stay * surv * comm * disc
    
    # To "mciof_c" (MCI off-treatment, community)
    a.TP["mcion_c", "mciof_c", t] <- p_mci_stay * surv * comm * (1 - disc)
    
    # To "mci_i" (MCI institutionalized)
    a.TP["mcion_c", "mci_i", t] <- p_mci_stay * surv * inst
    
    # To "milon_c" (mild dementia on-treatment, community)
    a.TP["mcion_c", "milon_c", t] <- p_mci_mil * surv * comm * disc
    
    # To "milof_c" (mild dementia off-treatment, community)
    a.TP["mcion_c", "milof_c", t] <- p_mci_mil * surv * comm * (1 - disc)
    
    # To "mil_i" (mild dementia institutionalized)
    a.TP["mcion_c", "mil_i", t] <- p_mci_mil * surv * inst
    
    # To "mod_c" (moderate dementia community)
    a.TP["mcion_c", "mod_c", t] <- p_mci_mod * surv
    
    # To "sev_c" (severe dementia community)
    a.TP["mcion_c", "sev_c", t] <- p_mci_sev * surv
    
    # Transitions from "milon_c" (mild on-treatment, community)
    if (strat == "soc") {
      p_mil_mci <- v.p.mil_mci_soc[t]
      p_mil_mod <- v.p.mil_mod_soc[t]
      p_mil_sev <- v.p.mil_sev_soc[t]
      p_mil_stay <- v.p.mil_stay_soc[t]
    } else {
      p_mil_mci <- v.p.mil_mci_int[t]
      p_mil_mod <- v.p.mil_mod_int[t]
      p_mil_sev <- v.p.mil_sev_int[t]
      p_mil_stay <- v.p.mil_stay_int[t]
    }
    
    surv <- 1 - a.TP["milon_c", "dth", t]
    disc <- 1 - v.p_discontinuation[t]
    inst <- v.p.mil_i[t]
    comm <- 1 - inst
    
    # To "milon_c" (stay)
    a.TP["milon_c", "milon_c", t] <- p_mil_stay * surv * comm * disc
    
    # To "milof_c" (off-treatment)
    a.TP["milon_c", "milof_c", t] <- p_mil_stay * surv * comm * (1 - disc)
    
    # To "mil_i"
    a.TP["milon_c", "mil_i", t] <- p_mil_stay * surv * inst
    
    # To "mciof_c" (backward, off-treatment only)
    a.TP["milon_c", "mciof_c", t] <- p_mil_mci * surv * comm
    
    # To "mci_i"
    a.TP["milon_c", "mci_i", t] <- p_mil_mci * surv * inst
    
    # To "mod_c"
    a.TP["milon_c", "mod_c", t] <- p_mil_mod * surv
    
    # To "sev_c"
    a.TP["milon_c", "sev_c", t] <- p_mil_sev * surv
    
    
    # Transitions from "mciof_c" (MCI off-treatment, community)
    if (strat == "soc") {
      p_mci_mil <- v.p.mci_mil_soc[t]
      p_mci_mod <- v.p.mci_mod_soc[t]
      p_mci_sev <- v.p.mci_sev_soc[t]
      p_mci_stay <- v.p.mci_stay_soc[t]
    } else {
      p_mci_mil <- v.p.mci_mil_int_dis[t]
      p_mci_mod <- v.p.mci_mod_int_dis[t]
      p_mci_sev <- v.p.mci_sev_int_dis[t]
      p_mci_stay <- v.p.mci_stay_int_dis[t]
    }
    
    surv <- 1 - a.TP["mciof_c", "dth", t]
    inst <- v.p.mci_i[t]
    comm <- 1 - inst
    
    a.TP["mciof_c", "mciof_c", t] <- p_mci_stay * surv * comm
    a.TP["mciof_c", "mci_i", t]   <- p_mci_stay * surv * inst
    a.TP["mciof_c", "milof_c", t] <- p_mci_mil * surv * comm
    a.TP["mciof_c", "mil_i", t]   <- p_mci_mil * surv * inst
    a.TP["mciof_c", "mod_c", t]   <- p_mci_mod * surv
    a.TP["mciof_c", "sev_c", t]   <- p_mci_sev * surv
    
    # Transitions from "milof_c" (mild off-treatment, community)
    if (strat == "soc") {
      p_mil_mci <- v.p.mil_mci_soc[t]
      p_mil_mod <- v.p.mil_mod_soc[t]
      p_mil_sev <- v.p.mil_sev_soc[t]
      p_mil_stay <- v.p.mil_stay_soc[t]
    } else {
      p_mil_mci <- v.p.mil_mci_int_dis[t]
      p_mil_mod <- v.p.mil_mod_int_dis[t]
      p_mil_sev <- v.p.mil_sev_int_dis[t]
      p_mil_stay <- v.p.mil_stay_int_dis[t]
    }
    
    surv <- 1 - a.TP["milof_c", "dth", t]
    inst <- v.p.mil_i[t]
    comm <- 1 - inst
    
    a.TP["milof_c", "milof_c", t] <- p_mil_stay * surv * comm
    a.TP["milof_c", "mil_i", t]   <- p_mil_stay * surv * inst
    a.TP["milof_c", "mciof_c", t] <- p_mil_mci * surv * comm
    a.TP["milof_c", "mci_i", t]   <- p_mil_mci * surv * inst
    a.TP["milof_c", "mod_c", t]   <- p_mil_mod * surv
    a.TP["milof_c", "sev_c", t]   <- p_mil_sev * surv
    
    # Transitions from "mod_c" (moderate, community)
    if (strat == "soc") {
      p_mod_mil <- v.p.mod_mil_soc[t]
      p_mod_sev <- v.p.mod_sev_soc[t]
      p_mod_stay <- v.p.mod_stay_soc[t]
    } else {
      p_mod_mil <- v.p.mod_mil_soc[t]
      p_mod_sev <- v.p.mod_sev_soc[t]
      p_mod_stay <- v.p.mod_stay_soc[t]
    }
    
    surv <- 1 - a.TP["mod_c", "dth", t]
    inst <- v.p.mod_i[t]
    comm <- 1 - inst
    
    a.TP["mod_c", "mod_c", t] <- p_mod_stay * surv * comm
    a.TP["mod_c", "mod_i", t] <- p_mod_stay * surv * inst
    a.TP["mod_c", "milof_c", t] <- p_mod_mil * surv * comm
    a.TP["mod_c", "mil_i", t]   <- p_mod_mil * surv * inst
    a.TP["mod_c", "sev_c", t]   <- p_mod_sev * surv
    
    # Transitions from "sev_c" (severe, community)
    if (strat == "soc") {
      p_sev_mil <- v.p.sev_mil_soc[t]
      p_sev_mod <- v.p.sev_mod_soc[t]
      p_sev_stay <- v.p.sev_stay_soc[t]
    } else {
      p_sev_mil <- v.p.sev_mil_soc[t]
      p_sev_mod <- v.p.sev_mod_soc[t]
      p_sev_stay <- v.p.sev_stay_soc[t]
    }
    
    surv <- 1 - a.TP["sev_c", "dth", t]
    inst <- v.p.sev_i[t]
    comm <- 1 - inst
    
    a.TP["sev_c", "sev_c", t] <- p_sev_stay * surv * comm
    a.TP["sev_c", "sev_i", t] <- p_sev_stay * surv * inst
    a.TP["sev_c", "milof_c", t] <- p_sev_mil * surv * comm
    a.TP["sev_c", "mil_i", t]   <- p_sev_mil * surv * inst
    a.TP["sev_c", "mod_c", t]   <- p_sev_mod * surv
    
    
    # Transitions from "mci_i" (institutionalized MCI)
    if (strat == "soc") {
      p_mci_mil <- v.p.mci_mil_soc[t]
      p_mci_mod <- v.p.mci_mod_soc[t]
      p_mci_sev <- v.p.mci_sev_soc[t]
      p_mci_stay <- v.p.mci_stay_soc[t]
    } else {
      p_mci_mil <- v.p.mci_mil_int_dis[t]
      p_mci_mod <- v.p.mci_mod_int_dis[t]
      p_mci_sev <- v.p.mci_sev_int_dis[t]
      p_mci_stay <- v.p.mci_stay_int_dis[t]
    }
    
    surv <- 1 - a.TP["mci_i", "dth", t]
    a.TP["mci_i", "mci_i", t] <- p_mci_stay * surv
    a.TP["mci_i", "mil_i", t] <- p_mci_mil * surv
    a.TP["mci_i", "mod_i", t] <- p_mci_mod * surv
    a.TP["mci_i", "sev_i", t] <- p_mci_sev * surv
    
    # Transitions from "mil_i"
    if (strat == "soc") {
      p_mil_mci <- v.p.mil_mci_soc[t]
      p_mil_mod <- v.p.mil_mod_soc[t]
      p_mil_sev <- v.p.mil_sev_soc[t]
      p_mil_stay <- v.p.mil_stay_soc[t]
    } else {
      p_mil_mci <- v.p.mil_mci_int_dis[t]
      p_mil_mod <- v.p.mil_mod_int_dis[t]
      p_mil_sev <- v.p.mil_sev_int_dis[t]
      p_mil_stay <- v.p.mil_stay_int_dis[t]
    }
    
    surv <- 1 - a.TP["mil_i", "dth", t]
    a.TP["mil_i", "mil_i", t] <- p_mil_stay * surv
    a.TP["mil_i", "mci_i", t] <- p_mil_mci * surv
    a.TP["mil_i", "mod_i", t] <- p_mil_mod * surv
    a.TP["mil_i", "sev_i", t] <- p_mil_sev * surv
    
    # Transitions from "mod_i"
    p_mod_mil <- v.p.mod_mil_soc[t]
    p_mod_sev <- v.p.mod_sev_soc[t]
    p_mod_stay <- v.p.mod_stay_soc[t]
    
    surv <- 1 - a.TP["mod_i", "dth", t]
    a.TP["mod_i", "mod_i", t] <- p_mod_stay * surv
    a.TP["mod_i", "mil_i", t] <- p_mod_mil * surv
    a.TP["mod_i", "sev_i", t] <- p_mod_sev * surv
    
    # Transitions from "sev_i"
    p_sev_mil <- v.p.sev_mil_soc[t]
    p_sev_mod <- v.p.sev_mod_soc[t]
    p_sev_stay <- v.p.sev_stay_soc[t]
    
    surv <- 1 - a.TP["sev_i", "dth", t]
    a.TP["sev_i", "sev_i", t] <- p_sev_stay * surv
    a.TP["sev_i", "mil_i", t] <- p_sev_mil * surv
    a.TP["sev_i", "mod_i", t] <- p_sev_mod * surv
    
    
    
    # Normalize rows to sum to 1 (excluding "dth" state, already normalized)
    for (s in setdiff(state_names, "dth")) {
      row_sum <- sum(a.TP[s, , t])
      if (row_sum > 0 && abs(row_sum - 1) > 1e-8) {
        a.TP[s, , t] <- a.TP[s, , t] / row_sum
      }
    }
  }
  
  return(a.TP)
}


# Function to compute Markov trace and total outcomes for a given strategy
compute_outcomes <- function(strat) {
  
  # Initialize Markov trace matrix
  m.M <- matrix(0, nrow = n_cycles, ncol = length(state_names),
                dimnames = list(1:n_cycles, state_names))
  
  # Initial state distribution
  m.M[1, ] <- c(
    mcion_c = ifelse(strat == "int", p_starting_state_mci, 0),
    mciof_c = ifelse(strat == "soc", p_starting_state_mci, 0),
    milon_c = ifelse(strat == "int", p_starting_state_mil, 0),
    milof_c = ifelse(strat == "soc", p_starting_state_mil, 0),
    mod_c = 0, sev_c = 0, mci_i = 0, mil_i = 0, mod_i = 0, sev_i = 0, dth = 0
  )
  
  # Transition matrix
  a.P <- build_transition_matrix(strat)
  
  # Initialize outcome matrices
  v.QALYs <- numeric(n_cycles)
  v.Costs <- numeric(n_cycles)
  v.LYs <- numeric(n_cycles)
  
  # Define utility and cost vectors (named)
  v.u <- c(
    mcion_c = u_mci_pt + u_mci_ic, mciof_c = u_mci_pt + u_mci_ic,
    milon_c = u_mil_pt + u_mil_ic, milof_c = u_mil_pt + u_mil_ic,
    mod_c = u_mod_pt + u_mod_ic, sev_c = u_sev_pt + u_sev_ic,
    mci_i = u_mci_pt_i + u_mci_ic_i, mil_i = u_mil_pt_i + u_mil_ic_i,
    mod_i = u_mod_pt_i + u_mod_ic_i, sev_i = u_sev_pt_i + u_sev_ic_i,
    dth = 0
  )
  
  v.c <- c(
    mcion_c = c_mci_hc + c_mci_sc + c_mci_ic,
    mciof_c = c_mci_hc + c_mci_sc + c_mci_ic,
    milon_c = c_mil_hc + c_mil_sc + c_mil_ic,
    milof_c = c_mil_hc + c_mil_sc + c_mil_ic,
    mod_c = c_mod_hc + c_mod_sc + c_mod_ic,
    sev_c = c_sev_hc + c_sev_sc + c_sev_ic,
    mci_i = c_mci_i_hc + c_mci_i_sc + c_mci_i_ic,
    mil_i = c_mil_i_hc + c_mil_i_sc + c_mil_i_ic,
    mod_i = c_mod_i_hc + c_mod_i_sc + c_mod_i_ic,
    sev_i = c_sev_i_hc + c_sev_i_sc + c_sev_i_ic,
    dth = 0
  )
  
  # Markov trace iteration
  for (t in 2:n_cycles) {
    m.M[t, ] <- m.M[t - 1, ] %*% a.P[, , t - 1]
  }
  
  # Compute undiscounted QALYs and costs per cycle
  for (t in 1:n_cycles) {
    v.QALYs[t] <- sum(m.M[t, ] * v.u)
    v.Costs[t] <- sum(m.M[t, ] * v.c)
    v.LYs[t] <- sum(m.M[t, state_names[state_names != "dth"]])
  }
  
  # Apply treatment cost and benefit for "int" strategy
  if (strat == "int") {
    treat_states <- c("mcion_c", "milon_c")
    # Recurring treatment cost
    v.Costs <- v.Costs + rowSums(m.M[, treat_states, drop = FALSE]) * c_Tx
    # Initial treatment QALY gain and cost (one-time)
    v.QALYs[1] <- v.QALYs[1] + sum(m.M[1, treat_states]) * u_Tx_start
    v.Costs[1] <- v.Costs[1] + c_Tx_start
  }
  
  # Apply half-cycle correction
  v.QALYs <- 0.5 * (v.QALYs[-n_cycles] + v.QALYs[-1])
  v.Costs <- 0.5 * (v.Costs[-n_cycles] + v.Costs[-1])
  v.LYs <- 0.5 * (v.LYs[-n_cycles] + v.LYs[-1])
  
  # Apply discounting
  discount_vector <- 1 / ((1 + discount_QALY) ^ (1:(n_cycles - 1)))
  QALYs_total <- sum(v.QALYs * discount_vector)
  Costs_total <- sum(v.Costs * discount_vector)
  LYs_total <- sum(v.LYs * discount_vector)
  
  return(list(QALYs = QALYs_total, Costs = Costs_total, LYs = LYs_total))
}

# Compute and compare outcomes
res_soc <- compute_outcomes("soc")
res_int <- compute_outcomes("int")

# Print results
cat("=== Total Outcomes ===\n")
cat("Strategy: Standard of Care (soc)\n")
cat(sprintf("QALYs: %.2f\nCosts: $%.2f\nLife Years: %.2f\n\n", res_soc$QALYs, res_soc$Costs, res_soc$LYs))
cat("Strategy: Intervention (int)\n")
cat(sprintf("QALYs: %.2f\nCosts: $%.2f\nLife Years: %.2f\n\n", res_int$QALYs, res_int$Costs, res_int$LYs))

cat("=== Incremental Results (int - soc) ===\n")
cat(sprintf("Incremental QALYs: %.2f\n", res_int$QALYs - res_soc$QALYs))
cat(sprintf("Incremental Costs: $%.2f\n", res_int$Costs - res_soc$Costs))
cat(sprintf("Incremental Life Years: %.2f\n", res_int$LYs - res_soc$LYs))
```
