## Supplementary material for "Replicating Health-Economic Simulation Models for Alzheimer’s Disease Using Artificial Intelligence": final R scripts: o3.R

```
#=======================
# INPUT PARAMETERS FOR IPECAD MARKOV MODEL
#=======================

# Fixed state names
state_names <- c("mcion_c", "mciof_c", "milon_c", "milof_c", "mod_c", "sev_c",
                 "mci_i", "mil_i", "mod_i", "sev_i", "dth")

# Strategy names
strategy_names <- c("soc", "int")

# Demographics
age_start <- 71
sex <- "weighted"  # 52% female

# Starting population distribution
p_starting_state_mci <- 0.55  # all others start in mild dementia, community setting

# Model time horizon
n_cycle <- 29  # 1-year cycles

# Transition probabilities (community setting, natural history)
p_mci_mil <- 0.23
p_mci_mod <- 0
p_mci_sev <- 0
p_mil_mci <- 0.03
p_mil_mod <- 0.35
p_mil_sev <- 0.04
p_mod_mil <- 0.03
p_mod_sev <- 0.42
p_sev_mil <- 0
p_sev_mod <- 0.02

# Transition probabilities to institutional care
p_mci_i <- 0.024
p_mil_i <- 0.038
p_mod_i <- 0.110
p_sev_i <- 0.259

# Mortality hazard ratios (HRs relative to general population)
hr_mort_mci <- 1.82
hr_mort_mil <- 2.92
hr_mort_mod <- 3.85
hr_mort_sev <- 9.52

# Treatment effects (on-treatment, relative risk for progression)
rr_tx_mci_mil <- 0.69
rr_tx_mci_mod <- 1
rr_tx_mci_sev <- 1
rr_tx_mil_mod <- 0.69
rr_tx_mil_sev <- 0.69

# Treatment effects after discontinuation
rr_tx_mci_mil_dis <- 1
rr_tx_mci_mod_dis <- 1
rr_tx_mci_sev_dis <- 1
rr_tx_mil_mod_dis <- 1
rr_tx_mil_sev_dis <- 1

# Treatment discontinuation
p_discontinuation1 <- 0.069
p_discontinuation2 <- 0
discontinuation2_begin <- 2
tx_duration <- 29  # max duration
tx_waning <- 0
tx_waning_dis <- 0

# Utilities – patient (community)
u_mci_pt <- 0.851 - 0.17
u_mil_pt <- 0.851 - 0.22
u_mod_pt <- 0.851 - 0.36
u_sev_pt <- 0.851 - 0.53

# Utilities – patient (institution)
u_mci_pt_i <- 0.851 - 0.17
u_mil_pt_i <- 0.851 - 0.19
u_mod_pt_i <- 0.851 - 0.42
u_sev_pt_i <- 0.851 - 0.59

# Utilities – informal caregiver (community)
u_mci_ic <- -0.03
u_mil_ic <- -0.05
u_mod_ic <- -0.08
u_sev_ic <- -0.10

# Utilities – informal caregiver (institution)
u_mci_ic_i <- -0.03
u_mil_ic_i <- -0.05
u_mod_ic_i <- -0.08
u_sev_ic_i <- -0.10

# Disutility from treatment start (e.g., ARIA)
u_Tx_start <- -0.14 * (12 / 52) * 0.035

# Costs – healthcare (community)
c_mci_hc <- 6042 * 1.12 + 460
c_mil_hc <- 6042 * 1.56 + 965 + 0.21 * 365 * 0.333
c_mod_hc <- 6042 * 1.93 + 1544 + 0.66 * 365 * 0.333
c_sev_hc <- 6042 * 1.93 + 1930

# Costs – healthcare (institution)
c_mci_i_hc <- c_mci_hc
c_mil_i_hc <- c_mil_hc
c_mod_i_hc <- c_mod_hc
c_sev_i_hc <- c_sev_hc

# Costs – social care (community)
c_mci_sc <- 0
c_mil_sc <- 0
c_mod_sc <- 0
c_sev_sc <- 0

# Costs – social care (institution)
c_mci_i_sc <- 7394 * 12
c_mil_i_sc <- 7394 * 12
c_mod_i_sc <- 7394 * 12
c_sev_i_sc <- 7394 * 12

# Costs – informal care (community)
c_mci_ic <- 69 * 12 * 32.46 + 0.204 * 0.049 * 20 * 52 * 32.46
c_mil_ic <- 113 * 12 * 32.46 + 0.112 * 0.086 * 20 * 52 * 32.46
c_mod_ic <- 169 * 12 * 32.46
c_sev_ic <- 298 * 12 * 32.46

# Costs – informal care (institution)
c_mci_i_ic <- 69 * 12 * 32.46 * 0.44 + 0.204 * 0.049 * 20 * 52 * 32.46
c_mil_i_ic <- 113 * 12 * 32.46 * 0.44 + 0.112 * 0.086 * 20 * 52 * 32.46
c_mod_i_ic <- 169 * 12 * 32.46 * 0.44
c_sev_i_ic <- 298 * 12 * 32.46 * 0.44

# Costs – treatment and first-cycle add-ons
c_Tx <- 26500 + (52 / 2) * 78.35
c_Tx_start <- 261.10 * 4 + 261.10 * 3 * 0.215

# Economic evaluation settings
discount_EFFECT <- 0.03
discount_QALY <- 0.03
discount_COST <- 0.03
wtp <- 100000
half_cycle_correction <- TRUE
#=======================
# LOAD AND PROCESS US LIFE TABLE DATA (2019)
#=======================

# Load life table CSV
lifetable <- read.csv("lifetable_US_2019_ssa.csv", stringsAsFactors = FALSE)

# Convert mortality probabilities (q) to mortality rates (r) using: r = -log(1 - q)
lifetable$male <- -log(1 - lifetable$male)
lifetable$female <- -log(1 - lifetable$female)

# Compute weighted average mortality rate (52% female, 48% male)
lifetable$weighted <- 0.48 * lifetable$male + 0.52 * lifetable$female

# View resulting data frame
lifetable <- lifetable[, c("age", "male", "female", "weighted")]

# Output for checking (optional)
head(lifetable)


#=======================
# TIME-DEPENDENT TRANSITION PROBABILITIES - "SOC" STRATEGY
#=======================

# Initialize cycle vector
cycles <- 0:(n_cycle - 1)

# Disease progression transition probabilities (time-invariant, repeated per cycle)
v.p.mci_mil_soc <- rep(p_mci_mil, n_cycle)
v.p.mci_mod_soc <- rep(p_mci_mod, n_cycle)
v.p.mci_sev_soc <- rep(p_mci_sev, n_cycle)

v.p.mil_mci_soc <- rep(p_mil_mci, n_cycle)
v.p.mil_mod_soc <- rep(p_mil_mod, n_cycle)
v.p.mil_sev_soc <- rep(p_mil_sev, n_cycle)

v.p.mod_mil_soc <- rep(p_mod_mil, n_cycle)
v.p.mod_sev_soc <- rep(p_mod_sev, n_cycle)

v.p.sev_mil_soc <- rep(p_sev_mil, n_cycle)
v.p.sev_mod_soc <- rep(p_sev_mod, n_cycle)

# Stay probabilities (excluding mortality and institutionalization)
v.p.mci_stay_soc <- 1 - (v.p.mci_mil_soc + v.p.mci_mod_soc + v.p.mci_sev_soc)
v.p.mil_stay_soc <- 1 - (v.p.mil_mci_soc + v.p.mil_mod_soc + v.p.mil_sev_soc)
v.p.mod_stay_soc <- 1 - (v.p.mod_mil_soc + v.p.mod_sev_soc)
v.p.sev_stay_soc <- 1 - (v.p.sev_mil_soc + v.p.sev_mod_soc)

# Institutionalization transitions (time-invariant)
v.p.mci_to_inst <- rep(p_mci_i, n_cycle)
v.p.mil_to_inst <- rep(p_mil_i, n_cycle)
v.p.mod_to_inst <- rep(p_mod_i, n_cycle)
v.p.sev_to_inst <- rep(p_sev_i, n_cycle)

# Discontinuation vector
v.p.discontinuation <- rep(0, n_cycle)
v.p.discontinuation[1] <- p_discontinuation1
v.p.discontinuation[2:(tx_duration)] <- p_discontinuation2
if (tx_duration < n_cycle) {
  v.p.discontinuation[(tx_duration + 1):n_cycle] <- 1
}

# Mortality vector (age- and sex-specific, weighted)
# Use preprocessed life table: "lifetable" with columns "age" and "weighted"
ages <- age_start + cycles
v.mort_rate <- lifetable$weighted[match(ages, lifetable$age)]


#=======================
# TIME-DEPENDENT TRANSITION PROBABILITIES - "INT" STRATEGY
#=======================

# Initialize vectors
v.p.mcion_mil_int <- numeric(n_cycle)
v.p.mcion_mod_int <- numeric(n_cycle)
v.p.mcion_sev_int <- numeric(n_cycle)
v.p.mciof_mil_int <- numeric(n_cycle)
v.p.mciof_mod_int <- numeric(n_cycle)
v.p.mciof_sev_int <- numeric(n_cycle)

v.p.milon_mci_int <- numeric(n_cycle)
v.p.milon_mod_int <- numeric(n_cycle)
v.p.milon_sev_int <- numeric(n_cycle)
v.p.milof_mci_int <- numeric(n_cycle)
v.p.milof_mod_int <- numeric(n_cycle)
v.p.milof_sev_int <- numeric(n_cycle)

# Loop through each cycle to compute time-dependent values
for (t in 1:n_cycle) {
  waning_factor <- (1 - tx_waning)^(t - 1)
  waning_dis_factor <- (1 - tx_waning_dis)^(t - 1)
  
  # On-treatment MCI transitions (community)
  r <- -log(1 - p_mci_mil)
  v.p.mcion_mil_int[t] <- 1 - exp(-r * rr_tx_mci_mil^waning_factor)
  
  r <- -log(1 - p_mci_mod)
  v.p.mcion_mod_int[t] <- 1 - exp(-r * rr_tx_mci_mod^waning_factor)
  
  r <- -log(1 - p_mci_sev)
  v.p.mcion_sev_int[t] <- 1 - exp(-r * rr_tx_mci_sev^waning_factor)
  
  # On-treatment Mild transitions (community)
  v.p.milon_mci_int[t] <- p_mil_mci  # backward transition (no treatment effect)
  
  r <- -log(1 - p_mil_mod)
  v.p.milon_mod_int[t] <- 1 - exp(-r * rr_tx_mil_mod^waning_factor)
  
  r <- -log(1 - p_mil_sev)
  v.p.milon_sev_int[t] <- 1 - exp(-r * rr_tx_mil_sev^waning_factor)
  
  # Post-discontinuation MCI transitions (community)
  r <- -log(1 - p_mci_mil)
  v.p.mciof_mil_int[t] <- 1 - exp(-r * rr_tx_mci_mil_dis^waning_dis_factor)
  
  r <- -log(1 - p_mci_mod)
  v.p.mciof_mod_int[t] <- 1 - exp(-r * rr_tx_mci_mod_dis^waning_dis_factor)
  
  r <- -log(1 - p_mci_sev)
  v.p.mciof_sev_int[t] <- 1 - exp(-r * rr_tx_mci_sev_dis^waning_dis_factor)
  
  # Post-discontinuation Mild transitions (community)
  v.p.milof_mci_int[t] <- p_mil_mci  # backward transition (no treatment effect)
  
  r <- -log(1 - p_mil_mod)
  v.p.milof_mod_int[t] <- 1 - exp(-r * rr_tx_mil_mod_dis^waning_dis_factor)
  
  r <- -log(1 - p_mil_sev)
  v.p.milof_sev_int[t] <- 1 - exp(-r * rr_tx_mil_sev_dis^waning_dis_factor)
}

# Stay probabilities for on-treatment
v.p.mcion_stay_int <- 1 - (v.p.mcion_mil_int + v.p.mcion_mod_int + v.p.mcion_sev_int)
v.p.milon_stay_int <- 1 - (v.p.milon_mci_int + v.p.milon_mod_int + v.p.milon_sev_int)

# Stay probabilities for off-treatment
v.p.mciof_stay_int <- 1 - (v.p.mciof_mil_int + v.p.mciof_mod_int + v.p.mciof_sev_int)
v.p.milof_stay_int <- 1 - (v.p.milof_mci_int + v.p.milof_mod_int + v.p.milof_sev_int)


#=======================
# FUNCTION TO BUILD TRANSITION MATRIX ARRAY a.TP
#=======================

build_transition_matrix <- function(strat) {
  
  # Define array dimensions
  n_states <- length(state_names)
  a.TP <- array(0, dim = c(n_states, n_states, n_cycle),
                dimnames = list(state_names, state_names, NULL))
  
  # Loop over each cycle
  for (t in 1:n_cycle) {
    
    # Get age-specific mortality rate for this cycle
    mort_rate <- v.mort_rate[t]
    
    # Compute transition probabilities to death from each state
    a.TP["mcion_c", "dth", t] <- 1 - exp(-mort_rate * hr_mort_mci)
    a.TP["mciof_c", "dth", t] <- 1 - exp(-mort_rate * hr_mort_mci)
    a.TP["milon_c", "dth", t] <- 1 - exp(-mort_rate * hr_mort_mil)
    a.TP["milof_c", "dth", t] <- 1 - exp(-mort_rate * hr_mort_mil)
    a.TP["mod_c",   "dth", t] <- 1 - exp(-mort_rate * hr_mort_mod)
    a.TP["sev_c",   "dth", t] <- 1 - exp(-mort_rate * hr_mort_sev)
    a.TP["mci_i",   "dth", t] <- 1 - exp(-mort_rate * hr_mort_mci)
    a.TP["mil_i",   "dth", t] <- 1 - exp(-mort_rate * hr_mort_mil)
    a.TP["mod_i",   "dth", t] <- 1 - exp(-mort_rate * hr_mort_mod)
    a.TP["sev_i",   "dth", t] <- 1 - exp(-mort_rate * hr_mort_sev)
    a.TP["dth",     "dth", t] <- 1  # absorbing state
    
    # Transitions from mcion_c (MCI on-treatment in community)
    if (strat == "soc") {
      p_mil <- v.p.mci_mil_soc[t]
      p_mod <- v.p.mci_mod_soc[t]
      p_sev <- v.p.mci_sev_soc[t]
      p_stay <- v.p.mci_stay_soc[t]
      p_inst <- v.p.mci_to_inst[t]
    } else {
      p_mil <- v.p.mcion_mil_int[t]
      p_mod <- v.p.mcion_mod_int[t]
      p_sev <- v.p.mcion_sev_int[t]
      p_stay <- v.p.mcion_stay_int[t]
      p_inst <- v.p.mci_to_inst[t]
    }
    
    p_mort <- a.TP["mcion_c", "dth", t]
    p_alive <- 1 - p_mort
    p_disc <- v.p.discontinuation[t]
    
    a.TP["mcion_c", "mcion_c", t] <- p_alive * p_stay * (1 - p_inst) * (1 - p_disc)
    a.TP["mcion_c", "mciof_c", t] <- p_alive * p_stay * (1 - p_inst) * p_disc
    a.TP["mcion_c", "mci_i",    t] <- p_alive * p_stay * p_inst
    
    a.TP["mcion_c", "milon_c",  t] <- p_alive * p_mil * (1 - p_inst) * (1 - p_disc)
    a.TP["mcion_c", "milof_c", t] <- p_alive * p_mil * (1 - p_inst) * p_disc
    a.TP["mcion_c", "mil_i",    t] <- p_alive * p_mil * p_inst
    
    a.TP["mcion_c", "mod_c",    t] <- p_alive * p_mod
    a.TP["mcion_c", "sev_c",    t] <- p_alive * p_sev
    
    # Transitions from milon_c (Mild on-treatment in community)
    if (strat == "soc") {
      p_mci <- v.p.mil_mci_soc[t]
      p_mod <- v.p.mil_mod_soc[t]
      p_sev <- v.p.mil_sev_soc[t]
      p_stay <- v.p.mil_stay_soc[t]
      p_inst <- v.p.mil_to_inst[t]
    } else {
      p_mci <- v.p.milon_mci_int[t]
      p_mod <- v.p.milon_mod_int[t]
      p_sev <- v.p.milon_sev_int[t]
      p_stay <- v.p.milon_stay_int[t]
      p_inst <- v.p.mil_to_inst[t]
    }
    
    p_mort <- a.TP["milon_c", "dth", t]
    p_alive <- 1 - p_mort
    p_disc <- v.p.discontinuation[t]
    
    a.TP["milon_c", "milon_c", t] <- p_alive * p_stay * (1 - p_inst) * (1 - p_disc)
    a.TP["milon_c", "milof_c", t] <- p_alive * p_stay * (1 - p_inst) * p_disc
    a.TP["milon_c", "mil_i",   t] <- p_alive * p_stay * p_inst
    
    a.TP["milon_c", "mcion_c", t] <- p_alive * p_mci * (1 - p_inst) * (1 - p_disc)
    a.TP["milon_c", "mciof_c", t] <- p_alive * p_mci * (1 - p_inst) * p_disc
    a.TP["milon_c", "mci_i",   t] <- p_alive * p_mci * p_inst
    
    a.TP["milon_c", "mod_c",   t] <- p_alive * p_mod
    a.TP["milon_c", "sev_c",   t] <- p_alive * p_sev
    
    # Transitions from mciof_c (MCI off-treatment in community)
    if (strat == "soc") {
      p_mil <- v.p.mci_mil_soc[t]
      p_mod <- v.p.mci_mod_soc[t]
      p_sev <- v.p.mci_sev_soc[t]
      p_stay <- v.p.mci_stay_soc[t]
      p_inst <- v.p.mci_to_inst[t]
    } else {
      p_mil <- v.p.mciof_mil_int[t]
      p_mod <- v.p.mciof_mod_int[t]
      p_sev <- v.p.mciof_sev_int[t]
      p_stay <- v.p.mciof_stay_int[t]
      p_inst <- v.p.mci_to_inst[t]
    }
    
    p_mort <- a.TP["mciof_c", "dth", t]
    p_alive <- 1 - p_mort
    
    a.TP["mciof_c", "mciof_c", t] <- p_alive * p_stay * (1 - p_inst)
    a.TP["mciof_c", "mci_i",    t] <- p_alive * p_stay * p_inst
    
    a.TP["mciof_c", "milof_c", t] <- p_alive * p_mil * (1 - p_inst)
    a.TP["mciof_c", "mil_i",   t] <- p_alive * p_mil * p_inst
    
    a.TP["mciof_c", "mod_c",   t] <- p_alive * p_mod
    a.TP["mciof_c", "sev_c",   t] <- p_alive * p_sev
    
    # Transitions from milof_c (Mild off-treatment in community)
    if (strat == "soc") {
      p_mci <- v.p.mil_mci_soc[t]
      p_mod <- v.p.mil_mod_soc[t]
      p_sev <- v.p.mil_sev_soc[t]
      p_stay <- v.p.mil_stay_soc[t]
      p_inst <- v.p.mil_to_inst[t]
    } else {
      p_mci <- v.p.milof_mci_int[t]
      p_mod <- v.p.milof_mod_int[t]
      p_sev <- v.p.milof_sev_int[t]
      p_stay <- v.p.milof_stay_int[t]
      p_inst <- v.p.mil_to_inst[t]
    }
    
    p_mort <- a.TP["milof_c", "dth", t]
    p_alive <- 1 - p_mort
    
    a.TP["milof_c", "milof_c", t] <- p_alive * p_stay * (1 - p_inst)
    a.TP["milof_c", "mil_i",   t] <- p_alive * p_stay * p_inst
    
    a.TP["milof_c", "mciof_c", t] <- p_alive * p_mci * (1 - p_inst)
    a.TP["milof_c", "mci_i",   t] <- p_alive * p_mci * p_inst
    
    a.TP["milof_c", "mod_c",   t] <- p_alive * p_mod
    a.TP["milof_c", "sev_c",   t] <- p_alive * p_sev
    
    # Transitions from mod_c (Moderate dementia in community)
    if (strat == "soc") {
      p_mil <- v.p.mod_mil_soc[t]
      p_sev <- v.p.mod_sev_soc[t]
      p_stay <- v.p.mod_stay_soc[t]
      p_inst <- v.p.mod_to_inst[t]
    } else {
      p_mil <- v.p.mod_mil_soc[t]  # No separate vectors for int
      p_sev <- v.p.mod_sev_soc[t]
      p_stay <- v.p.mod_stay_soc[t]
      p_inst <- v.p.mod_to_inst[t]
    }
    
    p_mort <- a.TP["mod_c", "dth", t]
    p_alive <- 1 - p_mort
    
    a.TP["mod_c", "mod_c", t] <- p_alive * p_stay * (1 - p_inst)
    a.TP["mod_c", "mod_i", t] <- p_alive * p_stay * p_inst
    
    a.TP["mod_c", "milof_c", t] <- p_alive * p_mil * (1 - p_inst)
    a.TP["mod_c", "mil_i",   t] <- p_alive * p_mil * p_inst
    
    a.TP["mod_c", "sev_c",   t] <- p_alive * p_sev
    
    # Transitions from sev_c (Severe dementia in community)
    if (strat == "soc") {
      p_mil <- v.p.sev_mil_soc[t]
      p_mod <- v.p.sev_mod_soc[t]
      p_stay <- v.p.sev_stay_soc[t]
      p_inst <- v.p.sev_to_inst[t]
    } else {
      p_mil <- v.p.sev_mil_soc[t]  # No separate vectors for int
      p_mod <- v.p.sev_mod_soc[t]
      p_stay <- v.p.sev_stay_soc[t]
      p_inst <- v.p.sev_to_inst[t]
    }
    
    p_mort <- a.TP["sev_c", "dth", t]
    p_alive <- 1 - p_mort
    
    a.TP["sev_c", "sev_c", t] <- p_alive * p_stay * (1 - p_inst)
    a.TP["sev_c", "sev_i", t] <- p_alive * p_stay * p_inst
    
    a.TP["sev_c", "milof_c", t] <- p_alive * p_mil * (1 - p_inst)
    a.TP["sev_c", "mil_i",   t] <- p_alive * p_mil * p_inst
    
    a.TP["sev_c", "mod_c",   t] <- p_alive * p_mod
    
    
    # Transitions from mci_i (MCI in institution)
    if (strat == "soc") {
      p_mil <- v.p.mci_mil_soc[t]
      p_mod <- v.p.mci_mod_soc[t]
      p_sev <- v.p.mci_sev_soc[t]
      p_stay <- v.p.mci_stay_soc[t]
    } else {
      p_mil <- v.p.mciof_mil_int[t]
      p_mod <- v.p.mciof_mod_int[t]
      p_sev <- v.p.mciof_sev_int[t]
      p_stay <- v.p.mciof_stay_int[t]
    }
    
    p_mort <- a.TP["mci_i", "dth", t]
    p_alive <- 1 - p_mort
    
    a.TP["mci_i", "mci_i", t] <- p_alive * p_stay
    a.TP["mci_i", "mil_i", t] <- p_alive * p_mil
    a.TP["mci_i", "mod_i", t] <- p_alive * p_mod
    a.TP["mci_i", "sev_i", t] <- p_alive * p_sev
    
    # Transitions from mil_i (Mild dementia in institution)
    if (strat == "soc") {
      p_mci <- v.p.mil_mci_soc[t]
      p_mod <- v.p.mil_mod_soc[t]
      p_sev <- v.p.mil_sev_soc[t]
      p_stay <- v.p.mil_stay_soc[t]
    } else {
      p_mci <- v.p.milof_mci_int[t]
      p_mod <- v.p.milof_mod_int[t]
      p_sev <- v.p.milof_sev_int[t]
      p_stay <- v.p.milof_stay_int[t]
    }
    
    p_mort <- a.TP["mil_i", "dth", t]
    p_alive <- 1 - p_mort
    
    a.TP["mil_i", "mil_i", t] <- p_alive * p_stay
    a.TP["mil_i", "mci_i", t] <- p_alive * p_mci
    a.TP["mil_i", "mod_i", t] <- p_alive * p_mod
    a.TP["mil_i", "sev_i", t] <- p_alive * p_sev
    
    # Transitions from mod_i (Moderate dementia in institution)
    p_mil <- v.p.mod_mil_soc[t]
    p_sev <- v.p.mod_sev_soc[t]
    p_stay <- v.p.mod_stay_soc[t]
    
    p_mort <- a.TP["mod_i", "dth", t]
    p_alive <- 1 - p_mort
    
    a.TP["mod_i", "mod_i", t] <- p_alive * p_stay
    a.TP["mod_i", "mil_i", t] <- p_alive * p_mil
    a.TP["mod_i", "sev_i", t] <- p_alive * p_sev
    
    # Transitions from sev_i (Severe dementia in institution)
    p_mil <- v.p.sev_mil_soc[t]
    p_mod <- v.p.sev_mod_soc[t]
    p_stay <- v.p.sev_stay_soc[t]
    
    p_mort <- a.TP["sev_i", "dth", t]
    p_alive <- 1 - p_mort
    
    a.TP["sev_i", "sev_i", t] <- p_alive * p_stay
    a.TP["sev_i", "mil_i", t] <- p_alive * p_mil
    a.TP["sev_i", "mod_i", t] <- p_alive * p_mod
    
    # Normalize rows to sum to 1 (if not absorbing state)
    for (s in state_names[state_names != "dth"]) {
      row_sum <- sum(a.TP[s, , t])
      if (row_sum > 0 && abs(row_sum - 1) > 1e-8) {
        a.TP[s, , t] <- a.TP[s, , t] / row_sum
      }
    }
  }
  
  return(a.TP)
}
#=======================
# FUNCTION TO COMPUTE MARKOV TRACE AND OUTCOMES
#=======================

compute_outcomes <- function(strat) {
  
  # Transition matrix
  a.TP <- build_transition_matrix(strat)
  
  # Initialize Markov trace matrix
  m.trace <- matrix(0, nrow = n_cycle, ncol = length(state_names), dimnames = list(NULL, state_names))
  
  # Initial state distribution
  v.init <- rep(0, length(state_names))
  names(v.init) <- state_names
  v.init["mciof_c"] <- 1 - p_starting_state_mci
  v.init["mcion_c"] <- p_starting_state_mci
  m.trace[1, ] <- v.init
  
  # Utility vector (patient + caregiver)
  v.utility <- c(
    "mcion_c" = u_mci_pt + u_mci_ic,
    "mciof_c" = u_mci_pt + u_mci_ic,
    "milon_c" = u_mil_pt + u_mil_ic,
    "milof_c" = u_mil_pt + u_mil_ic,
    "mod_c"   = u_mod_pt + u_mod_ic,
    "sev_c"   = u_sev_pt + u_sev_ic,
    "mci_i"   = u_mci_pt_i + u_mci_ic_i,
    "mil_i"   = u_mil_pt_i + u_mil_ic_i,
    "mod_i"   = u_mod_pt_i + u_mod_ic_i,
    "sev_i"   = u_sev_pt_i + u_sev_ic_i,
    "dth"     = 0
  )
  
  # Cost vector (health + social + informal care)
  v.cost <- c(
    "mcion_c" = c_mci_hc + c_mci_sc + c_mci_ic,
    "mciof_c" = c_mci_hc + c_mci_sc + c_mci_ic,
    "milon_c" = c_mil_hc + c_mil_sc + c_mil_ic,
    "milof_c" = c_mil_hc + c_mil_sc + c_mil_ic,
    "mod_c"   = c_mod_hc + c_mod_sc + c_mod_ic,
    "sev_c"   = c_sev_hc + c_sev_sc + c_sev_ic,
    "mci_i"   = c_mci_i_hc + c_mci_i_sc + c_mci_i_ic,
    "mil_i"   = c_mil_i_hc + c_mil_i_sc + c_mil_i_ic,
    "mod_i"   = c_mod_i_hc + c_mod_i_sc + c_mod_i_ic,
    "sev_i"   = c_sev_i_hc + c_sev_i_sc + c_sev_i_ic,
    "dth"     = 0
  )
  
  # Apply treatment cost to on-treatment states for "int"
  if (strat == "int") {
    v.cost["mcion_c"] <- v.cost["mcion_c"] + c_Tx
    v.cost["milon_c"] <- v.cost["milon_c"] + c_Tx
  }
  
  # Outcome matrices
  v.qalys <- numeric(n_cycle)
  v.costs <- numeric(n_cycle)
  v.lys <- numeric(n_cycle)
  
  for (t in 2:n_cycle) {
    m.trace[t, ] <- m.trace[t - 1, ] %*% a.TP[ , , t - 1]
  }
  
  for (t in 1:n_cycle) {
    v.qalys[t] <- sum(m.trace[t, ] * v.utility)
    v.costs[t] <- sum(m.trace[t, ] * v.cost)
    v.lys[t] <- sum(m.trace[t, state_names != "dth"])
  }
  
  # Apply initial treatment cost and utility in cycle 1 for "int"
  if (strat == "int") {
    v.qalys[1] <- v.qalys[1] + u_Tx_start
    v.costs[1] <- v.costs[1] + c_Tx_start
  }
  
  # Half-cycle correction
  v.qalys <- 0.5 * (v.qalys[-n_cycle] + v.qalys[-1])
  v.costs <- 0.5 * (v.costs[-n_cycle] + v.costs[-1])
  v.lys <- 0.5 * (v.lys[-n_cycle] + v.lys[-1])
  
  # Discounting
  v.discount_weights <- 1 / ((1 + discount_QALY) ^ (1:(n_cycle - 1)))
  qalys_total <- sum(v.qalys * v.discount_weights)
  
  v.discount_weights <- 1 / ((1 + discount_COST) ^ (1:(n_cycle - 1)))
  costs_total <- sum(v.costs * v.discount_weights)
  
  v.discount_weights <- 1 / ((1 + discount_EFFECT) ^ (1:(n_cycle - 1)))
  lys_total <- sum(v.lys * v.discount_weights)
  
  return(list(QALYs = qalys_total, Costs = costs_total, LYs = lys_total))
}

#=======================
# PRINT COMPARATIVE RESULTS
#=======================

res_soc <- compute_outcomes("soc")
res_int <- compute_outcomes("int")

cat("===== Comparative Results =====\n")
cat(sprintf("Strategy:        %8s   %8s\n", "SOC", "INT"))
cat(sprintf("Total QALYs:     %8.2f   %8.2f\n", res_soc$QALYs, res_int$QALYs))
cat(sprintf("Total Costs ($): %8.0f   %8.0f\n", res_soc$Costs, res_int$Costs))
cat(sprintf("Total LYs:       %8.2f   %8.2f\n", res_soc$LYs, res_int$LYs))
cat("================================\n")
```
