## Supplementary material for "Replicating Health-Economic Simulation Models for Alzheimer’s Disease Using Artificial Intelligence": final R scripts: o4.R

```
# ===============================
# INPUT PARAMETERS FOR IPECAD MODEL
# ===============================

# Required libraries
library(dplyr)

# State and strategy names
v.names_state <- c("mcion_c", "mciof_c", "milon_c", "milof_c", "mod_c", "sev_c", "mci_i", "mil_i", "mod_i", "sev_i", "dth")
v.names_strat <- c("soc", "int")

# Model settings
age_start <- 71
sex <- "weighted"  # 52% female
n.cycle <- 29
half_cycle_correction <- TRUE
discount_EFFECT <- 0.03
discount_QALY <- 0.03
discount_COST <- 0.03
wtp <- 100000

# Starting state proportions
p.starting_state_mci <- 0.55

# Transition probabilities (natural history)
p.mci_mil <- 0.23
p.mci_mod <- 0
p.mci_sev <- 0
p.mil_mci <- 0.03
p.mil_mod <- 0.35
p.mil_sev <- 0.04
p.mod_mil <- 0.03
p.mod_sev <- 0.42
p.sev_mil <- 0
p.sev_mod <- 0.02

# Transition probabilities to institutional care
p.mci_i <- 0.024
p.mil_i <- 0.038
p.mod_i <- 0.110
p.sev_i <- 0.259

# Hazard ratios for mortality
hr.mort_mci <- 1.82
hr.mort_mil <- 2.92
hr.mort_mod <- 3.85
hr.mort_sev <- 9.52

# Treatment relative risks (on-treatment)
rr.tx_mci_mil <- 0.69
rr.tx_mci_mod <- 1
rr.tx_mci_sev <- 1
rr.tx_mil_mod <- 0.69
rr.tx_mil_sev <- 0.69

# Treatment relative risks (post-discontinuation)
rr.tx_mci_mil_dis <- 1
rr.tx_mci_mod_dis <- 1
rr.tx_mci_sev_dis <- 1
rr.tx_mil_mod_dis <- 1
rr.tx_mil_sev_dis <- 1

# Discontinuation probabilities
p.discontinuation1 <- 0.069
p.discontinuation2 <- 0
discontinuation2_begin <- 2
tx_duration <- 29
tx_waning <- 0
tx_waning_dis <- 0

# Utility values (patient, community)
u.mci_pt <- 0.851 - 0.17
u.mil_pt <- 0.851 - 0.22
u.mod_pt <- 0.851 - 0.36
u.sev_pt <- 0.851 - 0.53

# Utility values (patient, institution)
u.mci_pt_i <- 0.851 - 0.17
u.mil_pt_i <- 0.851 - 0.19
u.mod_pt_i <- 0.851 - 0.42
u.sev_pt_i <- 0.851 - 0.59

# Utility values (caregiver, community)
u.mci_ic <- -0.03
u.mil_ic <- -0.05
u.mod_ic <- -0.08
u.sev_ic <- -0.10

# Utility values (caregiver, institution)
u.mci_ic_i <- -0.03
u.mil_ic_i <- -0.05
u.mod_ic_i <- -0.08
u.sev_ic_i <- -0.10

# Utility for treatment initiation (one-time, not half-cycle corrected)
u.Tx_start <- -0.14 * (12 / 52) * 0.035

# Costs (patient health care, community)
c.mci_hc <- 6042 * 1.12 + 460
c.mil_hc <- 6042 * 1.56 + 965 + 0.21 * 365 * 0.333
c.mod_hc <- 6042 * 1.93 + 1544 + 0.66 * 365 * 0.333
c.sev_hc <- 6042 * 1.93 + 1930

# Costs (patient health care, institution)
c.mci_i_hc <- c.mci_hc
c.mil_i_hc <- c.mil_hc
c.mod_i_hc <- c.mod_hc
c.sev_i_hc <- c.sev_hc

# Costs (patient social care, community)
c.mci_sc <- 0
c.mil_sc <- 0
c.mod_sc <- 0
c.sev_sc <- 0

# Costs (patient social care, institution)
c.mci_i_sc <- 7394 * 12
c.mil_i_sc <- 7394 * 12
c.mod_i_sc <- 7394 * 12
c.sev_i_sc <- 7394 * 12

# Costs (informal care, community)
c.mci_ic <- 69 * 12 * 32.46 + 0.204 * 0.049 * 20 * 52 * 32.46
c.mil_ic <- 113 * 12 * 32.46 + 0.112 * 0.086 * 20 * 52 * 32.46
c.mod_ic <- 169 * 12 * 32.46
c.sev_ic <- 298 * 12 * 32.46

# Costs (informal care, institution)
c.mci_i_ic <- 69 * 12 * 32.46 * 0.44 + 0.204 * 0.049 * 20 * 52 * 32.46
c.mil_i_ic <- 113 * 12 * 32.46 * 0.44 + 0.112 * 0.086 * 20 * 52 * 32.46
c.mod_i_ic <- 169 * 12 * 32.46 * 0.44
c.sev_i_ic <- 298 * 12 * 32.46 * 0.44

# Treatment costs
c.Tx <- 26500 + (52 / 2) * 78.35
c.Tx_start <- 261.10 * 4 + 261.10 * 3 * 0.215


# ===============================
# LOAD AND PROCESS LIFE TABLE DATA
# ===============================

# Load the life table CSV file
lifetable <- read.csv("lifetable_US_2019_ssa.csv")

# Convert annual mortality probabilities to annual mortality rates using:
# rate = -log(1 - probability)
lifetable$male <- -log(1 - lifetable$male)
lifetable$female <- -log(1 - lifetable$female)

# Compute the weighted average of the mortality rates
# Assuming 52% female and 48% male (from prior 'sex' parameter)
lifetable$weighted <- 0.48 * lifetable$male + 0.52 * lifetable$female

# Resulting data frame with required columns
lifetable_processed <- lifetable[, c("age", "male", "female", "weighted")]

# Print or return the processed life table
lifetable_processed

# ===============================
# TIME-DEPENDENT TRANSITION PROBABILITIES - "soc" STRATEGY
# ===============================

# Number of cycles
n_cycles <- n.cycle

# Disease progression probabilities per cycle (fixed over time for SOC strategy)
v.p.mci_mil_soc <- rep(p.mci_mil, n_cycles)
v.p.mci_mod_soc <- rep(p.mci_mod, n_cycles)
v.p.mci_sev_soc <- rep(p.mci_sev, n_cycles)
v.p.mil_mci_soc <- rep(p.mil_mci, n_cycles)
v.p.mil_mod_soc <- rep(p.mil_mod, n_cycles)
v.p.mil_sev_soc <- rep(p.mil_sev, n_cycles)
v.p.mod_mil_soc <- rep(p.mod_mil, n_cycles)
v.p.mod_sev_soc <- rep(p.mod_sev, n_cycles)
v.p.sev_mil_soc <- rep(p.sev_mil, n_cycles)
v.p.sev_mod_soc <- rep(p.sev_mod, n_cycles)

# Staying probabilities (excluding death and institutional transitions)
v.p.mci_stay_soc <- 1 - v.p.mci_mil_soc - v.p.mci_mod_soc - v.p.mci_sev_soc
v.p.mil_stay_soc <- 1 - v.p.mil_mci_soc - v.p.mil_mod_soc - v.p.mil_sev_soc
v.p.mod_stay_soc <- 1 - v.p.mod_mil_soc - v.p.mod_sev_soc
v.p.sev_stay_soc <- 1 - v.p.sev_mil_soc - v.p.sev_mod_soc

# Time-dependent institutionalization probabilities per cycle
v.p.mci_i <- rep(p.mci_i, n_cycles)
v.p.mil_i <- rep(p.mil_i, n_cycles)
v.p.mod_i <- rep(p.mod_i, n_cycles)
v.p.sev_i <- rep(p.sev_i, n_cycles)

# Discontinuation vector per cycle
v.p.discontinuation <- rep(0, n_cycles)
v.p.discontinuation[1] <- p.discontinuation1
v.p.discontinuation[2:(discontinuation2_begin - 1)] <- p.discontinuation1
v.p.discontinuation[discontinuation2_begin:min(tx_duration, n_cycles)] <- p.discontinuation2
if (tx_duration < n_cycles) {
  v.p.discontinuation[(tx_duration + 1):n_cycles] <- 1
}

# Mortality vector based on age- and sex-specific weighted mortality rates
# Assume 'lifetable_processed' is the processed dataframe from previous step
v.mort_soc <- lifetable_processed$weighted[match(age_start:(age_start + n_cycles - 1), lifetable_processed$age)]
# ===============================
# TIME-DEPENDENT TRANSITION PROBABILITIES - "int" STRATEGY
# ===============================

# On-treatment transitions (from "mcion_c" and "milon_c")
v.p.mci_mil_int <- numeric(n_cycles)
v.p.mci_mod_int <- numeric(n_cycles)
v.p.mci_sev_int <- numeric(n_cycles)
v.p.mci_stay_int <- numeric(n_cycles)

v.p.mil_mci_int <- rep(p.mil_mci, n_cycles)  # backward transition (no treatment effect)
v.p.mil_mod_int <- numeric(n_cycles)
v.p.mil_sev_int <- numeric(n_cycles)
v.p.mil_stay_int <- numeric(n_cycles)

# Off-treatment (post-discontinuation) transitions (from "mciof_c" and "milof_c")
v.p.mci_mil_post <- numeric(n_cycles)
v.p.mci_mod_post <- numeric(n_cycles)
v.p.mci_sev_post <- numeric(n_cycles)
v.p.mci_stay_post <- numeric(n_cycles)

v.p.mil_mci_post <- rep(p.mil_mci, n_cycles)  # backward transition (no treatment effect)
v.p.mil_mod_post <- numeric(n_cycles)
v.p.mil_sev_post <- numeric(n_cycles)
v.p.mil_stay_post <- numeric(n_cycles)

# Loop over each cycle to apply treatment effect and waning
for (cycle in 1:n_cycles) {
  
  # Waning factor
  waning_factor <- (1 - tx_waning)^(cycle - 1)
  waning_factor_dis <- (1 - tx_waning_dis)^(cycle - 1)
  
  # ============ ON-TREATMENT ============
  # MCI → mild
  rate <- -log(1 - p.mci_mil)
  rate_adj <- rate * rr.tx_mci_mil^waning_factor
  v.p.mci_mil_int[cycle] <- 1 - exp(-rate_adj)
  
  # MCI → moderate
  rate <- -log(1 - p.mci_mod)
  rate_adj <- rate * rr.tx_mci_mod^waning_factor
  v.p.mci_mod_int[cycle] <- 1 - exp(-rate_adj)
  
  # MCI → severe
  rate <- -log(1 - p.mci_sev)
  rate_adj <- rate * rr.tx_mci_sev^waning_factor
  v.p.mci_sev_int[cycle] <- 1 - exp(-rate_adj)
  
  # MCI stay
  v.p.mci_stay_int[cycle] <- 1 - (v.p.mci_mil_int[cycle] + v.p.mci_mod_int[cycle] + v.p.mci_sev_int[cycle])
  
  # Mild → moderate
  rate <- -log(1 - p.mil_mod)
  rate_adj <- rate * rr.tx_mil_mod^waning_factor
  v.p.mil_mod_int[cycle] <- 1 - exp(-rate_adj)
  
  # Mild → severe
  rate <- -log(1 - p.mil_sev)
  rate_adj <- rate * rr.tx_mil_sev^waning_factor
  v.p.mil_sev_int[cycle] <- 1 - exp(-rate_adj)
  
  # Mild stay
  v.p.mil_stay_int[cycle] <- 1 - (v.p.mil_mci_int[cycle] + v.p.mil_mod_int[cycle] + v.p.mil_sev_int[cycle])
  
  # ============ POST-DISCONTINUATION ============
  # MCI → mild
  rate <- -log(1 - p.mci_mil)
  rate_adj <- rate * rr.tx_mci_mil_dis^waning_factor_dis
  v.p.mci_mil_post[cycle] <- 1 - exp(-rate_adj)
  
  # MCI → moderate
  rate <- -log(1 - p.mci_mod)
  rate_adj <- rate * rr.tx_mci_mod_dis^waning_factor_dis
  v.p.mci_mod_post[cycle] <- 1 - exp(-rate_adj)
  
  # MCI → severe
  rate <- -log(1 - p.mci_sev)
  rate_adj <- rate * rr.tx_mci_sev_dis^waning_factor_dis
  v.p.mci_sev_post[cycle] <- 1 - exp(-rate_adj)
  
  # MCI stay (off-treatment)
  v.p.mci_stay_post[cycle] <- 1 - (v.p.mci_mil_post[cycle] + v.p.mci_mod_post[cycle] + v.p.mci_sev_post[cycle])
  
  # Mild → moderate
  rate <- -log(1 - p.mil_mod)
  rate_adj <- rate * rr.tx_mil_mod_dis^waning_factor_dis
  v.p.mil_mod_post[cycle] <- 1 - exp(-rate_adj)
  
  # Mild → severe
  rate <- -log(1 - p.mil_sev)
  rate_adj <- rate * rr.tx_mil_sev_dis^waning_factor_dis
  v.p.mil_sev_post[cycle] <- 1 - exp(-rate_adj)
  
  # Mild stay (off-treatment)
  v.p.mil_stay_post[cycle] <- 1 - (v.p.mil_mci_post[cycle] + v.p.mil_mod_post[cycle] + v.p.mil_sev_post[cycle])
}
# ===============================
# TRANSITION MATRIX INITIALIZATION FUNCTION
# ===============================

build_transition_matrix <- function(strat) {
  # Initialize transition probability array: states × states × cycles
  n_states <- length(v.names_state)
  a.TP <- array(0, dim = c(n_states, n_states, n.cycle),
                dimnames = list(v.names_state, v.names_state, NULL))
  
  # Loop over each cycle
  for (t in 1:n.cycle) {
    
    # Mortality rate for current cycle
    mort_rate <- v.mort_soc[t]
    
    # === Transitions to death from each state ===
    a.TP["mcion_c", "dth", t] <- 1 - exp(-mort_rate * hr.mort_mci)
    a.TP["mciof_c", "dth", t] <- 1 - exp(-mort_rate * hr.mort_mci)
    a.TP["milon_c", "dth", t] <- 1 - exp(-mort_rate * hr.mort_mil)
    a.TP["milof_c", "dth", t] <- 1 - exp(-mort_rate * hr.mort_mil)
    a.TP["mod_c",   "dth", t] <- 1 - exp(-mort_rate * hr.mort_mod)
    a.TP["sev_c",   "dth", t] <- 1 - exp(-mort_rate * hr.mort_sev)
    a.TP["mci_i",   "dth", t] <- 1 - exp(-mort_rate * hr.mort_mci)
    a.TP["mil_i",   "dth", t] <- 1 - exp(-mort_rate * hr.mort_mil)
    a.TP["mod_i",   "dth", t] <- 1 - exp(-mort_rate * hr.mort_mod)
    a.TP["sev_i",   "dth", t] <- 1 - exp(-mort_rate * hr.mort_sev)
    
    # === Death is an absorbing state ===
    a.TP["dth", "dth", t] <- 1
    
    # === Placeholders for future prompts ===
    
    # === [INSERT PROMPT 5B HERE] — Transitions from on-treatment community states ===
    
    # ---- From mcion_c ----
    p_surv <- 1 - a.TP["mcion_c", "dth", t]
    
    if (strat == "soc") {
      p_mci_mil <- v.p.mci_mil_soc[t]
      p_mci_mod <- v.p.mci_mod_soc[t]
      p_mci_sev <- v.p.mci_sev_soc[t]
      p_mci_stay <- v.p.mci_stay_soc[t]
      p_inst <- v.p.mci_i[t]
      p_disc <- v.p.discontinuation[t]
    } else if (strat == "int") {
      p_mci_mil <- v.p.mci_mil_int[t]
      p_mci_mod <- v.p.mci_mod_int[t]
      p_mci_sev <- v.p.mci_sev_int[t]
      p_mci_stay <- v.p.mci_stay_int[t]
      p_inst <- v.p.mci_i[t]
      p_disc <- v.p.discontinuation[t]
    }
    
    # To milon_c (on-treatment): MCI → Mild, stays on treatment
    a.TP["mcion_c", "milon_c", t] <- p_mci_mil * p_surv * (1 - p_inst) * (1 - p_disc)
    # To milof_c (off-treatment): MCI → Mild, discontinues
    a.TP["mcion_c", "milof_c", t] <- p_mci_mil * p_surv * (1 - p_inst) * p_disc
    # To mod_c: no discontinuation factor
    a.TP["mcion_c", "mod_c", t] <- p_mci_mod * p_surv * (1 - p_inst)
    a.TP["mcion_c", "sev_c", t] <- p_mci_sev * p_surv * (1 - p_inst)
    # To mci_i: institutionalization
    a.TP["mcion_c", "mci_i", t] <- (p_mci_mil + p_mci_mod + p_mci_sev + p_mci_stay) * p_surv * p_inst
    # To mcion_c: stay in MCI, on-treatment
    a.TP["mcion_c", "mcion_c", t] <- p_mci_stay * p_surv * (1 - p_inst) * (1 - p_disc)
    # To mciof_c: stay in MCI, but discontinues
    a.TP["mcion_c", "mciof_c", t] <- p_mci_stay * p_surv * (1 - p_inst) * p_disc
    
    # ---- From milon_c ----
    p_surv <- 1 - a.TP["milon_c", "dth", t]
    
    if (strat == "soc") {
      p_mil_mci <- v.p.mil_mci_soc[t]
      p_mil_mod <- v.p.mil_mod_soc[t]
      p_mil_sev <- v.p.mil_sev_soc[t]
      p_mil_stay <- v.p.mil_stay_soc[t]
      p_inst <- v.p.mil_i[t]
      p_disc <- v.p.discontinuation[t]
    } else if (strat == "int") {
      p_mil_mci <- v.p.mil_mci_int[t]
      p_mil_mod <- v.p.mil_mod_int[t]
      p_mil_sev <- v.p.mil_sev_int[t]
      p_mil_stay <- v.p.mil_stay_int[t]
      p_inst <- v.p.mil_i[t]
      p_disc <- v.p.discontinuation[t]
    }
    
    # To milon_c: stay in mild dementia, on-treatment
    a.TP["milon_c", "milon_c", t] <- p_mil_stay * p_surv * (1 - p_inst) * (1 - p_disc)
    # To milof_c: stay in mild dementia, discontinues
    a.TP["milon_c", "milof_c", t] <- p_mil_stay * p_surv * (1 - p_inst) * p_disc
    # To mciof_c: backward to MCI, discontinues
    a.TP["milon_c", "mciof_c", t] <- p_mil_mci * p_surv * (1 - p_inst)
    # To mod_c: no discontinuation factor
    a.TP["milon_c", "mod_c", t] <- p_mil_mod * p_surv * (1 - p_inst)
    a.TP["milon_c", "sev_c", t] <- p_mil_sev * p_surv * (1 - p_inst)
    # To mil_i: institutionalization
    a.TP["milon_c", "mil_i", t] <- (p_mil_mci + p_mil_mod + p_mil_sev + p_mil_stay) * p_surv * p_inst
    
    
    # === [INSERT PROMPT 5C HERE] — Transitions from off-treatment and moderate/severe community states ===
    
    # ---- From mciof_c ----
    p_surv <- 1 - a.TP["mciof_c", "dth", t]
    
    if (strat == "soc") {
      p_mci_mil <- v.p.mci_mil_soc[t]
      p_mci_mod <- v.p.mci_mod_soc[t]
      p_mci_sev <- v.p.mci_sev_soc[t]
      p_mci_stay <- v.p.mci_stay_soc[t]
      p_inst <- v.p.mci_i[t]
    } else if (strat == "int") {
      p_mci_mil <- v.p.mci_mil_post[t]
      p_mci_mod <- v.p.mci_mod_post[t]
      p_mci_sev <- v.p.mci_sev_post[t]
      p_mci_stay <- v.p.mci_stay_post[t]
      p_inst <- v.p.mci_i[t]
    }
    
    a.TP["mciof_c", "milof_c", t] <- p_mci_mil * p_surv * (1 - p_inst)
    a.TP["mciof_c", "mod_c",    t] <- p_mci_mod * p_surv * (1 - p_inst)
    a.TP["mciof_c", "sev_c",    t] <- p_mci_sev * p_surv * (1 - p_inst)
    a.TP["mciof_c", "mciof_c",  t] <- p_mci_stay * p_surv * (1 - p_inst)
    a.TP["mciof_c", "mci_i",    t] <- (p_mci_mil + p_mci_mod + p_mci_sev + p_mci_stay) * p_surv * p_inst
    
    # ---- From milof_c ----
    p_surv <- 1 - a.TP["milof_c", "dth", t]
    
    if (strat == "soc") {
      p_mil_mci <- v.p.mil_mci_soc[t]
      p_mil_mod <- v.p.mil_mod_soc[t]
      p_mil_sev <- v.p.mil_sev_soc[t]
      p_mil_stay <- v.p.mil_stay_soc[t]
      p_inst <- v.p.mil_i[t]
    } else if (strat == "int") {
      p_mil_mci <- v.p.mil_mci_post[t]
      p_mil_mod <- v.p.mil_mod_post[t]
      p_mil_sev <- v.p.mil_sev_post[t]
      p_mil_stay <- v.p.mil_stay_post[t]
      p_inst <- v.p.mil_i[t]
    }
    
    a.TP["milof_c", "mciof_c",  t] <- p_mil_mci * p_surv * (1 - p_inst)
    a.TP["milof_c", "mod_c",    t] <- p_mil_mod * p_surv * (1 - p_inst)
    a.TP["milof_c", "sev_c",    t] <- p_mil_sev * p_surv * (1 - p_inst)
    a.TP["milof_c", "milof_c",  t] <- p_mil_stay * p_surv * (1 - p_inst)
    a.TP["milof_c", "mil_i",    t] <- (p_mil_mci + p_mil_mod + p_mil_sev + p_mil_stay) * p_surv * p_inst
    
    # ---- From mod_c ----
    p_surv <- 1 - a.TP["mod_c", "dth", t]
    p_mod_mil <- v.p.mod_mil_soc[t]
    p_mod_sev <- v.p.mod_sev_soc[t]
    p_mod_stay <- v.p.mod_stay_soc[t]
    p_inst <- v.p.mod_i[t]
    
    a.TP["mod_c", "milof_c", t] <- p_mod_mil * p_surv * (1 - p_inst)
    a.TP["mod_c", "sev_c",   t] <- p_mod_sev * p_surv * (1 - p_inst)
    a.TP["mod_c", "mod_c",   t] <- p_mod_stay * p_surv * (1 - p_inst)
    a.TP["mod_c", "mod_i",   t] <- (p_mod_mil + p_mod_sev + p_mod_stay) * p_surv * p_inst
    
    # ---- From sev_c ----
    p_surv <- 1 - a.TP["sev_c", "dth", t]
    p_sev_mil <- v.p.sev_mil_soc[t]
    p_sev_mod <- v.p.sev_mod_soc[t]
    p_sev_stay <- v.p.sev_stay_soc[t]
    p_inst <- v.p.sev_i[t]
    
    a.TP["sev_c", "milof_c", t] <- p_sev_mil * p_surv * (1 - p_inst)
    a.TP["sev_c", "mod_c",   t] <- p_sev_mod * p_surv * (1 - p_inst)
    a.TP["sev_c", "sev_c",   t] <- p_sev_stay * p_surv * (1 - p_inst)
    a.TP["sev_c", "sev_i",   t] <- (p_sev_mil + p_sev_mod + p_sev_stay) * p_surv * p_inst
    
    # === [INSERT PROMPT 5D HERE] — Transitions from institutionalized states ===
    
    # ---- From mci_i ----
    p_surv <- 1 - a.TP["mci_i", "dth", t]
    
    if (strat == "soc") {
      p_mci_mil <- v.p.mci_mil_soc[t]
      p_mci_mod <- v.p.mci_mod_soc[t]
      p_mci_sev <- v.p.mci_sev_soc[t]
      p_mci_stay <- v.p.mci_stay_soc[t]
    } else if (strat == "int") {
      p_mci_mil <- v.p.mci_mil_post[t]
      p_mci_mod <- v.p.mci_mod_post[t]
      p_mci_sev <- v.p.mci_sev_post[t]
      p_mci_stay <- v.p.mci_stay_post[t]
    }
    
    a.TP["mci_i", "mil_i", t] <- p_mci_mil * p_surv
    a.TP["mci_i", "mod_i", t] <- p_mci_mod * p_surv
    a.TP["mci_i", "sev_i", t] <- p_mci_sev * p_surv
    a.TP["mci_i", "mci_i", t] <- p_mci_stay * p_surv
    
    # ---- From mil_i ----
    p_surv <- 1 - a.TP["mil_i", "dth", t]
    
    if (strat == "soc") {
      p_mil_mci <- v.p.mil_mci_soc[t]
      p_mil_mod <- v.p.mil_mod_soc[t]
      p_mil_sev <- v.p.mil_sev_soc[t]
      p_mil_stay <- v.p.mil_stay_soc[t]
    } else if (strat == "int") {
      p_mil_mci <- v.p.mil_mci_post[t]
      p_mil_mod <- v.p.mil_mod_post[t]
      p_mil_sev <- v.p.mil_sev_post[t]
      p_mil_stay <- v.p.mil_stay_post[t]
    }
    
    a.TP["mil_i", "mci_i", t] <- p_mil_mci * p_surv
    a.TP["mil_i", "mod_i", t] <- p_mil_mod * p_surv
    a.TP["mil_i", "sev_i", t] <- p_mil_sev * p_surv
    a.TP["mil_i", "mil_i", t] <- p_mil_stay * p_surv
    
    # ---- From mod_i ----
    p_surv <- 1 - a.TP["mod_i", "dth", t]
    p_mod_mil <- v.p.mod_mil_soc[t]
    p_mod_sev <- v.p.mod_sev_soc[t]
    p_mod_stay <- v.p.mod_stay_soc[t]
    
    a.TP["mod_i", "mil_i", t] <- p_mod_mil * p_surv
    a.TP["mod_i", "sev_i", t] <- p_mod_sev * p_surv
    a.TP["mod_i", "mod_i", t] <- p_mod_stay * p_surv
    
    # ---- From sev_i ----
    p_surv <- 1 - a.TP["sev_i", "dth", t]
    p_sev_mil <- v.p.sev_mil_soc[t]
    p_sev_mod <- v.p.sev_mod_soc[t]
    p_sev_stay <- v.p.sev_stay_soc[t]
    
    a.TP["sev_i", "mil_i", t] <- p_sev_mil * p_surv
    a.TP["sev_i", "mod_i", t] <- p_sev_mod * p_surv
    a.TP["sev_i", "sev_i", t] <- p_sev_stay * p_surv
    
    # === Normalize rows to ensure probabilities sum to 1 ===
    for (s in v.names_state) {
      row_sum <- sum(a.TP[s, , t])
      if (row_sum > 0 && row_sum != 1) {
        a.TP[s, , t] <- a.TP[s, , t] / row_sum
      }
    }
  }
  
  return(a.TP)
}
# ===============================
# MARKOV TRACE AND TOTAL OUTCOMES FUNCTION
# ===============================

compute_outcomes <- function(strat) {
  # Build transition probability matrix
  a.TP <- build_transition_matrix(strat)
  
  # Initialize trace matrix
  m.M <- matrix(0, nrow = n.cycle + 1, ncol = length(v.names_state),
                dimnames = list(0:n.cycle, v.names_state))
  
  # Initial distribution
  m.M[1, ] <- 0
  if (strat == "soc") {
    m.M[1, "mciof_c"] <- p.starting_state_mci
    m.M[1, "milof_c"] <- 1 - p.starting_state_mci
  } else if (strat == "int") {
    m.M[1, "mcion_c"] <- p.starting_state_mci
    m.M[1, "milon_c"] <- 1 - p.starting_state_mci
  }
  
  # Initialize outcome matrices
  v.QALYs <- numeric(n.cycle + 1)
  v.Costs <- numeric(n.cycle + 1)
  v.LYs   <- numeric(n.cycle + 1)
  
  # Define utility vectors
  v.u <- c(
    mcion_c = u.mci_pt,   mciof_c = u.mci_pt,
    milon_c = u.mil_pt,   milof_c = u.mil_pt,
    mod_c   = u.mod_pt,   sev_c   = u.sev_pt,
    mci_i   = u.mci_pt_i, mil_i   = u.mil_pt_i,
    mod_i   = u.mod_pt_i, sev_i   = u.sev_pt_i,
    dth     = 0
  )
  
  # Define cost vectors
  v.c <- c(
    mcion_c = c.mci_hc + c.mci_sc + c.mci_ic,
    mciof_c = c.mci_hc + c.mci_sc + c.mci_ic,
    milon_c = c.mil_hc + c.mil_sc + c.mil_ic,
    milof_c = c.mil_hc + c.mil_sc + c.mil_ic,
    mod_c   = c.mod_hc + c.mod_sc + c.mod_ic,
    sev_c   = c.sev_hc + c.sev_sc + c.sev_ic,
    mci_i   = c.mci_i_hc + c.mci_i_sc + c.mci_i_ic,
    mil_i   = c.mil_i_hc + c.mil_i_sc + c.mil_i_ic,
    mod_i   = c.mod_i_hc + c.mod_i_sc + c.mod_i_ic,
    sev_i   = c.sev_i_hc + c.sev_i_sc + c.sev_i_ic,
    dth     = 0
  )
  
  # Repeating treatment cost each year in on-treatment states
  on_treatment_states <- c("mcion_c", "milon_c")
  
  for (t in 1:n.cycle) {
    # Update Markov trace
    m.M[t + 1, ] <- m.M[t, ] %*% a.TP[ , , t]
    
    # Occupancy at time t
    state_occupancy <- m.M[t, ]
    
    # Apply state utilities and costs
    qalys_t <- sum(state_occupancy * v.u)
    costs_t <- sum(state_occupancy * v.c)
    lys_t   <- sum(state_occupancy)
    
    # Add treatment costs for intervention strategy
    if (strat == "int") {
      costs_t <- costs_t + sum(state_occupancy[on_treatment_states]) * c.Tx
    }
    
    # Add initial costs and utility once
    if (t == 1 && strat == "int") {
      costs_t <- costs_t + c.Tx_start
      qalys_t <- qalys_t + u.Tx_start
    }
    
    # Apply discounting
    v.QALYs[t] <- qalys_t / (1 + discount_QALY)^(t - 1)
    v.Costs[t] <- costs_t / (1 + discount_COST)^(t - 1)
    v.LYs[t]   <- lys_t / (1 + discount_EFFECT)^(t - 1)
  }
  
  # Half-cycle correction (average current and next cycle)
  v.QALYs <- (v.QALYs[-1] + v.QALYs[-length(v.QALYs)]) / 2
  v.Costs <- (v.Costs[-1] + v.Costs[-length(v.Costs)]) / 2
  v.LYs   <- (v.LYs[-1] + v.LYs[-length(v.LYs)]) / 2
  
  # Summarize total outcomes
  total_QALYs <- sum(v.QALYs)
  total_Costs <- sum(v.Costs)
  total_LYs   <- sum(v.LYs)
  
  return(c(QALYs = total_QALYs, Costs = total_Costs, LYs = total_LYs))
}

# ===============================
# COMPARATIVE RESULTS
# ===============================

# Compute outcomes
results_soc <- compute_outcomes("soc")
results_int <- compute_outcomes("int")

# Print results
cat("=== IPECAD MODEL RESULTS ===\n")
cat("Strategy: Standard of Care (SOC)\n")
print(results_soc)
cat("\nStrategy: Intervention (INT)\n")
print(results_int)

# Incremental outcomes
incremental <- results_int - results_soc
cat("\n=== Incremental Outcomes (INT - SOC) ===\n")
print(incremental)
```
