## Supplementary material for "Replicating Health-Economic Simulation Models for Alzheimer’s Disease Using Artificial Intelligence": final R scripts: o5.R

```
# IPECAD Markov Model - Input Parameters (Cross-validation with ICER model)
# Required packages
library(dampack)

# Descriptive names
state_names <- c("mcion_c", "mciof_c", "milon_c", "milof_c", "mod_c", "sev_c",
                 "mci_i", "mil_i", "mod_i", "sev_i", "dth")
strategy_names <- c("soc", "int")

# Population characteristics
age_start <- 71
sex <- "weighted"  # 52% female
p_starting_state_mci <- 0.55
n_cycle <- 29

# Disease progression probabilities
p_mci_mil <- 0.23
p_mci_mod <- 0
p_mci_sev <- 0
p_mil_mci <- 0.03
p_mil_mod <- 0.35
p_mil_sev <- 0.04
p_mod_mil <- 0.03
p_mod_sev <- 0.42
p_sev_mil <- 0
p_sev_mod <- 0.02

# Institution transition probabilities
p_mci_i <- 0.024
p_mil_i <- 0.038
p_mod_i <- 0.110
p_sev_i <- 0.259

# Mortality hazard ratios (relative to general population)
hr_mort_mci <- 1.82
hr_mort_mil <- 2.92
hr_mort_mod <- 3.85
hr_mort_sev <- 9.52

# Treatment effects (on-treatment)
rr_tx_mci_mil <- 0.69
rr_tx_mci_mod <- 1
rr_tx_mci_sev <- 1
rr_tx_mil_mod <- 0.69
rr_tx_mil_sev <- 0.69

# Treatment effects (off-treatment)
rr_tx_mci_mil_dis <- 1
rr_tx_mci_mod_dis <- 1
rr_tx_mci_sev_dis <- 1
rr_tx_mil_mod_dis <- 1
rr_tx_mil_sev_dis <- 1

# Discontinuation probabilities
p_discontinuation1 <- 0.069
p_discontinuation2 <- 0
discontinuation2_begin <- 2
tx_duration <- 29
tx_waning <- 0
tx_waning_dis <- 0

# Utilities (patients - community)
u_mci_pt <- 0.851 - 0.17
u_mil_pt <- 0.851 - 0.22
u_mod_pt <- 0.851 - 0.36
u_sev_pt <- 0.851 - 0.53

# Utilities (patients - institution)
u_mci_pt_i <- 0.851 - 0.17
u_mil_pt_i <- 0.851 - 0.19
u_mod_pt_i <- 0.851 - 0.42
u_sev_pt_i <- 0.851 - 0.59

# Utilities (caregivers - community)
u_mci_ic <- -0.03
u_mil_ic <- -0.05
u_mod_ic <- -0.08
u_sev_ic <- -0.10

# Utilities (caregivers - institution)
u_mci_ic_i <- -0.03
u_mil_ic_i <- -0.05
u_mod_ic_i <- -0.08
u_sev_ic_i <- -0.10

# Additional utility decrement due to ARIA (amyloid-related imaging abnormalities)
u_Tx_start <- -0.14 * (12/52) * 0.035

# Costs (healthcare - community)
c_mci_hc <- 6042 * 1.12 + 460
c_mil_hc <- 6042 * 1.56 + 965 + 0.21 * 365 * 0.333
c_mod_hc <- 6042 * 1.93 + 1544 + 0.66 * 365 * 0.333
c_sev_hc <- 6042 * 1.93 + 1930

# Costs (healthcare - institution)
c_mci_i_hc <- c_mci_hc
c_mil_i_hc <- c_mil_hc
c_mod_i_hc <- c_mod_hc
c_sev_i_hc <- c_sev_hc

# Costs (social care - community)
c_mci_sc <- 0
c_mil_sc <- 0
c_mod_sc <- 0
c_sev_sc <- 0

# Costs (social care - institution)
c_mci_i_sc <- 7394 * 12
c_mil_i_sc <- 7394 * 12
c_mod_i_sc <- 7394 * 12
c_sev_i_sc <- 7394 * 12

# Costs (informal care - community)
c_mci_ic <- 69 * 12 * 32.46 + 0.204 * 0.049 * 20 * 52 * 32.46
c_mil_ic <- 113 * 12 * 32.46 + 0.112 * 0.086 * 20 * 52 * 32.46
c_mod_ic <- 169 * 12 * 32.46
c_sev_ic <- 298 * 12 * 32.46

# Costs (informal care - institution)
c_mci_i_ic <- 69 * 12 * 32.46 * 0.44 + 0.204 * 0.049 * 20 * 52 * 32.46
c_mil_i_ic <- 113 * 12 * 32.46 * 0.44 + 0.112 * 0.086 * 20 * 52 * 32.46
c_mod_i_ic <- 169 * 12 * 32.46 * 0.44
c_sev_i_ic <- 298 * 12 * 32.46 * 0.44

# Cost of treatment
c_Tx <- 26500 + (52/2) * 78.35

# Additional first-cycle costs (monitoring and AE management)
c_Tx_start <- 261.10 * 4 + 261.10 * 3 * 0.215

# Discounting
discount_EFFECT <- 0.03
discount_QALY <- 0.03
discount_COST <- 0.03

# WTP threshold
wtp <- 100000

# Half-cycle correction
half_cycle_correction <- TRUE
# Load the 2019 US life table from CSV
life_table <- read.csv("lifetable_US_2019_ssa.csv")

# Convert annual mortality probabilities to annual mortality rates using:
# rate = -log(1 - probability)
life_table$male <- -log(1 - life_table$male)
life_table$female <- -log(1 - life_table$female)

# Compute sex-weighted average mortality rate (52% female, 48% male)
life_table$weighted <- 0.48 * life_table$male + 0.52 * life_table$female

# Resulting data frame with converted rates and weighted column
life_table <- life_table[, c("age", "male", "female", "weighted")]

# Display the resulting table
life_table
# Define number of cycles
cycles <- n_cycle  # From prompt 1

# Time-dependent transition probabilities for SOC (standard of care) strategy
v.p.mci_mil_soc <- rep(p_mci_mil, cycles)
v.p.mci_mod_soc <- rep(p_mci_mod, cycles)
v.p.mci_sev_soc <- rep(p_mci_sev, cycles)

v.p.mil_mci_soc <- rep(p_mil_mci, cycles)
v.p.mil_mod_soc <- rep(p_mil_mod, cycles)
v.p.mil_sev_soc <- rep(p_mil_sev, cycles)

v.p.mod_mil_soc <- rep(p_mod_mil, cycles)
v.p.mod_sev_soc <- rep(p_mod_sev, cycles)

v.p.sev_mil_soc <- rep(p_sev_mil, cycles)
v.p.sev_mod_soc <- rep(p_sev_mod, cycles)

# Staying probabilities (excluding institutional transitions)
v.p.mci_stay_soc <- 1 - (v.p.mci_mil_soc + v.p.mci_mod_soc + v.p.mci_sev_soc)
v.p.mil_stay_soc <- 1 - (v.p.mil_mci_soc + v.p.mil_mod_soc + v.p.mil_sev_soc)
v.p.mod_stay_soc <- 1 - (v.p.mod_mil_soc + v.p.mod_sev_soc)
v.p.sev_stay_soc <- 1 - (v.p.sev_mil_soc + v.p.sev_mod_soc)

# Time-dependent vectors for institutionalization transitions
v.p.mci_i <- rep(p_mci_i, cycles)
v.p.mil_i <- rep(p_mil_i, cycles)
v.p.mod_i <- rep(p_mod_i, cycles)
v.p.sev_i <- rep(p_sev_i, cycles)

# Discontinuation vector
v.p.discontinuation <- numeric(cycles)
v.p.discontinuation[1] <- p_discontinuation1
if (cycles > 1) {
  v.p.discontinuation[2:tx_duration] <- p_discontinuation2
}
if (tx_duration < cycles) {
  v.p.discontinuation[(tx_duration + 1):cycles] <- 1
}

# Age- and sex-specific mortality rates from weighted life table
v.mortality <- life_table$weighted[life_table$age >= age_start][1:cycles]
# Define intervention strategy transition vectors per cycle
v.p.mci_mil_int <- numeric(n_cycle)
v.p.mci_mod_int <- numeric(n_cycle)
v.p.mci_sev_int <- numeric(n_cycle)
v.p.mil_mci_int <- rep(p_mil_mci, n_cycle)  # backward transition, no treatment effect
v.p.mil_mod_int <- numeric(n_cycle)
v.p.mil_sev_int <- numeric(n_cycle)

v.p.mci_mil_dis_int <- numeric(n_cycle)
v.p.mci_mod_dis_int <- numeric(n_cycle)
v.p.mci_sev_dis_int <- numeric(n_cycle)
v.p.mil_mci_dis_int <- rep(p_mil_mci, n_cycle)  # backward transition, no treatment effect
v.p.mil_mod_dis_int <- numeric(n_cycle)
v.p.mil_sev_dis_int <- numeric(n_cycle)

# On-treatment transitions (mcion_c, milon_c)
for (i in 1:n_cycle) {
  waning_factor <- (1 - tx_waning)^(i - 1)
  
  # MCI → mild
  rate <- -log(1 - p_mci_mil)
  adj_rate <- rate * rr_tx_mci_mil^waning_factor
  v.p.mci_mil_int[i] <- 1 - exp(-adj_rate)
  
  # MCI → moderate
  rate <- -log(1 - p_mci_mod)
  adj_rate <- rate * rr_tx_mci_mod^waning_factor
  v.p.mci_mod_int[i] <- 1 - exp(-adj_rate)
  
  # MCI → severe
  rate <- -log(1 - p_mci_sev)
  adj_rate <- rate * rr_tx_mci_sev^waning_factor
  v.p.mci_sev_int[i] <- 1 - exp(-adj_rate)
  
  # Mild → moderate
  rate <- -log(1 - p_mil_mod)
  adj_rate <- rate * rr_tx_mil_mod^waning_factor
  v.p.mil_mod_int[i] <- 1 - exp(-adj_rate)
  
  # Mild → severe
  rate <- -log(1 - p_mil_sev)
  adj_rate <- rate * rr_tx_mil_sev^waning_factor
  v.p.mil_sev_int[i] <- 1 - exp(-adj_rate)
}

# Off-treatment transitions (mciof_c, milof_c)
for (i in 1:n_cycle) {
  waning_factor <- (1 - tx_waning_dis)^(i - 1)
  
  # MCI → mild
  rate <- -log(1 - p_mci_mil)
  adj_rate <- rate * rr_tx_mci_mil_dis^waning_factor
  v.p.mci_mil_dis_int[i] <- 1 - exp(-adj_rate)
  
  # MCI → moderate
  rate <- -log(1 - p_mci_mod)
  adj_rate <- rate * rr_tx_mci_mod_dis^waning_factor
  v.p.mci_mod_dis_int[i] <- 1 - exp(-adj_rate)
  
  # MCI → severe
  rate <- -log(1 - p_mci_sev)
  adj_rate <- rate * rr_tx_mci_sev_dis^waning_factor
  v.p.mci_sev_dis_int[i] <- 1 - exp(-adj_rate)
  
  # Mild → moderate
  rate <- -log(1 - p_mil_mod)
  adj_rate <- rate * rr_tx_mil_mod_dis^waning_factor
  v.p.mil_mod_dis_int[i] <- 1 - exp(-adj_rate)
  
  # Mild → severe
  rate <- -log(1 - p_mil_sev)
  adj_rate <- rate * rr_tx_mil_sev_dis^waning_factor
  v.p.mil_sev_dis_int[i] <- 1 - exp(-adj_rate)
}

# Staying probabilities - on-treatment
v.p.mci_stay_int <- 1 - (v.p.mci_mil_int + v.p.mci_mod_int + v.p.mci_sev_int)
v.p.mil_stay_int <- 1 - (v.p.mil_mci_int + v.p.mil_mod_int + v.p.mil_sev_int)

# Staying probabilities - off-treatment
v.p.mci_stay_dis_int <- 1 - (v.p.mci_mil_dis_int + v.p.mci_mod_dis_int + v.p.mci_sev_dis_int)
v.p.mil_stay_dis_int <- 1 - (v.p.mil_mci_dis_int + v.p.mil_mod_dis_int + v.p.mil_sev_dis_int)


# Function to build the transition probability array for a given strategy
build_transition_matrix <- function(strat) {
  
  # Initialize transition probability array (states × states × cycles)
  n_states <- length(state_names)
  a.TP <- array(0, dim = c(n_states, n_states, n_cycle),
                dimnames = list(state_names, state_names, NULL))
  
  # Loop over each cycle
  for (t in 1:n_cycle) {
    
    # Mortality rates for this cycle
    rate_mort <- v.mortality[t]
    
    # Transition to death from each state
    a.TP["mcion_c", "dth", t] <- 1 - exp(-rate_mort * hr_mort_mci)
    a.TP["mciof_c", "dth", t] <- 1 - exp(-rate_mort * hr_mort_mci)
    a.TP["milon_c", "dth", t] <- 1 - exp(-rate_mort * hr_mort_mil)
    a.TP["milof_c", "dth", t] <- 1 - exp(-rate_mort * hr_mort_mil)
    a.TP["mod_c", "dth", t]    <- 1 - exp(-rate_mort * hr_mort_mod)
    a.TP["sev_c", "dth", t]    <- 1 - exp(-rate_mort * hr_mort_sev)
    a.TP["mci_i", "dth", t]    <- 1 - exp(-rate_mort * hr_mort_mci)
    a.TP["mil_i", "dth", t]    <- 1 - exp(-rate_mort * hr_mort_mil)
    a.TP["mod_i", "dth", t]    <- 1 - exp(-rate_mort * hr_mort_mod)
    a.TP["sev_i", "dth", t]    <- 1 - exp(-rate_mort * hr_mort_sev)
    
    # Death is absorbing
    a.TP["dth", "dth", t] <- 1
    
    # ------------------- PLACEHOLDERS FOR PROMPTS 5B–5D -------------------
    
    # Transitions from "mcion_c" (MCI on-treatment, community)
    if (strat == "soc") {
      tp_mci_mil <- v.p.mci_mil_soc[t]
      tp_mci_mod <- v.p.mci_mod_soc[t]
      tp_mci_sev <- v.p.mci_sev_soc[t]
      tp_mci_stay <- v.p.mci_stay_soc[t]
    } else {
      tp_mci_mil <- v.p.mci_mil_int[t]
      tp_mci_mod <- v.p.mci_mod_int[t]
      tp_mci_sev <- v.p.mci_sev_int[t]
      tp_mci_stay <- v.p.mci_stay_int[t]
    }
    p_mort <- a.TP["mcion_c", "dth", t]
    p_surv <- 1 - p_mort
    p_disc <- v.p.discontinuation[t]
    p_inst <- v.p.mci_i[t]
    p_comm <- 1 - p_inst
    
    # To milon_c (mild, on-treatment)
    a.TP["mcion_c", "milon_c", t] <- tp_mci_mil * p_comm * p_surv * (1 - p_disc)
    # To milof_c (mild, off-treatment)
    a.TP["mcion_c", "milof_c", t] <- tp_mci_mil * p_comm * p_surv * p_disc
    # To mod_c
    a.TP["mcion_c", "mod_c", t] <- tp_mci_mod * p_surv
    # To sev_c
    a.TP["mcion_c", "sev_c", t] <- tp_mci_sev * p_surv
    # To mci_i
    a.TP["mcion_c", "mci_i", t] <- tp_mci_stay * p_inst * p_surv
    # To mcion_c (stay in state)
    a.TP["mcion_c", "mcion_c", t] <- tp_mci_stay * p_comm * p_surv * (1 - p_disc)
    # To mciof_c (off-treatment version of self)
    a.TP["mcion_c", "mciof_c", t] <- tp_mci_stay * p_comm * p_surv * p_disc
    
    # Transitions from "milon_c" (mild on-treatment, community)
    if (strat == "soc") {
      tp_mil_mci <- v.p.mil_mci_soc[t]
      tp_mil_mod <- v.p.mil_mod_soc[t]
      tp_mil_sev <- v.p.mil_sev_soc[t]
      tp_mil_stay <- v.p.mil_stay_soc[t]
    } else {
      tp_mil_mci <- v.p.mil_mci_int[t]
      tp_mil_mod <- v.p.mil_mod_int[t]
      tp_mil_sev <- v.p.mil_sev_int[t]
      tp_mil_stay <- v.p.mil_stay_int[t]
    }
    p_mort <- a.TP["milon_c", "dth", t]
    p_surv <- 1 - p_mort
    p_disc <- v.p.discontinuation[t]
    p_inst <- v.p.mil_i[t]
    p_comm <- 1 - p_inst
    
    # To mciof_c (MCI, off-treatment)
    a.TP["milon_c", "mciof_c", t] <- tp_mil_mci * p_comm * p_surv * p_disc
    # To mod_c
    a.TP["milon_c", "mod_c", t] <- tp_mil_mod * p_surv
    # To sev_c
    a.TP["milon_c", "sev_c", t] <- tp_mil_sev * p_surv
    # To mil_i
    a.TP["milon_c", "mil_i", t] <- tp_mil_stay * p_inst * p_surv
    # To milon_c (stay in state)
    a.TP["milon_c", "milon_c", t] <- tp_mil_stay * p_comm * p_surv * (1 - p_disc)
    # To milof_c (off-treatment version of self)
    a.TP["milon_c", "milof_c", t] <- tp_mil_stay * p_comm * p_surv * p_disc
    
    
    # Transitions from "mciof_c" (MCI off-treatment, community)
    if (strat == "soc") {
      tp_mci_mil <- v.p.mci_mil_soc[t]
      tp_mci_mod <- v.p.mci_mod_soc[t]
      tp_mci_sev <- v.p.mci_sev_soc[t]
      tp_mci_stay <- v.p.mci_stay_soc[t]
    } else {
      tp_mci_mil <- v.p.mci_mil_dis_int[t]
      tp_mci_mod <- v.p.mci_mod_dis_int[t]
      tp_mci_sev <- v.p.mci_sev_dis_int[t]
      tp_mci_stay <- v.p.mci_stay_dis_int[t]
    }
    p_mort <- a.TP["mciof_c", "dth", t]
    p_surv <- 1 - p_mort
    p_inst <- v.p.mci_i[t]
    p_comm <- 1 - p_inst
    
    a.TP["mciof_c", "milof_c", t] <- tp_mci_mil * p_comm * p_surv
    a.TP["mciof_c", "mod_c", t]   <- tp_mci_mod * p_surv
    a.TP["mciof_c", "sev_c", t]   <- tp_mci_sev * p_surv
    a.TP["mciof_c", "mci_i", t]   <- tp_mci_stay * p_inst * p_surv
    a.TP["mciof_c", "mciof_c", t] <- tp_mci_stay * p_comm * p_surv
    
    # Transitions from "milof_c" (Mild off-treatment, community)
    if (strat == "soc") {
      tp_mil_mci <- v.p.mil_mci_soc[t]
      tp_mil_mod <- v.p.mil_mod_soc[t]
      tp_mil_sev <- v.p.mil_sev_soc[t]
      tp_mil_stay <- v.p.mil_stay_soc[t]
    } else {
      tp_mil_mci <- v.p.mil_mci_dis_int[t]
      tp_mil_mod <- v.p.mil_mod_dis_int[t]
      tp_mil_sev <- v.p.mil_sev_dis_int[t]
      tp_mil_stay <- v.p.mil_stay_dis_int[t]
    }
    p_mort <- a.TP["milof_c", "dth", t]
    p_surv <- 1 - p_mort
    p_inst <- v.p.mil_i[t]
    p_comm <- 1 - p_inst
    
    a.TP["milof_c", "mciof_c", t] <- tp_mil_mci * p_comm * p_surv
    a.TP["milof_c", "mod_c", t]   <- tp_mil_mod * p_surv
    a.TP["milof_c", "sev_c", t]   <- tp_mil_sev * p_surv
    a.TP["milof_c", "mil_i", t]   <- tp_mil_stay * p_inst * p_surv
    a.TP["milof_c", "milof_c", t] <- tp_mil_stay * p_comm * p_surv
    
    # Transitions from "mod_c" (Moderate dementia, community)
    if (strat == "soc") {
      tp_mod_mil <- v.p.mod_mil_soc[t]
      tp_mod_sev <- v.p.mod_sev_soc[t]
      tp_mod_stay <- v.p.mod_stay_soc[t]
    } else {
      tp_mod_mil <- v.p.mod_mil_soc[t]
      tp_mod_sev <- v.p.mod_sev_soc[t]
      tp_mod_stay <- v.p.mod_stay_soc[t]
    }
    p_mort <- a.TP["mod_c", "dth", t]
    p_surv <- 1 - p_mort
    p_inst <- v.p.mod_i[t]
    p_comm <- 1 - p_inst
    
    a.TP["mod_c", "milof_c", t] <- tp_mod_mil * p_comm * p_surv
    a.TP["mod_c", "sev_c", t]   <- tp_mod_sev * p_surv
    a.TP["mod_c", "mod_i", t]   <- tp_mod_stay * p_inst * p_surv
    a.TP["mod_c", "mod_c", t]   <- tp_mod_stay * p_comm * p_surv
    
    # Transitions from "sev_c" (Severe dementia, community)
    if (strat == "soc") {
      tp_sev_mil <- v.p.sev_mil_soc[t]
      tp_sev_mod <- v.p.sev_mod_soc[t]
      tp_sev_stay <- v.p.sev_stay_soc[t]
    } else {
      tp_sev_mil <- v.p.sev_mil_soc[t]
      tp_sev_mod <- v.p.sev_mod_soc[t]
      tp_sev_stay <- v.p.sev_stay_soc[t]
    }
    p_mort <- a.TP["sev_c", "dth", t]
    p_surv <- 1 - p_mort
    p_inst <- v.p.sev_i[t]
    p_comm <- 1 - p_inst
    
    a.TP["sev_c", "milof_c", t] <- tp_sev_mil * p_comm * p_surv
    a.TP["sev_c", "mod_c", t]   <- tp_sev_mod * p_comm * p_surv
    a.TP["sev_c", "sev_i", t]   <- tp_sev_stay * p_inst * p_surv
    a.TP["sev_c", "sev_c", t]   <- tp_sev_stay * p_comm * p_surv
    
    # Transitions from "mci_i" (institutionalized MCI)
    if (strat == "soc") {
      tp_mci_mil <- v.p.mci_mil_soc[t]
      tp_mci_mod <- v.p.mci_mod_soc[t]
      tp_mci_sev <- v.p.mci_sev_soc[t]
      tp_mci_stay <- v.p.mci_stay_soc[t]
    } else {
      tp_mci_mil <- v.p.mci_mil_dis_int[t]
      tp_mci_mod <- v.p.mci_mod_dis_int[t]
      tp_mci_sev <- v.p.mci_sev_dis_int[t]
      tp_mci_stay <- v.p.mci_stay_dis_int[t]
    }
    p_mort <- a.TP["mci_i", "dth", t]
    p_surv <- 1 - p_mort
    
    a.TP["mci_i", "mil_i", t] <- tp_mci_mil * p_surv
    a.TP["mci_i", "mod_i", t] <- tp_mci_mod * p_surv
    a.TP["mci_i", "sev_i", t] <- tp_mci_sev * p_surv
    a.TP["mci_i", "mci_i", t] <- tp_mci_stay * p_surv
    
    # Transitions from "mil_i" (institutionalized mild)
    if (strat == "soc") {
      tp_mil_mci <- v.p.mil_mci_soc[t]
      tp_mil_mod <- v.p.mil_mod_soc[t]
      tp_mil_sev <- v.p.mil_sev_soc[t]
      tp_mil_stay <- v.p.mil_stay_soc[t]
    } else {
      tp_mil_mci <- v.p.mil_mci_dis_int[t]
      tp_mil_mod <- v.p.mil_mod_dis_int[t]
      tp_mil_sev <- v.p.mil_sev_dis_int[t]
      tp_mil_stay <- v.p.mil_stay_dis_int[t]
    }
    p_mort <- a.TP["mil_i", "dth", t]
    p_surv <- 1 - p_mort
    
    a.TP["mil_i", "mci_i", t] <- tp_mil_mci * p_surv
    a.TP["mil_i", "mod_i", t] <- tp_mil_mod * p_surv
    a.TP["mil_i", "sev_i", t] <- tp_mil_sev * p_surv
    a.TP["mil_i", "mil_i", t] <- tp_mil_stay * p_surv
    
    # Transitions from "mod_i" (institutionalized moderate)
    tp_mod_mil <- v.p.mod_mil_soc[t]
    tp_mod_sev <- v.p.mod_sev_soc[t]
    tp_mod_stay <- v.p.mod_stay_soc[t]
    p_mort <- a.TP["mod_i", "dth", t]
    p_surv <- 1 - p_mort
    
    a.TP["mod_i", "mil_i", t] <- tp_mod_mil * p_surv
    a.TP["mod_i", "sev_i", t] <- tp_mod_sev * p_surv
    a.TP["mod_i", "mod_i", t] <- tp_mod_stay * p_surv
    
    # Transitions from "sev_i" (institutionalized severe)
    tp_sev_mil <- v.p.sev_mil_soc[t]
    tp_sev_mod <- v.p.sev_mod_soc[t]
    tp_sev_stay <- v.p.sev_stay_soc[t]
    p_mort <- a.TP["sev_i", "dth", t]
    p_surv <- 1 - p_mort
    
    a.TP["sev_i", "mil_i", t] <- tp_sev_mil * p_surv
    a.TP["sev_i", "mod_i", t] <- tp_sev_mod * p_surv
    a.TP["sev_i", "sev_i", t] <- tp_sev_stay * p_surv
    
    # ----------------------------------------------------------------------
    
    # Normalize rows to ensure row sums = 1
    for (s in state_names) {
      row_sum <- sum(a.TP[s, , t])
      if (row_sum > 0 && abs(row_sum - 1) > 1e-8) {
        a.TP[s, , t] <- a.TP[s, , t] / row_sum
      }
    }
  }
  
  return(a.TP)
}


# Compute Markov trace and total outcomes for a given strategy ("soc" or "int")
run_model <- function(strat) {
  
  # Initialize transition matrix
  a.TP <- build_transition_matrix(strat)
  
  # Initialize Markov trace matrix (cycles × states)
  m.Trace <- matrix(0, nrow = n_cycle + 1, ncol = length(state_names),
                    dimnames = list(0:n_cycle, state_names))
  
  # Initial state distribution (everyone starts in MCI)
  m.Trace[1, ] <- 0
  if (strat == "soc") {
    m.Trace[1, "mciof_c"] <- p_starting_state_mci
    m.Trace[1, "milof_c"] <- 1 - p_starting_state_mci
  } else if (strat == "int") {
    m.Trace[1, "mcion_c"] <- p_starting_state_mci
    m.Trace[1, "milon_c"] <- 1 - p_starting_state_mci
  }
  
  # Utilities vector
  v.utilities <- c(
    "mcion_c" = u_mci_pt + u_mci_ic,
    "mciof_c" = u_mci_pt + u_mci_ic,
    "milon_c" = u_mil_pt + u_mil_ic,
    "milof_c" = u_mil_pt + u_mil_ic,
    "mod_c"   = u_mod_pt + u_mod_ic,
    "sev_c"   = u_sev_pt + u_sev_ic,
    "mci_i"   = u_mci_pt_i + u_mci_ic_i,
    "mil_i"   = u_mil_pt_i + u_mil_ic_i,
    "mod_i"   = u_mod_pt_i + u_mod_ic_i,
    "sev_i"   = u_sev_pt_i + u_sev_ic_i,
    "dth"     = 0
  )
  
  # Costs vector
  v.costs <- c(
    "mcion_c" = c_mci_hc + c_mci_sc + c_mci_ic,
    "mciof_c" = c_mci_hc + c_mci_sc + c_mci_ic,
    "milon_c" = c_mil_hc + c_mil_sc + c_mil_ic,
    "milof_c" = c_mil_hc + c_mil_sc + c_mil_ic,
    "mod_c"   = c_mod_hc + c_mod_sc + c_mod_ic,
    "sev_c"   = c_sev_hc + c_sev_sc + c_sev_ic,
    "mci_i"   = c_mci_i_hc + c_mci_i_sc + c_mci_i_ic,
    "mil_i"   = c_mil_i_hc + c_mil_i_sc + c_mil_i_ic,
    "mod_i"   = c_mod_i_hc + c_mod_i_sc + c_mod_i_ic,
    "sev_i"   = c_sev_i_hc + c_sev_i_sc + c_sev_i_ic,
    "dth"     = 0
  )
  
  # Track outcomes per cycle
  v.QALYs <- numeric(n_cycle + 1)
  v.Costs <- numeric(n_cycle + 1)
  v.LYs   <- numeric(n_cycle + 1)
  
  for (t in 1:n_cycle) {
    # Update state vector
    m.Trace[t + 1, ] <- m.Trace[t, ] %*% a.TP[, , t]
    
    # QALYs and costs
    v.QALYs[t + 1] <- sum(m.Trace[t + 1, ] * v.utilities)
    v.Costs[t + 1] <- sum(m.Trace[t + 1, ] * v.costs)
    v.LYs[t + 1]   <- sum(m.Trace[t + 1, state_names[state_names != "dth"]])
    
    # Add recurring treatment cost for "int"
    if (strat == "int") {
      v.Costs[t + 1] <- v.Costs[t + 1] + c_Tx * sum(m.Trace[t + 1, c("mcion_c", "milon_c")])
    }
  }
  
  # Add initial cycle outcomes
  v.QALYs[1] <- sum(m.Trace[1, ] * v.utilities)
  v.Costs[1] <- sum(m.Trace[1, ] * v.costs)
  v.LYs[1]   <- sum(m.Trace[1, state_names[state_names != "dth"]])
  
  # Add initial treatment cost and utility loss for ARIA (only in cycle 1)
  if (strat == "int") {
    v.Costs[1] <- v.Costs[1] + c_Tx_start
    v.QALYs[1] <- v.QALYs[1] + u_Tx_start
  }
  
  # Apply half-cycle correction
  v.QALYs <- (v.QALYs[-1] + v.QALYs[-length(v.QALYs)]) / 2
  v.Costs <- (v.Costs[-1] + v.Costs[-length(v.Costs)]) / 2
  v.LYs   <- (v.LYs[-1] + v.LYs[-length(v.LYs)]) / 2
  
  # Apply discounting
  discount_factors <- 1 / ((1 + discount_QALY)^(0:(n_cycle - 1)))
  QALYs_total <- sum(v.QALYs * discount_factors)
  LYs_total   <- sum(v.LYs * discount_factors)
  Costs_total <- sum(v.Costs * (1 / ((1 + discount_COST)^(0:(n_cycle - 1)))))
  
  return(list(QALYs = QALYs_total, Costs = Costs_total, LYs = LYs_total))
}

# Run both strategies
results_soc <- run_model("soc")
results_int <- run_model("int")

# Print results
cat("===== IPECAD Model Results =====\n")
cat("Strategy: Standard of Care\n")
cat(sprintf("  QALYs: %.2f\n  LYs: %.2f\n  Costs: $%0.2f\n\n",
            results_soc$QALYs, results_soc$LYs, results_soc$Costs))
cat("Strategy: Intervention\n")
cat(sprintf("  QALYs: %.2f\n  LYs: %.2f\n  Costs: $%0.2f\n\n",
            results_int$QALYs, results_int$LYs, results_int$Costs))

# Incremental results
cat("===== Incremental (Intervention - SOC) =====\n")
cat(sprintf("  ΔQALYs: %.2f\n  ΔLYs: %.2f\n  ΔCosts: $%0.2f\n",
            results_int$QALYs - results_soc$QALYs,
            results_int$LYs - results_soc$LYs,
            results_int$Costs - results_soc$Costs))
```
