## Supplementary material for "Replicating Health-Economic Simulation Models for Alzheimer’s Disease Using Artificial Intelligence": final R scripts: o6.R

```
# Required packages
library(dampack)

# Define strategy and state names
state_names <- c("mcion_c", "mciof_c", "milon_c", "milof_c", "mod_c", "sev_c",
                 "mci_i", "mil_i", "mod_i", "sev_i", "dth")
strategy_names <- c("soc", "int")

# Starting population characteristics
age_start <- 71
sex <- "weighted"  # 52% female
p_starting_state_mci <- 0.55

# Model settings
n_cycles <- 29
discount_EFFECT <- 0.03
discount_QALY <- 0.03
discount_COST <- 0.03
wtp <- 100000
half_cycle_correction <- TRUE

# Disease progression probabilities (per year)
p_mci_mil <- 0.23
p_mci_mod <- 0
p_mci_sev <- 0

p_mil_mci <- 0.03
p_mil_mod <- 0.35
p_mil_sev <- 0.04

p_mod_mil <- 0.03
p_mod_sev <- 0.42

p_sev_mil <- 0
p_sev_mod <- 0.02

# Institutionalization probabilities (per year)
p_mci_i <- 0.024
p_mil_i <- 0.038
p_mod_i <- 0.110
p_sev_i <- 0.259

# Mortality multipliers
hr_mort_mci <- 1.82
hr_mort_mil <- 2.92
hr_mort_mod <- 3.85
hr_mort_sev <- 9.52

# Treatment effect (relative risk per transition)
rr_tx_mci_mil <- 0.69
rr_tx_mci_mod <- 1
rr_tx_mci_sev <- 1
rr_tx_mil_mod <- 0.69
rr_tx_mil_sev <- 0.69

# Treatment effect after discontinuation (no effect)
rr_tx_mci_mil_dis <- 1
rr_tx_mci_mod_dis <- 1
rr_tx_mci_sev_dis <- 1
rr_tx_mil_mod_dis <- 1
rr_tx_mil_sev_dis <- 1

# Discontinuation
p_discontinuation1 <- 0.069
p_discontinuation2 <- 0
discontinuation2_begin <- 2
tx_duration <- 29
tx_waning <- 0
tx_waning_dis <- 0

# Utilities (patient)
u_mci_pt <- 0.851 - 0.17
u_mil_pt <- 0.851 - 0.22
u_mod_pt <- 0.851 - 0.36
u_sev_pt <- 0.851 - 0.53
u_mci_pt_i <- 0.851 - 0.17
u_mil_pt_i <- 0.851 - 0.19
u_mod_pt_i <- 0.851 - 0.42
u_sev_pt_i <- 0.851 - 0.59

# Utilities (informal caregiver)
u_mci_ic <- -0.03
u_mil_ic <- -0.05
u_mod_ic <- -0.08
u_sev_ic <- -0.10
u_mci_ic_i <- -0.03
u_mil_ic_i <- -0.05
u_mod_ic_i <- -0.08
u_sev_ic_i <- -0.10

# Treatment disutility in first cycle (not half-cycle corrected)
u_Tx_start <- -0.14 * (12/52) * 0.035

# Costs (health care - community)
c_mci_hc <- 6042 * 1.12 + 460
c_mil_hc <- 6042 * 1.56 + 965 + 0.21 * 365 * 0.333
c_mod_hc <- 6042 * 1.93 + 1544 + 0.66 * 365 * 0.333
c_sev_hc <- 6042 * 1.93 + 1930

# Costs (health care - institution)
c_mci_i_hc <- c_mci_hc
c_mil_i_hc <- c_mil_hc
c_mod_i_hc <- c_mod_hc
c_sev_i_hc <- c_sev_hc

# Costs (social care - community)
c_mci_sc <- 0
c_mil_sc <- 0
c_mod_sc <- 0
c_sev_sc <- 0

# Costs (social care - institution, monthly * 12)
c_mci_i_sc <- 7394 * 12
c_mil_i_sc <- 7394 * 12
c_mod_i_sc <- 7394 * 12
c_sev_i_sc <- 7394 * 12

# Costs (informal care - community)
c_mci_ic <- 69 * 12 * 32.46 + 0.204 * 0.049 * 20 * 52 * 32.46
c_mil_ic <- 113 * 12 * 32.46 + 0.112 * 0.086 * 20 * 52 * 32.46
c_mod_ic <- 169 * 12 * 32.46
c_sev_ic <- 298 * 12 * 32.46

# Costs (informal care - institution, 44% of community + same productivity loss)
c_mci_i_ic <- 69 * 12 * 32.46 * 0.44 + 0.204 * 0.049 * 20 * 52 * 32.46
c_mil_i_ic <- 113 * 12 * 32.46 * 0.44 + 0.112 * 0.086 * 20 * 52 * 32.46
c_mod_i_ic <- 169 * 12 * 32.46 * 0.44
c_sev_i_ic <- 298 * 12 * 32.46 * 0.44

# Treatment cost (annual + biweekly admin)
c_Tx <- 26500 + (52 / 2) * 78.35

# Additional cost in first cycle (monitoring MRI)
c_Tx_start <- 261.10 * 4 + 261.10 * 3 * 0.215
# Load the 2019 US life table CSV file
lifetable <- read.csv("lifetable_US_2019_ssa.csv", stringsAsFactors = FALSE)

# Convert annual mortality probabilities to annual mortality rates
# Rate = -log(1 - probability)
lifetable$male <- -log(1 - lifetable$male)
lifetable$female <- -log(1 - lifetable$female)

# Compute the weighted average of male and female mortality rates
# Using 52% female and 48% male as per earlier input
lifetable$weighted <- 0.48 * lifetable$male + 0.52 * lifetable$female

# Return only the relevant columns
lifetable <- lifetable[, c("age", "male", "female", "weighted")]

# Output the processed life table
lifetable
# Define time vector
v.cycles <- 0:(n_cycles - 1)

# Define age per cycle
v.age <- age_start + v.cycles

# Disease progression probabilities under "soc" strategy (time-independent, repeated for each cycle)
v.p.mci_mil_soc <- rep(p_mci_mil, n_cycles)
v.p.mci_mod_soc <- rep(p_mci_mod, n_cycles)
v.p.mci_sev_soc <- rep(p_mci_sev, n_cycles)

v.p.mil_mci_soc <- rep(p_mil_mci, n_cycles)
v.p.mil_mod_soc <- rep(p_mil_mod, n_cycles)
v.p.mil_sev_soc <- rep(p_mil_sev, n_cycles)

v.p.mod_mil_soc <- rep(p_mod_mil, n_cycles)
v.p.mod_sev_soc <- rep(p_mod_sev, n_cycles)

v.p.sev_mil_soc <- rep(p_sev_mil, n_cycles)
v.p.sev_mod_soc <- rep(p_sev_mod, n_cycles)

# Staying probabilities (excluding institutional transitions and mortality)
v.p.mci_stay_soc <- 1 - v.p.mci_mil_soc - v.p.mci_mod_soc - v.p.mci_sev_soc
v.p.mil_stay_soc <- 1 - v.p.mil_mci_soc - v.p.mil_mod_soc - v.p.mil_sev_soc
v.p.mod_stay_soc <- 1 - v.p.mod_mil_soc - v.p.mod_sev_soc
v.p.sev_stay_soc <- 1 - v.p.sev_mil_soc - v.p.sev_mod_soc

# Transition to institution probabilities (time-independent)
v.p.mci_i <- rep(p_mci_i, n_cycles)
v.p.mil_i <- rep(p_mil_i, n_cycles)
v.p.mod_i <- rep(p_mod_i, n_cycles)
v.p.sev_i <- rep(p_sev_i, n_cycles)

# Discontinuation probability vector (no cumulative calculation)
v.p_discontinuation <- ifelse(v.cycles == 0, 
                              p_discontinuation1,
                              ifelse(v.cycles < tx_duration, 
                                     p_discontinuation2, 
                                     1))

# Load mortality rates for corresponding ages from life table
# Assuming 'lifetable' is already loaded and formatted from Prompt 2
v.mortality_rate <- lifetable$weighted[match(v.age, lifetable$age)]
# Create cycle index (1-based)
v.cycle_index <- 1:n_cycles

# Treatment waning factors per cycle (on-treatment and off-treatment)
v.waning_factor <- (1 - tx_waning)^(v.cycle_index - 1)
v.waning_factor_dis <- (1 - tx_waning_dis)^(v.cycle_index - 1)

# Helper function to apply RR-adjusted probability transformation
apply_rr <- function(p, rr, waning) {
  rate <- -log(1 - p)
  adjusted_rate <- rate * rr^waning
  prob <- 1 - exp(-adjusted_rate)
  return(prob)
}

### On-treatment: mcion_c ###
v.p.mci_mil_int_on <- apply_rr(p_mci_mil, rr_tx_mci_mil, v.waning_factor)
v.p.mci_mod_int_on <- apply_rr(p_mci_mod, rr_tx_mci_mod, v.waning_factor)
v.p.mci_sev_int_on <- apply_rr(p_mci_sev, rr_tx_mci_sev, v.waning_factor)
v.p.mci_stay_int_on <- 1 - v.p.mci_mil_int_on - v.p.mci_mod_int_on - v.p.mci_sev_int_on

### On-treatment: milon_c ###
v.p.mil_mci_int_on <- rep(p_mil_mci, n_cycles)  # backward transition, no treatment effect
v.p.mil_mod_int_on <- apply_rr(p_mil_mod, rr_tx_mil_mod, v.waning_factor)
v.p.mil_sev_int_on <- apply_rr(p_mil_sev, rr_tx_mil_sev, v.waning_factor)
v.p.mil_stay_int_on <- 1 - v.p.mil_mci_int_on - v.p.mil_mod_int_on - v.p.mil_sev_int_on

### Off-treatment (discontinued): mciof_c ###
v.p.mci_mil_int_off <- apply_rr(p_mci_mil, rr_tx_mci_mil_dis, v.waning_factor_dis)
v.p.mci_mod_int_off <- apply_rr(p_mci_mod, rr_tx_mci_mod_dis, v.waning_factor_dis)
v.p.mci_sev_int_off <- apply_rr(p_mci_sev, rr_tx_mci_sev_dis, v.waning_factor_dis)
v.p.mci_stay_int_off <- 1 - v.p.mci_mil_int_off - v.p.mci_mod_int_off - v.p.mci_sev_int_off

### Off-treatment: milof_c ###
v.p.mil_mci_int_off <- rep(p_mil_mci, n_cycles)  # backward transition, no treatment effect
v.p.mil_mod_int_off <- apply_rr(p_mil_mod, rr_tx_mil_mod_dis, v.waning_factor_dis)
v.p.mil_sev_int_off <- apply_rr(p_mil_sev, rr_tx_mil_sev_dis, v.waning_factor_dis)
v.p.mil_stay_int_off <- 1 - v.p.mil_mci_int_off - v.p.mil_mod_int_off - v.p.mil_sev_int_off
# Function to build transition probability array for a given strategy
build_transition_matrix <- function(strat) {
  
  # Initialize transition probability array: [from state, to state, cycle]
  a.TP <- array(0, dim = c(length(state_names), length(state_names), n_cycles),
                dimnames = list(state_names, state_names, NULL))
  
  # Loop over each cycle to fill in death transitions
  for (t in 1:n_cycles) {
    
    # Extract age-specific mortality rate
    mort_rate <- v.mortality_rate[t]
    
    # Transitions to death from each non-death state
    a.TP["mcion_c", "dth", t] <- 1 - exp(-mort_rate * hr_mort_mci)
    a.TP["mciof_c", "dth", t] <- 1 - exp(-mort_rate * hr_mort_mci)
    a.TP["milon_c", "dth", t] <- 1 - exp(-mort_rate * hr_mort_mil)
    a.TP["milof_c", "dth", t] <- 1 - exp(-mort_rate * hr_mort_mil)
    a.TP["mod_c",   "dth", t] <- 1 - exp(-mort_rate * hr_mort_mod)
    a.TP["sev_c",   "dth", t] <- 1 - exp(-mort_rate * hr_mort_sev)
    a.TP["mci_i",   "dth", t] <- 1 - exp(-mort_rate * hr_mort_mci)
    a.TP["mil_i",   "dth", t] <- 1 - exp(-mort_rate * hr_mort_mil)
    a.TP["mod_i",   "dth", t] <- 1 - exp(-mort_rate * hr_mort_mod)
    a.TP["sev_i",   "dth", t] <- 1 - exp(-mort_rate * hr_mort_sev)
    
    # Death is an absorbing state
    a.TP["dth", "dth", t] <- 1
    
    # Transitions from "mcion_c" (on-treatment MCI community)
    if (strat == "soc") {
      p_mci_mil <- v.p.mci_mil_soc[t]
      p_mci_mod <- v.p.mci_mod_soc[t]
      p_mci_sev <- v.p.mci_sev_soc[t]
      p_mci_stay <- v.p.mci_stay_soc[t]
    } else {
      p_mci_mil <- v.p.mci_mil_int_on[t]
      p_mci_mod <- v.p.mci_mod_int_on[t]
      p_mci_sev <- v.p.mci_sev_int_on[t]
      p_mci_stay <- v.p.mci_stay_int_on[t]
    }
    
    surv_prob <- 1 - a.TP["mcion_c", "dth", t]
    p_inst <- v.p.mci_i[t]
    p_disc <- v.p_discontinuation[t]
    
    a.TP["mcion_c", "mcion_c", t] <- p_mci_stay * surv_prob * (1 - p_inst) * (1 - p_disc)
    a.TP["mcion_c", "mciof_c", t] <- p_mci_stay * surv_prob * (1 - p_inst) * p_disc
    a.TP["mcion_c", "milon_c", t] <- p_mci_mil * surv_prob * (1 - p_inst) * (1 - p_disc)
    a.TP["mcion_c", "mciof_c",  t] <- p_mci_mil * surv_prob * (1 - p_inst) * p_disc  # no such state, remove
    a.TP["mcion_c", "mod_c",   t] <- p_mci_mod * surv_prob * (1 - p_inst)
    a.TP["mcion_c", "sev_c",   t] <- p_mci_sev * surv_prob * (1 - p_inst)
    a.TP["mcion_c", "mci_i",   t] <- (p_mci_stay + p_mci_mil + p_mci_mod + p_mci_sev) * surv_prob * p_inst
    
    # Transitions from "milon_c" (on-treatment mild community)
    if (strat == "soc") {
      p_mil_mci <- v.p.mil_mci_soc[t]
      p_mil_mod <- v.p.mil_mod_soc[t]
      p_mil_sev <- v.p.mil_sev_soc[t]
      p_mil_stay <- v.p.mil_stay_soc[t]
    } else {
      p_mil_mci <- v.p.mil_mci_int_on[t]
      p_mil_mod <- v.p.mil_mod_int_on[t]
      p_mil_sev <- v.p.mil_sev_int_on[t]
      p_mil_stay <- v.p.mil_stay_int_on[t]
    }
    
    surv_prob <- 1 - a.TP["milon_c", "dth", t]
    p_inst <- v.p.mil_i[t]
    p_disc <- v.p_discontinuation[t]
    
    a.TP["milon_c", "milon_c", t] <- p_mil_stay * surv_prob * (1 - p_inst) * (1 - p_disc)
    a.TP["milon_c", "milof_c", t] <- p_mil_stay * surv_prob * (1 - p_inst) * p_disc
    a.TP["milon_c", "mciof_c", t] <- p_mil_mci * surv_prob * (1 - p_inst)  # backward, always off-treatment
    a.TP["milon_c", "mod_c",   t] <- p_mil_mod * surv_prob * (1 - p_inst)
    a.TP["milon_c", "sev_c",   t] <- p_mil_sev * surv_prob * (1 - p_inst)
    a.TP["milon_c", "mil_i",   t] <- (p_mil_stay + p_mil_mci + p_mil_mod + p_mil_sev) * surv_prob * p_inst
    
    # Transitions from "mciof_c"
    if (strat == "soc") {
      p_mci_mil <- v.p.mci_mil_soc[t]
      p_mci_mod <- v.p.mci_mod_soc[t]
      p_mci_sev <- v.p.mci_sev_soc[t]
      p_mci_stay <- v.p.mci_stay_soc[t]
    } else {
      p_mci_mil <- v.p.mci_mil_int_off[t]
      p_mci_mod <- v.p.mci_mod_int_off[t]
      p_mci_sev <- v.p.mci_sev_int_off[t]
      p_mci_stay <- v.p.mci_stay_int_off[t]
    }
    
    surv_prob <- 1 - a.TP["mciof_c", "dth", t]
    p_inst <- v.p.mci_i[t]
    
    a.TP["mciof_c", "mciof_c", t] <- p_mci_stay * surv_prob * (1 - p_inst)
    a.TP["mciof_c", "milof_c", t] <- p_mci_mil * surv_prob * (1 - p_inst)
    a.TP["mciof_c", "mod_c",   t] <- p_mci_mod * surv_prob * (1 - p_inst)
    a.TP["mciof_c", "sev_c",   t] <- p_mci_sev * surv_prob * (1 - p_inst)
    a.TP["mciof_c", "mci_i",   t] <- (p_mci_stay + p_mci_mil + p_mci_mod + p_mci_sev) * surv_prob * p_inst
    
    # Transitions from "milof_c"
    if (strat == "soc") {
      p_mil_mci <- v.p.mil_mci_soc[t]
      p_mil_mod <- v.p.mil_mod_soc[t]
      p_mil_sev <- v.p.mil_sev_soc[t]
      p_mil_stay <- v.p.mil_stay_soc[t]
    } else {
      p_mil_mci <- v.p.mil_mci_int_off[t]
      p_mil_mod <- v.p.mil_mod_int_off[t]
      p_mil_sev <- v.p.mil_sev_int_off[t]
      p_mil_stay <- v.p.mil_stay_int_off[t]
    }
    
    surv_prob <- 1 - a.TP["milof_c", "dth", t]
    p_inst <- v.p.mil_i[t]
    
    a.TP["milof_c", "milof_c", t] <- p_mil_stay * surv_prob * (1 - p_inst)
    a.TP["milof_c", "mciof_c", t] <- p_mil_mci * surv_prob * (1 - p_inst)
    a.TP["milof_c", "mod_c",   t] <- p_mil_mod * surv_prob * (1 - p_inst)
    a.TP["milof_c", "sev_c",   t] <- p_mil_sev * surv_prob * (1 - p_inst)
    a.TP["milof_c", "mil_i",   t] <- (p_mil_stay + p_mil_mci + p_mil_mod + p_mil_sev) * surv_prob * p_inst
    
    # Transitions from "mod_c"
    if (strat == "soc") {
      p_mod_mil <- v.p.mod_mil_soc[t]
      p_mod_sev <- v.p.mod_sev_soc[t]
      p_mod_stay <- v.p.mod_stay_soc[t]
    } else {
      p_mod_mil <- v.p.mod_mil_soc[t]  # same for int and soc
      p_mod_sev <- v.p.mod_sev_soc[t]
      p_mod_stay <- v.p.mod_stay_soc[t]
    }
    
    surv_prob <- 1 - a.TP["mod_c", "dth", t]
    p_inst <- v.p.mod_i[t]
    
    a.TP["mod_c", "mod_c", t] <- p_mod_stay * surv_prob * (1 - p_inst)
    a.TP["mod_c", "milof_c", t] <- p_mod_mil * surv_prob * (1 - p_inst)
    a.TP["mod_c", "sev_c",   t] <- p_mod_sev * surv_prob * (1 - p_inst)
    a.TP["mod_c", "mod_i",   t] <- (p_mod_stay + p_mod_mil + p_mod_sev) * surv_prob * p_inst
    
    # Transitions from "sev_c"
    if (strat == "soc") {
      p_sev_mil <- v.p.sev_mil_soc[t]
      p_sev_mod <- v.p.sev_mod_soc[t]
      p_sev_stay <- v.p.sev_stay_soc[t]
    } else {
      p_sev_mil <- v.p.sev_mil_soc[t]
      p_sev_mod <- v.p.sev_mod_soc[t]
      p_sev_stay <- v.p.sev_stay_soc[t]
    }
    
    surv_prob <- 1 - a.TP["sev_c", "dth", t]
    p_inst <- v.p.sev_i[t]
    
    a.TP["sev_c", "sev_c", t] <- p_sev_stay * surv_prob * (1 - p_inst)
    a.TP["sev_c", "milof_c", t] <- p_sev_mil * surv_prob * (1 - p_inst)
    a.TP["sev_c", "mod_c",   t] <- p_sev_mod * surv_prob * (1 - p_inst)
    a.TP["sev_c", "sev_i",   t] <- (p_sev_stay + p_sev_mil + p_sev_mod) * surv_prob * p_inst
    
    # Transitions from "mci_i"
    if (strat == "soc") {
      p_mci_mil <- v.p.mci_mil_soc[t]
      p_mci_mod <- v.p.mci_mod_soc[t]
      p_mci_sev <- v.p.mci_sev_soc[t]
      p_mci_stay <- v.p.mci_stay_soc[t]
    } else {
      p_mci_mil <- v.p.mci_mil_int_off[t]
      p_mci_mod <- v.p.mci_mod_int_off[t]
      p_mci_sev <- v.p.mci_sev_int_off[t]
      p_mci_stay <- v.p.mci_stay_int_off[t]
    }
    
    surv_prob <- 1 - a.TP["mci_i", "dth", t]
    
    a.TP["mci_i", "mci_i", t] <- p_mci_stay * surv_prob
    a.TP["mci_i", "mil_i", t] <- p_mci_mil * surv_prob
    a.TP["mci_i", "mod_i", t] <- p_mci_mod * surv_prob
    a.TP["mci_i", "sev_i", t] <- p_mci_sev * surv_prob
    
    # Transitions from "mil_i"
    if (strat == "soc") {
      p_mil_mci <- v.p.mil_mci_soc[t]
      p_mil_mod <- v.p.mil_mod_soc[t]
      p_mil_sev <- v.p.mil_sev_soc[t]
      p_mil_stay <- v.p.mil_stay_soc[t]
    } else {
      p_mil_mci <- v.p.mil_mci_int_off[t]
      p_mil_mod <- v.p.mil_mod_int_off[t]
      p_mil_sev <- v.p.mil_sev_int_off[t]
      p_mil_stay <- v.p.mil_stay_int_off[t]
    }
    
    surv_prob <- 1 - a.TP["mil_i", "dth", t]
    
    a.TP["mil_i", "mil_i", t] <- p_mil_stay * surv_prob
    a.TP["mil_i", "mci_i", t] <- p_mil_mci * surv_prob
    a.TP["mil_i", "mod_i", t] <- p_mil_mod * surv_prob
    a.TP["mil_i", "sev_i", t] <- p_mil_sev * surv_prob
    
    # Transitions from "mod_i"
    p_mod_mil <- v.p.mod_mil_soc[t]
    p_mod_sev <- v.p.mod_sev_soc[t]
    p_mod_stay <- v.p.mod_stay_soc[t]
    surv_prob <- 1 - a.TP["mod_i", "dth", t]
    
    a.TP["mod_i", "mod_i", t] <- p_mod_stay * surv_prob
    a.TP["mod_i", "mil_i", t] <- p_mod_mil * surv_prob
    a.TP["mod_i", "sev_i", t] <- p_mod_sev * surv_prob
    
    # Transitions from "sev_i"
    p_sev_mil <- v.p.sev_mil_soc[t]
    p_sev_mod <- v.p.sev_mod_soc[t]
    p_sev_stay <- v.p.sev_stay_soc[t]
    surv_prob <- 1 - a.TP["sev_i", "dth", t]
    
    a.TP["sev_i", "sev_i", t] <- p_sev_stay * surv_prob
    a.TP["sev_i", "mil_i", t] <- p_sev_mil * surv_prob
    a.TP["sev_i", "mod_i", t] <- p_sev_mod * surv_prob
    
    # Normalize each row to ensure total probability equals 1
    for (s in state_names) {
      row_sum <- sum(a.TP[s, , t])
      if (row_sum > 0 && row_sum != 1) {
        a.TP[s, , t] <- a.TP[s, , t] / row_sum
      }
    }
  }
  
  return(a.TP)
}
# Function to compute Markov trace and outcomes
compute_outcomes <- function(strat) {
  # Build transition matrix
  a.TP <- build_transition_matrix(strat)
  
  # Initialize trace matrix: rows = cycles, cols = states
  m.trace <- matrix(0, nrow = n_cycles, ncol = length(state_names),
                    dimnames = list(0:(n_cycles - 1), state_names))
  
  # Initial state distribution
  v.init <- rep(0, length(state_names))
  names(v.init) <- state_names
  if (strat == "soc") {
    v.init["mciof_c"] <- p_starting_state_mci
    v.init["milof_c"] <- 1 - p_starting_state_mci
  } else if (strat == "int") {
    v.init["mcion_c"] <- p_starting_state_mci
    v.init["milon_c"] <- 1 - p_starting_state_mci
  }
  m.trace[1, ] <- v.init
  
  # Named vectors for utilities (patients + informal caregivers)
  v.u <- c(
    mcion_c = u_mci_pt + u_mci_ic,
    mciof_c = u_mci_pt + u_mci_ic,
    milon_c = u_mil_pt + u_mil_ic,
    milof_c = u_mil_pt + u_mil_ic,
    mod_c   = u_mod_pt + u_mod_ic,
    sev_c   = u_sev_pt + u_sev_ic,
    mci_i   = u_mci_pt_i + u_mci_ic_i,
    mil_i   = u_mil_pt_i + u_mil_ic_i,
    mod_i   = u_mod_pt_i + u_mod_ic_i,
    sev_i   = u_sev_pt_i + u_sev_ic_i,
    dth     = 0
  )
  
  # Named vectors for costs (health + social + informal care)
  v.c <- c(
    mcion_c = c_mci_hc + c_mci_sc + c_mci_ic,
    mciof_c = c_mci_hc + c_mci_sc + c_mci_ic,
    milon_c = c_mil_hc + c_mil_sc + c_mil_ic,
    milof_c = c_mil_hc + c_mil_sc + c_mil_ic,
    mod_c   = c_mod_hc + c_mod_sc + c_mod_ic,
    sev_c   = c_sev_hc + c_sev_sc + c_sev_ic,
    mci_i   = c_mci_i_hc + c_mci_i_sc + c_mci_i_ic,
    mil_i   = c_mil_i_hc + c_mil_i_sc + c_mil_i_ic,
    mod_i   = c_mod_i_hc + c_mod_i_sc + c_mod_i_ic,
    sev_i   = c_sev_i_hc + c_sev_i_sc + c_sev_i_ic,
    dth     = 0
  )
  
  # Storage for outcomes
  v.ly <- numeric(n_cycles)
  v.qaly <- numeric(n_cycles)
  v.cost <- numeric(n_cycles)
  
  # Markov trace iteration and outcomes calculation
  for (t in 2:n_cycles) {
    m.trace[t, ] <- m.trace[t - 1, ] %*% a.TP[ , , t - 1]
  }
  
  for (t in 1:n_cycles) {
    v.ly[t] <- sum(m.trace[t, state_names != "dth"])
    v.qaly[t] <- sum(m.trace[t, ] * v.u)
    v.cost[t] <- sum(m.trace[t, ] * v.c)
    
    if (strat == "int") {
      v.cost[t] <- v.cost[t] +
        sum(m.trace[t, c("mcion_c", "milon_c")]) * c_Tx  # Recurring treatment cost
    }
    
    if (strat == "int" && t == 1) {
      v.cost[t] <- v.cost[t] + c_Tx_start
      v.qaly[t] <- v.qaly[t] + u_Tx_start
    }
  }
  
  # Half-cycle correction (exclude last cycle)
  v.ly     <- (v.ly[-n_cycles] + v.ly[-1]) / 2
  v.qaly   <- (v.qaly[-n_cycles] + v.qaly[-1]) / 2
  v.cost   <- (v.cost[-n_cycles] + v.cost[-1]) / 2
  
  # Apply discounting
  v.discount <- 1 / (1 + discount_QALY)^(1:(n_cycles - 1))
  total_ly   <- sum(v.ly * v.discount)
  total_qaly <- sum(v.qaly * v.discount)
  total_cost <- sum(v.cost * (1 / (1 + discount_COST)^(1:(n_cycles - 1))))
  
  return(list(total_ly = total_ly, total_qaly = total_qaly, total_cost = total_cost))
}

# Run model for both strategies
res_soc <- compute_outcomes("soc")
res_int <- compute_outcomes("int")

# Print comparative results
cat("===== Comparative Results =====\n")
cat(sprintf("Total Life Years (SOC): %.2f\n", res_soc$total_ly))
cat(sprintf("Total Life Years (INT): %.2f\n", res_int$total_ly))
cat(sprintf("Total QALYs (SOC): %.2f\n", res_soc$total_qaly))
cat(sprintf("Total QALYs (INT): %.2f\n", res_int$total_qaly))
cat(sprintf("Total Costs (SOC): $%.0f\n", res_soc$total_cost))
cat(sprintf("Total Costs (INT): $%.0f\n", res_int$total_cost))
cat("================================\n")
```
