## Supplementary material for "Replicating Health-Economic Simulation Models for Alzheimer’s Disease Using Artificial Intelligence": final R scripts: o7.R

```
# Load required package
library(dampack)

# Strategy and state names
v.names_strat <- c("soc", "int")
v.names_state <- c("mcion_c", "mciof_c", "milon_c", "milof_c", "mod_c", "sev_c",
                   "mci_i", "mil_i", "mod_i", "sev_i", "dth")

# Population characteristics
age_start <- 71
sex <- "weighted"  # 52% female
p.starting_state_mci <- 0.55
n.cycle <- 29

# Transition probabilities (soc strategy)
p.mci_mil <- 0.23
p.mci_mod <- 0
p.mci_sev <- 0
p.mil_mci <- 0.03
p.mil_mod <- 0.35
p.mil_sev <- 0.04
p.mod_mil <- 0.03
p.mod_sev <- 0.42
p.sev_mil <- 0
p.sev_mod <- 0.02

# Institution transitions
p.mci_i <- 0.024
p.mil_i <- 0.038
p.mod_i <- 0.110
p.sev_i <- 0.259

# Mortality (general population + hazard ratios)
m.r.mortality <- "from_life_table_csv"  # To be loaded later
hr.mort_mci <- 1.82
hr.mort_mil <- 2.92
hr.mort_mod <- 3.85
hr.mort_sev <- 9.52

# Treatment effect (relative risks)
rr.tx_mci_mil <- 0.69
rr.tx_mci_mod <- 1
rr.tx_mci_sev <- 1
rr.tx_mil_mod <- 0.69
rr.tx_mil_sev <- 0.69
rr.tx_mci_mil_dis <- 1
rr.tx_mci_mod_dis <- 1
rr.tx_mci_sev_dis <- 1
rr.tx_mil_mod_dis <- 1
rr.tx_mil_sev_dis <- 1

# Discontinuation
p.discontinuation1 <- 0.069
p.discontinuation2 <- 0
discontinuation2_begin <- 2
tx_duration <- 29
tx_waning <- 0
tx_waning_dis <- 0

# Utilities (patients, community)
u.mci_pt <- 0.851 - 0.17
u.mil_pt <- 0.851 - 0.22
u.mod_pt <- 0.851 - 0.36
u.sev_pt <- 0.851 - 0.53

# Utilities (patients, institution)
u.mci_pt_i <- 0.851 - 0.17
u.mil_pt_i <- 0.851 - 0.19
u.mod_pt_i <- 0.851 - 0.42
u.sev_pt_i <- 0.851 - 0.59

# Utilities (informal caregiver, community)
u.mci_ic <- -0.03
u.mil_ic <- -0.05
u.mod_ic <- -0.08
u.sev_ic <- -0.10

# Utilities (informal caregiver, institution)
u.mci_ic_i <- -0.03
u.mil_ic_i <- -0.05
u.mod_ic_i <- -0.08
u.sev_ic_i <- -0.10

# ARIA disutility in first cycle
u.Tx_start <- -0.14 * (12 / 52) * 0.035

# Costs (healthcare, patient, community)
c.mci_hc <- 6042 * 1.12 + 460
c.mil_hc <- 6042 * 1.56 + 965 + 0.21 * 365 * 0.333
c.mod_hc <- 6042 * 1.93 + 1544 + 0.66 * 365 * 0.333
c.sev_hc <- 6042 * 1.93 + 1930

# Costs (healthcare, patient, institution)
c.mci_i_hc <- c.mci_hc
c.mil_i_hc <- c.mil_hc
c.mod_i_hc <- c.mod_hc
c.sev_i_hc <- c.sev_hc

# Costs (social care, community)
c.mci_sc <- 0
c.mil_sc <- 0
c.mod_sc <- 0
c.sev_sc <- 0

# Costs (social care, institution)
c.mci_i_sc <- 7394 * 12
c.mil_i_sc <- 7394 * 12
c.mod_i_sc <- 7394 * 12
c.sev_i_sc <- 7394 * 12

# Costs (informal care, community)
c.mci_ic <- 69 * 12 * 32.46 + 0.204 * 0.049 * 20 * 52 * 32.46
c.mil_ic <- 113 * 12 * 32.46 + 0.112 * 0.086 * 20 * 52 * 32.46
c.mod_ic <- 169 * 12 * 32.46
c.sev_ic <- 298 * 12 * 32.46

# Costs (informal care, institution)
c.mci_i_ic <- 69 * 12 * 32.46 * 0.44 + 0.204 * 0.049 * 20 * 52 * 32.46
c.mil_i_ic <- 113 * 12 * 32.46 * 0.44 + 0.112 * 0.086 * 20 * 52 * 32.46
c.mod_i_ic <- 169 * 12 * 32.46 * 0.44
c.sev_i_ic <- 298 * 12 * 32.46 * 0.44

# Treatment costs
c.Tx <- 26500 + (52 / 2) * 78.35
c.Tx_start <- 261.10 * 4 + 261.10 * 3 * 0.215

# Discounting and economic settings
discount_EFFECT <- 0.03
discount_QALY <- 0.03
discount_COST <- 0.03
wtp <- 100000
half_cycle_correction <- TRUE
# Load the life table data
life_table <- read.csv("lifetable_US_2019_ssa.csv")

# Convert annual mortality probabilities to mortality rates using: rate = -log(1 - probability)
life_table$male <- -log(1 - life_table$male)
life_table$female <- -log(1 - life_table$female)

# Compute the weighted average of mortality rates (52% female, 48% male)
life_table$weighted <- 0.48 * life_table$male + 0.52 * life_table$female

# Return the result as a data frame with required columns
life_table <- life_table[, c("age", "male", "female", "weighted")]
# Define number of cycles
v.n_cycles <- n.cycle

# Initialize time-dependent vectors for disease progression (standard of care)
v.p.mci_mil_soc <- rep(p.mci_mil, v.n_cycles)
v.p.mci_mod_soc <- rep(p.mci_mod, v.n_cycles)
v.p.mci_sev_soc <- rep(p.mci_sev, v.n_cycles)

v.p.mil_mci_soc <- rep(p.mil_mci, v.n_cycles)
v.p.mil_mod_soc <- rep(p.mil_mod, v.n_cycles)
v.p.mil_sev_soc <- rep(p.mil_sev, v.n_cycles)

v.p.mod_mil_soc <- rep(p.mod_mil, v.n_cycles)
v.p.mod_sev_soc <- rep(p.mod_sev, v.n_cycles)

v.p.sev_mil_soc <- rep(p.sev_mil, v.n_cycles)
v.p.sev_mod_soc <- rep(p.sev_mod, v.n_cycles)

# Staying probabilities per state (excluding institutionalization and mortality)
v.p.stay_mci_soc <- 1 - (v.p.mci_mil_soc + v.p.mci_mod_soc + v.p.mci_sev_soc)
v.p.stay_mil_soc <- 1 - (v.p.mil_mci_soc + v.p.mil_mod_soc + v.p.mil_sev_soc)
v.p.stay_mod_soc <- 1 - (v.p.mod_mil_soc + v.p.mod_sev_soc)
v.p.stay_sev_soc <- 1 - (v.p.sev_mil_soc + v.p.sev_mod_soc)

# Institutionalization transition vectors (community states only)
v.p.mci_i <- rep(p.mci_i, v.n_cycles)
v.p.mil_i <- rep(p.mil_i, v.n_cycles)
v.p.mod_i <- rep(p.mod_i, v.n_cycles)
v.p.sev_i <- rep(p.sev_i, v.n_cycles)

# Discontinuation vector
v.p.discontinue <- rep(0, v.n_cycles)
v.p.discontinue[1] <- p.discontinuation1
v.p.discontinue[2:(tx_duration - 1)] <- p.discontinuation2
v.p.discontinue[tx_duration:v.n_cycles] <- 1

# Load mortality rates from precomputed life_table
# Assuming life_table is already loaded and formatted with a "weighted" column
v.age <- age_start + 0:(v.n_cycles - 1)
v.p.death <- life_table$weighted[match(v.age, life_table$age)]

# Total number of cycles
v.n_cycles <- n.cycle

# Create vectors for on-treatment transition probabilities (community states)
v.p.mci_mil_int_on <- numeric(v.n_cycles)
v.p.mci_mod_int_on <- numeric(v.n_cycles)
v.p.mci_sev_int_on <- numeric(v.n_cycles)

v.p.mil_mci_int_on <- rep(p.mil_mci, v.n_cycles)  # No treatment effect
v.p.mil_mod_int_on <- numeric(v.n_cycles)
v.p.mil_sev_int_on <- numeric(v.n_cycles)

# Create vectors for off-treatment transition probabilities (community states)
v.p.mci_mil_int_off <- numeric(v.n_cycles)
v.p.mci_mod_int_off <- numeric(v.n_cycles)
v.p.mci_sev_int_off <- numeric(v.n_cycles)

v.p.mil_mci_int_off <- rep(p.mil_mci, v.n_cycles)  # No treatment effect
v.p.mil_mod_int_off <- numeric(v.n_cycles)
v.p.mil_sev_int_off <- numeric(v.n_cycles)

# Compute time-dependent transitions
for (i in 1:v.n_cycles) {
  # Waning exponent
  w <- (1 - tx_waning)^(i - 1)
  w_dis <- (1 - tx_waning_dis)^(i - 1)
  
  # MCI transitions (on-treatment)
  rate_mci_mil <- -log(1 - p.mci_mil) * rr.tx_mci_mil^w
  rate_mci_mod <- -log(1 - p.mci_mod) * rr.tx_mci_mod^w
  rate_mci_sev <- -log(1 - p.mci_sev) * rr.tx_mci_sev^w
  
  v.p.mci_mil_int_on[i] <- 1 - exp(-rate_mci_mil)
  v.p.mci_mod_int_on[i] <- 1 - exp(-rate_mci_mod)
  v.p.mci_sev_int_on[i] <- 1 - exp(-rate_mci_sev)
  
  # MCI transitions (off-treatment)
  rate_mci_mil_dis <- -log(1 - p.mci_mil) * rr.tx_mci_mil_dis^w_dis
  rate_mci_mod_dis <- -log(1 - p.mci_mod) * rr.tx_mci_mod_dis^w_dis
  rate_mci_sev_dis <- -log(1 - p.mci_sev) * rr.tx_mci_sev_dis^w_dis
  
  v.p.mci_mil_int_off[i] <- 1 - exp(-rate_mci_mil_dis)
  v.p.mci_mod_int_off[i] <- 1 - exp(-rate_mci_mod_dis)
  v.p.mci_sev_int_off[i] <- 1 - exp(-rate_mci_sev_dis)
  
  # MIL transitions (on-treatment)
  rate_mil_mod <- -log(1 - p.mil_mod) * rr.tx_mil_mod^w
  rate_mil_sev <- -log(1 - p.mil_sev) * rr.tx_mil_sev^w
  
  v.p.mil_mod_int_on[i] <- 1 - exp(-rate_mil_mod)
  v.p.mil_sev_int_on[i] <- 1 - exp(-rate_mil_sev)
  
  # MIL transitions (off-treatment)
  rate_mil_mod_dis <- -log(1 - p.mil_mod) * rr.tx_mil_mod_dis^w_dis
  rate_mil_sev_dis <- -log(1 - p.mil_sev) * rr.tx_mil_sev_dis^w_dis
  
  v.p.mil_mod_int_off[i] <- 1 - exp(-rate_mil_mod_dis)
  v.p.mil_sev_int_off[i] <- 1 - exp(-rate_mil_sev_dis)
}

# Staying probabilities (on-treatment)
v.p.stay_mci_int_on <- 1 - (v.p.mci_mil_int_on + v.p.mci_mod_int_on + v.p.mci_sev_int_on)
v.p.stay_mil_int_on <- 1 - (v.p.mil_mci_int_on + v.p.mil_mod_int_on + v.p.mil_sev_int_on)

# Staying probabilities (off-treatment)
v.p.stay_mci_int_off <- 1 - (v.p.mci_mil_int_off + v.p.mci_mod_int_off + v.p.mci_sev_int_off)
v.p.stay_mil_int_off <- 1 - (v.p.mil_mci_int_off + v.p.mil_mod_int_off + v.p.mil_sev_int_off)
# Function to build transition probability array
build_transition_matrix <- function(strat) {
  # Number of states and cycles
  n.states <- length(v.names_state)
  n.cycles <- n.cycle
  
  # Initialize transition probability array
  a.TP <- array(0, dim = c(n.states, n.states, n.cycles),
                dimnames = list(v.names_state, v.names_state, NULL))
  
  # Loop over cycles to define transitions to death and placeholders
  for (t in 1:n.cycles) {
    # Mortality rate for current cycle
    rate_mort <- v.p.death[t]
    
    # Transitions to death from community states (hazard ratio applied)
    a.TP["mcion_c", "dth", t] <- 1 - exp(-rate_mort * hr.mort_mci)
    a.TP["mciof_c", "dth", t] <- 1 - exp(-rate_mort * hr.mort_mci)
    a.TP["milon_c", "dth", t] <- 1 - exp(-rate_mort * hr.mort_mil)
    a.TP["milof_c", "dth", t] <- 1 - exp(-rate_mort * hr.mort_mil)
    a.TP["mod_c",   "dth", t] <- 1 - exp(-rate_mort * hr.mort_mod)
    a.TP["sev_c",   "dth", t] <- 1 - exp(-rate_mort * hr.mort_sev)
    
    # Transitions to death from institutionalized states
    a.TP["mci_i", "dth", t] <- 1 - exp(-rate_mort * hr.mort_mci)
    a.TP["mil_i", "dth", t] <- 1 - exp(-rate_mort * hr.mort_mil)
    a.TP["mod_i", "dth", t] <- 1 - exp(-rate_mort * hr.mort_mod)
    a.TP["sev_i", "dth", t] <- 1 - exp(-rate_mort * hr.mort_sev)
    
    # Death state is absorbing
    a.TP["dth", "dth", t] <- 1
    
    # Transitions from "mcion_c"
    a.TP["mcion_c", "mcion_c", t] <- if (strat == "soc") {
      v.p.stay_mci_soc[t] * (1 - a.TP["mcion_c", "dth", t]) * (1 - v.p.mci_i[t])
    } else {
      v.p.stay_mci_int_on[t] * (1 - a.TP["mcion_c", "dth", t]) * (1 - v.p.mci_i[t]) * (1 - v.p.discontinue[t])
    }
    
    a.TP["mcion_c", "mciof_c", t] <- if (strat == "soc") {
      0
    } else {
      v.p.stay_mci_int_on[t] * (1 - a.TP["mcion_c", "dth", t]) * (1 - v.p.mci_i[t]) * v.p.discontinue[t]
    }
    
    a.TP["mcion_c", "milon_c", t] <- if (strat == "soc") {
      v.p.mci_mil_soc[t] * (1 - a.TP["mcion_c", "dth", t]) * (1 - v.p.mci_i[t])
    } else {
      v.p.mci_mil_int_on[t] * (1 - a.TP["mcion_c", "dth", t]) * (1 - v.p.mci_i[t]) * (1 - v.p.discontinue[t])
    }
    
    a.TP["mcion_c", "milof_c", t] <- if (strat == "soc") {
      0
    } else {
      v.p.mci_mil_int_on[t] * (1 - a.TP["mcion_c", "dth", t]) * (1 - v.p.mci_i[t]) * v.p.discontinue[t]
    }
    
    a.TP["mcion_c", "mod_c", t] <- if (strat == "soc") {
      v.p.mci_mod_soc[t] * (1 - a.TP["mcion_c", "dth", t])
    } else {
      v.p.mci_mod_int_on[t] * (1 - a.TP["mcion_c", "dth", t])
    }
    
    a.TP["mcion_c", "sev_c", t] <- if (strat == "soc") {
      v.p.mci_sev_soc[t] * (1 - a.TP["mcion_c", "dth", t])
    } else {
      v.p.mci_sev_int_on[t] * (1 - a.TP["mcion_c", "dth", t])
    }
    
    a.TP["mcion_c", "mci_i", t] <- if (strat == "soc") {
      v.p.mci_i[t] * (1 - a.TP["mcion_c", "dth", t])
    } else {
      v.p.mci_i[t] * (1 - a.TP["mcion_c", "dth", t])
    }
    
    # Transitions from "milon_c"
    a.TP["milon_c", "milon_c", t] <- if (strat == "soc") {
      v.p.stay_mil_soc[t] * (1 - a.TP["milon_c", "dth", t]) * (1 - v.p.mil_i[t])
    } else {
      v.p.stay_mil_int_on[t] * (1 - a.TP["milon_c", "dth", t]) * (1 - v.p.mil_i[t]) * (1 - v.p.discontinue[t])
    }
    
    a.TP["milon_c", "milof_c", t] <- if (strat == "soc") {
      0
    } else {
      v.p.stay_mil_int_on[t] * (1 - a.TP["milon_c", "dth", t]) * (1 - v.p.mil_i[t]) * v.p.discontinue[t]
    }
    
    a.TP["milon_c", "mciof_c", t] <- if (strat == "soc") {
      p.mil_mci * (1 - a.TP["milon_c", "dth", t]) * (1 - v.p.mil_i[t])
    } else {
      p.mil_mci * (1 - a.TP["milon_c", "dth", t]) * (1 - v.p.mil_i[t]) * v.p.discontinue[t]
    }
    
    a.TP["milon_c", "mod_c", t] <- if (strat == "soc") {
      v.p.mil_mod_soc[t] * (1 - a.TP["milon_c", "dth", t])
    } else {
      v.p.mil_mod_int_on[t] * (1 - a.TP["milon_c", "dth", t])
    }
    
    a.TP["milon_c", "sev_c", t] <- if (strat == "soc") {
      v.p.mil_sev_soc[t] * (1 - a.TP["milon_c", "dth", t])
    } else {
      v.p.mil_sev_int_on[t] * (1 - a.TP["milon_c", "dth", t])
    }
    
    a.TP["milon_c", "mil_i", t] <- if (strat == "soc") {
      v.p.mil_i[t] * (1 - a.TP["milon_c", "dth", t])
    } else {
      v.p.mil_i[t] * (1 - a.TP["milon_c", "dth", t])
    }
    
    # Transitions from "mciof_c"
    a.TP["mciof_c", "mciof_c", t] <- if (strat == "soc") {
      v.p.stay_mci_soc[t] * (1 - a.TP["mciof_c", "dth", t]) * (1 - v.p.mci_i[t])
    } else {
      v.p.stay_mci_int_off[t] * (1 - a.TP["mciof_c", "dth", t]) * (1 - v.p.mci_i[t])
    }
    
    a.TP["mciof_c", "milof_c", t] <- if (strat == "soc") {
      v.p.mci_mil_soc[t] * (1 - a.TP["mciof_c", "dth", t]) * (1 - v.p.mci_i[t])
    } else {
      v.p.mci_mil_int_off[t] * (1 - a.TP["mciof_c", "dth", t]) * (1 - v.p.mci_i[t])
    }
    
    a.TP["mciof_c", "mod_c", t] <- if (strat == "soc") {
      v.p.mci_mod_soc[t] * (1 - a.TP["mciof_c", "dth", t])
    } else {
      v.p.mci_mod_int_off[t] * (1 - a.TP["mciof_c", "dth", t])
    }
    
    a.TP["mciof_c", "sev_c", t] <- if (strat == "soc") {
      v.p.mci_sev_soc[t] * (1 - a.TP["mciof_c", "dth", t])
    } else {
      v.p.mci_sev_int_off[t] * (1 - a.TP["mciof_c", "dth", t])
    }
    
    a.TP["mciof_c", "mci_i", t] <- v.p.mci_i[t] * (1 - a.TP["mciof_c", "dth", t])
    
    # Transitions from "milof_c"
    a.TP["milof_c", "milof_c", t] <- if (strat == "soc") {
      v.p.stay_mil_soc[t] * (1 - a.TP["milof_c", "dth", t]) * (1 - v.p.mil_i[t])
    } else {
      v.p.stay_mil_int_off[t] * (1 - a.TP["milof_c", "dth", t]) * (1 - v.p.mil_i[t])
    }
    
    a.TP["milof_c", "mciof_c", t] <- p.mil_mci * (1 - a.TP["milof_c", "dth", t]) * (1 - v.p.mil_i[t])
    
    a.TP["milof_c", "mod_c", t] <- if (strat == "soc") {
      v.p.mil_mod_soc[t] * (1 - a.TP["milof_c", "dth", t])
    } else {
      v.p.mil_mod_int_off[t] * (1 - a.TP["milof_c", "dth", t])
    }
    
    a.TP["milof_c", "sev_c", t] <- if (strat == "soc") {
      v.p.mil_sev_soc[t] * (1 - a.TP["milof_c", "dth", t])
    } else {
      v.p.mil_sev_int_off[t] * (1 - a.TP["milof_c", "dth", t])
    }
    
    a.TP["milof_c", "mil_i", t] <- v.p.mil_i[t] * (1 - a.TP["milof_c", "dth", t])
    
    # Transitions from "mod_c"
    a.TP["mod_c", "mod_c", t] <- v.p.stay_mod_soc[t] * (1 - a.TP["mod_c", "dth", t]) * (1 - v.p.mod_i[t])
    a.TP["mod_c", "milof_c", t] <- p.mod_mil * (1 - a.TP["mod_c", "dth", t]) * (1 - v.p.mod_i[t])
    a.TP["mod_c", "sev_c", t] <- v.p.mod_sev_soc[t] * (1 - a.TP["mod_c", "dth", t])
    a.TP["mod_c", "mod_i", t] <- v.p.mod_i[t] * (1 - a.TP["mod_c", "dth", t])
    
    # Transitions from "sev_c"
    a.TP["sev_c", "sev_c", t] <- v.p.stay_sev_soc[t] * (1 - a.TP["sev_c", "dth", t]) * (1 - v.p.sev_i[t])
    a.TP["sev_c", "milof_c", t] <- p.sev_mil * (1 - a.TP["sev_c", "dth", t]) * (1 - v.p.sev_i[t])
    a.TP["sev_c", "mod_c", t] <- p.sev_mod * (1 - a.TP["sev_c", "dth", t]) * (1 - v.p.sev_i[t])
    a.TP["sev_c", "sev_i", t] <- v.p.sev_i[t] * (1 - a.TP["sev_c", "dth", t])
    
    # Transitions from "mci_i"
    a.TP["mci_i", "mci_i", t] <- v.p.stay_mci_soc[t] * (1 - a.TP["mci_i", "dth", t])
    a.TP["mci_i", "mil_i", t] <- v.p.mci_mil_soc[t] * (1 - a.TP["mci_i", "dth", t])
    a.TP["mci_i", "mod_i", t] <- v.p.mci_mod_soc[t] * (1 - a.TP["mci_i", "dth", t])
    a.TP["mci_i", "sev_i", t] <- v.p.mci_sev_soc[t] * (1 - a.TP["mci_i", "dth", t])
    
    # Transitions from "mil_i"
    a.TP["mil_i", "mil_i", t] <- v.p.stay_mil_soc[t] * (1 - a.TP["mil_i", "dth", t])
    a.TP["mil_i", "mci_i", t] <- p.mil_mci * (1 - a.TP["mil_i", "dth", t])
    a.TP["mil_i", "mod_i", t] <- v.p.mil_mod_soc[t] * (1 - a.TP["mil_i", "dth", t])
    a.TP["mil_i", "sev_i", t] <- v.p.mil_sev_soc[t] * (1 - a.TP["mil_i", "dth", t])
    
    # Transitions from "mod_i"
    a.TP["mod_i", "mod_i", t] <- v.p.stay_mod_soc[t] * (1 - a.TP["mod_i", "dth", t])
    a.TP["mod_i", "mil_i", t] <- p.mod_mil * (1 - a.TP["mod_i", "dth", t])
    a.TP["mod_i", "sev_i", t] <- v.p.mod_sev_soc[t] * (1 - a.TP["mod_i", "dth", t])
    
    # Transitions from "sev_i"
    a.TP["sev_i", "sev_i", t] <- v.p.stay_sev_soc[t] * (1 - a.TP["sev_i", "dth", t])
    a.TP["sev_i", "mil_i", t] <- p.sev_mil * (1 - a.TP["sev_i", "dth", t])
    a.TP["sev_i", "mod_i", t] <- p.sev_mod * (1 - a.TP["sev_i", "dth", t])
    
    # Normalize each row in the transition matrix at cycle t if row sum != 1
    for (s in v.names_state) {
      row_sum <- sum(a.TP[s, , t])
      if (row_sum > 0 && abs(row_sum - 1) > 1e-8) {
        a.TP[s, , t] <- a.TP[s, , t] / row_sum
      }
    }
  }
  
  return(a.TP)
}
# Function to compute Markov trace and total outcomes
run_markov_model <- function(strat, a.TP) {
  # Initialize state index vector
  v.states <- v.names_state
  n.states <- length(v.states)
  n.cycles <- n.cycle
  
  # Create named vectors for utility (patient + caregiver) and cost
  v.utility <- c(
    mcion_c = u.mci_pt + u.mci_ic,
    mciof_c = u.mci_pt + u.mci_ic,
    milon_c = u.mil_pt + u.mil_ic,
    milof_c = u.mil_pt + u.mil_ic,
    mod_c   = u.mod_pt + u.mod_ic,
    sev_c   = u.sev_pt + u.sev_ic,
    mci_i   = u.mci_pt_i + u.mci_ic_i,
    mil_i   = u.mil_pt_i + u.mil_ic_i,
    mod_i   = u.mod_pt_i + u.mod_ic_i,
    sev_i   = u.sev_pt_i + u.sev_ic_i,
    dth     = 0
  )
  
  v.cost <- c(
    mcion_c = c.mci_hc + c.mci_sc + c.mci_ic,
    mciof_c = c.mci_hc + c.mci_sc + c.mci_ic,
    milon_c = c.mil_hc + c.mil_sc + c.mil_ic,
    milof_c = c.mil_hc + c.mil_sc + c.mil_ic,
    mod_c   = c.mod_hc + c.mod_sc + c.mod_ic,
    sev_c   = c.sev_hc + c.sev_sc + c.sev_ic,
    mci_i   = c.mci_i_hc + c.mci_i_sc + c.mci_i_ic,
    mil_i   = c.mil_i_hc + c.mil_i_sc + c.mil_i_ic,
    mod_i   = c.mod_i_hc + c.mod_i_sc + c.mod_i_ic,
    sev_i   = c.sev_i_hc + c.sev_i_sc + c.sev_i_ic,
    dth     = 0
  )
  
  # Initialize initial state vector
  v.initial <- setNames(rep(0, n.states), v.states)
  if (strat == "soc") {
    v.initial["mciof_c"] <- 1 - p.starting_state_mci
    v.initial["mciof_c"] <- v.initial["mciof_c"] + 0  # No starting in institutional care
    v.initial["mcion_c"] <- p.starting_state_mci
  } else if (strat == "int") {
    v.initial["mcion_c"] <- p.starting_state_mci
    v.initial["milon_c"] <- 1 - p.starting_state_mci
  }
  
  # Initialize trace and outcomes matrices
  m.trace <- matrix(0, nrow = n.cycles, ncol = n.states, dimnames = list(NULL, v.states))
  v.QALYs <- numeric(n.cycles)
  v.Costs <- numeric(n.cycles)
  v.LYs   <- numeric(n.cycles)
  
  # Fill initial state
  m.trace[1, ] <- v.initial
  
  # Run model for each cycle
  for (t in 2:n.cycles) {
    m.trace[t, ] <- m.trace[t - 1, ] %*% a.TP[ , , t - 1]
  }
  
  # Apply outcomes per cycle
  for (t in 1:n.cycles) {
    state_vector <- m.trace[t, ]
    
    v.QALYs[t] <- sum(state_vector * v.utility)
    v.Costs[t] <- sum(state_vector * v.cost)
    v.LYs[t]   <- sum(state_vector * (v.states != "dth"))
    
    # Apply treatment costs and utility impact
    if (strat == "int") {
      v.Costs[t] <- v.Costs[t] + sum(state_vector[c("mcion_c", "milon_c")] * c.Tx)
      
      if (t == 1) {
        v.Costs[t] <- v.Costs[t] + c.Tx_start
        v.QALYs[t] <- v.QALYs[t] + u.Tx_start
      }
    }
  }
  
  # Apply half-cycle correction
  v.QALYs <- (v.QALYs[-n.cycles] + v.QALYs[-1]) / 2
  v.Costs <- (v.Costs[-n.cycles] + v.Costs[-1]) / 2
  v.LYs   <- (v.LYs[-n.cycles] + v.LYs[-1]) / 2
  
  # Apply discounting
  v.discount_weights <- 1 / ((1 + discount_QALY)^(1:(n.cycles - 1)))
  total.QALY <- sum(v.QALYs * v.discount_weights)
  total.Cost <- sum(v.Costs * v.discount_weights)
  total.LY   <- sum(v.LYs   * v.discount_weights)
  
  return(list(QALYs = total.QALY, Costs = total.Cost, LYs = total.LY))
}

# Run model for both strategies
res.soc <- run_markov_model("soc", build_transition_matrix("soc"))
res.int <- run_markov_model("int", build_transition_matrix("int"))

# Print results
cat("Strategy Comparison Results\n")
cat("===========================\n")
cat(sprintf("Standard of Care - QALYs: %.2f, Costs: $%.0f, LYs: %.2f\n", res.soc$QALYs, res.soc$Costs, res.soc$LYs))
cat(sprintf("Intervention     - QALYs: %.2f, Costs: $%.0f, LYs: %.2f\n", res.int$QALYs, res.int$Costs, res.int$LYs))
cat("===========================\n")
cat(sprintf("Incremental      - QALYs: %.2f, Costs: $%.0f, LYs: %.2f\n",
            res.int$QALYs - res.soc$QALYs,
            res.int$Costs - res.soc$Costs,
            res.int$LYs   - res.soc$LYs))
```
