## Supplementary material for "Replicating Health-Economic Simulation Models for Alzheimer’s Disease Using Artificial Intelligence": final R scripts: o8.R

```
# IPECAD Markov Model Inputs for dampack (Prompt 1)

# Load necessary libraries
library(dampack)

# Define state names and strategy names
v.names_state <- c("mcion_c","mciof_c","milon_c","milof_c","mod_c","sev_c","mci_i","mil_i","mod_i","sev_i","dth")
v.names_strat <- c("soc", "int")

# Starting population characteristics
age_start <- 71
sex <- "weighted"  # Represents 52% female
p.starting_state_mci <- 0.55

# Model time horizon
n.cycles <- 29

# Transition probabilities (from Table E4-E6)
p.mci_mil <- 0.23
p.mci_mod <- 0
p.mci_sev <- 0
p.mil_mci <- 0.03
p.mil_mod <- 0.35
p.mil_sev <- 0.04
p.mod_mil <- 0.03
p.mod_sev <- 0.42
p.sev_mil <- 0
p.sev_mod <- 0.02

# Transition to institution care
p.mci_i <- 0.024
p.mil_i <- 0.038
p.mod_i <- 0.110
p.sev_i <- 0.259

# Mortality hazard ratios by disease state
hr.mort_mci <- 1.82
hr.mort_mil <- 2.92
hr.mort_mod <- 3.85
hr.mort_sev <- 9.52

# Relative risks for treatment (on-treatment)
rr.tx_mci_mil <- 0.69
rr.tx_mci_mod <- 1
rr.tx_mci_sev <- 1
rr.tx_mil_mod <- 0.69
rr.tx_mil_sev <- 0.69

# Relative risks for treatment (off-treatment)
rr.tx_mci_mil_dis <- 1
rr.tx_mci_mod_dis <- 1
rr.tx_mci_sev_dis <- 1
rr.tx_mil_mod_dis <- 1
rr.tx_mil_sev_dis <- 1

# Discontinuation parameters
p.discontinuation1 <- 0.069
p.discontinuation2 <- 0
discontinuation2_begin <- 2
tx_duration <- 29

# Treatment effect waning
tx_waning <- 0
tx_waning_dis <- 0

# Utility values (patients - community)
u.mci_pt <- 0.851 - 0.17
u.mil_pt <- 0.851 - 0.22
u.mod_pt <- 0.851 - 0.36
u.sev_pt <- 0.851 - 0.53

# Utility values (patients - institution)
u.mci_pt_i <- 0.851 - 0.17
u.mil_pt_i <- 0.851 - 0.19
u.mod_pt_i <- 0.851 - 0.42
u.sev_pt_i <- 0.851 - 0.59

# Utility values (informal caregivers - community)
u.mci_ic <- -0.03
u.mil_ic <- -0.05
u.mod_ic <- -0.08
u.sev_ic <- -0.10

# Utility values (informal caregivers - institution)
u.mci_ic_i <- -0.03
u.mil_ic_i <- -0.05
u.mod_ic_i <- -0.08
u.sev_ic_i <- -0.10

# Utility decrement from ARIA in first cycle
u.Tx_start <- -0.14 * (12/52) * 0.035

# Cost inputs (healthcare - community)
c.mci_hc <- 6042 * 1.12 + 460
c.mil_hc <- 6042 * 1.56 + 965 + 0.21 * 365 * 0.333
c.mod_hc <- 6042 * 1.93 + 1544 + 0.66 * 365 * 0.333
c.sev_hc <- 6042 * 1.93 + 1930

# Cost inputs (healthcare - institution)
c.mci_i_hc <- c.mci_hc
c.mil_i_hc <- c.mil_hc
c.mod_i_hc <- c.mod_hc
c.sev_i_hc <- c.sev_hc

# Cost inputs (social care - community)
c.mci_sc <- 0
c.mil_sc <- 0
c.mod_sc <- 0
c.sev_sc <- 0

# Cost inputs (social care - institution)
c.mci_i_sc <- 7394 * 12
c.mil_i_sc <- 7394 * 12
c.mod_i_sc <- 7394 * 12
c.sev_i_sc <- 7394 * 12

# Cost inputs (informal care - community)
c.mci_ic <- 69 * 12 * 32.46 + 0.204 * 0.049 * 20 * 52 * 32.46
c.mil_ic <- 113 * 12 * 32.46 + 0.112 * 0.086 * 20 * 52 * 32.46
c.mod_ic <- 169 * 12 * 32.46
c.sev_ic <- 298 * 12 * 32.46

# Cost inputs (informal care - institution)
c.mci_i_ic <- 69 * 12 * 32.46 * 0.44 + 0.204 * 0.049 * 20 * 52 * 32.46
c.mil_i_ic <- 113 * 12 * 32.46 * 0.44 + 0.112 * 0.086 * 20 * 52 * 32.46
c.mod_i_ic <- 169 * 12 * 32.46 * 0.44
c.sev_i_ic <- 298 * 12 * 32.46 * 0.44

# Treatment costs
c.Tx <- 26500 + (52/2) * 78.35
c.Tx_start <- 261.10 * 4 + 261.10 * 3 * 0.215

# Discounting
discount_EFFECT <- 0.03
discount_QALY <- 0.03
discount_COST <- 0.03

# Willingness-to-pay threshold
wtp <- 100000

# Half-cycle correction flag
half_cycle_correction <- TRUE
# Load life table data
lifetable <- read.csv("lifetable_US_2019_ssa.csv")

# Convert annual mortality probabilities (q) to annual mortality rates (r)
# using the formula: r = -log(1 - q)
lifetable$male <- -log(1 - lifetable$male)
lifetable$female <- -log(1 - lifetable$female)

# Define sex proportion for weighting: 52% female and 48% male
prop_male <- 0.48
prop_female <- 0.52

# Calculate weighted average mortality rate
lifetable$weighted <- lifetable$male * prop_male + lifetable$female * prop_female

# Return the resulting dataframe with converted rates
lifetable <- lifetable[, c("age", "male", "female", "weighted")]
# Define total number of cycles
v.n_cycles <- n.cycles

# Initialize time-dependent transition probability vectors (standard of care)

# Disease progression (forward and backward where allowed)
v.p.mci_mil_soc <- rep(p.mci_mil, v.n_cycles)
v.p.mci_mod_soc <- rep(p.mci_mod, v.n_cycles)
v.p.mci_sev_soc <- rep(p.mci_sev, v.n_cycles)

v.p.mil_mci_soc <- rep(p.mil_mci, v.n_cycles)
v.p.mil_mod_soc <- rep(p.mil_mod, v.n_cycles)
v.p.mil_sev_soc <- rep(p.mil_sev, v.n_cycles)

v.p.mod_mil_soc <- rep(p.mod_mil, v.n_cycles)
v.p.mod_sev_soc <- rep(p.mod_sev, v.n_cycles)

v.p.sev_mil_soc <- rep(p.sev_mil, v.n_cycles)
v.p.sev_mod_soc <- rep(p.sev_mod, v.n_cycles)

# Staying probabilities (excluding transitions to institutional care and death)
v.p.mci_stay_soc <- 1 - (v.p.mci_mil_soc + v.p.mci_mod_soc + v.p.mci_sev_soc)
v.p.mil_stay_soc <- 1 - (v.p.mil_mci_soc + v.p.mil_mod_soc + v.p.mil_sev_soc)
v.p.mod_stay_soc <- 1 - (v.p.mod_mil_soc + v.p.mod_sev_soc)
v.p.sev_stay_soc <- 1 - (v.p.sev_mil_soc + v.p.sev_mod_soc)

# Institutionalization transition probabilities
v.p.mci_inst <- rep(p.mci_i, v.n_cycles)
v.p.mil_inst <- rep(p.mil_i, v.n_cycles)
v.p.mod_inst <- rep(p.mod_i, v.n_cycles)
v.p.sev_inst <- rep(p.sev_i, v.n_cycles)

# Discontinuation vector: p.discontinuation1 applies only at cycle 1,
# p.discontinuation2 from cycle 2 to tx_duration, then 1 afterwards
v.p.discontinuation <- c(
  rep(p.discontinuation1, 1),
  rep(p.discontinuation2, tx_duration - 1),
  rep(1, v.n_cycles - tx_duration)
)

# Vector of age- and sex-weighted mortality rates from the life table
# Use the 'lifetable' dataframe defined in Prompt 2
v.age <- age_start + 0:(v.n_cycles - 1)
v.p.mortality <- lifetable$weighted[match(v.age, lifetable$age)]
# Initialize index vector for cycles
v.cycles <- 1:n.cycles

# Waning factors per cycle (on-treatment and off-treatment)
v.waning_factor_on <- (1 - tx_waning)^(v.cycles - 1)
v.waning_factor_off <- (1 - tx_waning_dis)^(v.cycles - 1)

# ----- On-Treatment Transitions: mcion_c -----
rate.mci_mil <- -log(1 - p.mci_mil)
v.p.mci_mil_int <- 1 - exp(-rate.mci_mil * rr.tx_mci_mil^v.waning_factor_on)

rate.mci_mod <- -log(1 - p.mci_mod)
v.p.mci_mod_int <- 1 - exp(-rate.mci_mod * rr.tx_mci_mod^v.waning_factor_on)

rate.mci_sev <- -log(1 - p.mci_sev)
v.p.mci_sev_int <- 1 - exp(-rate.mci_sev * rr.tx_mci_sev^v.waning_factor_on)

# Staying probability
v.p.mci_stay_int <- 1 - (v.p.mci_mil_int + v.p.mci_mod_int + v.p.mci_sev_int)

# ----- On-Treatment Transitions: milon_c -----
rate.mil_mci <- -log(1 - p.mil_mci)
v.p.mil_mci_int <- rep(p.mil_mci, n.cycles)  # Backward transition, no treatment effect

rate.mil_mod <- -log(1 - p.mil_mod)
v.p.mil_mod_int <- 1 - exp(-rate.mil_mod * rr.tx_mil_mod^v.waning_factor_on)

rate.mil_sev <- -log(1 - p.mil_sev)
v.p.mil_sev_int <- 1 - exp(-rate.mil_sev * rr.tx_mil_sev^v.waning_factor_on)

# Staying probability
v.p.mil_stay_int <- 1 - (v.p.mil_mci_int + v.p.mil_mod_int + v.p.mil_sev_int)

# ----- Off-Treatment Transitions: mciof_c -----
v.p.mci_mil_int_dis <- 1 - exp(-rate.mci_mil * rr.tx_mci_mil_dis^v.waning_factor_off)
v.p.mci_mod_int_dis <- 1 - exp(-rate.mci_mod * rr.tx_mci_mod_dis^v.waning_factor_off)
v.p.mci_sev_int_dis <- 1 - exp(-rate.mci_sev * rr.tx_mci_sev_dis^v.waning_factor_off)

# Staying probability
v.p.mci_stay_int_dis <- 1 - (v.p.mci_mil_int_dis + v.p.mci_mod_int_dis + v.p.mci_sev_int_dis)

# ----- Off-Treatment Transitions: milof_c -----
v.p.mil_mci_int_dis <- rep(p.mil_mci, n.cycles)  # Backward transition, no treatment effect

v.p.mil_mod_int_dis <- 1 - exp(-rate.mil_mod * rr.tx_mil_mod_dis^v.waning_factor_off)
v.p.mil_sev_int_dis <- 1 - exp(-rate.mil_sev * rr.tx_mil_sev_dis^v.waning_factor_off)

# Staying probability
v.p.mil_stay_int_dis <- 1 - (v.p.mil_mci_int_dis + v.p.mil_mod_int_dis + v.p.mil_sev_int_dis)


# Function to build transition probability matrix for a given strategy
build_transition_matrix <- function(strat) {
  
  # Initialize transition probability array (states × states × cycles)
  n.states <- length(v.names_state)
  a.TP <- array(0, dim = c(n.states, n.states, n.cycles),
                dimnames = list(v.names_state, v.names_state, NULL))
  
  # Loop through each cycle to fill death transitions and placeholders
  for (t in 1:n.cycles) {
    
    # Extract mortality rate for current cycle
    mort_rate <- v.p.mortality[t]
    
    # ----- Transitions to death -----
    a.TP["mcion_c", "dth", t]   <- 1 - exp(-mort_rate * hr.mort_mci)
    a.TP["mciof_c", "dth", t]   <- 1 - exp(-mort_rate * hr.mort_mci)
    a.TP["milon_c", "dth", t]   <- 1 - exp(-mort_rate * hr.mort_mil)
    a.TP["milof_c", "dth", t]   <- 1 - exp(-mort_rate * hr.mort_mil)
    a.TP["mod_c", "dth", t]     <- 1 - exp(-mort_rate * hr.mort_mod)
    a.TP["sev_c", "dth", t]     <- 1 - exp(-mort_rate * hr.mort_sev)
    
    a.TP["mci_i", "dth", t]     <- 1 - exp(-mort_rate * hr.mort_mci)
    a.TP["mil_i", "dth", t]     <- 1 - exp(-mort_rate * hr.mort_mil)
    a.TP["mod_i", "dth", t]     <- 1 - exp(-mort_rate * hr.mort_mod)
    a.TP["sev_i", "dth", t]     <- 1 - exp(-mort_rate * hr.mort_sev)
    
    # Absorbing state: death
    a.TP["dth", "dth", t]       <- 1
    
    # ----- Placeholders for future transitions -----
    # ----- [INSERT PROMPT 5B HERE] — On-treatment community state transitions -----
    
    # Transitions from "mcion_c" (on-treatment MCI community)
    p_mort <- a.TP["mcion_c", "dth", t]
    surv_prob <- 1 - p_mort
    disc_prob <- v.p.discontinuation[t]
    inst_prob <- v.p.mci_inst[t]
    comm_prob <- 1 - inst_prob
    
    if (strat == "int") {
      # To "mcion_c" (stay)
      a.TP["mcion_c", "mcion_c", t] <- v.p.mci_stay_int[t] * surv_prob * comm_prob * (1 - disc_prob)
      # To "mciof_c" (discontinue)
      a.TP["mcion_c", "mciof_c", t] <- v.p.mci_stay_int[t] * surv_prob * comm_prob * disc_prob
      # To "milon_c"
      a.TP["mcion_c", "milon_c", t] <- v.p.mci_mil_int[t] * surv_prob * comm_prob * (1 - disc_prob)
      # To "milof_c"
      a.TP["mcion_c", "milof_c", t] <- v.p.mci_mil_int[t] * surv_prob * comm_prob * disc_prob
      # To "mod_c"
      a.TP["mcion_c", "mod_c", t]   <- v.p.mci_mod_int[t] * surv_prob * comm_prob
      # To "sev_c"
      a.TP["mcion_c", "sev_c", t]   <- v.p.mci_sev_int[t] * surv_prob * comm_prob
      # To "mci_i"
      a.TP["mcion_c", "mci_i", t]   <- v.p.mci_stay_int[t] * surv_prob * inst_prob
    } else {
      # Standard of care uses soc suffix vectors
      a.TP["mcion_c", "mcion_c", t] <- v.p.mci_stay_soc[t] * surv_prob * comm_prob * (1 - disc_prob)
      a.TP["mcion_c", "mciof_c", t] <- v.p.mci_stay_soc[t] * surv_prob * comm_prob * disc_prob
      a.TP["mcion_c", "milon_c", t] <- v.p.mci_mil_soc[t] * surv_prob * comm_prob * (1 - disc_prob)
      a.TP["mcion_c", "milof_c", t] <- v.p.mci_mil_soc[t] * surv_prob * comm_prob * disc_prob
      a.TP["mcion_c", "mod_c", t]   <- v.p.mci_mod_soc[t] * surv_prob * comm_prob
      a.TP["mcion_c", "sev_c", t]   <- v.p.mci_sev_soc[t] * surv_prob * comm_prob
      a.TP["mcion_c", "mci_i", t]   <- v.p.mci_stay_soc[t] * surv_prob * inst_prob
    }
    
    # Transitions from "milon_c" (on-treatment mild community)
    p_mort <- a.TP["milon_c", "dth", t]
    surv_prob <- 1 - p_mort
    disc_prob <- v.p.discontinuation[t]
    inst_prob <- v.p.mil_inst[t]
    comm_prob <- 1 - inst_prob
    
    if (strat == "int") {
      a.TP["milon_c", "milon_c", t] <- v.p.mil_stay_int[t] * surv_prob * comm_prob * (1 - disc_prob)
      a.TP["milon_c", "milof_c", t] <- v.p.mil_stay_int[t] * surv_prob * comm_prob * disc_prob
      a.TP["milon_c", "mciof_c", t] <- v.p.mil_mci_int[t] * surv_prob * comm_prob * disc_prob
      a.TP["milon_c", "mci_i", t]   <- v.p.mil_mci_int[t] * surv_prob * inst_prob
      a.TP["milon_c", "mod_c", t]   <- v.p.mil_mod_int[t] * surv_prob * comm_prob
      a.TP["milon_c", "sev_c", t]   <- v.p.mil_sev_int[t] * surv_prob * comm_prob
      a.TP["milon_c", "mil_i", t]   <- v.p.mil_stay_int[t] * surv_prob * inst_prob
    } else {
      a.TP["milon_c", "milon_c", t] <- v.p.mil_stay_soc[t] * surv_prob * comm_prob * (1 - disc_prob)
      a.TP["milon_c", "milof_c", t] <- v.p.mil_stay_soc[t] * surv_prob * comm_prob * disc_prob
      a.TP["milon_c", "mciof_c", t] <- v.p.mil_mci_soc[t] * surv_prob * comm_prob * disc_prob
      a.TP["milon_c", "mci_i", t]   <- v.p.mil_mci_soc[t] * surv_prob * inst_prob
      a.TP["milon_c", "mod_c", t]   <- v.p.mil_mod_soc[t] * surv_prob * comm_prob
      a.TP["milon_c", "sev_c", t]   <- v.p.mil_sev_soc[t] * surv_prob * comm_prob
      a.TP["milon_c", "mil_i", t]   <- v.p.mil_stay_soc[t] * surv_prob * inst_prob
    }
    
    # ----- [INSERT PROMPT 5C HERE] — Off-treatment, moderate/severe community state transitions -----
    
    # Transitions from "mciof_c"
    p_mort <- a.TP["mciof_c", "dth", t]
    surv_prob <- 1 - p_mort
    inst_prob <- v.p.mci_inst[t]
    comm_prob <- 1 - inst_prob
    
    if (strat == "int") {
      a.TP["mciof_c", "mciof_c", t] <- v.p.mci_stay_int_dis[t] * surv_prob * comm_prob
      a.TP["mciof_c", "milof_c", t] <- v.p.mci_mil_int_dis[t] * surv_prob * comm_prob
      a.TP["mciof_c", "mod_c", t]   <- v.p.mci_mod_int_dis[t] * surv_prob * comm_prob
      a.TP["mciof_c", "sev_c", t]   <- v.p.mci_sev_int_dis[t] * surv_prob * comm_prob
      a.TP["mciof_c", "mci_i", t]   <- v.p.mci_stay_int_dis[t] * surv_prob * inst_prob
    } else {
      a.TP["mciof_c", "mciof_c", t] <- v.p.mci_stay_soc[t] * surv_prob * comm_prob
      a.TP["mciof_c", "milof_c", t] <- v.p.mci_mil_soc[t] * surv_prob * comm_prob
      a.TP["mciof_c", "mod_c", t]   <- v.p.mci_mod_soc[t] * surv_prob * comm_prob
      a.TP["mciof_c", "sev_c", t]   <- v.p.mci_sev_soc[t] * surv_prob * comm_prob
      a.TP["mciof_c", "mci_i", t]   <- v.p.mci_stay_soc[t] * surv_prob * inst_prob
    }
    
    # Transitions from "milof_c"
    p_mort <- a.TP["milof_c", "dth", t]
    surv_prob <- 1 - p_mort
    inst_prob <- v.p.mil_inst[t]
    comm_prob <- 1 - inst_prob
    
    if (strat == "int") {
      a.TP["milof_c", "milof_c", t] <- v.p.mil_stay_int_dis[t] * surv_prob * comm_prob
      a.TP["milof_c", "mciof_c", t] <- v.p.mil_mci_int_dis[t] * surv_prob * comm_prob
      a.TP["milof_c", "mod_c", t]   <- v.p.mil_mod_int_dis[t] * surv_prob * comm_prob
      a.TP["milof_c", "sev_c", t]   <- v.p.mil_sev_int_dis[t] * surv_prob * comm_prob
      a.TP["milof_c", "mil_i", t]   <- v.p.mil_stay_int_dis[t] * surv_prob * inst_prob
    } else {
      a.TP["milof_c", "milof_c", t] <- v.p.mil_stay_soc[t] * surv_prob * comm_prob
      a.TP["milof_c", "mciof_c", t] <- v.p.mil_mci_soc[t] * surv_prob * comm_prob
      a.TP["milof_c", "mod_c", t]   <- v.p.mil_mod_soc[t] * surv_prob * comm_prob
      a.TP["milof_c", "sev_c", t]   <- v.p.mil_sev_soc[t] * surv_prob * comm_prob
      a.TP["milof_c", "mil_i", t]   <- v.p.mil_stay_soc[t] * surv_prob * inst_prob
    }
    
    # Transitions from "mod_c"
    p_mort <- a.TP["mod_c", "dth", t]
    surv_prob <- 1 - p_mort
    inst_prob <- v.p.mod_inst[t]
    comm_prob <- 1 - inst_prob
    
    a.TP["mod_c", "mod_c", t] <- v.p.mod_stay_soc[t] * surv_prob * comm_prob
    a.TP["mod_c", "milof_c", t] <- v.p.mod_mil_soc[t] * surv_prob * comm_prob
    a.TP["mod_c", "sev_c", t] <- v.p.mod_sev_soc[t] * surv_prob * comm_prob
    a.TP["mod_c", "mod_i", t] <- v.p.mod_stay_soc[t] * surv_prob * inst_prob
    
    # Transitions from "sev_c"
    p_mort <- a.TP["sev_c", "dth", t]
    surv_prob <- 1 - p_mort
    inst_prob <- v.p.sev_inst[t]
    comm_prob <- 1 - inst_prob
    
    a.TP["sev_c", "sev_c", t] <- v.p.sev_stay_soc[t] * surv_prob * comm_prob
    a.TP["sev_c", "milof_c", t] <- v.p.sev_mil_soc[t] * surv_prob * comm_prob
    a.TP["sev_c", "mod_c", t] <- v.p.sev_mod_soc[t] * surv_prob * comm_prob
    a.TP["sev_c", "sev_i", t] <- v.p.sev_stay_soc[t] * surv_prob * inst_prob
    
    # ----- [INSERT PROMPT 5D HERE] — Institutionalized state transitions -----
    
    # Transitions from "mci_i"
    p_mort <- a.TP["mci_i", "dth", t]
    surv_prob <- 1 - p_mort
    
    if (strat == "int") {
      a.TP["mci_i", "mci_i", t] <- v.p.mci_stay_int_dis[t] * surv_prob
      a.TP["mci_i", "mil_i", t] <- v.p.mci_mil_int_dis[t] * surv_prob
      a.TP["mci_i", "mod_i", t] <- v.p.mci_mod_int_dis[t] * surv_prob
      a.TP["mci_i", "sev_i", t] <- v.p.mci_sev_int_dis[t] * surv_prob
    } else {
      a.TP["mci_i", "mci_i", t] <- v.p.mci_stay_soc[t] * surv_prob
      a.TP["mci_i", "mil_i", t] <- v.p.mci_mil_soc[t] * surv_prob
      a.TP["mci_i", "mod_i", t] <- v.p.mci_mod_soc[t] * surv_prob
      a.TP["mci_i", "sev_i", t] <- v.p.mci_sev_soc[t] * surv_prob
    }
    
    # Transitions from "mil_i"
    p_mort <- a.TP["mil_i", "dth", t]
    surv_prob <- 1 - p_mort
    
    if (strat == "int") {
      a.TP["mil_i", "mil_i", t] <- v.p.mil_stay_int_dis[t] * surv_prob
      a.TP["mil_i", "mci_i", t] <- v.p.mil_mci_int_dis[t] * surv_prob
      a.TP["mil_i", "mod_i", t] <- v.p.mil_mod_int_dis[t] * surv_prob
      a.TP["mil_i", "sev_i", t] <- v.p.mil_sev_int_dis[t] * surv_prob
    } else {
      a.TP["mil_i", "mil_i", t] <- v.p.mil_stay_soc[t] * surv_prob
      a.TP["mil_i", "mci_i", t] <- v.p.mil_mci_soc[t] * surv_prob
      a.TP["mil_i", "mod_i", t] <- v.p.mil_mod_soc[t] * surv_prob
      a.TP["mil_i", "sev_i", t] <- v.p.mil_sev_soc[t] * surv_prob
    }
    
    # Transitions from "mod_i"
    p_mort <- a.TP["mod_i", "dth", t]
    surv_prob <- 1 - p_mort
    
    a.TP["mod_i", "mod_i", t] <- v.p.mod_stay_soc[t] * surv_prob
    a.TP["mod_i", "mil_i", t] <- v.p.mod_mil_soc[t] * surv_prob
    a.TP["mod_i", "sev_i", t] <- v.p.mod_sev_soc[t] * surv_prob
    
    # Transitions from "sev_i"
    p_mort <- a.TP["sev_i", "dth", t]
    surv_prob <- 1 - p_mort
    
    a.TP["sev_i", "sev_i", t] <- v.p.sev_stay_soc[t] * surv_prob
    a.TP["sev_i", "mil_i", t] <- v.p.sev_mil_soc[t] * surv_prob
    a.TP["sev_i", "mod_i", t] <- v.p.sev_mod_soc[t] * surv_prob
    
    # Normalize rows to ensure transition probabilities sum to 1
    for (s in v.names_state) {
      row_sum <- sum(a.TP[s, , t])
      if (row_sum > 0 && abs(row_sum - 1) > 1e-8) {
        a.TP[s, , t] <- a.TP[s, , t] / row_sum
      }
    }
  }
  
  return(a.TP)
}
# Function to compute Markov trace and total outcomes
compute_outcomes <- function(strat) {
  
  # Initialize structures
  a.TP <- build_transition_matrix(strat)
  v.states <- v.names_state
  n.states <- length(v.states)
  
  # Initialize Markov trace matrix
  m.trace <- matrix(0, nrow = n.cycles, ncol = n.states,
                    dimnames = list(NULL, v.states))
  
  # Set initial state distribution
  v.init <- setNames(rep(0, n.states), v.states)
  v.init["mciof_c"] <- 1 - p.starting_state_mci
  v.init["mcion_c"] <- p.starting_state_mci
  m.trace[1, ] <- v.init
  
  # Initialize utility and cost vectors (per state, named)
  v.utility <- setNames(c(
    u.mci_pt + u.mci_ic,   # mcion_c
    u.mci_pt + u.mci_ic,   # mciof_c
    u.mil_pt + u.mil_ic,   # milon_c
    u.mil_pt + u.mil_ic,   # milof_c
    u.mod_pt + u.mod_ic,   # mod_c
    u.sev_pt + u.sev_ic,   # sev_c
    u.mci_pt_i + u.mci_ic_i, # mci_i
    u.mil_pt_i + u.mil_ic_i, # mil_i
    u.mod_pt_i + u.mod_ic_i, # mod_i
    u.sev_pt_i + u.sev_ic_i, # sev_i
    0                       # dth
  ), v.states)
  
  v.cost <- setNames(c(
    c.mci_hc + c.mci_sc + c.mci_ic,       # mcion_c
    c.mci_hc + c.mci_sc + c.mci_ic,       # mciof_c
    c.mil_hc + c.mil_sc + c.mil_ic,       # milon_c
    c.mil_hc + c.mil_sc + c.mil_ic,       # milof_c
    c.mod_hc + c.mod_sc + c.mod_ic,       # mod_c
    c.sev_hc + c.sev_sc + c.sev_ic,       # sev_c
    c.mci_i_hc + c.mci_i_sc + c.mci_i_ic, # mci_i
    c.mil_i_hc + c.mil_i_sc + c.mil_i_ic, # mil_i
    c.mod_i_hc + c.mod_i_sc + c.mod_i_ic, # mod_i
    c.sev_i_hc + c.sev_i_sc + c.sev_i_ic, # sev_i
    0                                     # dth
  ), v.states)
  
  # Treatment-specific: add recurring treatment costs to on-treatment states
  if (strat == "int") {
    v.cost["mcion_c"] <- v.cost["mcion_c"] + c.Tx
    v.cost["milon_c"] <- v.cost["milon_c"] + c.Tx
  }
  
  # Initialize results for each cycle
  v.qalys <- numeric(n.cycles)
  v.costs <- numeric(n.cycles)
  v.lys <- numeric(n.cycles)
  
  # Compute outcomes per cycle
  for (t in 1:(n.cycles - 1)) {
    # Update state distribution
    m.trace[t + 1, ] <- m.trace[t, ] %*% a.TP[,,t]
    
    # Compute undiscounted outcomes
    v.qalys[t] <- sum(m.trace[t, ] * v.utility)
    v.costs[t] <- sum(m.trace[t, ] * v.cost)
    v.lys[t] <- sum(m.trace[t, v.states != "dth"])
  }
  
  # Final cycle
  v.qalys[n.cycles] <- sum(m.trace[n.cycles, ] * v.utility)
  v.costs[n.cycles] <- sum(m.trace[n.cycles, ] * v.cost)
  v.lys[n.cycles] <- sum(m.trace[n.cycles, v.states != "dth"])
  
  # Half-cycle correction (drop last cycle after averaging)
  v.qalys <- head((v.qalys[-1] + v.qalys[-n.cycles]) / 2, -1)
  v.costs <- head((v.costs[-1] + v.costs[-n.cycles]) / 2, -1)
  v.lys <- head((v.lys[-1] + v.lys[-n.cycles]) / 2, -1)
  
  # Apply one-time treatment utility gain and cost at cycle 1 for "int"
  if (strat == "int") {
    v.qalys[1] <- v.qalys[1] + u.Tx_start
    v.costs[1] <- v.costs[1] + c.Tx_start
  }
  
  # Apply discounting
  v.discount <- 1 / ((1 + discount_QALY)^(1:(n.cycles - 1)))
  qalys_total <- sum(v.qalys * v.discount)
  costs_total <- sum(v.costs * v.discount)
  lys_total <- sum(v.lys * v.discount)
  
  return(list(qalys = qalys_total, costs = costs_total, lys = lys_total))
}

# Run model for both strategies
results_soc <- compute_outcomes("soc")
results_int <- compute_outcomes("int")

# Print comparative results
cat("=== Total Outcomes Comparison ===\n")
cat(sprintf("Standard of Care (SOC):     QALYs = %.2f, Costs = $%.0f, LYs = %.2f\n",
            results_soc$qalys, results_soc$costs, results_soc$lys))
cat(sprintf("Intervention (INT):         QALYs = %.2f, Costs = $%.0f, LYs = %.2f\n",
            results_int$qalys, results_int$costs, results_int$lys))
cat(sprintf("Incremental (INT - SOC):    QALYs = %.2f, Costs = $%.0f, LYs = %.2f\n",
            results_int$qalys - results_soc$qalys,
            results_int$costs - results_soc$costs,
            results_int$lys - results_soc$lys))
```
