## Supplementary material for "Replicating Health-Economic Simulation Models for Alzheimer’s Disease Using Artificial Intelligence": final R scripts: o9.R

```
# ========================
# Input Parameters for IPECAD Markov Model
# Source: IPECAD Publication + Supplemental Material
# ========================

# --- Descriptive parameters ---
state_names <- c("mcion_c", "mciof_c", "milon_c", "milof_c", "mod_c", "sev_c",
                 "mci_i", "mil_i", "mod_i", "sev_i", "dth")
strategy_names <- c("soc", "int")

# --- Demographic and time parameters ---
age_start <- 71
sex <- "weighted"  # 52% female
p_starting_state_mci <- 0.55
n_cycle <- 29

# --- Transition probabilities (per year) ---
p_mci_mil <- 0.23
p_mci_mod <- 0
p_mci_sev <- 0
p_mil_mci <- 0.03
p_mil_mod <- 0.35
p_mil_sev <- 0.04
p_mod_mil <- 0.03
p_mod_sev <- 0.42
p_sev_mil <- 0
p_sev_mod <- 0.02

# --- Transitions to institution ---
p_mci_i <- 0.024
p_mil_i <- 0.038
p_mod_i <- 0.110
p_sev_i <- 0.259

# --- Mortality hazard ratios by state ---
hr_mort_mci <- 1.82
hr_mort_mil <- 2.92
hr_mort_mod <- 3.85
hr_mort_sev <- 9.52

# --- Relative risk of treatment effect (on treatment) ---
rr_tx_mci_mil <- 0.69
rr_tx_mci_mod <- 1
rr_tx_mci_sev <- 1
rr_tx_mil_mod <- 0.69
rr_tx_mil_sev <- 0.69

# --- Relative risk of treatment effect (off treatment) ---
rr_tx_mci_mil_dis <- 1
rr_tx_mci_mod_dis <- 1
rr_tx_mci_sev_dis <- 1
rr_tx_mil_mod_dis <- 1
rr_tx_mil_sev_dis <- 1

# --- Treatment discontinuation ---
p_discontinuation1 <- 0.069
p_discontinuation2 <- 0
discontinuation2_begin <- 2
tx_duration <- 29

# --- Treatment effect waning (no waning assumed) ---
tx_waning <- 0
tx_waning_dis <- 0

# --- Utilities (patient, community setting) ---
u_mci_pt <- 0.851 - 0.17
u_mil_pt <- 0.851 - 0.22
u_mod_pt <- 0.851 - 0.36
u_sev_pt <- 0.851 - 0.53

# --- Utilities (patient, institution setting) ---
u_mci_pt_i <- 0.851 - 0.17
u_mil_pt_i <- 0.851 - 0.19
u_mod_pt_i <- 0.851 - 0.42
u_sev_pt_i <- 0.851 - 0.59

# --- Utilities (caregiver, community setting) ---
u_mci_ic <- -0.03
u_mil_ic <- -0.05
u_mod_ic <- -0.08
u_sev_ic <- -0.10

# --- Utilities (caregiver, institution setting) ---
u_mci_ic_i <- -0.03
u_mil_ic_i <- -0.05
u_mod_ic_i <- -0.08
u_sev_ic_i <- -0.10

# --- Additional utility in first cycle due to ARIA (not half-cycle corrected) ---
u_Tx_start <- -0.14 * (12 / 52) * 0.035

# --- Costs (USD) per year ---
# Direct medical + informal care + social care, per setting

# Community setting costs
c_mci_hc <- 6042 * 1.12 + 460
c_mil_hc <- 6042 * 1.56 + 965 + 0.21 * 365 * 0.333
c_mod_hc <- 6042 * 1.93 + 1544 + 0.66 * 365 * 0.333
c_sev_hc <- 6042 * 1.93 + 1930

c_mci_sc <- 0
c_mil_sc <- 0
c_mod_sc <- 0
c_sev_sc <- 0

c_mci_ic <- 69 * 12 * 32.46 + 0.204 * 0.049 * 20 * 52 * 32.46
c_mil_ic <- 113 * 12 * 32.46 + 0.112 * 0.086 * 20 * 52 * 32.46
c_mod_ic <- 169 * 12 * 32.46
c_sev_ic <- 298 * 12 * 32.46

# Institution setting costs
c_mci_i_hc <- c_mci_hc
c_mil_i_hc <- c_mil_hc
c_mod_i_hc <- c_mod_hc
c_sev_i_hc <- c_sev_hc

c_mci_i_sc <- 7394 * 12
c_mil_i_sc <- 7394 * 12
c_mod_i_sc <- 7394 * 12
c_sev_i_sc <- 7394 * 12

c_mci_i_ic <- 69 * 12 * 32.46 * 0.44 + 0.204 * 0.049 * 20 * 52 * 32.46
c_mil_i_ic <- 113 * 12 * 32.46 * 0.44 + 0.112 * 0.086 * 20 * 52 * 32.46
c_mod_i_ic <- 169 * 12 * 32.46 * 0.44
c_sev_i_ic <- 298 * 12 * 32.46 * 0.44

# --- Treatment costs ---
c_Tx <- 26500 + (52 / 2) * 78.35
c_Tx_start <- 261.10 * 4 + 261.10 * 3 * 0.215

# --- Discount rates ---
discount_EFFECT <- 0.03
discount_QALY <- 0.03
discount_COST <- 0.03

# --- Economic evaluation threshold ---
wtp <- 100000

# --- Model control ---
half_cycle_correction <- TRUE
# Load the 2019 US life table CSV file
lifetable <- read.csv("lifetable_US_2019_ssa.csv")

# Convert annual mortality probabilities to annual mortality rates
# Formula: rate = -log(1 - prob)
lifetable$male <- -log(1 - lifetable$male)
lifetable$female <- -log(1 - lifetable$female)

# Compute sex-weighted average mortality rate (52% female, 48% male)
lifetable$weighted <- 0.48 * lifetable$male + 0.52 * lifetable$female

# Return the resulting dataframe with specified columns
lifetable <- lifetable[, c("age", "male", "female", "weighted")]

# View the result (optional)
# head(lifetable)
# Number of cycles and starting age
v.n_cycle <- n_cycle
v.age <- age_start + 0:(v.n_cycle - 1)

# --- Disease progression probabilities for standard of care (time-independent) ---
v.p.mci_mil_soc <- rep(p_mci_mil, v.n_cycle)
v.p.mci_mod_soc <- rep(p_mci_mod, v.n_cycle)
v.p.mci_sev_soc <- rep(p_mci_sev, v.n_cycle)
v.p.mil_mci_soc <- rep(p_mil_mci, v.n_cycle)
v.p.mil_mod_soc <- rep(p_mil_mod, v.n_cycle)
v.p.mil_sev_soc <- rep(p_mil_sev, v.n_cycle)
v.p.mod_mil_soc <- rep(p_mod_mil, v.n_cycle)
v.p.mod_sev_soc <- rep(p_mod_sev, v.n_cycle)
v.p.sev_mil_soc <- rep(p_sev_mil, v.n_cycle)
v.p.sev_mod_soc <- rep(p_sev_mod, v.n_cycle)

# --- Staying probabilities (exclude institutional transitions) ---
v.p.mci_mci_soc <- 1 - (v.p.mci_mil_soc + v.p.mci_mod_soc + v.p.mci_sev_soc)
v.p.mil_mil_soc <- 1 - (v.p.mil_mci_soc + v.p.mil_mod_soc + v.p.mil_sev_soc)
v.p.mod_mod_soc <- 1 - (v.p.mod_mil_soc + v.p.mod_sev_soc)
v.p.sev_sev_soc <- 1 - (v.p.sev_mil_soc + v.p.sev_mod_soc)

# --- Institutionalization probabilities (time-independent) ---
v.p.mci_inst <- rep(p_mci_i, v.n_cycle)
v.p.mil_inst <- rep(p_mil_i, v.n_cycle)
v.p.mod_inst <- rep(p_mod_i, v.n_cycle)
v.p.sev_inst <- rep(p_sev_i, v.n_cycle)

# --- Discontinuation probabilities per cycle ---
v.p_discontinuation <- numeric(v.n_cycle)
v.p_discontinuation[1] <- p_discontinuation1
v.p_discontinuation[2:(discontinuation2_begin - 1)] <- p_discontinuation1
v.p_discontinuation[discontinuation2_begin:tx_duration] <- p_discontinuation2
if (tx_duration < v.n_cycle) {
  v.p_discontinuation[(tx_duration + 1):v.n_cycle] <- 1
}

# --- Mortality rates by age and sex from pre-loaded life table ---
# Ensure that v.age exists in the life table age column
v.mort_rate <- rep(NA, v.n_cycle)
for (i in 1:v.n_cycle) {
  age_i <- v.age[i]
  v.mort_rate[i] <- lifetable$weighted[lifetable$age == age_i]
}
# Initialize empty vectors
v.p.mci_mil_int_on  <- numeric(v.n_cycle)
v.p.mci_mod_int_on  <- numeric(v.n_cycle)
v.p.mci_sev_int_on  <- numeric(v.n_cycle)
v.p.mil_mci_int_on  <- rep(p_mil_mci, v.n_cycle)  # no treatment effect
v.p.mil_mod_int_on  <- numeric(v.n_cycle)
v.p.mil_sev_int_on  <- numeric(v.n_cycle)

v.p.mci_mil_int_off <- numeric(v.n_cycle)
v.p.mci_mod_int_off <- numeric(v.n_cycle)
v.p.mci_sev_int_off <- numeric(v.n_cycle)
v.p.mil_mci_int_off <- rep(p_mil_mci, v.n_cycle)  # no treatment effect
v.p.mil_mod_int_off <- numeric(v.n_cycle)
v.p.mil_sev_int_off <- numeric(v.n_cycle)

# Define RR and waning parameters
RR_mci_mil      <- rr_tx_mci_mil
RR_mci_mod      <- rr_tx_mci_mod
RR_mci_sev      <- rr_tx_mci_sev
RR_mil_mod      <- rr_tx_mil_mod
RR_mil_sev      <- rr_tx_mil_sev

RR_mci_mil_dis  <- rr_tx_mci_mil_dis
RR_mci_mod_dis  <- rr_tx_mci_mod_dis
RR_mci_sev_dis  <- rr_tx_mci_sev_dis
RR_mil_mod_dis  <- rr_tx_mil_mod_dis
RR_mil_sev_dis  <- rr_tx_mil_sev_dis

waning_on       <- tx_waning
waning_off      <- tx_waning_dis

# Compute time-dependent transition probabilities for intervention strategy
for (i in 1:v.n_cycle) {
  waning_factor_on  <- (1 - waning_on)^(i - 1)
  waning_factor_off <- (1 - waning_off)^(i - 1)
  
  # On-treatment (mcion_c, milon_c)
  rate_mci_mil  <- -log(1 - p_mci_mil)
  rate_mil_mod  <- -log(1 - p_mil_mod)
  rate_mil_sev  <- -log(1 - p_mil_sev)
  
  v.p.mci_mil_int_on[i] <- 1 - exp(-rate_mci_mil * RR_mci_mil^waning_factor_on)
  v.p.mci_mod_int_on[i] <- 1 - exp(-0)  # p_mci_mod = 0
  v.p.mci_sev_int_on[i] <- 1 - exp(-0)  # p_mci_sev = 0
  v.p.mil_mod_int_on[i] <- 1 - exp(-rate_mil_mod * RR_mil_mod^waning_factor_on)
  v.p.mil_sev_int_on[i] <- 1 - exp(-rate_mil_sev * RR_mil_sev^waning_factor_on)
  
  # Off-treatment (mciof_c, milof_c)
  v.p.mci_mil_int_off[i] <- 1 - exp(-rate_mci_mil * RR_mci_mil_dis^waning_factor_off)
  v.p.mci_mod_int_off[i] <- 1 - exp(-0)  # p_mci_mod = 0
  v.p.mci_sev_int_off[i] <- 1 - exp(-0)  # p_mci_sev = 0
  v.p.mil_mod_int_off[i] <- 1 - exp(-rate_mil_mod * RR_mil_mod_dis^waning_factor_off)
  v.p.mil_sev_int_off[i] <- 1 - exp(-rate_mil_sev * RR_mil_sev_dis^waning_factor_off)
}

# Staying probabilities (excluding institutional transitions)
v.p.mci_mci_int_on  <- 1 - (v.p.mci_mil_int_on + v.p.mci_mod_int_on + v.p.mci_sev_int_on)
v.p.mil_mil_int_on  <- 1 - (v.p.mil_mci_int_on + v.p.mil_mod_int_on + v.p.mil_sev_int_on)

v.p.mci_mci_int_off <- 1 - (v.p.mci_mil_int_off + v.p.mci_mod_int_off + v.p.mci_sev_int_off)
v.p.mil_mil_int_off <- 1 - (v.p.mil_mci_int_off + v.p.mil_mod_int_off + v.p.mil_sev_int_off)

# Function to build transition probability array for a given strategy ("soc" or "int")
build_transition_matrix <- function(strat) {
  
  # Initialize transition probability array (states x states x cycles)
  a.TP <- array(0, dim = c(length(state_names), length(state_names), v.n_cycle),
                dimnames = list(state_names, state_names, NULL))
  
  for (t in 1:v.n_cycle) {
    
    # --- Transition to death from each state (cycle t) ---
    # Mortality rate at cycle t
    mort_rate <- v.mort_rate[t]
    
    a.TP["mcion_c", "dth", t]  <- 1 - exp(-mort_rate * hr_mort_mci)
    a.TP["mciof_c", "dth", t]  <- 1 - exp(-mort_rate * hr_mort_mci)
    a.TP["milon_c", "dth", t]  <- 1 - exp(-mort_rate * hr_mort_mil)
    a.TP["milof_c", "dth", t]  <- 1 - exp(-mort_rate * hr_mort_mil)
    a.TP["mod_c",  "dth", t]   <- 1 - exp(-mort_rate * hr_mort_mod)
    a.TP["sev_c",  "dth", t]   <- 1 - exp(-mort_rate * hr_mort_sev)
    a.TP["mci_i",  "dth", t]   <- 1 - exp(-mort_rate * hr_mort_mci)
    a.TP["mil_i",  "dth", t]   <- 1 - exp(-mort_rate * hr_mort_mil)
    a.TP["mod_i",  "dth", t]   <- 1 - exp(-mort_rate * hr_mort_mod)
    a.TP["sev_i",  "dth", t]   <- 1 - exp(-mort_rate * hr_mort_sev)
    
    # Death is absorbing state
    a.TP["dth", "dth", t] <- 1
    
    # --- Transitions from mcion_c ---
    if (strat == "soc") {
      p_mci_mil <- v.p.mci_mil_soc[t]
      p_mci_mci <- v.p.mci_mci_soc[t]
    } else {
      p_mci_mil <- v.p.mci_mil_int_on[t]
      p_mci_mci <- v.p.mci_mci_int_on[t]
    }
    
    p_inst_mci <- v.p.mci_inst[t]
    p_discont  <- v.p_discontinuation[t]
    p_surv     <- 1 - a.TP["mcion_c", "dth", t]
    
    # Stay in mcion_c (on-treatment)
    a.TP["mcion_c", "mcion_c", t] <- p_mci_mci * (1 - p_inst_mci) * (1 - p_discont) * p_surv
    
    # Progress to milon_c (on-treatment)
    a.TP["mcion_c", "milon_c", t] <- p_mci_mil * (1 - p_inst_mci) * (1 - p_discont) * p_surv
    
    # Discontinue and stay in mciof_c (off-treatment)
    a.TP["mcion_c", "mciof_c", t] <- p_mci_mci * (1 - p_inst_mci) * p_discont * p_surv
    
    # Discontinue and move to milof_c (off-treatment)
    a.TP["mcion_c", "milof_c", t] <- p_mci_mil * (1 - p_inst_mci) * p_discont * p_surv
    
    # Move to mod_c (off-treatment, no discontinuation applied)
    a.TP["mcion_c", "mod_c", t] <- v.p.mci_mod_soc[t] * (1 - p_inst_mci) * p_surv
    
    # Move to sev_c (off-treatment, no discontinuation applied)
    a.TP["mcion_c", "sev_c", t] <- v.p.mci_sev_soc[t] * (1 - p_inst_mci) * p_surv
    
    # Transition to institutionalized state mci_i (only from mcion_c)
    a.TP["mcion_c", "mci_i", t] <- (p_mci_mci + p_mci_mil + v.p.mci_mod_soc[t] + v.p.mci_sev_soc[t]) * p_inst_mci * p_surv
    
    # --- Transitions from milon_c ---
    if (strat == "soc") {
      p_mil_mod <- v.p.mil_mod_soc[t]
      p_mil_mil <- v.p.mil_mil_soc[t]
      p_mil_mci <- v.p.mil_mci_soc[t]
      p_mil_sev <- v.p.mil_sev_soc[t]
    } else {
      p_mil_mod <- v.p.mil_mod_int_on[t]
      p_mil_mil <- v.p.mil_mil_int_on[t]
      p_mil_mci <- v.p.mil_mci_int_on[t]
      p_mil_sev <- v.p.mil_sev_int_on[t]
    }
    
    p_inst_mil <- v.p.mil_inst[t]
    p_surv     <- 1 - a.TP["milon_c", "dth", t]
    
    # Stay in milon_c (on-treatment)
    a.TP["milon_c", "milon_c", t] <- p_mil_mil * (1 - p_inst_mil) * (1 - p_discont) * p_surv
    
    # Back-transition to mciof_c (off-treatment)
    a.TP["milon_c", "mciof_c", t] <- p_mil_mci * (1 - p_inst_mil) * p_discont * p_surv
    
    # Stay in milof_c (off-treatment)
    a.TP["milon_c", "milof_c", t] <- p_mil_mil * (1 - p_inst_mil) * p_discont * p_surv
    
    # Forward transition to mod_c (off-treatment)
    a.TP["milon_c", "mod_c", t] <- p_mil_mod * (1 - p_inst_mil) * p_surv
    
    # Forward transition to sev_c (off-treatment)
    a.TP["milon_c", "sev_c", t] <- p_mil_sev * (1 - p_inst_mil) * p_surv
    
    # Transition to institutionalized state mil_i
    a.TP["milon_c", "mil_i", t] <- (p_mil_mci + p_mil_mil + p_mil_mod + p_mil_sev) * p_inst_mil * p_surv
    
    
    # --- Transitions from mciof_c ---
    if (strat == "soc") {
      p_mci_mil <- v.p.mci_mil_soc[t]
      p_mci_mod <- v.p.mci_mod_soc[t]
      p_mci_sev <- v.p.mci_sev_soc[t]
      p_mci_mci <- v.p.mci_mci_soc[t]
    } else {
      p_mci_mil <- v.p.mci_mil_int_off[t]
      p_mci_mod <- v.p.mci_mod_int_off[t]
      p_mci_sev <- v.p.mci_sev_int_off[t]
      p_mci_mci <- v.p.mci_mci_int_off[t]
    }
    
    p_inst_mci <- v.p.mci_inst[t]
    p_surv     <- 1 - a.TP["mciof_c", "dth", t]
    
    a.TP["mciof_c", "mciof_c", t] <- p_mci_mci * (1 - p_inst_mci) * p_surv
    a.TP["mciof_c", "milof_c", t] <- p_mci_mil * (1 - p_inst_mci) * p_surv
    a.TP["mciof_c", "mod_c", t]   <- p_mci_mod * (1 - p_inst_mci) * p_surv
    a.TP["mciof_c", "sev_c", t]   <- p_mci_sev * (1 - p_inst_mci) * p_surv
    a.TP["mciof_c", "mci_i", t]   <- (p_mci_mci + p_mci_mil + p_mci_mod + p_mci_sev) * p_inst_mci * p_surv
    
    # --- Transitions from milof_c ---
    if (strat == "soc") {
      p_mil_mci <- v.p.mil_mci_soc[t]
      p_mil_mod <- v.p.mil_mod_soc[t]
      p_mil_sev <- v.p.mil_sev_soc[t]
      p_mil_mil <- v.p.mil_mil_soc[t]
    } else {
      p_mil_mci <- v.p.mil_mci_int_off[t]
      p_mil_mod <- v.p.mil_mod_int_off[t]
      p_mil_sev <- v.p.mil_sev_int_off[t]
      p_mil_mil <- v.p.mil_mil_int_off[t]
    }
    
    p_inst_mil <- v.p.mil_inst[t]
    p_surv     <- 1 - a.TP["milof_c", "dth", t]
    
    a.TP["milof_c", "mciof_c", t] <- p_mil_mci * (1 - p_inst_mil) * p_surv
    a.TP["milof_c", "milof_c", t] <- p_mil_mil * (1 - p_inst_mil) * p_surv
    a.TP["milof_c", "mod_c", t]   <- p_mil_mod * (1 - p_inst_mil) * p_surv
    a.TP["milof_c", "sev_c", t]   <- p_mil_sev * (1 - p_inst_mil) * p_surv
    a.TP["milof_c", "mil_i", t]   <- (p_mil_mci + p_mil_mil + p_mil_mod + p_mil_sev) * p_inst_mil * p_surv
    
    # --- Transitions from mod_c ---
    p_mod_mil <- v.p.mod_mil_soc[t]
    p_mod_sev <- v.p.mod_sev_soc[t]
    p_mod_mod <- v.p.mod_mod_soc[t]
    p_inst_mod <- v.p.mod_inst[t]
    p_surv     <- 1 - a.TP["mod_c", "dth", t]
    
    a.TP["mod_c", "milof_c", t] <- p_mod_mil * (1 - p_inst_mod) * p_surv
    a.TP["mod_c", "mod_c", t]   <- p_mod_mod * (1 - p_inst_mod) * p_surv
    a.TP["mod_c", "sev_c", t]   <- p_mod_sev * (1 - p_inst_mod) * p_surv
    a.TP["mod_c", "mod_i", t]   <- (p_mod_mil + p_mod_mod + p_mod_sev) * p_inst_mod * p_surv
    
    # --- Transitions from sev_c ---
    p_sev_mil <- v.p.sev_mil_soc[t]
    p_sev_mod <- v.p.sev_mod_soc[t]
    p_sev_sev <- v.p.sev_sev_soc[t]
    p_inst_sev <- v.p.sev_inst[t]
    p_surv     <- 1 - a.TP["sev_c", "dth", t]
    
    a.TP["sev_c", "milof_c", t] <- p_sev_mil * (1 - p_inst_sev) * p_surv
    a.TP["sev_c", "mod_c", t]   <- p_sev_mod * (1 - p_inst_sev) * p_surv
    a.TP["sev_c", "sev_c", t]   <- p_sev_sev * (1 - p_inst_sev) * p_surv
    a.TP["sev_c", "sev_i", t]   <- (p_sev_mil + p_sev_mod + p_sev_sev) * p_inst_sev * p_surv
    
    # --- Transitions from mci_i ---
    p_mci_mil <- if (strat == "soc") v.p.mci_mil_soc[t] else v.p.mci_mil_int_off[t]
    p_mci_mod <- if (strat == "soc") v.p.mci_mod_soc[t] else v.p.mci_mod_int_off[t]
    p_mci_sev <- if (strat == "soc") v.p.mci_sev_soc[t] else v.p.mci_sev_int_off[t]
    p_mci_mci <- if (strat == "soc") v.p.mci_mci_soc[t] else v.p.mci_mci_int_off[t]
    p_surv    <- 1 - a.TP["mci_i", "dth", t]
    
    a.TP["mci_i", "mci_i", t] <- p_mci_mci * p_surv
    a.TP["mci_i", "mil_i", t] <- p_mci_mil * p_surv
    a.TP["mci_i", "mod_i", t] <- p_mci_mod * p_surv
    a.TP["mci_i", "sev_i", t] <- p_mci_sev * p_surv
    
    # --- Transitions from mil_i ---
    p_mil_mci <- if (strat == "soc") v.p.mil_mci_soc[t] else v.p.mil_mci_int_off[t]
    p_mil_mod <- if (strat == "soc") v.p.mil_mod_soc[t] else v.p.mil_mod_int_off[t]
    p_mil_sev <- if (strat == "soc") v.p.mil_sev_soc[t] else v.p.mil_sev_int_off[t]
    p_mil_mil <- if (strat == "soc") v.p.mil_mil_soc[t] else v.p.mil_mil_int_off[t]
    p_surv    <- 1 - a.TP["mil_i", "dth", t]
    
    a.TP["mil_i", "mci_i", t] <- p_mil_mci * p_surv
    a.TP["mil_i", "mil_i", t] <- p_mil_mil * p_surv
    a.TP["mil_i", "mod_i", t] <- p_mil_mod * p_surv
    a.TP["mil_i", "sev_i", t] <- p_mil_sev * p_surv
    
    # --- Transitions from mod_i ---
    p_mod_mil <- v.p.mod_mil_soc[t]
    p_mod_sev <- v.p.mod_sev_soc[t]
    p_mod_mod <- v.p.mod_mod_soc[t]
    p_surv    <- 1 - a.TP["mod_i", "dth", t]
    
    a.TP["mod_i", "mil_i", t] <- p_mod_mil * p_surv
    a.TP["mod_i", "mod_i", t] <- p_mod_mod * p_surv
    a.TP["mod_i", "sev_i", t] <- p_mod_sev * p_surv
    
    # --- Transitions from sev_i ---
    p_sev_mil <- v.p.sev_mil_soc[t]
    p_sev_mod <- v.p.sev_mod_soc[t]
    p_sev_sev <- v.p.sev_sev_soc[t]
    p_surv    <- 1 - a.TP["sev_i", "dth", t]
    
    a.TP["sev_i", "mil_i", t] <- p_sev_mil * p_surv
    a.TP["sev_i", "mod_i", t] <- p_sev_mod * p_surv
    a.TP["sev_i", "sev_i", t] <- p_sev_sev * p_surv
    
    # --- Normalize rows (except for absorbing state "dth") ---
    for (s in state_names[-which(state_names == "dth")]) {
      row_sum <- sum(a.TP[s, , t])
      if (row_sum > 0 && abs(row_sum - 1) > 1e-8) {
        a.TP[s, , t] <- a.TP[s, , t] / row_sum
      }
    }
  }
  
  return(a.TP)
}


# Function to compute Markov trace and total outcomes for a strategy ("soc" or "int")
compute_outcomes <- function(strat) {
  
  # Get transition probability array
  a.TP <- build_transition_matrix(strat)
  
  # Initialize Markov trace matrix (cycles x states)
  m.M <- matrix(0, nrow = v.n_cycle, ncol = length(state_names), dimnames = list(NULL, state_names))
  
  # Initial state distribution
  m.M[1, ] <- 0
  m.M[1, "mciof_c"] <- 1 - p_starting_state_mci  # mild dementia off-treatment
  m.M[1, "mcion_c"] <- p_starting_state_mci      # MCI on-treatment
  
  # Utility vector (named)
  v.utility <- c(
    mcion_c = u_mci_pt,  mciof_c = u_mci_pt,
    milon_c = u_mil_pt,  milof_c = u_mil_pt,
    mod_c   = u_mod_pt,  sev_c   = u_sev_pt,
    mci_i   = u_mci_pt_i, mil_i  = u_mil_pt_i,
    mod_i   = u_mod_pt_i, sev_i  = u_sev_pt_i,
    dth     = 0
  )
  
  # Cost vector (named, excluding treatment cost)
  v.cost <- c(
    mcion_c = c_mci_hc + c_mci_sc + c_mci_ic,
    mciof_c = c_mci_hc + c_mci_sc + c_mci_ic,
    milon_c = c_mil_hc + c_mil_sc + c_mil_ic,
    milof_c = c_mil_hc + c_mil_sc + c_mil_ic,
    mod_c   = c_mod_hc + c_mod_sc + c_mod_ic,
    sev_c   = c_sev_hc + c_sev_sc + c_sev_ic,
    mci_i   = c_mci_i_hc + c_mci_i_sc + c_mci_i_ic,
    mil_i   = c_mil_i_hc + c_mil_i_sc + c_mil_i_ic,
    mod_i   = c_mod_i_hc + c_mod_i_sc + c_mod_i_ic,
    sev_i   = c_sev_i_hc + c_sev_i_sc + c_sev_i_ic,
    dth     = 0
  )
  
  # Initialize outcomes
  v.QALYs <- numeric(v.n_cycle)
  v.Costs <- numeric(v.n_cycle)
  v.LYs   <- numeric(v.n_cycle)
  
  # Apply treatment cost vector (recurrent) for "int"
  if (strat == "int") {
    treat_states <- c("mcion_c", "milon_c")
    for (s in treat_states) {
      v.cost[s] <- v.cost[s] + c_Tx
    }
  }
  
  # Markov trace updates and outcome calculations
  for (t in 2:v.n_cycle) {
    m.M[t, ] <- m.M[t - 1, ] %*% a.TP[, , t - 1]
  }
  
  for (t in 1:v.n_cycle) {
    v.QALYs[t] <- sum(m.M[t, ] * v.utility)
    v.Costs[t] <- sum(m.M[t, ] * v.cost)
    v.LYs[t]   <- sum(m.M[t, state_names[state_names != "dth"]])
  }
  
  # Half-cycle correction
  v.QALYs <- 0.5 * (v.QALYs[-v.n_cycle] + v.QALYs[-1])
  v.Costs <- 0.5 * (v.Costs[-v.n_cycle] + v.Costs[-1])
  v.LYs   <- 0.5 * (v.LYs[-v.n_cycle] + v.LYs[-1])
  
  # Apply initial treatment cost and disutility only for intervention
  if (strat == "int") {
    v.QALYs[1] <- v.QALYs[1] + u_Tx_start
    v.Costs[1] <- v.Costs[1] + c_Tx_start
  }
  
  # Apply discounting
  v.discount <- 1 / ((1 + discount_QALY) ^ (1:length(v.QALYs)))
  total_QALYs <- sum(v.QALYs * v.discount)
  total_LYs   <- sum(v.LYs * v.discount)
  
  v.discount_cost <- 1 / ((1 + discount_COST) ^ (1:length(v.Costs)))
  total_Costs <- sum(v.Costs * v.discount_cost)
  
  return(c(QALYs = total_QALYs, Costs = total_Costs, LYs = total_LYs))
}

# Run both strategies
results_soc <- compute_outcomes("soc")
results_int <- compute_outcomes("int")

# Print comparison results
cat("Strategy Comparison:\n")
cat(sprintf("QALYs (soc): %.3f | QALYs (int): %.3f | ΔQALYs: %.3f\n", results_soc["QALYs"], results_int["QALYs"], results_int["QALYs"] - results_soc["QALYs"]))
cat(sprintf("Costs  (soc): $%.2f | Costs  (int): $%.2f | ΔCosts : $%.2f\n", results_soc["Costs"], results_int["Costs"], results_int["Costs"] - results_soc["Costs"]))
cat(sprintf("LYs    (soc): %.3f | LYs    (int): %.3f | ΔLYs   : %.3f\n", results_soc["LYs"], results_int["LYs"], results_int["LYs"] - results_soc["LYs"]))
```
