## Supplementary material for "Replicating Health-Economic Simulation Models for Alzheimer’s Disease Using Artificial Intelligence": final R scripts: o10.R

```
# Load required package
library(dampack)

# Define model states and strategies
state_names <- c("mcion_c", "mciof_c", "milon_c", "milof_c", 
                 "mod_c", "sev_c", "mci_i", "mil_i", "mod_i", 
                 "sev_i", "dth")
strategy_names <- c("soc", "int")

# General model parameters
age_start <- 71
sex <- "weighted"  # Used for life table mortality, 52% female
p_starting_state_mci <- 0.55
n_cycles <- 29
half_cycle_correction <- TRUE
discount_EFFECT <- 0.03
discount_QALY <- 0.03
discount_COST <- 0.03
wtp <- 100000

# Transition probabilities
p.mci_mil <- 0.23
p.mci_mod <- 0
p.mci_sev <- 0
p.mil_mci <- 0.03
p.mil_mod <- 0.35
p.mil_sev <- 0.04
p.mod_mil <- 0.03
p.mod_sev <- 0.42
p.sev_mil <- 0
p.sev_mod <- 0.02

# Institutionalization
p.mci_i <- 0.024
p.mil_i <- 0.038
p.mod_i <- 0.110
p.sev_i <- 0.259

# Hazard ratios for mortality
hr.mort_mci <- 1.82
hr.mort_mil <- 2.92
hr.mort_mod <- 3.85
hr.mort_sev <- 9.52

# Treatment effect (relative risk)
rr.tx_mci_mil <- 0.69
rr.tx_mci_mod <- 1
rr.tx_mci_sev <- 1
rr.tx_mil_mod <- 0.69
rr.tx_mil_sev <- 0.69

# Treatment effect after discontinuation
rr.tx_mci_mil_dis <- 1
rr.tx_mci_mod_dis <- 1
rr.tx_mci_sev_dis <- 1
rr.tx_mil_mod_dis <- 1
rr.tx_mil_sev_dis <- 1

# Discontinuation
p.discontinuation1 <- 0.069
p.discontinuation2 <- 0
discontinuation2_begin <- 2
tx_duration <- 29
tx_waning <- 0
tx_waning_dis <- 0

# Utilities - patient (community)
u.mci_pt <- 0.851 - 0.17
u.mil_pt <- 0.851 - 0.22
u.mod_pt <- 0.851 - 0.36
u.sev_pt <- 0.851 - 0.53

# Utilities - patient (institution)
u.mci_pt_i <- 0.851 - 0.17
u.mil_pt_i <- 0.851 - 0.19
u.mod_pt_i <- 0.851 - 0.42
u.sev_pt_i <- 0.851 - 0.59

# Utilities - caregiver (community)
u.mci_ic <- -0.03
u.mil_ic <- -0.05
u.mod_ic <- -0.08
u.sev_ic <- -0.10

# Utilities - caregiver (institution)
u.mci_ic_i <- -0.03
u.mil_ic_i <- -0.05
u.mod_ic_i <- -0.08
u.sev_ic_i <- -0.10

# ARIA disutility (first cycle only)
u.Tx_start <- -0.14 * (12 / 52) * 0.035

# Costs - health care (community)
c.mci_hc <- 6042 * 1.12 + 460
c.mil_hc <- 6042 * 1.56 + 965 + 0.21 * 365 * 0.333
c.mod_hc <- 6042 * 1.93 + 1544 + 0.66 * 365 * 0.333
c.sev_hc <- 6042 * 1.93 + 1930

# Costs - health care (institution)
c.mci_i_hc <- c.mci_hc
c.mil_i_hc <- c.mil_hc
c.mod_i_hc <- c.mod_hc
c.sev_i_hc <- c.sev_hc

# Costs - social care (community)
c.mci_sc <- 0
c.mil_sc <- 0
c.mod_sc <- 0
c.sev_sc <- 0

# Costs - social care (institution)
c.mci_i_sc <- 7394 * 12
c.mil_i_sc <- 7394 * 12
c.mod_i_sc <- 7394 * 12
c.sev_i_sc <- 7394 * 12

# Costs - informal care (community)
c.mci_ic <- 69 * 12 * 32.46 + 0.204 * 0.049 * 20 * 52 * 32.46
c.mil_ic <- 113 * 12 * 32.46 + 0.112 * 0.086 * 20 * 52 * 32.46
c.mod_ic <- 169 * 12 * 32.46
c.sev_ic <- 298 * 12 * 32.46

# Costs - informal care (institution)
c.mci_i_ic <- 69 * 12 * 32.46 * 0.44 + 0.204 * 0.049 * 20 * 52 * 32.46
c.mil_i_ic <- 113 * 12 * 32.46 * 0.44 + 0.112 * 0.086 * 20 * 52 * 32.46
c.mod_i_ic <- 169 * 12 * 32.46 * 0.44
c.sev_i_ic <- 298 * 12 * 32.46 * 0.44

# Treatment cost
c.Tx <- 26500 + (52 / 2) * 78.35

# Additional costs in first cycle
c.Tx_start <- 261.10 * 4 + 261.10 * 3 * 0.215
# Load life table
life_table <- read.csv("lifetable_US_2019_ssa.csv")

# Convert annual mortality probabilities to rates using: rate = -log(1 - probability)
life_table$male <- -log(1 - life_table$male)
life_table$female <- -log(1 - life_table$female)

# Compute weighted average of mortality rates (52% female, 48% male)
life_table$weighted <- 0.48 * life_table$male + 0.52 * life_table$female

# Keep only required columns
life_table <- life_table[, c("age", "male", "female", "weighted")]

# Output the processed life table
life_table
# Number of cycles
n_cycles <- 29

# --- Disease progression transition probability vectors under SOC strategy (time-independent) ---
v.p.mci_mil_soc <- rep(p.mci_mil, n_cycles)
v.p.mci_mod_soc <- rep(p.mci_mod, n_cycles)
v.p.mci_sev_soc <- rep(p.mci_sev, n_cycles)

v.p.mil_mci_soc <- rep(p.mil_mci, n_cycles)
v.p.mil_mod_soc <- rep(p.mil_mod, n_cycles)
v.p.mil_sev_soc <- rep(p.mil_sev, n_cycles)

v.p.mod_mil_soc <- rep(p.mod_mil, n_cycles)
v.p.mod_sev_soc <- rep(p.mod_sev, n_cycles)

v.p.sev_mil_soc <- rep(p.sev_mil, n_cycles)
v.p.sev_mod_soc <- rep(p.sev_mod, n_cycles)

# --- Stay probabilities (not including death or institutional transitions) ---
v.p.mci_stay_soc <- 1 - v.p.mci_mil_soc - v.p.mci_mod_soc - v.p.mci_sev_soc
v.p.mil_stay_soc <- 1 - v.p.mil_mci_soc - v.p.mil_mod_soc - v.p.mil_sev_soc
v.p.mod_stay_soc <- 1 - v.p.mod_mil_soc - v.p.mod_sev_soc
v.p.sev_stay_soc <- 1 - v.p.sev_mil_soc - v.p.sev_mod_soc

# --- Institutionalization transition probability vectors (time-independent) ---
v.p.mci_to_inst_soc <- rep(p.mci_i, n_cycles)
v.p.mil_to_inst_soc <- rep(p.mil_i, n_cycles)
v.p.mod_to_inst_soc <- rep(p.mod_i, n_cycles)
v.p.sev_to_inst_soc <- rep(p.sev_i, n_cycles)

# --- Discontinuation vector per cycle (non-cumulative) ---
v.p_discontinuation_soc <- rep(0, n_cycles)
v.p_discontinuation_soc[1] <- p.discontinuation1
v.p_discontinuation_soc[2:(tx_duration)] <- p.discontinuation2
if (tx_duration < n_cycles) {
  v.p_discontinuation_soc[(tx_duration + 1):n_cycles] <- 1
}

# --- Age- and sex-weighted mortality vector ---
# Assume life_table is already loaded from Prompt 2 and named life_table
ages <- age_start + 0:(n_cycles - 1)
v.p_death_general <- life_table$weighted[match(ages, life_table$age)]
# --- Preallocate vectors ---
v.p.mci_mil_int_on <- numeric(n_cycles)
v.p.mci_mod_int_on <- numeric(n_cycles)
v.p.mci_sev_int_on <- numeric(n_cycles)

v.p.mil_mci_int_on <- rep(p.mil_mci, n_cycles)  # Backward transitions, unaffected by treatment
v.p.mil_mod_int_on <- numeric(n_cycles)
v.p.mil_sev_int_on <- numeric(n_cycles)

v.p.mci_stay_int_on <- numeric(n_cycles)
v.p.mil_stay_int_on <- numeric(n_cycles)

v.p.mci_mil_int_off <- numeric(n_cycles)
v.p.mci_mod_int_off <- numeric(n_cycles)
v.p.mci_sev_int_off <- numeric(n_cycles)

v.p.mil_mci_int_off <- rep(p.mil_mci, n_cycles)  # Backward transitions, unaffected by treatment
v.p.mil_mod_int_off <- numeric(n_cycles)
v.p.mil_sev_int_off <- numeric(n_cycles)

v.p.mci_stay_int_off <- numeric(n_cycles)
v.p.mil_stay_int_off <- numeric(n_cycles)

# --- Compute time-dependent transition probabilities ---
for (cycle in 1:n_cycles) {
  # Treatment waning factor
  waning_factor <- (1 - tx_waning)^(cycle - 1)
  waning_factor_dis <- (1 - tx_waning_dis)^(cycle - 1)
  
  # --- On-treatment (mcion_c and milon_c) ---
  # MCI → Mild
  rate <- -log(1 - p.mci_mil)
  adj_rate <- rate * rr.tx_mci_mil^waning_factor
  v.p.mci_mil_int_on[cycle] <- 1 - exp(-adj_rate)
  
  # MCI → Moderate
  rate <- -log(1 - p.mci_mod)
  adj_rate <- rate * rr.tx_mci_mod^waning_factor
  v.p.mci_mod_int_on[cycle] <- 1 - exp(-adj_rate)
  
  # MCI → Severe
  rate <- -log(1 - p.mci_sev)
  adj_rate <- rate * rr.tx_mci_sev^waning_factor
  v.p.mci_sev_int_on[cycle] <- 1 - exp(-adj_rate)
  
  # MCI Stay
  v.p.mci_stay_int_on[cycle] <- 1 - (
    v.p.mci_mil_int_on[cycle] +
      v.p.mci_mod_int_on[cycle] +
      v.p.mci_sev_int_on[cycle]
  )
  
  # Mild → Moderate
  rate <- -log(1 - p.mil_mod)
  adj_rate <- rate * rr.tx_mil_mod^waning_factor
  v.p.mil_mod_int_on[cycle] <- 1 - exp(-adj_rate)
  
  # Mild → Severe
  rate <- -log(1 - p.mil_sev)
  adj_rate <- rate * rr.tx_mil_sev^waning_factor
  v.p.mil_sev_int_on[cycle] <- 1 - exp(-adj_rate)
  
  # Mild Stay
  v.p.mil_stay_int_on[cycle] <- 1 - (
    v.p.mil_mci_int_on[cycle] +
      v.p.mil_mod_int_on[cycle] +
      v.p.mil_sev_int_on[cycle]
  )
  
  # --- Off-treatment (mciof_c and milof_c) ---
  # MCI → Mild
  rate <- -log(1 - p.mci_mil)
  adj_rate <- rate * rr.tx_mci_mil_dis^waning_factor_dis
  v.p.mci_mil_int_off[cycle] <- 1 - exp(-adj_rate)
  
  # MCI → Moderate
  rate <- -log(1 - p.mci_mod)
  adj_rate <- rate * rr.tx_mci_mod_dis^waning_factor_dis
  v.p.mci_mod_int_off[cycle] <- 1 - exp(-adj_rate)
  
  # MCI → Severe
  rate <- -log(1 - p.mci_sev)
  adj_rate <- rate * rr.tx_mci_sev_dis^waning_factor_dis
  v.p.mci_sev_int_off[cycle] <- 1 - exp(-adj_rate)
  
  # MCI Stay
  v.p.mci_stay_int_off[cycle] <- 1 - (
    v.p.mci_mil_int_off[cycle] +
      v.p.mci_mod_int_off[cycle] +
      v.p.mci_sev_int_off[cycle]
  )
  
  # Mild → Moderate
  rate <- -log(1 - p.mil_mod)
  adj_rate <- rate * rr.tx_mil_mod_dis^waning_factor_dis
  v.p.mil_mod_int_off[cycle] <- 1 - exp(-adj_rate)
  
  # Mild → Severe
  rate <- -log(1 - p.mil_sev)
  adj_rate <- rate * rr.tx_mil_sev_dis^waning_factor_dis
  v.p.mil_sev_int_off[cycle] <- 1 - exp(-adj_rate)
  
  # Mild Stay
  v.p.mil_stay_int_off[cycle] <- 1 - (
    v.p.mil_mci_int_off[cycle] +
      v.p.mil_mod_int_off[cycle] +
      v.p.mil_sev_int_off[cycle]
  )
}
build_transition_matrix <- function(strat) {
  # Initialize transition probability array
  a.TP <- array(0, dim = c(length(state_names), length(state_names), n_cycles),
                dimnames = list(state_names, state_names, NULL))
  
  for (t in 1:n_cycles) {
    # --- Transition to death for all non-death states ---
    a.TP["mcion_c", "dth", t] <- 1 - exp(-v.p_death_general[t] * hr.mort_mci)
    a.TP["mciof_c", "dth", t] <- 1 - exp(-v.p_death_general[t] * hr.mort_mci)
    a.TP["milon_c", "dth", t] <- 1 - exp(-v.p_death_general[t] * hr.mort_mil)
    a.TP["milof_c", "dth", t] <- 1 - exp(-v.p_death_general[t] * hr.mort_mil)
    a.TP["mod_c", "dth", t]   <- 1 - exp(-v.p_death_general[t] * hr.mort_mod)
    a.TP["sev_c", "dth", t]   <- 1 - exp(-v.p_death_general[t] * hr.mort_sev)
    a.TP["mci_i", "dth", t]   <- 1 - exp(-v.p_death_general[t] * hr.mort_mci)
    a.TP["mil_i", "dth", t]   <- 1 - exp(-v.p_death_general[t] * hr.mort_mil)
    a.TP["mod_i", "dth", t]   <- 1 - exp(-v.p_death_general[t] * hr.mort_mod)
    a.TP["sev_i", "dth", t]   <- 1 - exp(-v.p_death_general[t] * hr.mort_sev)
    
    # Death is an absorbing state
    a.TP["dth", "dth", t] <- 1
    
    # --- Transitions from mcion_c ---
    if (strat == "soc") {
      p_mci_mil <- v.p.mci_mil_soc[t]
      p_mci_mod <- v.p.mci_mod_soc[t]
      p_mci_sev <- v.p.mci_sev_soc[t]
      p_mci_stay <- v.p.mci_stay_soc[t]
    } else {
      p_mci_mil <- v.p.mci_mil_int_on[t]
      p_mci_mod <- v.p.mci_mod_int_on[t]
      p_mci_sev <- v.p.mci_sev_int_on[t]
      p_mci_stay <- v.p.mci_stay_int_on[t]
    }
    
    surv <- 1 - a.TP["mcion_c", "dth", t]
    p_inst <- v.p.mci_to_inst_soc[t]
    p_disc <- v.p_discontinuation_soc[t]
    
    # On-treatment to on-treatment transitions
    a.TP["mcion_c", "mcion_c", t] <- p_mci_stay * surv * (1 - p_inst) * (1 - p_disc)
    a.TP["mcion_c", "milon_c", t] <- p_mci_mil * surv * (1 - p_inst) * (1 - p_disc)
    
    # On-treatment to off-treatment (due to discontinuation)
    a.TP["mcion_c", "mciof_c", t] <- p_mci_stay * surv * (1 - p_inst) * p_disc
    a.TP["mcion_c", "milof_c", t] <- p_mci_mil * surv * (1 - p_inst) * p_disc
    
    # Progression to more severe states — no discontinuation factor
    a.TP["mcion_c", "mod_c", t] <- p_mci_mod * surv * (1 - p_inst)
    a.TP["mcion_c", "sev_c", t] <- p_mci_sev * surv * (1 - p_inst)
    
    # Institutionalization
    a.TP["mcion_c", "mci_i", t] <- p_mci_stay * surv * p_inst
    a.TP["mcion_c", "mil_i", t] <- p_mci_mil * surv * p_inst
    a.TP["mcion_c", "mod_i", t] <- p_mci_mod * surv * p_inst
    a.TP["mcion_c", "sev_i", t] <- p_mci_sev * surv * p_inst
    
    # --- Transitions from milon_c ---
    if (strat == "soc") {
      p_mil_mci <- v.p.mil_mci_soc[t]
      p_mil_mod <- v.p.mil_mod_soc[t]
      p_mil_sev <- v.p.mil_sev_soc[t]
      p_mil_stay <- v.p.mil_stay_soc[t]
    } else {
      p_mil_mci <- v.p.mil_mci_int_on[t]
      p_mil_mod <- v.p.mil_mod_int_on[t]
      p_mil_sev <- v.p.mil_sev_int_on[t]
      p_mil_stay <- v.p.mil_stay_int_on[t]
    }
    
    surv <- 1 - a.TP["milon_c", "dth", t]
    p_inst <- v.p.mil_to_inst_soc[t]
    p_disc <- v.p_discontinuation_soc[t]
    
    # On-treatment to on-treatment transitions
    a.TP["milon_c", "milon_c", t] <- p_mil_stay * surv * (1 - p_inst) * (1 - p_disc)
    a.TP["milon_c", "mcion_c", t] <- p_mil_mci * surv * (1 - p_inst) * (1 - p_disc)
    
    # On-treatment to off-treatment (due to discontinuation)
    a.TP["milon_c", "milof_c", t] <- p_mil_stay * surv * (1 - p_inst) * p_disc
    a.TP["milon_c", "mciof_c", t] <- p_mil_mci * surv * (1 - p_inst) * p_disc
    
    # Progression to more severe states — no discontinuation factor
    a.TP["milon_c", "mod_c", t] <- p_mil_mod * surv * (1 - p_inst)
    a.TP["milon_c", "sev_c", t] <- p_mil_sev * surv * (1 - p_inst)
    
    # Institutionalization
    a.TP["milon_c", "mil_i", t] <- p_mil_stay * surv * p_inst
    a.TP["milon_c", "mci_i", t] <- p_mil_mci * surv * p_inst
    a.TP["milon_c", "mod_i", t] <- p_mil_mod * surv * p_inst
    a.TP["milon_c", "sev_i", t] <- p_mil_sev * surv * p_inst
    
    
    # --- Transitions from mciof_c ---
    if (strat == "soc") {
      p_mci_mil <- v.p.mci_mil_soc[t]
      p_mci_mod <- v.p.mci_mod_soc[t]
      p_mci_sev <- v.p.mci_sev_soc[t]
      p_mci_stay <- v.p.mci_stay_soc[t]
    } else {
      p_mci_mil <- v.p.mci_mil_int_off[t]
      p_mci_mod <- v.p.mci_mod_int_off[t]
      p_mci_sev <- v.p.mci_sev_int_off[t]
      p_mci_stay <- v.p.mci_stay_int_off[t]
    }
    
    surv <- 1 - a.TP["mciof_c", "dth", t]
    p_inst <- v.p.mci_to_inst_soc[t]
    
    a.TP["mciof_c", "mciof_c", t] <- p_mci_stay * surv * (1 - p_inst)
    a.TP["mciof_c", "milof_c", t] <- p_mci_mil * surv * (1 - p_inst)
    a.TP["mciof_c", "mod_c", t]   <- p_mci_mod * surv * (1 - p_inst)
    a.TP["mciof_c", "sev_c", t]   <- p_mci_sev * surv * (1 - p_inst)
    
    a.TP["mciof_c", "mci_i", t]   <- p_mci_stay * surv * p_inst
    a.TP["mciof_c", "mil_i", t]   <- p_mci_mil * surv * p_inst
    a.TP["mciof_c", "mod_i", t]   <- p_mci_mod * surv * p_inst
    a.TP["mciof_c", "sev_i", t]   <- p_mci_sev * surv * p_inst
    
    # --- Transitions from milof_c ---
    if (strat == "soc") {
      p_mil_mci <- v.p.mil_mci_soc[t]
      p_mil_mod <- v.p.mil_mod_soc[t]
      p_mil_sev <- v.p.mil_sev_soc[t]
      p_mil_stay <- v.p.mil_stay_soc[t]
    } else {
      p_mil_mci <- v.p.mil_mci_int_off[t]
      p_mil_mod <- v.p.mil_mod_int_off[t]
      p_mil_sev <- v.p.mil_sev_int_off[t]
      p_mil_stay <- v.p.mil_stay_int_off[t]
    }
    
    surv <- 1 - a.TP["milof_c", "dth", t]
    p_inst <- v.p.mil_to_inst_soc[t]
    
    a.TP["milof_c", "milof_c", t] <- p_mil_stay * surv * (1 - p_inst)
    a.TP["milof_c", "mciof_c", t] <- p_mil_mci * surv * (1 - p_inst)
    a.TP["milof_c", "mod_c", t]   <- p_mil_mod * surv * (1 - p_inst)
    a.TP["milof_c", "sev_c", t]   <- p_mil_sev * surv * (1 - p_inst)
    
    a.TP["milof_c", "mil_i", t]   <- p_mil_stay * surv * p_inst
    a.TP["milof_c", "mci_i", t]   <- p_mil_mci * surv * p_inst
    a.TP["milof_c", "mod_i", t]   <- p_mil_mod * surv * p_inst
    a.TP["milof_c", "sev_i", t]   <- p_mil_sev * surv * p_inst
    
    # --- Transitions from mod_c ---
    if (strat == "soc") {
      p_mod_mil <- v.p.mod_mil_soc[t]
      p_mod_sev <- v.p.mod_sev_soc[t]
      p_mod_stay <- v.p.mod_stay_soc[t]
    } else {
      p_mod_mil <- v.p.mod_mil_soc[t]
      p_mod_sev <- v.p.mod_sev_soc[t]
      p_mod_stay <- v.p.mod_stay_soc[t]
    }
    
    surv <- 1 - a.TP["mod_c", "dth", t]
    p_inst <- v.p.mod_to_inst_soc[t]
    
    a.TP["mod_c", "mod_c", t] <- p_mod_stay * surv * (1 - p_inst)
    a.TP["mod_c", "milof_c", t] <- p_mod_mil * surv * (1 - p_inst)
    a.TP["mod_c", "sev_c", t]   <- p_mod_sev * surv * (1 - p_inst)
    
    a.TP["mod_c", "mod_i", t]   <- p_mod_stay * surv * p_inst
    a.TP["mod_c", "mil_i", t]   <- p_mod_mil * surv * p_inst
    a.TP["mod_c", "sev_i", t]   <- p_mod_sev * surv * p_inst
    
    # --- Transitions from sev_c ---
    if (strat == "soc") {
      p_sev_mil <- v.p.sev_mil_soc[t]
      p_sev_mod <- v.p.sev_mod_soc[t]
      p_sev_stay <- v.p.sev_stay_soc[t]
    } else {
      p_sev_mil <- v.p.sev_mil_soc[t]
      p_sev_mod <- v.p.sev_mod_soc[t]
      p_sev_stay <- v.p.sev_stay_soc[t]
    }
    
    surv <- 1 - a.TP["sev_c", "dth", t]
    p_inst <- v.p.sev_to_inst_soc[t]
    
    a.TP["sev_c", "sev_c", t] <- p_sev_stay * surv * (1 - p_inst)
    a.TP["sev_c", "milof_c", t] <- p_sev_mil * surv * (1 - p_inst)
    a.TP["sev_c", "mod_c", t]   <- p_sev_mod * surv * (1 - p_inst)
    
    a.TP["sev_c", "sev_i", t]   <- p_sev_stay * surv * p_inst
    a.TP["sev_c", "mil_i", t]   <- p_sev_mil * surv * p_inst
    a.TP["sev_c", "mod_i", t]   <- p_sev_mod * surv * p_inst
    
    
    # --- Transitions from mci_i ---
    if (strat == "soc") {
      p_mci_mil <- v.p.mci_mil_soc[t]
      p_mci_mod <- v.p.mci_mod_soc[t]
      p_mci_sev <- v.p.mci_sev_soc[t]
      p_mci_stay <- v.p.mci_stay_soc[t]
    } else {
      p_mci_mil <- v.p.mci_mil_int_off[t]
      p_mci_mod <- v.p.mci_mod_int_off[t]
      p_mci_sev <- v.p.mci_sev_int_off[t]
      p_mci_stay <- v.p.mci_stay_int_off[t]
    }
    
    surv <- 1 - a.TP["mci_i", "dth", t]
    
    a.TP["mci_i", "mci_i", t] <- p_mci_stay * surv
    a.TP["mci_i", "mil_i", t] <- p_mci_mil * surv
    a.TP["mci_i", "mod_i", t] <- p_mci_mod * surv
    a.TP["mci_i", "sev_i", t] <- p_mci_sev * surv
    
    # --- Transitions from mil_i ---
    if (strat == "soc") {
      p_mil_mci <- v.p.mil_mci_soc[t]
      p_mil_mod <- v.p.mil_mod_soc[t]
      p_mil_sev <- v.p.mil_sev_soc[t]
      p_mil_stay <- v.p.mil_stay_soc[t]
    } else {
      p_mil_mci <- v.p.mil_mci_int_off[t]
      p_mil_mod <- v.p.mil_mod_int_off[t]
      p_mil_sev <- v.p.mil_sev_int_off[t]
      p_mil_stay <- v.p.mil_stay_int_off[t]
    }
    
    surv <- 1 - a.TP["mil_i", "dth", t]
    
    a.TP["mil_i", "mil_i", t] <- p_mil_stay * surv
    a.TP["mil_i", "mci_i", t] <- p_mil_mci * surv
    a.TP["mil_i", "mod_i", t] <- p_mil_mod * surv
    a.TP["mil_i", "sev_i", t] <- p_mil_sev * surv
    
    # --- Transitions from mod_i ---
    if (strat == "soc") {
      p_mod_mil <- v.p.mod_mil_soc[t]
      p_mod_sev <- v.p.mod_sev_soc[t]
      p_mod_stay <- v.p.mod_stay_soc[t]
    } else {
      p_mod_mil <- v.p.mod_mil_soc[t]
      p_mod_sev <- v.p.mod_sev_soc[t]
      p_mod_stay <- v.p.mod_stay_soc[t]
    }
    
    surv <- 1 - a.TP["mod_i", "dth", t]
    
    a.TP["mod_i", "mod_i", t] <- p_mod_stay * surv
    a.TP["mod_i", "mil_i", t] <- p_mod_mil * surv
    a.TP["mod_i", "sev_i", t] <- p_mod_sev * surv
    
    # --- Transitions from sev_i ---
    if (strat == "soc") {
      p_sev_mil <- v.p.sev_mil_soc[t]
      p_sev_mod <- v.p.sev_mod_soc[t]
      p_sev_stay <- v.p.sev_stay_soc[t]
    } else {
      p_sev_mil <- v.p.sev_mil_soc[t]
      p_sev_mod <- v.p.sev_mod_soc[t]
      p_sev_stay <- v.p.sev_stay_soc[t]
    }
    
    surv <- 1 - a.TP["sev_i", "dth", t]
    
    a.TP["sev_i", "sev_i", t] <- p_sev_stay * surv
    a.TP["sev_i", "mil_i", t] <- p_sev_mil * surv
    a.TP["sev_i", "mod_i", t] <- p_sev_mod * surv
    
    
    # --- Row normalization if row sum ≠ 1 ---
    for (s in state_names) {
      row_sum <- sum(a.TP[s, , t])
      if (row_sum > 0 && row_sum != 1) {
        a.TP[s, , t] <- a.TP[s, , t] / row_sum
      }
    }
  }
  
  return(a.TP)
}
# Function to compute Markov trace and total outcomes for a strategy
run_markov_model <- function(strat) {
  # Build transition probability array
  a.TP <- build_transition_matrix(strat)
  
  # Initialize state occupancy matrix
  m.trace <- matrix(0, nrow = n_cycles, ncol = length(state_names),
                    dimnames = list(NULL, state_names))
  
  # Initial state distribution
  v.init <- setNames(rep(0, length(state_names)), state_names)
  v.init["mciof_c"] <- 1 - p_starting_state_mci
  v.init["mcion_c"] <- p_starting_state_mci
  m.trace[1, ] <- v.init
  
  # Predefine utility and cost vectors
  v.utility <- c(
    mcion_c = u.mci_pt + u.mci_ic,
    mciof_c = u.mci_pt + u.mci_ic,
    milon_c = u.mil_pt + u.mil_ic,
    milof_c = u.mil_pt + u.mil_ic,
    mod_c = u.mod_pt + u.mod_ic,
    sev_c = u.sev_pt + u.sev_ic,
    mci_i = u.mci_pt_i + u.mci_ic_i,
    mil_i = u.mil_pt_i + u.mil_ic_i,
    mod_i = u.mod_pt_i + u.mod_ic_i,
    sev_i = u.sev_pt_i + u.sev_ic_i,
    dth = 0
  )
  
  v.cost <- c(
    mcion_c = c.mci_hc + c.mci_sc + c.mci_ic,
    mciof_c = c.mci_hc + c.mci_sc + c.mci_ic,
    milon_c = c.mil_hc + c.mil_sc + c.mil_ic,
    milof_c = c.mil_hc + c.mil_sc + c.mil_ic,
    mod_c = c.mod_hc + c.mod_sc + c.mod_ic,
    sev_c = c.sev_hc + c.sev_sc + c.sev_ic,
    mci_i = c.mci_i_hc + c.mci_i_sc + c.mci_i_ic,
    mil_i = c.mil_i_hc + c.mil_i_sc + c.mil_i_ic,
    mod_i = c.mod_i_hc + c.mod_i_sc + c.mod_i_ic,
    sev_i = c.sev_i_hc + c.sev_i_sc + c.sev_i_ic,
    dth = 0
  )
  
  # Initialize outcomes
  v.qalys <- numeric(n_cycles)
  v.costs <- numeric(n_cycles)
  v.lys <- numeric(n_cycles)
  
  for (t in 1:(n_cycles - 1)) {
    m.trace[t + 1, ] <- m.trace[t, ] %*% a.TP[, , t]
    
    v.qalys[t] <- sum(m.trace[t, ] * v.utility)
    v.costs[t] <- sum(m.trace[t, ] * v.cost)
    v.lys[t] <- sum(m.trace[t, state_names != "dth"])
    
    # Apply recurring treatment cost for "int"
    if (strat == "int") {
      v.costs[t] <- v.costs[t] + c.Tx * sum(m.trace[t, c("mcion_c", "milon_c")])
    }
  }
  
  # Final cycle outcomes
  v.qalys[n_cycles] <- sum(m.trace[n_cycles, ] * v.utility)
  v.costs[n_cycles] <- sum(m.trace[n_cycles, ] * v.cost)
  v.lys[n_cycles] <- sum(m.trace[n_cycles, state_names != "dth"])
  if (strat == "int") {
    v.costs[n_cycles] <- v.costs[n_cycles] + c.Tx * sum(m.trace[n_cycles, c("mcion_c", "milon_c")])
  }
  
  # Half-cycle correction
  v.qalys <- 0.5 * (v.qalys[-n_cycles] + v.qalys[-1])
  v.costs <- 0.5 * (v.costs[-n_cycles] + v.costs[-1])
  v.lys <- 0.5 * (v.lys[-n_cycles] + v.lys[-1])
  
  # Apply initial treatment QALY and cost (only once at start)
  if (strat == "int") {
    v.qalys[1] <- v.qalys[1] + u.Tx_start
    v.costs[1] <- v.costs[1] + c.Tx_start
  }
  
  # Apply discounting
  v.qalys <- v.qalys / ((1 + discount_QALY)^(1:(n_cycles - 1)))
  v.costs <- v.costs / ((1 + discount_COST)^(1:(n_cycles - 1)))
  v.lys <- v.lys / ((1 + discount_EFFECT)^(1:(n_cycles - 1)))
  
  # Summarize total outcomes
  total_qalys <- sum(v.qalys)
  total_costs <- sum(v.costs)
  total_lys <- sum(v.lys)
  
  return(list(QALYs = total_qalys, Costs = total_costs, LYs = total_lys))
}

# Run model for both strategies
results_soc <- run_markov_model("soc")
results_int <- run_markov_model("int")

# Display results
cat("\n--- IPECAD MODEL RESULTS ---\n")
cat("Standard of Care:\n")
cat("  QALYs:", round(results_soc$QALYs, 3), "\n")
cat("  Costs:", round(results_soc$Costs, 2), "\n")
cat("  LYs:", round(results_soc$LYs, 3), "\n\n")

cat("Intervention:\n")
cat("  QALYs:", round(results_int$QALYs, 3), "\n")
cat("  Costs:", round(results_int$Costs, 2), "\n")
cat("  LYs:", round(results_int$LYs, 3), "\n\n")

cat("Incremental:\n")
cat("  ΔQALYs:", round(results_int$QALYs - results_soc$QALYs, 3), "\n")
cat("  ΔCosts:", round(results_int$Costs - results_soc$Costs, 2), "\n")
cat("  ΔLYs:", round(results_int$LYs - results_soc$LYs, 3), "\n")
```
