## Supplementary material for "Replicating Health-Economic Simulation Models for Alzheimer’s Disease Using Artificial Intelligence": final R scripts: o11.R

```
# =========================
# IPECAD Model Input Setup
# =========================

# ---- Descriptive Variables ----
state_names <- c("mcion_c", "mciof_c", "milon_c", "milof_c", 
                 "mod_c", "sev_c", "mci_i", "mil_i", "mod_i", "sev_i", "dth")

strategy_names <- c("soc", "int")

# ---- Demographics ----
age_start <- 71
sex <- "weighted"  # 52% female

# ---- Time Horizon ----
n_cycles <- 29

# ---- Initial State Distribution ----
p_starting_state_mci <- 0.55  # Implies 0.45 in mild dementia

# ---- Transition Probabilities (Disease Progression) ----
p_mci_mil <- 0.23
p_mci_mod <- 0
p_mci_sev <- 0

p_mil_mci <- 0.03
p_mil_mod <- 0.35
p_mil_sev <- 0.04

p_mod_mil <- 0.03
p_mod_sev <- 0.42

p_sev_mil <- 0
p_sev_mod <- 0.02

# ---- Transition Probabilities (Institutionalization) ----
p_mci_i <- 0.024
p_mil_i <- 0.038
p_mod_i <- 0.110
p_sev_i <- 0.259

# ---- Mortality ----
hr_mort_mci <- 1.82
hr_mort_mil <- 2.92
hr_mort_mod <- 3.85
hr_mort_sev <- 9.52

# ---- Treatment Effect (Relative Risks) ----
rr_tx_mci_mil <- 0.69
rr_tx_mci_mod <- 1
rr_tx_mci_sev <- 1
rr_tx_mil_mod <- 0.69
rr_tx_mil_sev <- 0.69

# ---- Treatment Effect After Discontinuation ----
rr_tx_mci_mil_dis <- 1
rr_tx_mci_mod_dis <- 1
rr_tx_mci_sev_dis <- 1
rr_tx_mil_mod_dis <- 1
rr_tx_mil_sev_dis <- 1

# ---- Treatment Discontinuation ----
p_discontinuation1 <- 0.069
p_discontinuation2 <- 0
discontinuation2_begin <- 2
tx_duration <- 29

# ---- Treatment Waning ----
tx_waning <- 0
tx_waning_dis <- 0

# ---- Utilities (Patient) ----
u_mci_pt <- 0.851 - 0.17
u_mil_pt <- 0.851 - 0.22
u_mod_pt <- 0.851 - 0.36
u_sev_pt <- 0.851 - 0.53

u_mci_pt_i <- 0.851 - 0.17
u_mil_pt_i <- 0.851 - 0.19
u_mod_pt_i <- 0.851 - 0.42
u_sev_pt_i <- 0.851 - 0.59

# ---- Utilities (Informal Caregiver) ----
u_mci_ic <- -0.03
u_mil_ic <- -0.05
u_mod_ic <- -0.08
u_sev_ic <- -0.10

u_mci_ic_i <- -0.03
u_mil_ic_i <- -0.05
u_mod_ic_i <- -0.08
u_sev_ic_i <- -0.10

# ---- Utility Impact of Treatment (ARIA) ----
u_Tx_start <- -0.14 * (12/52) * 0.035

# ---- Costs (USD/year) ----
c_mci_hc <- 6042 * 1.12 + 460
c_mil_hc <- 6042 * 1.56 + 965 + 0.21 * 365 * 0.333
c_mod_hc <- 6042 * 1.93 + 1544 + 0.66 * 365 * 0.333
c_sev_hc <- 6042 * 1.93 + 1930

c_mci_i_hc <- c_mci_hc
c_mil_i_hc <- c_mil_hc
c_mod_i_hc <- c_mod_hc
c_sev_i_hc <- c_sev_hc

c_mci_sc <- 0
c_mil_sc <- 0
c_mod_sc <- 0
c_sev_sc <- 0

c_mci_i_sc <- 7394 * 12
c_mil_i_sc <- 7394 * 12
c_mod_i_sc <- 7394 * 12
c_sev_i_sc <- 7394 * 12

c_mci_ic <- 69 * 12 * 32.46 + 0.204 * 0.049 * 20 * 52 * 32.46
c_mil_ic <- 113 * 12 * 32.46 + 0.112 * 0.086 * 20 * 52 * 32.46
c_mod_ic <- 169 * 12 * 32.46
c_sev_ic <- 298 * 12 * 32.46

c_mci_i_ic <- 69 * 12 * 32.46 * 0.44 + 0.204 * 0.049 * 20 * 52 * 32.46
c_mil_i_ic <- 113 * 12 * 32.46 * 0.44 + 0.112 * 0.086 * 20 * 52 * 32.46
c_mod_i_ic <- 169 * 12 * 32.46 * 0.44
c_sev_i_ic <- 298 * 12 * 32.46 * 0.44

# ---- Treatment Costs ----
c_Tx <- 26500 + (52/2) * 78.35
c_Tx_start <- 261.10 * 4 + 261.10 * 3 * 0.215

# ---- Discount Rates ----
discount_EFFECT <- 0.03
discount_QALY <- 0.03
discount_COST <- 0.03

# ---- Willingness-to-Pay Threshold ----
wtp <- 100000

# ---- Half-Cycle Correction ----
half_cycle_correction <- TRUE
# ===============================
# Load and Process 2019 US Life Table
# ===============================

# Load life table from CSV
life_table <- read.csv("lifetable_US_2019_ssa.csv")

# Convert mortality probabilities to mortality rates
# Formula: rate = -log(1 - probability)
life_table$male <- -log(1 - life_table$male)
life_table$female <- -log(1 - life_table$female)

# Compute sex-weighted average mortality rates
# 52% female as per IPECAD model input
life_table$weighted <- 0.48 * life_table$male + 0.52 * life_table$female

# Retain only relevant columns
life_table <- life_table[, c("age", "male", "female", "weighted")]

# Return the processed life table
life_table
# ===============================
# Time-dependent Transition Vectors (SOC Strategy)
# ===============================

# Assume `n_cycles` and `age_start` are already defined from previous code
# Assume `life_table` is already loaded and contains columns: "age", "male", "female", "weighted"

# ---- Disease Progression Probabilities for SOC Strategy ----
v.p.mci_mil_soc <- rep(p_mci_mil, n_cycles)
v.p.mci_mod_soc <- rep(p_mci_mod, n_cycles)
v.p.mci_sev_soc <- rep(p_mci_sev, n_cycles)

v.p.mil_mci_soc <- rep(p_mil_mci, n_cycles)
v.p.mil_mod_soc <- rep(p_mil_mod, n_cycles)
v.p.mil_sev_soc <- rep(p_mil_sev, n_cycles)

v.p.mod_mil_soc <- rep(p_mod_mil, n_cycles)
v.p.mod_sev_soc <- rep(p_mod_sev, n_cycles)

v.p.sev_mil_soc <- rep(p_sev_mil, n_cycles)
v.p.sev_mod_soc <- rep(p_sev_mod, n_cycles)

# ---- Staying Probabilities (excluding institutional and mortality) ----
v.p.mci_stay_soc <- 1 - (v.p.mci_mil_soc + v.p.mci_mod_soc + v.p.mci_sev_soc)
v.p.mil_stay_soc <- 1 - (v.p.mil_mci_soc + v.p.mil_mod_soc + v.p.mil_sev_soc)
v.p.mod_stay_soc <- 1 - (v.p.mod_mil_soc + v.p.mod_sev_soc)
v.p.sev_stay_soc <- 1 - (v.p.sev_mil_soc + v.p.sev_mod_soc)

# ---- Transition to Institution (Community States Only) ----
v.p.mci_i <- rep(p_mci_i, n_cycles)
v.p.mil_i <- rep(p_mil_i, n_cycles)
v.p.mod_i <- rep(p_mod_i, n_cycles)
v.p.sev_i <- rep(p_sev_i, n_cycles)

# ---- Treatment Discontinuation Vector ----
v.p.discontinuation <- numeric(n_cycles)
v.p.discontinuation[1:(discontinuation2_begin - 1)] <- p_discontinuation1
v.p.discontinuation[discontinuation2_begin:min(tx_duration, n_cycles)] <- p_discontinuation2
if (tx_duration < n_cycles) {
  v.p.discontinuation[(tx_duration + 1):n_cycles] <- 1
}

# ---- Mortality Vector (Weighted General Population Mortality) ----
v.mort_rate <- life_table$weighted[match(age_start:(age_start + n_cycles - 1), life_table$age)]
# ===============================
# Time-dependent Transition Vectors (INT Strategy)
# ===============================

# Define cycle indices
cycle_index <- 1:n_cycles

# ---- Helper function for treatment adjustment ----
adjusted_prob <- function(p, rr, waning, t) {
  rate <- -log(1 - p)
  rate_adj <- rate * rr ^ ((1 - waning) ^ (t - 1))
  p_adj <- 1 - exp(-rate_adj)
  return(p_adj)
}

# ---- On-Treatment Transitions (mcion_c) ----
v.p.mci_mil_int_on <- adjusted_prob(p_mci_mil, rr_tx_mci_mil, tx_waning, cycle_index)
v.p.mci_mod_int_on <- adjusted_prob(p_mci_mod, rr_tx_mci_mod, tx_waning, cycle_index)
v.p.mci_sev_int_on <- adjusted_prob(p_mci_sev, rr_tx_mci_sev, tx_waning, cycle_index)

# Staying probability
v.p.mci_stay_int_on <- 1 - (v.p.mci_mil_int_on + v.p.mci_mod_int_on + v.p.mci_sev_int_on)

# ---- On-Treatment Transitions (milon_c) ----
v.p.mil_mci_int_on <- rep(p_mil_mci, n_cycles)  # backward, no treatment effect
v.p.mil_mod_int_on <- adjusted_prob(p_mil_mod, rr_tx_mil_mod, tx_waning, cycle_index)
v.p.mil_sev_int_on <- adjusted_prob(p_mil_sev, rr_tx_mil_sev, tx_waning, cycle_index)

# Staying probability
v.p.mil_stay_int_on <- 1 - (v.p.mil_mci_int_on + v.p.mil_mod_int_on + v.p.mil_sev_int_on)

# ---- Off-Treatment Transitions (mciof_c) ----
v.p.mci_mil_int_off <- adjusted_prob(p_mci_mil, rr_tx_mci_mil_dis, tx_waning_dis, cycle_index)
v.p.mci_mod_int_off <- adjusted_prob(p_mci_mod, rr_tx_mci_mod_dis, tx_waning_dis, cycle_index)
v.p.mci_sev_int_off <- adjusted_prob(p_mci_sev, rr_tx_mci_sev_dis, tx_waning_dis, cycle_index)

# Staying probability
v.p.mci_stay_int_off <- 1 - (v.p.mci_mil_int_off + v.p.mci_mod_int_off + v.p.mci_sev_int_off)

# ---- Off-Treatment Transitions (milof_c) ----
v.p.mil_mci_int_off <- rep(p_mil_mci, n_cycles)  # backward, no treatment effect
v.p.mil_mod_int_off <- adjusted_prob(p_mil_mod, rr_tx_mil_mod_dis, tx_waning_dis, cycle_index)
v.p.mil_sev_int_off <- adjusted_prob(p_mil_sev, rr_tx_mil_sev_dis, tx_waning_dis, cycle_index)

# Staying probability
v.p.mil_stay_int_off <- 1 - (v.p.mil_mci_int_off + v.p.mil_mod_int_off + v.p.mil_sev_int_off)
# ===============================
# Transition Probability Matrix Builder
# ===============================

build_transition_matrix <- function(strat) {
  # Initialize 3D array: states × states × cycles
  a.TP <- array(0, dim = c(length(state_names), length(state_names), n_cycles),
                dimnames = list(state_names, state_names, NULL))
  
  for (t in 1:n_cycles) {
    # Mortality rate at cycle t
    mort_t <- v.mort_rate[t]
    
    # ---- Assign transition to death for all states (explicit lines) ----
    a.TP["mcion_c", "dth", t] <- 1 - exp(-mort_t * hr_mort_mci)
    a.TP["mciof_c", "dth", t] <- 1 - exp(-mort_t * hr_mort_mci)
    a.TP["milon_c", "dth", t] <- 1 - exp(-mort_t * hr_mort_mil)
    a.TP["milof_c", "dth", t] <- 1 - exp(-mort_t * hr_mort_mil)
    a.TP["mod_c", "dth", t]    <- 1 - exp(-mort_t * hr_mort_mod)
    a.TP["sev_c", "dth", t]    <- 1 - exp(-mort_t * hr_mort_sev)
    a.TP["mci_i", "dth", t]    <- 1 - exp(-mort_t * hr_mort_mci)
    a.TP["mil_i", "dth", t]    <- 1 - exp(-mort_t * hr_mort_mil)
    a.TP["mod_i", "dth", t]    <- 1 - exp(-mort_t * hr_mort_mod)
    a.TP["sev_i", "dth", t]    <- 1 - exp(-mort_t * hr_mort_sev)
    
    # ---- Death is absorbing ----
    a.TP["dth", "dth", t] <- 1
    
    # ---- Transitions from mcion_c (on-treatment MCI community) ----
    a.TP["mcion_c", "mcion_c", t] <- if (strat == "soc") {
      v.p.mci_stay_soc[t] * (1 - a.TP["mcion_c", "dth", t]) * (1 - v.p.mci_i[t]) * (1 - v.p.discontinuation[t])
    } else {
      v.p.mci_stay_int_on[t] * (1 - a.TP["mcion_c", "dth", t]) * (1 - v.p.mci_i[t]) * (1 - v.p.discontinuation[t])
    }
    
    a.TP["mcion_c", "mciof_c", t] <- if (strat == "soc") {
      v.p.mci_stay_soc[t] * (1 - a.TP["mcion_c", "dth", t]) * (1 - v.p.mci_i[t]) * v.p.discontinuation[t]
    } else {
      v.p.mci_stay_int_on[t] * (1 - a.TP["mcion_c", "dth", t]) * (1 - v.p.mci_i[t]) * v.p.discontinuation[t]
    }
    
    a.TP["mcion_c", "milon_c", t] <- if (strat == "soc") {
      v.p.mci_mil_soc[t] * (1 - a.TP["mcion_c", "dth", t]) * (1 - v.p.mci_i[t]) * (1 - v.p.discontinuation[t])
    } else {
      v.p.mci_mil_int_on[t] * (1 - a.TP["mcion_c", "dth", t]) * (1 - v.p.mci_i[t]) * (1 - v.p.discontinuation[t])
    }
    
    a.TP["mcion_c", "mciof_c", t] <- a.TP["mcion_c", "mciof_c", t] + if (strat == "soc") {
      v.p.mci_mil_soc[t] * (1 - a.TP["mcion_c", "dth", t]) * (1 - v.p.mci_i[t]) * v.p.discontinuation[t]
    } else {
      v.p.mci_mil_int_on[t] * (1 - a.TP["mcion_c", "dth", t]) * (1 - v.p.mci_i[t]) * v.p.discontinuation[t]
    }
    
    a.TP["mcion_c", "mod_c", t] <- if (strat == "soc") {
      v.p.mci_mod_soc[t] * (1 - a.TP["mcion_c", "dth", t])
    } else {
      v.p.mci_mod_int_on[t] * (1 - a.TP["mcion_c", "dth", t])
    }
    
    a.TP["mcion_c", "sev_c", t] <- if (strat == "soc") {
      v.p.mci_sev_soc[t] * (1 - a.TP["mcion_c", "dth", t])
    } else {
      v.p.mci_sev_int_on[t] * (1 - a.TP["mcion_c", "dth", t])
    }
    
    a.TP["mcion_c", "mci_i", t] <- if (strat == "soc") {
      v.p.mci_i[t] * (1 - a.TP["mcion_c", "dth", t])
    } else {
      v.p.mci_i[t] * (1 - a.TP["mcion_c", "dth", t])
    }
    
    # ---- Transitions from milon_c (on-treatment mild community) ----
    a.TP["milon_c", "milon_c", t] <- if (strat == "soc") {
      v.p.mil_stay_soc[t] * (1 - a.TP["milon_c", "dth", t]) * (1 - v.p.mil_i[t]) * (1 - v.p.discontinuation[t])
    } else {
      v.p.mil_stay_int_on[t] * (1 - a.TP["milon_c", "dth", t]) * (1 - v.p.mil_i[t]) * (1 - v.p.discontinuation[t])
    }
    
    a.TP["milon_c", "milof_c", t] <- if (strat == "soc") {
      v.p.mil_stay_soc[t] * (1 - a.TP["milon_c", "dth", t]) * (1 - v.p.mil_i[t]) * v.p.discontinuation[t]
    } else {
      v.p.mil_stay_int_on[t] * (1 - a.TP["milon_c", "dth", t]) * (1 - v.p.mil_i[t]) * v.p.discontinuation[t]
    }
    
    a.TP["milon_c", "mciof_c", t] <- if (strat == "soc") {
      v.p.mil_mci_soc[t] * (1 - a.TP["milon_c", "dth", t]) * (1 - v.p.mil_i[t]) * v.p.discontinuation[t]
    } else {
      v.p.mil_mci_int_on[t] * (1 - a.TP["milon_c", "dth", t]) * (1 - v.p.mil_i[t]) * v.p.discontinuation[t]
    }
    
    a.TP["milon_c", "milof_c", t] <- a.TP["milon_c", "milof_c", t] + if (strat == "soc") {
      v.p.mil_mci_soc[t] * (1 - a.TP["milon_c", "dth", t]) * (1 - v.p.mil_i[t]) * (1 - v.p.discontinuation[t])
    } else {
      v.p.mil_mci_int_on[t] * (1 - a.TP["milon_c", "dth", t]) * (1 - v.p.mil_i[t]) * (1 - v.p.discontinuation[t])
    }
    
    a.TP["milon_c", "mod_c", t] <- if (strat == "soc") {
      v.p.mil_mod_soc[t] * (1 - a.TP["milon_c", "dth", t])
    } else {
      v.p.mil_mod_int_on[t] * (1 - a.TP["milon_c", "dth", t])
    }
    
    a.TP["milon_c", "sev_c", t] <- if (strat == "soc") {
      v.p.mil_sev_soc[t] * (1 - a.TP["milon_c", "dth", t])
    } else {
      v.p.mil_sev_int_on[t] * (1 - a.TP["milon_c", "dth", t])
    }
    
    a.TP["milon_c", "mil_i", t] <- if (strat == "soc") {
      v.p.mil_i[t] * (1 - a.TP["milon_c", "dth", t])
    } else {
      v.p.mil_i[t] * (1 - a.TP["milon_c", "dth", t])
    }
    
    # ---- Transitions from mciof_c (off-treatment MCI community) ----
    if (strat == "soc") {
      a.TP["mciof_c", "mciof_c", t] <- v.p.mci_stay_soc[t] * (1 - a.TP["mciof_c", "dth", t]) * (1 - v.p.mci_i[t])
      a.TP["mciof_c", "milof_c", t] <- v.p.mci_mil_soc[t] * (1 - a.TP["mciof_c", "dth", t]) * (1 - v.p.mci_i[t])
      a.TP["mciof_c", "mod_c", t]   <- v.p.mci_mod_soc[t] * (1 - a.TP["mciof_c", "dth", t])
      a.TP["mciof_c", "sev_c", t]   <- v.p.mci_sev_soc[t] * (1 - a.TP["mciof_c", "dth", t])
    } else {
      a.TP["mciof_c", "mciof_c", t] <- v.p.mci_stay_int_off[t] * (1 - a.TP["mciof_c", "dth", t]) * (1 - v.p.mci_i[t])
      a.TP["mciof_c", "milof_c", t] <- v.p.mci_mil_int_off[t] * (1 - a.TP["mciof_c", "dth", t]) * (1 - v.p.mci_i[t])
      a.TP["mciof_c", "mod_c", t]   <- v.p.mci_mod_int_off[t] * (1 - a.TP["mciof_c", "dth", t])
      a.TP["mciof_c", "sev_c", t]   <- v.p.mci_sev_int_off[t] * (1 - a.TP["mciof_c", "dth", t])
    }
    a.TP["mciof_c", "mci_i", t] <- v.p.mci_i[t] * (1 - a.TP["mciof_c", "dth", t])
    
    # ---- Transitions from milof_c (off-treatment mild community) ----
    if (strat == "soc") {
      a.TP["milof_c", "milof_c", t] <- v.p.mil_stay_soc[t] * (1 - a.TP["milof_c", "dth", t]) * (1 - v.p.mil_i[t])
      a.TP["milof_c", "mciof_c", t] <- v.p.mil_mci_soc[t] * (1 - a.TP["milof_c", "dth", t]) * (1 - v.p.mil_i[t])
      a.TP["milof_c", "mod_c", t]   <- v.p.mil_mod_soc[t] * (1 - a.TP["milof_c", "dth", t])
      a.TP["milof_c", "sev_c", t]   <- v.p.mil_sev_soc[t] * (1 - a.TP["milof_c", "dth", t])
    } else {
      a.TP["milof_c", "milof_c", t] <- v.p.mil_stay_int_off[t] * (1 - a.TP["milof_c", "dth", t]) * (1 - v.p.mil_i[t])
      a.TP["milof_c", "mciof_c", t] <- v.p.mil_mci_int_off[t] * (1 - a.TP["milof_c", "dth", t]) * (1 - v.p.mil_i[t])
      a.TP["milof_c", "mod_c", t]   <- v.p.mil_mod_int_off[t] * (1 - a.TP["milof_c", "dth", t])
      a.TP["milof_c", "sev_c", t]   <- v.p.mil_sev_int_off[t] * (1 - a.TP["milof_c", "dth", t])
    }
    a.TP["milof_c", "mil_i", t] <- v.p.mil_i[t] * (1 - a.TP["milof_c", "dth", t])
    
    # ---- Transitions from mod_c (moderate dementia community) ----
    a.TP["mod_c", "mod_c", t] <- v.p.mod_stay_soc[t] * (1 - a.TP["mod_c", "dth", t]) * (1 - v.p.mod_i[t])
    a.TP["mod_c", "milof_c", t] <- v.p.mod_mil_soc[t] * (1 - a.TP["mod_c", "dth", t]) * (1 - v.p.mod_i[t])
    a.TP["mod_c", "sev_c", t] <- v.p.mod_sev_soc[t] * (1 - a.TP["mod_c", "dth", t])
    a.TP["mod_c", "mod_i", t] <- v.p.mod_i[t] * (1 - a.TP["mod_c", "dth", t])
    
    # ---- Transitions from sev_c (severe dementia community) ----
    a.TP["sev_c", "sev_c", t] <- v.p.sev_stay_soc[t] * (1 - a.TP["sev_c", "dth", t]) * (1 - v.p.sev_i[t])
    a.TP["sev_c", "mod_c", t] <- v.p.sev_mod_soc[t] * (1 - a.TP["sev_c", "dth", t])
    a.TP["sev_c", "sev_i", t] <- v.p.sev_i[t] * (1 - a.TP["sev_c", "dth", t])
    
    # ---- Transitions from mci_i (institutionalized MCI) ----
    a.TP["mci_i", "mci_i", t] <- v.p.mci_stay_soc[t] * (1 - a.TP["mci_i", "dth", t])
    a.TP["mci_i", "mil_i", t] <- v.p.mci_mil_soc[t] * (1 - a.TP["mci_i", "dth", t])
    a.TP["mci_i", "mod_i", t] <- v.p.mci_mod_soc[t] * (1 - a.TP["mci_i", "dth", t])
    a.TP["mci_i", "sev_i", t] <- v.p.mci_sev_soc[t] * (1 - a.TP["mci_i", "dth", t])
    
    # ---- Transitions from mil_i (institutionalized mild) ----
    a.TP["mil_i", "mil_i", t] <- v.p.mil_stay_soc[t] * (1 - a.TP["mil_i", "dth", t])
    a.TP["mil_i", "mci_i", t] <- v.p.mil_mci_soc[t] * (1 - a.TP["mil_i", "dth", t])
    a.TP["mil_i", "mod_i", t] <- v.p.mil_mod_soc[t] * (1 - a.TP["mil_i", "dth", t])
    a.TP["mil_i", "sev_i", t] <- v.p.mil_sev_soc[t] * (1 - a.TP["mil_i", "dth", t])
    
    # ---- Transitions from mod_i (institutionalized moderate) ----
    a.TP["mod_i", "mod_i", t] <- v.p.mod_stay_soc[t] * (1 - a.TP["mod_i", "dth", t])
    a.TP["mod_i", "mil_i", t] <- v.p.mod_mil_soc[t] * (1 - a.TP["mod_i", "dth", t])
    a.TP["mod_i", "sev_i", t] <- v.p.mod_sev_soc[t] * (1 - a.TP["mod_i", "dth", t])
    
    # ---- Transitions from sev_i (institutionalized severe) ----
    a.TP["sev_i", "sev_i", t] <- v.p.sev_stay_soc[t] * (1 - a.TP["sev_i", "dth", t])
    a.TP["sev_i", "mod_i", t] <- v.p.sev_mod_soc[t] * (1 - a.TP["sev_i", "dth", t])
    
    # ---- Row normalization ----
    for (s in state_names) {
      row_sum <- sum(a.TP[s, , t])
      if (row_sum > 0 && row_sum != 1) {
        a.TP[s, , t] <- a.TP[s, , t] / row_sum
      }
    }
  }
  
  return(a.TP)
}
# ===============================
# Compute Markov Trace and Outcomes
# ===============================

compute_outcomes <- function(strat) {
  # Get transition probabilities
  a.TP <- build_transition_matrix(strat)
  
  # ---- Initial State Distribution ----
  v.init_state <- setNames(rep(0, length(state_names)), state_names)
  v.init_state["mcion_c"] <- if (strat == "int") p_starting_state_mci else 0
  v.init_state["mciof_c"] <- if (strat == "soc") p_starting_state_mci else 0
  v.init_state["milon_c"] <- if (strat == "int") 1 - p_starting_state_mci else 0
  v.init_state["milof_c"] <- if (strat == "soc") 1 - p_starting_state_mci else 0
  
  # ---- Initialize Trace Matrix ----
  m.M <- matrix(0, nrow = n_cycles + 1, ncol = length(state_names),
                dimnames = list(0:n_cycles, state_names))
  m.M[1, ] <- v.init_state
  
  # ---- Outcome Vectors ----
  v.QALYs <- numeric(n_cycles + 1)
  v.Costs <- numeric(n_cycles + 1)
  v.LYs <- numeric(n_cycles + 1)
  
  # ---- Utility and Cost Vectors ----
  v.u <- setNames(c(u_mci_pt, u_mci_pt, u_mil_pt, u_mil_pt, u_mod_pt, u_sev_pt,
                    u_mci_pt_i, u_mil_pt_i, u_mod_pt_i, u_sev_pt_i, 0), state_names)
  
  v.c <- setNames(c(c_mci_hc + c_mci_sc + c_mci_ic,
                    c_mci_hc + c_mci_sc + c_mci_ic,
                    c_mil_hc + c_mil_sc + c_mil_ic,
                    c_mil_hc + c_mil_sc + c_mil_ic,
                    c_mod_hc + c_mod_sc + c_mod_ic,
                    c_sev_hc + c_sev_sc + c_sev_ic,
                    c_mci_i_hc + c_mci_i_sc + c_mci_i_ic,
                    c_mil_i_hc + c_mil_i_sc + c_mil_i_ic,
                    c_mod_i_hc + c_mod_i_sc + c_mod_i_ic,
                    c_sev_i_hc + c_sev_i_sc + c_sev_i_ic,
                    0), state_names)
  
  # ---- Markov Trace Simulation ----
  for (t in 1:n_cycles) {
    m.M[t + 1, ] <- m.M[t, ] %*% a.TP[, , t]
    
    # On-treatment states for "int"
    on_tx_states <- c("mcion_c", "milon_c")
    
    # Calculate QALYs and Costs
    v.QALYs[t] <- sum(m.M[t, ] * v.u)
    v.Costs[t] <- sum(m.M[t, ] * v.c)
    v.LYs[t] <- sum(m.M[t, ])
    
    # Add recurring treatment costs for "int"
    if (strat == "int") {
      v.Costs[t] <- v.Costs[t] + sum(m.M[t, on_tx_states]) * c_Tx
    }
    
    # Add first cycle initial treatment costs and disutility
    if (t == 1 && strat == "int") {
      v.Costs[t] <- v.Costs[t] + c_Tx_start
      v.QALYs[t] <- v.QALYs[t] + sum(m.M[t, on_tx_states]) * u_Tx_start
    }
  }
  
  # ---- Half-Cycle Correction ----
  v.QALYs <- (v.QALYs[-length(v.QALYs)] + v.QALYs[-1]) / 2
  v.Costs <- (v.Costs[-length(v.Costs)] + v.Costs[-1]) / 2
  v.LYs   <- (v.LYs[-length(v.LYs)]   + v.LYs[-1])   / 2
  
  # ---- Discounting ----
  discount_weights <- 1 / ((1 + discount_QALY) ^ (0:(n_cycles - 1)))
  total_QALYs <- sum(v.QALYs * discount_weights)
  
  discount_weights_cost <- 1 / ((1 + discount_COST) ^ (0:(n_cycles - 1)))
  total_Costs <- sum(v.Costs * discount_weights_cost)
  
  discount_weights_ly <- 1 / ((1 + discount_EFFECT) ^ (0:(n_cycles - 1)))
  total_LYs <- sum(v.LYs * discount_weights_ly)
  
  return(list(QALYs = total_QALYs, Costs = total_Costs, LYs = total_LYs))
}

# ---- Compute and Compare Results ----
results_soc <- compute_outcomes("soc")
results_int <- compute_outcomes("int")

cat("===== Comparative Results =====\n")
cat(sprintf("Standard of Care: QALYs = %.2f, Costs = $%.2f, LYs = %.2f\n",
            results_soc$QALYs, results_soc$Costs, results_soc$LYs))
cat(sprintf("Intervention    : QALYs = %.2f, Costs = $%.2f, LYs = %.2f\n",
            results_int$QALYs, results_int$Costs, results_int$LYs))
cat("================================\n")
```
