## Supplementary material for "Replicating Health-Economic Simulation Models for Alzheimer’s Disease Using Artificial Intelligence": final R scripts: o12.R

```
# Load necessary library
library(dampack)

# Fixed state names
state_names <- c("mcion_c", "mciof_c", "milon_c", "milof_c", 
                 "mod_c", "sev_c", "mci_i", "mil_i", 
                 "mod_i", "sev_i", "dth")

# Strategy names
strategy_names <- c("soc", "int")

# Starting population characteristics
age_start <- 71
sex <- "weighted"  # 52% female
p_starting_state_mci <- 0.55

# Time horizon
n_cycles <- 29

# Transition probabilities (per cycle)
p_mci_mil <- 0.23
p_mci_mod <- 0
p_mci_sev <- 0

p_mil_mci <- 0.03
p_mil_mod <- 0.35
p_mil_sev <- 0.04

p_mod_mil <- 0.03
p_mod_sev <- 0.42

p_sev_mil <- 0
p_sev_mod <- 0.02

# Transition to institution setting (per cycle)
p_mci_i <- 0.024
p_mil_i <- 0.038
p_mod_i <- 0.110
p_sev_i <- 0.259

# Hazard ratios for AD mortality vs general population
hr_mort_mci <- 1.82
hr_mort_mil <- 2.92
hr_mort_mod <- 3.85
hr_mort_sev <- 9.52

# Treatment effect relative risks (on-treatment)
rr_tx_mci_mil <- 0.69
rr_tx_mci_mod <- 1
rr_tx_mci_sev <- 1
rr_tx_mil_mod <- 0.69
rr_tx_mil_sev <- 0.69

# Treatment effect relative risks (post-discontinuation)
rr_tx_mci_mil_dis <- 1
rr_tx_mci_mod_dis <- 1
rr_tx_mci_sev_dis <- 1
rr_tx_mil_mod_dis <- 1
rr_tx_mil_sev_dis <- 1

# Treatment discontinuation
p_discontinuation1 <- 0.069
p_discontinuation2 <- 0
discontinuation2_begin <- 2
tx_duration <- 29

# Treatment effect waning
tx_waning <- 0
tx_waning_dis <- 0

# Utilities - Patient (community)
u_mci_pt <- 0.851 - 0.17
u_mil_pt <- 0.851 - 0.22
u_mod_pt <- 0.851 - 0.36
u_sev_pt <- 0.851 - 0.53

# Utilities - Patient (institution)
u_mci_pt_i <- 0.851 - 0.17
u_mil_pt_i <- 0.851 - 0.19
u_mod_pt_i <- 0.851 - 0.42
u_sev_pt_i <- 0.851 - 0.59

# Utilities - Informal caregiver (community)
u_mci_ic <- -0.03
u_mil_ic <- -0.05
u_mod_ic <- -0.08
u_sev_ic <- -0.10

# Utilities - Informal caregiver (institution)
u_mci_ic_i <- -0.03
u_mil_ic_i <- -0.05
u_mod_ic_i <- -0.08
u_sev_ic_i <- -0.10

# Additional utility decrement in first cycle
u_Tx_start <- -0.14 * (12 / 52) * 0.035

# Costs - Health care (community)
c_mci_hc <- 6042 * 1.12 + 460
c_mil_hc <- 6042 * 1.56 + 965 + 0.21 * 365 * 0.333
c_mod_hc <- 6042 * 1.93 + 1544 + 0.66 * 365 * 0.333
c_sev_hc <- 6042 * 1.93 + 1930

# Costs - Health care (institution)
c_mci_i_hc <- c_mci_hc
c_mil_i_hc <- c_mil_hc
c_mod_i_hc <- c_mod_hc
c_sev_i_hc <- c_sev_hc

# Costs - Social care (community)
c_mci_sc <- 0
c_mil_sc <- 0
c_mod_sc <- 0
c_sev_sc <- 0

# Costs - Social care (institution)
c_mci_i_sc <- 7394 * 12
c_mil_i_sc <- 7394 * 12
c_mod_i_sc <- 7394 * 12
c_sev_i_sc <- 7394 * 12

# Costs - Informal care (community)
c_mci_ic <- 69 * 12 * 32.46 + 0.204 * 0.049 * 20 * 52 * 32.46
c_mil_ic <- 113 * 12 * 32.46 + 0.112 * 0.086 * 20 * 52 * 32.46
c_mod_ic <- 169 * 12 * 32.46
c_sev_ic <- 298 * 12 * 32.46

# Costs - Informal care (institution, 44% of community)
c_mci_i_ic <- c_mci_ic * 0.44
c_mil_i_ic <- c_mil_ic * 0.44
c_mod_i_ic <- c_mod_ic * 0.44
c_sev_i_ic <- c_sev_ic * 0.44

# Treatment cost (annual)
c_Tx <- 26500 + (52 / 2) * 78.35

# Additional treatment costs (monitoring MRI scans, ARIA-related)
c_Tx_start <- 261.10 * 4 + 261.10 * 3 * 0.215

# Discounting
discount_EFFECT <- 0.03
discount_QALY <- 0.03
discount_COST <- 0.03

# Willingness to pay threshold
wtp <- 100000

# Half-cycle correction flag
half_cycle_correction <- TRUE
# Load the life table CSV file
lifetable <- read.csv("lifetable_US_2019_ssa.csv", stringsAsFactors = FALSE)

# Convert annual mortality probabilities to mortality rates using: rate = -log(1 - probability)
lifetable$male <- -log(1 - lifetable$male)
lifetable$female <- -log(1 - lifetable$female)

# Compute weighted average mortality rate (52% female, 48% male)
lifetable$weighted <- 0.48 * lifetable$male + 0.52 * lifetable$female

# Return the required columns
lifetable <- lifetable[, c("age", "male", "female", "weighted")]

# Display the first few rows (optional)
head(lifetable)
# Number of cycles
n_cycles <- 29

# Time-dependent disease progression probabilities for "soc" strategy
v.p.mci_mil_soc <- rep(p_mci_mil, n_cycles)
v.p.mci_mod_soc <- rep(p_mci_mod, n_cycles)
v.p.mci_sev_soc <- rep(p_mci_sev, n_cycles)

v.p.mil_mci_soc <- rep(p_mil_mci, n_cycles)
v.p.mil_mod_soc <- rep(p_mil_mod, n_cycles)
v.p.mil_sev_soc <- rep(p_mil_sev, n_cycles)

v.p.mod_mil_soc <- rep(p_mod_mil, n_cycles)
v.p.mod_sev_soc <- rep(p_mod_sev, n_cycles)

v.p.sev_mil_soc <- rep(p_sev_mil, n_cycles)
v.p.sev_mod_soc <- rep(p_sev_mod, n_cycles)

# Staying probabilities per state (excluding institutional transitions and death)
v.p.mci_stay_soc <- 1 - (v.p.mci_mil_soc + v.p.mci_mod_soc + v.p.mci_sev_soc)
v.p.mil_stay_soc <- 1 - (v.p.mil_mci_soc + v.p.mil_mod_soc + v.p.mil_sev_soc)
v.p.mod_stay_soc <- 1 - (v.p.mod_mil_soc + v.p.mod_sev_soc)
v.p.sev_stay_soc <- 1 - (v.p.sev_mil_soc + v.p.sev_mod_soc)

# Transition to institutional care setting (fixed over time)
v.p.mci_i <- rep(p_mci_i, n_cycles)
v.p.mil_i <- rep(p_mil_i, n_cycles)
v.p.mod_i <- rep(p_mod_i, n_cycles)
v.p.sev_i <- rep(p_sev_i, n_cycles)

# Discontinuation vector (applies only to intervention, but included here for completeness)
v.p.discontinuation <- rep(0, n_cycles)
v.p.discontinuation[1:(discontinuation2_begin - 1)] <- p_discontinuation1
v.p.discontinuation[discontinuation2_begin:(tx_duration - 1)] <- p_discontinuation2
if (tx_duration <= n_cycles) {
  v.p.discontinuation[tx_duration:n_cycles] <- 1
}

# Age-specific general population mortality vector from life table (weighted column)
# Assumes lifetable object has already been created and contains a column named "weighted"
v.mort_soc <- lifetable$weighted[match(age_start:(age_start + n_cycles - 1), lifetable$age)]
# Define time-dependent transition probabilities for intervention ("int") strategy

# Create cycle index vector
cycle_index <- 1:n_cycles

# ---- On-treatment: from "mcion_c" ----
rate_mci_mil <- -log(1 - p_mci_mil)
v.p.mci_mil_int <- 1 - exp(-rate_mci_mil * rr_tx_mci_mil ^ ((1 - tx_waning) ^ (cycle_index - 1)))

rate_mci_mod <- -log(1 - p_mci_mod)
v.p.mci_mod_int <- 1 - exp(-rate_mci_mod * rr_tx_mci_mod ^ ((1 - tx_waning) ^ (cycle_index - 1)))

rate_mci_sev <- -log(1 - p_mci_sev)
v.p.mci_sev_int <- 1 - exp(-rate_mci_sev * rr_tx_mci_sev ^ ((1 - tx_waning) ^ (cycle_index - 1)))

v.p.mci_stay_int <- 1 - (v.p.mci_mil_int + v.p.mci_mod_int + v.p.mci_sev_int)

# ---- On-treatment: from "milon_c" ----
rate_mil_mci <- -log(1 - p_mil_mci)  # backward transition, no treatment effect
v.p.mil_mci_int <- rep(1 - exp(-rate_mil_mci), n_cycles)

rate_mil_mod <- -log(1 - p_mil_mod)
v.p.mil_mod_int <- 1 - exp(-rate_mil_mod * rr_tx_mil_mod ^ ((1 - tx_waning) ^ (cycle_index - 1)))

rate_mil_sev <- -log(1 - p_mil_sev)
v.p.mil_sev_int <- 1 - exp(-rate_mil_sev * rr_tx_mil_sev ^ ((1 - tx_waning) ^ (cycle_index - 1)))

v.p.mil_stay_int <- 1 - (v.p.mil_mci_int + v.p.mil_mod_int + v.p.mil_sev_int)

# ---- Post-discontinuation: from "mciof_c" ----
v.p.mci_mil_int_dis <- 1 - exp(-rate_mci_mil * rr_tx_mci_mil_dis ^ ((1 - tx_waning_dis) ^ (cycle_index - 1)))
v.p.mci_mod_int_dis <- 1 - exp(-rate_mci_mod * rr_tx_mci_mod_dis ^ ((1 - tx_waning_dis) ^ (cycle_index - 1)))
v.p.mci_sev_int_dis <- 1 - exp(-rate_mci_sev * rr_tx_mci_sev_dis ^ ((1 - tx_waning_dis) ^ (cycle_index - 1)))

v.p.mci_stay_int_dis <- 1 - (v.p.mci_mil_int_dis + v.p.mci_mod_int_dis + v.p.mci_sev_int_dis)

# ---- Post-discontinuation: from "milof_c" ----
v.p.mil_mci_int_dis <- rep(1 - exp(-rate_mil_mci), n_cycles)

v.p.mil_mod_int_dis <- 1 - exp(-rate_mil_mod * rr_tx_mil_mod_dis ^ ((1 - tx_waning_dis) ^ (cycle_index - 1)))
v.p.mil_sev_int_dis <- 1 - exp(-rate_mil_sev * rr_tx_mil_sev_dis ^ ((1 - tx_waning_dis) ^ (cycle_index - 1)))

v.p.mil_stay_int_dis <- 1 - (v.p.mil_mci_int_dis + v.p.mil_mod_int_dis + v.p.mil_sev_int_dis)


# Function to build transition probability matrix array
build_transition_matrix <- function(strat) {
  # Initialize 3D array: states x states x cycles
  a.TP <- array(0, dim = c(length(state_names), length(state_names), n_cycles),
                dimnames = list(state_names, state_names, NULL))
  
  for (t in 1:n_cycles) {
    # Mortality rates for this cycle
    mort_t <- v.mort_soc[t]
    
    # Transitions to death from each state (explicitly)
    a.TP["mcion_c", "dth", t]   <- 1 - exp(-mort_t * hr_mort_mci)
    a.TP["mciof_c", "dth", t]   <- 1 - exp(-mort_t * hr_mort_mci)
    a.TP["milon_c", "dth", t]   <- 1 - exp(-mort_t * hr_mort_mil)
    a.TP["milof_c", "dth", t]   <- 1 - exp(-mort_t * hr_mort_mil)
    a.TP["mod_c", "dth", t]     <- 1 - exp(-mort_t * hr_mort_mod)
    a.TP["sev_c", "dth", t]     <- 1 - exp(-mort_t * hr_mort_sev)
    a.TP["mci_i", "dth", t]     <- 1 - exp(-mort_t * hr_mort_mci)
    a.TP["mil_i", "dth", t]     <- 1 - exp(-mort_t * hr_mort_mil)
    a.TP["mod_i", "dth", t]     <- 1 - exp(-mort_t * hr_mort_mod)
    a.TP["sev_i", "dth", t]     <- 1 - exp(-mort_t * hr_mort_sev)
    
    # Absorbing death state
    a.TP["dth", "dth", t] <- 1
    
    # Transitions from "mcion_c" (on-treatment MCI, community)
    a.TP["mcion_c", "mcion_c", t] <- if (strat == "soc") {
      v.p.mci_stay_soc[t] * (1 - a.TP["mcion_c", "dth", t]) * (1 - v.p.mci_i[t]) * (1 - v.p.discontinuation[t])
    } else {
      v.p.mci_stay_int[t] * (1 - a.TP["mcion_c", "dth", t]) * (1 - v.p.mci_i[t]) * (1 - v.p.discontinuation[t])
    }
    
    a.TP["mcion_c", "mciof_c", t] <- if (strat == "soc") {
      v.p.mci_stay_soc[t] * (1 - a.TP["mcion_c", "dth", t]) * (1 - v.p.mci_i[t]) * v.p.discontinuation[t]
    } else {
      v.p.mci_stay_int[t] * (1 - a.TP["mcion_c", "dth", t]) * (1 - v.p.mci_i[t]) * v.p.discontinuation[t]
    }
    
    a.TP["mcion_c", "milon_c", t] <- if (strat == "soc") {
      v.p.mci_mil_soc[t] * (1 - a.TP["mcion_c", "dth", t]) * (1 - v.p.mci_i[t]) * (1 - v.p.discontinuation[t])
    } else {
      v.p.mci_mil_int[t] * (1 - a.TP["mcion_c", "dth", t]) * (1 - v.p.mci_i[t]) * (1 - v.p.discontinuation[t])
    }
    
    a.TP["mcion_c", "mciof_c", t] <- a.TP["mcion_c", "mciof_c", t] + if (strat == "soc") {
      v.p.mci_mil_soc[t] * (1 - a.TP["mcion_c", "dth", t]) * (1 - v.p.mci_i[t]) * v.p.discontinuation[t]
    } else {
      v.p.mci_mil_int[t] * (1 - a.TP["mcion_c", "dth", t]) * (1 - v.p.mci_i[t]) * v.p.discontinuation[t]
    }
    
    a.TP["mcion_c", "mod_c", t] <- if (strat == "soc") {
      v.p.mci_mod_soc[t] * (1 - a.TP["mcion_c", "dth", t]) * (1 - v.p.mci_i[t])
    } else {
      v.p.mci_mod_int[t] * (1 - a.TP["mcion_c", "dth", t]) * (1 - v.p.mci_i[t])
    }
    
    a.TP["mcion_c", "sev_c", t] <- if (strat == "soc") {
      v.p.mci_sev_soc[t] * (1 - a.TP["mcion_c", "dth", t]) * (1 - v.p.mci_i[t])
    } else {
      v.p.mci_sev_int[t] * (1 - a.TP["mcion_c", "dth", t]) * (1 - v.p.mci_i[t])
    }
    
    a.TP["mcion_c", "mci_i", t] <- if (strat == "soc") {
      (v.p.mci_stay_soc[t] + v.p.mci_mil_soc[t] + v.p.mci_mod_soc[t] + v.p.mci_sev_soc[t]) *
        (1 - a.TP["mcion_c", "dth", t]) * v.p.mci_i[t]
    } else {
      (v.p.mci_stay_int[t] + v.p.mci_mil_int[t] + v.p.mci_mod_int[t] + v.p.mci_sev_int[t]) *
        (1 - a.TP["mcion_c", "dth", t]) * v.p.mci_i[t]
    }
    
    # Transitions from "milon_c" (on-treatment mild, community)
    a.TP["milon_c", "milon_c", t] <- if (strat == "soc") {
      v.p.mil_stay_soc[t] * (1 - a.TP["milon_c", "dth", t]) * (1 - v.p.mil_i[t]) * (1 - v.p.discontinuation[t])
    } else {
      v.p.mil_stay_int[t] * (1 - a.TP["milon_c", "dth", t]) * (1 - v.p.mil_i[t]) * (1 - v.p.discontinuation[t])
    }
    
    a.TP["milon_c", "milof_c", t] <- if (strat == "soc") {
      v.p.mil_stay_soc[t] * (1 - a.TP["milon_c", "dth", t]) * (1 - v.p.mil_i[t]) * v.p.discontinuation[t]
    } else {
      v.p.mil_stay_int[t] * (1 - a.TP["milon_c", "dth", t]) * (1 - v.p.mil_i[t]) * v.p.discontinuation[t]
    }
    
    a.TP["milon_c", "mciof_c", t] <- if (strat == "soc") {
      v.p.mil_mci_soc[t] * (1 - a.TP["milon_c", "dth", t]) * (1 - v.p.mil_i[t])
    } else {
      v.p.mil_mci_int[t] * (1 - a.TP["milon_c", "dth", t]) * (1 - v.p.mil_i[t])
    }
    
    a.TP["milon_c", "mod_c", t] <- if (strat == "soc") {
      v.p.mil_mod_soc[t] * (1 - a.TP["milon_c", "dth", t]) * (1 - v.p.mil_i[t])
    } else {
      v.p.mil_mod_int[t] * (1 - a.TP["milon_c", "dth", t]) * (1 - v.p.mil_i[t])
    }
    
    a.TP["milon_c", "sev_c", t] <- if (strat == "soc") {
      v.p.mil_sev_soc[t] * (1 - a.TP["milon_c", "dth", t]) * (1 - v.p.mil_i[t])
    } else {
      v.p.mil_sev_int[t] * (1 - a.TP["milon_c", "dth", t]) * (1 - v.p.mil_i[t])
    }
    
    a.TP["milon_c", "mil_i", t] <- if (strat == "soc") {
      (v.p.mil_stay_soc[t] + v.p.mil_mci_soc[t] + v.p.mil_mod_soc[t] + v.p.mil_sev_soc[t]) *
        (1 - a.TP["milon_c", "dth", t]) * v.p.mil_i[t]
    } else {
      (v.p.mil_stay_int[t] + v.p.mil_mci_int[t] + v.p.mil_mod_int[t] + v.p.mil_sev_int[t]) *
        (1 - a.TP["milon_c", "dth", t]) * v.p.mil_i[t]
    }
    
    
    # Transitions from "mciof_c" (off-treatment MCI, community)
    a.TP["mciof_c", "mciof_c", t] <- if (strat == "soc") {
      v.p.mci_stay_soc[t] * (1 - a.TP["mciof_c", "dth", t]) * (1 - v.p.mci_i[t])
    } else {
      v.p.mci_stay_int_dis[t] * (1 - a.TP["mciof_c", "dth", t]) * (1 - v.p.mci_i[t])
    }
    
    a.TP["mciof_c", "milof_c", t] <- if (strat == "soc") {
      v.p.mci_mil_soc[t] * (1 - a.TP["mciof_c", "dth", t]) * (1 - v.p.mci_i[t])
    } else {
      v.p.mci_mil_int_dis[t] * (1 - a.TP["mciof_c", "dth", t]) * (1 - v.p.mci_i[t])
    }
    
    a.TP["mciof_c", "mod_c", t] <- if (strat == "soc") {
      v.p.mci_mod_soc[t] * (1 - a.TP["mciof_c", "dth", t]) * (1 - v.p.mci_i[t])
    } else {
      v.p.mci_mod_int_dis[t] * (1 - a.TP["mciof_c", "dth", t]) * (1 - v.p.mci_i[t])
    }
    
    a.TP["mciof_c", "sev_c", t] <- if (strat == "soc") {
      v.p.mci_sev_soc[t] * (1 - a.TP["mciof_c", "dth", t]) * (1 - v.p.mci_i[t])
    } else {
      v.p.mci_sev_int_dis[t] * (1 - a.TP["mciof_c", "dth", t]) * (1 - v.p.mci_i[t])
    }
    
    a.TP["mciof_c", "mci_i", t] <- if (strat == "soc") {
      (v.p.mci_stay_soc[t] + v.p.mci_mil_soc[t] + v.p.mci_mod_soc[t] + v.p.mci_sev_soc[t]) *
        (1 - a.TP["mciof_c", "dth", t]) * v.p.mci_i[t]
    } else {
      (v.p.mci_stay_int_dis[t] + v.p.mci_mil_int_dis[t] + v.p.mci_mod_int_dis[t] + v.p.mci_sev_int_dis[t]) *
        (1 - a.TP["mciof_c", "dth", t]) * v.p.mci_i[t]
    }
    
    # Transitions from "milof_c" (off-treatment mild, community)
    a.TP["milof_c", "milof_c", t] <- if (strat == "soc") {
      v.p.mil_stay_soc[t] * (1 - a.TP["milof_c", "dth", t]) * (1 - v.p.mil_i[t])
    } else {
      v.p.mil_stay_int_dis[t] * (1 - a.TP["milof_c", "dth", t]) * (1 - v.p.mil_i[t])
    }
    
    a.TP["milof_c", "mciof_c", t] <- if (strat == "soc") {
      v.p.mil_mci_soc[t] * (1 - a.TP["milof_c", "dth", t]) * (1 - v.p.mil_i[t])
    } else {
      v.p.mil_mci_int_dis[t] * (1 - a.TP["milof_c", "dth", t]) * (1 - v.p.mil_i[t])
    }
    
    a.TP["milof_c", "mod_c", t] <- if (strat == "soc") {
      v.p.mil_mod_soc[t] * (1 - a.TP["milof_c", "dth", t]) * (1 - v.p.mil_i[t])
    } else {
      v.p.mil_mod_int_dis[t] * (1 - a.TP["milof_c", "dth", t]) * (1 - v.p.mil_i[t])
    }
    
    a.TP["milof_c", "sev_c", t] <- if (strat == "soc") {
      v.p.mil_sev_soc[t] * (1 - a.TP["milof_c", "dth", t]) * (1 - v.p.mil_i[t])
    } else {
      v.p.mil_sev_int_dis[t] * (1 - a.TP["milof_c", "dth", t]) * (1 - v.p.mil_i[t])
    }
    
    a.TP["milof_c", "mil_i", t] <- if (strat == "soc") {
      (v.p.mil_stay_soc[t] + v.p.mil_mci_soc[t] + v.p.mil_mod_soc[t] + v.p.mil_sev_soc[t]) *
        (1 - a.TP["milof_c", "dth", t]) * v.p.mil_i[t]
    } else {
      (v.p.mil_stay_int_dis[t] + v.p.mil_mci_int_dis[t] + v.p.mil_mod_int_dis[t] + v.p.mil_sev_int_dis[t]) *
        (1 - a.TP["milof_c", "dth", t]) * v.p.mil_i[t]
    }
    
    # Transitions from "mod_c" (moderate, community)
    a.TP["mod_c", "mod_c", t] <- v.p.mod_stay_soc[t] * (1 - a.TP["mod_c", "dth", t]) * (1 - v.p.mod_i[t])
    a.TP["mod_c", "milof_c", t] <- v.p.mod_mil_soc[t] * (1 - a.TP["mod_c", "dth", t]) * (1 - v.p.mod_i[t])
    a.TP["mod_c", "sev_c", t] <- v.p.mod_sev_soc[t] * (1 - a.TP["mod_c", "dth", t]) * (1 - v.p.mod_i[t])
    a.TP["mod_c", "mod_i", t] <- (v.p.mod_stay_soc[t] + v.p.mod_mil_soc[t] + v.p.mod_sev_soc[t]) *
      (1 - a.TP["mod_c", "dth", t]) * v.p.mod_i[t]
    
    # Transitions from "sev_c" (severe, community)
    a.TP["sev_c", "sev_c", t] <- v.p.sev_stay_soc[t] * (1 - a.TP["sev_c", "dth", t]) * (1 - v.p.sev_i[t])
    a.TP["sev_c", "milof_c", t] <- v.p.sev_mil_soc[t] * (1 - a.TP["sev_c", "dth", t]) * (1 - v.p.sev_i[t])
    a.TP["sev_c", "mod_c", t] <- v.p.sev_mod_soc[t] * (1 - a.TP["sev_c", "dth", t]) * (1 - v.p.sev_i[t])
    a.TP["sev_c", "sev_i", t] <- (v.p.sev_stay_soc[t] + v.p.sev_mil_soc[t] + v.p.sev_mod_soc[t]) *
      (1 - a.TP["sev_c", "dth", t]) * v.p.sev_i[t]
    
    
    # Transitions from "mci_i" (institutionalized MCI)
    a.TP["mci_i", "mci_i", t] <- v.p.mci_stay_soc[t] * (1 - a.TP["mci_i", "dth", t])
    a.TP["mci_i", "mil_i", t] <- v.p.mci_mil_soc[t] * (1 - a.TP["mci_i", "dth", t])
    a.TP["mci_i", "mod_i", t] <- v.p.mci_mod_soc[t] * (1 - a.TP["mci_i", "dth", t])
    a.TP["mci_i", "sev_i", t] <- v.p.mci_sev_soc[t] * (1 - a.TP["mci_i", "dth", t])
    
    # Transitions from "mil_i" (institutionalized mild)
    a.TP["mil_i", "mil_i", t] <- v.p.mil_stay_soc[t] * (1 - a.TP["mil_i", "dth", t])
    a.TP["mil_i", "mci_i", t] <- v.p.mil_mci_soc[t] * (1 - a.TP["mil_i", "dth", t])
    a.TP["mil_i", "mod_i", t] <- v.p.mil_mod_soc[t] * (1 - a.TP["mil_i", "dth", t])
    a.TP["mil_i", "sev_i", t] <- v.p.mil_sev_soc[t] * (1 - a.TP["mil_i", "dth", t])
    
    # Transitions from "mod_i" (institutionalized moderate)
    a.TP["mod_i", "mod_i", t] <- v.p.mod_stay_soc[t] * (1 - a.TP["mod_i", "dth", t])
    a.TP["mod_i", "mil_i", t] <- v.p.mod_mil_soc[t] * (1 - a.TP["mod_i", "dth", t])
    a.TP["mod_i", "sev_i", t] <- v.p.mod_sev_soc[t] * (1 - a.TP["mod_i", "dth", t])
    
    # Transitions from "sev_i" (institutionalized severe)
    a.TP["sev_i", "sev_i", t] <- v.p.sev_stay_soc[t] * (1 - a.TP["sev_i", "dth", t])
    a.TP["sev_i", "mil_i", t] <- v.p.sev_mil_soc[t] * (1 - a.TP["sev_i", "dth", t])
    a.TP["sev_i", "mod_i", t] <- v.p.sev_mod_soc[t] * (1 - a.TP["sev_i", "dth", t])
    
    # Normalize each row to sum to 1 if necessary
    for (s in state_names) {
      row_sum <- sum(a.TP[s, , t])
      if (row_sum > 0 && abs(row_sum - 1) > 1e-8) {
        a.TP[s, , t] <- a.TP[s, , t] / row_sum
      }
    }
  }
  
  return(a.TP)
}


# Function to compute outcomes for a given strategy
compute_outcomes <- function(strat) {
  # Build transition matrix
  a.TP <- build_transition_matrix(strat)
  
  # Initialize trace matrix
  m.trace <- matrix(0, nrow = n_cycles + 1, ncol = length(state_names),
                    dimnames = list(0:n_cycles, state_names))
  
  # Initial state distribution
  v.init <- rep(0, length(state_names))
  names(v.init) <- state_names
  v.init["mciof_c"] <- 1 - p_starting_state_mci
  v.init["mcion_c"] <- p_starting_state_mci
  m.trace[1, ] <- v.init
  
  # Utility vector per state
  v.u <- c(mcion_c = u_mci_pt, mciof_c = u_mci_pt,
           milon_c = u_mil_pt, milof_c = u_mil_pt,
           mod_c = u_mod_pt, sev_c = u_sev_pt,
           mci_i = u_mci_pt_i, mil_i = u_mil_pt_i,
           mod_i = u_mod_pt_i, sev_i = u_sev_pt_i,
           dth = 0)
  
  # Cost vector per state (excluding treatment cost)
  v.c <- c(mcion_c = c_mci_hc + c_mci_sc + c_mci_ic,
           mciof_c = c_mci_hc + c_mci_sc + c_mci_ic,
           milon_c = c_mil_hc + c_mil_sc + c_mil_ic,
           milof_c = c_mil_hc + c_mil_sc + c_mil_ic,
           mod_c = c_mod_hc + c_mod_sc + c_mod_ic,
           sev_c = c_sev_hc + c_sev_sc + c_sev_ic,
           mci_i = c_mci_i_hc + c_mci_i_sc + c_mci_i_ic,
           mil_i = c_mil_i_hc + c_mil_i_sc + c_mil_i_ic,
           mod_i = c_mod_i_hc + c_mod_i_sc + c_mod_i_ic,
           sev_i = c_sev_i_hc + c_sev_i_sc + c_sev_i_ic,
           dth = 0)
  
  # Treatment cost vector (recurring each cycle, intervention only)
  v.c_tx <- rep(0, length(state_names))
  names(v.c_tx) <- state_names
  if (strat == "int") {
    v.c_tx["mcion_c"] <- c_Tx
    v.c_tx["milon_c"] <- c_Tx
  }
  
  # Initialize vectors to store outcomes
  v.QALYs <- numeric(n_cycles + 1)
  v.Costs <- numeric(n_cycles + 1)
  v.LYs <- numeric(n_cycles + 1)
  
  # Loop over each cycle to compute outcomes
  for (t in 1:n_cycles) {
    m.trace[t + 1, ] <- m.trace[t, ] %*% a.TP[ , , t]
  }
  
  # Compute outcomes per cycle
  for (t in 1:(n_cycles + 1)) {
    v.QALYs[t] <- sum(m.trace[t, ] * v.u)
    v.Costs[t] <- sum(m.trace[t, ] * (v.c + v.c_tx))
    v.LYs[t] <- sum(m.trace[t, state_names != "dth"])
  }
  
  # Apply initial treatment disutility and cost in first cycle (if intervention)
  if (strat == "int") {
    v.QALYs[1] <- v.QALYs[1] + u_Tx_start
    v.Costs[1] <- v.Costs[1] + c_Tx_start
  }
  
  # Apply half-cycle correction (average each cycle with next cycle)
  v.QALYs <- (v.QALYs[-length(v.QALYs)] + v.QALYs[-1]) / 2
  v.Costs <- (v.Costs[-length(v.Costs)] + v.Costs[-1]) / 2
  v.LYs <- (v.LYs[-length(v.LYs)] + v.LYs[-1]) / 2
  
  # Apply discounting
  v.disc <- 1 / ((1 + discount_QALY) ^ (0:(n_cycles - 1)))
  total_QALYs <- sum(v.QALYs * v.disc)
  total_LYs <- sum(v.LYs * v.disc)
  total_Costs <- sum(v.Costs * (1 / ((1 + discount_COST) ^ (0:(n_cycles - 1)))))
  
  # Return total outcomes
  return(list(QALYs = total_QALYs, LYs = total_LYs, Costs = total_Costs))
}

# Run both strategies and compare
res_soc <- compute_outcomes("soc")
res_int <- compute_outcomes("int")

# Print results
cat("----- RESULTS -----\n")
cat("Standard of Care (SOC):\n")
cat(sprintf("  QALYs: %.2f\n  LYs: %.2f\n  Costs: $%.0f\n", res_soc$QALYs, res_soc$LYs, res_soc$Costs))

cat("\nIntervention (INT):\n")
cat(sprintf("  QALYs: %.2f\n  LYs: %.2f\n  Costs: $%.0f\n", res_int$QALYs, res_int$LYs, res_int$Costs))

cat("\nIncremental (INT - SOC):\n")
cat(sprintf("  ΔQALYs: %.2f\n  ΔLYs: %.2f\n  ΔCosts: $%.0f\n",
            res_int$QALYs - res_soc$QALYs,
            res_int$LYs - res_soc$LYs,
            res_int$Costs - res_soc$Costs))
```
