## Supplementary material for "Replicating Health-Economic Simulation Models for Alzheimer’s Disease Using Artificial Intelligence": final R scripts: o13.R

```
# IPECAD Markov Model Input Parameters for Use with dampack

# Strategy and state names
v.names_state <- c("mcion_c", "mciof_c", "milon_c", "milof_c", "mod_c", "sev_c", "mci_i", "mil_i", "mod_i", "sev_i", "dth")
v.names_strat <- c("soc", "int")

# Demographic and model settings
age_start <- 71
sex <- "weighted"  # 52% female
p.starting_state_mci <- 0.55
n.cycle <- 29

# Transition probabilities for disease progression (community)
p.mci_mil <- 0.23
p.mci_mod <- 0
p.mci_sev <- 0
p.mil_mci <- 0.03
p.mil_mod <- 0.35
p.mil_sev <- 0.04
p.mod_mil <- 0.03
p.mod_sev <- 0.42
p.sev_mil <- 0
p.sev_mod <- 0.02

# Transition probabilities to institutional care
p.mci_i <- 0.024
p.mil_i <- 0.038
p.mod_i <- 0.110
p.sev_i <- 0.259

# Mortality multipliers relative to general population
hr.mort_mci <- 1.82
hr.mort_mil <- 2.92
hr.mort_mod <- 3.85
hr.mort_sev <- 9.52

# Treatment effect relative risks (on-treatment)
rr.tx_mci_mil <- 0.69
rr.tx_mci_mod <- 1
rr.tx_mci_sev <- 1
rr.tx_mil_mod <- 0.69
rr.tx_mil_sev <- 0.69

# Treatment effect relative risks (off-treatment / post-discontinuation)
rr.tx_mci_mil_dis <- 1
rr.tx_mci_mod_dis <- 1
rr.tx_mci_sev_dis <- 1
rr.tx_mil_mod_dis <- 1
rr.tx_mil_sev_dis <- 1

# Discontinuation probabilities
p.discontinuation1 <- 0.069
p.discontinuation2 <- 0
discontinuation2_begin <- 2
tx_duration <- 29
tx_waning <- 0
tx_waning_dis <- 0

# Utilities for patients (community and institutional)
u.mci_pt <- 0.851 - 0.17
u.mil_pt <- 0.851 - 0.22
u.mod_pt <- 0.851 - 0.36
u.sev_pt <- 0.851 - 0.53
u.mci_pt_i <- 0.851 - 0.17
u.mil_pt_i <- 0.851 - 0.19
u.mod_pt_i <- 0.851 - 0.42
u.sev_pt_i <- 0.851 - 0.59

# Utilities for informal caregivers (community and institutional)
u.mci_ic <- -0.03
u.mil_ic <- -0.05
u.mod_ic <- -0.08
u.sev_ic <- -0.10
u.mci_ic_i <- -0.03
u.mil_ic_i <- -0.05
u.mod_ic_i <- -0.08
u.sev_ic_i <- -0.10

# Additional utility loss from treatment adverse events in the first cycle
u.Tx_start <- -0.14 * (12/52) * 0.035

# Costs for health care (community)
c.mci_hc <- 6042*1.12 + 460
c.mil_hc <- 6042*1.56 + 965 + 0.21*365*0.333
c.mod_hc <- 6042*1.93 + 1544 + 0.66*365*0.333
c.sev_hc <- 6042*1.93 + 1930

# Costs for health care (institutional)
c.mci_i_hc <- c.mci_hc
c.mil_i_hc <- c.mil_hc
c.mod_i_hc <- c.mod_hc
c.sev_i_hc <- c.sev_hc

# Social care costs (institution only)
c.mci_sc <- 0
c.mil_sc <- 0
c.mod_sc <- 0
c.sev_sc <- 0
c.mci_i_sc <- 7394 * 12
c.mil_i_sc <- 7394 * 12
c.mod_i_sc <- 7394 * 12
c.sev_i_sc <- 7394 * 12

# Informal care costs (community)
c.mci_ic <- 69*12*32.46 + 0.204*0.049*20*52*32.46
c.mil_ic <- 113*12*32.46 + 0.112*0.086*20*52*32.46
c.mod_ic <- 169*12*32.46
c.sev_ic <- 298*12*32.46

# Informal care costs (institution, 44% of community)
c.mci_i_ic <- 69*12*32.46*0.44 + 0.204*0.049*20*52*32.46
c.mil_i_ic <- 113*12*32.46*0.44 + 0.112*0.086*20*52*32.46
c.mod_i_ic <- 169*12*32.46*0.44
c.sev_i_ic <- 298*12*32.46*0.44

# Treatment and monitoring costs
c.Tx <- 26500 + (52/2)*78.35
c.Tx_start <- 261.10*4 + 261.10*3*0.215

# Discounting parameters
discount_EFFECT <- 0.03
discount_QALY <- 0.03
discount_COST <- 0.03

# Willingness-to-pay threshold
wtp <- 100000

# Half-cycle correction flag
half_cycle_correction <- TRUE
# Load the 2019 US life table from CSV
lifetable <- read.csv("lifetable_US_2019_ssa.csv", stringsAsFactors = FALSE)

# Convert annual mortality probabilities to annual mortality rates using:
# rate = -log(1 - probability)
lifetable$male <- -log(1 - lifetable$male)
lifetable$female <- -log(1 - lifetable$female)

# Define weighted average based on 52% female, 48% male (from the main model parameter)
lifetable$weighted <- 0.48 * lifetable$male + 0.52 * lifetable$female

# Return the processed dataframe with rates
lifetable <- lifetable[, c("age", "male", "female", "weighted")]
# Define time-dependent transition probability vectors for the "soc" strategy
v.p.mci_mil_soc <- rep(p.mci_mil, n.cycle)
v.p.mci_mod_soc <- rep(p.mci_mod, n.cycle)
v.p.mci_sev_soc <- rep(p.mci_sev, n.cycle)

v.p.mil_mci_soc <- rep(p.mil_mci, n.cycle)
v.p.mil_mod_soc <- rep(p.mil_mod, n.cycle)
v.p.mil_sev_soc <- rep(p.mil_sev, n.cycle)

v.p.mod_mil_soc <- rep(p.mod_mil, n.cycle)
v.p.mod_sev_soc <- rep(p.mod_sev, n.cycle)

v.p.sev_mil_soc <- rep(p.sev_mil, n.cycle)
v.p.sev_mod_soc <- rep(p.sev_mod, n.cycle)

# Staying probabilities per state (excluding institutional transitions and death)
v.p.mci_mci_soc <- 1 - v.p.mci_mil_soc - v.p.mci_mod_soc - v.p.mci_sev_soc
v.p.mil_mil_soc <- 1 - v.p.mil_mci_soc - v.p.mil_mod_soc - v.p.mil_sev_soc
v.p.mod_mod_soc <- 1 - v.p.mod_mil_soc - v.p.mod_sev_soc
v.p.sev_sev_soc <- 1 - v.p.sev_mil_soc - v.p.sev_mod_soc

# Transition probabilities to institutional care (time-independent)
v.p.mci_i_soc <- rep(p.mci_i, n.cycle)
v.p.mil_i_soc <- rep(p.mil_i, n.cycle)
v.p.mod_i_soc <- rep(p.mod_i, n.cycle)
v.p.sev_i_soc <- rep(p.sev_i, n.cycle)

# Discontinuation vector per cycle (applies only to on-treatment community MCI and mild)
v.p.discontinuation_soc <- c(
  rep(p.discontinuation1, discontinuation2_begin - 1),
  rep(p.discontinuation2, tx_duration - discontinuation2_begin + 1),
  rep(1, max(0, n.cycle - tx_duration))
)

# Vector of age-specific mortality rates from weighted life table
# Assumes variable `lifetable` from prompt 2 is available and includes age and weighted rate
v.mort_rate <- lifetable$weighted[match(age_start:(age_start + n.cycle - 1), lifetable$age)]
# Initialize on-treatment transition vectors for "int" strategy (from mcion_c)
v.p.mci_mil_int <- numeric(n.cycle)
v.p.mci_mod_int <- numeric(n.cycle)
v.p.mci_sev_int <- numeric(n.cycle)
v.p.mci_mci_int <- numeric(n.cycle)

# Initialize post-discontinuation transition vectors (from mciof_c)
v.p.mci_mil_int_dis <- numeric(n.cycle)
v.p.mci_mod_int_dis <- numeric(n.cycle)
v.p.mci_sev_int_dis <- numeric(n.cycle)
v.p.mci_mci_int_dis <- numeric(n.cycle)

# Initialize on-treatment transition vectors (from milon_c)
v.p.mil_mci_int <- rep(p.mil_mci, n.cycle)  # backward transition, no treatment effect
v.p.mil_mod_int <- numeric(n.cycle)
v.p.mil_sev_int <- numeric(n.cycle)
v.p.mil_mil_int <- numeric(n.cycle)

# Initialize post-discontinuation transition vectors (from milof_c)
v.p.mil_mci_int_dis <- rep(p.mil_mci, n.cycle)  # backward transition, no treatment effect
v.p.mil_mod_int_dis <- numeric(n.cycle)
v.p.mil_sev_int_dis <- numeric(n.cycle)
v.p.mil_mil_int_dis <- numeric(n.cycle)

# Compute on-treatment and post-discontinuation transition probabilities for each cycle
for (t in 1:n.cycle) {
  # Waning factor
  waning_factor <- (1 - tx_waning)^(t - 1)
  
  # --- MCI on-treatment ---
  rate_mci_mil <- -log(1 - p.mci_mil) * rr.tx_mci_mil^waning_factor
  v.p.mci_mil_int[t] <- 1 - exp(-rate_mci_mil)
  v.p.mci_mod_int[t] <- 0  # original probability = 0
  v.p.mci_sev_int[t] <- 0  # original probability = 0
  v.p.mci_mci_int[t] <- 1 - v.p.mci_mil_int[t]
  
  # --- MCI post-discontinuation ---
  rate_mci_mil_dis <- -log(1 - p.mci_mil) * rr.tx_mci_mil_dis^((1 - tx_waning_dis)^(t - 1))
  v.p.mci_mil_int_dis[t] <- 1 - exp(-rate_mci_mil_dis)
  v.p.mci_mod_int_dis[t] <- 0
  v.p.mci_sev_int_dis[t] <- 0
  v.p.mci_mci_int_dis[t] <- 1 - v.p.mci_mil_int_dis[t]
  
  # --- Mild on-treatment ---
  rate_mil_mod <- -log(1 - p.mil_mod) * rr.tx_mil_mod^waning_factor
  rate_mil_sev <- -log(1 - p.mil_sev) * rr.tx_mil_sev^waning_factor
  v.p.mil_mod_int[t] <- 1 - exp(-rate_mil_mod)
  v.p.mil_sev_int[t] <- 1 - exp(-rate_mil_sev)
  v.p.mil_mil_int[t] <- 1 - v.p.mil_mci_int[t] - v.p.mil_mod_int[t] - v.p.mil_sev_int[t]
  
  # --- Mild post-discontinuation ---
  rate_mil_mod_dis <- -log(1 - p.mil_mod) * rr.tx_mil_mod_dis^((1 - tx_waning_dis)^(t - 1))
  rate_mil_sev_dis <- -log(1 - p.mil_sev) * rr.tx_mil_sev_dis^((1 - tx_waning_dis)^(t - 1))
  v.p.mil_mod_int_dis[t] <- 1 - exp(-rate_mil_mod_dis)
  v.p.mil_sev_int_dis[t] <- 1 - exp(-rate_mil_sev_dis)
  v.p.mil_mil_int_dis[t] <- 1 - v.p.mil_mci_int_dis[t] - v.p.mil_mod_int_dis[t] - v.p.mil_sev_int_dis[t]
}
# Function to build transition probability matrix array
build_transition_matrix <- function(strat) {
  
  # Initialize array: [from_state, to_state, cycle]
  a.TP <- array(0, dim = c(length(v.names_state), length(v.names_state), n.cycle),
                dimnames = list(v.names_state, v.names_state, NULL))
  
  # Loop over each cycle
  for (t in 1:n.cycle) {
    
    # Mortality rate for current cycle
    mort_t <- v.mort_rate[t]
    
    # Transition to death from each state (explicitly listed)
    a.TP["mcion_c", "dth", t] <- 1 - exp(-mort_t * hr.mort_mci)
    a.TP["mciof_c", "dth", t] <- 1 - exp(-mort_t * hr.mort_mci)
    a.TP["milon_c", "dth", t] <- 1 - exp(-mort_t * hr.mort_mil)
    a.TP["milof_c", "dth", t] <- 1 - exp(-mort_t * hr.mort_mil)
    a.TP["mod_c", "dth", t]    <- 1 - exp(-mort_t * hr.mort_mod)
    a.TP["sev_c", "dth", t]    <- 1 - exp(-mort_t * hr.mort_sev)
    a.TP["mci_i", "dth", t]    <- 1 - exp(-mort_t * hr.mort_mci)
    a.TP["mil_i", "dth", t]    <- 1 - exp(-mort_t * hr.mort_mil)
    a.TP["mod_i", "dth", t]    <- 1 - exp(-mort_t * hr.mort_mod)
    a.TP["sev_i", "dth", t]    <- 1 - exp(-mort_t * hr.mort_sev)
    a.TP["dth",    "dth", t]   <- 1  # Death is an absorbing state
    
    # === Placeholders for strategy-specific transition assignments ===
    
    # === [INSERT PROMPT 5B HERE] — transitions from on-treatment community states ===
    
    if (strat == "soc") {
      # Use *_soc vectors for standard of care
      # --- From mcion_c ---
      a.TP["mcion_c", "mcion_c", t] <- v.p.mci_mci_soc[t] * (1 - a.TP["mcion_c", "dth", t]) * (1 - v.p.mci_i_soc[t]) * (1 - v.p.discontinuation_soc[t])
      a.TP["mcion_c", "mciof_c", t] <- v.p.mci_mci_soc[t] * (1 - a.TP["mcion_c", "dth", t]) * (1 - v.p.mci_i_soc[t]) * v.p.discontinuation_soc[t]
      a.TP["mcion_c", "milon_c", t] <- v.p.mci_mil_soc[t] * (1 - a.TP["mcion_c", "dth", t]) * (1 - v.p.mci_i_soc[t]) * (1 - v.p.discontinuation_soc[t])
      a.TP["mcion_c", "mci_i",   t] <- v.p.mci_mil_soc[t] * (1 - a.TP["mcion_c", "dth", t]) * v.p.mci_i_soc[t]
      
      # --- From milon_c ---
      a.TP["milon_c", "milon_c", t] <- v.p.mil_mil_soc[t] * (1 - a.TP["milon_c", "dth", t]) * (1 - v.p.mil_i_soc[t]) * (1 - v.p.discontinuation_soc[t])
      a.TP["milon_c", "milof_c", t] <- v.p.mil_mil_soc[t] * (1 - a.TP["milon_c", "dth", t]) * (1 - v.p.mil_i_soc[t]) * v.p.discontinuation_soc[t]
      a.TP["milon_c", "milof_c", t] <- a.TP["milon_c", "milof_c", t] + v.p.mil_mod_soc[t] * (1 - a.TP["milon_c", "dth", t]) * (1 - v.p.mil_i_soc[t])
      a.TP["milon_c", "mil_i",   t] <- v.p.mil_mod_soc[t] * (1 - a.TP["milon_c", "dth", t]) * v.p.mil_i_soc[t]
      
    } else if (strat == "int") {
      # Use *_int and *_int_dis vectors for intervention
      
      # --- From mcion_c ---
      a.TP["mcion_c", "mcion_c", t] <- v.p.mci_mci_int[t] * (1 - a.TP["mcion_c", "dth", t]) * (1 - v.p.mci_i_soc[t]) * (1 - v.p.discontinuation_soc[t])
      a.TP["mcion_c", "mciof_c", t] <- v.p.mci_mci_int[t] * (1 - a.TP["mcion_c", "dth", t]) * (1 - v.p.mci_i_soc[t]) * v.p.discontinuation_soc[t]
      a.TP["mcion_c", "milon_c", t] <- v.p.mci_mil_int[t] * (1 - a.TP["mcion_c", "dth", t]) * (1 - v.p.mci_i_soc[t]) * (1 - v.p.discontinuation_soc[t])
      a.TP["mcion_c", "mci_i",   t] <- v.p.mci_mil_int[t] * (1 - a.TP["mcion_c", "dth", t]) * v.p.mci_i_soc[t]
      
      # --- From milon_c ---
      a.TP["milon_c", "milon_c", t] <- v.p.mil_mil_int[t] * (1 - a.TP["milon_c", "dth", t]) * (1 - v.p.mil_i_soc[t]) * (1 - v.p.discontinuation_soc[t])
      a.TP["milon_c", "milof_c", t] <- v.p.mil_mil_int[t] * (1 - a.TP["milon_c", "dth", t]) * (1 - v.p.mil_i_soc[t]) * v.p.discontinuation_soc[t]
      a.TP["milon_c", "milof_c", t] <- a.TP["milon_c", "milof_c", t] + v.p.mil_mod_int[t] * (1 - a.TP["milon_c", "dth", t]) * (1 - v.p.mil_i_soc[t])
      a.TP["milon_c", "mil_i",   t] <- v.p.mil_mod_int[t] * (1 - a.TP["milon_c", "dth", t]) * v.p.mil_i_soc[t]
    }
    
    # === [INSERT PROMPT 5C HERE] — transitions from off-treatment, moderate/severe community states ===
    
    if (strat == "soc") {
      # --- From mciof_c ---
      a.TP["mciof_c", "mciof_c", t] <- v.p.mci_mci_soc[t] * (1 - a.TP["mciof_c", "dth", t]) * (1 - v.p.mci_i_soc[t])
      a.TP["mciof_c", "milof_c", t] <- v.p.mci_mil_soc[t] * (1 - a.TP["mciof_c", "dth", t]) * (1 - v.p.mci_i_soc[t])
      a.TP["mciof_c", "mci_i",    t] <- v.p.mci_mil_soc[t] * (1 - a.TP["mciof_c", "dth", t]) * v.p.mci_i_soc[t]
      
      # --- From milof_c ---
      a.TP["milof_c", "milof_c", t] <- v.p.mil_mil_soc[t] * (1 - a.TP["milof_c", "dth", t]) * (1 - v.p.mil_i_soc[t])
      a.TP["milof_c", "mod_c",   t] <- v.p.mil_mod_soc[t] * (1 - a.TP["milof_c", "dth", t]) * (1 - v.p.mil_i_soc[t])
      a.TP["milof_c", "mil_i",   t] <- v.p.mil_mod_soc[t] * (1 - a.TP["milof_c", "dth", t]) * v.p.mil_i_soc[t]
      
      # --- From mod_c ---
      a.TP["mod_c", "mod_c", t] <- v.p.mod_mod_soc[t] * (1 - a.TP["mod_c", "dth", t]) * (1 - v.p.mod_i_soc[t])
      a.TP["mod_c", "sev_c", t] <- v.p.mod_sev_soc[t] * (1 - a.TP["mod_c", "dth", t]) * (1 - v.p.mod_i_soc[t])
      a.TP["mod_c", "mod_i", t] <- v.p.mod_sev_soc[t] * (1 - a.TP["mod_c", "dth", t]) * v.p.mod_i_soc[t]
      
      # --- From sev_c ---
      a.TP["sev_c", "sev_c", t] <- v.p.sev_sev_soc[t] * (1 - a.TP["sev_c", "dth", t]) * (1 - v.p.sev_i_soc[t])
      a.TP["sev_c", "mod_c", t] <- v.p.sev_mod_soc[t] * (1 - a.TP["sev_c", "dth", t]) * (1 - v.p.sev_i_soc[t])
      a.TP["sev_c", "sev_i", t] <- v.p.sev_mod_soc[t] * (1 - a.TP["sev_c", "dth", t]) * v.p.sev_i_soc[t]
      
    } else if (strat == "int") {
      # --- From mciof_c ---
      a.TP["mciof_c", "mciof_c", t] <- v.p.mci_mci_int_dis[t] * (1 - a.TP["mciof_c", "dth", t]) * (1 - v.p.mci_i_soc[t])
      a.TP["mciof_c", "milof_c", t] <- v.p.mci_mil_int_dis[t] * (1 - a.TP["mciof_c", "dth", t]) * (1 - v.p.mci_i_soc[t])
      a.TP["mciof_c", "mci_i",    t] <- v.p.mci_mil_int_dis[t] * (1 - a.TP["mciof_c", "dth", t]) * v.p.mci_i_soc[t]
      
      # --- From milof_c ---
      a.TP["milof_c", "milof_c", t] <- v.p.mil_mil_int_dis[t] * (1 - a.TP["milof_c", "dth", t]) * (1 - v.p.mil_i_soc[t])
      a.TP["milof_c", "mod_c",   t] <- v.p.mil_mod_int_dis[t] * (1 - a.TP["milof_c", "dth", t]) * (1 - v.p.mil_i_soc[t])
      a.TP["milof_c", "mil_i",   t] <- v.p.mil_mod_int_dis[t] * (1 - a.TP["milof_c", "dth", t]) * v.p.mil_i_soc[t]
      
      # --- From mod_c ---
      a.TP["mod_c", "mod_c", t] <- v.p.mod_mod_soc[t] * (1 - a.TP["mod_c", "dth", t]) * (1 - v.p.mod_i_soc[t])
      a.TP["mod_c", "sev_c", t] <- v.p.mod_sev_soc[t] * (1 - a.TP["mod_c", "dth", t]) * (1 - v.p.mod_i_soc[t])
      a.TP["mod_c", "mod_i", t] <- v.p.mod_sev_soc[t] * (1 - a.TP["mod_c", "dth", t]) * v.p.mod_i_soc[t]
      
      # --- From sev_c ---
      a.TP["sev_c", "sev_c", t] <- v.p.sev_sev_soc[t] * (1 - a.TP["sev_c", "dth", t]) * (1 - v.p.sev_i_soc[t])
      a.TP["sev_c", "mod_c", t] <- v.p.sev_mod_soc[t] * (1 - a.TP["sev_c", "dth", t]) * (1 - v.p.sev_i_soc[t])
      a.TP["sev_c", "sev_i", t] <- v.p.sev_mod_soc[t] * (1 - a.TP["sev_c", "dth", t]) * v.p.sev_i_soc[t]
    }
    
    # === [INSERT PROMPT 5D HERE] — transitions from institutionalized states ===
    
    # Transitions from mci_i
    a.TP["mci_i", "mci_i", t] <- v.p.mci_mci_soc[t] * (1 - a.TP["mci_i", "dth", t])
    a.TP["mci_i", "mil_i", t] <- v.p.mci_mil_soc[t] * (1 - a.TP["mci_i", "dth", t])
    a.TP["mci_i", "mod_i", t] <- v.p.mci_mod_soc[t] * (1 - a.TP["mci_i", "dth", t])
    a.TP["mci_i", "sev_i", t] <- v.p.mci_sev_soc[t] * (1 - a.TP["mci_i", "dth", t])
    
    # Transitions from mil_i
    a.TP["mil_i", "mil_i", t] <- v.p.mil_mil_soc[t] * (1 - a.TP["mil_i", "dth", t])
    a.TP["mil_i", "mod_i", t] <- v.p.mil_mod_soc[t] * (1 - a.TP["mil_i", "dth", t])
    a.TP["mil_i", "sev_i", t] <- v.p.mil_sev_soc[t] * (1 - a.TP["mil_i", "dth", t])
    
    # Transitions from mod_i
    a.TP["mod_i", "mod_i", t] <- v.p.mod_mod_soc[t] * (1 - a.TP["mod_i", "dth", t])
    a.TP["mod_i", "sev_i", t] <- v.p.mod_sev_soc[t] * (1 - a.TP["mod_i", "dth", t])
    
    # Transitions from sev_i
    a.TP["sev_i", "sev_i", t] <- v.p.sev_sev_soc[t] * (1 - a.TP["sev_i", "dth", t])
    a.TP["sev_i", "mod_i", t] <- v.p.sev_mod_soc[t] * (1 - a.TP["sev_i", "dth", t])
    
    # === Normalize rows to ensure each row sums to 1 ===
    for (s in v.names_state) {
      row_sum <- sum(a.TP[s, , t])
      if (row_sum > 0 && abs(row_sum - 1) > 1e-8) {
        a.TP[s, , t] <- a.TP[s, , t] / row_sum
      }
    }
  }
  
  return(a.TP)
}
# Function to run Markov trace and compute total QALYs, costs, and LYs
run_markov <- function(strat) {
  # Build transition matrix
  a.TP <- build_transition_matrix(strat)
  
  # Initialize trace matrix
  m.M <- matrix(0, nrow = n.cycle, ncol = length(v.names_state),
                dimnames = list(1:n.cycle, v.names_state))
  
  # Set initial state distribution
  m.M[1, ] <- 0
  m.M[1, "mcion_c"] <- ifelse(strat == "int", p.starting_state_mci, 0)
  m.M[1, "mciof_c"] <- ifelse(strat == "soc", p.starting_state_mci, 0)
  m.M[1, "milon_c"] <- ifelse(strat == "int", 1 - p.starting_state_mci, 0)
  m.M[1, "milof_c"] <- ifelse(strat == "soc", 1 - p.starting_state_mci, 0)
  
  # Initialize vectors for utilities and costs (named for safe indexing)
  v.u.pt <- c(mcion_c = u.mci_pt, mciof_c = u.mci_pt, milon_c = u.mil_pt, milof_c = u.mil_pt,
              mod_c = u.mod_pt, sev_c = u.sev_pt, mci_i = u.mci_pt_i, mil_i = u.mil_pt_i,
              mod_i = u.mod_pt_i, sev_i = u.sev_pt_i, dth = 0)
  
  v.u.ic <- c(mcion_c = u.mci_ic, mciof_c = u.mci_ic, milon_c = u.mil_ic, milof_c = u.mil_ic,
              mod_c = u.mod_ic, sev_c = u.sev_ic, mci_i = u.mci_ic_i, mil_i = u.mil_ic_i,
              mod_i = u.mod_ic_i, sev_i = u.sev_ic_i, dth = 0)
  
  v.c <- c(mcion_c = c.mci_hc + c.mci_sc + c.mci_ic,
           mciof_c = c.mci_hc + c.mci_sc + c.mci_ic,
           milon_c = c.mil_hc + c.mil_sc + c.mil_ic,
           milof_c = c.mil_hc + c.mil_sc + c.mil_ic,
           mod_c = c.mod_hc + c.mod_sc + c.mod_ic,
           sev_c = c.sev_hc + c.sev_sc + c.sev_ic,
           mci_i = c.mci_i_hc + c.mci_i_sc + c.mci_i_ic,
           mil_i = c.mil_i_hc + c.mil_i_sc + c.mil_i_ic,
           mod_i = c.mod_i_hc + c.mod_i_sc + c.mod_i_ic,
           sev_i = c.sev_i_hc + c.sev_i_sc + c.sev_i_ic,
           dth = 0)
  
  # Initialize output matrices
  v.QALY <- numeric(n.cycle)
  v.Cost <- numeric(n.cycle)
  v.LY <- numeric(n.cycle)
  
  # Markov trace loop
  for (t in 1:(n.cycle - 1)) {
    m.M[t + 1, ] <- m.M[t, ] %*% a.TP[ , , t]
  }
  
  # Compute undiscounted outcomes
  for (t in 1:n.cycle) {
    state_occupancy <- m.M[t, ]
    v.QALY[t] <- sum(state_occupancy * (v.u.pt + v.u.ic))
    v.Cost[t] <- sum(state_occupancy * v.c)
    v.LY[t] <- sum(state_occupancy[setdiff(names(state_occupancy), "dth")])
    
    # Apply recurring treatment cost to on-treatment states
    if (strat == "int") {
      v.Cost[t] <- v.Cost[t] + c.Tx * sum(state_occupancy[c("mcion_c", "milon_c")])
    }
    
    # First cycle only: apply initial cost and disutility from treatment initiation
    if (t == 1 && strat == "int") {
      v.QALY[t] <- v.QALY[t] + u.Tx_start
      v.Cost[t] <- v.Cost[t] + c.Tx_start
    }
  }
  
  # Half-cycle correction (excluding final cycle)
  v.QALY <- 0.5 * (v.QALY[-1] + v.QALY[-n.cycle])
  v.Cost <- 0.5 * (v.Cost[-1] + v.Cost[-n.cycle])
  v.LY <- 0.5 * (v.LY[-1] + v.LY[-n.cycle])
  
  # Apply discounting
  discount_vector <- 1 / ((1 + discount_QALY) ^ (1:(n.cycle - 1)))
  total.QALY <- sum(v.QALY * discount_vector)
  total.LY <- sum(v.LY * discount_vector)
  total.Cost <- sum(v.Cost * (1 / ((1 + discount_COST) ^ (1:(n.cycle - 1)))))
  
  return(list(QALY = total.QALY, LY = total.LY, Cost = total.Cost))
}

# Run model for both strategies
res.soc <- run_markov("soc")
res.int <- run_markov("int")

# Print results comparison
cat("===== IPECAD Model Results Comparison =====\n")
cat(sprintf("Standard of Care:\n  QALYs: %.2f\n  LYs: %.2f\n  Costs: $%.2f\n\n",
            res.soc$QALY, res.soc$LY, res.soc$Cost))
cat(sprintf("Intervention:\n  QALYs: %.2f\n  LYs: %.2f\n  Costs: $%.2f\n\n",
            res.int$QALY, res.int$LY, res.int$Cost))
cat(sprintf("Incremental (Int - Soc):\n  ΔQALYs: %.2f\n  ΔLYs: %.2f\n  ΔCosts: $%.2f\n",
            res.int$QALY - res.soc$QALY,
            res.int$LY - res.soc$LY,
            res.int$Cost - res.soc$Cost))
```
