## Supplementary material for "Replicating Health-Economic Simulation Models for Alzheimer’s Disease Using Artificial Intelligence": final R scripts: o14.R

```
# ----------------------------
# IPECAD Markov Model Inputs
# ----------------------------

# Load required package
library(dampack)

# Fixed state and strategy names
v.names_state <- c("mcion_c", "mciof_c", "milon_c", "milof_c",
                   "mod_c", "sev_c", "mci_i", "mil_i", "mod_i", "sev_i", "dth")
v.names_strat <- c("soc", "int")

# Starting population characteristics
age_start <- 71
sex <- "weighted"  # 52% female
p.starting_state_mci <- 0.55

# Time horizon
n.cycle <- 29

# Transition probabilities (per cycle)
p.mci_mil <- 0.23
p.mci_mod <- 0
p.mci_sev <- 0
p.mil_mci <- 0.03
p.mil_mod <- 0.35
p.mil_sev <- 0.04
p.mod_mil <- 0.03
p.mod_sev <- 0.42
p.sev_mil <- 0
p.sev_mod <- 0.02

# Transition to institutional care
p.mci_i <- 0.024
p.mil_i <- 0.038
p.mod_i <- 0.110
p.sev_i <- 0.259

# Mortality hazard ratios
hr.mort_mci <- 1.82
hr.mort_mil <- 2.92
hr.mort_mod <- 3.85
hr.mort_sev <- 9.52

# Treatment effects (on-treatment relative risks)
rr.tx_mci_mil <- 0.69
rr.tx_mci_mod <- 1
rr.tx_mci_sev <- 1
rr.tx_mil_mod <- 0.69
rr.tx_mil_sev <- 0.69

# Treatment effects (post-discontinuation relative risks)
rr.tx_mci_mil_dis <- 1
rr.tx_mci_mod_dis <- 1
rr.tx_mci_sev_dis <- 1
rr.tx_mil_mod_dis <- 1
rr.tx_mil_sev_dis <- 1

# Discontinuation parameters
p.discontinuation1 <- 0.069
p.discontinuation2 <- 0
discontinuation2_begin <- 2
tx_duration <- 29

# Treatment waning parameters
tx_waning <- 0
tx_waning_dis <- 0

# Utility values - patient (community)
u.mci_pt <- 0.851 - 0.17
u.mil_pt <- 0.851 - 0.22
u.mod_pt <- 0.851 - 0.36
u.sev_pt <- 0.851 - 0.53

# Utility values - patient (institution)
u.mci_pt_i <- 0.851 - 0.17
u.mil_pt_i <- 0.851 - 0.19
u.mod_pt_i <- 0.851 - 0.42
u.sev_pt_i <- 0.851 - 0.59

# Utility values - informal caregiver (community)
u.mci_ic <- -0.03
u.mil_ic <- -0.05
u.mod_ic <- -0.08
u.sev_ic <- -0.10

# Utility values - informal caregiver (institution)
u.mci_ic_i <- -0.03
u.mil_ic_i <- -0.05
u.mod_ic_i <- -0.08
u.sev_ic_i <- -0.10

# One-time disutility in first cycle (e.g., ARIA)
u.Tx_start <- -0.14 * (12 / 52) * 0.035

# Cost calculations (community health care)
c.mci_hc <- 6042 * 1.12 + 460
c.mil_hc <- 6042 * 1.56 + 965 + 0.21 * 365 * 0.333
c.mod_hc <- 6042 * 1.93 + 1544 + 0.66 * 365 * 0.333
c.sev_hc <- 6042 * 1.93 + 1930

# Cost calculations (institutional health care)
c.mci_i_hc <- c.mci_hc
c.mil_i_hc <- c.mil_hc
c.mod_i_hc <- c.mod_hc
c.sev_i_hc <- c.sev_hc

# Cost calculations (community social care)
c.mci_sc <- 0
c.mil_sc <- 0
c.mod_sc <- 0
c.sev_sc <- 0

# Cost calculations (institution social care)
c.mci_i_sc <- 7394 * 12
c.mil_i_sc <- 7394 * 12
c.mod_i_sc <- 7394 * 12
c.sev_i_sc <- 7394 * 12

# Informal care (community)
c.mci_ic <- 69 * 12 * 32.46 + 0.204 * 0.049 * 20 * 52 * 32.46
c.mil_ic <- 113 * 12 * 32.46 + 0.112 * 0.086 * 20 * 52 * 32.46
c.mod_ic <- 169 * 12 * 32.46
c.sev_ic <- 298 * 12 * 32.46

# Informal care (institution) - 44% of community values + productivity
c.mci_i_ic <- 69 * 12 * 32.46 * 0.44 + 0.204 * 0.049 * 20 * 52 * 32.46
c.mil_i_ic <- 113 * 12 * 32.46 * 0.44 + 0.112 * 0.086 * 20 * 52 * 32.46
c.mod_i_ic <- 169 * 12 * 32.46 * 0.44
c.sev_i_ic <- 298 * 12 * 32.46 * 0.44

# Treatment cost per year
c.Tx <- 26500 + (52 / 2) * 78.35

# First cycle additional costs (e.g., MRI scans)
c.Tx_start <- 261.10 * 4 + 261.10 * 3 * 0.215

# Discount rates
discount_EFFECT <- 0.03
discount_QALY <- 0.03
discount_COST <- 0.03

# WTP threshold
wtp <- 100000

# Half-cycle correction flag
half_cycle_correction <- TRUE
# ----------------------------
# Load and Process 2019 US Life Table
# ----------------------------

# Load the life table CSV
lifetable <- read.csv("lifetable_US_2019_ssa.csv", stringsAsFactors = FALSE)

# Convert annual mortality probabilities (q) to rates (r) using: r = -log(1 - q)
lifetable$male <- -log(1 - lifetable$male)
lifetable$female <- -log(1 - lifetable$female)

# Define population sex weights (52% female, 48% male)
weight_female <- 0.52
weight_male <- 0.48

# Compute weighted average of mortality rates
lifetable$weighted <- lifetable$male * weight_male + lifetable$female * weight_female

# Resulting data frame
lifetable_processed <- lifetable[, c("age", "male", "female", "weighted")]

# Print first few rows (optional)
# head(lifetable_processed)

# Return the processed life table
lifetable_processed
# ----------------------------
# Time-dependent Transition Vectors for SOC Strategy
# ----------------------------

# Set time horizon
cycles <- n.cycle

# ----------------------------
# Disease progression probabilities (SOC strategy, constant over time)
# ----------------------------
v.p.mci_mil_soc  <- rep(p.mci_mil,  cycles)
v.p.mci_mod_soc  <- rep(p.mci_mod,  cycles)
v.p.mci_sev_soc  <- rep(p.mci_sev,  cycles)
v.p.mil_mci_soc  <- rep(p.mil_mci,  cycles)
v.p.mil_mod_soc  <- rep(p.mil_mod,  cycles)
v.p.mil_sev_soc  <- rep(p.mil_sev,  cycles)
v.p.mod_mil_soc  <- rep(p.mod_mil,  cycles)
v.p.mod_sev_soc  <- rep(p.mod_sev,  cycles)
v.p.sev_mil_soc  <- rep(p.sev_mil,  cycles)
v.p.sev_mod_soc  <- rep(p.sev_mod,  cycles)

# ----------------------------
# Staying probabilities (excluding mortality and institutionalization)
# ----------------------------
v.p.stay_mci_soc <- 1 - (v.p.mci_mil_soc + v.p.mci_mod_soc + v.p.mci_sev_soc)
v.p.stay_mil_soc <- 1 - (v.p.mil_mci_soc + v.p.mil_mod_soc + v.p.mil_sev_soc)
v.p.stay_mod_soc <- 1 - (v.p.mod_mil_soc + v.p.mod_sev_soc)
v.p.stay_sev_soc <- 1 - (v.p.sev_mil_soc + v.p.sev_mod_soc)

# ----------------------------
# Transition to institutional care (time-independent)
# ----------------------------
v.p.mci_i <- rep(p.mci_i, cycles)
v.p.mil_i <- rep(p.mil_i, cycles)
v.p.mod_i <- rep(p.mod_i, cycles)
v.p.sev_i <- rep(p.sev_i, cycles)

# ----------------------------
# Treatment discontinuation vector (SOC strategy is off-treatment, but structure included for completeness)
# For general use (e.g., applied to INT strategy where needed)
# ----------------------------
v.p.discontinuation <- numeric(cycles)
v.p.discontinuation[1] <- p.discontinuation1
v.p.discontinuation[2:(tx_duration)] <- p.discontinuation2
if (tx_duration < cycles) {
  v.p.discontinuation[(tx_duration + 1):cycles] <- 1
}

# ----------------------------
# Mortality rates per cycle based on age and sex (weighted)
# ----------------------------
# Ensure life table data has been processed and named 'lifetable_processed'
v.r.mortality <- lifetable_processed$weighted[age_start + 0:(cycles - 1)]

# Result vectors are now defined for disease progression, staying, institutionalization,
# treatment discontinuation, and background mortality.
# ----------------------------
# Time-dependent Transition Vectors for INT Strategy
# ----------------------------

# Create index vector for cycles
cycle_index <- 1:n.cycle

# ----------------------------
# MCI on-treatment (mcion_c) → mil/mod/sev
# ----------------------------
r.p.mci_mil_int_on <- -log(1 - p.mci_mil) * rr.tx_mci_mil^( (1 - tx_waning)^(cycle_index - 1) )
v.p.mci_mil_int_on <- 1 - exp(-r.p.mci_mil_int_on)

r.p.mci_mod_int_on <- -log(1 - p.mci_mod) * rr.tx_mci_mod^( (1 - tx_waning)^(cycle_index - 1) )
v.p.mci_mod_int_on <- 1 - exp(-r.p.mci_mod_int_on)

r.p.mci_sev_int_on <- -log(1 - p.mci_sev) * rr.tx_mci_sev^( (1 - tx_waning)^(cycle_index - 1) )
v.p.mci_sev_int_on <- 1 - exp(-r.p.mci_sev_int_on)

v.p.stay_mcion_c <- 1 - (v.p.mci_mil_int_on + v.p.mci_mod_int_on + v.p.mci_sev_int_on)

# ----------------------------
# Mild on-treatment (milon_c) → mci/mod/sev
# ----------------------------
# Mild → MCI (backward): no treatment effect
v.p.mil_mci_int_on <- rep(p.mil_mci, n.cycle)

r.p.mil_mod_int_on <- -log(1 - p.mil_mod) * rr.tx_mil_mod^( (1 - tx_waning)^(cycle_index - 1) )
v.p.mil_mod_int_on <- 1 - exp(-r.p.mil_mod_int_on)

r.p.mil_sev_int_on <- -log(1 - p.mil_sev) * rr.tx_mil_sev^( (1 - tx_waning)^(cycle_index - 1) )
v.p.mil_sev_int_on <- 1 - exp(-r.p.mil_sev_int_on)

v.p.stay_milon_c <- 1 - (v.p.mil_mci_int_on + v.p.mil_mod_int_on + v.p.mil_sev_int_on)

# ----------------------------
# MCI off-treatment (mciof_c) → mil/mod/sev
# ----------------------------
r.p.mci_mil_int_off <- -log(1 - p.mci_mil) * rr.tx_mci_mil_dis^( (1 - tx_waning_dis)^(cycle_index - 1) )
v.p.mci_mil_int_off <- 1 - exp(-r.p.mci_mil_int_off)

r.p.mci_mod_int_off <- -log(1 - p.mci_mod) * rr.tx_mci_mod_dis^( (1 - tx_waning_dis)^(cycle_index - 1) )
v.p.mci_mod_int_off <- 1 - exp(-r.p.mci_mod_int_off)

r.p.mci_sev_int_off <- -log(1 - p.mci_sev) * rr.tx_mci_sev_dis^( (1 - tx_waning_dis)^(cycle_index - 1) )
v.p.mci_sev_int_off <- 1 - exp(-r.p.mci_sev_int_off)

v.p.stay_mciof_c <- 1 - (v.p.mci_mil_int_off + v.p.mci_mod_int_off + v.p.mci_sev_int_off)

# ----------------------------
# Mild off-treatment (milof_c) → mci/mod/sev
# ----------------------------
# Mild → MCI (backward): no treatment effect
v.p.mil_mci_int_off <- rep(p.mil_mci, n.cycle)

r.p.mil_mod_int_off <- -log(1 - p.mil_mod) * rr.tx_mil_mod_dis^( (1 - tx_waning_dis)^(cycle_index - 1) )
v.p.mil_mod_int_off <- 1 - exp(-r.p.mil_mod_int_off)

r.p.mil_sev_int_off <- -log(1 - p.mil_sev) * rr.tx_mil_sev_dis^( (1 - tx_waning_dis)^(cycle_index - 1) )
v.p.mil_sev_int_off <- 1 - exp(-r.p.mil_sev_int_off)

v.p.stay_milof_c <- 1 - (v.p.mil_mci_int_off + v.p.mil_mod_int_off + v.p.mil_sev_int_off)


# ----------------------------
# Transition Matrix Constructor for IPECAD Model
# ----------------------------

build_transition_matrix <- function(strat) {
  # Number of states and cycles
  n.states <- length(v.names_state)
  n.cycles <- n.cycle
  
  # Initialize transition probability array: states × states × cycles
  a.TP <- array(0, dim = c(n.states, n.states, n.cycles),
                dimnames = list(v.names_state, v.names_state, NULL))
  
  for (t in 1:n.cycles) {
    
    # Mortality rates for each state at time t
    r.mort_mci <- v.r.mortality[t] * hr.mort_mci
    r.mort_mil <- v.r.mortality[t] * hr.mort_mil
    r.mort_mod <- v.r.mortality[t] * hr.mort_mod
    r.mort_sev <- v.r.mortality[t] * hr.mort_sev
    
    p.death_mci <- 1 - exp(-r.mort_mci)
    p.death_mil <- 1 - exp(-r.mort_mil)
    p.death_mod <- 1 - exp(-r.mort_mod)
    p.death_sev <- 1 - exp(-r.mort_sev)
    
    # ----------------------------
    # Explicit death transitions per state
    # ----------------------------
    a.TP["mcion_c", "dth", t]   <- p.death_mci
    a.TP["mciof_c", "dth", t]   <- p.death_mci
    a.TP["milon_c", "dth", t]   <- p.death_mil
    a.TP["milof_c", "dth", t]   <- p.death_mil
    a.TP["mod_c", "dth", t]     <- p.death_mod
    a.TP["sev_c", "dth", t]     <- p.death_sev
    a.TP["mci_i", "dth", t]     <- p.death_mci
    a.TP["mil_i", "dth", t]     <- p.death_mil
    a.TP["mod_i", "dth", t]     <- p.death_mod
    a.TP["sev_i", "dth", t]     <- p.death_sev
    a.TP["dth", "dth", t]       <- 1  # Absorbing state
    
    # ----------------------------
    # Transitions from "mcion_c"
    # ----------------------------
    p_survive_mci <- 1 - a.TP["mcion_c", "dth", t]
    
    # To "mcion_c" (stay on-treatment)
    a.TP["mcion_c", "mcion_c", t] <-
      v.p.stay_mcion_c[t] * (1 - v.p.mci_i[t]) * (1 - v.p.discontinuation[t]) * p_survive_mci
    
    # To "milon_c" (progress on-treatment)
    a.TP["mcion_c", "milon_c", t] <-
      v.p.mci_mil_int_on[t] * (1 - v.p.mci_i[t]) * (1 - v.p.discontinuation[t]) * p_survive_mci
    
    # To "mciof_c" (discontinue and stay)
    a.TP["mcion_c", "mciof_c", t] <-
      v.p.stay_mcion_c[t] * (1 - v.p.mci_i[t]) * v.p.discontinuation[t] * p_survive_mci
    
    # To "milof_c" (discontinue and progress)
    a.TP["mcion_c", "milof_c", t] <-
      v.p.mci_mil_int_on[t] * (1 - v.p.mci_i[t]) * v.p.discontinuation[t] * p_survive_mci
    
    # To "mod_c" (progress, no discontinuation applied)
    a.TP["mcion_c", "mod_c", t] <-
      v.p.mci_mod_int_on[t] * (1 - v.p.mci_i[t]) * p_survive_mci
    
    # To "sev_c" (progress, no discontinuation applied)
    a.TP["mcion_c", "sev_c", t] <-
      v.p.mci_sev_int_on[t] * (1 - v.p.mci_i[t]) * p_survive_mci
    
    # To "mci_i" (institution)
    a.TP["mcion_c", "mci_i", t] <-
      (v.p.stay_mcion_c[t] + v.p.mci_mil_int_on[t] + v.p.mci_mod_int_on[t] + v.p.mci_sev_int_on[t]) *
      v.p.mci_i[t] * p_survive_mci
    
    # ----------------------------
    # Transitions from "milon_c"
    # ----------------------------
    p_survive_mil <- 1 - a.TP["milon_c", "dth", t]
    
    # To "milon_c" (stay on-treatment)
    a.TP["milon_c", "milon_c", t] <-
      v.p.stay_milon_c[t] * (1 - v.p.mil_i[t]) * (1 - v.p.discontinuation[t]) * p_survive_mil
    
    # To "mciof_c" (discontinue and regress)
    a.TP["milon_c", "mciof_c", t] <-
      v.p.mil_mci_int_on[t] * (1 - v.p.mil_i[t]) * v.p.discontinuation[t] * p_survive_mil
    
    # To "milof_c" (discontinue and stay)
    a.TP["milon_c", "milof_c", t] <-
      v.p.stay_milon_c[t] * (1 - v.p.mil_i[t]) * v.p.discontinuation[t] * p_survive_mil
    
    # To "mci_i" (institution, from regress)
    a.TP["milon_c", "mci_i", t] <-
      v.p.mil_mci_int_on[t] * v.p.mil_i[t] * p_survive_mil
    
    # To "mil_i" (institution, from stay or progression)
    a.TP["milon_c", "mil_i", t] <-
      (v.p.stay_milon_c[t] + v.p.mil_mod_int_on[t] + v.p.mil_sev_int_on[t]) * v.p.mil_i[t] * p_survive_mil
    
    # To "mod_c" (progress, no discontinuation)
    a.TP["milon_c", "mod_c", t] <-
      v.p.mil_mod_int_on[t] * (1 - v.p.mil_i[t]) * p_survive_mil
    
    # To "sev_c" (progress, no discontinuation)
    a.TP["milon_c", "sev_c", t] <-
      v.p.mil_sev_int_on[t] * (1 - v.p.mil_i[t]) * p_survive_mil
    
    # ----------------------------
    # Transitions from "mciof_c"
    # ----------------------------
    p_survive_mci <- 1 - a.TP["mciof_c", "dth", t]
    
    if (strat == "soc") {
      a.TP["mciof_c", "mciof_c", t] <- v.p.stay_mci_soc[t] * (1 - v.p.mci_i[t]) * p_survive_mci
      a.TP["mciof_c", "milof_c", t] <- v.p.mci_mil_soc[t] * (1 - v.p.mci_i[t]) * p_survive_mci
      a.TP["mciof_c", "mod_c",    t] <- v.p.mci_mod_soc[t] * (1 - v.p.mci_i[t]) * p_survive_mci
      a.TP["mciof_c", "sev_c",    t] <- v.p.mci_sev_soc[t] * (1 - v.p.mci_i[t]) * p_survive_mci
      
      # Institutionalization
      a.TP["mciof_c", "mci_i", t] <- 
        (v.p.stay_mci_soc[t] + v.p.mci_mil_soc[t] + v.p.mci_mod_soc[t] + v.p.mci_sev_soc[t]) * v.p.mci_i[t] * p_survive_mci
    } else if (strat == "int") {
      a.TP["mciof_c", "mciof_c", t] <- v.p.stay_mciof_c[t] * (1 - v.p.mci_i[t]) * p_survive_mci
      a.TP["mciof_c", "milof_c", t] <- v.p.mci_mil_int_off[t] * (1 - v.p.mci_i[t]) * p_survive_mci
      a.TP["mciof_c", "mod_c",    t] <- v.p.mci_mod_int_off[t] * (1 - v.p.mci_i[t]) * p_survive_mci
      a.TP["mciof_c", "sev_c",    t] <- v.p.mci_sev_int_off[t] * (1 - v.p.mci_i[t]) * p_survive_mci
      
      # Institutionalization
      a.TP["mciof_c", "mci_i", t] <- 
        (v.p.stay_mciof_c[t] + v.p.mci_mil_int_off[t] + v.p.mci_mod_int_off[t] + v.p.mci_sev_int_off[t]) * v.p.mci_i[t] * p_survive_mci
    }
    
    # ----------------------------
    # Transitions from "milof_c"
    # ----------------------------
    p_survive_mil <- 1 - a.TP["milof_c", "dth", t]
    
    if (strat == "soc") {
      a.TP["milof_c", "mciof_c", t] <- v.p.mil_mci_soc[t] * (1 - v.p.mil_i[t]) * p_survive_mil
      a.TP["milof_c", "milof_c", t] <- v.p.stay_mil_soc[t] * (1 - v.p.mil_i[t]) * p_survive_mil
      a.TP["milof_c", "mod_c",    t] <- v.p.mil_mod_soc[t] * (1 - v.p.mil_i[t]) * p_survive_mil
      a.TP["milof_c", "sev_c",    t] <- v.p.mil_sev_soc[t] * (1 - v.p.mil_i[t]) * p_survive_mil
      
      a.TP["milof_c", "mci_i", t] <- v.p.mil_mci_soc[t] * v.p.mil_i[t] * p_survive_mil
      a.TP["milof_c", "mil_i", t] <- 
        (v.p.stay_mil_soc[t] + v.p.mil_mod_soc[t] + v.p.mil_sev_soc[t]) * v.p.mil_i[t] * p_survive_mil
    } else if (strat == "int") {
      a.TP["milof_c", "mciof_c", t] <- v.p.mil_mci_int_off[t] * (1 - v.p.mil_i[t]) * p_survive_mil
      a.TP["milof_c", "milof_c", t] <- v.p.stay_milof_c[t] * (1 - v.p.mil_i[t]) * p_survive_mil
      a.TP["milof_c", "mod_c",    t] <- v.p.mil_mod_int_off[t] * (1 - v.p.mil_i[t]) * p_survive_mil
      a.TP["milof_c", "sev_c",    t] <- v.p.mil_sev_int_off[t] * (1 - v.p.mil_i[t]) * p_survive_mil
      
      a.TP["milof_c", "mci_i", t] <- v.p.mil_mci_int_off[t] * v.p.mil_i[t] * p_survive_mil
      a.TP["milof_c", "mil_i", t] <- 
        (v.p.stay_milof_c[t] + v.p.mil_mod_int_off[t] + v.p.mil_sev_int_off[t]) * v.p.mil_i[t] * p_survive_mil
    }
    
    # ----------------------------
    # Transitions from "mod_c"
    # ----------------------------
    p_survive_mod <- 1 - a.TP["mod_c", "dth", t]
    
    if (strat == "soc") {
      a.TP["mod_c", "mod_c", t] <- v.p.stay_mod_soc[t] * (1 - v.p.mod_i[t]) * p_survive_mod
      a.TP["mod_c", "milof_c", t] <- v.p.mod_mil_soc[t] * (1 - v.p.mod_i[t]) * p_survive_mod
      a.TP["mod_c", "sev_c", t] <- v.p.mod_sev_soc[t] * (1 - v.p.mod_i[t]) * p_survive_mod
      
      a.TP["mod_c", "mil_i", t] <- v.p.mod_mil_soc[t] * v.p.mod_i[t] * p_survive_mod
      a.TP["mod_c", "mod_i", t] <- 
        (v.p.stay_mod_soc[t] + v.p.mod_sev_soc[t]) * v.p.mod_i[t] * p_survive_mod
    } else if (strat == "int") {
      a.TP["mod_c", "mod_c", t] <- v.p.stay_mod_soc[t] * (1 - v.p.mod_i[t]) * p_survive_mod
      a.TP["mod_c", "milof_c", t] <- v.p.mod_mil_soc[t] * (1 - v.p.mod_i[t]) * p_survive_mod
      a.TP["mod_c", "sev_c", t] <- v.p.mod_sev_soc[t] * (1 - v.p.mod_i[t]) * p_survive_mod
      
      a.TP["mod_c", "mil_i", t] <- v.p.mod_mil_soc[t] * v.p.mod_i[t] * p_survive_mod
      a.TP["mod_c", "mod_i", t] <- 
        (v.p.stay_mod_soc[t] + v.p.mod_sev_soc[t]) * v.p.mod_i[t] * p_survive_mod
    }
    
    # ----------------------------
    # Transitions from "sev_c"
    # ----------------------------
    p_survive_sev <- 1 - a.TP["sev_c", "dth", t]
    
    if (strat == "soc") {
      a.TP["sev_c", "sev_c", t] <- v.p.stay_sev_soc[t] * (1 - v.p.sev_i[t]) * p_survive_sev
      a.TP["sev_c", "mod_c", t] <- v.p.sev_mod_soc[t] * (1 - v.p.sev_i[t]) * p_survive_sev
      
      a.TP["sev_c", "mod_i", t] <- v.p.sev_mod_soc[t] * v.p.sev_i[t] * p_survive_sev
      a.TP["sev_c", "sev_i", t] <- v.p.stay_sev_soc[t] * v.p.sev_i[t] * p_survive_sev
    } else if (strat == "int") {
      a.TP["sev_c", "sev_c", t] <- v.p.stay_sev_soc[t] * (1 - v.p.sev_i[t]) * p_survive_sev
      a.TP["sev_c", "mod_c", t] <- v.p.sev_mod_soc[t] * (1 - v.p.sev_i[t]) * p_survive_sev
      
      a.TP["sev_c", "mod_i", t] <- v.p.sev_mod_soc[t] * v.p.sev_i[t] * p_survive_sev
      a.TP["sev_c", "sev_i", t] <- v.p.stay_sev_soc[t] * v.p.sev_i[t] * p_survive_sev
    }
    
    # ----------------------------
    # Transitions from "mci_i"
    # ----------------------------
    p_survive_mci <- 1 - a.TP["mci_i", "dth", t]
    
    if (strat == "soc" || strat == "int") {
      a.TP["mci_i", "mci_i", t] <- v.p.stay_mci_soc[t] * p_survive_mci
      a.TP["mci_i", "mil_i", t] <- v.p.mci_mil_soc[t] * p_survive_mci
      a.TP["mci_i", "mod_i", t] <- v.p.mci_mod_soc[t] * p_survive_mci
      a.TP["mci_i", "sev_i", t] <- v.p.mci_sev_soc[t] * p_survive_mci
    }
    
    # ----------------------------
    # Transitions from "mil_i"
    # ----------------------------
    p_survive_mil <- 1 - a.TP["mil_i", "dth", t]
    
    if (strat == "soc" || strat == "int") {
      a.TP["mil_i", "mci_i", t] <- v.p.mil_mci_soc[t] * p_survive_mil
      a.TP["mil_i", "mil_i", t] <- v.p.stay_mil_soc[t] * p_survive_mil
      a.TP["mil_i", "mod_i", t] <- v.p.mil_mod_soc[t] * p_survive_mil
      a.TP["mil_i", "sev_i", t] <- v.p.mil_sev_soc[t] * p_survive_mil
    }
    
    # ----------------------------
    # Transitions from "mod_i"
    # ----------------------------
    p_survive_mod <- 1 - a.TP["mod_i", "dth", t]
    
    if (strat == "soc" || strat == "int") {
      a.TP["mod_i", "mil_i", t] <- v.p.mod_mil_soc[t] * p_survive_mod
      a.TP["mod_i", "mod_i", t] <- v.p.stay_mod_soc[t] * p_survive_mod
      a.TP["mod_i", "sev_i", t] <- v.p.mod_sev_soc[t] * p_survive_mod
    }
    
    # ----------------------------
    # Transitions from "sev_i"
    # ----------------------------
    p_survive_sev <- 1 - a.TP["sev_i", "dth", t]
    
    if (strat == "soc" || strat == "int") {
      a.TP["sev_i", "mil_i", t] <- v.p.sev_mil_soc[t] * p_survive_sev
      a.TP["sev_i", "mod_i", t] <- v.p.sev_mod_soc[t] * p_survive_sev
      a.TP["sev_i", "sev_i", t] <- v.p.stay_sev_soc[t] * p_survive_sev
    }
    
    # ----------------------------
    # Normalize rows so that each state's row sums to 1
    # ----------------------------
    for (s in v.names_state) {
      row_sum <- sum(a.TP[s, , t])
      if (row_sum > 0 && row_sum != 1) {
        a.TP[s, , t] <- a.TP[s, , t] / row_sum
      }
    }
  }
  
  return(a.TP)
}
# ----------------------------
# Markov Trace and Outcome Calculator for IPECAD
# ----------------------------

run_markov_model <- function(strat) {
  # Get transition matrix
  a.TP <- build_transition_matrix(strat)
  
  # Initialize state trace matrix
  m.trace <- matrix(0, nrow = n.cycle + 1, ncol = length(v.names_state),
                    dimnames = list(0:n.cycle, v.names_state))
  
  # Set initial distribution
  m.trace[1, ] <- 0
  if (strat == "soc") {
    m.trace[1, "mciof_c"] <- p.starting_state_mci
    m.trace[1, "milof_c"] <- 1 - p.starting_state_mci
  } else if (strat == "int") {
    m.trace[1, "mcion_c"] <- p.starting_state_mci
    m.trace[1, "milon_c"] <- 1 - p.starting_state_mci
  }
  
  # Named vectors for utilities and costs (per state)
  v.u <- c(
    mcion_c = u.mci_pt,
    mciof_c = u.mci_pt,
    milon_c = u.mil_pt,
    milof_c = u.mil_pt,
    mod_c   = u.mod_pt,
    sev_c   = u.sev_pt,
    mci_i   = u.mci_pt_i,
    mil_i   = u.mil_pt_i,
    mod_i   = u.mod_pt_i,
    sev_i   = u.sev_pt_i,
    dth     = 0
  )
  
  v.c <- c(
    mcion_c = c.mci_hc + c.mci_sc + c.mci_ic,
    mciof_c = c.mci_hc + c.mci_sc + c.mci_ic,
    milon_c = c.mil_hc + c.mil_sc + c.mil_ic,
    milof_c = c.mil_hc + c.mil_sc + c.mil_ic,
    mod_c   = c.mod_hc + c.mod_sc + c.mod_ic,
    sev_c   = c.sev_hc + c.sev_sc + c.sev_ic,
    mci_i   = c.mci_i_hc + c.mci_i_sc + c.mci_i_ic,
    mil_i   = c.mil_i_hc + c.mil_i_sc + c.mil_i_ic,
    mod_i   = c.mod_i_hc + c.mod_i_sc + c.mod_i_ic,
    sev_i   = c.sev_i_hc + c.sev_i_sc + c.sev_i_ic,
    dth     = 0
  )
  
  # Vectors to store outcomes
  v.qalys <- numeric(n.cycle + 1)
  v.costs <- numeric(n.cycle + 1)
  v.lys   <- numeric(n.cycle + 1)
  
  # Evaluate outcomes each cycle
  for (t in 1:n.cycle) {
    # Update trace
    m.trace[t + 1, ] <- m.trace[t, ] %*% a.TP[, , t]
    
    # Apply treatment cost if intervention
    c.adj <- v.c
    if (strat == "int") {
      c.adj["mcion_c"] <- c.adj["mcion_c"] + c.Tx
      c.adj["milon_c"] <- c.adj["milon_c"] + c.Tx
    }
    
    # Utilities and outcomes
    v.qalys[t] <- sum(m.trace[t, ] * v.u)
    v.costs[t] <- sum(m.trace[t, ] * c.adj)
    v.lys[t]   <- sum(m.trace[t, v.names_state != "dth"])
  }
  
  # Last cycle outcomes
  t <- n.cycle + 1
  v.qalys[t] <- sum(m.trace[t, ] * v.u)
  v.costs[t] <- sum(m.trace[t, ] * v.c)
  v.lys[t]   <- sum(m.trace[t, v.names_state != "dth"])
  
  # Add initial treatment cost and QALY adjustment
  if (strat == "int") {
    v.costs[1] <- v.costs[1] + c.Tx_start
    v.qalys[1] <- v.qalys[1] + u.Tx_start
  }
  
  # Apply half-cycle correction
  v.qalys <- (v.qalys[-length(v.qalys)] + v.qalys[-1]) / 2
  v.costs <- (v.costs[-length(v.costs)] + v.costs[-1]) / 2
  v.lys   <- (v.lys[-length(v.lys)] + v.lys[-1]) / 2
  
  # Apply discounting
  v.discount <- 1 / ((1 + discount_QALY) ^ (0:(n.cycle - 1)))
  qalys_disc <- sum(v.qalys * v.discount)
  costs_disc <- sum(v.costs * v.discount)
  lys_disc   <- sum(v.lys   * v.discount)
  
  return(list(qalys = qalys_disc, costs = costs_disc, lys = lys_disc))
}

# ----------------------------
# Run and Compare Strategies
# ----------------------------

res_soc <- run_markov_model("soc")
res_int <- run_markov_model("int")

cat("Comparison of Strategies:\n")
cat("--------------------------------------------------\n")
cat(sprintf("Standard of Care:   QALYs = %.2f, Costs = $%.0f, LYs = %.2f\n", 
            res_soc$qalys, res_soc$costs, res_soc$lys))
cat(sprintf("Intervention:       QALYs = %.2f, Costs = $%.0f, LYs = %.2f\n", 
            res_int$qalys, res_int$costs, res_int$lys))
cat("--------------------------------------------------\n")
cat(sprintf("Incremental:        QALYs = %.2f, Costs = $%.0f, LYs = %.2f\n", 
            res_int$qalys - res_soc$qalys,
            res_int$costs - res_soc$costs,
            res_int$lys   - res_soc$lys))
```
