## Supplementary material for "Replicating Health-Economic Simulation Models for Alzheimer’s Disease Using Artificial Intelligence": final R scripts: o15.R

```
# Load required package
library(dampack)

#=========================#
# Descriptive Parameters  #
#=========================#

# Health states
v.names_state <- c("mcion_c", "mciof_c", "milon_c", "milof_c", "mod_c", 
                   "sev_c", "mci_i", "mil_i", "mod_i", "sev_i", "dth")

# Strategy names
v.names_strat <- c("soc", "int")

#=========================#
# Population Parameters   #
#=========================#

age_start <- 71
sex <- "weighted"  # 52% female
p.starting_state_mci <- 0.55
n.cycle <- 29

#=========================#
# Transition Probabilities#
#=========================#

p.mci_mil <- 0.23
p.mci_mod <- 0
p.mci_sev <- 0

p.mil_mci <- 0.03
p.mil_mod <- 0.35
p.mil_sev <- 0.04

p.mod_mil <- 0.03
p.mod_sev <- 0.42

p.sev_mil <- 0
p.sev_mod <- 0.02

# Transition to institution
p.mci_i <- 0.024
p.mil_i <- 0.038
p.mod_i <- 0.110
p.sev_i <- 0.259

#=========================#
# Mortality Parameters    #
#=========================#

# Mortality hazard ratios
hr.mort_mci <- 1.82
hr.mort_mil <- 2.92
hr.mort_mod <- 3.85
hr.mort_sev <- 9.52

# General mortality table loaded later

#=========================#
# Treatment Effect        #
#=========================#

# On treatment
rr.tx_mci_mil <- 0.69
rr.tx_mci_mod <- 1
rr.tx_mci_sev <- 1
rr.tx_mil_mod <- 0.69
rr.tx_mil_sev <- 0.69

# Off treatment
rr.tx_mci_mil_dis <- 1
rr.tx_mci_mod_dis <- 1
rr.tx_mci_sev_dis <- 1
rr.tx_mil_mod_dis <- 1
rr.tx_mil_sev_dis <- 1

#=========================#
# Discontinuation         #
#=========================#

p.discontinuation1 <- 0.069
p.discontinuation2 <- 0
discontinuation2_begin <- 2
tx_duration <- 29

#=========================#
# Treatment Waning        #
#=========================#

tx_waning <- 0
tx_waning_dis <- 0

#=========================#
# Utility (QALY) Inputs   #
#=========================#

# Utilities – Patient
u.mci_pt <- 0.851 - 0.17
u.mil_pt <- 0.851 - 0.22
u.mod_pt <- 0.851 - 0.36
u.sev_pt <- 0.851 - 0.53

u.mci_pt_i <- 0.851 - 0.17
u.mil_pt_i <- 0.851 - 0.19
u.mod_pt_i <- 0.851 - 0.42
u.sev_pt_i <- 0.851 - 0.59

# Utilities – Informal Caregiver
u.mci_ic <- -0.03
u.mil_ic <- -0.05
u.mod_ic <- -0.08
u.sev_ic <- -0.10

u.mci_ic_i <- -0.03
u.mil_ic_i <- -0.05
u.mod_ic_i <- -0.08
u.sev_ic_i <- -0.10

# One-time disutility for treatment adverse event (ARIA)
u.Tx_start <- -0.14 * (12/52) * 0.035

#=========================#
# Cost Inputs             #
#=========================#

# Health Care Costs – Community
c.mci_hc <- 6042 * 1.12 + 460
c.mil_hc <- 6042 * 1.56 + 965 + 0.21 * 365 * 0.333
c.mod_hc <- 6042 * 1.93 + 1544 + 0.66 * 365 * 0.333
c.sev_hc <- 6042 * 1.93 + 1930

# Health Care Costs – Institution
c.mci_i_hc <- c.mci_hc
c.mil_i_hc <- c.mil_hc
c.mod_i_hc <- c.mod_hc
c.sev_i_hc <- c.sev_hc

# Social Care Costs – Community
c.mci_sc <- 0
c.mil_sc <- 0
c.mod_sc <- 0
c.sev_sc <- 0

# Social Care Costs – Institution
c.mci_i_sc <- 7394 * 12
c.mil_i_sc <- 7394 * 12
c.mod_i_sc <- 7394 * 12
c.sev_i_sc <- 7394 * 12

# Informal Care Costs – Community
c.mci_ic <- 69 * 12 * 32.46 + 0.204 * 0.049 * 20 * 52 * 32.46
c.mil_ic <- 113 * 12 * 32.46 + 0.112 * 0.086 * 20 * 52 * 32.46
c.mod_ic <- 169 * 12 * 32.46
c.sev_ic <- 298 * 12 * 32.46

# Informal Care Costs – Institution
c.mci_i_ic <- 69 * 12 * 32.46 * 0.44 + 0.204 * 0.049 * 20 * 52 * 32.46
c.mil_i_ic <- 113 * 12 * 32.46 * 0.44 + 0.112 * 0.086 * 20 * 52 * 32.46
c.mod_i_ic <- 169 * 12 * 32.46 * 0.44
c.sev_i_ic <- 298 * 12 * 32.46 * 0.44

# Treatment Costs
c.Tx <- 26500 + (52/2) * 78.35
c.Tx_start <- 261.10 * 4 + 261.10 * 3 * 0.215

#=========================#
# Economic Evaluation     #
#=========================#

discount_EFFECT <- 0.03
discount_QALY <- 0.03
discount_COST <- 0.03
wtp <- 100000
half_cycle_correction <- TRUE
# Load life table CSV file
lifetable <- read.csv("lifetable_US_2019_ssa.csv")

# Convert annual mortality probabilities to annual mortality rates
# Formula: rate = -log(1 - probability)
lifetable$male <- -log(1 - lifetable$male)
lifetable$female <- -log(1 - lifetable$female)

# Compute weighted average mortality rate based on population sex distribution (52% female, 48% male)
lifetable$weighted <- 0.48 * lifetable$male + 0.52 * lifetable$female

# Select and return the desired columns
lifetable <- lifetable[, c("age", "male", "female", "weighted")]

# Return the final dataframe
lifetable
#=========================#
# Define number of cycles #
#=========================#

# Already defined in earlier code
# n.cycle <- 29

#=========================#
# Disease progression     #
#=========================#

# Time-dependent transition probability vectors for "soc" strategy (constant values replicated over cycles)
v.p.mci_mil_soc <- rep(p.mci_mil, n.cycle)
v.p.mci_mod_soc <- rep(p.mci_mod, n.cycle)
v.p.mci_sev_soc <- rep(p.mci_sev, n.cycle)

v.p.mil_mci_soc <- rep(p.mil_mci, n.cycle)
v.p.mil_mod_soc <- rep(p.mil_mod, n.cycle)
v.p.mil_sev_soc <- rep(p.mil_sev, n.cycle)

v.p.mod_mil_soc <- rep(p.mod_mil, n.cycle)
v.p.mod_sev_soc <- rep(p.mod_sev, n.cycle)

v.p.sev_mil_soc <- rep(p.sev_mil, n.cycle)
v.p.sev_mod_soc <- rep(p.sev_mod, n.cycle)

#=========================#
# Staying probabilities   #
#=========================#

v.p.mci_stay_soc <- 1 - (v.p.mci_mil_soc + v.p.mci_mod_soc + v.p.mci_sev_soc)
v.p.mil_stay_soc <- 1 - (v.p.mil_mci_soc + v.p.mil_mod_soc + v.p.mil_sev_soc)
v.p.mod_stay_soc <- 1 - (v.p.mod_mil_soc + v.p.mod_sev_soc)
v.p.sev_stay_soc <- 1 - (v.p.sev_mil_soc + v.p.sev_mod_soc)

#=========================#
# Institutionalization    #
#=========================#

v.p.mci_i <- rep(p.mci_i, n.cycle)
v.p.mil_i <- rep(p.mil_i, n.cycle)
v.p.mod_i <- rep(p.mod_i, n.cycle)
v.p.sev_i <- rep(p.sev_i, n.cycle)

#=========================#
# Discontinuation vector  #
#=========================#

v.p.discontinue <- numeric(n.cycle)
v.p.discontinue[1:(discontinuation2_begin - 1)] <- p.discontinuation1
v.p.discontinue[discontinuation2_begin:min(tx_duration, n.cycle)] <- p.discontinuation2
if (tx_duration < n.cycle) {
  v.p.discontinue[(tx_duration + 1):n.cycle] <- 1
}

#=========================#
# Mortality vector        #
#=========================#

# Create mortality vector for starting age (age_start to age_start + n.cycle - 1)
# Assuming 'lifetable' was previously loaded and processed
v.mort_rate <- lifetable$weighted[match(age_start:(age_start + n.cycle - 1), lifetable$age)]


#=============================#
# Helper: Apply treatment effect
#=============================#
apply_rr <- function(p, rr, waning, cycle) {
  rate <- -log(1 - p)
  adj_rate <- rate * rr^((1 - waning)^(cycle - 1))
  1 - exp(-adj_rate)
}

#=============================#
# Create time-dependent vectors
#=============================#
cycles <- 1:n.cycle

# On-treatment transition vectors (mcion_c)
v.p.mci_mil_int_on  <- sapply(cycles, function(c) apply_rr(p.mci_mil, rr.tx_mci_mil, tx_waning, c))
v.p.mci_mod_int_on  <- sapply(cycles, function(c) apply_rr(p.mci_mod, rr.tx_mci_mod, tx_waning, c))
v.p.mci_sev_int_on  <- sapply(cycles, function(c) apply_rr(p.mci_sev, rr.tx_mci_sev, tx_waning, c))

# Staying probability for mcion_c
v.p.mci_stay_int_on <- 1 - (v.p.mci_mil_int_on + v.p.mci_mod_int_on + v.p.mci_sev_int_on)

# On-treatment transition vectors (milon_c)
v.p.mil_mci_int_on  <- rep(p.mil_mci, n.cycle)  # backward transition, no RR
v.p.mil_mod_int_on  <- sapply(cycles, function(c) apply_rr(p.mil_mod, rr.tx_mil_mod, tx_waning, c))
v.p.mil_sev_int_on  <- sapply(cycles, function(c) apply_rr(p.mil_sev, rr.tx_mil_sev, tx_waning, c))

# Staying probability for milon_c
v.p.mil_stay_int_on <- 1 - (v.p.mil_mci_int_on + v.p.mil_mod_int_on + v.p.mil_sev_int_on)

# Off-treatment transition vectors (mciof_c)
v.p.mci_mil_int_off  <- sapply(cycles, function(c) apply_rr(p.mci_mil, rr.tx_mci_mil_dis, tx_waning_dis, c))
v.p.mci_mod_int_off  <- sapply(cycles, function(c) apply_rr(p.mci_mod, rr.tx_mci_mod_dis, tx_waning_dis, c))
v.p.mci_sev_int_off  <- sapply(cycles, function(c) apply_rr(p.mci_sev, rr.tx_mci_sev_dis, tx_waning_dis, c))

# Staying probability for mciof_c
v.p.mci_stay_int_off <- 1 - (v.p.mci_mil_int_off + v.p.mci_mod_int_off + v.p.mci_sev_int_off)

# Off-treatment transition vectors (milof_c)
v.p.mil_mci_int_off  <- rep(p.mil_mci, n.cycle)  # backward transition, no RR
v.p.mil_mod_int_off  <- sapply(cycles, function(c) apply_rr(p.mil_mod, rr.tx_mil_mod_dis, tx_waning_dis, c))
v.p.mil_sev_int_off  <- sapply(cycles, function(c) apply_rr(p.mil_sev, rr.tx_mil_sev_dis, tx_waning_dis, c))

# Staying probability for milof_c
v.p.mil_stay_int_off <- 1 - (v.p.mil_mci_int_off + v.p.mil_mod_int_off + v.p.mil_sev_int_off)


#=============================#
# Transition Matrix Builder   #
#=============================#

build_transition_matrix <- function(strat) {
  
  # Initialize transition probability array: states x states x cycles
  a.TP <- array(0, dim = c(length(v.names_state), length(v.names_state), n.cycle),
                dimnames = list(v.names_state, v.names_state, NULL))
  
  for (t in 1:n.cycle) {
    
    # Mortality rates per state
    rate_mci     <- v.mort_rate[t] * hr.mort_mci
    rate_mil     <- v.mort_rate[t] * hr.mort_mil
    rate_mod     <- v.mort_rate[t] * hr.mort_mod
    rate_sev     <- v.mort_rate[t] * hr.mort_sev
    
    # Convert to probabilities
    p_dth_mci <- 1 - exp(-rate_mci)
    p_dth_mil <- 1 - exp(-rate_mil)
    p_dth_mod <- 1 - exp(-rate_mod)
    p_dth_sev <- 1 - exp(-rate_sev)
    
    # Assign death transition probabilities (explicit for each state)
    a.TP["mcion_c", "dth", t] <- p_dth_mci
    a.TP["mciof_c", "dth", t] <- p_dth_mci
    a.TP["milon_c", "dth", t] <- p_dth_mil
    a.TP["milof_c", "dth", t] <- p_dth_mil
    a.TP["mod_c",   "dth", t] <- p_dth_mod
    a.TP["sev_c",   "dth", t] <- p_dth_sev
    a.TP["mci_i",   "dth", t] <- p_dth_mci
    a.TP["mil_i",   "dth", t] <- p_dth_mil
    a.TP["mod_i",   "dth", t] <- p_dth_mod
    a.TP["sev_i",   "dth", t] <- p_dth_sev
    
    # Death is absorbing
    a.TP["dth", "dth", t] <- 1
    
    #------------------------------------------------------------
    # [INSERT PROMPT 5B HERE] — On-treatment community transitions
    #------------------------------------------------------------
    
    # Transitions from "mcion_c"
    p_mci_mil      <- if (strat == "soc") v.p.mci_mil_soc[t] else v.p.mci_mil_int_on[t]
    p_mci_mod      <- if (strat == "soc") v.p.mci_mod_soc[t] else v.p.mci_mod_int_on[t]
    p_mci_sev      <- if (strat == "soc") v.p.mci_sev_soc[t] else v.p.mci_sev_int_on[t]
    p_mci_stay     <- if (strat == "soc") v.p.mci_stay_soc[t] else v.p.mci_stay_int_on[t]
    p_inst_mci     <- v.p.mci_i[t]
    p_disc         <- v.p.discontinue[t]
    p_surv         <- 1 - a.TP["mcion_c", "dth", t]
    
    a.TP["mcion_c", "mcion_c", t] <- p_mci_stay * p_surv * (1 - p_inst_mci) * (1 - p_disc)
    a.TP["mcion_c", "mciof_c", t] <- p_mci_stay * p_surv * (1 - p_inst_mci) * p_disc
    a.TP["mcion_c", "mci_i",   t] <- p_mci_stay * p_surv * p_inst_mci
    
    a.TP["mcion_c", "milon_c", t] <- p_mci_mil * p_surv * (1 - p_inst_mci) * (1 - p_disc)
    a.TP["mcion_c", "milof_c", t] <- p_mci_mil * p_surv * (1 - p_inst_mci) * p_disc #miliof_c
    a.TP["mcion_c", "mil_i",   t] <- p_mci_mil * p_surv * p_inst_mci
    
    a.TP["mcion_c", "mod_c",   t] <- p_mci_mod * p_surv
    a.TP["mcion_c", "sev_c",   t] <- p_mci_sev * p_surv
    
    # Transitions from "milon_c"
    p_mil_mci      <- if (strat == "soc") v.p.mil_mci_soc[t] else v.p.mil_mci_int_on[t]
    p_mil_mod      <- if (strat == "soc") v.p.mil_mod_soc[t] else v.p.mil_mod_int_on[t]
    p_mil_sev      <- if (strat == "soc") v.p.mil_sev_soc[t] else v.p.mil_sev_int_on[t]
    p_mil_stay     <- if (strat == "soc") v.p.mil_stay_soc[t] else v.p.mil_stay_int_on[t]
    p_inst_mil     <- v.p.mil_i[t]
    
    a.TP["milon_c", "milon_c", t] <- p_mil_stay * p_surv * (1 - p_inst_mil) * (1 - p_disc)
    a.TP["milon_c", "milof_c", t] <- p_mil_stay * p_surv * (1 - p_inst_mil) * p_disc
    a.TP["milon_c", "mil_i",   t] <- p_mil_stay * p_surv * p_inst_mil
    
    a.TP["milon_c", "mcion_c", t] <- p_mil_mci * p_surv * (1 - p_inst_mil) * (1 - p_disc)
    a.TP["milon_c", "mciof_c", t] <- p_mil_mci * p_surv * (1 - p_inst_mil) * p_disc
    a.TP["milon_c", "mci_i",   t] <- p_mil_mci * p_surv * p_inst_mil
    
    a.TP["milon_c", "mod_c",   t] <- p_mil_mod * p_surv
    a.TP["milon_c", "sev_c",   t] <- p_mil_sev * p_surv
    
    #------------------------------------------------------------
    # [INSERT PROMPT 5C HERE] — Off-treatment/moderate/severe community transitions
    #------------------------------------------------------------
    
    # Transitions from "mciof_c"
    p_mci_mil      <- if (strat == "soc") v.p.mci_mil_soc[t] else v.p.mci_mil_int_off[t]
    p_mci_mod      <- if (strat == "soc") v.p.mci_mod_soc[t] else v.p.mci_mod_int_off[t]
    p_mci_sev      <- if (strat == "soc") v.p.mci_sev_soc[t] else v.p.mci_sev_int_off[t]
    p_mci_stay     <- if (strat == "soc") v.p.mci_stay_soc[t] else v.p.mci_stay_int_off[t]
    p_inst_mci     <- v.p.mci_i[t]
    p_surv         <- 1 - a.TP["mciof_c", "dth", t]
    
    a.TP["mciof_c", "mciof_c", t] <- p_mci_stay * p_surv * (1 - p_inst_mci)
    a.TP["mciof_c", "mci_i",    t] <- p_mci_stay * p_surv * p_inst_mci
    
    a.TP["mciof_c", "milof_c",  t] <- p_mci_mil * p_surv * (1 - p_inst_mci)
    a.TP["mciof_c", "mil_i",    t] <- p_mci_mil * p_surv * p_inst_mci
    
    a.TP["mciof_c", "mod_c",    t] <- p_mci_mod * p_surv
    a.TP["mciof_c", "sev_c",    t] <- p_mci_sev * p_surv
    
    # Transitions from "milof_c"
    p_mil_mci      <- if (strat == "soc") v.p.mil_mci_soc[t] else v.p.mil_mci_int_off[t]
    p_mil_mod      <- if (strat == "soc") v.p.mil_mod_soc[t] else v.p.mil_mod_int_off[t]
    p_mil_sev      <- if (strat == "soc") v.p.mil_sev_soc[t] else v.p.mil_sev_int_off[t]
    p_mil_stay     <- if (strat == "soc") v.p.mil_stay_soc[t] else v.p.mil_stay_int_off[t]
    p_inst_mil     <- v.p.mil_i[t]
    p_surv         <- 1 - a.TP["milof_c", "dth", t]
    
    a.TP["milof_c", "milof_c", t] <- p_mil_stay * p_surv * (1 - p_inst_mil)
    a.TP["milof_c", "mil_i",    t] <- p_mil_stay * p_surv * p_inst_mil
    
    a.TP["milof_c", "mciof_c",  t] <- p_mil_mci * p_surv * (1 - p_inst_mil)
    a.TP["milof_c", "mci_i",    t] <- p_mil_mci * p_surv * p_inst_mil
    
    a.TP["milof_c", "mod_c",    t] <- p_mil_mod * p_surv
    a.TP["milof_c", "sev_c",    t] <- p_mil_sev * p_surv
    
    # Transitions from "mod_c"
    p_mod_mil   <- if (strat == "soc") v.p.mod_mil_soc[t] else v.p.mod_mil_soc[t]
    p_mod_sev   <- if (strat == "soc") v.p.mod_sev_soc[t] else v.p.mod_sev_soc[t]
    p_mod_stay  <- if (strat == "soc") v.p.mod_stay_soc[t] else v.p.mod_stay_soc[t]
    p_inst_mod  <- v.p.mod_i[t]
    p_surv      <- 1 - a.TP["mod_c", "dth", t]
    
    a.TP["mod_c", "mod_c", t] <- p_mod_stay * p_surv * (1 - p_inst_mod)
    a.TP["mod_c", "mod_i", t] <- p_mod_stay * p_surv * p_inst_mod
    
    a.TP["mod_c", "milof_c", t] <- p_mod_mil * p_surv * (1 - p_inst_mod)
    a.TP["mod_c", "mil_i",   t] <- p_mod_mil * p_surv * p_inst_mod
    
    a.TP["mod_c", "sev_c",   t] <- p_mod_sev * p_surv
    
    # Transitions from "sev_c"
    p_sev_mil   <- if (strat == "soc") v.p.sev_mil_soc[t] else v.p.sev_mil_soc[t]
    p_sev_mod   <- if (strat == "soc") v.p.sev_mod_soc[t] else v.p.sev_mod_soc[t]
    p_sev_stay  <- if (strat == "soc") v.p.sev_stay_soc[t] else v.p.sev_stay_soc[t]
    p_inst_sev  <- v.p.sev_i[t]
    p_surv      <- 1 - a.TP["sev_c", "dth", t]
    
    a.TP["sev_c", "sev_c", t] <- p_sev_stay * p_surv * (1 - p_inst_sev)
    a.TP["sev_c", "sev_i", t] <- p_sev_stay * p_surv * p_inst_sev
    
    a.TP["sev_c", "milof_c", t] <- p_sev_mil * p_surv * (1 - p_inst_sev)
    a.TP["sev_c", "mil_i",   t] <- p_sev_mil * p_surv * p_inst_sev
    
    a.TP["sev_c", "mod_c",   t] <- p_sev_mod * p_surv
    
    #------------------------------------------------------------
    # [INSERT PROMPT 5D HERE] — Institutionalized state transitions
    #------------------------------------------------------------
    
    # Transitions from "mci_i"
    p_mci_mil <- if (strat == "soc") v.p.mci_mil_soc[t] else v.p.mci_mil_int_off[t]
    p_mci_mod <- if (strat == "soc") v.p.mci_mod_soc[t] else v.p.mci_mod_int_off[t]
    p_mci_sev <- if (strat == "soc") v.p.mci_sev_soc[t] else v.p.mci_sev_int_off[t]
    p_mci_stay <- if (strat == "soc") v.p.mci_stay_soc[t] else v.p.mci_stay_int_off[t]
    p_surv <- 1 - a.TP["mci_i", "dth", t]
    
    a.TP["mci_i", "mci_i", t] <- p_mci_stay * p_surv
    a.TP["mci_i", "mil_i", t] <- p_mci_mil * p_surv
    a.TP["mci_i", "mod_i", t] <- p_mci_mod * p_surv
    a.TP["mci_i", "sev_i", t] <- p_mci_sev * p_surv
    
    # Transitions from "mil_i"
    p_mil_mci <- if (strat == "soc") v.p.mil_mci_soc[t] else v.p.mil_mci_int_off[t]
    p_mil_mod <- if (strat == "soc") v.p.mil_mod_soc[t] else v.p.mil_mod_int_off[t]
    p_mil_sev <- if (strat == "soc") v.p.mil_sev_soc[t] else v.p.mil_sev_int_off[t]
    p_mil_stay <- if (strat == "soc") v.p.mil_stay_soc[t] else v.p.mil_stay_int_off[t]
    p_surv <- 1 - a.TP["mil_i", "dth", t]
    
    a.TP["mil_i", "mil_i", t] <- p_mil_stay * p_surv
    a.TP["mil_i", "mci_i", t] <- p_mil_mci * p_surv
    a.TP["mil_i", "mod_i", t] <- p_mil_mod * p_surv
    a.TP["mil_i", "sev_i", t] <- p_mil_sev * p_surv
    
    # Transitions from "mod_i"
    p_mod_mil <- v.p.mod_mil_soc[t]
    p_mod_sev <- v.p.mod_sev_soc[t]
    p_mod_stay <- v.p.mod_stay_soc[t]
    p_surv <- 1 - a.TP["mod_i", "dth", t]
    
    a.TP["mod_i", "mod_i", t] <- p_mod_stay * p_surv
    a.TP["mod_i", "mil_i", t] <- p_mod_mil * p_surv
    a.TP["mod_i", "sev_i", t] <- p_mod_sev * p_surv
    
    # Transitions from "sev_i"
    p_sev_mil <- v.p.sev_mil_soc[t]
    p_sev_mod <- v.p.sev_mod_soc[t]
    p_sev_stay <- v.p.sev_stay_soc[t]
    p_surv <- 1 - a.TP["sev_i", "dth", t]
    
    a.TP["sev_i", "sev_i", t] <- p_sev_stay * p_surv
    a.TP["sev_i", "mil_i", t] <- p_sev_mil * p_surv
    a.TP["sev_i", "mod_i", t] <- p_sev_mod * p_surv
    
    # Normalize rows to sum to 1 (excluding absorbing state)
    for (s in v.names_state) {
      row_sum <- sum(a.TP[s, , t])
      if (row_sum > 0 && row_sum != 1) {
        a.TP[s, , t] <- a.TP[s, , t] / row_sum
      }
    }
  }
  
  return(a.TP)
}
#=============================#
# Markov Trace and Outcomes   #
#=============================#

compute_outcomes <- function(strat) {
  # Build transition matrix
  a.TP <- build_transition_matrix(strat)
  
  # Initialize Markov trace matrix (states x cycles)
  m.M <- matrix(0, nrow = length(v.names_state), ncol = n.cycle,
                dimnames = list(v.names_state, NULL))
  
  # Set initial state distribution
  v.init <- rep(0, length(v.names_state))
  names(v.init) <- v.names_state
  v.init["mciof_c"] <- p.starting_state_mci
  v.init["milof_c"] <- 1 - p.starting_state_mci
  m.M[, 1] <- v.init
  
  # Named utility vector (patient + caregiver)
  v.u <- c(
    "mcion_c" = u.mci_pt + u.mci_ic,
    "mciof_c" = u.mci_pt + u.mci_ic,
    "milon_c" = u.mil_pt + u.mil_ic,
    "milof_c" = u.mil_pt + u.mil_ic,
    "mod_c"   = u.mod_pt + u.mod_ic,
    "sev_c"   = u.sev_pt + u.sev_ic,
    "mci_i"   = u.mci_pt_i + u.mci_ic_i,
    "mil_i"   = u.mil_pt_i + u.mil_ic_i,
    "mod_i"   = u.mod_pt_i + u.mod_ic_i,
    "sev_i"   = u.sev_pt_i + u.sev_ic_i,
    "dth"     = 0
  )
  
  # Named cost vector (health care + social + informal care)
  v.c <- c(
    "mcion_c" = c.mci_hc + c.mci_sc + c.mci_ic,
    "mciof_c" = c.mci_hc + c.mci_sc + c.mci_ic,
    "milon_c" = c.mil_hc + c.mil_sc + c.mil_ic,
    "milof_c" = c.mil_hc + c.mil_sc + c.mil_ic,
    "mod_c"   = c.mod_hc + c.mod_sc + c.mod_ic,
    "sev_c"   = c.sev_hc + c.sev_sc + c.sev_ic,
    "mci_i"   = c.mci_i_hc + c.mci_i_sc + c.mci_i_ic,
    "mil_i"   = c.mil_i_hc + c.mil_i_sc + c.mil_i_ic,
    "mod_i"   = c.mod_i_hc + c.mod_i_sc + c.mod_i_ic,
    "sev_i"   = c.sev_i_hc + c.sev_i_sc + c.sev_i_ic,
    "dth"     = 0
  )
  
  # Initialize output vectors
  v.QALY <- numeric(n.cycle)
  v.Cost <- numeric(n.cycle)
  v.LY   <- numeric(n.cycle)
  
  # Compute outcomes over time
  for (t in 1:(n.cycle - 1)) {
    m.M[, t + 1] <- m.M[, t] %*% a.TP[, , t]
    
    v.QALY[t] <- sum(m.M[, t] * v.u)
    v.Cost[t] <- sum(m.M[, t] * v.c)
    v.LY[t]   <- sum(m.M[, t] * (v.names_state != "dth"))
    
    # Add recurring treatment cost for on-treatment states (if int)
    if (strat == "int") {
      v.Cost[t] <- v.Cost[t] + c.Tx * (m.M["mcion_c", t] + m.M["milon_c", t])
    }
  }
  
  # Last cycle outcomes (no Markov update needed)
  v.QALY[n.cycle] <- sum(m.M[, n.cycle] * v.u)
  v.Cost[n.cycle] <- sum(m.M[, n.cycle] * v.c)
  v.LY[n.cycle]   <- sum(m.M[, n.cycle] * (v.names_state != "dth"))
  if (strat == "int") {
    v.Cost[n.cycle] <- v.Cost[n.cycle] + c.Tx * (m.M["mcion_c", n.cycle] + m.M["milon_c", n.cycle])
  }
  
  # Apply initial treatment effects (first cycle only)
  if (strat == "int") {
    v.QALY[1] <- v.QALY[1] + u.Tx_start
    v.Cost[1] <- v.Cost[1] + c.Tx_start
  }
  
  # Apply half-cycle correction
  v.QALY <- 0.5 * (v.QALY[-1] + v.QALY[-n.cycle])
  v.Cost <- 0.5 * (v.Cost[-1] + v.Cost[-n.cycle])
  v.LY   <- 0.5 * (v.LY[-1]   + v.LY[-n.cycle])
  
  # Apply discounting
  v.discount_weights <- 1 / ((1 + discount_QALY) ^ (1:(n.cycle - 1)))
  total.QALY <- sum(v.QALY * v.discount_weights)
  total.Cost <- sum(v.Cost * v.discount_weights)
  total.LY   <- sum(v.LY   * v.discount_weights)
  
  return(c(QALYs = total.QALY, Costs = total.Cost, LYs = total.LY))
}

#=============================#
# Run and compare strategies #
#=============================#

res.soc <- compute_outcomes("soc")
res.int <- compute_outcomes("int")

cat("===== IPECAD Strategy Comparison =====\n")
cat(sprintf("Standard of Care:    QALYs = %.2f | Costs = $%.0f | LYs = %.2f\n", 
            res.soc["QALYs"], res.soc["Costs"], res.soc["LYs"]))
cat(sprintf("Intervention:        QALYs = %.2f | Costs = $%.0f | LYs = %.2f\n", 
            res.int["QALYs"], res.int["Costs"], res.int["LYs"]))
cat(sprintf("Incremental (INT - SOC):\n"))
cat(sprintf("  ΔQALYs = %.2f\n", res.int["QALYs"] - res.soc["QALYs"]))
cat(sprintf("  ΔCosts = $%.0f\n", res.int["Costs"] - res.soc["Costs"]))
cat(sprintf("  ΔLYs = %.2f\n", res.int["LYs"] - res.soc["LYs"]))
```
