## Supplementary material for "Replicating Health-Economic Simulation Models for Alzheimer’s Disease Using Artificial Intelligence": final R scripts: o.R

```
# ----------------------------
# IPECAD Markov Model Inputs
# ----------------------------

# Load required package
library(dampack)

# 1. Define State and Strategy Names
state_names <- c("mcion_c", "mciof_c", "milon_c", "milof_c", "mod_c", "sev_c",
                 "mci_i", "mil_i", "mod_i", "sev_i", "dth")
strategy_names <- c("soc", "int")

# 2. Cohort Characteristics
age_start <- 71
sex <- "weighted"  # 52% female
p_starting_state_mci <- 0.55
n_cycles <- 29

# 3. Transition Probabilities (per year)
p_mci_mil <- 0.23
p_mci_mod <- 0
p_mci_sev <- 0
p_mil_mci <- 0.03
p_mil_mod <- 0.35
p_mil_sev <- 0.04
p_mod_mil <- 0.03
p_mod_sev <- 0.42
p_sev_mil <- 0
p_sev_mod <- 0.02

# Transitions to institution
p_mci_i <- 0.024
p_mil_i <- 0.038
p_mod_i <- 0.110
p_sev_i <- 0.259

# 4. Mortality Hazard Ratios
hr_mort_mci <- 1.82
hr_mort_mil <- 2.92
hr_mort_mod <- 3.85
hr_mort_sev <- 9.52

# 5. Relative Risks for Treatment Effects (on treatment)
rr_tx_mci_mil <- 0.69
rr_tx_mci_mod <- 1
rr_tx_mci_sev <- 1
rr_tx_mil_mod <- 0.69
rr_tx_mil_sev <- 0.69

# Relative Risks for Treatment Effects (off treatment)
rr_tx_mci_mil_dis <- 1
rr_tx_mci_mod_dis <- 1
rr_tx_mci_sev_dis <- 1
rr_tx_mil_mod_dis <- 1
rr_tx_mil_sev_dis <- 1

# 6. Treatment Discontinuation
p_discontinuation1 <- 0.069
p_discontinuation2 <- 0
discontinuation2_begin <- 2
tx_duration <- 29
tx_waning <- 0
tx_waning_dis <- 0

# 7. Utilities (Patients - Community)
u_mci_pt <- 0.851 - 0.17
u_mil_pt <- 0.851 - 0.22
u_mod_pt <- 0.851 - 0.36
u_sev_pt <- 0.851 - 0.53

# Utilities (Patients - Institution)
u_mci_pt_i <- 0.851 - 0.17
u_mil_pt_i <- 0.851 - 0.19
u_mod_pt_i <- 0.851 - 0.42
u_sev_pt_i <- 0.851 - 0.59

# Utilities (Informal Caregivers - Community)
u_mci_ic <- -0.03
u_mil_ic <- -0.05
u_mod_ic <- -0.08
u_sev_ic <- -0.10

# Utilities (Informal Caregivers - Institution)
u_mci_ic_i <- -0.03
u_mil_ic_i <- -0.05
u_mod_ic_i <- -0.08
u_sev_ic_i <- -0.10

# Utility Loss from ARIA (one-time, first cycle)
u_Tx_start <- -0.14 * (12/52) * 0.035

# 8. Costs (Healthcare, Social, Informal)

# Community Healthcare Costs
c_mci_hc <- 6042 * 1.12 + 460
c_mil_hc <- 6042 * 1.56 + 965 + 0.21 * 365 * 0.333
c_mod_hc <- 6042 * 1.93 + 1544 + 0.66 * 365 * 0.333
c_sev_hc <- 6042 * 1.93 + 1930

# Institution Healthcare Costs
c_mci_i_hc <- c_mci_hc
c_mil_i_hc <- c_mil_hc
c_mod_i_hc <- c_mod_hc
c_sev_i_hc <- c_sev_hc

# Community Social Costs
c_mci_sc <- 0
c_mil_sc <- 0
c_mod_sc <- 0
c_sev_sc <- 0

# Institution Social Costs
c_mci_i_sc <- 7394 * 12
c_mil_i_sc <- c_mci_i_sc
c_mod_i_sc <- c_mci_i_sc
c_sev_i_sc <- c_mci_i_sc

# Informal Care Costs (Community)
c_mci_ic <- 69 * 12 * 32.46 + 0.204 * 0.049 * 20 * 52 * 32.46
c_mil_ic <- 113 * 12 * 32.46 + 0.112 * 0.086 * 20 * 52 * 32.46
c_mod_ic <- 169 * 12 * 32.46
c_sev_ic <- 298 * 12 * 32.46

# Informal Care Costs (Institution)
c_mci_i_ic <- 69 * 12 * 32.46 * 0.44 + 0.204 * 0.049 * 20 * 52 * 32.46
c_mil_i_ic <- 113 * 12 * 32.46 * 0.44 + 0.112 * 0.086 * 20 * 52 * 32.46
c_mod_i_ic <- 169 * 12 * 32.46 * 0.44
c_sev_i_ic <- 298 * 12 * 32.46 * 0.44

# Treatment Costs
c_Tx <- 26500 + (52 / 2) * 78.35
c_Tx_start <- 261.10 * 4 + 261.10 * 3 * 0.215

# 9. Discount Rates
discount_EFFECT <- 0.03
discount_QALY <- 0.03
discount_COST <- 0.03

# 10. WTP Threshold
wtp <- 100000

# 11. Half-cycle correction
half_cycle_correction <- TRUE
# ----------------------------
# Load and Process Life Table
# ----------------------------

# Load the CSV file
life_table <- read.csv("lifetable_US_2019_ssa.csv")

# Convert annual mortality probabilities to mortality rates using: rate = -log(1 - prob)
life_table$male <- -log(1 - life_table$male)
life_table$female <- -log(1 - life_table$female)

# Define weighted average using 52% female and 48% male (from earlier parameter)
life_table$weighted <- life_table$male * 0.48 + life_table$female * 0.52

# Retain only required columns
life_table <- life_table[, c("age", "male", "female", "weighted")]

# Return processed life table
life_table


# --------------------------------------------
# Time-Dependent Transition Vectors for SOC
# --------------------------------------------

# 1. Initialize time index
cycles <- 1:n_cycles

# 2. Disease progression vectors for SOC (same each year unless specified)
v.p.mci_mil_soc <- rep(p_mci_mil, n_cycles)
v.p.mci_mod_soc <- rep(p_mci_mod, n_cycles)
v.p.mci_sev_soc <- rep(p_mci_sev, n_cycles)

v.p.mil_mci_soc <- rep(p_mil_mci, n_cycles)
v.p.mil_mod_soc <- rep(p_mil_mod, n_cycles)
v.p.mil_sev_soc <- rep(p_mil_sev, n_cycles)

v.p.mod_mil_soc <- rep(p_mod_mil, n_cycles)
v.p.mod_sev_soc <- rep(p_mod_sev, n_cycles)

v.p.sev_mil_soc <- rep(p_sev_mil, n_cycles)
v.p.sev_mod_soc <- rep(p_sev_mod, n_cycles)

# 3. Stay probabilities (exclude transitions to institution and death)
v.p.mci_stay_soc <- 1 - (v.p.mci_mil_soc + v.p.mci_mod_soc + v.p.mci_sev_soc)
v.p.mil_stay_soc <- 1 - (v.p.mil_mci_soc + v.p.mil_mod_soc + v.p.mil_sev_soc)
v.p.mod_stay_soc <- 1 - (v.p.mod_mil_soc + v.p.mod_sev_soc)
v.p.sev_stay_soc <- 1 - (v.p.sev_mil_soc + v.p.sev_mod_soc)

# 4. Institutionalization probabilities (same each year)
v.p.mci_i <- rep(p_mci_i, n_cycles)
v.p.mil_i <- rep(p_mil_i, n_cycles)
v.p.mod_i <- rep(p_mod_i, n_cycles)
v.p.sev_i <- rep(p_sev_i, n_cycles)

# 5. Discontinuation vector per cycle (not cumulative)
v.p.discontinue <- rep(NA, n_cycles)
v.p.discontinue[1:(discontinuation2_begin - 1)] <- p_discontinuation1
v.p.discontinue[discontinuation2_begin:(tx_duration)] <- p_discontinuation2
v.p.discontinue[(tx_duration + 1):n_cycles] <- 1

# 6. Mortality vector (age- and sex-weighted) from mortality table
# Assume 'life_table' (from Prompt 2) is already loaded and processed
v.mortality <- life_table$weighted[match(age_start + cycles - 1, life_table$age)]


# ------------------------------------------------------------
# Time-Dependent Transition Vectors for Intervention Strategy
# ------------------------------------------------------------

# Initialize cycle index
cycles <- 1:n_cycles

# Waning factors
waning_factor <- (1 - tx_waning)^(cycles - 1)
waning_factor_dis <- (1 - tx_waning_dis)^(cycles - 1)

# 1. On-Treatment Transitions (mcion_c)
# Apply treatment effect and waning to forward transitions only
r_mci_mil <- -log(1 - p_mci_mil)
r_mci_mod <- -log(1 - p_mci_mod)
r_mci_sev <- -log(1 - p_mci_sev)

v.p.mci_mil_int_on <- 1 - exp(-r_mci_mil * rr_tx_mci_mil ^ waning_factor)
v.p.mci_mod_int_on <- 1 - exp(-r_mci_mod * rr_tx_mci_mod ^ waning_factor)
v.p.mci_sev_int_on <- 1 - exp(-r_mci_sev * rr_tx_mci_sev ^ waning_factor)

# Stay probability (excluding institutional transitions)
v.p.mci_stay_int_on <- 1 - (v.p.mci_mil_int_on + v.p.mci_mod_int_on + v.p.mci_sev_int_on)

# 2. On-Treatment Transitions (milon_c)
r_mil_mci <- -log(1 - p_mil_mci)  # No treatment effect
r_mil_mod <- -log(1 - p_mil_mod)
r_mil_sev <- -log(1 - p_mil_sev)

v.p.mil_mci_int_on <- rep(p_mil_mci, n_cycles)  # Backward transition, no treatment effect
v.p.mil_mod_int_on <- 1 - exp(-r_mil_mod * rr_tx_mil_mod ^ waning_factor)
v.p.mil_sev_int_on <- 1 - exp(-r_mil_sev * rr_tx_mil_sev ^ waning_factor)

v.p.mil_stay_int_on <- 1 - (v.p.mil_mci_int_on + v.p.mil_mod_int_on + v.p.mil_sev_int_on)

# 3. Off-Treatment Transitions (mciof_c)
v.p.mci_mil_int_off <- 1 - exp(-r_mci_mil * rr_tx_mci_mil_dis ^ waning_factor_dis)
v.p.mci_mod_int_off <- 1 - exp(-r_mci_mod * rr_tx_mci_mod_dis ^ waning_factor_dis)
v.p.mci_sev_int_off <- 1 - exp(-r_mci_sev * rr_tx_mci_sev_dis ^ waning_factor_dis)

v.p.mci_stay_int_off <- 1 - (v.p.mci_mil_int_off + v.p.mci_mod_int_off + v.p.mci_sev_int_off)

# 4. Off-Treatment Transitions (milof_c)
v.p.mil_mci_int_off <- rep(p_mil_mci, n_cycles)  # No treatment effect
v.p.mil_mod_int_off <- 1 - exp(-r_mil_mod * rr_tx_mil_mod_dis ^ waning_factor_dis)
v.p.mil_sev_int_off <- 1 - exp(-r_mil_sev * rr_tx_mil_sev_dis ^ waning_factor_dis)

v.p.mil_stay_int_off <- 1 - (v.p.mil_mci_int_off + v.p.mil_mod_int_off + v.p.mil_sev_int_off)


# -------------------------------------------------------------
# Function to Initialize Transition Probability Matrix: build_transition_matrix
# -------------------------------------------------------------

build_transition_matrix <- function(strat) {
  
  # Define dimensions
  n_states <- length(state_names)
  
  # Initialize 3D transition probability array: states × states × cycles
  a.TP <- array(0, dim = c(n_states, n_states, n_cycles),
                dimnames = list(state_names, state_names, NULL))
  
  # Loop over each cycle
  for (t in 1:n_cycles) {
    
    # -------------------------
    # Transitions to Death
    # -------------------------
    
    # Calculate mortality rates per state (adjusted by HR for each health state)
    rate_mci <- v.mortality[t] * hr_mort_mci
    rate_mil <- v.mortality[t] * hr_mort_mil
    rate_mod <- v.mortality[t] * hr_mort_mod
    rate_sev <- v.mortality[t] * hr_mort_sev
    
    # Compute transition probabilities to death from each living state
    a.TP["mcion_c", "dth", t] <- 1 - exp(-rate_mci)
    a.TP["mciof_c", "dth", t] <- 1 - exp(-rate_mci)
    a.TP["milon_c", "dth", t] <- 1 - exp(-rate_mil)
    a.TP["milof_c", "dth", t] <- 1 - exp(-rate_mil)
    a.TP["mod_c",   "dth", t] <- 1 - exp(-rate_mod)
    a.TP["sev_c",   "dth", t] <- 1 - exp(-rate_sev)
    a.TP["mci_i",   "dth", t] <- 1 - exp(-rate_mci)
    a.TP["mil_i",   "dth", t] <- 1 - exp(-rate_mil)
    a.TP["mod_i",   "dth", t] <- 1 - exp(-rate_mod)
    a.TP["sev_i",   "dth", t] <- 1 - exp(-rate_sev)
    
    # Absorbing state: remain in death
    a.TP["dth", "dth", t] <- 1
    
    # -------------------------
    # Placeholders for Prompts
    # -------------------------
    
    # -------------------------
    # Transitions from "mcion_c"
    # -------------------------
    if (strat == "soc") {
      p_mci_mil <- v.p.mci_mil_soc[t]
      p_mci_mod <- v.p.mci_mod_soc[t]
      p_mci_sev <- v.p.mci_sev_soc[t]
      p_mci_stay <- v.p.mci_stay_soc[t]
    } else if (strat == "int") {
      p_mci_mil <- v.p.mci_mil_int_on[t]
      p_mci_mod <- v.p.mci_mod_int_on[t]
      p_mci_sev <- v.p.mci_sev_int_on[t]
      p_mci_stay <- v.p.mci_stay_int_on[t]
    }
    
    surv <- 1 - a.TP["mcion_c", "dth", t]
    p_inst <- v.p.mci_i[t]
    p_disc <- v.p.discontinue[t]
    
    # On-treatment transitions (only to on-treatment and off-treatment states)
    a.TP["mcion_c", "mcion_c", t] <- p_mci_stay * (1 - p_inst) * surv * (1 - p_disc)
    a.TP["mcion_c", "mciof_c", t] <- p_mci_stay * (1 - p_inst) * surv * p_disc
    a.TP["mcion_c", "milon_c", t] <- p_mci_mil * (1 - p_inst) * surv * (1 - p_disc)
    a.TP["mcion_c", "milof_c", t] <- p_mci_mil * (1 - p_inst) * surv * p_disc
    a.TP["mcion_c", "mod_c", t]   <- p_mci_mod * (1 - p_inst) * surv  # no discontinuation adjustment
    a.TP["mcion_c", "sev_c", t]   <- p_mci_sev * (1 - p_inst) * surv  # no discontinuation adjustment
    a.TP["mcion_c", "mci_i", t]   <- p_mci_stay * p_inst * surv       # no discontinuation adjustment
    a.TP["mcion_c", "mil_i", t]   <- p_mci_mil * p_inst * surv        # no discontinuation adjustment
    a.TP["mcion_c", "mod_i", t]   <- p_mci_mod * p_inst * surv        # no discontinuation adjustment
    a.TP["mcion_c", "sev_i", t]   <- p_mci_sev * p_inst * surv        # no discontinuation adjustment
    
    # -------------------------
    # Transitions from "milon_c"
    # -------------------------
    if (strat == "soc") {
      p_mil_mci <- v.p.mil_mci_soc[t]
      p_mil_mod <- v.p.mil_mod_soc[t]
      p_mil_sev <- v.p.mil_sev_soc[t]
      p_mil_stay <- v.p.mil_stay_soc[t]
    } else if (strat == "int") {
      p_mil_mci <- v.p.mil_mci_int_on[t]
      p_mil_mod <- v.p.mil_mod_int_on[t]
      p_mil_sev <- v.p.mil_sev_int_on[t]
      p_mil_stay <- v.p.mil_stay_int_on[t]
    }
    
    surv <- 1 - a.TP["milon_c", "dth", t]
    p_inst <- v.p.mil_i[t]
    p_disc <- v.p.discontinue[t]
    
    a.TP["milon_c", "milon_c", t] <- p_mil_stay * (1 - p_inst) * surv * (1 - p_disc)
    a.TP["milon_c", "milof_c", t] <- p_mil_stay * (1 - p_inst) * surv * p_disc
    a.TP["milon_c", "mcion_c", t] <- p_mil_mci * (1 - p_inst) * surv * (1 - p_disc)
    a.TP["milon_c", "mciof_c", t] <- p_mil_mci * (1 - p_inst) * surv * p_disc
    a.TP["milon_c", "mod_c", t]   <- p_mil_mod * (1 - p_inst) * surv
    a.TP["milon_c", "sev_c", t]   <- p_mil_sev * (1 - p_inst) * surv
    a.TP["milon_c", "mci_i", t]   <- p_mil_mci * p_inst * surv
    a.TP["milon_c", "mil_i", t]   <- p_mil_stay * p_inst * surv
    a.TP["milon_c", "mod_i", t]   <- p_mil_mod * p_inst * surv
    a.TP["milon_c", "sev_i", t]   <- p_mil_sev * p_inst * surv
    
    # -------------------------
    # Transitions from "mciof_c"
    # -------------------------
    if (strat == "soc") {
      p_mci_mil <- v.p.mci_mil_soc[t]
      p_mci_mod <- v.p.mci_mod_soc[t]
      p_mci_sev <- v.p.mci_sev_soc[t]
      p_mci_stay <- v.p.mci_stay_soc[t]
    } else {
      p_mci_mil <- v.p.mci_mil_int_off[t]
      p_mci_mod <- v.p.mci_mod_int_off[t]
      p_mci_sev <- v.p.mci_sev_int_off[t]
      p_mci_stay <- v.p.mci_stay_int_off[t]
    }
    
    surv <- 1 - a.TP["mciof_c", "dth", t]
    p_inst <- v.p.mci_i[t]
    
    a.TP["mciof_c", "mciof_c", t] <- p_mci_stay * (1 - p_inst) * surv
    a.TP["mciof_c", "milof_c", t] <- p_mci_mil * (1 - p_inst) * surv
    a.TP["mciof_c", "mod_c", t]   <- p_mci_mod * (1 - p_inst) * surv
    a.TP["mciof_c", "sev_c", t]   <- p_mci_sev * (1 - p_inst) * surv
    a.TP["mciof_c", "mci_i", t]   <- p_mci_stay * p_inst * surv
    a.TP["mciof_c", "mil_i", t]   <- p_mci_mil * p_inst * surv
    a.TP["mciof_c", "mod_i", t]   <- p_mci_mod * p_inst * surv
    a.TP["mciof_c", "sev_i", t]   <- p_mci_sev * p_inst * surv
    
    # -------------------------
    # Transitions from "milof_c"
    # -------------------------
    if (strat == "soc") {
      p_mil_mci <- v.p.mil_mci_soc[t]
      p_mil_mod <- v.p.mil_mod_soc[t]
      p_mil_sev <- v.p.mil_sev_soc[t]
      p_mil_stay <- v.p.mil_stay_soc[t]
    } else {
      p_mil_mci <- v.p.mil_mci_int_off[t]
      p_mil_mod <- v.p.mil_mod_int_off[t]
      p_mil_sev <- v.p.mil_sev_int_off[t]
      p_mil_stay <- v.p.mil_stay_int_off[t]
    }
    
    surv <- 1 - a.TP["milof_c", "dth", t]
    p_inst <- v.p.mil_i[t]
    
    a.TP["milof_c", "milof_c", t] <- p_mil_stay * (1 - p_inst) * surv
    a.TP["milof_c", "mciof_c", t] <- p_mil_mci * (1 - p_inst) * surv
    a.TP["milof_c", "mod_c", t]   <- p_mil_mod * (1 - p_inst) * surv
    a.TP["milof_c", "sev_c", t]   <- p_mil_sev * (1 - p_inst) * surv
    a.TP["milof_c", "mci_i", t]   <- p_mil_mci * p_inst * surv
    a.TP["milof_c", "mil_i", t]   <- p_mil_stay * p_inst * surv
    a.TP["milof_c", "mod_i", t]   <- p_mil_mod * p_inst * surv
    a.TP["milof_c", "sev_i", t]   <- p_mil_sev * p_inst * surv
    
    # -------------------------
    # Transitions from "mod_c"
    # -------------------------
    p_mod_mil <- v.p.mod_mil_soc[t]
    p_mod_sev <- v.p.mod_sev_soc[t]
    p_mod_stay <- v.p.mod_stay_soc[t]
    surv <- 1 - a.TP["mod_c", "dth", t]
    p_inst <- v.p.mod_i[t]
    
    a.TP["mod_c", "mod_c", t] <- p_mod_stay * (1 - p_inst) * surv
    a.TP["mod_c", "milof_c", t] <- p_mod_mil * (1 - p_inst) * surv
    a.TP["mod_c", "sev_c", t]   <- p_mod_sev * (1 - p_inst) * surv
    a.TP["mod_c", "mil_i", t]   <- p_mod_mil * p_inst * surv
    a.TP["mod_c", "mod_i", t]   <- p_mod_stay * p_inst * surv
    a.TP["mod_c", "sev_i", t]   <- p_mod_sev * p_inst * surv
    
    # -------------------------
    # Transitions from "sev_c"
    # -------------------------
    p_sev_mil <- v.p.sev_mil_soc[t]
    p_sev_mod <- v.p.sev_mod_soc[t]
    p_sev_stay <- v.p.sev_stay_soc[t]
    surv <- 1 - a.TP["sev_c", "dth", t]
    p_inst <- v.p.sev_i[t]
    
    a.TP["sev_c", "sev_c", t] <- p_sev_stay * (1 - p_inst) * surv
    a.TP["sev_c", "milof_c", t] <- p_sev_mil * (1 - p_inst) * surv
    a.TP["sev_c", "mod_c", t]   <- p_sev_mod * (1 - p_inst) * surv
    a.TP["sev_c", "mil_i", t]   <- p_sev_mil * p_inst * surv
    a.TP["sev_c", "mod_i", t]   <- p_sev_mod * p_inst * surv
    a.TP["sev_c", "sev_i", t]   <- p_sev_stay * p_inst * surv
    
    # -------------------------
    # Transitions from "mci_i"
    # -------------------------
    if (strat == "soc") {
      p_mci_mil <- v.p.mci_mil_soc[t]
      p_mci_mod <- v.p.mci_mod_soc[t]
      p_mci_sev <- v.p.mci_sev_soc[t]
      p_mci_stay <- v.p.mci_stay_soc[t]
    } else {
      p_mci_mil <- v.p.mci_mil_int_off[t]
      p_mci_mod <- v.p.mci_mod_int_off[t]
      p_mci_sev <- v.p.mci_sev_int_off[t]
      p_mci_stay <- v.p.mci_stay_int_off[t]
    }
    
    surv <- 1 - a.TP["mci_i", "dth", t]
    
    a.TP["mci_i", "mci_i", t] <- p_mci_stay * surv
    a.TP["mci_i", "mil_i", t] <- p_mci_mil * surv
    a.TP["mci_i", "mod_i", t] <- p_mci_mod * surv
    a.TP["mci_i", "sev_i", t] <- p_mci_sev * surv
    
    # -------------------------
    # Transitions from "mil_i"
    # -------------------------
    if (strat == "soc") {
      p_mil_mci <- v.p.mil_mci_soc[t]
      p_mil_mod <- v.p.mil_mod_soc[t]
      p_mil_sev <- v.p.mil_sev_soc[t]
      p_mil_stay <- v.p.mil_stay_soc[t]
    } else {
      p_mil_mci <- v.p.mil_mci_int_off[t]
      p_mil_mod <- v.p.mil_mod_int_off[t]
      p_mil_sev <- v.p.mil_sev_int_off[t]
      p_mil_stay <- v.p.mil_stay_int_off[t]
    }
    
    surv <- 1 - a.TP["mil_i", "dth", t]
    
    a.TP["mil_i", "mil_i", t] <- p_mil_stay * surv
    a.TP["mil_i", "mci_i", t] <- p_mil_mci * surv
    a.TP["mil_i", "mod_i", t] <- p_mil_mod * surv
    a.TP["mil_i", "sev_i", t] <- p_mil_sev * surv
    
    # -------------------------
    # Transitions from "mod_i"
    # -------------------------
    p_mod_mil <- v.p.mod_mil_soc[t]
    p_mod_sev <- v.p.mod_sev_soc[t]
    p_mod_stay <- v.p.mod_stay_soc[t]
    surv <- 1 - a.TP["mod_i", "dth", t]
    
    a.TP["mod_i", "mod_i", t] <- p_mod_stay * surv
    a.TP["mod_i", "mil_i", t] <- p_mod_mil * surv
    a.TP["mod_i", "sev_i", t] <- p_mod_sev * surv
    
    # -------------------------
    # Transitions from "sev_i"
    # -------------------------
    p_sev_mil <- v.p.sev_mil_soc[t]
    p_sev_mod <- v.p.sev_mod_soc[t]
    p_sev_stay <- v.p.sev_stay_soc[t]
    surv <- 1 - a.TP["sev_i", "dth", t]
    
    a.TP["sev_i", "sev_i", t] <- p_sev_stay * surv
    a.TP["sev_i", "mil_i", t] <- p_sev_mil * surv
    a.TP["sev_i", "mod_i", t] <- p_sev_mod * surv
    
    # -------------------------
    # Normalize each row if sum ≠ 1
    # -------------------------
    for (s in state_names) {
      row_sum <- sum(a.TP[s, , t])
      if (row_sum > 0 && row_sum != 1) {
        a.TP[s, , t] <- a.TP[s, , t] / row_sum
      }
    }
  }
  
  return(a.TP)
}


# ---------------------------------------------------------------
# Compute Markov Trace and Total Outcomes for a Given Strategy
# ---------------------------------------------------------------

compute_outcomes <- function(strat) {
  
  # Build transition matrix array
  a.TP <- build_transition_matrix(strat)
  
  # Initialize state trace matrix (rows = cycles, cols = states)
  m.Trace <- matrix(0, nrow = n_cycles + 1, ncol = length(state_names),
                    dimnames = list(0:n_cycles, state_names))
  
  # Set initial distribution: all patients start in MCI (community, on-treatment or off)
  m.Trace[1, ] <- 0
  if (strat == "soc") {
    m.Trace[1, "mciof_c"] <- p_starting_state_mci
    m.Trace[1, "milof_c"] <- 1 - p_starting_state_mci
  } else {
    m.Trace[1, "mcion_c"] <- p_starting_state_mci
    m.Trace[1, "milon_c"] <- 1 - p_starting_state_mci
  }
  
  # Define utility and cost vectors
  v.utility <- c(
    "mcion_c" = u_mci_pt + u_mci_ic,
    "mciof_c" = u_mci_pt + u_mci_ic,
    "milon_c" = u_mil_pt + u_mil_ic,
    "milof_c" = u_mil_pt + u_mil_ic,
    "mod_c"   = u_mod_pt + u_mod_ic,
    "sev_c"   = u_sev_pt + u_sev_ic,
    "mci_i"   = u_mci_pt_i + u_mci_ic_i,
    "mil_i"   = u_mil_pt_i + u_mil_ic_i,
    "mod_i"   = u_mod_pt_i + u_mod_ic_i,
    "sev_i"   = u_sev_pt_i + u_sev_ic_i,
    "dth"     = 0
  )
  
  v.cost <- c(
    "mcion_c" = c_mci_hc + c_mci_sc + c_mci_ic,
    "mciof_c" = c_mci_hc + c_mci_sc + c_mci_ic,
    "milon_c" = c_mil_hc + c_mil_sc + c_mil_ic,
    "milof_c" = c_mil_hc + c_mil_sc + c_mil_ic,
    "mod_c"   = c_mod_hc + c_mod_sc + c_mod_ic,
    "sev_c"   = c_sev_hc + c_sev_sc + c_sev_ic,
    "mci_i"   = c_mci_i_hc + c_mci_i_sc + c_mci_i_ic,
    "mil_i"   = c_mil_i_hc + c_mil_i_sc + c_mil_i_ic,
    "mod_i"   = c_mod_i_hc + c_mod_i_sc + c_mod_i_ic,
    "sev_i"   = c_sev_i_hc + c_sev_i_sc + c_sev_i_ic,
    "dth"     = 0
  )
  
  # Identify on-treatment states (for recurring treatment costs in "int")
  on_tx_states <- c("mcion_c", "milon_c")
  
  # Initialize vectors for outcomes
  v.QALYs <- numeric(n_cycles + 1)
  v.Costs <- numeric(n_cycles + 1)
  v.LYs   <- numeric(n_cycles + 1)
  
  # Loop through cycles to update trace and calculate outcomes
  for (t in 1:n_cycles) {
    m.Trace[t + 1, ] <- m.Trace[t, ] %*% a.TP[ , , t]
    
    # QALYs and Costs per state
    v.QALYs[t] <- sum(m.Trace[t, ] * v.utility)
    v.Costs[t] <- sum(m.Trace[t, ] * v.cost)
    v.LYs[t]   <- sum(m.Trace[t, state_names != "dth"])
    
    # Add recurring treatment costs for intervention strategy
    if (strat == "int") {
      v.Costs[t] <- v.Costs[t] + sum(m.Trace[t, on_tx_states]) * c_Tx
    }
    
    # Add one-time treatment start cost and disutility (only at cycle 1)
    if (strat == "int" && t == 1) {
      v.Costs[t] <- v.Costs[t] + c_Tx_start
      v.QALYs[t] <- v.QALYs[t] + u_Tx_start
    }
  }
  
  # Half-cycle correction: average adjacent cycles and drop last
  v.QALYs <- (v.QALYs[-length(v.QALYs)] + v.QALYs[-1]) / 2
  v.Costs <- (v.Costs[-length(v.Costs)] + v.Costs[-1]) / 2
  v.LYs   <- (v.LYs[-length(v.LYs)] + v.LYs[-1]) / 2
  
  # Apply discounting
  v.discount_weights <- 1 / ((1 + discount_QALY) ^ (1:n_cycles))
  QALYs_total <- sum(v.QALYs * v.discount_weights)
  Costs_total <- sum(v.Costs * v.discount_weights)
  LYs_total   <- sum(v.LYs   * v.discount_weights)
  
  return(list(QALYs = QALYs_total, Costs = Costs_total, LYs = LYs_total))
}

# -----------------------------
# Run Model and Print Results
# -----------------------------

res_soc <- compute_outcomes("soc")
res_int <- compute_outcomes("int")

cat("Comparative Results:\n")
cat("Strategy        | QALYs  | Costs (USD) | Life Years\n")
cat("----------------------------------------------------\n")
cat(sprintf("Standard of Care| %6.2f | %12.0f | %10.2f\n",
            res_soc$QALYs, res_soc$Costs, res_soc$LYs))
cat(sprintf("Intervention    | %6.2f | %12.0f | %10.2f\n",
            res_int$QALYs, res_int$Costs, res_int$LYs))
cat("----------------------------------------------------\n")
cat(sprintf("Incremental     | %+6.2f | %+12.0f | %+10.2f\n",
            res_int$QALYs - res_soc$QALYs,
            res_int$Costs - res_soc$Costs,
            res_int$LYs   - res_soc$LYs))
```
