## Supplementary material for "Replicating Health-Economic Simulation Models for Alzheimer’s Disease Using Artificial Intelligence": final R scripts: FinalPrompts_IPECAD.pdf

**Attached Documents** | **Sequence of Prompts to replicate the IPECAD Model** | **Validation Criteria**

### Prompt 0 – General Instructions and Workflow Overview

*"Do not generate any output in response to this prompt. This message is only to inform you that, in this discussion, you will receive a sequence of 6 prompts aimed at sequentially generating all components of a health economic Markov model in R. The objective of the upcoming prompts will be:*
*1. Load and define all input parameters from a publication (shared later).*
*2. Load and process mortality rate data from a life table CSV file.*
*3. Generate all time-dependent transition probability vectors for the standard of care ("soc") strategy, as well as mortality and discontinuation.*
*4. Generate all time-dependent transition probability vectors for the intervention ("int") strategy.*
*5. Build the transition probability matrix using structured logic sections.*
*6. Compute the Markov trace and total outcomes (QALYs, LYs, and Costs).*
*Global instructions to follow in all prompts:*
*- Use only the 11 fixed state names (they will be provided with the first prompt).*
*- Use consistent and explicit variable names across prompts; do not invent or abbreviate names.*
*- Always reuse the exact same variable names defined in earlier prompts.*
*- Never repeat code that was defined in a previous prompt unless explicitly asked.*
*- Return only a fully functional and copy-paste-ready R code block.*
*- Wait for the first actual prompt before generating any output."*

### Prompt 1 – Load and Define Model Parameters

Attached documents: IPECAD main publication PDF; IPECAD Supplemental material PDF

*"(Prompt 1): Please extract all input parameters required to code the IPECAD Markov model using the dampack package, based on the content of the IPECAD publication and its supplemental material. Include all variables, including descriptive ones (e.g., state names and strategy names) directly as named R objects in the script. Restrict state names to the 11 that are actually used in the publication ("mcion_c","mciof_c","milon_c","milof_c","mod_c","sev_c","mci_i","mil_i","mod_i","sev_i","dth"). Do not omit any parameter, even if its value is zero. If a parameter is derived from a formula, preserve the original formula in the R script rather than just showing the final numeric value. Display the full R script directly in the response (do not put it in a downloadable file)."*

Validation criteria:
- All parameters (including zero) are extracted from Table S1 of the Supplementary Material.
- Formulas are preserved in code.

### Prompt 2 – Create Age-Specific Mortality Rates Table

Attached document: lifetable_US_2019_ssa.csv

*"(Prompt 2): Please write a R code that loads the 2019 US life table from the CSV file named "lifetable_US_2019_ssa.csv". The CSV contains the following columns: "age", "male", and "female", which represent age-specific annual mortality probabilities for males and females, respectively. First, convert these annual mortality probabilities to annual mortality rates. Then, use these columns to compute a weighted average of these mortality rates and place them into a new column "weighted". Return a matrix or dataframe with columns: "age", "male", "female", and "weighted", where "male" and "female" contain the converted rates (not the original probabilities). Do not add any other packages. Assume the file is in the working directory."*

Validation criteria:
- Mortality probabilities are correctly converted to rates.
- "*weighted*" column is correctly computed as weighted average.
- Output includes columns: age, male, female, weighted.

### Prompt 3 – Generate Transition Vectors for Standard of Care (SOC)

*"(Prompt 3): Please write a R code that defines all time-dependent transition probability vectors for the standard of care ("soc") strategy, without using assign() or get(), or dynamic variable names. Define the following vectors:*
*- Transition probability vectors per cycle for disease progression under the "soc" strategy ("v.p.mci_mil_soc", "v.p.mci_mod_soc", "v.p.mci_sev_soc", etc.)*
*- Staying probabilities per state as 1 minus the sum of other disease progression probabilities only (exclude institutional transitions). Survival adjustment will be applied later.*
*- Time-dependent vectors for transitions from each community state to institutionalized settings.*
*- A discontinuation vector per cycle, with the first discontinuation probability value for the first time interval and the second discontinuation probability from the second time point until the maximum treatment duration. After reaching the maximum treatment duration, the value should be set to 1. Each cycle's value must be treated independently (no cumulative calculation).*
*- A vector of age- and sex-specific mortality rates using the predefined mortality matrix."*

Validation criteria:
- One named vector per transition, no *assign()/get()*.
- All vectors are correctly defined.
- Stay probabilities are computed as *1 - sum(other transitions)*.
- Discontinuation vector uses correct fixed values for each of the three phases.
- Mortality vector is correctly extracted per age and cycle.

### Prompt 4 – Generate Transition Vectors for Intervention (INT)

*"(Prompt 4): Please write a R code that defines all time-dependent transition probability vectors for the intervention ("int") strategy of the IPECAD model. Use only explicit variable names.*
*For each cycle, create vectors for:*
*- On-treatment transition vectors from community states "mcion_c" and "milon_c".*
*- Post-discontinuation transitions from off-treatment states "mciof_c" and "milof_c".*
*- For each transition:*
  *- Convert probabilities to rates: `-log(1 - p)`*
  *- Multiply rate by the relative risk (RR or RR_dis), raised to the power of `(1 - waning)^(cycle_index - 1)`*
  *- Convert back to probability: `1 - exp(-rate)`*
  *- For backward transitions (e.g., mild → mci), no treatment effect should be applied (just use original probabilities).*
*- Also create staying probabilities per state for both phases, defined as: `1 - (sum of other disease progression transitions)` (excluding institutional transitions)."*

Validation criteria:
- Treatment effect (*RR* or *RR_dis*) is applied using correct waning formula.
- No treatment effect applied for backward transitions.
- Probabilities converted: p → rate → adjusted rate → p.
- Stay probabilities recomputed per cycle and phase.

### Prompt 5: Build Transition Probability Matrix (TPM)

#### Prompt 5A: Initialize TPM Structure with Placeholders

*"(Prompt 5A): Please write a R code that initializes the transition probability array named a.TP (dimensions: states × states × cycles) for the IPECAD model. The function must be structured as build_transition_matrix(strat), where strat is either "soc" or "int". Inside the function:*
*- Initialize the array a.TP and loop over each cycle*
*- For each cycle, explicitly assign the transition probabilities from each state to death without using loops (Write one assignment line explicitly for each state).*
*- For each state, compute the transition probability to death using the formula: 1 - exp(-rate), where rate = v.mortality_rate[t] * HR and HR is the appropriate hazard ratio for that state.*
*- For the death state "dth", set the probability of remaining in this state to 1 at all cycles, since death is an absorbing state.*
*- Leave three clearly visible placeholders in the function body:*
  *[INSERT PROMPT 5B HERE] — to define the transitions from on-treatment community states later.*
  *[INSERT PROMPT 5C HERE] — to define the transitions from off-treatment, moderate/severe community states later.*
  *[INSERT PROMPT 5D HERE] — to define the transitions from institutionalized states later.*
*- After filling the transitions for each state at each cycle (5B, 5C and 5D), normalize the rows if necessary: if the row sum differs from 1, divide each element of the row by the row sum.*
*Do not yet define transitions for community or institutionalized states here — only set up the structure and placeholders."*

**Validation Criteria Prompt 5A**
- *a.TP* array correctly initialized with dimensions and names.
- Loop over cycles implemented.
- All transitions to death are computed using: *1 - exp(-rate)*.
- "*dth*" state set as absorbing.
- Placeholder comments for 5B, 5C, 5D are clearly included.
- Row sums are normalized if needed at the end of each cycle.

#### Prompt 5B: Define Transitions from On-Treatment Community States

*"(Prompt 5B): Please extend the previously initialized build_transition_matrix(strat) function by filling all transitions from the community states on-treatment ("mcion_c" and "milon_c") for each cycle. Use only explicit variable names without using assign() or get(), or dynamic variable names.*
*- Use only valid 11 states and pre-defined time-dependent vectors (*_soc for "soc", no suffix for "int").*
*- For each valid destination state (toward community, institutionalized and staying), assign probabilities by multiplying:*
  *- the appropriate transition probability vector,*
  *- (1 - mortality probability) for all living state transitions,*
  *- (1 - p.institution) or p.institution as appropriate,*
  *- and (1 - p.discontinuation) from on-treatment to on-treatment (e.g. from "mcion_c" to "milon_c" or "mcion_c", etc.) or p.discontinuation from on-treatment to off-treatment. However, do not apply the discontinuation factor for transitions toward "mod_c", "sev_c", and institutionalized states.*
*- Even though "mcion_c" and "milon_c" represent on-treatment states, define their transitions in "soc" as well to ensure consistency across strategies.*
*Do not repeat the initialization part of the function. Return only the R code for these transitions."*

**Validation Criteria Prompt 5B**
- All transitions from *"mcion_c"* and *"milon_c"* are explicitly defined, no *assign()/get()*.
- Discontinuation only affects transitions to *"mciof_c"* and *"milof_c"*.
- All living transitions are adjusted by *(1 - mortality)*.
- Institutional transitions use correct *(1 - p.institution)* or *p.institution*.
- No return from off-treatment to on-treatment (except *"mcion_c"* → *"milon_c"*).

#### Prompt 5C: Define Transitions from Off-Treatment and Moderate/Severe States

*"(Prompt 5C): Please extend the previously initialized build_transition_matrix(strat) function by filling all transitions from the community states off-treatment and moderate/severe ("mciof_c", "milof_c", "mod_c", "sev_c") for each cycle.*
*- Assign transition probabilities by multiplying:*
  *- the appropriate transition probability vector,*
  *- (1 - mortality probability) for all living state transitions,*
  *- and (1 - p.institution) or p.institution as appropriate.*
*- For "soc", use *_soc vectors.*
*- For "int", always use the post-discontinuation vectors for these states (no discontinuation factor applies for these states).*
*- No transitions back to on-treatment states are allowed."*

**Validation Criteria Prompt 5C**
- All transitions from *"mciof_c"*, *"milof_c"*, *"mod_c"* and *"sev_c"* are explicitly defined, no *assign()/get()*.
- All living transitions are adjusted by *(1 - mortality)*.
- Institutional transitions use correct *(1 - p.institution)* or *p.institution*.
- No return from off-treatment to on-treatment.

#### Prompt 5D: Define Transitions from Institutionalized States

*"(Prompt 5D): Please extend the build_transition_matrix(strat) function by filling all transitions for the institutionalized states: "mci_i", "mil_i", "mod_i", and "sev_i". For each institutionalized state at each cycle, define transitions toward other institutionalized states (including remaining in the same state) using only time-dependent vectors previously defined. For mci_i state forward transitions are allowed, such as to mil_i, mod_i and sev_i."*

**Validation Criteria Prompt 5D**
- All transitions from *"mci_i"*, *"mil_i"*, *"mod_i"* and *"sev_i"* are explicitly defined, no *assign()/get()*.
- Only institutional states are used as destination states.
- All living transitions are adjusted by *(1 - mortality)*.
- No treatment effect (*RR/waning*).

**Global validation for Prompt 5 (5A + 5B + 5C)**
- Implemented R script to compare generated vs. IPECAD TPMs per cycle and transition.
- Calculated absolute differences (state × state × cycle).
- Flagged any discrepancy exceeding 10% threshold.

### Prompt 6: Simulate Markov Model and Compute Outcomes

*"(Prompt 6): Please generate a fully functional and copy-paste-ready R code block that initializes and computes the Markov trace matrix and total outcomes (QUALYs and Costs) for a given strategy ("soc" or "int") over all cycles. Use predefined state names to define named vectors for state occupancy, utilities, and costs to ensure safe indexing. Start by setting the initial state distribution. At each cycle, update the state distribution using Markov multiplication with the corresponding cycle's transition probability matrix. Calculates primary outcomes (QALYs and costs) by multiplying named state occupancy at each cycle with utility and cost vectors, and storing them in the output matrix. Apply half-cycle correction by averaging each cycle's outcomes with the next cycle's values, then remove the last cycle from all outcome vectors. For the "int" strategy, apply recurring treatment costs to on-treatment states every cycle. Apply the initial treatment cost and QUALY gain (c.Tx_start and u.Tx_start) only once, during the first cycle. Apply discounting to both QALYs and costs. Calculates total outcomes across all cycles, including life-years, total QALYs and total costs. Return only the total (aggregate) QALYs, costs and LYs across all cycles. At the end, write a script that prints the comparative results clearly."*

Validation criteria:
- Implemented R script to compare final outcomes (QALYs, Costs, LYs) for each strategy (*"soc"* and *"int"*) vs. IPECAD reference.
- Calculated absolute and relative differences for each outcome.
- Flagged discrepancies exceeding pre-defined thresholds: QALYs/LYs >5%, Costs >10%.
