## Supplementary material for "Replicating Health-Economic Simulation Models for Alzheimer’s Disease Using Artificial Intelligence": final R scripts: validation15Trials_IPECAD.R

```
##### Script: Compare Replicated Model Results to IPECAD Reference #####

# Clear console and environment
cat("\014")
rm(list = ls())

# Load required libraries
library(knitr)
library(ggplot2)
library(tidyr)
library(dplyr)
library(extrafont)

# Reference values from the original IPECAD model
IPECAD_soc <- c(QALYs = 2.94, Costs = 484963.13, LYs = 6.15)
IPECAD_int <- c(QALYs = 3.29, Costs = 580333.59, LYs = 6.50)

# List of results from 15 replications
results_list <- list(
  list(soc = c(QALYs = 2.85, Costs = 470838, LYs = 5.98), int = c(QALYs = 3.19, Costs = 562842, LYs = 6.31)), #O.R
  list(soc = c(QALYs = 2.86, Costs = 465902.80, LYs = 5.98), int = c(QALYs = 3.18, Costs = 552612.39, LYs = 6.29)), #O2.R
  list(soc = c(QALYs = 3.50, Costs = 444403, LYs = 6.60), int = c(QALYs = 3.73, Costs = 512352, LYs = 6.83)), #O3.R
  list(soc = c(QALYs = 3.323684e+00, Costs = 4.830820e+05, LYs = 1.926411e+01), int = c(QALYs = 3.635874e+00, Costs = 5.696718e+05, LYs = 1.926411e+01)), #O4.R
  list(soc = c(QALYs = 2.95, Costs = 475542.45, LYs = 6.15), int = c(QALYs = 3.20, Costs = 555304.24, LYs = 6.39)), #O5.R
  list(soc = c(QALYs = 2.90, Costs = 475729, LYs = 6.03), int = c(QALYs = 3.23, Costs = 563876, LYs = 6.34)), #O6.R
  list(soc = c(QALYs = 3.53, Costs = 454383, LYs = 6.65), int = c(QALYs = 3.12, Costs = 565084, LYs = 6.24)), #O7.R
  list(soc = c(QALYs = 3.77, Costs = 453179, LYs = 7.01), int = c(QALYs = 4.00, Costs = 528637, LYs = 7.23)), #O8.R
  list(soc = c(QALYs = 3.860, Costs = 453332.91, LYs = 6.624), int = c(QALYs = 4.075, Costs = 519073.69, LYs = 6.841)), #O9.R
  list(soc = c(QALYs = 3.492, Costs = 448098, LYs = 6.602), int = c(QALYs = 3.722, Costs = 516120.1, LYs = 6.83)), #O10.R
  list(soc = c(QALYs = 3.35, Costs = 492280.09, LYs = 19.55), int = c(QALYs = 3.62, Costs = 580953.37, LYs = 19.55)), #O11.R
  list(soc = c(QALYs = 4.02, Costs = 465847, LYs = 6.87), int = c(QALYs = 4.24, Costs = 532458, LYs = 7.09)), #O12.R
  list(soc = c(QALYs = 2.79, Costs = 412237.81, LYs = 5.71), int = c(QALYs = 3.56, Costs = 500659.46, LYs = 6.50)), #O13.R
  list(soc = c(QALYs = 3.38, Costs = 490543, LYs = 6.22), int = c(QALYs = 3.61, Costs = 581521, LYs = 6.45)), #O14.R
  list(soc = c(QALYs = 2.86, Costs = 465903 , LYs = 5.98), int = c(QALYs = 2.86 , Costs = 466492, LYs = 5.98)) #O15.R
)


# Initialize results dataframe
detailed_results <- data.frame()

# Loop through each simulation
for (i in seq_along(results_list)) {
  res <- results_list[[i]]
  
  soc <- res$soc
  int <- res$int
  
  # Absolute differences
  abs_soc <- abs(soc - IPECAD_soc)
  abs_int <- abs(int - IPECAD_int)
  
  # Relative differences (in %)
  rel_soc <- 100 * abs((soc - IPECAD_soc) / IPECAD_soc)
  rel_int <- 100 * abs((int - IPECAD_int) / IPECAD_int)
  
  # Combine into one row
  row <- data.frame(
    Sim = paste0("Sim_", i),
    QALYs_SOC = soc["QALYs"], QALYs_INT = int["QALYs"],
    AbsDiff_Q_SOC = abs_soc["QALYs"], AbsDiff_Q_INT = abs_int["QALYs"],
    RelDiff_Q_SOC = rel_soc["QALYs"], RelDiff_Q_INT = rel_int["QALYs"],
    
    Costs_SOC = soc["Costs"], Costs_INT = int["Costs"],
    AbsDiff_C_SOC = abs_soc["Costs"], AbsDiff_C_INT = abs_int["Costs"],
    RelDiff_C_SOC = rel_soc["Costs"], RelDiff_C_INT = rel_int["Costs"],
    
    LYs_SOC = soc["LYs"], LYs_INT = int["LYs"],
    AbsDiff_LY_SOC = abs_soc["LYs"], AbsDiff_LY_INT = abs_int["LYs"],
    RelDiff_LY_SOC = rel_soc["LYs"], RelDiff_LY_INT = rel_int["LYs"]
  )
  
  detailed_results <- rbind(detailed_results, row)
}


# Compute MAD and SD summary statistics
summary_table <- data.frame(
  Metric = c("MAD", "SD"),
  QALYs_SOC = c(mad(detailed_results$QALYs_SOC, constant = 1), sd(detailed_results$QALYs_SOC)),
  QALYs_INT = c(mad(detailed_results$QALYs_INT, constant = 1), sd(detailed_results$QALYs_INT)),
  Costs_SOC = c(mad(detailed_results$Costs_SOC, constant = 1), sd(detailed_results$Costs_SOC)),
  Costs_INT = c(mad(detailed_results$Costs_INT, constant = 1), sd(detailed_results$Costs_INT)),
  LYs_SOC = c(mad(detailed_results$LYs_SOC, constant = 1), sd(detailed_results$LYs_SOC)),
  LYs_INT = c(mad(detailed_results$LYs_INT, constant = 1), sd(detailed_results$LYs_INT))
)

# Display table
print(kable(summary_table, digits = 3, caption = "Summary of Differences vs. IPECAD (MAD and SD)"))

# View all detailed results
View(detailed_results, title = "Detailed simulation results vs. IPECAD")


##### ────────────────────────────────────────────────────────────
#####  Box-plots: distribution, mean, median, SD & MAD
##### ────────────────────────────────────────────────────────────

# Reshape to long format
long_values <- detailed_results %>%
  select(Sim, QALYs_SOC, QALYs_INT, Costs_SOC, Costs_INT, LYs_SOC, LYs_INT) %>%
  pivot_longer(-Sim, names_to = "Metric_Strategy", values_to = "Value") %>%
  separate(Metric_Strategy, into = c("Metric", "Strategy"), sep = "_")

# Rename strategies
long_values$Strategy <- recode(long_values$Strategy,
                               SOC = "Standard of Care",
                               INT = "Intervention")

# Colour palette
custom_cols <- c("Standard of Care" = "#ffbd59",
                 "Intervention"    = "#b9d8de")

# Loop through QALYs | Costs | LYs
for (metric in unique(long_values$Metric)) {
  
  df_metric <- filter(long_values, Metric == metric)
  
  
  # Summary stats per strategy
  df_sum <- df_metric %>%
    group_by(Strategy) %>%
    summarise(mean   = mean(Value),
              median = median(Value),
              sd     = sd(Value),
              mad    = mad(Value, constant = 1),
              .groups = "drop")
  
  
  # Data for mean/median horizontal ticks
  df_lines <- df_sum %>%
    pivot_longer(c(mean, median), names_to = "LineType", values_to = "Y") %>%
    mutate(x_pos = as.numeric(factor(Strategy,
                                     levels = c("Intervention",
                                                "Standard of Care"))),
           LineType = recode(LineType, mean = "Mean", median = "Median"))
  
  # Box-plot
  p <- ggplot(df_metric, aes(Strategy, Value, fill = Strategy)) +
    geom_boxplot(width = 0.4, alpha = 0.6, outlier.shape = NA) +
    geom_segment(data = df_lines,
                 aes(x = x_pos - 0.2, xend = x_pos + 0.2,
                     y = Y, yend = Y,
                     linetype = LineType, colour = LineType),
                 linewidth = 0.8, inherit.aes = FALSE) +
    geom_text(data = df_sum,
              aes(x = Strategy, y = max(df_metric$Value) * 1.02,
                  label = paste0("SD = ", round(sd, 2),
                                 "\nMAD = ", round(mad, 2))),
              size = 3, family = "Verdana") +
    scale_fill_manual(values = custom_cols, name = "Strategy") +
    scale_colour_manual(values = c(Mean = "red", Median = "black"),
                        name = "Measure") +
    scale_linetype_manual(values = c(Mean = "solid", Median = "solid"),
                          name   = "Measure") +
    labs(title = paste("Distribution of", metric,
                       "by Strategy  (n =", nrow(df_metric) / 2, ")"),
         x = NULL, y = metric) +
    theme_minimal(base_family = "Verdana") +
    theme(plot.title   = element_text(size = 10, face = "bold", hjust = 0.5),
          legend.title = element_text(face = "bold"),
          legend.position = "right")
  
  print(p)
}


##### ────────────────────────────────────────────────────────────
#####  Optional: Bar-plots of raw values vs. reference
##### ────────────────────────────────────────────────────────────
if (TRUE) {
  
  # Add IPECAD reference row
  ref_row <- data.frame(
    Sim          = "IPECAD",
    QALYs_SOC    = IPECAD_soc["QALYs"],
    QALYs_INT    = IPECAD_int["QALYs"],
    Costs_SOC    = IPECAD_soc["Costs"],
    Costs_INT    = IPECAD_int["Costs"],
    LYs_SOC      = IPECAD_soc["LYs"],
    LYs_INT      = IPECAD_int["LYs"]
  )
  
  detailed_vals <- detailed_results %>%
    select(Sim, QALYs_SOC, QALYs_INT, Costs_SOC, Costs_INT, LYs_SOC, LYs_INT)
  
  detailed_vals <- rbind(ref_row, detailed_vals) %>%
    mutate(SimID = factor(Sim, levels = c("IPECAD", paste0("Sim_", 1:15))))
  
  # Long format
  plot_data <- detailed_vals %>%
    pivot_longer(-SimID, names_to = "Metric_Strategy", values_to = "Value") %>%
    separate(Metric_Strategy, into = c("Metric", "Strategy"), sep = "_") %>%
    mutate(Strategy  = ifelse(Strategy == "SOC", "Standard of Care", "Intervention"),
           ref_flag  = ifelse(SimID == "IPECAD", "ref", "sim"),
           fill_grp  = interaction(Strategy, ref_flag, sep = "_"))
  
  # Colours
  strat_cols <- c("Standard of Care_sim"  = "#ffbd59",
                  "Intervention_sim"      = "#b9d8de",
                  "Standard of Care_ref"  = "#cc8400",
                  "Intervention_ref"      = "#6396a6")
  
  for (m in c("QALYs", "Costs", "LYs")) {
    p <- ggplot(filter(plot_data, Metric == m),
                aes(SimID, Value, fill = fill_grp)) +
      geom_bar(stat = "identity", position = "dodge") +
      scale_fill_manual(values = strat_cols,
                        name   = "Strategy",
                        labels = c("Standard of Care_sim" = "Standard of Care",
                                   "Intervention_sim"     = "Intervention",
                                   "Standard of Care_ref" = "Standard of Care (IPECAD)",
                                   "Intervention_ref"     = "Intervention (IPECAD)")) +
      labs(title = paste("Replicated", m, "by Simulation"),
           x = "Simulation", y = m) +
      theme_minimal(base_family = "Verdana") +
      theme(plot.title = element_text(face = "bold", hjust = 0.5),
            axis.text.x = element_text(angle = 45, hjust = 1))
    
    print(p)
  }
}

##### ────────────────────────────────────────────────────────────
#####  Optional: MAD ± SD bar-plot (set if(FALSE) to skip)
##### ────────────────────────────────────────────────────────────
if (FALSE) {
  
  # Exclude reference row
  detailed_no_ref <- filter(detailed_results, Sim != "IPECAD")
  
  mad_sd <- data.frame(
    Metric   = rep(c("QALYs", "Costs", "LYs"), each = 2),
    Strategy = rep(c("SOC", "INT"), 3),
    MAD = c(mean(detailed_no_ref$AbsDiff_Q_SOC),
            mean(detailed_no_ref$AbsDiff_Q_INT),
            mean(detailed_no_ref$AbsDiff_C_SOC),
            mean(detailed_no_ref$AbsDiff_C_INT),
            mean(detailed_no_ref$AbsDiff_LY_SOC),
            mean(detailed_no_ref$AbsDiff_LY_INT)),
    SD  = c(sd(detailed_no_ref$AbsDiff_Q_SOC),
            sd(detailed_no_ref$AbsDiff_Q_INT),
            sd(detailed_no_ref$AbsDiff_C_SOC),
            sd(detailed_no_ref$AbsDiff_C_INT),
            sd(detailed_no_ref$AbsDiff_LY_SOC),
            sd(detailed_no_ref$AbsDiff_LY_INT))
  )
  
  ggplot(mad_sd, aes(Strategy, MAD, fill = Strategy)) +
    geom_bar(stat = "identity", width = 0.6) +
    geom_errorbar(aes(ymin = MAD - SD, ymax = MAD + SD), width = 0.2) +
    facet_wrap(~ Metric, scales = "free_y") +
    scale_fill_manual(values = c(SOC = "#ffbd59", INT = "#b9d8de")) +
    labs(title = "MAD ± SD of Absolute Differences vs. IPECAD",
         y = "MAD (± SD)", x = NULL) +
    theme_minimal(base_family = "Times New Roman") +
    theme(plot.title = element_text(face = "bold", hjust = 0.5),
          strip.text  = element_text(face = "bold"),
          legend.position = "none")
}
```
