## Supplementary material for "Replicating Health-Economic Simulation Models for Alzheimer’s Disease Using Artificial Intelligence": final R scripts: validationPrompt4.R

```
##### Code to compare the obtained matrices from the prompt 4 to the Original IPECAD matrices #####


cat("\014") # clear console
rm(list = ls()) # clear environment


# Load the original IPECAD matrices for both strategies
source("IPECAD_modelOnly.R")
IPECAD_TP_soc <- l.out_icer[["l.out"]][["soc"]][["a.TP"]]
IPECAD_TP_int <- l.out_icer[["l.out"]][["int"]][["a.TP"]]


# Load the prompt 4 generated matrices for both strategies
rm(list = setdiff(ls(), c("IPECAD_TP_soc", "IPECAD_TP_int"))) # To clean the environment except IPECAD matrices
source("o15.R")
TP_soc <- build_transition_matrix("soc")
TP_int <- build_transition_matrix("int")


# Parameters
validation_threshold = 0.1 # = 10% of absolute error
n_cycles = 29
state_names <- dimnames(IPECAD_TP_soc)[[1]]


# Fonction that identifies absolute errors above the threshold
validate_transition_matrix <- function(original_matrix, new_matrix, threshold) {
  exceedances <- list()

  for (cycle in 1:n_cycles) {
    for (from_state in state_names) {
      for (to_state in state_names) {
        original_value <- original_matrix[from_state, to_state, cycle]
        new_value <- new_matrix[from_state, to_state, cycle]
        abs_error <- abs(original_value - new_value)
        
        if (abs_error > threshold) {
          exceedances[[length(exceedances) + 1]] <- list(
            cycle = cycle,
            from = from_state,
            to = to_state,
            original = round(original_value, 4),
            new = round(new_value, 4),
            abs_error = round(abs_error, 4)
          )
        }
      }
    }
  }
  
  if (length(exceedances) == 0) {
    cat("Validation passed: all transitions are within the absolute error threshold of", threshold, "\n")
  } else {
    cat("Validation failed: some transitions exceed the error threshold (", threshold, ").\n", sep = "")
    cat("Details of the differences:\n")
    df <- do.call(rbind, lapply(exceedances, as.data.frame))
    print(df)
  }
}


# Apply the previous fonction to validate both strategies
cat("\n--- Checking SOC strategy ---\n")
validate_transition_matrix(IPECAD_TP_soc, TP_soc, validation_threshold)

cat("\n--- Checking INT strategy ---\n")
validate_transition_matrix(IPECAD_TP_int, TP_int, validation_threshold)
```
