## Supplementary material for "Replicating Health-Economic Simulation Models for Alzheimer’s Disease Using Artificial Intelligence": final R scripts: Wimo_O1.R

```
# Load input parameters for the Markov model from Wimo et al. (2020)

# Descriptive Parameters
state_names <- c("ad_mci", "mild", "moderate", "severe", "death")
strategy_names <- c("no_intervention", "intervention")
cycle_length <- 1       # years
n_cycles <- 40          # 40-year simulation
start_age <- 60
discount_rate <- 0.03
cohort_size <- 100000

# Transition Probabilities (Base Case Estimates)
# Note: These will be used in the matrix logic later
p_ad_mci_to_mild <- 0.206  # Annualized from 3-year 50% risk: 1 - (1 - 0.50)^(1/3)
p_ad_mci_to_death <- NA    # To be derived from life table in next step

p_mild_to_mild <- 0.706
p_mild_to_moderate <- 0.293
p_mild_to_severe <- 0.001

p_moderate_to_mild <- 0.087
p_moderate_to_moderate <- 0.803
p_moderate_to_severe <- 0.109

p_severe_to_moderate <- 0.196
p_severe_to_severe <- 0.804
p_severe_to_death <- NA    # Age-specific mortality will be added later

# Hazard Ratios for Mortality by State (Relative to AD-MCI/Very Mild)
hr_mild <- 1.318
hr_moderate <- 2.419
hr_severe <- 4.267

# Base Case Hypothetical Intervention Effects
rr_ad_mci_to_mild <- 0.75     # 25% reduction in progression
rr_mild_to_moderate <- 0.75   # 25% reduction in progression
dmt_treatment_cost <- 50000   # SEK/year

# QALY Utility Functions
u_ad_mci <- function(age) {
  -0.002464 * age + 0.962679
}

u_mild <- function(age) {
  -0.0025 * age + 0.8293
}

u_moderate <- function(age) {
  -4e-07 * age^2 - 0.0024 * age + 0.5828
}

u_severe <- function(age) {
  -4e-07 * age^2 - 0.0025 * age + 0.4337
}

u_death <- function(age) {
  0
}


# Load mortality rates and weights
mortality_rates <- read.csv("mortalityRates_Swedish_2016.csv", stringsAsFactors = FALSE)
weights <- read.csv("weightsMortalityRates_Swedish_2016.csv", stringsAsFactors = FALSE)

# Define age ranges corresponding to each group
age_group_map <- list(
  "60-64" = 60:64,
  "65-69" = 65:69,
  "70-74" = 70:74,
  "75-79" = 75:79,
  "80-84" = 80:84,
  "85-89" = 85:89,
  "90+"   = 90:99
)

# Expand grouped mortality rates to individual ages
expanded_rates <- data.frame(age = 60:99, male = NA, female = NA)

for (group in names(age_group_map)) {
  ages <- age_group_map[[group]]
  male_rate <- mortality_rates$male[mortality_rates$age == group]
  female_rate <- mortality_rates$female[mortality_rates$age == group]
  expanded_rates$male[expanded_rates$age %in% ages] <- male_rate
  expanded_rates$female[expanded_rates$age %in% ages] <- female_rate
}

# Merge with weights
merged_data <- merge(expanded_rates, weights, by = "age")

# Compute sex-weighted average mortality rate
merged_data$rate_avg <- with(merged_data, 
                             (male * weight_male + female * weight_female) / (weight_male + weight_female)
)

# Use hazard ratios from previously defined variables
# hr_mild, hr_moderate, hr_severe (defined in Prompt 1)

# Compute probabilities of death for each state
merged_data$prob_death_ad_mci <- 1 - exp(-merged_data$rate_avg)
merged_data$prob_death_mild <- 1 - exp(-merged_data$rate_avg * hr_mild)
merged_data$prob_death_moderate <- 1 - exp(-merged_data$rate_avg * hr_moderate)
merged_data$prob_death_severe <- 1 - exp(-merged_data$rate_avg * hr_severe)

# Final data frame
death_probs <- merged_data[, c("age", "rate_avg", "prob_death_ad_mci",
                               "prob_death_mild", "prob_death_moderate",
                               "prob_death_severe")]

# Ensure column types
death_probs$age <- as.integer(death_probs$age)
# Time-dependent transition probabilities for no intervention strategy
p_ad_mci_to_mild_no_int <- rep(p_ad_mci_to_mild, n_cycles)
p_ad_mci_stay_no_int <- 1 - p_ad_mci_to_mild_no_int

p_mild_to_moderate_no_int <- rep(p_mild_to_moderate, n_cycles)
p_mild_to_severe_no_int <- rep(p_mild_to_severe, n_cycles)
p_mild_stay_no_int <- 1 - (p_mild_to_moderate_no_int + p_mild_to_severe_no_int)

p_moderate_to_mild_no_int <- rep(p_moderate_to_mild, n_cycles)
p_moderate_to_severe_no_int <- rep(p_moderate_to_severe, n_cycles)
p_moderate_stay_no_int <- 1 - (p_moderate_to_mild_no_int + p_moderate_to_severe_no_int)

p_severe_to_moderate_no_int <- rep(p_severe_to_moderate, n_cycles)
p_severe_stay_no_int <- 1 - p_severe_to_moderate_no_int

# Time-dependent transition probabilities for intervention strategy (DMT applied in ad_mci and mild)
# Adjust AD-MCI to mild
rate_ad_mci_to_mild <- -log(1 - p_ad_mci_to_mild)
rate_ad_mci_to_mild_adj <- rate_ad_mci_to_mild * rr_ad_mci_to_mild
p_ad_mci_to_mild_int <- rep(1 - exp(-rate_ad_mci_to_mild_adj), n_cycles)
p_ad_mci_stay_int <- 1 - p_ad_mci_to_mild_int

# Adjust Mild to Moderate
rate_mild_to_moderate <- -log(1 - p_mild_to_moderate)
rate_mild_to_moderate_adj <- rate_mild_to_moderate * rr_mild_to_moderate
p_mild_to_moderate_int <- rep(1 - exp(-rate_mild_to_moderate_adj), n_cycles)

# Severe transition remains unchanged
p_mild_to_severe_int <- rep(p_mild_to_severe, n_cycles)
p_mild_stay_int <- 1 - (p_mild_to_moderate_int + p_mild_to_severe_int)

# Moderate and severe transitions are unaffected by intervention
p_moderate_to_mild_int <- p_moderate_to_mild_no_int
p_moderate_to_severe_int <- p_moderate_to_severe_no_int
p_moderate_stay_int <- p_moderate_stay_no_int

p_severe_to_moderate_int <- p_severe_to_moderate_no_int
p_severe_stay_int <- p_severe_stay_no_int
# Function to build transition probability array for a given strategy
build_transition_matrix <- function(strat) {
  # Define states and initialize array
  states <- c("ad_mci", "mild", "moderate", "severe", "death")
  n_states <- length(states)
  tp_array <- array(0, dim = c(n_states, n_states, n_cycles),
                    dimnames = list(from = states, to = states, cycle = 1:n_cycles))
  
  for (t in 1:n_cycles) {
    # Assign age-specific death probabilities (from Prompt 2)
    age <- start_age + t - 1
    tp_array["ad_mci", "death", t] <- death_probs$prob_death_ad_mci[death_probs$age == age]
    tp_array["mild", "death", t] <- death_probs$prob_death_mild[death_probs$age == age]
    tp_array["moderate", "death", t] <- death_probs$prob_death_moderate[death_probs$age == age]
    tp_array["severe", "death", t] <- death_probs$prob_death_severe[death_probs$age == age]
    tp_array["death", "death", t] <- 1  # Absorbing state
    
    if (strat == "no_intervention") {
      tp_array["ad_mci", "mild", t] <- p_ad_mci_to_mild_no_int[t] * (1 - tp_array["ad_mci", "death", t])
      tp_array["ad_mci", "ad_mci", t] <- p_ad_mci_stay_no_int[t] * (1 - tp_array["ad_mci", "death", t])
      
      tp_array["mild", "moderate", t] <- p_mild_to_moderate_no_int[t] * (1 - tp_array["mild", "death", t])
      tp_array["mild", "severe", t] <- p_mild_to_severe_no_int[t] * (1 - tp_array["mild", "death", t])
      tp_array["mild", "mild", t] <- p_mild_stay_no_int[t] * (1 - tp_array["mild", "death", t])
      
      tp_array["moderate", "mild", t] <- p_moderate_to_mild_no_int[t] * (1 - tp_array["moderate", "death", t])
      tp_array["moderate", "severe", t] <- p_moderate_to_severe_no_int[t] * (1 - tp_array["moderate", "death", t])
      tp_array["moderate", "moderate", t] <- p_moderate_stay_no_int[t] * (1 - tp_array["moderate", "death", t])
      
      tp_array["severe", "moderate", t] <- p_severe_to_moderate_no_int[t] * (1 - tp_array["severe", "death", t])
      tp_array["severe", "severe", t] <- p_severe_stay_no_int[t] * (1 - tp_array["severe", "death", t])
      
    } else if (strat == "intervention") {
      tp_array["ad_mci", "mild", t] <- p_ad_mci_to_mild_int[t] * (1 - tp_array["ad_mci", "death", t])
      tp_array["ad_mci", "ad_mci", t] <- p_ad_mci_stay_int[t] * (1 - tp_array["ad_mci", "death", t])
      
      tp_array["mild", "moderate", t] <- p_mild_to_moderate_int[t] * (1 - tp_array["mild", "death", t])
      tp_array["mild", "severe", t] <- p_mild_to_severe_int[t] * (1 - tp_array["mild", "death", t])
      tp_array["mild", "mild", t] <- p_mild_stay_int[t] * (1 - tp_array["mild", "death", t])
      
      tp_array["moderate", "mild", t] <- p_moderate_to_mild_int[t] * (1 - tp_array["moderate", "death", t])
      tp_array["moderate", "severe", t] <- p_moderate_to_severe_int[t] * (1 - tp_array["moderate", "death", t])
      tp_array["moderate", "moderate", t] <- p_moderate_stay_int[t] * (1 - tp_array["moderate", "death", t])
      
      tp_array["severe", "moderate", t] <- p_severe_to_moderate_int[t] * (1 - tp_array["severe", "death", t])
      tp_array["severe", "severe", t] <- p_severe_stay_int[t] * (1 - tp_array["severe", "death", t])
    }
    
    # Normalize rows to sum to 1
    for (s in states) {
      row_sum <- sum(tp_array[s, , t])
      if (row_sum > 0 && row_sum != 1) {
        tp_array[s, , t] <- tp_array[s, , t] / row_sum
      }
    }
  }
  
  return(tp_array)
}
# Define age range
ages <- 60:100

# Compute costs for each cognitive state using provided GLM equations
cost_ad_mci <- 37.961 * ages^2 - 1234.5 * ages + 124934
cost_mild <- 49.314 * ages^2 - 1828.7 * ages + 166239
cost_moderate <- 61.412 * ages^2 - 2086.3 * ages + 205578
cost_severe <- 131670 * exp(0.0176 * ages)
cost_death <- rep(0, length(ages))

# Create data frame
cost_by_age <- data.frame(
  age = ages,
  ad_mci = cost_ad_mci,
  mild = cost_mild,
  moderate = cost_moderate,
  severe = cost_severe,
  death = cost_death
)

# Function to run the Markov model for a given strategy
run_markov_model <- function(strat) {
  # Define states
  states <- c("ad_mci", "mild", "moderate", "severe", "death")
  n_states <- length(states)
  
  # Initialize transition matrix
  tp_matrix <- build_transition_matrix(strat)
  
  # Initialize state trace matrix
  trace <- matrix(0, nrow = n_cycles + 1, ncol = n_states,
                  dimnames = list(cycle = 0:n_cycles, state = states))
  trace[1, "ad_mci"] <- cohort_size  # Start entire cohort in AD-MCI
  
  # Initialize outputs
  qalys <- numeric(n_cycles)
  costs <- numeric(n_cycles)
  pys <- matrix(0, nrow = n_cycles, ncol = 4,
                dimnames = list(cycle = 1:n_cycles, state = states[1:4]))
  deaths_total <- numeric(n_cycles)
  deaths_ad_mci <- numeric(n_cycles)
  
  for (t in 1:n_cycles) {
    # Advance one cycle using transition matrix multiplication
    trace[t + 1, ] <- trace[t, ] %*% tp_matrix[, , t]
    
    # Current age
    age <- start_age + t - 1
    
    # Compute QALYs
    qalys[t] <- trace[t, "ad_mci"] * u_ad_mci(age) +
      trace[t, "mild"]   * u_mild(age) +
      trace[t, "moderate"] * u_moderate(age) +
      trace[t, "severe"] * u_severe(age)
    
    # Compute Costs
    age_costs <- cost_by_age[cost_by_age$age == age, ]
    total_cost <- trace[t, "ad_mci"] * age_costs$ad_mci +
      trace[t, "mild"]   * age_costs$mild +
      trace[t, "moderate"] * age_costs$moderate +
      trace[t, "severe"] * age_costs$severe
    
    # Add DMT cost for intervention strategy
    if (strat == "intervention") {
      total_cost <- total_cost +
        trace[t, "ad_mci"] * dmt_treatment_cost +
        trace[t, "mild"] * dmt_treatment_cost
    }
    
    costs[t] <- total_cost
    
    # Compute Person-Years for non-death states
    pys[t, "ad_mci"]   <- trace[t, "ad_mci"]
    pys[t, "mild"]     <- trace[t, "mild"]
    pys[t, "moderate"] <- trace[t, "moderate"]
    pys[t, "severe"]   <- trace[t, "severe"]
    
    # Total deaths this cycle
    deaths_total[t] <- trace[t + 1, "death"] - trace[t, "death"]
    
    # Cumulative deaths directly from AD-MCI
    deaths_ad_mci[t] <- trace[t, "ad_mci"] * tp_matrix["ad_mci", "death", t]
  }
  
  # Half-cycle correction
  qalys_hcc <- 0.5 * (qalys[-n_cycles] + qalys[-1])
  costs_hcc <- 0.5 * (costs[-n_cycles] + costs[-1])
  pys_hcc <- 0.5 * (pys[-n_cycles, ] + pys[-1, ])
  trace_hcc <- trace[-(n_cycles + 1), ]  # Remove last cycle
  
  # Discounting
  discount_vector <- 1 / (1 + discount_rate) ^ (0:(length(qalys_hcc) - 1))
  qalys_discounted <- sum(qalys_hcc * discount_vector) / cohort_size
  costs_discounted <- sum(costs_hcc * discount_vector) / cohort_size
  
  # Return results
  return(list(
    qalys = qalys_discounted,
    final_costs = costs_discounted,
    pys_ad_mci = sum(pys_hcc[, "ad_mci"]) / cohort_size,
    pys_mild = sum(pys_hcc[, "mild"]) / cohort_size,
    pys_mod = sum(pys_hcc[, "moderate"]) / cohort_size,
    pys_sev = sum(pys_hcc[, "severe"]) / cohort_size,
    deaths_total = sum(deaths_total),
    deaths_ad_mci = sum(deaths_ad_mci)
  ))
}

# Run model for both strategies
results <- list()
results$no_intervention <- run_markov_model("no_intervention")
results$intervention <- run_markov_model("intervention")

# Print summary table
summary_table <- data.frame(
  Strategy = c("No Intervention", "Intervention"),
  QALYs = c(results$no_intervention$qalys, results$intervention$qalys),
  Costs_SEK = c(results$no_intervention$final_costs, results$intervention$final_costs),
  PYs_AD_MCI = c(results$no_intervention$pys_ad_mci, results$intervention$pys_ad_mci),
  PYs_Mild = c(results$no_intervention$pys_mild, results$intervention$pys_mild),
  PYs_Moderate = c(results$no_intervention$pys_mod, results$intervention$pys_mod),
  PYs_Severe = c(results$no_intervention$pys_sev, results$intervention$pys_sev),
  Total_Deaths = c(results$no_intervention$deaths_total, results$intervention$deaths_total),
  Deaths_AD_MCI = c(results$no_intervention$deaths_ad_mci, results$intervention$deaths_ad_mci)
)
print(summary_table)
```
