## Supplementary material for "Replicating Health-Economic Simulation Models for Alzheimer’s Disease Using Artificial Intelligence": final R scripts: Wimo_O2.R

```
# Input parameters for Wimo et al. (2020) health-economic Markov model

# Descriptive parameters
state_names <- c("ad_mci", "mild", "moderate", "severe", "death")
strategy_names <- c("no_intervention", "intervention")
start_age <- 60
cycle_length <- 1  # in years
n_cycles <- 40
cohort_size <- 100000
discount_rate <- 0.03

# Disease-modifying treatment (DMT) cost (annual, in SEK)
cost_dmt <- 50000  # hypothetical anchor price:contentReference[oaicite:0]{index=0}

# Base case transition probabilities (without time dependency)
# From: AD-MCI
p_ad_mci_to_mild <- 0.206  # derived from 50% 3-year risk:contentReference[oaicite:1]{index=1}
p_ad_mci_to_death <- NA    # to be derived from life table (handled in prompt 2)
p_ad_mci_to_ad_mci <- 1 - p_ad_mci_to_mild  # assuming no direct death transitions here

# From: Mild AD
p_mild_to_mild <- 0.706
p_mild_to_moderate <- 0.293
p_mild_to_severe <- 0.001

# From: Moderate AD
p_moderate_to_mild <- 0.087
p_moderate_to_moderate <- 0.803
p_moderate_to_severe <- 0.109

# From: Severe AD
p_severe_to_moderate <- 0.196
p_severe_to_severe <- 0.804

# Transition to death for mild/moderate/severe AD will be age- and severity-specific
# using HRs below applied to population mortality (handled in prompt 2)

# Hazard ratios (HRs) for mortality based on MMSE categories:contentReference[oaicite:2]{index=2}
hr_mild <- 1.318
hr_moderate <- 2.419
hr_severe <- 4.267

# QALY utility functions (age-dependent, based on regression formulas)
u_ad_mci <- function(age) {
  -0.002464 * age + 0.962679
}

u_mild <- function(age) {
  -0.0025 * age + 0.8293
}

u_moderate <- function(age) {
  -4e-07 * age^2 - 0.0024 * age + 0.5828
}

u_severe <- function(age) {
  -4e-07 * age^2 - 0.0025 * age + 0.4337
}

# Utility for death state
u_death <- 0

# Relative risk reductions (RRs) applied in base case scenario (intervention strategy):contentReference[oaicite:3]{index=3}
rr_ad_mci_to_mild <- 0.75      # 25% risk reduction
rr_mild_to_moderate <- 0.75    # 25% risk reduction
# Load mortality rate data
mort_rates <- read.csv("mortalityRates_Swedish_2016.csv", stringsAsFactors = FALSE)
weights <- read.csv("weightsMortalityRates_Swedish_2016.csv", stringsAsFactors = FALSE)

# Create a mapping of age groups to individual ages
age_groups <- list(
  "60-64" = 60:64,
  "65-69" = 65:69,
  "70-74" = 70:74,
  "75-79" = 75:79,
  "80-84" = 80:84,
  "85-89" = 85:89,
  "90+"   = 90:99
)

# Expand the grouped mortality rates to individual ages
expanded_rates <- data.frame(age = integer(), male = numeric(), female = numeric())

for (i in seq_len(nrow(mort_rates))) {
  group <- mort_rates$age[i]
  ages <- age_groups[[group]]
  male_rate <- mort_rates$male[i]
  female_rate <- mort_rates$female[i]
  
  temp <- data.frame(age = ages, male = rep(male_rate, length(ages)), female = rep(female_rate, length(ages)))
  expanded_rates <- rbind(expanded_rates, temp)
}

# Merge with weights
merged_data <- merge(expanded_rates, weights, by = "age")

# Compute sex-weighted average mortality rate
merged_data$rate_avg <- with(merged_data, (male * weight_male + female * weight_female) / (weight_male + weight_female))

# Define hazard ratios from previous prompt
hr_mild <- 1.318
hr_moderate <- 2.419
hr_severe <- 4.267

# Compute probability of death for each severity state
merged_data$prob_death_ad_mci <- 1 - exp(-merged_data$rate_avg * 1)  # HR = 1
merged_data$prob_death_mild <- 1 - exp(-merged_data$rate_avg * hr_mild)
merged_data$prob_death_moderate <- 1 - exp(-merged_data$rate_avg * hr_moderate)
merged_data$prob_death_severe <- 1 - exp(-merged_data$rate_avg * hr_severe)

# Create final data frame
mortality_df <- merged_data[, c("age", "rate_avg", "prob_death_ad_mci", "prob_death_mild", "prob_death_moderate", "prob_death_severe")]
# Define time-dependent transition probability vectors for n_cycles
# Note: p_ad_mci_to_mild, p_mild_to_moderate, etc., and RRs already defined previously

# Define number of cycles
# (already defined earlier, but included here for clarity)
# n_cycles <- 40

# Relative risks for intervention (from previous prompt)
rr_ad_mci_to_mild <- 0.75
rr_mild_to_moderate <- 0.75

# ---- NO INTERVENTION STRATEGY ----

# From "ad_mci"
p_ad_mci_to_mild_vec_no_int <- rep(p_ad_mci_to_mild, n_cycles)
p_ad_mci_to_ad_mci_vec_no_int <- 1 - p_ad_mci_to_mild_vec_no_int

# From "mild"
p_mild_to_moderate_vec_no_int <- rep(p_mild_to_moderate, n_cycles)
p_mild_to_severe_vec_no_int <- rep(p_mild_to_severe, n_cycles)
p_mild_to_mild_vec_no_int <- 1 - p_mild_to_moderate_vec_no_int - p_mild_to_severe_vec_no_int

# From "moderate"
p_moderate_to_mild_vec_no_int <- rep(p_moderate_to_mild, n_cycles)
p_moderate_to_severe_vec_no_int <- rep(p_moderate_to_severe, n_cycles)
p_moderate_to_moderate_vec_no_int <- 1 - p_moderate_to_mild_vec_no_int - p_moderate_to_severe_vec_no_int

# From "severe"
p_severe_to_moderate_vec_no_int <- rep(p_severe_to_moderate, n_cycles)
p_severe_to_severe_vec_no_int <- 1 - p_severe_to_moderate_vec_no_int

# ---- INTERVENTION STRATEGY ----

# From "ad_mci" (adjusted for RR)
rate_ad_mci_to_mild <- -log(1 - p_ad_mci_to_mild)
adj_rate_ad_mci_to_mild <- rate_ad_mci_to_mild * rr_ad_mci_to_mild
adj_p_ad_mci_to_mild <- 1 - exp(-adj_rate_ad_mci_to_mild)

p_ad_mci_to_mild_vec_int <- rep(adj_p_ad_mci_to_mild, n_cycles)
p_ad_mci_to_ad_mci_vec_int <- 1 - p_ad_mci_to_mild_vec_int

# From "mild" (adjusted for RR)
rate_mild_to_moderate <- -log(1 - p_mild_to_moderate)
adj_rate_mild_to_moderate <- rate_mild_to_moderate * rr_mild_to_moderate
adj_p_mild_to_moderate <- 1 - exp(-adj_rate_mild_to_moderate)

p_mild_to_moderate_vec_int <- rep(adj_p_mild_to_moderate, n_cycles)
p_mild_to_severe_vec_int <- p_mild_to_severe_vec_no_int  # unchanged
p_mild_to_mild_vec_int <- 1 - p_mild_to_moderate_vec_int - p_mild_to_severe_vec_int

# From "moderate" (not affected by intervention)
p_moderate_to_mild_vec_int <- p_moderate_to_mild_vec_no_int
p_moderate_to_severe_vec_int <- p_moderate_to_severe_vec_no_int
p_moderate_to_moderate_vec_int <- p_moderate_to_moderate_vec_no_int

# From "severe" (not affected by intervention)
p_severe_to_moderate_vec_int <- p_severe_to_moderate_vec_no_int
p_severe_to_severe_vec_int <- p_severe_to_severe_vec_no_int


# Define state names and number of states
state_names <- c("ad_mci", "mild", "moderate", "severe", "death")
n_states <- length(state_names)

# Function to build the transition probability matrix array
build_transition_matrix <- function(strat) {
  # Initialize transition probability array: states × states × cycles
  tpm_array <- array(0, dim = c(n_states, n_states, n_cycles),
                     dimnames = list(state_names, state_names, NULL))
  
  for (t in 1:n_cycles) {
    # Transition to death for each state from mortality_df (previously created)
    age <- start_age + t - 1
    p_death_ad_mci <- mortality_df$prob_death_ad_mci[mortality_df$age == age]
    p_death_mild <- mortality_df$prob_death_mild[mortality_df$age == age]
    p_death_moderate <- mortality_df$prob_death_moderate[mortality_df$age == age]
    p_death_severe <- mortality_df$prob_death_severe[mortality_df$age == age]
    
    # Assign death probabilities for each non-death state
    tpm_array["ad_mci", "death", t] <- p_death_ad_mci
    tpm_array["mild", "death", t] <- p_death_mild
    tpm_array["moderate", "death", t] <- p_death_moderate
    tpm_array["severe", "death", t] <- p_death_severe
    
    # Death is absorbing
    tpm_array["death", "death", t] <- 1
    
    # [INSERT PROMPT 4B HERE] — define transitions from on-treatment and off-treatment states
    
    if (strat == "no_intervention") {
      # From ad_mci
      tpm_array["ad_mci", "mild", t] <- p_ad_mci_to_mild_vec_no_int[t] * (1 - p_death_ad_mci)
      tpm_array["ad_mci", "ad_mci", t] <- p_ad_mci_to_ad_mci_vec_no_int[t] * (1 - p_death_ad_mci)
      
      # From mild
      tpm_array["mild", "moderate", t] <- p_mild_to_moderate_vec_no_int[t] * (1 - p_death_mild)
      tpm_array["mild", "severe", t]   <- p_mild_to_severe_vec_no_int[t] * (1 - p_death_mild)
      tpm_array["mild", "mild", t]     <- p_mild_to_mild_vec_no_int[t] * (1 - p_death_mild)
      
      # From moderate
      tpm_array["moderate", "mild", t]     <- p_moderate_to_mild_vec_no_int[t] * (1 - p_death_moderate)
      tpm_array["moderate", "severe", t]   <- p_moderate_to_severe_vec_no_int[t] * (1 - p_death_moderate)
      tpm_array["moderate", "moderate", t] <- p_moderate_to_moderate_vec_no_int[t] * (1 - p_death_moderate)
      
      # From severe
      tpm_array["severe", "moderate", t] <- p_severe_to_moderate_vec_no_int[t] * (1 - p_death_severe)
      tpm_array["severe", "severe", t]   <- p_severe_to_severe_vec_no_int[t] * (1 - p_death_severe)
      
    } else if (strat == "intervention") {
      # From ad_mci
      tpm_array["ad_mci", "mild", t] <- p_ad_mci_to_mild_vec_int[t] * (1 - p_death_ad_mci)
      tpm_array["ad_mci", "ad_mci", t] <- p_ad_mci_to_ad_mci_vec_int[t] * (1 - p_death_ad_mci)
      
      # From mild
      tpm_array["mild", "moderate", t] <- p_mild_to_moderate_vec_int[t] * (1 - p_death_mild)
      tpm_array["mild", "severe", t]   <- p_mild_to_severe_vec_int[t] * (1 - p_death_mild)
      tpm_array["mild", "mild", t]     <- p_mild_to_mild_vec_int[t] * (1 - p_death_mild)
      
      # From moderate
      tpm_array["moderate", "mild", t]     <- p_moderate_to_mild_vec_int[t] * (1 - p_death_moderate)
      tpm_array["moderate", "severe", t]   <- p_moderate_to_severe_vec_int[t] * (1 - p_death_moderate)
      tpm_array["moderate", "moderate", t] <- p_moderate_to_moderate_vec_int[t] * (1 - p_death_moderate)
      
      # From severe
      tpm_array["severe", "moderate", t] <- p_severe_to_moderate_vec_int[t] * (1 - p_death_severe)
      tpm_array["severe", "severe", t]   <- p_severe_to_severe_vec_int[t] * (1 - p_death_severe)
    }
    
    # Normalize each row so that row sums to 1 (if needed)
    for (s in state_names) {
      row_sum <- sum(tpm_array[s, , t])
      if (row_sum > 0 && abs(row_sum - 1) > 1e-8) {
        tpm_array[s, , t] <- tpm_array[s, , t] / row_sum
      }
    }
  }
  
  return(tpm_array)
}
# Define the vector of ages
ages <- 60:100

# Compute cost per cognitive state using provided equations
cost_ad_mci <- 37.961 * ages^2 - 1234.5 * ages + 124934
cost_mild <- 49.314 * ages^2 - 1828.7 * ages + 166239
cost_moderate <- 61.412 * ages^2 - 2086.3 * ages + 205578
cost_severe <- 131670 * exp(0.0176 * ages)
cost_death <- rep(0, length(ages))

# Combine into a data frame
annual_costs_df <- data.frame(
  age = ages,
  ad_mci = cost_ad_mci,
  mild = cost_mild,
  moderate = cost_moderate,
  severe = cost_severe,
  death = cost_death
)

# Function to run the Markov model for a given strategy
run_markov_model <- function(strategy) {
  # Initialize variables
  trace <- matrix(0, nrow = n_cycles + 1, ncol = 5,
                  dimnames = list(0:n_cycles, state_names))
  trace[1, "ad_mci"] <- cohort_size
  
  qalys <- numeric(n_cycles)
  costs <- numeric(n_cycles)
  pys <- matrix(0, nrow = n_cycles, ncol = 4,
                dimnames = list(1:n_cycles, c("ad_mci", "mild", "moderate", "severe")))
  deaths_total <- numeric(n_cycles)
  deaths_ad_mci <- numeric(n_cycles)
  
  # Build transition matrix
  tpm <- build_transition_matrix(strategy)
  
  for (t in 1:n_cycles) {
    # Update cohort distribution
    trace[t + 1, ] <- trace[t, ] %*% tpm[, , t]
    
    # Current age
    current_age <- start_age + t - 1
    
    # Compute QALYs (using utility functions defined earlier)
    qalys[t] <- trace[t, "ad_mci"] * u_ad_mci(current_age) +
      trace[t, "mild"]   * u_mild(current_age) +
      trace[t, "moderate"] * u_moderate(current_age) +
      trace[t, "severe"] * u_severe(current_age)
    # No utility for death state
    
    # Compute Costs using precomputed cost table
    costs[t] <- trace[t, "ad_mci"] * annual_costs_df$ad_mci[annual_costs_df$age == current_age] +
      trace[t, "mild"]   * annual_costs_df$mild[annual_costs_df$age == current_age] +
      trace[t, "moderate"] * annual_costs_df$moderate[annual_costs_df$age == current_age] +
      trace[t, "severe"] * annual_costs_df$severe[annual_costs_df$age == current_age]
    # No cost for death
    
    # Add DMT cost if strategy is intervention
    if (strategy == "intervention") {
      costs[t] <- costs[t] +
        trace[t, "ad_mci"] * cost_dmt +
        trace[t, "mild"] * cost_dmt
    }
    
    # Store person-years (state occupancies before transitions)
    pys[t, "ad_mci"]   <- trace[t, "ad_mci"]
    pys[t, "mild"]     <- trace[t, "mild"]
    pys[t, "moderate"] <- trace[t, "moderate"]
    pys[t, "severe"]   <- trace[t, "severe"]
    
    # Track total deaths in current cycle
    deaths_total[t] <- trace[t + 1, "death"] - trace[t, "death"]
    
    # Track cumulative deaths from AD-MCI
    deaths_ad_mci[t] <- trace[t, "ad_mci"] * tpm["ad_mci", "death", t]
  }
  
  # Half-cycle correction: average each cycle with next, drop last
  qalys <- (qalys[-1] + qalys[-length(qalys)]) / 2
  costs <- (costs[-1] + costs[-length(costs)]) / 2
  pys <- (pys[-1, , drop = FALSE] + pys[-n_cycles, , drop = FALSE]) / 2
  deaths_total <- deaths_total[-n_cycles]
  deaths_ad_mci <- deaths_ad_mci[-n_cycles]
  trace <- trace[-(n_cycles + 1), , drop = FALSE]
  
  # Apply discounting
  discount_factors <- 1 / (1 + discount_rate)^(1:(n_cycles - 1))
  qalys_discounted <- sum(qalys * discount_factors) / cohort_size
  costs_discounted <- sum(costs * discount_factors) / cohort_size
  
  # Return results
  return(list(
    qalys = qalys_discounted,
    final_costs = costs_discounted,
    pys_ad_mci = sum(pys[, "ad_mci"]) / cohort_size,
    pys_mild = sum(pys[, "mild"]) / cohort_size,
    pys_mod = sum(pys[, "moderate"]) / cohort_size,
    pys_sev = sum(pys[, "severe"]) / cohort_size,
    deaths_total = sum(deaths_total),
    deaths_ad_mci = sum(deaths_ad_mci)
  ))
}

# Run model for both strategies
results <- list()
results$no_intervention <- run_markov_model("no_intervention")
results$intervention <- run_markov_model("intervention")
```
