## Supplementary material for "Replicating Health-Economic Simulation Models for Alzheimer’s Disease Using Artificial Intelligence": final R scripts: Wimo_O3.R

```
# ===============================
# Input Parameters for Markov Model
# Based on Wimo et al. (2020)
# ===============================

# ---- State Names ----
state_names <- c("ad_mci", "mild", "moderate", "severe", "death")

# ---- Strategy Names ----
strategies <- c("no_intervention", "intervention")

# ---- Model Settings ----
start_age <- 60
cycle_length <- 1
n_cycles <- 40
discount_rate <- 0.03
cohort_size <- 100000

# ---- Disease Progression Probabilities (Base Case - No Intervention) ----
# Annual probability derived from a 3-year cumulative risk of 50%
p_ad_mci_to_mild <- 1 - (1 - 0.50)^(1 / 3)  # ≈ 0.206

# ---- Hazard Ratios for Mortality (Compared to Very Mild AD) ----
hr_mild <- 1.318
hr_moderate <- 2.419
hr_severe <- 4.267

# ---- Hypothetical DMT Intervention Effects ----
# 25% reduction in progression risk
rr_ad_mci_to_mild <- 0.75
rr_mild_to_moderate <- 0.75

# ---- Treatment Cost ----
dmt_cost_per_year <- 50000  # SEK/year

# ---- Utility Functions (QALY) ----
u_ad_mci <- function(age) {
  -0.002464 * age + 0.962679
}

u_mild <- function(age) {
  -0.0025 * age + 0.8293
}

u_moderate <- function(age) {
  -4e-07 * age^2 - 0.0024 * age + 0.5828
}

u_severe <- function(age) {
  -4e-07 * age^2 - 0.0025 * age + 0.4337
}

u_death <- function(age) {
  return(0)
}

# ---- Transition Probabilities (Other than AD-MCI to Mild) ----
# These will be used in the transition matrix later, just define now as variables
p_mild_to_moderate <- 0.293
p_mild_to_severe <- 0.001
p_mild_stay <- 0.706

p_moderate_to_mild <- 0.087
p_moderate_to_severe <- 0.109
p_moderate_stay <- 0.803

p_severe_to_moderate <- 0.196
p_severe_stay <- 0.804

# Placeholder for age-specific death probabilities; will be defined from life tables later
# p_death_ad_mci, p_death_mild, p_death_moderate, p_death_severe to be computed later
# ===============================
# Mortality Rate Processing Script
# ===============================

# ---- Load Data ----
mortality <- read.csv("mortalityRates_Swedish_2016.csv", stringsAsFactors = FALSE)
weights <- read.csv("weightsMortalityRates_Swedish_2016.csv", stringsAsFactors = FALSE)

# ---- Define Age Groups ----
age_groups <- c("60-64", "65-69", "70-74", "75-79", "80-84", "85-89", "90+")
age_group_ranges <- list(
  "60-64" = 60:64,
  "65-69" = 65:69,
  "70-74" = 70:74,
  "75-79" = 75:79,
  "80-84" = 80:84,
  "85-89" = 85:89,
  "90+"   = 90:99
)

# ---- Expand Grouped Mortality Rates to Individual Ages ----
expanded <- data.frame(age = integer(), rate_male = numeric(), rate_female = numeric())

for (i in 1:nrow(mortality)) {
  group <- mortality$age[i]
  ages <- age_group_ranges[[group]]
  rate_male <- rep(mortality$male[i], length(ages))
  rate_female <- rep(mortality$female[i], length(ages))
  expanded <- rbind(expanded, data.frame(age = ages, rate_male = rate_male, rate_female = rate_female))
}

# ---- Merge with Weights ----
merged <- merge(expanded, weights, by = "age")

# ---- Compute Weighted Average Mortality Rate ----
merged$rate_avg <- with(merged, (rate_male * weight_male + rate_female * weight_female) /
                          (weight_male + weight_female))

# ---- Hazard Ratios from Previous Prompt ----
hr_mild <- 1.318
hr_moderate <- 2.419
hr_severe <- 4.267

# ---- Compute Death Probabilities ----
merged$prob_death_ad_mci <- 1 - exp(-merged$rate_avg)
merged$prob_death_mild <- 1 - exp(-merged$rate_avg * hr_mild)
merged$prob_death_moderate <- 1 - exp(-merged$rate_avg * hr_moderate)
merged$prob_death_severe <- 1 - exp(-merged$rate_avg * hr_severe)

# ---- Select Final Columns ----
mortality_final <- merged[, c("age", "rate_avg", "prob_death_ad_mci", "prob_death_mild",
                              "prob_death_moderate", "prob_death_severe")]

# ---- Result: mortality_final data frame with all required columns ----
# mortality_final is ready for use in subsequent model components.
# ===============================
# Time-dependent Transition Probabilities
# ===============================

# ---- No Intervention Strategy ----
# From AD-MCI
p_ad_mci_to_mild_vec_noint <- rep(p_ad_mci_to_mild, n_cycles)
p_ad_mci_stay_vec_noint <- 1 - p_ad_mci_to_mild_vec_noint

# From Mild
p_mild_to_moderate_vec_noint <- rep(p_mild_to_moderate, n_cycles)
p_mild_to_severe_vec_noint <- rep(p_mild_to_severe, n_cycles)
p_mild_stay_vec_noint <- 1 - (p_mild_to_moderate_vec_noint + p_mild_to_severe_vec_noint)

# From Moderate
p_moderate_to_mild_vec_noint <- rep(p_moderate_to_mild, n_cycles)
p_moderate_to_severe_vec_noint <- rep(p_moderate_to_severe, n_cycles)
p_moderate_stay_vec_noint <- 1 - (p_moderate_to_mild_vec_noint + p_moderate_to_severe_vec_noint)

# From Severe
p_severe_to_moderate_vec_noint <- rep(p_severe_to_moderate, n_cycles)
p_severe_stay_vec_noint <- 1 - p_severe_to_moderate_vec_noint

# ---- Intervention Strategy ----
# From AD-MCI (adjusted)
rate_ad_mci_to_mild <- -log(1 - p_ad_mci_to_mild)
adjusted_rate_ad_mci_to_mild <- rate_ad_mci_to_mild * rr_ad_mci_to_mild
p_ad_mci_to_mild_vec_int <- rep(1 - exp(-adjusted_rate_ad_mci_to_mild), n_cycles)
p_ad_mci_stay_vec_int <- 1 - p_ad_mci_to_mild_vec_int

# From Mild (adjusted)
rate_mild_to_moderate <- -log(1 - p_mild_to_moderate)
rate_mild_to_severe <- -log(1 - p_mild_to_severe)
adjusted_rate_mild_to_moderate <- rate_mild_to_moderate * rr_mild_to_moderate
adjusted_rate_mild_to_severe <- rate_mild_to_severe  # unchanged (not targeted)
p_mild_to_moderate_vec_int <- rep(1 - exp(-adjusted_rate_mild_to_moderate), n_cycles)
p_mild_to_severe_vec_int <- rep(1 - exp(-adjusted_rate_mild_to_severe), n_cycles)
p_mild_stay_vec_int <- 1 - (p_mild_to_moderate_vec_int + p_mild_to_severe_vec_int)

# From Moderate (unchanged)
p_moderate_to_mild_vec_int <- p_moderate_to_mild_vec_noint
p_moderate_to_severe_vec_int <- p_moderate_to_severe_vec_noint
p_moderate_stay_vec_int <- p_moderate_stay_vec_noint

# From Severe (unchanged)
p_severe_to_moderate_vec_int <- p_severe_to_moderate_vec_noint
p_severe_stay_vec_int <- p_severe_stay_vec_noint


# ===============================
# Transition Probability Matrix Builder
# ===============================

build_transition_matrix <- function(strat) {
  # Initialize the transition probability array
  tpm <- array(0, dim = c(length(state_names), length(state_names), n_cycles),
               dimnames = list(state_names, state_names, NULL))
  
  for (cycle in 1:n_cycles) {
    # --- Assign probabilities to death state ---
    tpm["ad_mci", "death", cycle] <- mortality_final$prob_death_ad_mci[cycle]
    tpm["mild", "death", cycle] <- mortality_final$prob_death_mild[cycle]
    tpm["moderate", "death", cycle] <- mortality_final$prob_death_moderate[cycle]
    tpm["severe", "death", cycle] <- mortality_final$prob_death_severe[cycle]
    tpm["death", "death", cycle] <- 1  # Absorbing state
    
    if (strat == "no_intervention") {
      # From AD-MCI
      tpm["ad_mci", "mild", cycle] <- p_ad_mci_to_mild_vec_noint[cycle] * (1 - tpm["ad_mci", "death", cycle])
      tpm["ad_mci", "ad_mci", cycle] <- p_ad_mci_stay_vec_noint[cycle] * (1 - tpm["ad_mci", "death", cycle])
      
      # From Mild
      tpm["mild", "moderate", cycle] <- p_mild_to_moderate_vec_noint[cycle] * (1 - tpm["mild", "death", cycle])
      tpm["mild", "severe", cycle] <- p_mild_to_severe_vec_noint[cycle] * (1 - tpm["mild", "death", cycle])
      tpm["mild", "mild", cycle] <- p_mild_stay_vec_noint[cycle] * (1 - tpm["mild", "death", cycle])
      
      # From Moderate
      tpm["moderate", "mild", cycle] <- p_moderate_to_mild_vec_noint[cycle] * (1 - tpm["moderate", "death", cycle])
      tpm["moderate", "severe", cycle] <- p_moderate_to_severe_vec_noint[cycle] * (1 - tpm["moderate", "death", cycle])
      tpm["moderate", "moderate", cycle] <- p_moderate_stay_vec_noint[cycle] * (1 - tpm["moderate", "death", cycle])
      
      # From Severe
      tpm["severe", "moderate", cycle] <- p_severe_to_moderate_vec_noint[cycle] * (1 - tpm["severe", "death", cycle])
      tpm["severe", "severe", cycle] <- p_severe_stay_vec_noint[cycle] * (1 - tpm["severe", "death", cycle])
      
    } else if (strat == "intervention") {
      # From AD-MCI
      tpm["ad_mci", "mild", cycle] <- p_ad_mci_to_mild_vec_int[cycle] * (1 - tpm["ad_mci", "death", cycle])
      tpm["ad_mci", "ad_mci", cycle] <- p_ad_mci_stay_vec_int[cycle] * (1 - tpm["ad_mci", "death", cycle])
      
      # From Mild
      tpm["mild", "moderate", cycle] <- p_mild_to_moderate_vec_int[cycle] * (1 - tpm["mild", "death", cycle])
      tpm["mild", "severe", cycle] <- p_mild_to_severe_vec_int[cycle] * (1 - tpm["mild", "death", cycle])
      tpm["mild", "mild", cycle] <- p_mild_stay_vec_int[cycle] * (1 - tpm["mild", "death", cycle])
      
      # From Moderate
      tpm["moderate", "mild", cycle] <- p_moderate_to_mild_vec_int[cycle] * (1 - tpm["moderate", "death", cycle])
      tpm["moderate", "severe", cycle] <- p_moderate_to_severe_vec_int[cycle] * (1 - tpm["moderate", "death", cycle])
      tpm["moderate", "moderate", cycle] <- p_moderate_stay_vec_int[cycle] * (1 - tpm["moderate", "death", cycle])
      
      # From Severe
      tpm["severe", "moderate", cycle] <- p_severe_to_moderate_vec_int[cycle] * (1 - tpm["severe", "death", cycle])
      tpm["severe", "severe", cycle] <- p_severe_stay_vec_int[cycle] * (1 - tpm["severe", "death", cycle])
    }
    
    # --- Normalize rows if needed ---
    for (state in state_names) {
      row_sum <- sum(tpm[state, , cycle])
      if (abs(row_sum - 1) > 1e-8 && row_sum > 0) {
        tpm[state, , cycle] <- tpm[state, , cycle] / row_sum
      }
    }
  }
  
  return(tpm)
}
# ===============================
# Annual Societal Costs by Age and Cognitive State
# ===============================

# Define age range
ages <- 60:100

# Calculate costs for each state using the provided GLM-fitted equations
cost_ad_mci     <- 37.961 * ages^2 - 1234.5 * ages + 124934
cost_mild       <- 49.314 * ages^2 - 1828.7 * ages + 166239
cost_moderate   <- 61.412 * ages^2 - 2086.3 * ages + 205578
cost_severe     <- 131670 * exp(0.0176 * ages)
cost_death      <- rep(0, length(ages))

# Combine into a data frame
annual_costs <- data.frame(
  age = ages,
  ad_mci = cost_ad_mci,
  mild = cost_mild,
  moderate = cost_moderate,
  severe = cost_severe,
  death = cost_death
)

# annual_costs is now ready for use in outcome calculations
# ===============================
# Run Markov Model for Both Strategies
# ===============================

run_markov_model <- function(strat) {
  # Transition matrix
  tpm <- build_transition_matrix(strat)
  
  # Initialize state trace matrix
  state_trace <- matrix(0, nrow = n_cycles + 1, ncol = length(state_names))
  colnames(state_trace) <- state_names
  state_trace[1, "ad_mci"] <- cohort_size  # All individuals start in ad_mci
  
  # Initialize outcome vectors
  qalys <- numeric(n_cycles)
  costs <- numeric(n_cycles)
  
  # Person-years in each alive state
  pys <- matrix(0, nrow = n_cycles, ncol = 4)
  colnames(pys) <- c("ad_mci", "mild", "moderate", "severe")
  
  # Death trackers
  deaths_total <- numeric(n_cycles)
  deaths_ad_mci <- numeric(n_cycles)
  
  for (cycle in 1:n_cycles) {
    # Advance the cohort using transition matrix for the current cycle
    state_trace[cycle + 1, ] <- state_trace[cycle, ] %*% tpm[, , cycle]
    
    # Calculate current age
    age <- start_age + cycle - 1
    
    # QALYs: utility × population in each state
    qalys[cycle] <- state_trace[cycle, "ad_mci"] * u_ad_mci(age) +
      state_trace[cycle, "mild"] * u_mild(age) +
      state_trace[cycle, "moderate"] * u_moderate(age) +
      state_trace[cycle, "severe"] * u_severe(age)
    
    # Costs: per-person cost × number of people in each state
    cost_row <- annual_costs[annual_costs$age == age, ]
    costs[cycle] <- state_trace[cycle, "ad_mci"] * cost_row$ad_mci +
      state_trace[cycle, "mild"] * cost_row$mild +
      state_trace[cycle, "moderate"] * cost_row$moderate +
      state_trace[cycle, "severe"] * cost_row$severe
    if (strat == "intervention") {
      costs[cycle] <- costs[cycle] +
        state_trace[cycle, "ad_mci"] * dmt_cost_per_year +
        state_trace[cycle, "mild"] * dmt_cost_per_year
    }
    
    # Person-years (PYs) in each living state
    pys[cycle, "ad_mci"] <- state_trace[cycle, "ad_mci"]
    pys[cycle, "mild"] <- state_trace[cycle, "mild"]
    pys[cycle, "moderate"] <- state_trace[cycle, "moderate"]
    pys[cycle, "severe"] <- state_trace[cycle, "severe"]
    
    # Total deaths this cycle
    deaths_total[cycle] <- state_trace[cycle + 1, "death"] - state_trace[cycle, "death"]
    
    # Cumulative deaths from AD-MCI (only direct transitions in current cycle)
    deaths_ad_mci[cycle] <- state_trace[cycle, "ad_mci"] * tpm["ad_mci", "death", cycle]
  }
  
  # Half-cycle correction for QALYs, costs, and PYs
  qalys_hcc <- 0.5 * (qalys[-n_cycles] + qalys[-1])
  costs_hcc <- 0.5 * (costs[-n_cycles] + costs[-1])
  pys_hcc <- 0.5 * (pys[-n_cycles, ] + pys[-1, ])
  
  # Apply discounting (excluding last cycle after correction)
  discount_factors <- 1 / ((1 + discount_rate) ^ (0:(length(qalys_hcc) - 1)))
  qalys_discounted <- sum(qalys_hcc * discount_factors) / cohort_size
  costs_discounted <- sum(costs_hcc * discount_factors) / cohort_size
  
  # Final outputs
  return(list(
    qalys = qalys_discounted,
    final_costs = costs_discounted,
    pys_ad_mci = sum(pys_hcc[, "ad_mci"]) / cohort_size,
    pys_mild = sum(pys_hcc[, "mild"]) / cohort_size,
    pys_mod = sum(pys_hcc[, "moderate"]) / cohort_size,
    pys_sev = sum(pys_hcc[, "severe"]) / cohort_size,
    deaths_total = sum(deaths_total),
    deaths_ad_mci = sum(deaths_ad_mci)
  ))
}

# Run model for both strategies
results <- list()
results$no_intervention <- run_markov_model("no_intervention")
results$intervention <- run_markov_model("intervention")
```
