## Supplementary material for "Replicating Health-Economic Simulation Models for Alzheimer’s Disease Using Artificial Intelligence": final R scripts: Wimo_O4.R

```
# =============================
# Wimo et al. (2020) - Markov Model Inputs
# =============================

# --- State and Strategy Names ---
states <- c("ad_mci", "mild", "moderate", "severe", "death")
strategies <- c("no_intervention", "intervention")

# --- General Model Parameters ---
start_age <- 60
cycle_length <- 1  # years
n_cycles <- 40
discount_rate <- 0.03
cohort_size <- 100000

# --- Disease Progression Probabilities (Annual, Base Case) ---
# Derived from Supplementary Table 3
p_mild_to_mild     <- 0.706
p_mild_to_moderate <- 0.293
p_mild_to_severe   <- 0.001

p_moderate_to_mild     <- 0.087
p_moderate_to_moderate <- 0.803
p_moderate_to_severe   <- 0.109

p_severe_to_moderate <- 0.196
p_severe_to_severe   <- 0.804

# From core text: Annual transition from AD-MCI to mild
p_ad_mci_to_mild <- 0.206

# Base case mortality handled separately using mortality data (Prompt 2)

# --- Treatment Effect Parameters ---
# Intervention assumed to reduce progression from AD-MCI and mild by 25%
rr_ad_mci_to_mild <- 0.75
rr_mild_to_moderate <- 0.75

# --- Hazard Ratios for Mortality by State (vs. Very mild/normal) ---
# From Supplementary Table 4
hr_mild     <- 1.318
hr_moderate <- 2.419
hr_severe   <- 4.267

# --- DMT Annual Cost (Base Case Anchor Price) ---
dmt_cost_per_year <- 50000  # SEK

# --- QALY Utility Functions (from Supplementary Table 6 regression models) ---
u_ad_mci <- function(age) {
  -0.002464 * age + 0.962679
}

u_mild <- function(age) {
  -0.0025 * age + 0.8293
}

u_moderate <- function(age) {
  -4e-07 * age^2 - 0.0024 * age + 0.5828
}

u_severe <- function(age) {
  -4e-07 * age^2 - 0.0025 * age + 0.4337
}

u_death <- function(age) {
  return(0)
}
# =============================
# Prompt 2: Load and Process Mortality Data
# =============================

# --- Load Mortality Rates and Weights ---
mort_rates <- read.csv("mortalityRates_Swedish_2016.csv", stringsAsFactors = FALSE)
weights <- read.csv("weightsMortalityRates_Swedish_2016.csv", stringsAsFactors = FALSE)

# --- Expand Grouped Mortality Rates to Individual Ages (60–99) ---
# Define age group mapping
age_groups <- list(
  "60-64" = 60:64,
  "65-69" = 65:69,
  "70-74" = 70:74,
  "75-79" = 75:79,
  "80-84" = 80:84,
  "85-89" = 85:89,
  "90+"   = 90:99
)

# Create an empty list to store per-age rates
expanded_rates <- data.frame()

# Loop through each age group and replicate rates
for (i in 1:nrow(mort_rates)) {
  group <- mort_rates$age[i]
  ages <- age_groups[[group]]
  temp <- data.frame(
    age = ages,
    male_rate = rep(mort_rates$male[i], length(ages)),
    female_rate = rep(mort_rates$female[i], length(ages))
  )
  expanded_rates <- rbind(expanded_rates, temp)
}

# --- Merge with Weights ---
merged <- merge(expanded_rates, weights, by = "age")

# --- Compute Sex-Weighted Average Mortality Rate ---
merged$rate_avg <- with(merged, 
                        (male_rate * weight_male + female_rate * weight_female) / 
                          (weight_male + weight_female))

# --- Compute Probability of Death for Each State ---
# Using defined hazard ratios from Prompt 1
hr_mild     <- 1.318
hr_moderate <- 2.419
hr_severe   <- 4.267

merged$prob_death_ad_mci   <- 1 - exp(-merged$rate_avg * 1)  # HR = 1
merged$prob_death_mild     <- 1 - exp(-merged$rate_avg * hr_mild)
merged$prob_death_moderate <- 1 - exp(-merged$rate_avg * hr_moderate)
merged$prob_death_severe   <- 1 - exp(-merged$rate_avg * hr_severe)

# --- Final Output Data Frame ---
mortality_df <- merged[, c("age", "rate_avg", "prob_death_ad_mci",
                           "prob_death_mild", "prob_death_moderate", "prob_death_severe")]

# Result: mortality_df with all required columns
# =============================
# Prompt 3: Time-dependent Transition Probability Vectors
# =============================

# --- Initialize cycle length ---
# Ensure all vectors have length n_cycles
len <- n_cycles

# --- No Intervention Strategy ---

# AD-MCI: transitions
p_ad_mci_to_mild_vec_no_int <- rep(p_ad_mci_to_mild, len)
p_ad_mci_to_ad_mci_vec_no_int <- 1 - p_ad_mci_to_mild_vec_no_int

# Mild: transitions
p_mild_to_moderate_vec_no_int <- rep(p_mild_to_moderate, len)
p_mild_to_severe_vec_no_int <- rep(p_mild_to_severe, len)
p_mild_to_mild_vec_no_int <- 1 - (p_mild_to_moderate_vec_no_int + p_mild_to_severe_vec_no_int)

# Moderate: transitions
p_moderate_to_mild_vec_no_int <- rep(p_moderate_to_mild, len)
p_moderate_to_severe_vec_no_int <- rep(p_moderate_to_severe, len)
p_moderate_to_moderate_vec_no_int <- 1 - (p_moderate_to_mild_vec_no_int + p_moderate_to_severe_vec_no_int)

# Severe: transitions
p_severe_to_moderate_vec_no_int <- rep(p_severe_to_moderate, len)
p_severe_to_severe_vec_no_int <- 1 - p_severe_to_moderate_vec_no_int

# --- Intervention Strategy ---

# Apply DMT effect: RR = 0.75 for relevant transitions

# AD-MCI to Mild
rate_ad_mci_to_mild <- -log(1 - p_ad_mci_to_mild)
rate_ad_mci_to_mild_adj <- rate_ad_mci_to_mild * rr_ad_mci_to_mild
p_ad_mci_to_mild_vec_int <- rep(1 - exp(-rate_ad_mci_to_mild_adj), len)
p_ad_mci_to_ad_mci_vec_int <- 1 - p_ad_mci_to_mild_vec_int

# Mild to Moderate
rate_mild_to_moderate <- -log(1 - p_mild_to_moderate)
rate_mild_to_moderate_adj <- rate_mild_to_moderate * rr_mild_to_moderate
p_mild_to_moderate_vec_int <- rep(1 - exp(-rate_mild_to_moderate_adj), len)

# Mild to Severe remains unchanged
p_mild_to_severe_vec_int <- rep(p_mild_to_severe, len)

# Staying in Mild
p_mild_to_mild_vec_int <- 1 - (p_mild_to_moderate_vec_int + p_mild_to_severe_vec_int)

# Other transitions are unaffected by intervention
p_moderate_to_mild_vec_int <- p_moderate_to_mild_vec_no_int
p_moderate_to_severe_vec_int <- p_moderate_to_severe_vec_no_int
p_moderate_to_moderate_vec_int <- p_moderate_to_moderate_vec_no_int

p_severe_to_moderate_vec_int <- p_severe_to_moderate_vec_no_int
p_severe_to_severe_vec_int <- p_severe_to_severe_vec_no_int
# =============================
# Prompt 4A: Initialize Transition Probability Matrix
# =============================

build_transition_matrix <- function(strat) {
  # Define state index mapping
  state_names <- c("ad_mci", "mild", "moderate", "severe", "death")
  n_states <- length(state_names)
  tpm <- array(0, dim = c(n_states, n_states, n_cycles),
               dimnames = list(state_names, state_names, NULL))
  
  for (cycle in 1:n_cycles) {
    
    # Transition to death for all non-death states using prompt 2 data
    age_index <- cycle  # Assume starting at age 60, 1 year per cycle
    current_age <- start_age + cycle - 1
    death_prob <- mortality_df$prob_death_ad_mci[mortality_df$age == current_age]
    
    tpm["ad_mci", "death", cycle]   <- death_prob
    tpm["mild", "death", cycle]     <- mortality_df$prob_death_mild[mortality_df$age == current_age]
    tpm["moderate", "death", cycle] <- mortality_df$prob_death_moderate[mortality_df$age == current_age]
    tpm["severe", "death", cycle]   <- mortality_df$prob_death_severe[mortality_df$age == current_age]
    tpm["death", "death", cycle]    <- 1  # Absorbing state
    
    # === Begin PROMPT 4B insert ===
    
    # Get mortality probabilities at current age
    p_death_ad_mci   <- mortality_df$prob_death_ad_mci[mortality_df$age == current_age]
    p_death_mild     <- mortality_df$prob_death_mild[mortality_df$age == current_age]
    p_death_moderate <- mortality_df$prob_death_moderate[mortality_df$age == current_age]
    p_death_severe   <- mortality_df$prob_death_severe[mortality_df$age == current_age]
    
    if (strat == "no_intervention") {
      # AD-MCI transitions
      tpm["ad_mci", "mild", cycle]   <- p_ad_mci_to_mild_vec_no_int[cycle] * (1 - p_death_ad_mci)
      tpm["ad_mci", "ad_mci", cycle] <- p_ad_mci_to_ad_mci_vec_no_int[cycle] * (1 - p_death_ad_mci)
      
      # Mild transitions
      tpm["mild", "moderate", cycle] <- p_mild_to_moderate_vec_no_int[cycle] * (1 - p_death_mild)
      tpm["mild", "severe", cycle]   <- p_mild_to_severe_vec_no_int[cycle]   * (1 - p_death_mild)
      tpm["mild", "mild", cycle]     <- p_mild_to_mild_vec_no_int[cycle]     * (1 - p_death_mild)
      
    } else if (strat == "intervention") {
      # AD-MCI transitions
      tpm["ad_mci", "mild", cycle]   <- p_ad_mci_to_mild_vec_int[cycle] * (1 - p_death_ad_mci)
      tpm["ad_mci", "ad_mci", cycle] <- p_ad_mci_to_ad_mci_vec_int[cycle] * (1 - p_death_ad_mci)
      
      # Mild transitions
      tpm["mild", "moderate", cycle] <- p_mild_to_moderate_vec_int[cycle] * (1 - p_death_mild)
      tpm["mild", "severe", cycle]   <- p_mild_to_severe_vec_int[cycle]   * (1 - p_death_mild)
      tpm["mild", "mild", cycle]     <- p_mild_to_mild_vec_int[cycle]     * (1 - p_death_mild)
    }
    
    # Moderate transitions (same in both strategies)
    tpm["moderate", "mild", cycle]     <- p_moderate_to_mild_vec_no_int[cycle] * (1 - p_death_moderate)
    tpm["moderate", "severe", cycle]   <- p_moderate_to_severe_vec_no_int[cycle] * (1 - p_death_moderate)
    tpm["moderate", "moderate", cycle] <- p_moderate_to_moderate_vec_no_int[cycle] * (1 - p_death_moderate)
    
    # Severe transitions (same in both strategies)
    tpm["severe", "moderate", cycle] <- p_severe_to_moderate_vec_no_int[cycle] * (1 - p_death_severe)
    tpm["severe", "severe", cycle]   <- p_severe_to_severe_vec_no_int[cycle] * (1 - p_death_severe)
    
    # === End PROMPT 4B insert ===
    
    # Normalize rows to ensure row sums = 1
    for (state in state_names) {
      row_sum <- sum(tpm[state, , cycle])
      if (row_sum > 0 && abs(row_sum - 1) > 1e-8) {
        tpm[state, , cycle] <- tpm[state, , cycle] / row_sum
      }
    }
  }
  
  return(tpm)
}
# =============================
# Prompt 5: Annual Societal Cost by Age and State
# =============================

# Define age range
ages <- 60:100

# Compute costs per state using provided GLM equations
cost_ad_mci <- 37.961 * ages^2 - 1234.5 * ages + 124934
cost_mild <- 49.314 * ages^2 - 1828.7 * ages + 166239
cost_moderate <- 61.412 * ages^2 - 2086.3 * ages + 205578
cost_severe <- 131670 * exp(0.0176 * ages)
cost_death <- rep(0, length(ages))

# Combine into a data frame
cost_by_age_state <- data.frame(
  age = ages,
  ad_mci = cost_ad_mci,
  mild = cost_mild,
  moderate = cost_moderate,
  severe = cost_severe,
  death = cost_death
)

# Result: cost_by_age_state has rows for age 60–100 and columns for each cognitive state


# =============================
# Prompt 6A: Markov Model Structure
# =============================

run_markov_model <- function(strat) {
  # State names
  states <- c("ad_mci", "mild", "moderate", "severe", "death")
  n_states <- length(states)
  
  # Initialize transition matrix
  tpm <- build_transition_matrix(strat)
  
  # Initialize state trace matrix
  trace <- matrix(0, nrow = n_cycles + 1, ncol = n_states)
  colnames(trace) <- states
  trace[1, "ad_mci"] <- cohort_size
  
  # Initialize outcome trackers
  qalys <- numeric(n_cycles)
  costs <- numeric(n_cycles)
  pys <- matrix(0, nrow = n_cycles, ncol = 4)
  colnames(pys) <- c("ad_mci", "mild", "moderate", "severe")
  deaths_total <- numeric(n_cycles)
  deaths_ad_mci <- numeric(n_cycles)
  
  for (cycle in 1:n_cycles) {
    # Update cohort distribution
    trace[cycle + 1, ] <- trace[cycle, ] %*% tpm[, , cycle]
    
    # Compute current age
    current_age <- start_age + cycle - 1
    
    # QALYs calculation
    qalys[cycle] <- sum(
      trace[cycle, "ad_mci"]   * u_ad_mci(current_age) +
        trace[cycle, "mild"]     * u_mild(current_age) +
        trace[cycle, "moderate"] * u_moderate(current_age) +
        trace[cycle, "severe"]   * u_severe(current_age)
    )
    
    # Cost calculation
    age_row <- cost_by_age_state[cost_by_age_state$age == current_age, ]
    cost_cycle <- sum(
      trace[cycle, "ad_mci"]   * age_row$ad_mci +
        trace[cycle, "mild"]     * age_row$mild +
        trace[cycle, "moderate"] * age_row$moderate +
        trace[cycle, "severe"]   * age_row$severe
    )
    
    # Add DMT cost for intervention strategy
    if (strat == "intervention") {
      cost_cycle <- cost_cycle +
        (trace[cycle, "ad_mci"] + trace[cycle, "mild"]) * dmt_cost_per_year
    }
    
    costs[cycle] <- cost_cycle
    
    # Person-years (non-death states)
    pys[cycle, "ad_mci"]   <- trace[cycle, "ad_mci"]
    pys[cycle, "mild"]     <- trace[cycle, "mild"]
    pys[cycle, "moderate"] <- trace[cycle, "moderate"]
    pys[cycle, "severe"]   <- trace[cycle, "severe"]
    
    # Total deaths this cycle
    deaths_total[cycle] <- trace[cycle + 1, "death"] - trace[cycle, "death"]
    
    # Cumulative deaths from AD-MCI
    deaths_ad_mci[cycle] <- trace[cycle, "ad_mci"] * tpm["ad_mci", "death", cycle]
  }
  
  # Apply half-cycle correction (QALYs, costs, PYs)
  qalys_hcc <- 0.5 * (qalys[-n_cycles] + qalys[-1])
  costs_hcc <- 0.5 * (costs[-n_cycles] + costs[-1])
  pys_hcc <- 0.5 * (pys[-n_cycles, ] + pys[-1, ])
  trace_hcc <- trace[-(n_cycles + 1), ]  # remove final row (post-last transition)
  
  # Apply discounting
  discount_factors <- 1 / ((1 + discount_rate) ^ (0:(n_cycles - 2)))
  qalys_discounted <- sum(qalys_hcc * discount_factors) / cohort_size
  costs_discounted <- sum(costs_hcc * discount_factors) / cohort_size
  
  # Return results
  return(list(
    qalys = qalys_discounted,
    final_costs = costs_discounted,
    pys_ad_mci = sum(pys_hcc[, "ad_mci"]) / cohort_size,
    pys_mild = sum(pys_hcc[, "mild"]) / cohort_size,
    pys_mod = sum(pys_hcc[, "moderate"]) / cohort_size,
    pys_sev = sum(pys_hcc[, "severe"]) / cohort_size,
    deaths_total = sum(deaths_total),
    deaths_ad_mci = sum(deaths_ad_mci)
  ))
}

# Run the model for both strategies
results <- list()
results$no_intervention <- run_markov_model("no_intervention")
results$intervention <- run_markov_model("intervention")
```
