## Supplementary material for "Replicating Health-Economic Simulation Models for Alzheimer’s Disease Using Artificial Intelligence": final R scripts: Wimo_O5.R

```
#===========================
# Input Parameters from Wimo et al. (2020)
#===========================

# State names
states <- c("ad_mci", "mild", "moderate", "severe", "death")

# Strategy names
strategies <- c("no_intervention", "intervention")

# Model settings
start_age <- 60
n_cycles <- 40
cycle_length <- 1
discount_rate <- 0.03
cohort_size <- 100000

# Base case: Annual transition probabilities (from Supplementary Table 3)
p.mild_to_mild <- 0.706
p.mild_to_moderate <- 0.293
p.mild_to_severe <- 0.001

p.moderate_to_mild <- 0.087
p.moderate_to_moderate <- 0.803
p.moderate_to_severe <- 0.109

p.severe_to_moderate <- 0.196
p.severe_to_severe <- 0.804

# Progression risk from AD-MCI to AD-dementia (fixed)
p_ad_mci_to_mild <- 0.206  # derived from 3-year 50% risk: 1 - (1 - 0.5)^(1/3)

# Mortality is age- and severity-specific, to be processed in later prompt

# DMT treatment cost (hypothetical annual cost in SEK)
cost_dmt <- 50000

# Hazard ratios for mortality (from Supplementary Table 4, vs. very mild reference)
hr_mild <- 1.318
hr_moderate <- 2.419
hr_severe <- 4.267

# Relative risks for progression under intervention
rr_ad_mci_to_mild <- 0.75  # 25% reduction
rr_mild_to_moderate <- 0.75  # 25% reduction

#===========================
# Utility Functions (from Supplementary Table 6 Regression Equations)
#===========================

u_ad_mci <- function(age) {
  return(-0.002464 * age + 0.962679)
}

u_mild <- function(age) {
  return(-0.0025 * age + 0.8293)
}

u_moderate <- function(age) {
  return(-4e-07 * age^2 - 0.0024 * age + 0.5828)
}

u_severe <- function(age) {
  return(-4e-07 * age^2 - 0.0025 * age + 0.4337)
}

u_death <- function(age) {
  return(0)
}
#===========================
# Load and Process Mortality Rate Data
#===========================

# Load mortality rates and weights
mortality_rates <- read.csv("mortalityRates_Swedish_2016.csv", stringsAsFactors = FALSE)
weights <- read.csv("weightsMortalityRates_Swedish_2016.csv", stringsAsFactors = FALSE)

# Create a mapping of age ranges to individual ages
age_map <- list(
  "60-64" = 60:64,
  "65-69" = 65:69,
  "70-74" = 70:74,
  "75-79" = 75:79,
  "80-84" = 80:84,
  "85-89" = 85:89,
  "90+"   = 90:99
)

# Initialize vectors to store expanded data
age_expanded <- c()
rate_male <- c()
rate_female <- c()

# Expand grouped rates to individual ages
for (i in 1:nrow(mortality_rates)) {
  age_range <- mortality_rates$age[i]
  ages <- age_map[[age_range]]
  age_expanded <- c(age_expanded, ages)
  rate_male <- c(rate_male, rep(mortality_rates$male[i], length(ages)))
  rate_female <- c(rate_female, rep(mortality_rates$female[i], length(ages)))
}

# Create a data frame for expanded rates
expanded_rates <- data.frame(age = age_expanded, rate_male = rate_male, rate_female = rate_female)

# Merge with weights
weights$age <- as.integer(weights$age)
merged <- merge(expanded_rates, weights, by = "age")

# Compute weighted average mortality rate per age
merged$rate_avg <- (merged$rate_male * merged$weight_male + merged$rate_female * merged$weight_female) /
  (merged$weight_male + merged$weight_female)

# Compute death probabilities by severity
merged$prob_death_ad_mci    <- 1 - exp(-merged$rate_avg)                      # HR = 1 (baseline)
merged$prob_death_mild      <- 1 - exp(-merged$rate_avg * hr_mild)
merged$prob_death_moderate  <- 1 - exp(-merged$rate_avg * hr_moderate)
merged$prob_death_severe    <- 1 - exp(-merged$rate_avg * hr_severe)

# Select and reorder relevant columns
mortality_df <- merged[, c("age", "rate_avg", "prob_death_ad_mci", 
                           "prob_death_mild", "prob_death_moderate", "prob_death_severe")]

# Final output: mortality_df (ages 60–99)
#===========================
# Time-Dependent Transition Probabilities
#===========================

# Number of cycles
cycles <- n_cycles

#---- No Intervention ----#

# From ad_mci
p_ad_mci_to_mild_noint <- rep(p_ad_mci_to_mild, cycles)
p_ad_mci_to_ad_mci_noint <- 1 - p_ad_mci_to_mild_noint

# From mild
p_mild_to_moderate_noint <- rep(p.mild_to_moderate, cycles)
p_mild_to_severe_noint <- rep(p.mild_to_severe, cycles)
p_mild_to_mild_noint <- 1 - p_mild_to_moderate_noint - p_mild_to_severe_noint

# From moderate
p_moderate_to_mild_noint <- rep(p.moderate_to_mild, cycles)
p_moderate_to_severe_noint <- rep(p.moderate_to_severe, cycles)
p_moderate_to_moderate_noint <- 1 - p_moderate_to_mild_noint - p_moderate_to_severe_noint

# From severe
p_severe_to_moderate_noint <- rep(p.severe_to_moderate, cycles)
p_severe_to_severe_noint <- 1 - p_severe_to_moderate_noint

#---- Intervention ----#

# AD-MCI transition adjustment
rate_ad_mci_to_mild <- -log(1 - p_ad_mci_to_mild)
adjusted_rate_ad_mci_to_mild <- rate_ad_mci_to_mild * rr_ad_mci_to_mild
p_ad_mci_to_mild_int <- rep(1 - exp(-adjusted_rate_ad_mci_to_mild), cycles)
p_ad_mci_to_ad_mci_int <- 1 - p_ad_mci_to_mild_int

# Mild transition adjustment
rate_mild_to_moderate <- -log(1 - p.mild_to_moderate)
adjusted_rate_mild_to_moderate <- rate_mild_to_moderate * rr_mild_to_moderate
p_mild_to_moderate_int <- rep(1 - exp(-adjusted_rate_mild_to_moderate), cycles)
p_mild_to_severe_int <- rep(p.mild_to_severe, cycles)  # unchanged
p_mild_to_mild_int <- 1 - p_mild_to_moderate_int - p_mild_to_severe_int

# From moderate (unchanged)
p_moderate_to_mild_int <- p_moderate_to_mild_noint
p_moderate_to_severe_int <- p_moderate_to_severe_noint
p_moderate_to_moderate_int <- p_moderate_to_moderate_noint

# From severe (unchanged)
p_severe_to_moderate_int <- p_severe_to_moderate_noint
p_severe_to_severe_int <- p_severe_to_severe_noint
#===========================
# Build Transition Matrix Function
#===========================

build_transition_matrix <- function(strat) {
  
  # Define state names
  states <- c("ad_mci", "mild", "moderate", "severe", "death")
  n_states <- length(states)
  
  # Initialize transition array: [from_state, to_state, cycle]
  tpm <- array(0, dim = c(n_states, n_states, n_cycles),
               dimnames = list(from = states, to = states, cycle = 1:n_cycles))
  
  # Loop over cycles to populate transitions
  for (cycle in 1:n_cycles) {
    
    age <- start_age + cycle - 1
    
    # Get mortality probabilities for current age
    death_probs <- mortality_df[mortality_df$age == age, ]
    
    # Assign death transition probabilities
    tpm["ad_mci", "death", cycle]    <- death_probs$prob_death_ad_mci
    tpm["mild", "death", cycle]      <- death_probs$prob_death_mild
    tpm["moderate", "death", cycle]  <- death_probs$prob_death_moderate
    tpm["severe", "death", cycle]    <- death_probs$prob_death_severe
    tpm["death", "death", cycle]     <- 1  # Absorbing state
    
    #================================================
    # Transitions from ad_mci
    #================================================
    if (strat == "no_intervention") {
      p_mci_to_mild <- p_ad_mci_to_mild_noint[cycle]
      p_mci_to_mci <- p_ad_mci_to_ad_mci_noint[cycle]
    } else if (strat == "intervention") {
      p_mci_to_mild <- p_ad_mci_to_mild_int[cycle]
      p_mci_to_mci <- p_ad_mci_to_ad_mci_int[cycle]
    }
    tpm["ad_mci", "mild", cycle] <- p_mci_to_mild * (1 - tpm["ad_mci", "death", cycle])
    tpm["ad_mci", "ad_mci", cycle] <- p_mci_to_mci * (1 - tpm["ad_mci", "death", cycle])
    
    #================================================
    # Transitions from mild
    #================================================
    if (strat == "no_intervention") {
      p_mild_mod <- p_mild_to_moderate_noint[cycle]
      p_mild_sev <- p_mild_to_severe_noint[cycle]
      p_mild_mild <- p_mild_to_mild_noint[cycle]
    } else if (strat == "intervention") {
      p_mild_mod <- p_mild_to_moderate_int[cycle]
      p_mild_sev <- p_mild_to_severe_int[cycle]
      p_mild_mild <- p_mild_to_mild_int[cycle]
    }
    tpm["mild", "moderate", cycle] <- p_mild_mod * (1 - tpm["mild", "death", cycle])
    tpm["mild", "severe", cycle] <- p_mild_sev * (1 - tpm["mild", "death", cycle])
    tpm["mild", "mild", cycle] <- p_mild_mild * (1 - tpm["mild", "death", cycle])
    
    #================================================
    # Transitions from moderate
    #================================================
    tpm["moderate", "mild", cycle] <- p_moderate_to_mild_noint[cycle] * (1 - tpm["moderate", "death", cycle])
    tpm["moderate", "severe", cycle] <- p_moderate_to_severe_noint[cycle] * (1 - tpm["moderate", "death", cycle])
    tpm["moderate", "moderate", cycle] <- p_moderate_to_moderate_noint[cycle] * (1 - tpm["moderate", "death", cycle])
    
    #================================================
    # Transitions from severe
    #================================================
    tpm["severe", "moderate", cycle] <- p_severe_to_moderate_noint[cycle] * (1 - tpm["severe", "death", cycle])
    tpm["severe", "severe", cycle] <- p_severe_to_severe_noint[cycle] * (1 - tpm["severe", "death", cycle])
    
    # Normalize rows (if row sum differs from 1)
    for (state in states[1:4]) {  # exclude death row
      row_sum <- sum(tpm[state, , cycle])
      if (abs(row_sum - 1) > 1e-8) {
        tpm[state, , cycle] <- tpm[state, , cycle] / row_sum
      }
    }
  }
  
  return(tpm)
}
#===========================
# Annual Societal Per-Person Costs by Age and Cognitive State
#===========================

# Define age range
ages <- 60:100

# Compute costs using fitted equations
cost_ad_mci <- 37.961 * ages^2 - 1234.5 * ages + 124934
cost_mild <- 49.314 * ages^2 - 1828.7 * ages + 166239
cost_moderate <- 61.412 * ages^2 - 2086.3 * ages + 205578
cost_severe <- 131670 * exp(0.0176 * ages)
cost_death <- rep(0, length(ages))

# Create data frame
costs_df <- data.frame(
  age = ages,
  ad_mci = cost_ad_mci,
  mild = cost_mild,
  moderate = cost_moderate,
  severe = cost_severe,
  death = cost_death
)
#===========================
# Run Markov Model Function
#===========================

run_markov_model <- function(strat) {
  
  # Define state names
  states <- c("ad_mci", "mild", "moderate", "severe", "death")
  n_states <- length(states)
  
  # Build transition matrix for strategy
  tpm <- build_transition_matrix(strat)
  
  # Initialize trace matrix [cycle x state]
  trace <- matrix(0, nrow = n_cycles + 1, ncol = n_states,
                  dimnames = list(cycle = 0:n_cycles, state = states))
  trace[1, "ad_mci"] <- cohort_size
  
  # Initialize outcome trackers
  qalys <- numeric(n_cycles)
  costs <- numeric(n_cycles)
  pys <- matrix(0, nrow = n_cycles, ncol = 4)
  colnames(pys) <- c("pys_ad_mci", "pys_mild", "pys_mod", "pys_sev")
  deaths_total <- numeric(n_cycles)
  deaths_ad_mci <- numeric(n_cycles)
  
  for (cycle in 1:n_cycles) {
    
    # Age in this cycle
    age <- start_age + cycle - 1
    
    # Update trace for next cycle
    trace[cycle + 1, ] <- trace[cycle, ] %*% tpm[ , , cycle]
    
    # Get current state distribution
    current_state <- trace[cycle, ]
    
    # QALY calculation using utility functions
    qalys[cycle] <- current_state["ad_mci"] * u_ad_mci(age) +
      current_state["mild"]   * u_mild(age) +
      current_state["moderate"] * u_moderate(age) +
      current_state["severe"] * u_severe(age)
    
    # Cost calculation using age-specific costs
    costs_this_age <- costs_df[costs_df$age == age, ]
    costs[cycle] <- current_state["ad_mci"] * costs_this_age$ad_mci +
      current_state["mild"]   * costs_this_age$mild +
      current_state["moderate"] * costs_this_age$moderate +
      current_state["severe"] * costs_this_age$severe
    
    # Add DMT treatment costs in intervention strategy
    if (strat == "intervention") {
      costs[cycle] <- costs[cycle] + (current_state["ad_mci"] + current_state["mild"]) * cost_dmt
    }
    
    # Store person-years
    pys[cycle, "pys_ad_mci"] <- current_state["ad_mci"]
    pys[cycle, "pys_mild"]   <- current_state["mild"]
    pys[cycle, "pys_mod"]    <- current_state["moderate"]
    pys[cycle, "pys_sev"]    <- current_state["severe"]
    
    # Track total deaths
    deaths_total[cycle] <- trace[cycle + 1, "death"] - trace[cycle, "death"]
    
    # Track deaths from AD-MCI
    deaths_ad_mci[cycle] <- current_state["ad_mci"] * tpm["ad_mci", "death", cycle]
  }
  
  # Half-cycle correction (average adjacent cycles and drop last)
  qalys <- 0.5 * (qalys[-n_cycles] + qalys[-1])
  costs <- 0.5 * (costs[-n_cycles] + costs[-1])
  pys <- 0.5 * (pys[-n_cycles, ] + pys[-1, ])
  trace <- trace[-(n_cycles + 1), ]  # remove last row
  
  # Discounting
  discount_factors <- 1 / ((1 + discount_rate) ^ (1:(n_cycles - 1)))
  qalys_disc <- sum(qalys * discount_factors) / cohort_size
  costs_disc <- sum(costs * discount_factors) / cohort_size
  
  # Return named list
  return(list(
    qalys = qalys_disc,
    final_costs = costs_disc,
    pys_ad_mci = sum(pys[, "pys_ad_mci"]) / cohort_size,
    pys_mild = sum(pys[, "pys_mild"]) / cohort_size,
    pys_mod = sum(pys[, "pys_mod"]) / cohort_size,
    pys_sev = sum(pys[, "pys_sev"]) / cohort_size,
    deaths_total = sum(deaths_total),
    deaths_ad_mci = sum(deaths_ad_mci)
  ))
}

#===========================
# Run Model for Both Strategies
#===========================

results <- list()
results$no_intervention <- run_markov_model("no_intervention")
results$intervention <- run_markov_model("intervention")
```
