## Supplementary material for "Replicating Health-Economic Simulation Models for Alzheimer’s Disease Using Artificial Intelligence": final R scripts: Wimo_O6.R

```
# Health Economic Markov Model Parameters from Wimo et al. (2020)

# --- Descriptive Parameters ---
state_names <- c("ad_mci", "mild", "moderate", "severe", "death")
strategy_names <- c("no_intervention", "intervention")

# --- Model Setup Parameters ---
start_age <- 60
cohort_size <- 100000
n_cycles <- 40
cycle_length <- 1
discount_rate <- 0.03
cost_year <- 2016

# --- Disease Progression Probabilities (Base Case, No Time Dependence) ---
# Annual transition probability from AD-MCI to mild AD
p.ad_mci_to_mild <- 0.206

# Transition probabilities within dementia states (from Supplementary Table 3)
p.mild_to_mild <- 0.706
p.mild_to_moderate <- 0.293
p.mild_to_severe <- 0.001

p.moderate_to_mild <- 0.087
p.moderate_to_moderate <- 0.803
p.moderate_to_severe <- 0.109

p.severe_to_moderate <- 0.196
p.severe_to_severe <- 0.804

# Mortality hazard ratios from Supplementary Table 4 (relative to very mild)
hr_mild <- 1.318
hr_moderate <- 2.419
hr_severe <- 4.267

# --- Relative Risks and Hazard Ratios for Intervention Effect (Base Case) ---
# Applied in the intervention strategy:
# 25% risk reduction for MCI to mild
rr.ad_mci_to_mild <- 0.75

# 25% risk reduction for mild to moderate
rr.mild_to_moderate <- 0.75

# --- Treatment Cost for Hypothetical DMT (Base Case) ---
dmt_annual_cost <- 50000  # in SEK

# --- Utility Functions (QALY estimates) ---
u_ad_mci <- function(age) {
  -0.002464 * age + 0.962679
}

u_mild <- function(age) {
  -0.0025 * age + 0.8293
}

u_moderate <- function(age) {
  -4e-07 * age^2 - 0.0024 * age + 0.5828
}

u_severe <- function(age) {
  -4e-07 * age^2 - 0.0025 * age + 0.4337
}

u_death <- function(age) {
  return(0)
}
# --- Load mortality rates and weights ---
mortality_rates <- read.csv("mortalityRates_Swedish_2016.csv", stringsAsFactors = FALSE)
weights <- read.csv("weightsMortalityRates_Swedish_2016.csv", stringsAsFactors = FALSE)

# --- Define mapping of age group to individual ages ---
age_group_mapping <- list(
  "60-64" = 60:64,
  "65-69" = 65:69,
  "70-74" = 70:74,
  "75-79" = 75:79,
  "80-84" = 80:84,
  "85-89" = 85:89,
  "90+"   = 90:99
)

# --- Expand grouped mortality rates to individual ages ---
expanded_rates <- data.frame()

for (group in names(age_group_mapping)) {
  ages <- age_group_mapping[[group]]
  rate_male <- mortality_rates[mortality_rates$age == group, "male"]
  rate_female <- mortality_rates[mortality_rates$age == group, "female"]
  
  temp <- data.frame(
    age = ages,
    rate_male = rep(rate_male, length(ages)),
    rate_female = rep(rate_female, length(ages))
  )
  
  expanded_rates <- rbind(expanded_rates, temp)
}

# --- Merge with weights ---
merged <- merge(expanded_rates, weights, by = "age")

# --- Compute sex-weighted average mortality rate ---
merged$rate_avg <- with(merged, (rate_male * weight_male + rate_female * weight_female) / (weight_male + weight_female))

# --- Compute probability of death for each state using: p_death = 1 - exp(-rate * HR) ---
merged$prob_death_ad_mci <- 1 - exp(-merged$rate_avg * 1)  # HR = 1 for AD-MCI (general population)
merged$prob_death_mild <- 1 - exp(-merged$rate_avg * hr_mild)
merged$prob_death_moderate <- 1 - exp(-merged$rate_avg * hr_moderate)
merged$prob_death_severe <- 1 - exp(-merged$rate_avg * hr_severe)

# --- Final output: data frame with relevant columns ---
mortality_table <- merged[, c("age", "rate_avg", "prob_death_ad_mci", "prob_death_mild", "prob_death_moderate", "prob_death_severe")]
# --- Transition probabilities for no intervention strategy (length = n_cycles) ---

# AD-MCI
p_ad_mci_to_mild_ni <- rep(p.ad_mci_to_mild, n_cycles)
p_ad_mci_to_ad_mci_ni <- 1 - p_ad_mci_to_mild_ni

# Mild
p_mild_to_moderate_ni <- rep(p.mild_to_moderate, n_cycles)
p_mild_to_severe_ni <- rep(p.mild_to_severe, n_cycles)
p_mild_to_mild_ni <- 1 - (p_mild_to_moderate_ni + p_mild_to_severe_ni)

# Moderate
p_moderate_to_mild_ni <- rep(p.moderate_to_mild, n_cycles)
p_moderate_to_severe_ni <- rep(p.moderate_to_severe, n_cycles)
p_moderate_to_moderate_ni <- 1 - (p_moderate_to_mild_ni + p_moderate_to_severe_ni)

# Severe
p_severe_to_moderate_ni <- rep(p.severe_to_moderate, n_cycles)
p_severe_to_severe_ni <- 1 - p_severe_to_moderate_ni

# --- Transition probabilities for intervention strategy (adjusted) ---

# AD-MCI
rate_ad_mci_to_mild <- -log(1 - p.ad_mci_to_mild)
adjusted_rate_ad_mci_to_mild <- rate_ad_mci_to_mild * rr.ad_mci_to_mild
p_ad_mci_to_mild_int <- rep(1 - exp(-adjusted_rate_ad_mci_to_mild), n_cycles)
p_ad_mci_to_ad_mci_int <- 1 - p_ad_mci_to_mild_int

# Mild
rate_mild_to_moderate <- -log(1 - p.mild_to_moderate)
adjusted_rate_mild_to_moderate <- rate_mild_to_moderate * rr.mild_to_moderate
p_mild_to_moderate_int <- rep(1 - exp(-adjusted_rate_mild_to_moderate), n_cycles)
p_mild_to_severe_int <- rep(p.mild_to_severe, n_cycles)
p_mild_to_mild_int <- 1 - (p_mild_to_moderate_int + p_mild_to_severe_int)

# Moderate (unchanged)
p_moderate_to_mild_int <- p_moderate_to_mild_ni
p_moderate_to_severe_int <- p_moderate_to_severe_ni
p_moderate_to_moderate_int <- p_moderate_to_moderate_ni

# Severe (unchanged)
p_severe_to_moderate_int <- p_severe_to_moderate_ni
p_severe_to_severe_int <- p_severe_to_severe_ni
# --- Transition probability matrix builder ---
build_transition_matrix <- function(strat) {
  
  # Define state indices for readability
  state_indices <- list(
    ad_mci = 1,
    mild = 2,
    moderate = 3,
    severe = 4,
    death = 5
  )
  
  # Initialize empty transition probability array: [from, to, cycle]
  tmat <- array(0, dim = c(length(state_names), length(state_names), n_cycles),
                dimnames = list(state_names, state_names, NULL))
  
  for (cycle in 1:n_cycles) {
    age <- start_age + cycle - 1  # current age
    
    # Probability of death from general population for AD-MCI (HR = 1)
    tmat["ad_mci", "death", cycle] <- mortality_table$prob_death_ad_mci[mortality_table$age == age]
    
    # Probability of death for Mild AD
    tmat["mild", "death", cycle] <- mortality_table$prob_death_mild[mortality_table$age == age]
    
    # Probability of death for Moderate AD
    tmat["moderate", "death", cycle] <- mortality_table$prob_death_moderate[mortality_table$age == age]
    
    # Probability of death for Severe AD
    tmat["severe", "death", cycle] <- mortality_table$prob_death_severe[mortality_table$age == age]
    
    # Death is an absorbing state
    tmat["death", "death", cycle] <- 1
    
    # --- Living state transitions (adjusted for survival probability) ---
    if (strat == "no_intervention") {
      # AD-MCI
      tmat["ad_mci", "mild", cycle] <- p_ad_mci_to_mild_ni[cycle] * (1 - tmat["ad_mci", "death", cycle])
      tmat["ad_mci", "ad_mci", cycle] <- p_ad_mci_to_ad_mci_ni[cycle] * (1 - tmat["ad_mci", "death", cycle])
      
      # Mild
      tmat["mild", "moderate", cycle] <- p_mild_to_moderate_ni[cycle] * (1 - tmat["mild", "death", cycle])
      tmat["mild", "severe", cycle] <- p_mild_to_severe_ni[cycle] * (1 - tmat["mild", "death", cycle])
      tmat["mild", "mild", cycle] <- p_mild_to_mild_ni[cycle] * (1 - tmat["mild", "death", cycle])
      
      # Moderate
      tmat["moderate", "mild", cycle] <- p_moderate_to_mild_ni[cycle] * (1 - tmat["moderate", "death", cycle])
      tmat["moderate", "severe", cycle] <- p_moderate_to_severe_ni[cycle] * (1 - tmat["moderate", "death", cycle])
      tmat["moderate", "moderate", cycle] <- p_moderate_to_moderate_ni[cycle] * (1 - tmat["moderate", "death", cycle])
      
      # Severe
      tmat["severe", "moderate", cycle] <- p_severe_to_moderate_ni[cycle] * (1 - tmat["severe", "death", cycle])
      tmat["severe", "severe", cycle] <- p_severe_to_severe_ni[cycle] * (1 - tmat["severe", "death", cycle])
      
    } else if (strat == "intervention") {
      # AD-MCI
      tmat["ad_mci", "mild", cycle] <- p_ad_mci_to_mild_int[cycle] * (1 - tmat["ad_mci", "death", cycle])
      tmat["ad_mci", "ad_mci", cycle] <- p_ad_mci_to_ad_mci_int[cycle] * (1 - tmat["ad_mci", "death", cycle])
      
      # Mild
      tmat["mild", "moderate", cycle] <- p_mild_to_moderate_int[cycle] * (1 - tmat["mild", "death", cycle])
      tmat["mild", "severe", cycle] <- p_mild_to_severe_int[cycle] * (1 - tmat["mild", "death", cycle])
      tmat["mild", "mild", cycle] <- p_mild_to_mild_int[cycle] * (1 - tmat["mild", "death", cycle])
      
      # Moderate
      tmat["moderate", "mild", cycle] <- p_moderate_to_mild_int[cycle] * (1 - tmat["moderate", "death", cycle])
      tmat["moderate", "severe", cycle] <- p_moderate_to_severe_int[cycle] * (1 - tmat["moderate", "death", cycle])
      tmat["moderate", "moderate", cycle] <- p_moderate_to_moderate_int[cycle] * (1 - tmat["moderate", "death", cycle])
      
      # Severe
      tmat["severe", "moderate", cycle] <- p_severe_to_moderate_int[cycle] * (1 - tmat["severe", "death", cycle])
      tmat["severe", "severe", cycle] <- p_severe_to_severe_int[cycle] * (1 - tmat["severe", "death", cycle])
    }
    
    # --- Normalize rows (ensure each row sums to 1) ---
    for (state in state_names) {
      row_sum <- sum(tmat[state, , cycle])
      if (row_sum > 0 && row_sum != 1) {
        tmat[state, , cycle] <- tmat[state, , cycle] / row_sum
      }
    }
  }
  
  return(tmat)
}
# --- Define age range ---
ages <- 60:100

# --- Define cost functions for each state ---
cost_ad_mci <- function(age) {
  37.961 * age^2 - 1234.5 * age + 124934
}

cost_mild <- function(age) {
  49.314 * age^2 - 1828.7 * age + 166239
}

cost_moderate <- function(age) {
  61.412 * age^2 - 2086.3 * age + 205578
}

cost_severe <- function(age) {
  131670 * exp(0.0176 * age)
}

cost_death <- function(age) {
  0
}

# --- Compute costs per age and cognitive state ---
cost_table <- data.frame(
  age = ages,
  ad_mci = sapply(ages, cost_ad_mci),
  mild = sapply(ages, cost_mild),
  moderate = sapply(ages, cost_moderate),
  severe = sapply(ages, cost_severe),
  death = sapply(ages, cost_death)
)
# --- Discount factor vector ---
discount_factors <- 1 / (1 + discount_rate)^(0:(n_cycles - 2))

# --- Run Markov simulation function ---
run_markov_model <- function(strat) {
  
  # Transition matrix for strategy
  tmat <- build_transition_matrix(strat)
  
  # Initialize trace matrix: rows = cycles, cols = states
  trace <- matrix(0, nrow = n_cycles, ncol = length(state_names))
  colnames(trace) <- state_names
  trace[1, "ad_mci"] <- cohort_size  # all start in AD-MCI
  
  # Initialize outputs
  qalys <- numeric(n_cycles)
  costs <- numeric(n_cycles)
  pys <- matrix(0, nrow = n_cycles, ncol = 4)
  colnames(pys) <- c("ad_mci", "mild", "moderate", "severe")
  deaths_total <- numeric(n_cycles)
  deaths_ad_mci <- numeric(n_cycles)
  
  for (cycle in 2:n_cycles) {
    # Update state vector using transition matrix
    trace[cycle, ] <- trace[cycle - 1, ] %*% tmat[, , cycle - 1]
    
    # Current age
    age <- start_age + cycle - 1
    
    # QALYs
    qalys[cycle] <- trace[cycle, "ad_mci"] * u_ad_mci(age) +
      trace[cycle, "mild"]   * u_mild(age) +
      trace[cycle, "moderate"] * u_moderate(age) +
      trace[cycle, "severe"] * u_severe(age)
    
    # Costs
    cost_age_row <- cost_table[cost_table$age == age, ]
    costs[cycle] <- trace[cycle, "ad_mci"]   * cost_age_row$ad_mci +
      trace[cycle, "mild"]     * cost_age_row$mild +
      trace[cycle, "moderate"] * cost_age_row$moderate +
      trace[cycle, "severe"]   * cost_age_row$severe
    if (strat == "intervention") {
      costs[cycle] <- costs[cycle] + (trace[cycle, "ad_mci"] + trace[cycle, "mild"]) * dmt_annual_cost
    }
    
    # Person-years
    pys[cycle, "ad_mci"]   <- trace[cycle, "ad_mci"]
    pys[cycle, "mild"]     <- trace[cycle, "mild"]
    pys[cycle, "moderate"] <- trace[cycle, "moderate"]
    pys[cycle, "severe"]   <- trace[cycle, "severe"]
    
    # Total deaths (change in death state)
    deaths_total[cycle] <- trace[cycle, "death"] - trace[cycle - 1, "death"]
    
    # Cumulative deaths from AD-MCI (transition from ad_mci to death)
    deaths_ad_mci[cycle] <- trace[cycle - 1, "ad_mci"] * tmat["ad_mci", "death", cycle - 1]
  }
  
  # --- Half-cycle correction ---
  qalys_hc <- 0.5 * (qalys[-n_cycles] + qalys[-1])
  costs_hc <- 0.5 * (costs[-n_cycles] + costs[-1])
  pys_hc <- 0.5 * (pys[-n_cycles, ] + pys[-1, ])
  
  # Remove last cycle from trace
  trace_hc <- trace[-n_cycles, ]
  
  # --- Apply discounting ---
  qalys_disc <- sum(qalys_hc * discount_factors) / cohort_size
  costs_disc <- sum(costs_hc * discount_factors) / cohort_size
  
  # --- Return named list of outcomes ---
  return(list(
    qalys = qalys_disc,
    final_costs = costs_disc,
    pys_ad_mci = sum(pys_hc[, "ad_mci"]) / cohort_size,
    pys_mild = sum(pys_hc[, "mild"]) / cohort_size,
    pys_mod = sum(pys_hc[, "moderate"]) / cohort_size,
    pys_sev = sum(pys_hc[, "severe"]) / cohort_size,
    deaths_total = sum(deaths_total),
    deaths_ad_mci = sum(deaths_ad_mci)
  ))
}

# --- Run model for both strategies ---
results <- list()
results$no_intervention <- run_markov_model("no_intervention")
results$intervention <- run_markov_model("intervention")
```
