## Supplementary material for "Replicating Health-Economic Simulation Models for Alzheimer’s Disease Using Artificial Intelligence": final R scripts: Wimo_O7.R

```
# Input Parameters for Markov Model (Wimo et al., 2020)

# -----------------------------
# State and Strategy Names
# -----------------------------
states <- c("ad_mci", "mild", "moderate", "severe", "death")
strategies <- c("no_intervention", "intervention")

# -----------------------------
# General Parameters
# -----------------------------
start_age <- 60
cycle_length <- 1  # in years
n_cycles <- 40
discount_rate <- 0.03
cohort_size <- 100000

# -----------------------------
# Disease Progression Probabilities (1-year)
# -----------------------------
# From AD-MCI
p.ad_mci_to_mild <- 0.206  # derived from 50% over 3 years: 1 - (1 - 0.5)^(1/3)
p.ad_mci_stay <- 1 - p.ad_mci_to_mild  # ignoring mortality here, will be accounted separately

# From Mild AD
p.mild_to_moderate <- 0.293
p.mild_stay <- 0.706
p.mild_to_severe <- 0.001

# From Moderate AD
p.moderate_to_severe <- 0.109
p.moderate_stay <- 0.803
p.moderate_to_mild <- 0.087

# From Severe AD
p.severe_stay <- 0.804
p.severe_to_moderate <- 0.196

# -----------------------------
# Hypothetical DMT Effectiveness (Intervention Arm)
# -----------------------------
rr.ad_mci_to_mild <- 0.75  # 25% risk reduction
rr.mild_to_moderate <- 0.75  # 25% risk reduction

# -----------------------------
# Hypothetical DMT Cost (Annual)
# -----------------------------
cost_dmt <- 50000  # SEK per year

# -----------------------------
# QALY Utility Functions (Based on Supplementary Table 6 Regressions)
# -----------------------------
u_ad_mci <- function(age) {
  -0.002464 * age + 0.962679
}

u_mild <- function(age) {
  -0.0025 * age + 0.8293
}

u_moderate <- function(age) {
  -4e-07 * age^2 - 0.0024 * age + 0.5828
}

u_severe <- function(age) {
  -4e-07 * age^2 - 0.0025 * age + 0.4337
}

u_death <- function(age) {
  0
}
# Load mortality rate and weight data
mort_rates <- read.csv("mortalityRates_Swedish_2016.csv", stringsAsFactors = FALSE)
weights <- read.csv("weightsMortalityRates_Swedish_2016.csv", stringsAsFactors = FALSE)

# Define age group mapping
age_groups <- list(
  "60-64" = 60:64,
  "65-69" = 65:69,
  "70-74" = 70:74,
  "75-79" = 75:79,
  "80-84" = 80:84,
  "85-89" = 85:89,
  "90+"   = 90:99
)

# Expand group-level mortality rates to individual ages
expanded_rates <- data.frame()

for (group in names(age_groups)) {
  ages <- age_groups[[group]]
  male_rate <- mort_rates$male[mort_rates$age == group]
  female_rate <- mort_rates$female[mort_rates$age == group]
  temp <- data.frame(
    age = ages,
    male_rate = rep(male_rate, length(ages)),
    female_rate = rep(female_rate, length(ages))
  )
  expanded_rates <- rbind(expanded_rates, temp)
}

# Merge expanded rates with weights
merged <- merge(expanded_rates, weights, by = "age")

# Compute sex-weighted average mortality rate
merged$rate_avg <- with(merged, male_rate * weight_male + female_rate * weight_female)

# Define hazard ratios from Wimo et al. 2020 (Supplementary Table 4)
hr_mild <- 1.318
hr_moderate <- 2.419
hr_severe <- 4.267

# Compute probabilities of death for each severity state
merged$prob_death_ad_mci <- 1 - exp(-merged$rate_avg * 1)
merged$prob_death_mild <- 1 - exp(-merged$rate_avg * hr_mild)
merged$prob_death_moderate <- 1 - exp(-merged$rate_avg * hr_moderate)
merged$prob_death_severe <- 1 - exp(-merged$rate_avg * hr_severe)

# Final data frame
mortality_df <- merged[, c("age", "rate_avg", "prob_death_ad_mci", "prob_death_mild", "prob_death_moderate", "prob_death_severe")]
# Initialize time-dependent transition probability vectors (length = n_cycles)
p_ad_mci_to_mild_ni <- rep(p.ad_mci_to_mild, n_cycles)
p_ad_mci_stay_ni <- rep(1 - p.ad_mci_to_mild, n_cycles)

p_mild_to_moderate_ni <- rep(p.mild_to_moderate, n_cycles)
p_mild_to_severe_ni <- rep(p.mild_to_severe, n_cycles)
p_mild_stay_ni <- rep(1 - (p.mild_to_moderate + p.mild_to_severe), n_cycles)

p_moderate_to_mild_ni <- rep(p.moderate_to_mild, n_cycles)
p_moderate_to_severe_ni <- rep(p.moderate_to_severe, n_cycles)
p_moderate_stay_ni <- rep(1 - (p.moderate_to_mild + p.moderate_to_severe), n_cycles)

p_severe_to_moderate_ni <- rep(p.severe_to_moderate, n_cycles)
p_severe_stay_ni <- rep(1 - p.severe_to_moderate, n_cycles)

# Intervention strategy — apply RR for transitions in "ad_mci" and "mild"
# Adjusted probabilities using rate transformation
p_ad_mci_to_mild_i <- rep({
  rate <- -log(1 - p.ad_mci_to_mild)
  adjusted_rate <- rate * rr.ad_mci_to_mild
  1 - exp(-adjusted_rate)
}, n_cycles)
p_ad_mci_stay_i <- rep(1 - p_ad_mci_to_mild_i[1], n_cycles)

p_mild_to_moderate_i <- rep({
  rate <- -log(1 - p.mild_to_moderate)
  adjusted_rate <- rate * rr.mild_to_moderate
  1 - exp(-adjusted_rate)
}, n_cycles)
p_mild_to_severe_i <- rep(p.mild_to_severe, n_cycles)  # unaffected
p_mild_stay_i <- rep(1 - (p_mild_to_moderate_i[1] + p.mild_to_severe), n_cycles)

# Unchanged from no intervention for moderate and severe
p_moderate_to_mild_i <- p_moderate_to_mild_ni
p_moderate_to_severe_i <- p_moderate_to_severe_ni
p_moderate_stay_i <- p_moderate_stay_ni

p_severe_to_moderate_i <- p_severe_to_moderate_ni
p_severe_stay_i <- p_severe_stay_ni
# Define function to build transition probability matrix
build_transition_matrix <- function(strat) {
  n_states <- length(states)
  trans_mat <- array(0, dim = c(n_states, n_states, n_cycles),
                     dimnames = list(from = states, to = states, cycle = 1:n_cycles))
  
  for (t in 1:n_cycles) {
    
    # Probabilities of death (precomputed in mortality_df)
    p_death_ad_mci <- mortality_df$prob_death_ad_mci[t]
    p_death_mild <- mortality_df$prob_death_mild[t]
    p_death_moderate <- mortality_df$prob_death_moderate[t]
    p_death_severe <- mortality_df$prob_death_severe[t]
    
    # Assign death transition probabilities explicitly
    trans_mat["ad_mci", "death", t] <- p_death_ad_mci
    trans_mat["mild", "death", t] <- p_death_mild
    trans_mat["moderate", "death", t] <- p_death_moderate
    trans_mat["severe", "death", t] <- p_death_severe
    trans_mat["death", "death", t] <- 1  # absorbing state
    
    # Transitions for 'ad_mci'
    if (strat == "no_intervention") {
      trans_mat["ad_mci", "mild", t]    <- p_ad_mci_to_mild_ni[t] * (1 - p_death_ad_mci)
      trans_mat["ad_mci", "ad_mci", t] <- p_ad_mci_stay_ni[t] * (1 - p_death_ad_mci)
    } else if (strat == "intervention") {
      trans_mat["ad_mci", "mild", t]    <- p_ad_mci_to_mild_i[t] * (1 - p_death_ad_mci)
      trans_mat["ad_mci", "ad_mci", t] <- p_ad_mci_stay_i[t] * (1 - p_death_ad_mci)
    }
    
    # Transitions for 'mild'
    if (strat == "no_intervention") {
      trans_mat["mild", "moderate", t] <- p_mild_to_moderate_ni[t] * (1 - p_death_mild)
      trans_mat["mild", "severe", t]   <- p_mild_to_severe_ni[t] * (1 - p_death_mild)
      trans_mat["mild", "mild", t]     <- p_mild_stay_ni[t] * (1 - p_death_mild)
    } else if (strat == "intervention") {
      trans_mat["mild", "moderate", t] <- p_mild_to_moderate_i[t] * (1 - p_death_mild)
      trans_mat["mild", "severe", t]   <- p_mild_to_severe_i[t] * (1 - p_death_mild)
      trans_mat["mild", "mild", t]     <- p_mild_stay_i[t] * (1 - p_death_mild)
    }
    
    # Transitions for 'moderate'
    trans_mat["moderate", "mild", t]     <- p_moderate_to_mild_i[t] * (1 - p_death_moderate)
    trans_mat["moderate", "severe", t]   <- p_moderate_to_severe_i[t] * (1 - p_death_moderate)
    trans_mat["moderate", "moderate", t] <- p_moderate_stay_i[t] * (1 - p_death_moderate)
    
    # Transitions for 'severe'
    trans_mat["severe", "moderate", t] <- p_severe_to_moderate_i[t] * (1 - p_death_severe)
    trans_mat["severe", "severe", t]   <- p_severe_stay_i[t] * (1 - p_death_severe)
    
    # Normalize rows if necessary
    for (s in states) {
      row_sum <- sum(trans_mat[s, , t])
      if (row_sum > 0 && row_sum != 1) {
        trans_mat[s, , t] <- trans_mat[s, , t] / row_sum
      }
    }
  }
  
  return(trans_mat)
}
# Define age range
ages <- 60:100

# Calculate costs per age for each cognitive state using the provided equations
cost_ad_mci <- 37.961 * ages^2 - 1234.5 * ages + 124934
cost_mild <- 49.314 * ages^2 - 1828.7 * ages + 166239
cost_moderate <- 61.412 * ages^2 - 2086.3 * ages + 205578
cost_severe <- 131670 * exp(0.0176 * ages)
cost_death <- rep(0, length(ages))

# Combine into a data frame
annual_costs_df <- data.frame(
  age = ages,
  ad_mci = cost_ad_mci,
  mild = cost_mild,
  moderate = cost_moderate,
  severe = cost_severe,
  death = cost_death
)
# Function to run Markov model for a given strategy
run_markov_model <- function(strat) {
  # Initialize transition matrix
  trans_mat <- build_transition_matrix(strat)
  
  # Initialize state trace matrix (rows = cycles, cols = states)
  state_trace <- matrix(0, nrow = n_cycles + 1, ncol = length(states), dimnames = list(0:n_cycles, states))
  state_trace[1, "ad_mci"] <- cohort_size  # all individuals start in ad_mci
  
  # Initialize outcome containers
  qalys <- numeric(n_cycles)
  costs <- numeric(n_cycles)
  pys <- matrix(0, nrow = n_cycles, ncol = 4, dimnames = list(NULL, c("ad_mci", "mild", "moderate", "severe")))
  deaths_total <- numeric(n_cycles)
  deaths_ad_mci <- numeric(n_cycles)
  
  # Discount factors (start from cycle 1 to n_cycles - 1 after half-cycle correction)
  discount_factors <- 1 / ((1 + discount_rate) ^ (0:(n_cycles - 2))) #Needed to replace the 1 by 2!!!!!!!!!!!!
  
  for (t in 1:n_cycles) {
    # Current age
    current_age <- start_age + (t - 1)
    
    # Project next state distribution using Markov transition matrix
    state_trace[t + 1, ] <- state_trace[t, ] %*% trans_mat[, , t]
    
    # QALYs: state occupancy × utility at current age
    qalys[t] <- state_trace[t, "ad_mci"] * u_ad_mci(current_age) +
      state_trace[t, "mild"]   * u_mild(current_age) +
      state_trace[t, "moderate"] * u_moderate(current_age) +
      state_trace[t, "severe"] * u_severe(current_age)
    
    # Costs: state occupancy × annual cost from precomputed table
    cost_row <- annual_costs_df[annual_costs_df$age == current_age, ]
    state_costs <- state_trace[t, "ad_mci"] * cost_row$ad_mci +
      state_trace[t, "mild"]   * cost_row$mild +
      state_trace[t, "moderate"] * cost_row$moderate +
      state_trace[t, "severe"] * cost_row$severe
    
    # Add DMT costs for intervention strategy in ad_mci and mild states
    if (strat == "intervention") {
      state_costs <- state_costs +
        cost_dmt * (state_trace[t, "ad_mci"] + state_trace[t, "mild"])
    }
    costs[t] <- state_costs
    
    # Person-years in each non-death state
    pys[t, "ad_mci"]   <- state_trace[t, "ad_mci"]
    pys[t, "mild"]     <- state_trace[t, "mild"]
    pys[t, "moderate"] <- state_trace[t, "moderate"]
    pys[t, "severe"]   <- state_trace[t, "severe"]
    
    # Total deaths in this cycle
    deaths_total[t] <- state_trace[t + 1, "death"] - state_trace[t, "death"]
    
    # Deaths from AD-MCI: occupancy × transition probability to death
    p_death_ad_mci <- mortality_df$prob_death_ad_mci[t]
    deaths_ad_mci[t] <- state_trace[t, "ad_mci"] * p_death_ad_mci
  }
  
  # Apply half-cycle correction
  qalys <- 0.5 * (qalys[-n_cycles] + qalys[-1])
  costs <- 0.5 * (costs[-n_cycles] + costs[-1])
  pys <- 0.5 * (pys[-n_cycles, ] + pys[-1, ])
  deaths_total <- deaths_total[-1]
  deaths_ad_mci <- deaths_ad_mci[-1]
  state_trace <- state_trace[-(n_cycles + 1), ]
  
  # Apply discounting
  qalys_discounted <- qalys * discount_factors
  costs_discounted <- costs * discount_factors
  
  # Return results
  return(list(
    qalys = sum(qalys_discounted) / cohort_size,
    final_costs = sum(costs_discounted) / cohort_size,
    pys_ad_mci = sum(pys[, "ad_mci"]) / cohort_size,
    pys_mild = sum(pys[, "mild"]) / cohort_size,
    pys_mod = sum(pys[, "moderate"]) / cohort_size,
    pys_sev = sum(pys[, "severe"]) / cohort_size,
    deaths_total = sum(deaths_total),
    deaths_ad_mci = sum(deaths_ad_mci)
  ))
}

# Run model for both strategies
results <- list()
results$no_intervention <- run_markov_model("no_intervention")
results$intervention <- run_markov_model("intervention")
```
