## Supplementary material for "Replicating Health-Economic Simulation Models for Alzheimer’s Disease Using Artificial Intelligence": final R scripts: Wimo_O8.R

```
# ===============================
# Wimo et al. (2020) - Input Parameters for Markov Model
# ===============================

# ---- State names ----
states <- c("ad_mci", "mild", "moderate", "severe", "death")

# ---- Strategy names ----
strategies <- c("no_intervention", "intervention")

# ---- Model settings ----
start_age <- 60
cohort_size <- 100000
cycle_length <- 1  # year
n_cycles <- 40
discount_rate <- 0.03

# ---- Treatment cost for DMT (hypothetical) ----
dmt_annual_cost <- 50000  # SEK

# ---- Hazard ratios for mortality (from Supplementary Table 4) ----
hr_mild <- 1.318
hr_moderate <- 2.419
hr_severe <- 4.267

# ---- Relative risks for intervention (25% reduction in base case) ----
rr_ad_mci_to_mild <- 0.75
rr_mild_to_moderate <- 0.75

# ---- Transition probabilities (base case, from Supplementary Table 3) ----
# From mild
p_mild_to_mild <- 0.706
p_mild_to_moderate <- 0.293
p_mild_to_severe <- 0.001

# From moderate
p_moderate_to_mild <- 0.087
p_moderate_to_moderate <- 0.803
p_moderate_to_severe <- 0.109

# From severe
p_severe_to_moderate <- 0.196
p_severe_to_severe <- 0.804

# From ad_mci to mild dementia (derived from annualized 3-year risk of 50%)
p_ad_mci_to_mild <- 1 - (1 - 0.50)^(1/3)  # = 0.206 (approx)

# ---- QALY utility functions ----
u_ad_mci <- function(age) {
  -0.002464 * age + 0.962679
}

u_mild <- function(age) {
  -0.0025 * age + 0.8293
}

u_moderate <- function(age) {
  -4e-07 * age^2 - 0.0024 * age + 0.5828
}

u_severe <- function(age) {
  -4e-07 * age^2 - 0.0025 * age + 0.4337
}

# Utility for death state
u_death <- 0
# ===============================
# Load and Process Mortality Rates
# ===============================

# ---- Load data ----
mortality_rates <- read.csv("mortalityRates_Swedish_2016.csv", stringsAsFactors = FALSE)
weights <- read.csv("weightsMortalityRates_Swedish_2016.csv", stringsAsFactors = FALSE)

# ---- Define age group mapping ----
age_groups <- list(
  "60-64" = 60:64,
  "65-69" = 65:69,
  "70-74" = 70:74,
  "75-79" = 75:79,
  "80-84" = 80:84,
  "85-89" = 85:89,
  "90+"   = 90:99
)

# ---- Expand grouped mortality rates to individual ages ----
expanded_rates <- data.frame(age = integer(), male = numeric(), female = numeric(), stringsAsFactors = FALSE)

for (i in 1:nrow(mortality_rates)) {
  group <- mortality_rates$age[i]
  ages <- age_groups[[group]]
  male_rate <- mortality_rates$male[i]
  female_rate <- mortality_rates$female[i]
  expanded <- data.frame(age = ages, male = rep(male_rate, length(ages)), female = rep(female_rate, length(ages)))
  expanded_rates <- rbind(expanded_rates, expanded)
}

# ---- Merge with weights ----
weights$age <- as.integer(weights$age)
merged <- merge(expanded_rates, weights, by = "age")

# ---- Compute weighted average mortality rate per age ----
merged$rate_avg <- (merged$male * merged$weight_male + merged$female * merged$weight_female) /
  (merged$weight_male + merged$weight_female)

# ---- Compute probability of death per severity state ----
merged$prob_death_ad_mci <- merged$rate_avg  # HR = 1 (general population)
merged$prob_death_mild <- 1 - exp(-merged$rate_avg * hr_mild)
merged$prob_death_moderate <- 1 - exp(-merged$rate_avg * hr_moderate)
merged$prob_death_severe <- 1 - exp(-merged$rate_avg * hr_severe)

# ---- Create final output dataframe ----
mortality_by_age <- merged[, c("age", "rate_avg", "prob_death_ad_mci", "prob_death_mild",
                               "prob_death_moderate", "prob_death_severe")]
# ===============================
# Time-Dependent Transition Probabilities
# ===============================

# ---- No intervention: AD-MCI ----
p_ad_mci_to_mild_ni <- rep(p_ad_mci_to_mild, n_cycles)
p_ad_mci_to_ad_mci_ni <- 1 - p_ad_mci_to_mild_ni  # remainder stays in AD-MCI

# ---- No intervention: Mild ----
p_mild_to_moderate_ni <- rep(p_mild_to_moderate, n_cycles)
p_mild_to_severe_ni <- rep(p_mild_to_severe, n_cycles)
p_mild_to_mild_ni <- 1 - (p_mild_to_moderate_ni + p_mild_to_severe_ni)

# ---- No intervention: Moderate ----
p_moderate_to_mild_ni <- rep(p_moderate_to_mild, n_cycles)
p_moderate_to_severe_ni <- rep(p_moderate_to_severe, n_cycles)
p_moderate_to_moderate_ni <- 1 - (p_moderate_to_mild_ni + p_moderate_to_severe_ni)

# ---- No intervention: Severe ----
p_severe_to_moderate_ni <- rep(p_severe_to_moderate, n_cycles)
p_severe_to_severe_ni <- 1 - p_severe_to_moderate_ni

# ---- Intervention: AD-MCI ----
rate_ad_mci_to_mild <- -log(1 - p_ad_mci_to_mild)
adjusted_rate_ad_mci_to_mild <- rate_ad_mci_to_mild * rr_ad_mci_to_mild
p_ad_mci_to_mild_i <- rep(1 - exp(-adjusted_rate_ad_mci_to_mild), n_cycles)
p_ad_mci_to_ad_mci_i <- 1 - p_ad_mci_to_mild_i

# ---- Intervention: Mild ----
rate_mild_to_moderate <- -log(1 - p_mild_to_moderate)
adjusted_rate_mild_to_moderate <- rate_mild_to_moderate * rr_mild_to_moderate
p_mild_to_moderate_i <- rep(1 - exp(-adjusted_rate_mild_to_moderate), n_cycles)

# p_mild_to_severe remains the same
p_mild_to_severe_i <- rep(p_mild_to_severe, n_cycles)

# Recalculate staying probability
p_mild_to_mild_i <- 1 - (p_mild_to_moderate_i + p_mild_to_severe_i)

# ---- Intervention: Moderate (same as no intervention) ----
p_moderate_to_mild_i <- p_moderate_to_mild_ni
p_moderate_to_severe_i <- p_moderate_to_severe_ni
p_moderate_to_moderate_i <- p_moderate_to_moderate_ni

# ---- Intervention: Severe (same as no intervention) ----
p_severe_to_moderate_i <- p_severe_to_moderate_ni
p_severe_to_severe_i <- p_severe_to_severe_ni
# ===============================
# Transition Probability Matrix Initialization
# ===============================

build_transition_matrix <- function(strat) {
  
  # Initialize array: states x states x cycles
  tpm <- array(0, dim = c(length(states), length(states), n_cycles),
               dimnames = list(from = states, to = states, cycle = 1:n_cycles))
  
  for (cycle in 1:n_cycles) {
    
    # ---- Assign death transition probabilities from each state ----
    tpm["ad_mci", "death", cycle]     <- mortality_by_age$prob_death_ad_mci[cycle]
    tpm["mild", "death", cycle]       <- mortality_by_age$prob_death_mild[cycle]
    tpm["moderate", "death", cycle]   <- mortality_by_age$prob_death_moderate[cycle]
    tpm["severe", "death", cycle]     <- mortality_by_age$prob_death_severe[cycle]
    tpm["death", "death", cycle]      <- 1  # Death is absorbing
    
    # ---- Living state transitions adjusted for mortality ----
    if (strat == "no_intervention") {
      
      # From ad_mci
      tpm["ad_mci", "mild", cycle]    <- p_ad_mci_to_mild_ni[cycle] * (1 - mortality_by_age$prob_death_ad_mci[cycle])
      tpm["ad_mci", "ad_mci", cycle]  <- p_ad_mci_to_ad_mci_ni[cycle] * (1 - mortality_by_age$prob_death_ad_mci[cycle])
      
      # From mild
      tpm["mild", "moderate", cycle]  <- p_mild_to_moderate_ni[cycle] * (1 - mortality_by_age$prob_death_mild[cycle])
      tpm["mild", "severe", cycle]    <- p_mild_to_severe_ni[cycle] * (1 - mortality_by_age$prob_death_mild[cycle])
      tpm["mild", "mild", cycle]      <- p_mild_to_mild_ni[cycle] * (1 - mortality_by_age$prob_death_mild[cycle])
      
      # From moderate
      tpm["moderate", "mild", cycle]     <- p_moderate_to_mild_ni[cycle] * (1 - mortality_by_age$prob_death_moderate[cycle])
      tpm["moderate", "severe", cycle]   <- p_moderate_to_severe_ni[cycle] * (1 - mortality_by_age$prob_death_moderate[cycle])
      tpm["moderate", "moderate", cycle] <- p_moderate_to_moderate_ni[cycle] * (1 - mortality_by_age$prob_death_moderate[cycle])
      
      # From severe
      tpm["severe", "moderate", cycle] <- p_severe_to_moderate_ni[cycle] * (1 - mortality_by_age$prob_death_severe[cycle])
      tpm["severe", "severe", cycle]   <- p_severe_to_severe_ni[cycle] * (1 - mortality_by_age$prob_death_severe[cycle])
      
    } else if (strat == "intervention") {
      
      # From ad_mci
      tpm["ad_mci", "mild", cycle]    <- p_ad_mci_to_mild_i[cycle] * (1 - mortality_by_age$prob_death_ad_mci[cycle])
      tpm["ad_mci", "ad_mci", cycle]  <- p_ad_mci_to_ad_mci_i[cycle] * (1 - mortality_by_age$prob_death_ad_mci[cycle])
      
      # From mild
      tpm["mild", "moderate", cycle]  <- p_mild_to_moderate_i[cycle] * (1 - mortality_by_age$prob_death_mild[cycle])
      tpm["mild", "severe", cycle]    <- p_mild_to_severe_i[cycle] * (1 - mortality_by_age$prob_death_mild[cycle])
      tpm["mild", "mild", cycle]      <- p_mild_to_mild_i[cycle] * (1 - mortality_by_age$prob_death_mild[cycle])
      
      # From moderate
      tpm["moderate", "mild", cycle]     <- p_moderate_to_mild_i[cycle] * (1 - mortality_by_age$prob_death_moderate[cycle])
      tpm["moderate", "severe", cycle]   <- p_moderate_to_severe_i[cycle] * (1 - mortality_by_age$prob_death_moderate[cycle])
      tpm["moderate", "moderate", cycle] <- p_moderate_to_moderate_i[cycle] * (1 - mortality_by_age$prob_death_moderate[cycle])
      
      # From severe
      tpm["severe", "moderate", cycle] <- p_severe_to_moderate_i[cycle] * (1 - mortality_by_age$prob_death_severe[cycle])
      tpm["severe", "severe", cycle]   <- p_severe_to_severe_i[cycle] * (1 - mortality_by_age$prob_death_severe[cycle])
    }
    
    # ---- Normalize rows if necessary ----
    for (state in states) {
      row_sum <- sum(tpm[state, , cycle])
      if (row_sum != 0 && row_sum != 1) {
        tpm[state, , cycle] <- tpm[state, , cycle] / row_sum
      }
    }
  }
  
  return(tpm)
}
# ===============================
# Annual Societal Costs by Age and Cognitive State (SEK)
# ===============================

# Define age range
ages <- 60:100

# Calculate costs per state using fitted GLM formulas
cost_ad_mci    <- 37.961 * ages^2 - 1234.5 * ages + 124934
cost_mild      <- 49.314 * ages^2 - 1828.7 * ages + 166239
cost_moderate  <- 61.412 * ages^2 - 2086.3 * ages + 205578
cost_severe    <- 131670 * exp(0.0176 * ages)
cost_death     <- rep(0, length(ages))

# Combine into a data frame
annual_costs <- data.frame(
  age = ages,
  ad_mci = cost_ad_mci,
  mild = cost_mild,
  moderate = cost_moderate,
  severe = cost_severe,
  death = cost_death
)
# ===============================
# Markov Model Execution Function
# ===============================

run_markov_model <- function(strat) {
  
  # Load transition probability matrix for strategy
  tpm <- build_transition_matrix(strat)
  
  # ---- Initialize matrices and vectors ----
  trace <- matrix(0, nrow = n_cycles + 1, ncol = length(states),
                  dimnames = list(cycle = 0:n_cycles, state = states))
  trace[1, "ad_mci"] <- cohort_size
  
  qaly_vector <- numeric(n_cycles)
  cost_vector <- numeric(n_cycles)
  
  pys_matrix <- matrix(0, nrow = n_cycles, ncol = 4,
                       dimnames = list(cycle = 1:n_cycles,
                                       state = c("ad_mci", "mild", "moderate", "severe")))
  
  deaths_vector <- numeric(n_cycles)
  deaths_ad_mci <- numeric(n_cycles)
  
  for (cycle in 1:n_cycles) {
    # ---- Update state distribution ----
    trace[cycle + 1, ] <- trace[cycle, ] %*% tpm[, , cycle]
    
    # ---- Current age ----
    age <- start_age + cycle - 1
    
    # ---- QALYs ----
    qaly_vector[cycle] <- trace[cycle, "ad_mci"] * u_ad_mci(age) +
      trace[cycle, "mild"]   * u_mild(age) +
      trace[cycle, "moderate"] * u_moderate(age) +
      trace[cycle, "severe"] * u_severe(age)
    
    # ---- Costs ----
    age_index <- age - 59  # for indexing from 1 (60 = row 1)
    cost_vector[cycle] <- trace[cycle, "ad_mci"] * annual_costs$ad_mci[age_index] +
      trace[cycle, "mild"]   * annual_costs$mild[age_index] +
      trace[cycle, "moderate"] * annual_costs$moderate[age_index] +
      trace[cycle, "severe"] * annual_costs$severe[age_index]
    
    if (strat == "intervention") {
      cost_vector[cycle] <- cost_vector[cycle] +
        trace[cycle, "ad_mci"] * dmt_annual_cost +
        trace[cycle, "mild"]   * dmt_annual_cost
    }
    
    # ---- Person-years ----
    pys_matrix[cycle, "ad_mci"]   <- trace[cycle, "ad_mci"]
    pys_matrix[cycle, "mild"]     <- trace[cycle, "mild"]
    pys_matrix[cycle, "moderate"] <- trace[cycle, "moderate"]
    pys_matrix[cycle, "severe"]   <- trace[cycle, "severe"]
    
    # ---- Total deaths in cycle ----
    deaths_vector[cycle] <- trace[cycle + 1, "death"] - trace[cycle, "death"]
    
    # ---- Cumulative deaths from AD-MCI ----
    deaths_ad_mci[cycle] <- trace[cycle, "ad_mci"] * tpm["ad_mci", "death", cycle]
  }
  
  # ---- Half-cycle correction ----
  qaly_vector <- 0.5 * (qaly_vector[-n_cycles] + qaly_vector[-1])
  cost_vector <- 0.5 * (cost_vector[-n_cycles] + cost_vector[-1])
  pys_matrix <- 0.5 * (pys_matrix[-n_cycles, ] + pys_matrix[-1, ])
  deaths_vector <- deaths_vector[-n_cycles]
  deaths_ad_mci <- deaths_ad_mci[-n_cycles]
  trace <- trace[-(n_cycles + 1), ]
  
  # ---- Discounting ----
  discount_factors <- 1 / ((1 + discount_rate) ^ (0:(length(qaly_vector) - 1)))
  qaly_discounted <- sum(qaly_vector * discount_factors) / cohort_size
  cost_discounted <- sum(cost_vector * discount_factors) / cohort_size
  
  # ---- Return results ----
  return(list(
    qalys = qaly_discounted,
    final_costs = cost_discounted,
    pys_ad_mci = sum(pys_matrix[, "ad_mci"]) / cohort_size,
    pys_mild = sum(pys_matrix[, "mild"]) / cohort_size,
    pys_mod = sum(pys_matrix[, "moderate"]) / cohort_size,
    pys_sev = sum(pys_matrix[, "severe"]) / cohort_size,
    deaths_total = sum(deaths_vector),
    deaths_ad_mci = sum(deaths_ad_mci)
  ))
}

# ---- Run the model for both strategies ----
results <- list()
results$no_intervention <- run_markov_model("no_intervention")
results$intervention <- run_markov_model("intervention")
```
