## Supplementary material for "Replicating Health-Economic Simulation Models for Alzheimer’s Disease Using Artificial Intelligence": final R scripts: Wimo_O9.R

```
# Input Parameters for Markov Model from Wimo et al. (2020)

# Descriptive variables
state_names <- c("ad_mci", "mild", "moderate", "severe", "death")
strategy_names <- c("no_intervention", "intervention")
start_age <- 60
n_cycles <- 40
cycle_length <- 1
discount_rate <- 0.03
cohort_size <- 100000

# Annual treatment cost for the hypothetical DMT (SEK)
cost_dmt <- 50000

# Transition probabilities (Base Case - No Intervention)
p_ad_mci_to_mild <- 0.206    # Derived from 3-year risk of 50% progression to AD-dementia
p_ad_mci_to_death <- NA      # To be computed from general life tables
p_ad_mci_stay <- 1 - p_ad_mci_to_mild  # Will be adjusted later to account for death

# Transition probabilities between AD-dementia states (from Supplementary Table 3)
p_mild_to_mild <- 0.706
p_mild_to_moderate <- 0.293
p_mild_to_severe <- 0.001

p_moderate_to_mild <- NA  # Not modeled
p_moderate_to_moderate <- 0.803
p_moderate_to_severe <- 0.109
p_moderate_to_death <- NA  # Age- and severity-dependent

p_severe_to_moderate <- NA  # Not modeled
p_severe_to_severe <- 0.804
p_severe_to_death <- NA     # Age- and severity-dependent

# Hazard ratios for mortality relative to "very mild" (from Supplementary Table 4)
hr_mild <- 1.318
hr_moderate <- 2.419
hr_severe <- 4.267

# Relative risks (intervention effect, base case) from main paper text:
rr_ad_mci_to_mild <- 0.75         # 25% reduction in progression
rr_mild_to_moderate <- 0.75       # 25% reduction in progression

# QALY utility functions from Supplementary Table 6 regression formulas
u_ad_mci <- function(age) {
  -0.002464 * age + 0.962679
}

u_mild <- function(age) {
  -0.0025 * age + 0.8293
}

u_moderate <- function(age) {
  -4e-07 * age^2 - 0.0024 * age + 0.5828
}

u_severe <- function(age) {
  -4e-07 * age^2 - 0.0025 * age + 0.4337
}

u_death <- function(age) {
  return(0)
}
# Load required mortality and weight data
mortality_data <- read.csv("mortalityRates_Swedish_2016.csv", stringsAsFactors = FALSE)
weights_data <- read.csv("weightsMortalityRates_Swedish_2016.csv", stringsAsFactors = FALSE)

# Define age mapping from group labels to individual ages
age_group_mapping <- list(
  "60-64" = 60:64,
  "65-69" = 65:69,
  "70-74" = 70:74,
  "75-79" = 75:79,
  "80-84" = 80:84,
  "85-89" = 85:89,
  "90+"   = 90:99
)

# Expand grouped mortality rates to individual ages
expanded_mortality <- data.frame()

for (i in 1:nrow(mortality_data)) {
  group <- mortality_data$age[i]
  ages <- age_group_mapping[[group]]
  male_rate <- mortality_data$male[i]
  female_rate <- mortality_data$female[i]
  
  expanded_rows <- data.frame(
    age = ages,
    male_rate = rep(male_rate, length(ages)),
    female_rate = rep(female_rate, length(ages))
  )
  
  expanded_mortality <- rbind(expanded_mortality, expanded_rows)
}

# Merge with weight data
merged_data <- merge(expanded_mortality, weights_data, by = "age")

# Calculate sex-weighted average mortality rate
merged_data$rate_avg <- with(merged_data, (male_rate * weight_male + female_rate * weight_female) / 
                               (weight_male + weight_female))

# Define hazard ratios (from previous prompt)
hr_mild <- 1.318
hr_moderate <- 2.419
hr_severe <- 4.267

# Compute death probabilities per age and state
merged_data$prob_death_ad_mci <- 1 - exp(-merged_data$rate_avg)
merged_data$prob_death_mild <- 1 - exp(-merged_data$rate_avg * hr_mild)
merged_data$prob_death_moderate <- 1 - exp(-merged_data$rate_avg * hr_moderate)
merged_data$prob_death_severe <- 1 - exp(-merged_data$rate_avg * hr_severe)

# Final output: select and reorder columns
final_mortality_df <- merged_data[, c("age", "rate_avg", 
                                      "prob_death_ad_mci", 
                                      "prob_death_mild", 
                                      "prob_death_moderate", 
                                      "prob_death_severe")]

# Display or return the data frame
final_mortality_df
# Initialize vectors for each transition probability over all cycles
p_ad_mci_to_mild_ni <- rep(p_ad_mci_to_mild, n_cycles)
p_ad_mci_to_mild_i <- rep({
  rate <- -log(1 - p_ad_mci_to_mild)
  adj_rate <- rate * rr_ad_mci_to_mild
  1 - exp(-adj_rate)
}, n_cycles)

p_ad_mci_stay_ni <- 1 - p_ad_mci_to_mild_ni
p_ad_mci_stay_i <- 1 - p_ad_mci_to_mild_i

p_mild_to_moderate_ni <- rep(p_mild_to_moderate, n_cycles)
p_mild_to_moderate_i <- rep({
  rate <- -log(1 - p_mild_to_moderate)
  adj_rate <- rate * rr_mild_to_moderate
  1 - exp(-adj_rate)
}, n_cycles)

p_mild_to_severe_ni <- rep(p_mild_to_severe, n_cycles)
p_mild_to_severe_i <- p_mild_to_severe_ni  # Not affected by intervention

p_mild_stay_ni <- 1 - (p_mild_to_moderate_ni + p_mild_to_severe_ni)
p_mild_stay_i <- 1 - (p_mild_to_moderate_i + p_mild_to_severe_i)

p_moderate_to_severe <- rep(p_moderate_to_severe, n_cycles)
p_moderate_stay <- rep(p_moderate_to_moderate, n_cycles)
# No backward transition modeled from moderate to mild

p_severe_stay <- rep(p_severe_to_severe, n_cycles)
# No backward transition modeled from severe to moderate
# Function to build the transition probability array
build_transition_matrix <- function(strat) {
  n_states <- length(state_names)
  tp_matrix <- array(0, dim = c(n_states, n_states, n_cycles),
                     dimnames = list(state_names, state_names, NULL))
  
  for (cycle in 1:n_cycles) {
    age <- start_age + (cycle - 1)
    
    # Set probabilities of transition to death
    tp_matrix["ad_mci", "death", cycle] <- final_mortality_df$prob_death_ad_mci[final_mortality_df$age == age]
    tp_matrix["mild", "death", cycle] <- final_mortality_df$prob_death_mild[final_mortality_df$age == age]
    tp_matrix["moderate", "death", cycle] <- final_mortality_df$prob_death_moderate[final_mortality_df$age == age]
    tp_matrix["severe", "death", cycle] <- final_mortality_df$prob_death_severe[final_mortality_df$age == age]
    tp_matrix["death", "death", cycle] <- 1
    
    # Define transitions from "ad_mci"
    if (strat == "no_intervention") {
      tp_matrix["ad_mci", "mild", cycle] <- p_ad_mci_to_mild_ni[cycle] * 
        (1 - tp_matrix["ad_mci", "death", cycle])
      tp_matrix["ad_mci", "ad_mci", cycle] <- p_ad_mci_stay_ni[cycle] * 
        (1 - tp_matrix["ad_mci", "death", cycle])
    } else if (strat == "intervention") {
      tp_matrix["ad_mci", "mild", cycle] <- p_ad_mci_to_mild_i[cycle] * 
        (1 - tp_matrix["ad_mci", "death", cycle])
      tp_matrix["ad_mci", "ad_mci", cycle] <- p_ad_mci_stay_i[cycle] * 
        (1 - tp_matrix["ad_mci", "death", cycle])
    }
    
    # Define transitions from "mild"
    if (strat == "no_intervention") {
      tp_matrix["mild", "moderate", cycle] <- p_mild_to_moderate_ni[cycle] * 
        (1 - tp_matrix["mild", "death", cycle])
      tp_matrix["mild", "severe", cycle] <- p_mild_to_severe_ni[cycle] * 
        (1 - tp_matrix["mild", "death", cycle])
      tp_matrix["mild", "mild", cycle] <- p_mild_stay_ni[cycle] * 
        (1 - tp_matrix["mild", "death", cycle])
    } else if (strat == "intervention") {
      tp_matrix["mild", "moderate", cycle] <- p_mild_to_moderate_i[cycle] * 
        (1 - tp_matrix["mild", "death", cycle])
      tp_matrix["mild", "severe", cycle] <- p_mild_to_severe_i[cycle] * 
        (1 - tp_matrix["mild", "death", cycle])
      tp_matrix["mild", "mild", cycle] <- p_mild_stay_i[cycle] * 
        (1 - tp_matrix["mild", "death", cycle])
    }
    
    # Define transitions from "moderate"
    tp_matrix["moderate", "severe", cycle] <- p_moderate_to_severe[cycle] * 
      (1 - tp_matrix["moderate", "death", cycle])
    tp_matrix["moderate", "moderate", cycle] <- p_moderate_stay[cycle] * 
      (1 - tp_matrix["moderate", "death", cycle])
    
    # Define transitions from "severe"
    tp_matrix["severe", "severe", cycle] <- p_severe_stay[cycle] * 
      (1 - tp_matrix["severe", "death", cycle])
    
    # Normalize rows (excluding "death" row which is already valid)
    for (state in setdiff(state_names, "death")) {
      row_sum <- sum(tp_matrix[state, , cycle])
      if (row_sum > 0 && row_sum != 1) {
        tp_matrix[state, , cycle] <- tp_matrix[state, , cycle] / row_sum
      }
    }
  }
  
  return(tp_matrix)
}
# Define age range
ages <- 60:100

# Compute costs for each state using the provided GLM-derived equations
cost_ad_mci <- 37.961 * ages^2 - 1234.5 * ages + 124934
cost_mild <- 49.314 * ages^2 - 1828.7 * ages + 166239
cost_moderate <- 61.412 * ages^2 - 2086.3 * ages + 205578
cost_severe <- 131670 * exp(0.0176 * ages)
cost_death <- rep(0, length(ages))

# Combine into a data frame
costs_by_age <- data.frame(
  age = ages,
  ad_mci = cost_ad_mci,
  mild = cost_mild,
  moderate = cost_moderate,
  severe = cost_severe,
  death = cost_death
)

# View the result
costs_by_age
# Function to run the Markov model
run_markov_model <- function(strat) {
  tp <- build_transition_matrix(strat)
  
  state_trace <- matrix(0, nrow = n_cycles + 1, ncol = length(state_names),
                        dimnames = list(0:n_cycles, state_names))
  state_trace[1, "ad_mci"] <- cohort_size
  
  qalys <- numeric(n_cycles)
  costs <- numeric(n_cycles)
  pys <- matrix(0, nrow = n_cycles, ncol = 4,
                dimnames = list(NULL, c("ad_mci", "mild", "moderate", "severe")))
  deaths_total <- numeric(n_cycles)
  deaths_ad_mci <- numeric(n_cycles)
  
  for (cycle in 1:n_cycles) {
    # Update cohort distribution using matrix multiplication
    state_trace[cycle + 1, ] <- state_trace[cycle, ] %*% tp[, , cycle]
    
    # Determine age for current cycle
    age <- start_age + (cycle - 1)
    
    # Calculate QALYs using utility functions
    qalys[cycle] <- sum(
      state_trace[cycle, "ad_mci"] * u_ad_mci(age) +
        state_trace[cycle, "mild"]   * u_mild(age) +
        state_trace[cycle, "moderate"] * u_moderate(age) +
        state_trace[cycle, "severe"] * u_severe(age)
    )
    
    # Calculate costs using age-specific values
    costs_this_cycle <- costs_by_age[costs_by_age$age == age, ]
    costs[cycle] <- sum(
      state_trace[cycle, "ad_mci"] * (costs_this_cycle$ad_mci + if (strat == "intervention") cost_dmt else 0) +
        state_trace[cycle, "mild"]   * (costs_this_cycle$mild + if (strat == "intervention") cost_dmt else 0) +
        state_trace[cycle, "moderate"] * costs_this_cycle$moderate +
        state_trace[cycle, "severe"] * costs_this_cycle$severe
    )
    
    # Record person-years in non-death states
    pys[cycle, "ad_mci"]   <- state_trace[cycle, "ad_mci"]
    pys[cycle, "mild"]     <- state_trace[cycle, "mild"]
    pys[cycle, "moderate"] <- state_trace[cycle, "moderate"]
    pys[cycle, "severe"]   <- state_trace[cycle, "severe"]
    
    # Record total deaths (incremental deaths in this cycle)
    deaths_total[cycle] <- state_trace[cycle + 1, "death"] - state_trace[cycle, "death"]
    
    # Record cumulative deaths from AD-MCI
    deaths_ad_mci[cycle] <- state_trace[cycle, "ad_mci"] * tp["ad_mci", "death", cycle]
  }
  
  # Half-cycle correction
  qalys <- head((qalys + c(tail(qalys, -1), 0)) / 2, -1)
  costs <- head((costs + c(tail(costs, -1), 0)) / 2, -1)
  pys <- head((pys + rbind(tail(pys, -1), 0)) / 2, -1)
  state_trace <- state_trace[1:n_cycles, ]
  deaths_total <- head(deaths_total, -1)
  deaths_ad_mci <- head(deaths_ad_mci, -1)
  
  # Apply discounting
  discount_factors <- 1 / ((1 + discount_rate) ^ (0:(length(qalys) - 1)))
  qalys_discounted <- sum(qalys * discount_factors) / cohort_size
  costs_discounted <- sum(costs * discount_factors) / cohort_size
  
  # Return final results
  return(list(
    qalys = qalys_discounted,
    final_costs = costs_discounted,
    pys_ad_mci = sum(pys[, "ad_mci"]) / cohort_size,
    pys_mild = sum(pys[, "mild"]) / cohort_size,
    pys_mod = sum(pys[, "moderate"]) / cohort_size,
    pys_sev = sum(pys[, "severe"]) / cohort_size,
    deaths_total = sum(deaths_total),
    deaths_ad_mci = sum(deaths_ad_mci)
  ))
}

# ---- Run the model for both strategies ----
results <- list()
results$no_intervention <- run_markov_model("no_intervention")
results$intervention <- run_markov_model("intervention")
```
