## Supplementary material for "Replicating Health-Economic Simulation Models for Alzheimer’s Disease Using Artificial Intelligence": final R scripts: Wimo_O10.R

```
# Input Parameters for Health-Economic Markov Model based on Wimo et al. (2020)

# ---- Descriptive Parameters ----
state_names <- c("ad_mci", "mild", "moderate", "severe", "death")
strategy_names <- c("no_intervention", "intervention")
cycle_length <- 1  # in years
n_cycles <- 40
start_age <- 60
discount_rate <- 0.03
cohort_size <- 100000

# ---- Transition Probabilities (Base Case Estimates) ----
# Annual transition probabilities without intervention (base case)
p_ad_mci_to_mild <- 0.206     # Derived from 3-year 50% risk: 1 - (1 - 0.5)^(1/3)
p_ad_mci_to_death <- NA       # From general life table, defined in next prompt
p_ad_mci_stay <- 1 - p_ad_mci_to_mild  # Will be adjusted for mortality later

# From ordered probit model (Supplementary Table 3)
p_mild_to_mild <- 0.706
p_mild_to_moderate <- 0.293
p_mild_to_severe <- 0.001

p_moderate_to_mild <- 0.087
p_moderate_to_moderate <- 0.803
p_moderate_to_severe <- 0.109

p_severe_to_moderate <- 0.196
p_severe_to_severe <- 0.804

# ---- Treatment Effect Parameters ----
# Base case: 25% reduction in risk for specified transitions
rr_ad_mci_to_mild <- 0.75    # Relative risk of progression from AD-MCI to mild
rr_mild_to_moderate <- 0.75  # Relative risk of progression from mild to moderate

# ---- Hazard Ratios for Mortality (Supplementary Table 4) ----
hr_mild <- 1.318
hr_moderate <- 2.419
hr_severe <- 4.267

# ---- Hypothetical DMT Treatment Cost ----
dmt_annual_cost <- 50000  # SEK/year

# ---- Utility Functions (Supplementary Table 6) ----
u_ad_mci <- function(age) {
  -0.002464 * age + 0.962679
}
u_mild <- function(age) {
  -0.0025 * age + 0.8293
}
u_moderate <- function(age) {
  -4e-07 * age^2 - 0.0024 * age + 0.5828
}
u_severe <- function(age) {
  -4e-07 * age^2 - 0.0025 * age + 0.4337
}
u_death <- function(age) {
  return(0)
}
# ---- Load mortality and weight data ----
mortality_rates <- read.csv("mortalityRates_Swedish_2016.csv", stringsAsFactors = FALSE)
weights <- read.csv("weightsMortalityRates_Swedish_2016.csv", stringsAsFactors = FALSE)

# ---- Define age group mapping ----
age_groups <- list(
  "60-64" = 60:64,
  "65-69" = 65:69,
  "70-74" = 70:74,
  "75-79" = 75:79,
  "80-84" = 80:84,
  "85-89" = 85:89,
  "90+"   = 90:99
)

# ---- Expand mortality rates to single ages ----
expanded_rates <- data.frame(age = 60:99, male = NA, female = NA)

for (group in names(age_groups)) {
  ages <- age_groups[[group]]
  rate_row <- mortality_rates[mortality_rates$age == group, ]
  expanded_rates[expanded_rates$age %in% ages, "male"] <- rate_row$male
  expanded_rates[expanded_rates$age %in% ages, "female"] <- rate_row$female
}

# ---- Merge with weights ----
merged_data <- merge(expanded_rates, weights, by = "age")

# ---- Compute sex-weighted average mortality rate per age ----
merged_data$rate_avg <- with(merged_data,
                             (male * weight_male + female * weight_female) / (weight_male + weight_female)
)

# ---- Define hazard ratios (from Prompt 1) ----
# Note: these variables must already be defined in the environment
# hr_mild <- 1.318
# hr_moderate <- 2.419
# hr_severe <- 4.267

# ---- Compute death probabilities per state ----
merged_data$prob_death_ad_mci <- 1 - exp(-merged_data$rate_avg * 1)
merged_data$prob_death_mild <- 1 - exp(-merged_data$rate_avg * hr_mild)
merged_data$prob_death_moderate <- 1 - exp(-merged_data$rate_avg * hr_moderate)
merged_data$prob_death_severe <- 1 - exp(-merged_data$rate_avg * hr_severe)

# ---- Final output ----
mortality_by_age <- merged_data[, c("age", "rate_avg", "prob_death_ad_mci",
                                    "prob_death_mild", "prob_death_moderate", "prob_death_severe")]
# ---- Time-dependent transition probability vectors ----

# Preallocate vectors for n_cycles
p_ad_mci_to_mild_no_int <- rep(p_ad_mci_to_mild, n_cycles)
p_ad_mci_stay_no_int <- 1 - p_ad_mci_to_mild_no_int

# Mild state (no intervention)
p_mild_to_moderate_no_int <- rep(p_mild_to_moderate, n_cycles)
p_mild_to_severe_no_int <- rep(p_mild_to_severe, n_cycles)
p_mild_stay_no_int <- 1 - (p_mild_to_moderate_no_int + p_mild_to_severe_no_int)

# Moderate state (no intervention)
p_moderate_to_mild_no_int <- rep(p_moderate_to_mild, n_cycles)
p_moderate_to_severe_no_int <- rep(p_moderate_to_severe, n_cycles)
p_moderate_stay_no_int <- 1 - (p_moderate_to_mild_no_int + p_moderate_to_severe_no_int)

# Severe state (no intervention)
p_severe_to_moderate_no_int <- rep(p_severe_to_moderate, n_cycles)
p_severe_stay_no_int <- 1 - p_severe_to_moderate_no_int

# ---- Intervention strategy adjustments ----

# AD-MCI to Mild (intervention)
rate_ad_mci_to_mild <- -log(1 - p_ad_mci_to_mild)
adj_rate_ad_mci_to_mild <- rate_ad_mci_to_mild * rr_ad_mci_to_mild
p_ad_mci_to_mild_int <- rep(1 - exp(-adj_rate_ad_mci_to_mild), n_cycles)
p_ad_mci_stay_int <- 1 - p_ad_mci_to_mild_int

# Mild to Moderate (intervention)
rate_mild_to_moderate <- -log(1 - p_mild_to_moderate)
adj_rate_mild_to_moderate <- rate_mild_to_moderate * rr_mild_to_moderate
p_mild_to_moderate_int <- rep(1 - exp(-adj_rate_mild_to_moderate), n_cycles)

# Mild to Severe (unchanged)
p_mild_to_severe_int <- rep(p_mild_to_severe, n_cycles)

# Mild stay (intervention)
p_mild_stay_int <- 1 - (p_mild_to_moderate_int + p_mild_to_severe_int)

# Moderate state (unchanged under intervention)
p_moderate_to_mild_int <- p_moderate_to_mild_no_int
p_moderate_to_severe_int <- p_moderate_to_severe_no_int
p_moderate_stay_int <- p_moderate_stay_no_int

# Severe state (unchanged under intervention)
p_severe_to_moderate_int <- p_severe_to_moderate_no_int
p_severe_stay_int <- p_severe_stay_no_int
# ---- Function to build transition probability matrix ----
build_transition_matrix <- function(strat) {
  
  # Define state index mapping
  states <- c("ad_mci", "mild", "moderate", "severe", "death")
  n_states <- length(states)
  trans_mat <- array(0, dim = c(n_states, n_states, n_cycles),
                     dimnames = list(states, states, NULL))
  
  for (cycle in 1:n_cycles) {
    # ---- Assign probabilities to death (from prompt 2) ----
    trans_mat["ad_mci", "death", cycle]     <- mortality_by_age$prob_death_ad_mci[cycle]
    trans_mat["mild", "death", cycle]       <- mortality_by_age$prob_death_mild[cycle]
    trans_mat["moderate", "death", cycle]   <- mortality_by_age$prob_death_moderate[cycle]
    trans_mat["severe", "death", cycle]     <- mortality_by_age$prob_death_severe[cycle]
    trans_mat["death", "death", cycle]      <- 1  # Absorbing state
    
    # ---- Placeholder for intervention-specific transitions ----
    # ---- Time-dependent transitions (adjusted for mortality) ----
    
    if (strat == "no_intervention") {
      # AD-MCI
      trans_mat["ad_mci", "mild", cycle]    <- p_ad_mci_to_mild_no_int[cycle] * (1 - mortality_by_age$prob_death_ad_mci[cycle])
      trans_mat["ad_mci", "ad_mci", cycle]  <- p_ad_mci_stay_no_int[cycle] * (1 - mortality_by_age$prob_death_ad_mci[cycle])
      
      # Mild
      trans_mat["mild", "moderate", cycle]  <- p_mild_to_moderate_no_int[cycle] * (1 - mortality_by_age$prob_death_mild[cycle])
      trans_mat["mild", "severe", cycle]    <- p_mild_to_severe_no_int[cycle] * (1 - mortality_by_age$prob_death_mild[cycle])
      trans_mat["mild", "mild", cycle]      <- p_mild_stay_no_int[cycle] * (1 - mortality_by_age$prob_death_mild[cycle])
      
      # Moderate
      trans_mat["moderate", "mild", cycle]     <- p_moderate_to_mild_no_int[cycle] * (1 - mortality_by_age$prob_death_moderate[cycle])
      trans_mat["moderate", "severe", cycle]   <- p_moderate_to_severe_no_int[cycle] * (1 - mortality_by_age$prob_death_moderate[cycle])
      trans_mat["moderate", "moderate", cycle] <- p_moderate_stay_no_int[cycle] * (1 - mortality_by_age$prob_death_moderate[cycle])
      
      # Severe
      trans_mat["severe", "moderate", cycle] <- p_severe_to_moderate_no_int[cycle] * (1 - mortality_by_age$prob_death_severe[cycle])
      trans_mat["severe", "severe", cycle]   <- p_severe_stay_no_int[cycle] * (1 - mortality_by_age$prob_death_severe[cycle])
      
    } else if (strat == "intervention") {
      # AD-MCI
      trans_mat["ad_mci", "mild", cycle]    <- p_ad_mci_to_mild_int[cycle] * (1 - mortality_by_age$prob_death_ad_mci[cycle])
      trans_mat["ad_mci", "ad_mci", cycle]  <- p_ad_mci_stay_int[cycle] * (1 - mortality_by_age$prob_death_ad_mci[cycle])
      
      # Mild
      trans_mat["mild", "moderate", cycle]  <- p_mild_to_moderate_int[cycle] * (1 - mortality_by_age$prob_death_mild[cycle])
      trans_mat["mild", "severe", cycle]    <- p_mild_to_severe_int[cycle] * (1 - mortality_by_age$prob_death_mild[cycle])
      trans_mat["mild", "mild", cycle]      <- p_mild_stay_int[cycle] * (1 - mortality_by_age$prob_death_mild[cycle])
      
      # Moderate
      trans_mat["moderate", "mild", cycle]     <- p_moderate_to_mild_int[cycle] * (1 - mortality_by_age$prob_death_moderate[cycle])
      trans_mat["moderate", "severe", cycle]   <- p_moderate_to_severe_int[cycle] * (1 - mortality_by_age$prob_death_moderate[cycle])
      trans_mat["moderate", "moderate", cycle] <- p_moderate_stay_int[cycle] * (1 - mortality_by_age$prob_death_moderate[cycle])
      
      # Severe
      trans_mat["severe", "moderate", cycle] <- p_severe_to_moderate_int[cycle] * (1 - mortality_by_age$prob_death_severe[cycle])
      trans_mat["severe", "severe", cycle]   <- p_severe_stay_int[cycle] * (1 - mortality_by_age$prob_death_severe[cycle])
    }
    
    
    # ---- Normalize rows (if row sum != 1) ----
    for (s in states) {
      row_sum <- sum(trans_mat[s, , cycle])
      if (row_sum > 0 && abs(row_sum - 1) > 1e-8) {
        trans_mat[s, , cycle] <- trans_mat[s, , cycle] / row_sum
      }
    }
  }
  
  return(trans_mat)
}
# ---- Calculate per-person annual societal costs by age and cognitive state ----

# Age range
ages <- 60:100

# Cost formulas
cost_ad_mci <- 37.961 * ages^2 - 1234.5 * ages + 124934
cost_mild <- 49.314 * ages^2 - 1828.7 * ages + 166239
cost_moderate <- 61.412 * ages^2 - 2086.3 * ages + 205578
cost_severe <- 131670 * exp(0.0176 * ages)
cost_death <- rep(0, length(ages))

# Construct data frame
annual_costs <- data.frame(
  age = ages,
  ad_mci = cost_ad_mci,
  mild = cost_mild,
  moderate = cost_moderate,
  severe = cost_severe,
  death = cost_death
)
# ---- Function to run the Markov model for a given strategy ----
run_markov_model <- function(strat) {
  states <- c("ad_mci", "mild", "moderate", "severe", "death")
  n_states <- length(states)
  
  # Initialize transition matrix
  trans_mat <- build_transition_matrix(strat)
  
  # Initialize state trace
  trace <- matrix(0, nrow = n_cycles + 1, ncol = n_states, dimnames = list(NULL, states))
  trace[1, "ad_mci"] <- cohort_size
  
  # Initialize output vectors/matrices
  qalys <- numeric(n_cycles)
  costs <- numeric(n_cycles)
  pys <- matrix(0, nrow = n_cycles, ncol = 4, dimnames = list(NULL, states[1:4]))
  deaths_total <- numeric(n_cycles)
  deaths_ad_mci <- numeric(n_cycles)
  
  # ---- Placeholder for simulation loop ----
  for (t in 1:n_cycles) {
    # Advance the cohort
    trace[t + 1, ] <- trace[t, ] %*% trans_mat[, , t]
    
    # Current age
    current_age <- start_age + (t - 1)
    
    # Compute QALYs
    qalys[t] <- trace[t, "ad_mci"]    * u_ad_mci(current_age) +
      trace[t, "mild"]      * u_mild(current_age) +
      trace[t, "moderate"]  * u_moderate(current_age) +
      trace[t, "severe"]    * u_severe(current_age)
    
    # Compute Costs
    age_index <- current_age - 59  # index for rows in annual_costs (starts at age 60)
    cost_vector <- c(
      annual_costs$ad_mci[age_index],
      annual_costs$mild[age_index],
      annual_costs$moderate[age_index],
      annual_costs$severe[age_index],
      annual_costs$death[age_index]
    )
    
    # Add DMT costs in intervention for ad_mci and mild
    if (strat == "intervention") {
      cost_vector[1] <- cost_vector[1] + dmt_annual_cost  # ad_mci
      cost_vector[2] <- cost_vector[2] + dmt_annual_cost  # mild
    }
    
    costs[t] <- sum(trace[t, ] * cost_vector)
    
    # Record person-years in non-death states
    pys[t, "ad_mci"]   <- trace[t, "ad_mci"]
    pys[t, "mild"]     <- trace[t, "mild"]
    pys[t, "moderate"] <- trace[t, "moderate"]
    pys[t, "severe"]   <- trace[t, "severe"]
    
    # Track new deaths
    deaths_total[t] <- trace[t + 1, "death"] - trace[t, "death"]
    
    # Cumulative deaths from AD-MCI
    deaths_ad_mci[t] <- trace[t, "ad_mci"] * trans_mat["ad_mci", "death", t]
  }
  
  
  # ---- Half-cycle correction ----
  qalys_hcc <- 0.5 * (qalys[-n_cycles] + qalys[-1])
  costs_hcc <- 0.5 * (costs[-n_cycles] + costs[-1])
  pys_hcc <- 0.5 * (pys[-n_cycles, ] + pys[-1, ])
  deaths_total <- deaths_total[-1]
  deaths_ad_mci <- deaths_ad_mci[-1]
  trace <- trace[-(n_cycles + 1), ]
  
  # ---- Discounting ----
  discount_factors <- 1 / ((1 + discount_rate) ^ (1:(n_cycles - 1)))
  qalys_disc <- sum(qalys_hcc * discount_factors) / cohort_size
  costs_disc <- sum(costs_hcc * discount_factors) / cohort_size
  pys_avg <- colSums(pys_hcc) / cohort_size
  deaths_total_sum <- sum(deaths_total)
  deaths_ad_mci_sum <- sum(deaths_ad_mci)
  
  return(list(
    qalys = qalys_disc,
    final_costs = costs_disc,
    pys_ad_mci = pys_avg["ad_mci"],
    pys_mild = pys_avg["mild"],
    pys_mod = pys_avg["moderate"],
    pys_sev = pys_avg["severe"],
    deaths_total = deaths_total_sum,
    deaths_ad_mci = deaths_ad_mci_sum
  ))
}

# ---- Run model for both strategies ----
results <- list()
results$no_intervention <- run_markov_model("no_intervention")
results$intervention <- run_markov_model("intervention")
```
