## Supplementary material for "Replicating Health-Economic Simulation Models for Alzheimer’s Disease Using Artificial Intelligence": final R scripts: Wimo_O11.R

```
# =======================
# Markov Model Parameters
# From Wimo et al. (2020)
# =======================

# Descriptive Variables
state_names <- c("ad_mci", "mild", "moderate", "severe", "death")
strategy_names <- c("no_intervention", "intervention")

# Model Settings
start_age <- 60
discount_rate <- 0.03
n_cycles <- 40
cycle_length <- 1
cohort_size <- 100000

# Hypothetical Disease-Modifying Treatment (DMT) Cost
dmt_annual_cost <- 50000  # SEK

# Transition Probabilities (Base Case, extracted from Supp Table 3)
p_mild_to_mild     <- 0.706
p_mild_to_moderate <- 0.293
p_mild_to_severe   <- 0.001

p_moderate_to_mild     <- 0.087
p_moderate_to_moderate <- 0.803
p_moderate_to_severe   <- 0.109

p_severe_to_moderate <- 0.196
p_severe_to_severe   <- 0.804

# Progression probability from AD-MCI to Mild (constant annual risk derived from 3-year 50% rate)
p_ad_mci_to_mild <- 1 - (1 - 0.50)^(1/3)  # ≈ 0.206

# Hazard Ratios for Mortality (from Supplementary Table 4)
hr_mild     <- 1.318
hr_moderate <- 2.419
hr_severe   <- 4.267

# Relative Risks for Intervention (Base Case: 25% risk reduction)
rr_ad_mci_to_mild_intervention <- 0.75
rr_mild_to_moderate_intervention <- 0.75

# QALY Utility Functions (based on Supplementary Table 6 regression fits)
u_ad_mci <- function(age) {
  -0.002464 * age + 0.962679
}

u_mild <- function(age) {
  -0.0025 * age + 0.8293
}

u_moderate <- function(age) {
  -4e-07 * age^2 - 0.0024 * age + 0.5828
}

u_severe <- function(age) {
  -4e-07 * age^2 - 0.0025 * age + 0.4337
}

# Death state utility (fixed)
u_death <- function(age) {
  return(0)
}
# ======================================
# Load and Process Mortality Rate Data
# ======================================

# Load input files
mortality_rates <- read.csv("mortalityRates_Swedish_2016.csv", stringsAsFactors = FALSE)
weights <- read.csv("weightsMortalityRates_Swedish_2016.csv", stringsAsFactors = FALSE)

# Define age group mapping
group_to_ages <- list(
  "60-64" = 60:64,
  "65-69" = 65:69,
  "70-74" = 70:74,
  "75-79" = 75:79,
  "80-84" = 80:84,
  "85-89" = 85:89,
  "90+"   = 90:99
)

# Expand grouped mortality rates to single ages
expanded_rates <- data.frame()
for (i in 1:nrow(mortality_rates)) {
  group <- mortality_rates$age[i]
  ages <- group_to_ages[[group]]
  n_ages <- length(ages)
  df <- data.frame(
    age = ages,
    male = rep(mortality_rates$male[i], n_ages),
    female = rep(mortality_rates$female[i], n_ages)
  )
  expanded_rates <- rbind(expanded_rates, df)
}

# Merge expanded rates with weights
weights$age <- as.integer(weights$age)
expanded_rates$age <- as.integer(expanded_rates$age)
merged <- merge(expanded_rates, weights, by = "age")

# Compute sex-weighted average mortality rate per age
merged$rate_avg <- with(merged, male * weight_male + female * weight_female)

# Compute probability of death by severity state using hazard ratios
hr_mild <- 1.318
hr_moderate <- 2.419
hr_severe <- 4.267

output <- data.frame(
  age = merged$age,
  rate_avg = merged$rate_avg,
  prob_death_ad_mci = 1 - exp(-merged$rate_avg),  # HR = 1
  prob_death_mild = 1 - exp(-merged$rate_avg * hr_mild),
  prob_death_moderate = 1 - exp(-merged$rate_avg * hr_moderate),
  prob_death_severe = 1 - exp(-merged$rate_avg * hr_severe)
)

# Final mortality data frame: output
# ==============================================
# Time-Dependent Transition Probabilities (Vectors)
# ==============================================

# Initialize vectors for no intervention strategy (length = n_cycles)
p_ad_mci_to_mild <- rep(p_ad_mci_to_mild, n_cycles)
p_ad_mci_stay <- 1 - p_ad_mci_to_mild

p_mild_to_moderate <- rep(p_mild_to_moderate, n_cycles)
p_mild_to_severe <- rep(p_mild_to_severe, n_cycles)
p_mild_stay <- 1 - p_mild_to_moderate - p_mild_to_severe

p_moderate_to_mild <- rep(p_moderate_to_mild, n_cycles)
p_moderate_to_severe <- rep(p_moderate_to_severe, n_cycles)
p_moderate_stay <- 1 - p_moderate_to_mild - p_moderate_to_severe

p_severe_to_moderate <- rep(p_severe_to_moderate, n_cycles)
p_severe_stay <- 1 - p_severe_to_moderate

# Initialize vectors for intervention strategy
# Apply RR to transitions from "ad_mci" and "mild"
rr_ad_mci_to_mild <- rr_ad_mci_to_mild_intervention
rr_mild_to_moderate <- rr_mild_to_moderate_intervention

# Convert p -> rate -> adjusted rate -> adjusted p
rate_ad_mci_to_mild <- -log(1 - p_ad_mci_to_mild)
rate_mild_to_moderate <- -log(1 - p_mild_to_moderate)

adj_rate_ad_mci_to_mild <- rate_ad_mci_to_mild * rr_ad_mci_to_mild
adj_rate_mild_to_moderate <- rate_mild_to_moderate * rr_mild_to_moderate

p_ad_mci_to_mild_int <- 1 - exp(-adj_rate_ad_mci_to_mild)
p_ad_mci_stay_int <- 1 - p_ad_mci_to_mild_int

p_mild_to_moderate_int <- 1 - exp(-adj_rate_mild_to_moderate)
p_mild_to_severe_int <- p_mild_to_severe  # unchanged
p_mild_stay_int <- 1 - p_mild_to_moderate_int - p_mild_to_severe_int

# Transitions not affected by intervention
p_moderate_to_mild_int <- p_moderate_to_mild
p_moderate_to_severe_int <- p_moderate_to_severe
p_moderate_stay_int <- p_moderate_stay

p_severe_to_moderate_int <- p_severe_to_moderate
p_severe_stay_int <- p_severe_stay
# ================================================
# Transition Probability Matrix Initialization
# ================================================

build_transition_matrix <- function(strat) {
  # Define state order
  states <- c("ad_mci", "mild", "moderate", "severe", "death")
  n_states <- length(states)
  
  # Initialize 3D array: [from_state, to_state, cycle]
  tpm <- array(0, dim = c(n_states, n_states, n_cycles),
               dimnames = list(from = states, to = states, cycle = NULL))
  
  # Loop over cycles
  for (t in 1:n_cycles) {
    # Death probabilities from prompt 2
    tpm["ad_mci", "death", t] <- output$prob_death_ad_mci[t]
    tpm["mild", "death", t] <- output$prob_death_mild[t]
    tpm["moderate", "death", t] <- output$prob_death_moderate[t]
    tpm["severe", "death", t] <- output$prob_death_severe[t]
    tpm["death", "death", t] <- 1  # Absorbing state
    
    # =============================
    # [INSERT PROMPT 4B HERE]
    # =============================
    
    if (strat == "no_intervention") {
      # AD-MCI transitions
      tpm["ad_mci", "mild", t] <- p_ad_mci_to_mild[t] * (1 - output$prob_death_ad_mci[t])
      tpm["ad_mci", "ad_mci", t] <- p_ad_mci_stay[t] * (1 - output$prob_death_ad_mci[t])
      
      # Mild transitions
      tpm["mild", "moderate", t] <- p_mild_to_moderate[t] * (1 - output$prob_death_mild[t])
      tpm["mild", "severe", t] <- p_mild_to_severe[t] * (1 - output$prob_death_mild[t])
      tpm["mild", "mild", t] <- p_mild_stay[t] * (1 - output$prob_death_mild[t])
      
      # Moderate transitions
      tpm["moderate", "mild", t] <- p_moderate_to_mild[t] * (1 - output$prob_death_moderate[t])
      tpm["moderate", "severe", t] <- p_moderate_to_severe[t] * (1 - output$prob_death_moderate[t])
      tpm["moderate", "moderate", t] <- p_moderate_stay[t] * (1 - output$prob_death_moderate[t])
      
      # Severe transitions
      tpm["severe", "moderate", t] <- p_severe_to_moderate[t] * (1 - output$prob_death_severe[t])
      tpm["severe", "severe", t] <- p_severe_stay[t] * (1 - output$prob_death_severe[t])
      
    } else if (strat == "intervention") {
      # AD-MCI transitions (adjusted)
      tpm["ad_mci", "mild", t] <- p_ad_mci_to_mild_int[t] * (1 - output$prob_death_ad_mci[t])
      tpm["ad_mci", "ad_mci", t] <- p_ad_mci_stay_int[t] * (1 - output$prob_death_ad_mci[t])
      
      # Mild transitions (adjusted)
      tpm["mild", "moderate", t] <- p_mild_to_moderate_int[t] * (1 - output$prob_death_mild[t])
      tpm["mild", "severe", t] <- p_mild_to_severe_int[t] * (1 - output$prob_death_mild[t])
      tpm["mild", "mild", t] <- p_mild_stay_int[t] * (1 - output$prob_death_mild[t])
      
      # Moderate transitions (unchanged)
      tpm["moderate", "mild", t] <- p_moderate_to_mild_int[t] * (1 - output$prob_death_moderate[t])
      tpm["moderate", "severe", t] <- p_moderate_to_severe_int[t] * (1 - output$prob_death_moderate[t])
      tpm["moderate", "moderate", t] <- p_moderate_stay_int[t] * (1 - output$prob_death_moderate[t])
      
      # Severe transitions (unchanged)
      tpm["severe", "moderate", t] <- p_severe_to_moderate_int[t] * (1 - output$prob_death_severe[t])
      tpm["severe", "severe", t] <- p_severe_stay_int[t] * (1 - output$prob_death_severe[t])
    }
    
    # Normalize rows if they do not sum to 1
    for (s in states) {
      row_sum <- sum(tpm[s, , t])
      if (row_sum > 0 && abs(row_sum - 1) > 1e-8) {
        tpm[s, , t] <- tpm[s, , t] / row_sum
      }
    }
  }
  
  return(tpm)
}
# ==========================================
# Annual Societal Costs per Person (in SEK)
# ==========================================

# Define age range
ages <- 60:100

# Compute costs using the fitted GLM equations
cost_ad_mci <- 37.961 * ages^2 - 1234.5 * ages + 124934
cost_mild <- 49.314 * ages^2 - 1828.7 * ages + 166239
cost_moderate <- 61.412 * ages^2 - 2086.3 * ages + 205578
cost_severe <- 131670 * exp(0.0176 * ages)
cost_death <- rep(0, length(ages))

# Combine into data frame
costs_df <- data.frame(
  age = ages,
  ad_mci = cost_ad_mci,
  mild = cost_mild,
  moderate = cost_moderate,
  severe = cost_severe,
  death = cost_death
)
# ===============================================
# Markov Model Execution Function (Structure)
# ===============================================

run_markov_model <- function(strat) {
  states <- c("ad_mci", "mild", "moderate", "severe", "death")
  n_states <- length(states)
  
  # Build transition probability array for given strategy
  tpm <- build_transition_matrix(strat)
  
  # Initialize state trace: rows = cycles, cols = states
  trace <- matrix(0, nrow = n_cycles + 1, ncol = n_states,
                  dimnames = list(0:n_cycles, states))
  trace[1, "ad_mci"] <- cohort_size  # Entire cohort starts in ad_mci
  
  # Initialize output structures
  qalys <- numeric(n_cycles + 1)
  costs <- numeric(n_cycles + 1)
  pys <- matrix(0, nrow = n_cycles + 1, ncol = 4)  # Non-death states
  colnames(pys) <- c("ad_mci", "mild", "moderate", "severe")
  
  deaths_total <- numeric(n_cycles + 1)
  deaths_ad_mci <- numeric(n_cycles + 1)
  
  for (t in 1:n_cycles) {
    # Update cohort distribution
    trace[t + 1, ] <- trace[t, ] %*% tpm[, , t]
    
    # Calculate current age
    age <- start_age + t - 1
    
    # QALYs: apply age-specific utility functions to living states
    qalys[t + 1] <- trace[t + 1, "ad_mci"] * u_ad_mci(age) +
      trace[t + 1, "mild"]   * u_mild(age) +
      trace[t + 1, "moderate"] * u_moderate(age) +
      trace[t + 1, "severe"] * u_severe(age)
    
    # Costs: apply state- and age-specific costs
    age_index <- which(costs_df$age == age)
    costs[t + 1] <- trace[t + 1, "ad_mci"]   * costs_df$ad_mci[age_index] +
      trace[t + 1, "mild"]     * costs_df$mild[age_index] +
      trace[t + 1, "moderate"] * costs_df$moderate[age_index] +
      trace[t + 1, "severe"]   * costs_df$severe[age_index]
    
    # Add DMT cost for intervention strategy
    if (strat == "intervention") {
      costs[t + 1] <- costs[t + 1] +
        trace[t + 1, "ad_mci"] * dmt_annual_cost +
        trace[t + 1, "mild"]   * dmt_annual_cost
    }
    
    # Person-years (PYs)
    pys[t + 1, "ad_mci"]   <- trace[t + 1, "ad_mci"]
    pys[t + 1, "mild"]     <- trace[t + 1, "mild"]
    pys[t + 1, "moderate"] <- trace[t + 1, "moderate"]
    pys[t + 1, "severe"]   <- trace[t + 1, "severe"]
    
    # Total deaths this cycle (difference in death state between t+1 and t)
    deaths_total[t + 1] <- trace[t + 1, "death"] - trace[t, "death"]
    
    # Deaths from AD-MCI this cycle
    deaths_ad_mci[t + 1] <- trace[t, "ad_mci"] * tpm["ad_mci", "death", t]
  }
  
  # Half-cycle correction (apply to QALYs, costs, and PYs)
  qalys_hc <- 0.5 * (qalys[-(n_cycles + 1)] + qalys[-1])
  costs_hc <- 0.5 * (costs[-(n_cycles + 1)] + costs[-1])
  pys_hc <- 0.5 * (pys[-(n_cycles + 1), ] + pys[-1, ])
  trace <- trace[-(n_cycles + 1), ]
  deaths_total <- deaths_total[-(n_cycles + 1)]
  deaths_ad_mci <- deaths_ad_mci[-(n_cycles + 1)]
  
  # Apply discounting
  discount_vector <- 1 / (1 + discount_rate)^(0:(n_cycles - 1))
  qalys_discounted <- sum(qalys_hc * discount_vector) / cohort_size
  costs_discounted <- sum(costs_hc * discount_vector) / cohort_size
  
  # Return named list
  return(list(
    qalys = qalys_discounted,
    final_costs = costs_discounted,
    pys_ad_mci = sum(pys_hc[, "ad_mci"]) / cohort_size,
    pys_mild = sum(pys_hc[, "mild"]) / cohort_size,
    pys_mod = sum(pys_hc[, "moderate"]) / cohort_size,
    pys_sev = sum(pys_hc[, "severe"]) / cohort_size,
    deaths_total = sum(deaths_total),
    deaths_ad_mci = sum(deaths_ad_mci)
  ))
}

# ===============================
# Run Markov model for strategies
# ===============================
results <- list()
results$no_intervention <- run_markov_model("no_intervention")
results$intervention <- run_markov_model("intervention")
```
