## Supplementary material for "Replicating Health-Economic Simulation Models for Alzheimer’s Disease Using Artificial Intelligence": final R scripts: Wimo_O12.R

```
# =========================
# Prompt 1: Input Parameters
# =========================

# --- Descriptive Parameters ---
state_names <- c("ad_mci", "mild", "moderate", "severe", "death")
strategy_names <- c("no_intervention", "intervention")
start_age <- 60
cycle_length <- 1  # year
n_cycles <- 40
cohort_size <- 100000
discount_rate <- 0.03  # for both costs and QALYs

# --- DMT Treatment Cost (illustrative) ---
dmt_annual_cost <- 50000  # SEK/year

# --- Transition Probabilities (Base Case: No intervention) ---
# From Supplementary Table 3
p_mild_to_mild <- 0.706
p_mild_to_moderate <- 0.293
p_mild_to_severe <- 0.001

p_moderate_to_mild <- 0.087
p_moderate_to_moderate <- 0.803
p_moderate_to_severe <- 0.109

p_severe_to_moderate <- 0.196
p_severe_to_severe <- 0.804

# AD-MCI transition from main paper (20.6% annual progression to AD-dementia)
p_ad_mci_to_mild <- 0.206
p_ad_mci_stay <- 1 - p_ad_mci_to_mild  # no other transitions allowed

# --- Transition Adjustments for Intervention Strategy ---
# 25% relative risk reduction in AD-MCI to mild and mild to moderate
rr_mci_to_mild <- 0.75  # 25% reduction
rr_mild_to_moderate <- 0.75  # 25% reduction

# --- Hazard Ratios for Mortality by Severity (from Supplementary Table 4) ---
# Relative to "very mild" (used as baseline)
hr_mild <- 1.318
hr_moderate <- 2.419
hr_severe <- 4.267

# --- QALY Utility Functions ---
u_ad_mci <- function(age) {
  return(-0.002464 * age + 0.962679)
}

u_mild <- function(age) {
  return(-0.0025 * age + 0.8293)
}

u_moderate <- function(age) {
  return(-4e-07 * age^2 - 0.0024 * age + 0.5828)
}

u_severe <- function(age) {
  return(-4e-07 * age^2 - 0.0025 * age + 0.4337)
}

u_death <- function(age) {
  return(0)
}
# =========================
# Prompt 2: Mortality Processing
# =========================

# --- Load mortality rate and weights files ---
mortality_rates <- read.csv("mortalityRates_Swedish_2016.csv", stringsAsFactors = FALSE)
weights <- read.csv("weightsMortalityRates_Swedish_2016.csv", stringsAsFactors = FALSE)

# --- Define age groups mapping ---
age_groups <- list(
  "60-64" = 60:64,
  "65-69" = 65:69,
  "70-74" = 70:74,
  "75-79" = 75:79,
  "80-84" = 80:84,
  "85-89" = 85:89,
  "90+"   = 90:99
)

# --- Expand mortality rates to individual ages ---
expanded_rates <- data.frame(age = integer(), rate_male = numeric(), rate_female = numeric(), stringsAsFactors = FALSE)

for (i in 1:nrow(mortality_rates)) {
  group <- mortality_rates$age[i]
  ages <- age_groups[[group]]
  male_rate <- mortality_rates$male[i]
  female_rate <- mortality_rates$female[i]
  temp <- data.frame(age = ages, rate_male = rep(male_rate, length(ages)), rate_female = rep(female_rate, length(ages)))
  expanded_rates <- rbind(expanded_rates, temp)
}

# --- Merge with weights ---
merged <- merge(expanded_rates, weights, by = "age")

# --- Compute sex-weighted average mortality rate per age ---
merged$rate_avg <- with(merged, (rate_male * weight_male + rate_female * weight_female) / (weight_male + weight_female))

# --- Compute age-specific death probabilities per state ---
merged$prob_death_ad_mci    <- 1 - exp(-merged$rate_avg * 1)          # HR = 1 for AD-MCI
merged$prob_death_mild      <- 1 - exp(-merged$rate_avg * hr_mild)
merged$prob_death_moderate  <- 1 - exp(-merged$rate_avg * hr_moderate)
merged$prob_death_severe    <- 1 - exp(-merged$rate_avg * hr_severe)

# --- Final output data frame ---
mortality_by_age <- merged[, c("age", "rate_avg", "prob_death_ad_mci", "prob_death_mild", "prob_death_moderate", "prob_death_severe")]
# =========================
# Prompt 3: Time-Dependent Transition Vectors
# =========================

# --- No Intervention Strategy ---

# AD-MCI transitions
p_ad_mci_to_mild_vec <- rep(p_ad_mci_to_mild, n_cycles)
p_ad_mci_stay_vec <- rep(1 - p_ad_mci_to_mild, n_cycles)

# Mild transitions
p_mild_to_moderate_vec <- rep(p_mild_to_moderate, n_cycles)
p_mild_to_severe_vec <- rep(p_mild_to_severe, n_cycles)
p_mild_stay_vec <- rep(1 - (p_mild_to_moderate + p_mild_to_severe), n_cycles)

# Moderate transitions
p_moderate_to_mild_vec <- rep(p_moderate_to_mild, n_cycles)
p_moderate_to_severe_vec <- rep(p_moderate_to_severe, n_cycles)
p_moderate_stay_vec <- rep(1 - (p_moderate_to_mild + p_moderate_to_severe), n_cycles)

# Severe transitions
p_severe_to_moderate_vec <- rep(p_severe_to_moderate, n_cycles)
p_severe_stay_vec <- rep(1 - p_severe_to_moderate, n_cycles)

# --- Intervention Strategy (adjusted for RR where applicable) ---

# AD-MCI transitions
rate_ad_mci_to_mild <- -log(1 - p_ad_mci_to_mild)
adj_rate_ad_mci_to_mild <- rate_ad_mci_to_mild * rr_mci_to_mild
p_ad_mci_to_mild_int_vec <- rep(1 - exp(-adj_rate_ad_mci_to_mild), n_cycles)
p_ad_mci_stay_int_vec <- rep(1 - (1 - exp(-adj_rate_ad_mci_to_mild)), n_cycles)

# Mild transitions
rate_mild_to_moderate <- -log(1 - p_mild_to_moderate)
adj_rate_mild_to_moderate <- rate_mild_to_moderate * rr_mild_to_moderate
p_mild_to_moderate_int <- 1 - exp(-adj_rate_mild_to_moderate)
p_mild_to_moderate_int_vec <- rep(p_mild_to_moderate_int, n_cycles)
p_mild_to_severe_int_vec <- rep(p_mild_to_severe, n_cycles)
p_mild_stay_int_vec <- rep(1 - (p_mild_to_moderate_int + p_mild_to_severe), n_cycles)

# Moderate transitions (no adjustment)
p_moderate_to_mild_int_vec <- rep(p_moderate_to_mild, n_cycles)
p_moderate_to_severe_int_vec <- rep(p_moderate_to_severe, n_cycles)
p_moderate_stay_int_vec <- rep(1 - (p_moderate_to_mild + p_moderate_to_severe), n_cycles)

# Severe transitions (no adjustment)
p_severe_to_moderate_int_vec <- rep(p_severe_to_moderate, n_cycles)
p_severe_stay_int_vec <- rep(1 - p_severe_to_moderate, n_cycles)
# =========================
# Prompt 4A: Initialize Transition Matrix Array
# =========================

build_transition_matrix <- function(strat) {
  states <- state_names
  n_states <- length(states)
  tp_array <- array(0, dim = c(n_states, n_states, n_cycles),
                    dimnames = list(states, states, NULL))
  
  for (cycle in 1:n_cycles) {
    # --- Transition to death state ---
    tp_array["ad_mci", "death", cycle] <- mortality_by_age$prob_death_ad_mci[cycle]
    tp_array["mild", "death", cycle] <- mortality_by_age$prob_death_mild[cycle]
    tp_array["moderate", "death", cycle] <- mortality_by_age$prob_death_moderate[cycle]
    tp_array["severe", "death", cycle] <- mortality_by_age$prob_death_severe[cycle]
    tp_array["death", "death", cycle] <- 1  # Death is absorbing
    
    # --- [INSERT PROMPT 4B HERE] --- to define the transitions from on-treatment and off-treatment states later.
    
    if (strat == "no_intervention") {
      # AD-MCI transitions
      tp_array["ad_mci", "mild", cycle] <- p_ad_mci_to_mild_vec[cycle] * (1 - mortality_by_age$prob_death_ad_mci[cycle])
      tp_array["ad_mci", "ad_mci", cycle] <- p_ad_mci_stay_vec[cycle] * (1 - mortality_by_age$prob_death_ad_mci[cycle])
      
      # Mild transitions
      tp_array["mild", "moderate", cycle] <- p_mild_to_moderate_vec[cycle] * (1 - mortality_by_age$prob_death_mild[cycle])
      tp_array["mild", "severe", cycle]   <- p_mild_to_severe_vec[cycle] * (1 - mortality_by_age$prob_death_mild[cycle])
      tp_array["mild", "mild", cycle]     <- p_mild_stay_vec[cycle] * (1 - mortality_by_age$prob_death_mild[cycle])
      
      # Moderate transitions
      tp_array["moderate", "mild", cycle]     <- p_moderate_to_mild_vec[cycle] * (1 - mortality_by_age$prob_death_moderate[cycle])
      tp_array["moderate", "severe", cycle]   <- p_moderate_to_severe_vec[cycle] * (1 - mortality_by_age$prob_death_moderate[cycle])
      tp_array["moderate", "moderate", cycle] <- p_moderate_stay_vec[cycle] * (1 - mortality_by_age$prob_death_moderate[cycle])
      
      # Severe transitions
      tp_array["severe", "moderate", cycle] <- p_severe_to_moderate_vec[cycle] * (1 - mortality_by_age$prob_death_severe[cycle])
      tp_array["severe", "severe", cycle]   <- p_severe_stay_vec[cycle] * (1 - mortality_by_age$prob_death_severe[cycle])
      
    } else if (strat == "intervention") {
      # AD-MCI transitions
      tp_array["ad_mci", "mild", cycle] <- p_ad_mci_to_mild_int_vec[cycle] * (1 - mortality_by_age$prob_death_ad_mci[cycle])
      tp_array["ad_mci", "ad_mci", cycle] <- p_ad_mci_stay_int_vec[cycle] * (1 - mortality_by_age$prob_death_ad_mci[cycle])
      
      # Mild transitions
      tp_array["mild", "moderate", cycle] <- p_mild_to_moderate_int_vec[cycle] * (1 - mortality_by_age$prob_death_mild[cycle])
      tp_array["mild", "severe", cycle]   <- p_mild_to_severe_int_vec[cycle] * (1 - mortality_by_age$prob_death_mild[cycle])
      tp_array["mild", "mild", cycle]     <- p_mild_stay_int_vec[cycle] * (1 - mortality_by_age$prob_death_mild[cycle])
      
      # Moderate transitions
      tp_array["moderate", "mild", cycle]     <- p_moderate_to_mild_int_vec[cycle] * (1 - mortality_by_age$prob_death_moderate[cycle])
      tp_array["moderate", "severe", cycle]   <- p_moderate_to_severe_int_vec[cycle] * (1 - mortality_by_age$prob_death_moderate[cycle])
      tp_array["moderate", "moderate", cycle] <- p_moderate_stay_int_vec[cycle] * (1 - mortality_by_age$prob_death_moderate[cycle])
      
      # Severe transitions
      tp_array["severe", "moderate", cycle] <- p_severe_to_moderate_int_vec[cycle] * (1 - mortality_by_age$prob_death_severe[cycle])
      tp_array["severe", "severe", cycle]   <- p_severe_stay_int_vec[cycle] * (1 - mortality_by_age$prob_death_severe[cycle])
    }
    
    # --- Normalize each row (excluding death) ---
    for (state in states[-which(states == "death")]) {
      row_sum <- sum(tp_array[state, , cycle])
      if (row_sum != 1 && row_sum > 0) {
        tp_array[state, , cycle] <- tp_array[state, , cycle] / row_sum
      }
    }
  }
  
  return(tp_array)
}
# =========================
# Prompt 5: Annual Societal Per-Person Costs by Age and State
# =========================

# --- Age vector ---
ages <- 60:100

# --- Compute costs per cognitive state using fitted equations ---
cost_ad_mci <- 37.961 * ages^2 - 1234.5 * ages + 124934
cost_mild <- 49.314 * ages^2 - 1828.7 * ages + 166239
cost_moderate <- 61.412 * ages^2 - 2086.3 * ages + 205578
cost_severe <- 131670 * exp(0.0176 * ages)
cost_death <- rep(0, length(ages))

# --- Create data frame ---
cost_by_age_state <- data.frame(
  age = ages,
  ad_mci = cost_ad_mci,
  mild = cost_mild,
  moderate = cost_moderate,
  severe = cost_severe,
  death = cost_death
)
# =========================
# Prompt 6A: Markov Model Simulation Structure
# =========================

run_markov_model <- function(strategy) {
  states <- state_names
  n_states <- length(states)
  
  # --- Initialize transition matrix ---
  tp <- build_transition_matrix(strategy)
  
  # --- Initialize outputs ---
  state_trace <- matrix(0, nrow = n_cycles + 1, ncol = n_states, dimnames = list(NULL, states))
  state_trace[1, "ad_mci"] <- cohort_size
  
  qalys <- numeric(n_cycles)
  costs <- numeric(n_cycles)
  pys <- matrix(0, nrow = n_cycles, ncol = 4, dimnames = list(NULL, states[1:4]))  # excluding death
  
  deaths_total <- numeric(n_cycles)
  deaths_ad_mci <- numeric(n_cycles)
  
  # --- Simulation Loop ---
  for (cycle in 1:n_cycles) {
    # --- Age in current cycle ---
    current_age <- start_age + cycle - 1
    
    # --- Project to next state distribution ---
    state_trace[cycle + 1, ] <- state_trace[cycle, ] %*% tp[, , cycle]
    
    # --- QALYs ---
    qalys[cycle] <- sum(
      state_trace[cycle, "ad_mci"] * u_ad_mci(current_age),
      state_trace[cycle, "mild"] * u_mild(current_age),
      state_trace[cycle, "moderate"] * u_moderate(current_age),
      state_trace[cycle, "severe"] * u_severe(current_age)
    )
    
    # --- Costs ---
    age_index <- which(cost_by_age_state$age == current_age)
    cost_vector <- as.numeric(cost_by_age_state[age_index, c("ad_mci", "mild", "moderate", "severe", "death")])
    
    if (strategy == "intervention") {
      cost_vector["ad_mci"] <- cost_vector["ad_mci"] + dmt_annual_cost
      cost_vector["mild"] <- cost_vector["mild"] + dmt_annual_cost
    }
    
    costs[cycle] <- sum(state_trace[cycle, ] * cost_vector)
    
    # --- Person-years in each non-death state ---
    pys[cycle, "ad_mci"] <- state_trace[cycle, "ad_mci"]
    pys[cycle, "mild"] <- state_trace[cycle, "mild"]
    pys[cycle, "moderate"] <- state_trace[cycle, "moderate"]
    pys[cycle, "severe"] <- state_trace[cycle, "severe"]
    
    # --- Total deaths this cycle ---
    deaths_total[cycle] <- state_trace[cycle + 1, "death"] - state_trace[cycle, "death"]
    
    # --- Cumulative deaths from AD-MCI ---
    deaths_ad_mci[cycle] <- state_trace[cycle, "ad_mci"] * tp["ad_mci", "death", cycle]
    
  }
  
  # --- Half-cycle correction ---
  qalys_hcc <- 0.5 * (qalys[-n_cycles] + qalys[-1])
  costs_hcc <- 0.5 * (costs[-n_cycles] + costs[-1])
  pys_hcc <- 0.5 * (pys[-n_cycles, ] + pys[-1, ])
  
  # --- Discounting ---
  discount_factors <- 1 / ((1 + discount_rate) ^ (1:(n_cycles - 1)))
  qalys_disc <- sum(qalys_hcc * discount_factors) / cohort_size
  costs_disc <- sum(costs_hcc * discount_factors) / cohort_size
  
  # --- Results ---
  results <- list(
    qalys = qalys_disc,
    final_costs = costs_disc,
    pys_ad_mci = sum(pys_hcc[, "ad_mci"]) / cohort_size,
    pys_mild = sum(pys_hcc[, "mild"]) / cohort_size,
    pys_mod = sum(pys_hcc[, "moderate"]) / cohort_size,
    pys_sev = sum(pys_hcc[, "severe"]) / cohort_size,
    deaths_total = sum(deaths_total),
    deaths_ad_mci = sum(deaths_ad_mci)
  )
  
  return(results)
}

# --- Run model for both strategies ---
results <- list()
results$no_intervention <- run_markov_model("no_intervention")
results$intervention <- run_markov_model("intervention")
```
