## Supplementary material for "Replicating Health-Economic Simulation Models for Alzheimer’s Disease Using Artificial Intelligence": final R scripts: Wimo_O13.R

```
# Health-economic Markov Model Input Parameters from Wimo et al. (2020)

# --- Descriptive variables ---
states <- c("ad_mci", "mild", "moderate", "severe", "death")
strategies <- c("no_intervention", "intervention")

# --- Model configuration ---
start_age <- 60
cycle_length <- 1  # years
n_cycles <- 40
discount_rate <- 0.03
cohort_size <- 100000

# --- QALY utility functions per state (from Supplementary Table 6 regression formulas) ---
u_ad_mci <- function(age) {
  -0.002464 * age + 0.962679
}

u_mild <- function(age) {
  -0.0025 * age + 0.8293
}

u_moderate <- function(age) {
  -4e-07 * age^2 - 0.0024 * age + 0.5828
}

u_severe <- function(age) {
  -4e-07 * age^2 - 0.0025 * age + 0.4337
}

u_death <- function(age) {
  0
}

# --- Treatment cost for hypothetical disease-modifying treatment (DMT) ---
cost_dmt_per_year <- 50000  # in SEK

# --- Base case transition probabilities (defined separately as named variables) ---
# These are fixed for coding; actual time-dependency will be implemented later

# Transition probabilities (1-year) from Supplementary Table 3
p_mild_to_mild <- 0.706
p_mild_to_moderate <- 0.293
p_mild_to_severe <- 0.001

p_moderate_to_mild <- 0.087
p_moderate_to_moderate <- 0.803
p_moderate_to_severe <- 0.109

p_severe_to_moderate <- 0.196
p_severe_to_severe <- 0.804

# Annual progression probability from AD-MCI to mild dementia (base case: fixed)
# Derived from 3-year risk of 50% using p_1year = 1 - (1 - p_3year)^(1/3)
p_ad_mci_to_mild <- 1 - (1 - 0.50)^(1 / 3)  # ≈ 0.206

# --- Hazard ratios for mortality by cognitive state (Supplementary Table 4) ---
# These are relative to "very mild" AD (MMSE 27–30), approximated as baseline
hr_mild <- 1.318
hr_moderate <- 2.419
hr_severe <- 4.267

# --- Relative risks (RRs) applied for intervention effect in base case ---
# 25% reduction in risk = RR = 0.75
rr_ad_mci_to_mild <- 0.75
rr_mild_to_moderate <- 0.75
# Load mortality and weight data
mortality_rates <- read.csv("mortalityRates_Swedish_2016.csv", stringsAsFactors = FALSE)
weights <- read.csv("weightsMortalityRates_Swedish_2016.csv", stringsAsFactors = FALSE)

# Define age mapping from age groups to individual years
age_groups <- list(
  "60-64" = 60:64,
  "65-69" = 65:69,
  "70-74" = 70:74,
  "75-79" = 75:79,
  "80-84" = 80:84,
  "85-89" = 85:89,
  "90+"   = 90:99
)

# Expand grouped mortality rates to individual ages
expanded_rates <- data.frame()
for (i in 1:nrow(mortality_rates)) {
  group <- mortality_rates$age[i]
  ages <- age_groups[[group]]
  for (a in ages) {
    expanded_rates <- rbind(
      expanded_rates,
      data.frame(
        age = a,
        rate_male = mortality_rates$male[i],
        rate_female = mortality_rates$female[i]
      )
    )
  }
}

# Merge with weights
weights$age <- as.integer(weights$age)
expanded_rates$age <- as.integer(expanded_rates$age)
merged <- merge(expanded_rates, weights, by = "age")

# Compute sex-weighted average mortality rate per age
merged$rate_avg <- (merged$rate_male * merged$weight_male + merged$rate_female * merged$weight_female)

# Apply hazard ratios to compute death probabilities for each severity
# Hazard ratios previously defined:
# hr_mild <- 1.318
# hr_moderate <- 2.419
# hr_severe <- 4.267

merged$prob_death_ad_mci <- 1 - exp(-merged$rate_avg * 1)  # HR = 1
merged$prob_death_mild <- 1 - exp(-merged$rate_avg * 1.318)
merged$prob_death_moderate <- 1 - exp(-merged$rate_avg * 2.419)
merged$prob_death_severe <- 1 - exp(-merged$rate_avg * 4.267)

# Select final columns
mortality_df <- merged[, c("age", "rate_avg", "prob_death_ad_mci", "prob_death_mild", "prob_death_moderate", "prob_death_severe")]

# Resulting data frame: mortality_df
# --- Time-dependent transition probabilities ---

# Initialize vectors of length n_cycles
p_ad_mci_to_mild_no_int <- rep(p_ad_mci_to_mild, n_cycles)
p_ad_mci_stay_no_int <- 1 - p_ad_mci_to_mild_no_int

p_mild_to_moderate_no_int <- rep(p_mild_to_moderate, n_cycles)
p_mild_to_severe_no_int <- rep(p_mild_to_severe, n_cycles)
p_mild_stay_no_int <- 1 - (p_mild_to_moderate_no_int + p_mild_to_severe_no_int)

p_moderate_to_mild_no_int <- rep(p_moderate_to_mild, n_cycles)
p_moderate_to_severe_no_int <- rep(p_moderate_to_severe, n_cycles)
p_moderate_stay_no_int <- 1 - (p_moderate_to_mild_no_int + p_moderate_to_severe_no_int)

p_severe_to_moderate_no_int <- rep(p_severe_to_moderate, n_cycles)
p_severe_stay_no_int <- 1 - p_severe_to_moderate_no_int

# --- Intervention strategy: apply RR adjustments to relevant transitions ---

# From "ad_mci" to "mild"
rate_ad_mci_to_mild <- -log(1 - p_ad_mci_to_mild_no_int)
adj_rate_ad_mci_to_mild <- rate_ad_mci_to_mild * rr_ad_mci_to_mild
p_ad_mci_to_mild_int <- rep(1 - exp(-adj_rate_ad_mci_to_mild), n_cycles)
p_ad_mci_stay_int <- 1 - p_ad_mci_to_mild_int

# From "mild" to "moderate"
rate_mild_to_moderate <- -log(1 - p_mild_to_moderate)
adj_rate_mild_to_moderate <- rate_mild_to_moderate * rr_mild_to_moderate
p_mild_to_moderate_int <- rep(1 - exp(-adj_rate_mild_to_moderate), n_cycles)

# "mild" to "severe" unchanged
p_mild_to_severe_int <- rep(p_mild_to_severe, n_cycles)

# Stay in "mild"
p_mild_stay_int <- 1 - (p_mild_to_moderate_int + p_mild_to_severe_int)

# Transitions from "moderate" and "severe" remain the same in intervention
p_moderate_to_mild_int <- p_moderate_to_mild_no_int
p_moderate_to_severe_int <- p_moderate_to_severe_no_int
p_moderate_stay_int <- p_moderate_stay_no_int

p_severe_to_moderate_int <- p_severe_to_moderate_no_int
p_severe_stay_int <- p_severe_stay_no_int
# Function to build the transition probability array for a given strategy
build_transition_matrix <- function(strat) {
  n_states <- length(states)
  trans_mat <- array(0, dim = c(n_states, n_states, n_cycles),
                     dimnames = list(states, states, NULL))
  
  for (t in 1:n_cycles) {
    
    # Initialize transitions to death based on mortality probabilities (from prompt 2)
    trans_mat["ad_mci", "death", t]    <- mortality_df$prob_death_ad_mci[t]
    trans_mat["mild", "death", t]      <- mortality_df$prob_death_mild[t]
    trans_mat["moderate", "death", t]  <- mortality_df$prob_death_moderate[t]
    trans_mat["severe", "death", t]    <- mortality_df$prob_death_severe[t]
    trans_mat["death", "death", t]     <- 1  # Death is absorbing
    
    # Transitions from "ad_mci"
    if (strat == "no_intervention") {
      trans_mat["ad_mci", "mild", t] <- p_ad_mci_to_mild_no_int[t] * (1 - mortality_df$prob_death_ad_mci[t])
      trans_mat["ad_mci", "ad_mci", t] <- p_ad_mci_stay_no_int[t] * (1 - mortality_df$prob_death_ad_mci[t])
    } else if (strat == "intervention") {
      trans_mat["ad_mci", "mild", t] <- p_ad_mci_to_mild_int[t] * (1 - mortality_df$prob_death_ad_mci[t])
      trans_mat["ad_mci", "ad_mci", t] <- p_ad_mci_stay_int[t] * (1 - mortality_df$prob_death_ad_mci[t])
    }
    
    # Transitions from "mild"
    if (strat == "no_intervention") {
      trans_mat["mild", "moderate", t] <- p_mild_to_moderate_no_int[t] * (1 - mortality_df$prob_death_mild[t])
      trans_mat["mild", "severe", t]   <- p_mild_to_severe_no_int[t] * (1 - mortality_df$prob_death_mild[t])
      trans_mat["mild", "mild", t]     <- p_mild_stay_no_int[t] * (1 - mortality_df$prob_death_mild[t])
    } else if (strat == "intervention") {
      trans_mat["mild", "moderate", t] <- p_mild_to_moderate_int[t] * (1 - mortality_df$prob_death_mild[t])
      trans_mat["mild", "severe", t]   <- p_mild_to_severe_int[t] * (1 - mortality_df$prob_death_mild[t])
      trans_mat["mild", "mild", t]     <- p_mild_stay_int[t] * (1 - mortality_df$prob_death_mild[t])
    }
    
    # Transitions from "moderate"
    trans_mat["moderate", "mild", t]     <- p_moderate_to_mild_no_int[t] * (1 - mortality_df$prob_death_moderate[t])
    trans_mat["moderate", "severe", t]   <- p_moderate_to_severe_no_int[t] * (1 - mortality_df$prob_death_moderate[t])
    trans_mat["moderate", "moderate", t] <- p_moderate_stay_no_int[t] * (1 - mortality_df$prob_death_moderate[t])
    
    # Transitions from "severe"
    trans_mat["severe", "moderate", t] <- p_severe_to_moderate_no_int[t] * (1 - mortality_df$prob_death_severe[t])
    trans_mat["severe", "severe", t]   <- p_severe_stay_no_int[t] * (1 - mortality_df$prob_death_severe[t])
    
    # Normalize rows to ensure each sums to 1
    for (s in states) {
      row_sum <- sum(trans_mat[s, , t])
      if (row_sum != 1 && row_sum > 0) {
        trans_mat[s, , t] <- trans_mat[s, , t] / row_sum
      }
    }
  }
  
  return(trans_mat)
}
# Define age vector
ages <- 60:100

# Compute costs for each cognitive state using provided equations
cost_ad_mci     <- 37.961 * ages^2 - 1234.5 * ages + 124934
cost_mild       <- 49.314 * ages^2 - 1828.7 * ages + 166239
cost_moderate   <- 61.412 * ages^2 - 2086.3 * ages + 205578
cost_severe     <- 131670 * exp(0.0176 * ages)
cost_death      <- rep(0, length(ages))

# Combine into a data frame
annual_costs_df <- data.frame(
  age = ages,
  ad_mci = cost_ad_mci,
  mild = cost_mild,
  moderate = cost_moderate,
  severe = cost_severe,
  death = cost_death
)
# Function to run Markov model for a given strategy
run_markov_model <- function(strat) {
  # Initialize transition matrix for the strategy
  trans_mat <- build_transition_matrix(strat)
  
  # Initialize state trace matrix
  trace <- matrix(0, nrow = n_cycles + 1, ncol = length(states), dimnames = list(0:n_cycles, states))
  trace[1, "ad_mci"] <- cohort_size
  
  # Initialize outcome tracking
  qalys <- rep(0, n_cycles)
  costs <- rep(0, n_cycles)
  pys <- matrix(0, nrow = n_cycles, ncol = 4, dimnames = list(NULL, states[1:4]))
  deaths_total <- rep(0, n_cycles)
  deaths_ad_mci <- rep(0, n_cycles)
  
  for (t in 1:n_cycles) {
    # Age at current cycle
    age <- start_age + t - 1
    
    # Project state distribution to next cycle
    trace[t + 1, ] <- trace[t, ] %*% trans_mat[, , t]
    
    # QALYs (per state × state utility)
    qalys[t] <- trace[t, "ad_mci"] * u_ad_mci(age) +
      trace[t, "mild"]   * u_mild(age) +
      trace[t, "moderate"] * u_moderate(age) +
      trace[t, "severe"] * u_severe(age)
    
    # Costs using precomputed annual_costs_df
    cost_row <- annual_costs_df[annual_costs_df$age == age, ]
    cycle_cost <- trace[t, "ad_mci"] * cost_row$ad_mci +
      trace[t, "mild"]   * cost_row$mild +
      trace[t, "moderate"] * cost_row$moderate +
      trace[t, "severe"] * cost_row$severe
    
    # Add treatment cost for intervention strategy during "ad_mci" and "mild"
    if (strat == "intervention") {
      cycle_cost <- cycle_cost +
        trace[t, "ad_mci"] * cost_dmt_per_year +
        trace[t, "mild"]   * cost_dmt_per_year
    }
    costs[t] <- cycle_cost
    
    # Person-years per health state
    pys[t, "ad_mci"]   <- trace[t, "ad_mci"]
    pys[t, "mild"]     <- trace[t, "mild"]
    pys[t, "moderate"] <- trace[t, "moderate"]
    pys[t, "severe"]   <- trace[t, "severe"]
    
    # Deaths from all causes
    deaths_total[t] <- trace[t + 1, "death"] - trace[t, "death"]
    
    # Deaths directly from AD-MCI
    deaths_ad_mci[t] <- trace[t, "ad_mci"] * trans_mat["ad_mci", "death", t]
  }
  
  # Half-cycle correction
  qalys <- (qalys[-n_cycles] + qalys[-1]) / 2
  costs <- (costs[-n_cycles] + costs[-1]) / 2
  pys <- (pys[-n_cycles, , drop = FALSE] + pys[-1, , drop = FALSE]) / 2
  deaths_total <- deaths_total[-n_cycles]
  deaths_ad_mci <- deaths_ad_mci[-n_cycles]
  trace <- trace[-(n_cycles + 1), , drop = FALSE]
  
  # Apply discounting
  discount_factors <- 1 / ((1 + discount_rate) ^ (0:(n_cycles - 2))) #Needed to change the -1 into -2 !!
  qalys_discounted <- sum(qalys * discount_factors) / cohort_size
  costs_discounted <- sum(costs * discount_factors) / cohort_size
  
  # Return results
  return(list(
    qalys = qalys_discounted,
    final_costs = costs_discounted,
    pys_ad_mci = sum(pys[, "ad_mci"]) / cohort_size,
    pys_mild = sum(pys[, "mild"]) / cohort_size,
    pys_mod = sum(pys[, "moderate"]) / cohort_size,
    pys_sev = sum(pys[, "severe"]) / cohort_size,
    deaths_total = sum(deaths_total),
    deaths_ad_mci = sum(deaths_ad_mci)
  ))
}

# Run model for both strategies
results <- list()
results$no_intervention <- run_markov_model("no_intervention")
results$intervention <- run_markov_model("intervention")
```
