## Supplementary material for "Replicating Health-Economic Simulation Models for Alzheimer’s Disease Using Artificial Intelligence": final R scripts: Wimo_O14.R

```
# Load required parameters for the Wimo et al. (2020) health economic Markov model

# ---- Descriptive Variables ----
state_names <- c("ad_mci", "mild", "moderate", "severe", "death")
strategy_names <- c("no_intervention", "intervention")

# ---- Model Settings ----
start_age <- 60
cycle_length <- 1        # in years
n_cycles <- 40
discount_rate <- 0.03
cohort_size <- 100000

# ---- Transition Probabilities (from base case and ordered probit model) ----
# From Supplementary Table 3
p.mild_to_mild <- 0.706
p.mild_to_moderate <- 0.293
p.mild_to_severe <- 0.001

p.moderate_to_mild <- 0.087
p.moderate_to_moderate <- 0.803
p.moderate_to_severe <- 0.109

p.severe_to_moderate <- 0.196
p.severe_to_severe <- 0.804

# From core article: annual transition probability from AD-MCI to mild
p.ad_mci_to_mild <- 0.206
# Mortality from AD-MCI assumed to match general population (to be derived from life table)

# ---- Hazard Ratios for Mortality (from Supplementary Table 4) ----
HR_mild <- 1.318
HR_moderate <- 2.419
HR_severe <- 4.267

# ---- Hypothetical Disease Modifying Treatment (DMT) ----
dmt_annual_cost_sek <- 50000

# ---- Relative Risks (RRs) for Intervention (25% risk reduction in base case) ----
rr.ad_mci_to_mild <- 0.75
rr.mild_to_moderate <- 0.75

# ---- QALY Utility Functions (from Supplementary Table 6 regression equations) ----
u_ad_mci <- function(age) {
  -0.002464 * age + 0.962679
}

u_mild <- function(age) {
  -0.0025 * age + 0.8293
}

u_moderate <- function(age) {
  -4e-07 * age^2 - 0.0024 * age + 0.5828
}

u_severe <- function(age) {
  -4e-07 * age^2 - 0.0025 * age + 0.4337
}

u_death <- function(age) {
  0
}
# ---- Load Mortality Rate and Weight Files ----
mortality_rates <- read.csv("mortalityRates_Swedish_2016.csv", stringsAsFactors = FALSE)
weights <- read.csv("weightsMortalityRates_Swedish_2016.csv", stringsAsFactors = FALSE)

# ---- Define Age Groups Mapping ----
age_groups <- list(
  "60-64" = 60:64,
  "65-69" = 65:69,
  "70-74" = 70:74,
  "75-79" = 75:79,
  "80-84" = 80:84,
  "85-89" = 85:89,
  "90+"   = 90:99
)

# ---- Expand Grouped Mortality Rates to Individual Ages ----
mortality_expanded <- data.frame(age = integer(0), male = numeric(0), female = numeric(0))

for (group in names(age_groups)) {
  ages <- age_groups[[group]]
  rate_male <- mortality_rates[mortality_rates$age == group, "male"]
  rate_female <- mortality_rates[mortality_rates$age == group, "female"]
  temp <- data.frame(age = ages, male = rep(rate_male, length(ages)), female = rep(rate_female, length(ages)))
  mortality_expanded <- rbind(mortality_expanded, temp)
}

# ---- Merge Weights with Expanded Mortality ----
merged <- merge(mortality_expanded, weights, by = "age")

# ---- Compute Sex-Weighted Average Mortality Rate per Age ----
merged$rate_avg <- with(merged, male * weight_male + female * weight_female)

# ---- Compute Death Probabilities by Disease Severity ----
HR_mild <- 1.318
HR_moderate <- 2.419
HR_severe <- 4.267

merged$prob_death_ad_mci <- 1 - exp(-merged$rate_avg)
merged$prob_death_mild <- 1 - exp(-merged$rate_avg * HR_mild)
merged$prob_death_moderate <- 1 - exp(-merged$rate_avg * HR_moderate)
merged$prob_death_severe <- 1 - exp(-merged$rate_avg * HR_severe)

# ---- Final Data Frame ----
mortality_final <- merged[, c("age", "rate_avg", "prob_death_ad_mci", "prob_death_mild", "prob_death_moderate", "prob_death_severe")]
# ---- Time-Dependent Transition Probability Vectors ----

# Vectors for "no intervention" strategy
p_ad_mci_to_mild_vec <- rep(p.ad_mci_to_mild, n_cycles)
p_ad_mci_to_ad_mci_vec <- 1 - p_ad_mci_to_mild_vec

p_mild_to_moderate_vec <- rep(p.mild_to_moderate, n_cycles)
p_mild_to_severe_vec <- rep(p.mild_to_severe, n_cycles)
p_mild_to_mild_vec <- 1 - (p_mild_to_moderate_vec + p_mild_to_severe_vec)

p_moderate_to_mild_vec <- rep(p.moderate_to_mild, n_cycles)
p_moderate_to_severe_vec <- rep(p.moderate_to_severe, n_cycles)
p_moderate_to_moderate_vec <- 1 - (p_moderate_to_mild_vec + p_moderate_to_severe_vec)

p_severe_to_moderate_vec <- rep(p.severe_to_moderate, n_cycles)
p_severe_to_severe_vec <- 1 - p_severe_to_moderate_vec

# Vectors for "intervention" strategy (adjusting only ad_mci and mild transitions)
rate_ad_mci_to_mild <- -log(1 - p.ad_mci_to_mild)
adjusted_rate_ad_mci_to_mild <- rate_ad_mci_to_mild * rr.ad_mci_to_mild
p_ad_mci_to_mild_int_vec <- rep(1 - exp(-adjusted_rate_ad_mci_to_mild), n_cycles)
p_ad_mci_to_ad_mci_int_vec <- 1 - p_ad_mci_to_mild_int_vec

rate_mild_to_moderate <- -log(1 - p.mild_to_moderate)
adjusted_rate_mild_to_moderate <- rate_mild_to_moderate * rr.mild_to_moderate
p_mild_to_moderate_int_vec <- rep(1 - exp(-adjusted_rate_mild_to_moderate), n_cycles)
p_mild_to_severe_int_vec <- p_mild_to_severe_vec  # unaffected by DMT
p_mild_to_mild_int_vec <- 1 - (p_mild_to_moderate_int_vec + p_mild_to_severe_int_vec)

# Moderate and severe transitions remain unchanged
p_moderate_to_mild_int_vec <- p_moderate_to_mild_vec
p_moderate_to_severe_int_vec <- p_moderate_to_severe_vec
p_moderate_to_moderate_int_vec <- p_moderate_to_moderate_vec

p_severe_to_moderate_int_vec <- p_severe_to_moderate_vec
p_severe_to_severe_int_vec <- p_severe_to_severe_vec
# ---- Function to Build Transition Probability Matrix ----

build_transition_matrix <- function(strat) {
  states <- c("ad_mci", "mild", "moderate", "severe", "death")
  n_states <- length(states)
  tpm <- array(0, dim = c(n_states, n_states, n_cycles),
               dimnames = list(from = states, to = states, cycle = 1:n_cycles))
  
  for (cycle in 1:n_cycles) {
    
    # Assign probabilities to death (absorbing state)
    tpm["ad_mci", "death", cycle]     <- mortality_final$prob_death_ad_mci[cycle]
    tpm["mild", "death", cycle]       <- mortality_final$prob_death_mild[cycle]
    tpm["moderate", "death", cycle]   <- mortality_final$prob_death_moderate[cycle]
    tpm["severe", "death", cycle]     <- mortality_final$prob_death_severe[cycle]
    tpm["death", "death", cycle]      <- 1
    
    # ---- Transition Definitions for Living States ----
    if (strat == "no_intervention") {
      # AD-MCI
      tpm["ad_mci", "mild", cycle]   <- p_ad_mci_to_mild_vec[cycle] * (1 - tpm["ad_mci", "death", cycle])
      tpm["ad_mci", "ad_mci", cycle] <- p_ad_mci_to_ad_mci_vec[cycle] * (1 - tpm["ad_mci", "death", cycle])
      
      # Mild
      tpm["mild", "moderate", cycle] <- p_mild_to_moderate_vec[cycle] * (1 - tpm["mild", "death", cycle])
      tpm["mild", "severe", cycle]   <- p_mild_to_severe_vec[cycle]   * (1 - tpm["mild", "death", cycle])
      tpm["mild", "mild", cycle]     <- p_mild_to_mild_vec[cycle]     * (1 - tpm["mild", "death", cycle])
      
    } else if (strat == "intervention") {
      # AD-MCI
      tpm["ad_mci", "mild", cycle]   <- p_ad_mci_to_mild_int_vec[cycle]   * (1 - tpm["ad_mci", "death", cycle])
      tpm["ad_mci", "ad_mci", cycle] <- p_ad_mci_to_ad_mci_int_vec[cycle] * (1 - tpm["ad_mci", "death", cycle])
      
      # Mild
      tpm["mild", "moderate", cycle] <- p_mild_to_moderate_int_vec[cycle] * (1 - tpm["mild", "death", cycle])
      tpm["mild", "severe", cycle]   <- p_mild_to_severe_int_vec[cycle]   * (1 - tpm["mild", "death", cycle])
      tpm["mild", "mild", cycle]     <- p_mild_to_mild_int_vec[cycle]     * (1 - tpm["mild", "death", cycle])
    }
    
    # Moderate (same for both strategies)
    tpm["moderate", "mild", cycle]     <- p_moderate_to_mild_vec[cycle]     * (1 - tpm["moderate", "death", cycle])
    tpm["moderate", "severe", cycle]   <- p_moderate_to_severe_vec[cycle]   * (1 - tpm["moderate", "death", cycle])
    tpm["moderate", "moderate", cycle] <- p_moderate_to_moderate_vec[cycle] * (1 - tpm["moderate", "death", cycle])
    
    # Severe (same for both strategies)
    tpm["severe", "moderate", cycle] <- p_severe_to_moderate_vec[cycle] * (1 - tpm["severe", "death", cycle])
    tpm["severe", "severe", cycle]   <- p_severe_to_severe_vec[cycle]   * (1 - tpm["severe", "death", cycle])
    
    # Normalize rows (ensure sum = 1)
    for (s in states) {
      row_sum <- sum(tpm[s, , cycle])
      if (row_sum > 0 && abs(row_sum - 1) > 1e-8) {
        tpm[s, , cycle] <- tpm[s, , cycle] / row_sum
      }
    }
  }
  
  return(tpm)
}
# ---- Compute Age- and State-Specific Annual Societal Costs (SEK) ----

ages <- 60:100

# Calculate costs per state using the given equations
cost_ad_mci     <- 37.961 * ages^2 - 1234.5 * ages + 124934
cost_mild       <- 49.314 * ages^2 - 1828.7 * ages + 166239
cost_moderate   <- 61.412 * ages^2 - 2086.3 * ages + 205578
cost_severe     <- 131670 * exp(0.0176 * ages)
cost_death      <- rep(0, length(ages))

# Create the data frame
annual_costs <- data.frame(
  age       = ages,
  ad_mci    = cost_ad_mci,
  mild      = cost_mild,
  moderate  = cost_moderate,
  severe    = cost_severe,
  death     = cost_death
)
# ---- Function to Run Markov Model for a Given Strategy ----

run_markov_model <- function(strategy_name) {
  states <- c("ad_mci", "mild", "moderate", "severe", "death")
  n_states <- length(states)
  
  # Initialize trace matrix
  trace <- matrix(0, nrow = n_cycles + 1, ncol = n_states,
                  dimnames = list(0:n_cycles, states))
  trace[1, "ad_mci"] <- cohort_size
  
  # Initialize outcomes
  qalys <- numeric(n_cycles + 1)
  costs <- numeric(n_cycles + 1)
  pys <- matrix(0, nrow = n_cycles + 1, ncol = 4,
                dimnames = list(0:n_cycles, c("ad_mci", "mild", "moderate", "severe")))
  deaths_total <- numeric(n_cycles + 1)
  deaths_ad_mci <- numeric(n_cycles + 1)
  
  # Get transition matrix
  tpm <- build_transition_matrix(strategy_name)
  
  for (cycle in 1:n_cycles) {
    # Advance state
    trace[cycle + 1, ] <- trace[cycle, ] %*% tpm[, , cycle]
    
    # Compute current age
    current_age <- start_age + cycle - 1
    
    # Compute QALYs (utility weighted by state distribution)
    qalys[cycle + 1] <- trace[cycle, "ad_mci"] * u_ad_mci(current_age) +
      trace[cycle, "mild"]   * u_mild(current_age) +
      trace[cycle, "moderate"] * u_moderate(current_age) +
      trace[cycle, "severe"] * u_severe(current_age)
    
    # Compute Costs using pre-computed annual_costs table
    age_row <- which(annual_costs$age == current_age)
    cost_vec <- c(
      ad_mci  = annual_costs$ad_mci[age_row],
      mild    = annual_costs$mild[age_row],
      moderate= annual_costs$moderate[age_row],
      severe  = annual_costs$severe[age_row],
      death   = 0
    )
    
    # Add DMT cost in ad_mci and mild for intervention strategy
    if (strategy_name == "intervention") {
      cost_vec["ad_mci"] <- cost_vec["ad_mci"] + dmt_annual_cost_sek
      cost_vec["mild"]   <- cost_vec["mild"]   + dmt_annual_cost_sek
    }
    
    costs[cycle + 1] <- sum(trace[cycle, ] * cost_vec)
    
    # Record person-years
    pys[cycle + 1, "ad_mci"]   <- trace[cycle, "ad_mci"]
    pys[cycle + 1, "mild"]     <- trace[cycle, "mild"]
    pys[cycle + 1, "moderate"] <- trace[cycle, "moderate"]
    pys[cycle + 1, "severe"]   <- trace[cycle, "severe"]
    
    # Death tracking
    deaths_total[cycle + 1] <- trace[cycle + 1, "death"] - trace[cycle, "death"]
    deaths_ad_mci[cycle + 1] <- trace[cycle, "ad_mci"] * tpm["ad_mci", "death", cycle]
  }
  
  # ---- Half-cycle correction ----
  qalys_hcc <- 0.5 * (qalys[-length(qalys)] + qalys[-1])
  costs_hcc <- 0.5 * (costs[-length(costs)] + costs[-1])
  pys_hcc <- 0.5 * (pys[-nrow(pys), ] + pys[-1, ])
  deaths_total <- deaths_total[-1]
  deaths_ad_mci <- deaths_ad_mci[-1]
  trace <- trace[-1, ]
  
  # ---- Apply Discounting ----
  discount_vector <- 1 / ((1 + discount_rate) ^ (1:n_cycles))
  qalys_discounted <- sum(qalys_hcc * discount_vector)
  costs_discounted <- sum(costs_hcc * discount_vector)
  
  # ---- Return Results ----
  return(list(
    qalys = qalys_discounted / cohort_size,
    final_costs = costs_discounted / cohort_size,
    pys_ad_mci = sum(pys_hcc[, "ad_mci"]) / cohort_size,
    pys_mild = sum(pys_hcc[, "mild"]) / cohort_size,
    pys_mod = sum(pys_hcc[, "moderate"]) / cohort_size,
    pys_sev = sum(pys_hcc[, "severe"]) / cohort_size,
    deaths_total = sum(deaths_total),
    deaths_ad_mci = sum(deaths_ad_mci)
  ))
}

# ---- Run the Model for Both Strategies ----
results <- list()
results$no_intervention <- run_markov_model("no_intervention")
results$intervention <- run_markov_model("intervention")
```
