## Supplementary material for "Replicating Health-Economic Simulation Models for Alzheimer’s Disease Using Artificial Intelligence": final R scripts: Wimo_O15.R

```
# =============================
# Wimo et al. (2020) - Input Parameters for Markov Model
# =============================

# State names
state_names <- c("ad_mci", "mild", "moderate", "severe", "death")

# Strategy names
strategies <- c("no_intervention", "intervention")

# Model assumptions
start_age <- 60
cycle_length <- 1      # 1 year
n_cycles <- 40         # total 40 years
discount_rate <- 0.03  # for both costs and QALYs
cohort_size <- 100000

# Disease progression: Annual probability of progression from AD-MCI to mild AD (base case)
# Derived from 3-year risk of 50% using: p1year = 1 - (1 - p3year)^(1/3)
p.ad_mci_to_mild <- 1 - (1 - 0.50)^(1 / 3)  # ≈ 0.206

# Transition probabilities between dementia states (from Supplementary Table 3)
p.mild_to_mild <- 0.706
p.mild_to_moderate <- 0.293
p.mild_to_severe <- 0.001

p.moderate_to_mild <- 0.087
p.moderate_to_moderate <- 0.803
p.moderate_to_severe <- 0.109

p.severe_to_moderate <- 0.196
p.severe_to_severe <- 0.804

# Hypothetical intervention effect (Base case scenario)
# 25% reduction in progression from AD-MCI to Mild and from Mild to Moderate
rr.ad_mci_to_mild <- 0.75
rr.mild_to_moderate <- 0.75

# Hypothetical annual treatment cost for the Disease Modifying Treatment (DMT)
dmt_annual_cost <- 50000  # in SEK

# Hazard ratios for mortality (from Supplementary Table 4)
# Reference: Very mild (MMSE 27–30)
hr.mild <- 1.318
hr.moderate <- 2.419
hr.severe <- 4.267

# =============================
# Age-dependent utility functions (from Supplementary Table 6)
# =============================

u_ad_mci <- function(age) {
  -0.002464 * age + 0.962679
}

u_mild <- function(age) {
  -0.0025 * age + 0.8293
}

u_moderate <- function(age) {
  -4e-07 * age^2 - 0.0024 * age + 0.5828
}

u_severe <- function(age) {
  -4e-07 * age^2 - 0.0025 * age + 0.4337
}

u_death <- function(age) {
  0
}
# =============================
# Mortality Rate Processing for Markov Model
# =============================

# Load mortality rate files
mort_rates <- read.csv("mortalityRates_Swedish_2016.csv", stringsAsFactors = FALSE)
weights <- read.csv("weightsMortalityRates_Swedish_2016.csv", stringsAsFactors = FALSE)

# Define age group to individual age mapping
age_groups <- list(
  "60-64" = 60:64,
  "65-69" = 65:69,
  "70-74" = 70:74,
  "75-79" = 75:79,
  "80-84" = 80:84,
  "85-89" = 85:89,
  "90+"   = 90:99
)

# Expand grouped mortality rates to individual ages
expanded <- data.frame()
for (group in names(age_groups)) {
  ages <- age_groups[[group]]
  row <- mort_rates[mort_rates$age == group, ]
  df <- data.frame(
    age = ages,
    male = rep(row$male, length(ages)),
    female = rep(row$female, length(ages))
  )
  expanded <- rbind(expanded, df)
}

# Merge with weights data
merged <- merge(expanded, weights, by = "age")

# Calculate weighted average mortality rate per age
merged$rate_avg <- with(merged,
                        (male * weight_male + female * weight_female) /
                          (weight_male + weight_female))

# Define hazard ratios (already provided in previous prompt)
# hr.mild, hr.moderate, hr.severe are assumed to be already defined

# Compute death probabilities using the formula:
# p_death = 1 - exp(-rate * HR)
merged$prob_death_ad_mci <- 1 - exp(-merged$rate_avg * 1)           # HR = 1 for AD-MCI
merged$prob_death_mild   <- 1 - exp(-merged$rate_avg * hr.mild)
merged$prob_death_moderate <- 1 - exp(-merged$rate_avg * hr.moderate)
merged$prob_death_severe <- 1 - exp(-merged$rate_avg * hr.severe)

# Final data frame
mortality_df <- merged[, c("age", "rate_avg",
                           "prob_death_ad_mci",
                           "prob_death_mild",
                           "prob_death_moderate",
                           "prob_death_severe")]
# =============================
# Time-Dependent Transition Probability Vectors
# =============================

# Number of cycles
# Assumed previously defined: n_cycles

# === No Intervention Strategy ===

# AD-MCI state
p_ad_mci_to_mild_noint <- rep(p.ad_mci_to_mild, n_cycles)
p_ad_mci_to_ad_mci_noint <- 1 - p_ad_mci_to_mild_noint

# Mild state
p_mild_to_moderate_noint <- rep(p.mild_to_moderate, n_cycles)
p_mild_to_severe_noint <- rep(p.mild_to_severe, n_cycles)
p_mild_to_mild_noint <- 1 - p_mild_to_moderate_noint - p_mild_to_severe_noint

# Moderate state
p_moderate_to_mild_noint <- rep(p.moderate_to_mild, n_cycles)
p_moderate_to_severe_noint <- rep(p.moderate_to_severe, n_cycles)
p_moderate_to_moderate_noint <- 1 - p_moderate_to_mild_noint - p_moderate_to_severe_noint

# Severe state
p_severe_to_moderate_noint <- rep(p.severe_to_moderate, n_cycles)
p_severe_to_severe_noint <- 1 - p_severe_to_moderate_noint

# === Intervention Strategy ===

# AD-MCI state with intervention (apply RR)
rate_ad_mci_to_mild <- -log(1 - p.ad_mci_to_mild)
rate_ad_mci_to_mild_int <- rate_ad_mci_to_mild * rr.ad_mci_to_mild
p_ad_mci_to_mild_int <- rep(1 - exp(-rate_ad_mci_to_mild_int), n_cycles)
p_ad_mci_to_ad_mci_int <- 1 - p_ad_mci_to_mild_int

# Mild state with intervention (apply RR)
rate_mild_to_moderate <- -log(1 - p.mild_to_moderate)
rate_mild_to_moderate_int <- rate_mild_to_moderate * rr.mild_to_moderate
p_mild_to_moderate_int <- rep(1 - exp(-rate_mild_to_moderate_int), n_cycles)
p_mild_to_severe_int <- rep(p.mild_to_severe, n_cycles)  # unchanged
p_mild_to_mild_int <- 1 - p_mild_to_moderate_int - p_mild_to_severe_int

# Moderate state unchanged
p_moderate_to_mild_int <- p_moderate_to_mild_noint
p_moderate_to_severe_int <- p_moderate_to_severe_noint
p_moderate_to_moderate_int <- p_moderate_to_moderate_noint

# Severe state unchanged
p_severe_to_moderate_int <- p_severe_to_moderate_noint
p_severe_to_severe_int <- p_severe_to_severe_noint
# =============================
# Transition Probability Matrix Builder
# =============================

build_transition_matrix <- function(strat) {
  # Define state names
  states <- c("ad_mci", "mild", "moderate", "severe", "death")
  n_states <- length(states)
  
  # Initialize transition probability array: [from, to, cycle]
  tpm <- array(0, dim = c(n_states, n_states, n_cycles),
               dimnames = list(from = states, to = states, cycle = 1:n_cycles))
  
  for (t in 1:n_cycles) {
    # Probability of death from each state
    tpm["ad_mci", "death", t]    <- mortality_df$prob_death_ad_mci[t]
    tpm["mild", "death", t]      <- mortality_df$prob_death_mild[t]
    tpm["moderate", "death", t]  <- mortality_df$prob_death_moderate[t]
    tpm["severe", "death", t]    <- mortality_df$prob_death_severe[t]
    tpm["death", "death", t]     <- 1  # death is absorbing
    
    # === Transition probabilities for living states (adjusted for mortality) ===
    
    if (strat == "no_intervention") {
      # AD-MCI transitions
      tpm["ad_mci", "mild", t]   <- p_ad_mci_to_mild_noint[t] * (1 - mortality_df$prob_death_ad_mci[t])
      tpm["ad_mci", "ad_mci", t] <- p_ad_mci_to_ad_mci_noint[t] * (1 - mortality_df$prob_death_ad_mci[t])
      
      # Mild transitions
      tpm["mild", "moderate", t] <- p_mild_to_moderate_noint[t] * (1 - mortality_df$prob_death_mild[t])
      tpm["mild", "severe", t]   <- p_mild_to_severe_noint[t] * (1 - mortality_df$prob_death_mild[t])
      tpm["mild", "mild", t]     <- p_mild_to_mild_noint[t] * (1 - mortality_df$prob_death_mild[t])
      
      # Moderate transitions
      tpm["moderate", "mild", t]     <- p_moderate_to_mild_noint[t] * (1 - mortality_df$prob_death_moderate[t])
      tpm["moderate", "severe", t]   <- p_moderate_to_severe_noint[t] * (1 - mortality_df$prob_death_moderate[t])
      tpm["moderate", "moderate", t] <- p_moderate_to_moderate_noint[t] * (1 - mortality_df$prob_death_moderate[t])
      
      # Severe transitions
      tpm["severe", "moderate", t] <- p_severe_to_moderate_noint[t] * (1 - mortality_df$prob_death_severe[t])
      tpm["severe", "severe", t]   <- p_severe_to_severe_noint[t] * (1 - mortality_df$prob_death_severe[t])
      
    } else if (strat == "intervention") {
      # AD-MCI transitions
      tpm["ad_mci", "mild", t]   <- p_ad_mci_to_mild_int[t] * (1 - mortality_df$prob_death_ad_mci[t])
      tpm["ad_mci", "ad_mci", t] <- p_ad_mci_to_ad_mci_int[t] * (1 - mortality_df$prob_death_ad_mci[t])
      
      # Mild transitions
      tpm["mild", "moderate", t] <- p_mild_to_moderate_int[t] * (1 - mortality_df$prob_death_mild[t])
      tpm["mild", "severe", t]   <- p_mild_to_severe_int[t] * (1 - mortality_df$prob_death_mild[t])
      tpm["mild", "mild", t]     <- p_mild_to_mild_int[t] * (1 - mortality_df$prob_death_mild[t])
      
      # Moderate transitions
      tpm["moderate", "mild", t]     <- p_moderate_to_mild_int[t] * (1 - mortality_df$prob_death_moderate[t])
      tpm["moderate", "severe", t]   <- p_moderate_to_severe_int[t] * (1 - mortality_df$prob_death_moderate[t])
      tpm["moderate", "moderate", t] <- p_moderate_to_moderate_int[t] * (1 - mortality_df$prob_death_moderate[t])
      
      # Severe transitions
      tpm["severe", "moderate", t] <- p_severe_to_moderate_int[t] * (1 - mortality_df$prob_death_severe[t])
      tpm["severe", "severe", t]   <- p_severe_to_severe_int[t] * (1 - mortality_df$prob_death_severe[t])
    }
    
    # Normalize rows to ensure probabilities sum to 1
    for (s in states) {
      row_sum <- sum(tpm[s, , t])
      if (row_sum > 0 && row_sum != 1) {
        tpm[s, , t] <- tpm[s, , t] / row_sum
      }
    }
  }
  
  return(tpm)
}
# =============================
# Age- and Cognitive State-Specific Annual Costs (SEK)
# =============================

# Define age range
ages <- 60:100

# Compute costs for each state using the given GLM equations
cost_ad_mci    <-  37.961 * ages^2 - 1234.5 * ages + 124934
cost_mild      <-  49.314 * ages^2 - 1828.7 * ages + 166239
cost_moderate  <-  61.412 * ages^2 - 2086.3 * ages + 205578
cost_severe    <- 131670 * exp(0.0176 * ages)
cost_death     <- rep(0, length(ages))

# Combine into a data frame
costs_df <- data.frame(
  age       = ages,
  ad_mci    = cost_ad_mci,
  mild      = cost_mild,
  moderate  = cost_moderate,
  severe    = cost_severe,
  death     = cost_death
)
# =============================
# Markov Model Runner Function (Structure)
# =============================

run_markov_model <- function(strat, tpm, utility_functions, cost_table) {
  # Define states
  states <- c("ad_mci", "mild", "moderate", "severe", "death")
  living_states <- states[states != "death"]
  
  # Initialize state trace matrix
  trace <- matrix(0, nrow = n_cycles + 1, ncol = length(states),
                  dimnames = list(0:n_cycles, states))
  trace[1, "ad_mci"] <- cohort_size  # start entire cohort in ad_mci
  
  # Initialize outputs
  qalys <- numeric(n_cycles)
  costs <- numeric(n_cycles)
  pys <- matrix(0, nrow = n_cycles, ncol = length(living_states),
                dimnames = list(1:n_cycles, living_states))
  deaths_total <- numeric(n_cycles)
  deaths_ad_mci <- numeric(n_cycles)
  
  # Discount vector (excluding last cycle due to HCC)
  discount_factors <- 1 / (1 + discount_rate)^(0:(n_cycles - 2)) #Needed to change the minus -1 into -2 !!
  
  for (t in 1:n_cycles) {
    # Update cohort distribution
    trace[t + 1, ] <- trace[t, ] %*% tpm[, , t]
    
    # Current age
    current_age <- start_age + t - 1
    
    # --- QALYs ---
    qalys[t] <- trace[t, "ad_mci"]   * utility_functions[[1]](current_age) +
      trace[t, "mild"]     * utility_functions[[2]](current_age) +
      trace[t, "moderate"] * utility_functions[[3]](current_age) +
      trace[t, "severe"]   * utility_functions[[4]](current_age)
    
    # --- Costs ---
    age_row <- cost_table[cost_table$age == current_age, ]
    costs[t] <- trace[t, "ad_mci"]   * age_row$ad_mci +
      trace[t, "mild"]     * age_row$mild +
      trace[t, "moderate"] * age_row$moderate +
      trace[t, "severe"]   * age_row$severe
    
    if (strat == "intervention") {
      costs[t] <- costs[t] + 
        trace[t, "ad_mci"] * dmt_annual_cost +
        trace[t, "mild"]   * dmt_annual_cost
    }
    
    # --- Person-Years ---
    for (state in living_states) {
      pys[t, state] <- trace[t, state]
    }
    
    # --- Total Deaths ---
    deaths_total[t] <- trace[t + 1, "death"] - trace[t, "death"]
    
    # --- Cumulative Deaths from AD-MCI ---
    deaths_ad_mci[t] <- trace[t, "ad_mci"] * tpm["ad_mci", "death", t]
  }
  
  # Half-cycle correction: average current and next cycle, then drop last
  qalys <- 0.5 * (qalys[-1] + qalys[-length(qalys)])
  costs <- 0.5 * (costs[-1] + costs[-length(costs)])
  pys <- 0.5 * (pys[-1, , drop = FALSE] + pys[-n_cycles, , drop = FALSE])
  deaths_total <- deaths_total[-n_cycles]
  deaths_ad_mci <- deaths_ad_mci[-n_cycles]
  trace <- trace[-(n_cycles + 1), , drop = FALSE]
  
  # Apply discounting
  qalys_disc <- qalys * discount_factors
  costs_disc <- costs * discount_factors
  
  # Return results
  return(list(
    qalys = sum(qalys_disc) / cohort_size,
    final_costs = sum(costs_disc) / cohort_size,
    pys_ad_mci = sum(pys[, "ad_mci"]) / cohort_size,
    pys_mild = sum(pys[, "mild"]) / cohort_size,
    pys_mod = sum(pys[, "moderate"]) / cohort_size,
    pys_sev = sum(pys[, "severe"]) / cohort_size,
    deaths_total = sum(deaths_total),
    deaths_ad_mci = sum(deaths_ad_mci)
  ))
}

# Run the model for both strategies and store outputs
results <- list()
results$no_intervention <- run_markov_model("no_intervention", build_transition_matrix("no_intervention"), 
                                            utility_functions = list(u_ad_mci, u_mild, u_moderate, u_severe), 
                                            cost_table = costs_df)

results$intervention <- run_markov_model("intervention", build_transition_matrix("intervention"), 
                                         utility_functions = list(u_ad_mci, u_mild, u_moderate, u_severe), 
                                         cost_table = costs_df)
```
