## Supplementary material for "Replicating Health-Economic Simulation Models for Alzheimer’s Disease Using Artificial Intelligence": final R scripts: validation15Trials_Wimo.R

```
##### ──────────────────────────────────────────────────────────────
#####  Wimo Model Replication – Comparison with Reference Results
##### ──────────────────────────────────────────────────────────────

# Reference results from Wimo et al. (per individual)
reference_values <- list(
  no_intervention = list(
    qalys        = 7.90,
    final_costs  = 4503751,
    pys_ad_mci   = 4.20,
    pys_mild     = 4.97,
    pys_mod      = 7.31,
    pys_sev      = 2.50,
    deaths_total = 99987,
    deaths_ad_mci = 4030
  ),
  intervention = list(
    qalys        = 8.63,
    final_costs  = 4893703,
    pys_ad_mci   = 5.64,
    pys_mild     = 5.83,
    pys_mod      = 6.14,
    pys_sev      = 2.02,
    deaths_total = 99969,
    deaths_ad_mci = 6473
  )
)

# Vector of simulation script names
sim_names <- paste0("Wimo_O", 1:15)

# Metrics to compare
metrics <- c("qalys", "final_costs",
             "pys_ad_mci", "pys_mild", "pys_mod", "pys_sev",
             "deaths_total", "deaths_ad_mci")

# Function to compare simulation outputs against reference
compare_simulation <- function(res, ref) {
  comparison <- c()
  for (strategy in c("no_intervention", "intervention")) {
    for (metric in metrics) {
      value    <- res[[strategy]][[metric]]
      diff_abs <- value - ref[[strategy]][[metric]]
      diff_rel <- diff_abs / ref[[strategy]][[metric]]
      comparison <- c(comparison, value, diff_abs, diff_rel)
    }
  }
  return(comparison)
}

# Run each simulation file and compare against reference values
results_matrix <- t(sapply(sim_names, function(file_name) {
  env <- new.env()
  sys.source(paste0(file_name, ".R"), envir = env)
  compare_simulation(env$results, reference_values)
}))

# Generate column names for results
column_names <- c()
for (strategy in c("no_intervention", "intervention")) {
  for (metric in metrics) {
    column_names <- c(column_names,
                      paste0(strategy, "_", metric),           # raw value
                      paste0(strategy, "_", metric, "_diff_abs"), # absolute diff
                      paste0(strategy, "_", metric, "_diff_rel")) # relative diff
  }
}

# Final results as data frame
comparison_df <- as.data.frame(results_matrix)
colnames(comparison_df) <- column_names
rownames(comparison_df) <- sim_names

# View in RStudio
View(comparison_df)
```
