## Supplementary material for "Replicating Health-Economic Simulation Models for Alzheimer’s Disease Using Artificial Intelligence": final R scripts: Ross_O1.R

```
# Markov Model Input Parameters from Ross et al. (2022)

# ----------------------------
# State and Strategy Names
# ----------------------------
state_names <- c("mci", "mild", "moderate", "severe", "death")
strategy_names <- c("standard_care", "aducanumab", "donanemab")

# ----------------------------
# Cohort Characteristics
# ----------------------------
initial_age <- 75.2
cohort_size <- 1000
prop_mci <- 0.65
prop_mild <- 0.35

# ----------------------------
# Disease Progression Probabilities (monthly)
# ----------------------------
p.mci_to_mild <- 0.007
p.mild_to_moderate <- 0.016
p.moderate_to_severe <- 0.026

# ----------------------------
# Treatment Effects (Hazard Ratios)
# ----------------------------
hr_progression_aducanumab <- 0.89
hr_progression_donanemab <- 0.68

# ----------------------------
# Utilities (converted from annual to monthly)
# ----------------------------
u.mci <- 0.73 / 12
u.mild <- 0.69 / 12
u.moderate <- 0.53 / 12
u.severe <- 0.34 / 12

# ARIA-related utility decrement (applied for 3 months when symptomatic ARIA occurs)
aria_utility_decrement <- -0.065 / 12

# ----------------------------
# Background Healthcare Costs (monthly, in 2020 USD)
# ----------------------------
cost.healthcare.mci <- 691
cost.healthcare.mild <- 1049
cost.healthcare.moderate <- 1274
cost.healthcare.severe <- 3764

# Additional societal costs
cost.societal.mci <- 49
cost.societal.mild <- 1483
cost.societal.moderate <- 1932
cost.societal.severe <- 2153

# ----------------------------
# Drug Costs
# ----------------------------
# Annual drug costs
cost.drug.aducanumab.year1 <- 20500
cost.drug.aducanumab.subsequent <- 28200
cost.drug.donanemab <- 28200

# ----------------------------
# Infusion and Monitoring Costs (monthly)
# ----------------------------
# Aducanumab
cost.infusion.aducanumab.m1_6 <- 112
cost.infusion.aducanumab.m7_12 <- 87
cost.infusion.aducanumab.m13_plus <- 80

# Donanemab
cost.infusion.donanemab.m1_6 <- 599
cost.infusion.donanemab.m7_12 <- 302
cost.infusion.donanemab.m13_plus <- 287

# ----------------------------
# Screening Costs (one-time per eligible patient)
# ----------------------------
cost.screening.aducanumab <- 6957
cost.screening.donanemab <- 17096

# ----------------------------
# ARIA-Related Costs
# ----------------------------
cost.aria.imaging <- 1060           # follow-up MRI
cost.aria.hospitalization <- 35934  # 11.6 days * $3092/day
cost.aria.outpatient <- 128         # one physician visit

# ----------------------------
# ARIA Event Probabilities
# ----------------------------
prob.aria.aducanumab <- 0.41
prob.aria.donanemab <- 0.39
prob.aria.symptomatic.aducanumab <- 0.41 * 0.24
prob.aria.symptomatic.donanemab <- 0.39 * 0.16


# Load mortality rate data
mortality_data <- read.csv("mortalityRates_US_2017.csv")

# Define state-specific hazard ratios
hr.mci <- 1.61
hr.mild <- 2.23
hr.moderate <- 3.10
hr.severe <- 4.98

# Compute age-specific probabilities of death for each state
mortality_processed <- data.frame(
  age = mortality_data$age,
  prob_death_mci = 1 - exp(-mortality_data$monthly_rate * hr.mci),
  prob_death_mild = 1 - exp(-mortality_data$monthly_rate * hr.mild),
  prob_death_moderate = 1 - exp(-mortality_data$monthly_rate * hr.moderate),
  prob_death_severe = 1 - exp(-mortality_data$monthly_rate * hr.severe)
)


# Define number of cycles
n_cycles <- 480  # 40 years * 12 months

# ---- Standard Care (no intervention) ----
p_mci_to_mild_std <- rep(p.mci_to_mild, n_cycles)
p_mci_stay_std <- rep(1 - p.mci_to_mild, n_cycles)

p_mild_to_mod_std <- rep(p.mild_to_moderate, n_cycles)
p_mild_stay_std <- rep(1 - p.mild_to_moderate, n_cycles)

p_mod_to_sev_std <- rep(p.moderate_to_severe, n_cycles)
p_mod_stay_std <- rep(1 - p.moderate_to_severe, n_cycles)

p_sev_stay_std <- rep(1.0, n_cycles)  # No progression from severe (excluding death)

# ---- Aducanumab Intervention ----
rate_mci_to_mild <- -log(1 - p.mci_to_mild)
rate_mild_to_mod <- -log(1 - p.mild_to_moderate)
rate_mod_to_sev <- -log(1 - p.moderate_to_severe)

adj_rate_mci_to_mild_adu <- rate_mci_to_mild * hr_progression_aducanumab
adj_rate_mild_to_mod_adu <- rate_mild_to_mod * hr_progression_aducanumab
adj_rate_mod_to_sev_adu <- rate_mod_to_sev * hr_progression_aducanumab

p_mci_to_mild_adu <- rep(1 - exp(-adj_rate_mci_to_mild_adu), n_cycles)
p_mci_stay_adu <- rep(1 - (1 - exp(-adj_rate_mci_to_mild_adu)), n_cycles)

p_mild_to_mod_adu <- rep(1 - exp(-adj_rate_mild_to_mod_adu), n_cycles)
p_mild_stay_adu <- rep(1 - (1 - exp(-adj_rate_mild_to_mod_adu)), n_cycles)

p_mod_to_sev_adu <- rep(1 - exp(-adj_rate_mod_to_sev_adu), n_cycles)
p_mod_stay_adu <- rep(1 - (1 - exp(-adj_rate_mod_to_sev_adu)), n_cycles)

p_sev_stay_adu <- rep(1.0, n_cycles)

# ---- Donanemab Intervention ----
adj_rate_mci_to_mild_don <- rate_mci_to_mild * hr_progression_donanemab
adj_rate_mild_to_mod_don <- rate_mild_to_mod * hr_progression_donanemab
adj_rate_mod_to_sev_don <- rate_mod_to_sev * hr_progression_donanemab

p_mci_to_mild_don <- rep(1 - exp(-adj_rate_mci_to_mild_don), n_cycles)
p_mci_stay_don <- rep(1 - (1 - exp(-adj_rate_mci_to_mild_don)), n_cycles)

p_mild_to_mod_don <- rep(1 - exp(-adj_rate_mild_to_mod_don), n_cycles)
p_mild_stay_don <- rep(1 - (1 - exp(-adj_rate_mild_to_mod_don)), n_cycles)

p_mod_to_sev_don <- rep(1 - exp(-adj_rate_mod_to_sev_don), n_cycles)
p_mod_stay_don <- rep(1 - (1 - exp(-adj_rate_mod_to_sev_don)), n_cycles)

p_sev_stay_don <- rep(1.0, n_cycles)


# Function to build transition matrix array
build_transition_matrix <- function(strat) {
  n_states <- length(state_names)
  P <- array(0, dim = c(n_states, n_states, n_cycles),
             dimnames = list(state_names, state_names, NULL))
  
  for (t in 1:n_cycles) {
    # Current age for this cycle
    current_age <- initial_age + (t - 1) / 12
    current_age_floor <- floor(current_age)
    
    # Ensure age is within bounds
    if (current_age_floor < min(mortality_processed$age)) {
      age_idx <- which.min(mortality_processed$age)
    } else if (current_age_floor > max(mortality_processed$age)) {
      age_idx <- which.max(mortality_processed$age)
    } else {
      age_idx <- which(mortality_processed$age == current_age_floor)
    }
    
    # Transition probabilities to death by state
    p_death_mci <- mortality_processed$prob_death_mci[age_idx]
    p_death_mild <- mortality_processed$prob_death_mild[age_idx]
    p_death_moderate <- mortality_processed$prob_death_moderate[age_idx]
    p_death_severe <- mortality_processed$prob_death_severe[age_idx]
    
    # Assign death transition probabilities
    P["mci", "death", t] <- p_death_mci
    P["mild", "death", t] <- p_death_mild
    P["moderate", "death", t] <- p_death_moderate
    P["severe", "death", t] <- p_death_severe
    P["death", "death", t] <- 1
    
    # Transitions from "mci"
    if (strat == "no intervention") {
      p_prog <- p_mci_to_mild_std[t]
      p_stay <- p_mci_stay_std[t]
    } else if (strat == "aducanumab") {
      p_prog <- p_mci_to_mild_adu[t]
      p_stay <- p_mci_stay_adu[t]
    } else if (strat == "donanemab") {
      p_prog <- p_mci_to_mild_don[t]
      p_stay <- p_mci_stay_don[t]
    }
    P["mci", "mild", t] <- p_prog * (1 - p_death_mci)
    P["mci", "mci", t] <- p_stay * (1 - p_death_mci)
    
    # Transitions from "mild"
    if (strat == "no intervention") {
      p_prog <- p_mild_to_mod_std[t]
      p_stay <- p_mild_stay_std[t]
    } else if (strat == "aducanumab") {
      p_prog <- p_mild_to_mod_adu[t]
      p_stay <- p_mild_stay_adu[t]
    } else if (strat == "donanemab") {
      p_prog <- p_mild_to_mod_don[t]
      p_stay <- p_mild_stay_don[t]
    }
    P["mild", "moderate", t] <- p_prog * (1 - p_death_mild)
    P["mild", "mild", t] <- p_stay * (1 - p_death_mild)
    
    # Transitions from "moderate"
    if (strat == "no intervention") {
      p_prog <- p_mod_to_sev_std[t]
      p_stay <- p_mod_stay_std[t]
    } else if (strat == "aducanumab") {
      p_prog <- p_mod_to_sev_adu[t]
      p_stay <- p_mod_stay_adu[t]
    } else if (strat == "donanemab") {
      p_prog <- p_mod_to_sev_don[t]
      p_stay <- p_mod_stay_don[t]
    }
    P["moderate", "severe", t] <- p_prog * (1 - p_death_moderate)
    P["moderate", "moderate", t] <- p_stay * (1 - p_death_moderate)
    
    # Transitions from "severe" (only staying state is allowed besides death)
    if (strat == "no intervention") {
      p_stay <- p_sev_stay_std[t]
    } else if (strat == "aducanumab") {
      p_stay <- p_sev_stay_adu[t]
    } else if (strat == "donanemab") {
      p_stay <- p_sev_stay_don[t]
    }
    P["severe", "severe", t] <- p_stay * (1 - p_death_severe)
    
    # Normalize each row if necessary
    for (s in state_names) {
      row_sum <- sum(P[s, , t])
      if (row_sum > 0 && row_sum != 1) {
        P[s, , t] <- P[s, , t] / row_sum
      }
    }
  }
  
  return(P)
}


# ------------------------------
# Set Discounting Parameters
# ------------------------------
annual_discount_rate <- 0.03
monthly_discount_factor <- 1 / ((1 + annual_discount_rate)^(1 / 12))
monthly_discount_weights <- monthly_discount_factor ^ (0:(n_cycles - 1))

# ------------------------------
# Initial State Distribution
# ------------------------------
initial_distribution <- c(mci = prop_mci, mild = prop_mild, moderate = 0, severe = 0, death = 0)

# ------------------------------
# ARIA Costs and QALY Decrement
# ------------------------------
cost_aria_total <- cost.aria.outpatient + cost.aria.imaging + (cost.aria.hospitalization * 0.04)
aria_qaly_decrement <- aria_utility_decrement

# ------------------------------
# Results Storage
# ------------------------------
results <- list()

# ------------------------------
# Loop Over Strategies
# ------------------------------
for (strategy in c("no intervention", "aducanumab", "donanemab")) {
  
  P <- build_transition_matrix(strategy)
  trace <- matrix(0, nrow = n_cycles, ncol = length(state_names),
                  dimnames = list(NULL, state_names))
  trace[1, ] <- initial_distribution * cohort_size
  
  on_treatment <- rep(0, n_cycles)
  if (strategy != "no intervention") {
    on_treatment[1] <- sum(trace[1, c("mci", "mild", "moderate")])
  }
  
  qalys <- rep(0, n_cycles)
  healthcare_costs <- rep(0, n_cycles)
  societal_costs <- rep(0, n_cycles)
  
  # One-time screening costs
  if (strategy == "aducanumab") {
    healthcare_costs[1] <- healthcare_costs[1] + cost.screening.aducanumab * cohort_size
    societal_costs[1] <- societal_costs[1] + cost.screening.aducanumab * cohort_size
  } else if (strategy == "donanemab") {
    healthcare_costs[1] <- healthcare_costs[1] + cost.screening.donanemab * cohort_size
    societal_costs[1] <- societal_costs[1] + cost.screening.donanemab * cohort_size
  }
  
  for (t in 2:n_cycles) {
    trace[t, ] <- trace[t - 1, ] %*% P[, , t - 1]
    
    living_states <- trace[t, c("mci", "mild", "moderate", "severe")]
    utilities <- c(u.mci, u.mild, u.moderate, u.severe)
    qaly_base <- sum(living_states * utilities)
    
    if (strategy != "no intervention" && t <= 3) {
      if (strategy == "aducanumab") {
        aria_symp_rate <- prob.aria.symptomatic.aducanumab / 3
      } else {
        aria_symp_rate <- prob.aria.symptomatic.donanemab / 3
      }
      n_aria_cases <- on_treatment[t - 1] * aria_symp_rate
      qaly_base <- qaly_base + (n_aria_cases * aria_qaly_decrement)
    }
    qalys[t] <- qaly_base
    
    # Background costs
    hc_state_costs <- c(cost.healthcare.mci, cost.healthcare.mild,
                        cost.healthcare.moderate, cost.healthcare.severe)
    sc_state_costs <- c(cost.societal.mci, cost.societal.mild,
                        cost.societal.moderate, cost.societal.severe)
    
    healthcare_costs[t] <- healthcare_costs[t] + sum(living_states * hc_state_costs)
    societal_costs[t] <- societal_costs[t] + sum(living_states * (hc_state_costs + sc_state_costs))
    
    # Treatment status
    if (strategy == "aducanumab") {
      pct_remaining <- 1 - (0.38 * min(t - 1, 12) / 12)
      on_treatment[t] <- sum(trace[t, c("mci", "mild", "moderate")]) * pct_remaining
      drug_cost <- ifelse(t <= 12, cost.drug.aducanumab.year1 / 12, cost.drug.aducanumab.subsequent / 12)
      inf_cost <- ifelse(t <= 6, cost.infusion.aducanumab.m1_6,
                         ifelse(t <= 12, cost.infusion.aducanumab.m7_12, cost.infusion.aducanumab.m13_plus))
      
    } else if (strategy == "donanemab") {
      if (t <= 6) {
        pct_remaining <- 1 - 0.27 * (t - 1) / 6
      } else if (t <= 12) {
        pct_remaining <- 0.73 - 0.55 * (t - 6) / 6
      } else {
        pct_remaining <- 0.18
      }
      on_treatment[t] <- sum(trace[t, c("mci", "mild", "moderate")]) * pct_remaining
      drug_cost <- cost.drug.donanemab / 12
      inf_cost <- ifelse(t <= 6, cost.infusion.donanemab.m1_6,
                         ifelse(t <= 12, cost.infusion.donanemab.m7_12, cost.infusion.donanemab.m13_plus))
      
    } else {
      next
    }
    
    healthcare_costs[t] <- healthcare_costs[t] + on_treatment[t] * (drug_cost + inf_cost)
    societal_costs[t] <- societal_costs[t] + on_treatment[t] * (drug_cost + inf_cost)
    
    # ARIA-related costs
    if (t <= 3) {
      n_aria_cases <- on_treatment[t] * aria_symp_rate
      healthcare_costs[t] <- healthcare_costs[t] + n_aria_cases * cost_aria_total
      societal_costs[t] <- societal_costs[t] + n_aria_cases * cost_aria_total
    }
  }
  
  # Apply discounting and aggregate
  results[[gsub(" ", "_", strategy)]] <- list(
    qalys = sum(qalys * monthly_discount_weights) / cohort_size,
    healthcare_costs = sum(healthcare_costs * monthly_discount_weights) / cohort_size,
    societal_costs = sum(societal_costs * monthly_discount_weights) / cohort_size
  )
}

# Display results
results
```
