## Supplementary material for "Replicating Health-Economic Simulation Models for Alzheimer’s Disease Using Artificial Intelligence": final R scripts: Ross_O2.R

```
# Input Parameters for Health Economic Markov Model (based on Ross et al. 2022)

# State names
states <- c("mci", "mild", "moderate", "severe", "death")

# Strategy names
strategies <- c("standard_care", "aducanumab", "donanemab")

# Simulation settings
initial_age <- 75.2
cohort_size <- 1000
discount_rate <- 0.03
cycle_length <- 1 / 12  # monthly cycles

# Starting distribution
start_mci <- 0.65
start_mild <- 0.35

# Monthly disease progression probabilities (standard care)
p.mci_to_mild <- 0.007
p.mild_to_moderate <- 0.016
p.moderate_to_severe <- 0.026

# Hazard ratios for disease progression
hr.aducanumab <- 0.89
hr.donanemab <- 0.68

# Mortality hazard ratios
hr.mci <- 1.61
hr.mild <- 2.23
hr.moderate <- 3.10
hr.severe <- 4.98

# Annual utilities (converted to monthly)
u.mci <- 0.73 / 12
u.mild <- 0.69 / 12
u.moderate <- 0.53 / 12
u.severe <- 0.34 / 12
u.aria_decrement <- -0.065 / 12

# Monthly background healthcare costs (USD, 2020)
c.healthcare.mci <- 691
c.healthcare.mild <- 1049
c.healthcare.moderate <- 1274
c.healthcare.severe <- 3764

# Monthly additional societal costs (USD, 2020)
c.societal.mci <- 49
c.societal.mild <- 1483
c.societal.moderate <- 1932
c.societal.severe <- 2153

# Annual drug costs (USD)
c.drug.aducanumab.year1 <- 20500
c.drug.aducanumab.later <- 28200
c.drug.donanemab <- 28200

# Monthly infusion and monitoring costs (USD)
c.infusion.aducanumab.m1_6 <- 112
c.infusion.aducanumab.m7_12 <- 87
c.infusion.aducanumab.later <- 80

c.infusion.donanemab.m1_6 <- 599
c.infusion.donanemab.m7_12 <- 302
c.infusion.donanemab.later <- 287

# One-time screening costs (USD)
c.screening.aducanumab <- 6957
c.screening.donanemab <- 17096

# ARIA-related costs
c.aria.followup_imaging <- 1060
c.aria.hospitalization <- 35934
c.aria.outpatient_visit <- 128
p.aria.aducanumab <- 0.41
p.aria.donanemab <- 0.39
p.aria.symptomatic.aducanumab <- 0.24
p.aria.symptomatic.donanemab <- 0.16


# Load necessary library
library(readr)
library(dplyr)

# Load mortality rates from CSV
mortality_data <- read_csv("mortalityRates_US_2017.csv")

# Compute age-specific probability of death for each disease severity state
mortality_processed <- mortality_data %>%
  mutate(
    prob_death_mci = 1 - exp(-monthly_rate * hr.mci),
    prob_death_mild = 1 - exp(-monthly_rate * hr.mild),
    prob_death_moderate = 1 - exp(-monthly_rate * hr.moderate),
    prob_death_severe = 1 - exp(-monthly_rate * hr.severe)
  )

# View first few rows of the processed mortality data (optional)
# head(mortality_processed)


# Number of cycles (e.g., 30 years * 12 months = 360 cycles)
n_cycles <- 360

# --- Standard Care ---
p_mci_to_mild_std <- rep(p.mci_to_mild, n_cycles)
p_mild_to_moderate_std <- rep(p.mild_to_moderate, n_cycles)
p_moderate_to_severe_std <- rep(p.moderate_to_severe, n_cycles)

p_mci_stay_std <- 1 - p_mci_to_mild_std
p_mild_stay_std <- 1 - p_mild_to_moderate_std
p_moderate_stay_std <- 1 - p_moderate_to_severe_std
p_severe_stay_std <- rep(1, n_cycles)

# --- Aducanumab ---
rate_mci_to_mild <- -log(1 - p.mci_to_mild)
rate_mild_to_moderate <- -log(1 - p.mild_to_moderate)
rate_moderate_to_severe <- -log(1 - p.moderate_to_severe)

adj_rate_mci_to_mild_adu <- rate_mci_to_mild * hr.aducanumab
adj_rate_mild_to_moderate_adu <- rate_mild_to_moderate * hr.aducanumab
adj_rate_moderate_to_severe_adu <- rate_moderate_to_severe * hr.aducanumab

p_mci_to_mild_adu <- rep(1 - exp(-adj_rate_mci_to_mild_adu), n_cycles)
p_mild_to_moderate_adu <- rep(1 - exp(-adj_rate_mild_to_moderate_adu), n_cycles)
p_moderate_to_severe_adu <- rep(1 - exp(-adj_rate_moderate_to_severe_adu), n_cycles)

p_mci_stay_adu <- 1 - p_mci_to_mild_adu
p_mild_stay_adu <- 1 - p_mild_to_moderate_adu
p_moderate_stay_adu <- 1 - p_moderate_to_severe_adu
p_severe_stay_adu <- rep(1, n_cycles)

# --- Donanemab ---
adj_rate_mci_to_mild_don <- rate_mci_to_mild * hr.donanemab
adj_rate_mild_to_moderate_don <- rate_mild_to_moderate * hr.donanemab
adj_rate_moderate_to_severe_don <- rate_moderate_to_severe * hr.donanemab

p_mci_to_mild_don <- rep(1 - exp(-adj_rate_mci_to_mild_don), n_cycles)
p_mild_to_moderate_don <- rep(1 - exp(-adj_rate_mild_to_moderate_don), n_cycles)
p_moderate_to_severe_don <- rep(1 - exp(-adj_rate_moderate_to_severe_don), n_cycles)

p_mci_stay_don <- 1 - p_mci_to_mild_don
p_mild_stay_don <- 1 - p_mild_to_moderate_don
p_moderate_stay_don <- 1 - p_moderate_to_severe_don
p_severe_stay_don <- rep(1, n_cycles)


# Function to build transition probability array for a given strategy
build_transition_matrix <- function(strat) {
  n_states <- length(states)
  trans_mat <- array(0, dim = c(n_states, n_states, n_cycles),
                     dimnames = list(from = states, to = states, cycle = 1:n_cycles))
  
  for (t in 1:n_cycles) {
    current_age <- floor(initial_age + (t - 1) / 12)
    
    if (current_age < 60) current_age <- 60
    if (current_age > 99) current_age <- 99
    
    age_row <- mortality_processed %>% filter(age == current_age)
    
    # Assign mortality probabilities
    trans_mat["mci", "death", t]      <- age_row$prob_death_mci
    trans_mat["mild", "death", t]     <- age_row$prob_death_mild
    trans_mat["moderate", "death", t] <- age_row$prob_death_moderate
    trans_mat["severe", "death", t]   <- age_row$prob_death_severe
    trans_mat["death", "death", t]    <- 1
    
    if (strat == "no intervention") {
      trans_mat["mci", "mild", t]      <- p_mci_to_mild_std[t] * (1 - trans_mat["mci", "death", t])
      trans_mat["mci", "mci", t]       <- p_mci_stay_std[t] * (1 - trans_mat["mci", "death", t])
      
      trans_mat["mild", "moderate", t] <- p_mild_to_moderate_std[t] * (1 - trans_mat["mild", "death", t])
      trans_mat["mild", "mild", t]     <- p_mild_stay_std[t] * (1 - trans_mat["mild", "death", t])
      
      trans_mat["moderate", "severe", t] <- p_moderate_to_severe_std[t] * (1 - trans_mat["moderate", "death", t])
      trans_mat["moderate", "moderate", t] <- p_moderate_stay_std[t] * (1 - trans_mat["moderate", "death", t])
      
      trans_mat["severe", "severe", t] <- p_severe_stay_std[t] * (1 - trans_mat["severe", "death", t])
      
    } else if (strat == "aducanumab") {
      trans_mat["mci", "mild", t]      <- p_mci_to_mild_adu[t] * (1 - trans_mat["mci", "death", t])
      trans_mat["mci", "mci", t]       <- p_mci_stay_adu[t] * (1 - trans_mat["mci", "death", t])
      
      trans_mat["mild", "moderate", t] <- p_mild_to_moderate_adu[t] * (1 - trans_mat["mild", "death", t])
      trans_mat["mild", "mild", t]     <- p_mild_stay_adu[t] * (1 - trans_mat["mild", "death", t])
      
      trans_mat["moderate", "severe", t] <- p_moderate_to_severe_adu[t] * (1 - trans_mat["moderate", "death", t])
      trans_mat["moderate", "moderate", t] <- p_moderate_stay_adu[t] * (1 - trans_mat["moderate", "death", t])
      
      trans_mat["severe", "severe", t] <- p_severe_stay_adu[t] * (1 - trans_mat["severe", "death", t])
      
    } else if (strat == "donanemab") {
      trans_mat["mci", "mild", t]      <- p_mci_to_mild_don[t] * (1 - trans_mat["mci", "death", t])
      trans_mat["mci", "mci", t]       <- p_mci_stay_don[t] * (1 - trans_mat["mci", "death", t])
      
      trans_mat["mild", "moderate", t] <- p_mild_to_moderate_don[t] * (1 - trans_mat["mild", "death", t])
      trans_mat["mild", "mild", t]     <- p_mild_stay_don[t] * (1 - trans_mat["mild", "death", t])
      
      trans_mat["moderate", "severe", t] <- p_moderate_to_severe_don[t] * (1 - trans_mat["moderate", "death", t])
      trans_mat["moderate", "moderate", t] <- p_moderate_stay_don[t] * (1 - trans_mat["moderate", "death", t])
      
      trans_mat["severe", "severe", t] <- p_severe_stay_don[t] * (1 - trans_mat["severe", "death", t])
    }
    
    # Normalize rows if row sum != 1
    for (s in states) {
      row_sum <- sum(trans_mat[s, , t])
      if (row_sum > 0 && abs(row_sum - 1) > 1e-8) {
        trans_mat[s, , t] <- trans_mat[s, , t] / row_sum
      }
    }
  }
  
  return(trans_mat)
}

# Required library
library(dplyr)

# Constants
monthly_discount_rate <- (1 + discount_rate)^(1 / 12) - 1
discount_weights <- 1 / ((1 + monthly_discount_rate) ^ (0:(n_cycles - 1)))

# Initial state distribution
initial_dist <- numeric(length(states))
names(initial_dist) <- states
initial_dist["mci"] <- start_mci
initial_dist["mild"] <- start_mild
initial_dist["moderate"] <- 0
initial_dist["severe"] <- 0
initial_dist["death"] <- 0

# Function to simulate one strategy
simulate_strategy <- function(strat) {
  trans <- build_transition_matrix(strat)
  trace <- matrix(0, nrow = n_cycles + 1, ncol = length(states), dimnames = list(0:n_cycles, states))
  trace[1, ] <- initial_dist * cohort_size
  
  # Treatment continuation
  treat_status <- rep(1, n_cycles + 1)  # 1 = treated, 0 = discontinued
  if (strat == "aducanumab") {
    treat_status[1:12] <- 1 - seq(0, 0.38, length.out = 12 + 1)[-1]
    treat_status[13:(n_cycles + 1)] <- treat_status[12]
  } else if (strat == "donanemab") {
    treat_status[1:6] <- 1
    treat_status[7:12] <- 1 - 0.27
    treat_status[13:(n_cycles + 1)] <- 1 - 0.55
  }
  
  qalys <- numeric(n_cycles)
  hc_costs <- numeric(n_cycles)
  sc_costs <- numeric(n_cycles)
  
  for (t in 1:n_cycles) {
    trace[t + 1, ] <- trace[t, ] %*% trans[, , t]
    
    # Determine utility and costs
    u_vec <- c(
      "mci" = u.mci,
      "mild" = u.mild,
      "moderate" = u.moderate,
      "severe" = u.severe,
      "death" = 0
    )
    
    hc_vec <- c(
      "mci" = c.healthcare.mci,
      "mild" = c.healthcare.mild,
      "moderate" = c.healthcare.moderate,
      "severe" = c.healthcare.severe,
      "death" = 0
    )
    
    sc_vec <- hc_vec + c(
      "mci" = c.societal.mci,
      "mild" = c.societal.mild,
      "moderate" = c.societal.moderate,
      "severe" = c.societal.severe,
      "death" = 0
    )
    
    # Treatment-specific costs
    treat_cost <- 0
    if (strat == "aducanumab" && treat_status[t] > 0) {
      if (t <= 12) {
        if (t <= 6) {
          infusion_cost <- c.infusion.aducanumab.m1_6
        } else {
          infusion_cost <- c.infusion.aducanumab.m7_12
        }
      } else {
        infusion_cost <- c.infusion.aducanumab.later
      }
      drug_cost <- ifelse(t <= 12, c.drug.aducanumab.year1 / 12, c.drug.aducanumab.later / 12)
      n_treated <- sum(trace[t, c("mci", "mild", "moderate")])
      treat_cost <- treat_status[t] * n_treated * (drug_cost + infusion_cost)
    }
    
    if (strat == "donanemab" && treat_status[t] > 0) {
      if (t <= 6) {
        infusion_cost <- c.infusion.donanemab.m1_6
      } else if (t <= 12) {
        infusion_cost <- c.infusion.donanemab.m7_12
      } else {
        infusion_cost <- c.infusion.donanemab.later
      }
      drug_cost <- c.drug.donanemab / 12
      n_treated <- sum(trace[t, c("mci", "mild", "moderate")])
      treat_cost <- treat_status[t] * n_treated * (drug_cost + infusion_cost)
    }
    
    # ARIA-related costs (symptomatic only, outpatient + MRI + hospitalization for 4%)
    aria_cost <- 0
    if (strat %in% c("aducanumab", "donanemab") && t <= 3) {
      p_sympt_aria <- ifelse(strat == "aducanumab", p.aria.symptomatic.aducanumab, p.aria.symptomatic.donanemab)
      n_sympt_aria <- treat_status[t] * sum(trace[t, c("mci", "mild", "moderate")]) * p_sympt_aria
      aria_cost <- n_sympt_aria * (
        c.aria.outpatient_visit + c.aria.followup_imaging + (0.04 * c.aria.hospitalization)
      )
    }
    
    # QALY decrement from symptomatic ARIA
    aria_qalys <- 0
    if (strat %in% c("aducanumab", "donanemab") && t <= 3) {
      p_sympt_aria <- ifelse(strat == "aducanumab", p.aria.symptomatic.aducanumab, p.aria.symptomatic.donanemab)
      n_sympt_aria <- treat_status[t] * sum(trace[t, c("mci", "mild", "moderate")]) * p_sympt_aria
      aria_qalys <- n_sympt_aria * u.aria_decrement
    }
    
    # Total QALYs and costs for this cycle
    qalys[t] <- (sum(trace[t, ] * u_vec) + aria_qalys) * discount_weights[t]
    hc_costs[t] <- (sum(trace[t, ] * hc_vec) + treat_cost + aria_cost) * discount_weights[t]
    sc_costs[t] <- (sum(trace[t, ] * sc_vec) + treat_cost + aria_cost) * discount_weights[t]
  }
  
  # Add one-time screening costs in first cycle
  screen_cost <- ifelse(strat == "aducanumab", c.screening.aducanumab, ifelse(strat == "donanemab", c.screening.donanemab, 0))
  hc_costs[1] <- hc_costs[1] + screen_cost * cohort_size * discount_weights[1]
  sc_costs[1] <- sc_costs[1] + screen_cost * cohort_size * discount_weights[1]
  
  return(list(
    qalys = sum(qalys) / cohort_size,
    healthcare_costs = sum(hc_costs) / cohort_size,
    societal_costs = sum(sc_costs) / cohort_size
  ))
}

# Run simulation for all strategies
results <- list()
results$no_intervention <- simulate_strategy("no intervention")
results$aducanumab <- simulate_strategy("aducanumab")
results$donanemab <- simulate_strategy("donanemab")
```
