## Supplementary material for "Replicating Health-Economic Simulation Models for Alzheimer’s Disease Using Artificial Intelligence": final R scripts: Ross_O3.R

```
# MARKOV MODEL INPUT PARAMETERS FROM ROSS ET AL. (2022)

# State names (fixed)
state_names <- c("mci", "mild", "moderate", "severe", "death")

# Strategy names
strategy_names <- c("standard_care", "aducanumab", "donanemab")

# Simulation settings
initial_age <- 75.2
cohort_size <- 1000

# Baseline health state distribution
initial_mci_prop <- 0.65
initial_mild_prop <- 0.35

# Disease progression probabilities (monthly)
p.mci_to_mild <- 0.007
p.mild_to_moderate <- 0.016
p.moderate_to_severe <- 0.026

# Treatment effect on progression (hazard ratios)
hr_aducanumab <- 0.89
hr_donanemab <- 0.68

# Utility values (converted from annual to monthly)
u.mci <- 0.73 / 12
u.mild <- 0.69 / 12
u.moderate <- 0.53 / 12
u.severe <- 0.34 / 12
u.aria_decrement <- -0.065 / 12

# Background healthcare costs (monthly, USD 2020)
cost.healthcare.mci <- 691
cost.healthcare.mild <- 1049
cost.healthcare.moderate <- 1274
cost.healthcare.severe <- 3764

# Background additional societal costs (monthly, USD 2020)
cost.societal.mci <- 49
cost.societal.mild <- 1483
cost.societal.moderate <- 1932
cost.societal.severe <- 2153

# Annual drug costs
cost.aducanumab.year1 <- 20500
cost.aducanumab.year2plus <- 28200
cost.donanemab <- 28200

# Monthly infusion and monitoring costs (by time period)
cost.aducanumab.m1_6 <- 112
cost.aducanumab.m7_12 <- 87
cost.aducanumab.m13plus <- 80
cost.donanemab.m1_6 <- 599
cost.donanemab.m7_12 <- 302
cost.donanemab.m13plus <- 287

# One-time screening costs per eligible patient
cost.screening.aducanumab <- 6957
cost.screening.donanemab <- 17096

# ARIA (amyloid-related imaging abnormality) event probabilities
p.aria_aducanumab <- 0.41
p.aria_donanemab <- 0.39
p.aria_symptomatic_aducanumab <- 0.41 * 0.24
p.aria_symptomatic_donanemab <- 0.39 * 0.16

# ARIA-related costs
cost.aria_followup_imaging <- 1060
cost.aria_outpatient_visit <- 128
cost.aria_hospitalization <- 35934

# Mortality hazard ratios by state
hr_mortality.mci <- 1.61
hr_mortality.mild <- 2.23
hr_mortality.moderate <- 3.10
hr_mortality.severe <- 4.98


# Load mortality rates CSV
mortality_data <- read.csv("mortalityRates_US_2017.csv")

# Compute age-specific death probabilities for each disease severity
mortality_data$prob_death_mci <- 1 - exp(-mortality_data$monthly_rate * hr_mortality.mci)
mortality_data$prob_death_mild <- 1 - exp(-mortality_data$monthly_rate * hr_mortality.mild)
mortality_data$prob_death_moderate <- 1 - exp(-mortality_data$monthly_rate * hr_mortality.moderate)
mortality_data$prob_death_severe <- 1 - exp(-mortality_data$monthly_rate * hr_mortality.severe)


# Number of model cycles (define as needed for full time horizon)
n_cycles <- 480  # for example, 40 years with monthly cycles

# --- STANDARD CARE (no intervention) ---

# Progression probabilities (constant per month, repeated over time)
p_mci_to_mild_standard <- rep(p.mci_to_mild, n_cycles)
p_mild_to_moderate_standard <- rep(p.mild_to_moderate, n_cycles)
p_moderate_to_severe_standard <- rep(p.moderate_to_severe, n_cycles)

# Staying probabilities
p_mci_stay_standard <- 1 - p_mci_to_mild_standard
p_mild_stay_standard <- 1 - p_mild_to_moderate_standard
p_moderate_stay_standard <- 1 - p_moderate_to_severe_standard
p_severe_stay_standard <- rep(1, n_cycles)

# --- ADUCANUMAB ---

# Apply HR to progression probabilities
r_mci_to_mild <- -log(1 - p.mci_to_mild)
r_mild_to_moderate <- -log(1 - p.mild_to_moderate)
r_moderate_to_severe <- -log(1 - p.moderate_to_severe)

r_mci_to_mild_adu <- r_mci_to_mild * hr_aducanumab
r_mild_to_moderate_adu <- r_mild_to_moderate * hr_aducanumab
r_moderate_to_severe_adu <- r_moderate_to_severe * hr_aducanumab

p_mci_to_mild_adu <- rep(1 - exp(-r_mci_to_mild_adu), n_cycles)
p_mild_to_moderate_adu <- rep(1 - exp(-r_mild_to_moderate_adu), n_cycles)
p_moderate_to_severe_adu <- rep(1 - exp(-r_moderate_to_severe_adu), n_cycles)

p_mci_stay_adu <- 1 - p_mci_to_mild_adu
p_mild_stay_adu <- 1 - p_mild_to_moderate_adu
p_moderate_stay_adu <- 1 - p_moderate_to_severe_adu
p_severe_stay_adu <- rep(1, n_cycles)

# --- DONANEMAB ---

# Apply HR to progression probabilities
r_mci_to_mild_dona <- r_mci_to_mild * hr_donanemab
r_mild_to_moderate_dona <- r_mild_to_moderate * hr_donanemab
r_moderate_to_severe_dona <- r_moderate_to_severe * hr_donanemab

p_mci_to_mild_dona <- rep(1 - exp(-r_mci_to_mild_dona), n_cycles)
p_mild_to_moderate_dona <- rep(1 - exp(-r_mild_to_moderate_dona), n_cycles)
p_moderate_to_severe_dona <- rep(1 - exp(-r_moderate_to_severe_dona), n_cycles)

p_mci_stay_dona <- 1 - p_mci_to_mild_dona
p_mild_stay_dona <- 1 - p_mild_to_moderate_dona
p_moderate_stay_dona <- 1 - p_moderate_to_severe_dona
p_severe_stay_dona <- rep(1, n_cycles)


# Function to build transition probability matrix for a given strategy
build_transition_matrix <- function(strat) {
  # Define dimensions
  n_states <- length(state_names)
  trans_matrix <- array(0, dim = c(n_states, n_states, n_cycles),
                        dimnames = list(state_names, state_names, NULL))
  
  for (t in 1:n_cycles) {
    age_t <- initial_age + (t - 1) / 12
    age_index <- which.min(abs(mortality_data$age - age_t))
    
    # Assign death probabilities per state for this cycle
    trans_matrix["mci", "death", t] <- mortality_data$prob_death_mci[age_index]
    trans_matrix["mild", "death", t] <- mortality_data$prob_death_mild[age_index]
    trans_matrix["moderate", "death", t] <- mortality_data$prob_death_moderate[age_index]
    trans_matrix["severe", "death", t] <- mortality_data$prob_death_severe[age_index]
    
    # Absorbing state
    trans_matrix["death", "death", t] <- 1
    
    if (strat == "no intervention") {
      trans_matrix["mci", "mild", t] <- p_mci_to_mild_standard[t] * (1 - trans_matrix["mci", "death", t])
      trans_matrix["mci", "mci", t] <- p_mci_stay_standard[t] * (1 - trans_matrix["mci", "death", t])
      
      trans_matrix["mild", "moderate", t] <- p_mild_to_moderate_standard[t] * (1 - trans_matrix["mild", "death", t])
      trans_matrix["mild", "mild", t] <- p_mild_stay_standard[t] * (1 - trans_matrix["mild", "death", t])
      
      trans_matrix["moderate", "severe", t] <- p_moderate_to_severe_standard[t] * (1 - trans_matrix["moderate", "death", t])
      trans_matrix["moderate", "moderate", t] <- p_moderate_stay_standard[t] * (1 - trans_matrix["moderate", "death", t])
      
      trans_matrix["severe", "severe", t] <- p_severe_stay_standard[t] * (1 - trans_matrix["severe", "death", t])
      
    } else if (strat == "aducanumab") {
      trans_matrix["mci", "mild", t] <- p_mci_to_mild_adu[t] * (1 - trans_matrix["mci", "death", t])
      trans_matrix["mci", "mci", t] <- p_mci_stay_adu[t] * (1 - trans_matrix["mci", "death", t])
      
      trans_matrix["mild", "moderate", t] <- p_mild_to_moderate_adu[t] * (1 - trans_matrix["mild", "death", t])
      trans_matrix["mild", "mild", t] <- p_mild_stay_adu[t] * (1 - trans_matrix["mild", "death", t])
      
      trans_matrix["moderate", "severe", t] <- p_moderate_to_severe_adu[t] * (1 - trans_matrix["moderate", "death", t])
      trans_matrix["moderate", "moderate", t] <- p_moderate_stay_adu[t] * (1 - trans_matrix["moderate", "death", t])
      
      trans_matrix["severe", "severe", t] <- p_severe_stay_adu[t] * (1 - trans_matrix["severe", "death", t])
      
    } else if (strat == "donanemab") {
      trans_matrix["mci", "mild", t] <- p_mci_to_mild_dona[t] * (1 - trans_matrix["mci", "death", t])
      trans_matrix["mci", "mci", t] <- p_mci_stay_dona[t] * (1 - trans_matrix["mci", "death", t])
      
      trans_matrix["mild", "moderate", t] <- p_mild_to_moderate_dona[t] * (1 - trans_matrix["mild", "death", t])
      trans_matrix["mild", "mild", t] <- p_mild_stay_dona[t] * (1 - trans_matrix["mild", "death", t])
      
      trans_matrix["moderate", "severe", t] <- p_moderate_to_severe_dona[t] * (1 - trans_matrix["moderate", "death", t])
      trans_matrix["moderate", "moderate", t] <- p_moderate_stay_dona[t] * (1 - trans_matrix["moderate", "death", t])
      
      trans_matrix["severe", "severe", t] <- p_severe_stay_dona[t] * (1 - trans_matrix["severe", "death", t])
    }
    
    # Normalize rows to sum to 1
    for (s in state_names) {
      row_sum <- sum(trans_matrix[s, , t])
      if (row_sum > 0 && row_sum != 1) {
        trans_matrix[s, , t] <- trans_matrix[s, , t] / row_sum
      }
    }
  }
  
  return(trans_matrix)
}


# Define discount factor (monthly, 3% annual)
discount_rate_monthly <- (1 - 0.03)^(1 / 12)
discount_vector <- discount_rate_monthly^(0:(n_cycles - 1))

# Initial state vector for 1000 individuals
initial_state <- c(mci = initial_mci_prop, mild = initial_mild_prop, moderate = 0, severe = 0, death = 0)
initial_state <- initial_state * cohort_size

# Function to simulate one strategy
simulate_strategy <- function(strategy_name) {
  tp_matrix <- build_transition_matrix(strategy_name)
  trace <- matrix(0, nrow = n_cycles + 1, ncol = length(state_names),
                  dimnames = list(NULL, state_names))
  trace[1, ] <- initial_state
  
  # Treatment tracking
  on_treatment <- rep(1, n_cycles + 1)
  if (strategy_name == "aducanumab") {
    on_treatment[1:12] <- 1 - (0.38 / 12) * (0:11)
    on_treatment[13:(n_cycles + 1)] <- on_treatment[12]
  } else if (strategy_name == "donanemab") {
    on_treatment[1:6] <- 1
    on_treatment[7:12] <- 1 - (0.55 - 0.27) / 6 * (1:6)
    on_treatment[13:(n_cycles + 1)] <- 1 - 0.55
  }
  
  qalys <- numeric(n_cycles)
  costs_healthcare <- numeric(n_cycles)
  costs_societal <- numeric(n_cycles)
  
  for (t in 1:n_cycles) {
    trace[t + 1, ] <- trace[t, ] %*% tp_matrix[, , t]
    
    # Utilities and QALYs
    qaly <- (trace[t, "mci"] * u.mci +
               trace[t, "mild"] * u.mild +
               trace[t, "moderate"] * u.moderate +
               trace[t, "severe"] * u.severe)
    
    # Apply ARIA utility decrement (months 1-3)
    if (strategy_name != "no intervention" && t <= 3) {
      p_symptomatic_aria <- if (strategy_name == "aducanumab") p.aria_symptomatic_aducanumab else p.aria_symptomatic_donanemab
      qaly <- qaly + (trace[t, "mci"] + trace[t, "mild"]) * p_symptomatic_aria * u.aria_decrement
    }
    qalys[t] <- qaly * discount_vector[t]
    
    # Costs (healthcare and societal)
    hc <- trace[t, "mci"] * cost.healthcare.mci +
      trace[t, "mild"] * cost.healthcare.mild +
      trace[t, "moderate"] * cost.healthcare.moderate +
      trace[t, "severe"] * cost.healthcare.severe
    
    sc <- hc + trace[t, "mci"] * cost.societal.mci +
      trace[t, "mild"] * cost.societal.mild +
      trace[t, "moderate"] * cost.societal.moderate +
      trace[t, "severe"] * cost.societal.severe
    
    # Drug costs (annual split monthly)
    if (strategy_name == "aducanumab" && t <= 12) {
      drug_cost <- (if (t <= 12) cost.aducanumab.year1 else cost.aducanumab.year2plus) / 12
    } else if (strategy_name == "donanemab") {
      drug_cost <- cost.donanemab / 12
    } else {
      drug_cost <- 0
    }
    
    # Infusions & monitoring (adjust by treatment and time)
    infusion_cost <- 0
    if (strategy_name == "aducanumab") {
      if (t <= 6) infusion_cost <- cost.aducanumab.m1_6
      else if (t <= 12) infusion_cost <- cost.aducanumab.m7_12
      else infusion_cost <- cost.aducanumab.m13plus
    } else if (strategy_name == "donanemab") {
      if (t <= 6) infusion_cost <- cost.donanemab.m1_6
      else if (t <= 12) infusion_cost <- cost.donanemab.m7_12
      else infusion_cost <- cost.donanemab.m13plus
    }
    
    # ARIA costs in cycles 1-3
    aria_cost <- 0
    if (strategy_name != "no intervention" && t <= 3) {
      p_symptomatic_aria <- if (strategy_name == "aducanumab") p.aria_symptomatic_aducanumab else p.aria_symptomatic_donanemab
      aria_cases <- (trace[t, "mci"] + trace[t, "mild"]) * p_symptomatic_aria
      aria_cost <- aria_cases * (cost.aria_outpatient_visit + cost.aria_followup_imaging + 0.04 * cost.aria_hospitalization)
    }
    
    # Active treatment population (excluding severe)
    active_patients <- sum(trace[t, c("mci", "mild", "moderate")])
    drug_cost_total <- drug_cost * active_patients * on_treatment[t]
    infusion_cost_total <- infusion_cost * active_patients * on_treatment[t]
    
    total_healthcare <- hc + drug_cost_total + infusion_cost_total + aria_cost
    total_societal <- sc + drug_cost_total + infusion_cost_total + aria_cost
    
    costs_healthcare[t] <- total_healthcare * discount_vector[t]
    costs_societal[t] <- total_societal * discount_vector[t]
  }
  
  # One-time screening cost (only for treatment arms)
  screening_cost <- if (strategy_name == "aducanumab") cost.screening.aducanumab else if (strategy_name == "donanemab") cost.screening.donanemab else 0
  
  list(
    qalys = sum(qalys) / cohort_size,
    healthcare_costs = (sum(costs_healthcare) + screening_cost) / cohort_size,
    societal_costs = (sum(costs_societal) + screening_cost) / cohort_size
  )
}

# Run the model for each strategy
results <- list()
results$no_intervention <- simulate_strategy("no intervention")
results$aducanumab <- simulate_strategy("aducanumab")
results$donanemab <- simulate_strategy("donanemab")
```
