## Supplementary material for "Replicating Health-Economic Simulation Models for Alzheimer’s Disease Using Artificial Intelligence": final R scripts: Ross_O4.R

```
# ===========================
# MARKOV MODEL INPUTS (ROSS ET AL., 2022)
# ===========================

# --- Descriptive Inputs ---
state_names <- c("mci", "mild", "moderate", "severe", "death")
strategy_names <- c("standard_care", "aducanumab", "donanemab")
initial_age <- 75.2
cohort_size <- 1000

# --- Initial Distribution ---
init_dist <- c(mci = 0.65, mild = 0.35, moderate = 0, severe = 0, death = 0)

# --- Transition Probabilities (Natural History) ---
p.mci_to_mild <- 0.007
p.mild_to_moderate <- 0.016
p.moderate_to_severe <- 0.026

# --- Mortality Hazard Ratios (HRs) ---
hr_mci <- 1.61
hr_mild <- 2.23
hr_moderate <- 3.10
hr_severe <- 4.98

# --- Treatment Effects (Hazard Ratios vs. placebo) ---
hr_progression_aducanumab <- 0.89
hr_progression_donanemab <- 0.68

# --- Utilities (Annual to Monthly) ---
u_mci <- 0.73 / 12
u_mild <- 0.69 / 12
u_moderate <- 0.53 / 12
u_severe <- 0.34 / 12
u_aria_decrement <- -0.065 / 12

# --- ARIA Probabilities ---
p_aria_aducanumab <- 0.41
p_aria_donanemab <- 0.39

# --- Monthly Healthcare Costs (2020 USD) ---
c_hc_mci <- 691
c_hc_mild <- 1049
c_hc_moderate <- 1274
c_hc_severe <- 3764

# --- Monthly Societal Costs (additional to healthcare) ---
c_soc_mci <- 49
c_soc_mild <- 1483
c_soc_moderate <- 1932
c_soc_severe <- 2153

# --- Annual Drug Costs ---
c_annual_aducanumab_year1 <- 20500
c_annual_aducanumab_subsequent <- 28200
c_annual_donanemab <- 28200

# --- Monthly Infusion and Monitoring Costs ---
# Aducanumab
c_aducanumab_1_6mo <- 112
c_aducanumab_7_12mo <- 87
c_aducanumab_13mo_plus <- 80

# Donanemab
c_donanemab_1_6mo <- 599
c_donanemab_7_12mo <- 302
c_donanemab_13mo_plus <- 287

# --- Screening Costs (one-time per eligible patient) ---
c_screening_aducanumab <- 6957
c_screening_donanemab <- 17096

# --- ARIA-Related Costs ---
c_aria_followup_imaging <- 1060
c_aria_hospitalization <- 35934
c_aria_outpatient_visit <- 128


# ===========================
# LOAD AND PROCESS MORTALITY DATA
# ===========================

# Load mortality rates CSV
mortality_df <- read.csv("mortalityRates_US_2017.csv")

# Compute age-specific death probabilities by severity using HRs
mortality_df$prob_death_mci <- 1 - exp(-mortality_df$monthly_rate * hr_mci)
mortality_df$prob_death_mild <- 1 - exp(-mortality_df$monthly_rate * hr_mild)
mortality_df$prob_death_moderate <- 1 - exp(-mortality_df$monthly_rate * hr_moderate)
mortality_df$prob_death_severe <- 1 - exp(-mortality_df$monthly_rate * hr_severe)

# Resulting data frame includes: age, prob_death_mci, prob_death_mild, prob_death_moderate, prob_death_severe
mortality_df <- mortality_df[, c("age", "prob_death_mci", "prob_death_mild", "prob_death_moderate", "prob_death_severe")]


# ===========================
# TIME-DEPENDENT TRANSITION PROBABILITIES
# ===========================

# Define number of cycles
n_cycles <- 480  # e.g., 40 years * 12 months

# --- Standard care (no intervention) ---
p_mci_to_mild_std <- rep(p.mci_to_mild, n_cycles)
p_mci_stay_std <- 1 - p_mci_to_mild_std

p_mild_to_moderate_std <- rep(p.mild_to_moderate, n_cycles)
p_mild_stay_std <- 1 - p_mild_to_moderate_std

p_moderate_to_severe_std <- rep(p.moderate_to_severe, n_cycles)
p_moderate_stay_std <- 1 - p_moderate_to_severe_std

p_severe_stay_std <- rep(1, n_cycles)  # No progression from severe except to death

# --- Aducanumab intervention ---
# MCI to Mild
rate_mci <- -log(1 - p.mci_to_mild)
adj_rate_mci_aducanumab <- rate_mci * hr_progression_aducanumab
p_mci_to_mild_adu <- rep(1 - exp(-adj_rate_mci_aducanumab), n_cycles)
p_mci_stay_adu <- 1 - p_mci_to_mild_adu

# Mild to Moderate
rate_mild <- -log(1 - p.mild_to_moderate)
adj_rate_mild_aducanumab <- rate_mild * hr_progression_aducanumab
p_mild_to_moderate_adu <- rep(1 - exp(-adj_rate_mild_aducanumab), n_cycles)
p_mild_stay_adu <- 1 - p_mild_to_moderate_adu

# Moderate to Severe
rate_moderate <- -log(1 - p.moderate_to_severe)
adj_rate_moderate_aducanumab <- rate_moderate * hr_progression_aducanumab
p_moderate_to_severe_adu <- rep(1 - exp(-adj_rate_moderate_aducanumab), n_cycles)
p_moderate_stay_adu <- 1 - p_moderate_to_severe_adu

# Severe
p_severe_stay_adu <- rep(1, n_cycles)

# --- Donanemab intervention ---
adj_rate_mci_donanemab <- rate_mci * hr_progression_donanemab
p_mci_to_mild_don <- rep(1 - exp(-adj_rate_mci_donanemab), n_cycles)
p_mci_stay_don <- 1 - p_mci_to_mild_don

adj_rate_mild_donanemab <- rate_mild * hr_progression_donanemab
p_mild_to_moderate_don <- rep(1 - exp(-adj_rate_mild_donanemab), n_cycles)
p_mild_stay_don <- 1 - p_mild_to_moderate_don

adj_rate_moderate_donanemab <- rate_moderate * hr_progression_donanemab
p_moderate_to_severe_don <- rep(1 - exp(-adj_rate_moderate_donanemab), n_cycles)
p_moderate_stay_don <- 1 - p_moderate_to_severe_don

p_severe_stay_don <- rep(1, n_cycles)


# ===========================
# TRANSITION MATRIX INITIALIZATION FUNCTION
# ===========================

build_transition_matrix <- function(strat) {
  # Create an empty array: states x states x cycles
  trans_mat <- array(0, dim = c(length(state_names), length(state_names), n_cycles),
                     dimnames = list(state_names, state_names, NULL))
  
  for (cycle in 1:n_cycles) {
    # Determine age for this cycle (starting age + floor(month/12))
    age_index <- min(99, max(60, floor(initial_age + (cycle - 1) / 12)))
    age_row <- mortality_df[mortality_df$age == age_index, ]
    
    # --- Assign death probabilities ---
    trans_mat["mci",     "death", cycle] <- age_row$prob_death_mci
    trans_mat["mild",    "death", cycle] <- age_row$prob_death_mild
    trans_mat["moderate","death", cycle] <- age_row$prob_death_moderate
    trans_mat["severe",  "death", cycle] <- age_row$prob_death_severe
    trans_mat["death",   "death", cycle] <- 1
    
    # === [INSERT PROMPT 4B HERE] — define transitions to next disease stages (with or without treatment) ===
    
    if (strat == "no intervention") {
      trans_mat["mci", "mild", cycle]      <- p_mci_to_mild_std[cycle] * (1 - trans_mat["mci", "death", cycle])
      trans_mat["mci", "mci", cycle]       <- p_mci_stay_std[cycle] * (1 - trans_mat["mci", "death", cycle])
      
      trans_mat["mild", "moderate", cycle] <- p_mild_to_moderate_std[cycle] * (1 - trans_mat["mild", "death", cycle])
      trans_mat["mild", "mild", cycle]     <- p_mild_stay_std[cycle] * (1 - trans_mat["mild", "death", cycle])
      
      trans_mat["moderate", "severe", cycle] <- p_moderate_to_severe_std[cycle] * (1 - trans_mat["moderate", "death", cycle])
      trans_mat["moderate", "moderate", cycle] <- p_moderate_stay_std[cycle] * (1 - trans_mat["moderate", "death", cycle])
      
      trans_mat["severe", "severe", cycle] <- p_severe_stay_std[cycle] * (1 - trans_mat["severe", "death", cycle])
      
    } else if (strat == "aducanumab") {
      trans_mat["mci", "mild", cycle]      <- p_mci_to_mild_adu[cycle] * (1 - trans_mat["mci", "death", cycle])
      trans_mat["mci", "mci", cycle]       <- p_mci_stay_adu[cycle] * (1 - trans_mat["mci", "death", cycle])
      
      trans_mat["mild", "moderate", cycle] <- p_mild_to_moderate_adu[cycle] * (1 - trans_mat["mild", "death", cycle])
      trans_mat["mild", "mild", cycle]     <- p_mild_stay_adu[cycle] * (1 - trans_mat["mild", "death", cycle])
      
      trans_mat["moderate", "severe", cycle] <- p_moderate_to_severe_adu[cycle] * (1 - trans_mat["moderate", "death", cycle])
      trans_mat["moderate", "moderate", cycle] <- p_moderate_stay_adu[cycle] * (1 - trans_mat["moderate", "death", cycle])
      
      trans_mat["severe", "severe", cycle] <- p_severe_stay_adu[cycle] * (1 - trans_mat["severe", "death", cycle])
      
    } else if (strat == "donanemab") {
      trans_mat["mci", "mild", cycle]      <- p_mci_to_mild_don[cycle] * (1 - trans_mat["mci", "death", cycle])
      trans_mat["mci", "mci", cycle]       <- p_mci_stay_don[cycle] * (1 - trans_mat["mci", "death", cycle])
      
      trans_mat["mild", "moderate", cycle] <- p_mild_to_moderate_don[cycle] * (1 - trans_mat["mild", "death", cycle])
      trans_mat["mild", "mild", cycle]     <- p_mild_stay_don[cycle] * (1 - trans_mat["mild", "death", cycle])
      
      trans_mat["moderate", "severe", cycle] <- p_moderate_to_severe_don[cycle] * (1 - trans_mat["moderate", "death", cycle])
      trans_mat["moderate", "moderate", cycle] <- p_moderate_stay_don[cycle] * (1 - trans_mat["moderate", "death", cycle])
      
      trans_mat["severe", "severe", cycle] <- p_severe_stay_don[cycle] * (1 - trans_mat["severe", "death", cycle])
    }
    
    # --- Normalize each row to sum to 1 ---
    for (state in state_names) {
      row_sum <- sum(trans_mat[state, , cycle])
      if (row_sum > 0 && row_sum != 1) {
        trans_mat[state, , cycle] <- trans_mat[state, , cycle] / row_sum
      }
    }
  }
  
  return(trans_mat)
}


# ===========================
# MARKOV SIMULATION AND OUTCOME COMPUTATION
# ===========================

# Discount rate (monthly)
discount_rate <- 0.03 / 12
discount_factors <- 1 / ((1 + discount_rate) ^ (0:(n_cycles - 1)))

# Initialize results list
results <- list()

# Run model for each strategy
for (strategy in c("no intervention", "aducanumab", "donanemab")) {
  strat_id <- gsub(" ", "_", strategy)
  trans_mat <- build_transition_matrix(strategy)
  
  # Initialize trace matrix: cycles x states
  trace <- matrix(0, nrow = n_cycles + 1, ncol = length(state_names),
                  dimnames = list(0:n_cycles, state_names))
  trace[1, ] <- cohort_size * init_dist
  
  # Initialize treatment status vectors
  treated <- rep(TRUE, cohort_size)
  pct_treated <- rep(1, n_cycles + 1)
  
  if (strategy == "aducanumab") {
    for (m in 1:12) pct_treated[m + 1] <- 1 - (0.38 / 12) * m
    pct_treated[13:(n_cycles + 1)] <- pct_treated[13]
  } else if (strategy == "donanemab") {
    pct_treated[2:7]   <- 1
    pct_treated[8:13]  <- 1 - 0.27
    pct_treated[14:(n_cycles + 1)] <- 1 - 0.55
  }
  
  # Initialize vectors for outcomes
  qalys <- numeric(n_cycles)
  cost_hc <- numeric(n_cycles)
  cost_soc <- numeric(n_cycles)
  
  # One-time screening cost in cycle 1
  if (strategy == "aducanumab") {
    cost_hc[1] <- cohort_size * c_screening_aducanumab
    cost_soc[1] <- cohort_size * c_screening_aducanumab
  } else if (strategy == "donanemab") {
    cost_hc[1] <- cohort_size * c_screening_donanemab
    cost_soc[1] <- cohort_size * c_screening_donanemab
  }
  
  # Simulate Markov process
  for (cycle in 1:n_cycles) {
    trace[cycle + 1, ] <- trace[cycle, ] %*% trans_mat[, , cycle]
    
    n_mci <- trace[cycle, "mci"]
    n_mild <- trace[cycle, "mild"]
    n_mod <- trace[cycle, "moderate"]
    n_sev <- trace[cycle, "severe"]
    
    # Utilities and QALYs
    qaly_cycle <- (n_mci * u_mci + n_mild * u_mild + n_mod * u_moderate + n_sev * u_severe)
    
    # ARIA (applied to first 3 months only)
    if (cycle <= 3 && strategy != "no intervention") {
      p_aria <- ifelse(strategy == "aducanumab", p_aria_aducanumab, p_aria_donanemab)
      symptomatic_aria <- 0.24 * p_aria * (n_mci + n_mild) * pct_treated[cycle]
      qaly_cycle <- qaly_cycle + symptomatic_aria * u_aria_decrement
    }
    
    qalys[cycle] <- qaly_cycle
    
    # Background healthcare and societal costs
    c_hc_cycle <- n_mci * c_hc_mci + n_mild * c_hc_mild + n_mod * c_hc_moderate + n_sev * c_hc_severe
    c_soc_cycle <- c_hc_cycle + (n_mci * c_soc_mci + n_mild * c_soc_mild +
                                   n_mod * c_soc_moderate + n_sev * c_soc_severe)
    
    # Treatment-specific costs
    if (strategy == "aducanumab") {
      drug_cost <- ifelse(cycle <= 12, c_annual_aducanumab_year1 / 12, c_annual_aducanumab_subsequent / 12)
      mon_cost <- ifelse(cycle <= 6, c_aducanumab_1_6mo,
                         ifelse(cycle <= 12, c_aducanumab_7_12mo, c_aducanumab_13mo_plus))
      n_treated <- (n_mci + n_mild + n_mod) * pct_treated[cycle]
    } else if (strategy == "donanemab") {
      drug_cost <- c_annual_donanemab / 12
      mon_cost <- ifelse(cycle <= 6, c_donanemab_1_6mo,
                         ifelse(cycle <= 12, c_donanemab_7_12mo, c_donanemab_13mo_plus))
      n_treated <- (n_mci + n_mild + n_mod)
    } else {
      drug_cost <- 0
      mon_cost <- 0
      n_treated <- 0
      cost_hc[cycle] <- c_hc_cycle
      cost_soc[cycle] <- c_soc_cycle
    }
    
    # Apply treatment only to non-severe states
    cost_hc[cycle] <- cost_hc[cycle] + n_treated * (drug_cost + mon_cost)
    cost_soc[cycle] <- cost_soc[cycle] + n_treated * (drug_cost + mon_cost)
    
    # ARIA-related costs (outpatient visit + follow-up MRI for all; hospitalization for 4%)
    if (cycle <= 3 && strategy != "no intervention") {
      symptomatic_aria <- 0.24 * ifelse(strategy == "aducanumab", p_aria_aducanumab, p_aria_donanemab) *
        (n_mci + n_mild) * pct_treated[cycle]
      cost_aria <- symptomatic_aria * (c_aria_outpatient_visit + c_aria_followup_imaging +
                                         0.04 * c_aria_hospitalization)
      cost_hc[cycle] <- cost_hc[cycle] + cost_aria
      cost_soc[cycle] <- cost_soc[cycle] + cost_aria
    }
  }
  
  # Discounted totals
  results[[strat_id]] <- list(
    qalys = sum(qalys * discount_factors) / cohort_size,
    healthcare_costs = sum(cost_hc * discount_factors) / cohort_size,
    societal_costs = sum(cost_soc * discount_factors) / cohort_size
  )
}
```
