## Supplementary material for "Replicating Health-Economic Simulation Models for Alzheimer’s Disease Using Artificial Intelligence": final R scripts: Ross_O5.R

```
# Input Parameters for Health-Economic Markov Model from Ross et al. (2022)

# 1. State and Strategy Names
state_names <- c("mci", "mild", "moderate", "severe", "death")
strategy_names <- c("standard_care", "aducanumab", "donanemab")

# 2. Cohort Characteristics
initial_age <- 75.2
cohort_size <- 1000
initial_state_distribution <- c(mci = 0.65, mild = 0.35, moderate = 0, severe = 0, death = 0)

# 3. Monthly Disease Progression Probabilities (Standard Care)
p.mci_to_mild <- 0.007
p.mild_to_moderate <- 0.016
p.moderate_to_severe <- 0.026

# 4. Disease Progression Hazard Ratios (HRs)
hr.aducanumab <- 0.89
hr.donanemab <- 0.68

# 5. Utilities (converted from annual to monthly)
u.mci <- 0.73 / 12
u.mild <- 0.69 / 12
u.moderate <- 0.53 / 12
u.severe <- 0.34 / 12
u.death <- 0

# 6. ARIA-related disutility (symptomatic only, applied over 3 months)
aria_utility_decrement <- -0.065 / 12

# 7. Monthly Healthcare Sector Costs
cost.healthcare.mci <- 691
cost.healthcare.mild <- 1049
cost.healthcare.moderate <- 1274
cost.healthcare.severe <- 3764
cost.healthcare.death <- 0

# 8. Monthly Additional Societal Costs
cost.societal.mci <- 49
cost.societal.mild <- 1483
cost.societal.moderate <- 1932
cost.societal.severe <- 2153
cost.societal.death <- 0

# 9. Annual Drug Costs
cost.annual.aducanumab.year1 <- 20500
cost.annual.aducanumab.year2plus <- 28200
cost.annual.donanemab <- 28200

# 10. Monthly Infusion and Monitoring Costs
# Aducanumab
cost.infusion.aducanumab.m1_6 <- 112
cost.infusion.aducanumab.m7_12 <- 87
cost.infusion.aducanumab.m13plus <- 80
# Donanemab
cost.infusion.donanemab.m1_6 <- 599
cost.infusion.donanemab.m7_12 <- 302
cost.infusion.donanemab.m13plus <- 287

# 11. Screening Costs (one-time per eligible patient)
cost.screening.aducanumab <- 6957
cost.screening.donanemab <- 17096

# 12. ARIA-related Costs
cost.aria_followup_imaging <- 1060
cost.aria_hospitalization <- 35934
cost.aria_outpatient_visit <- 128

# 13. ARIA Probability (for monitoring and AE calculation)
prob.aria.aducanumab <- 0.41
prob.aria.donanemab <- 0.39
prob.symptomatic_aria.aducanumab <- 0.24
prob.symptomatic_aria.donanemab <- 0.16

# 14. Discount rate and time horizon
discount_rate <- 0.03  # Annual
time_horizon_years <- 100 - initial_age  # simulate until age 100
cycle_length_months <- 1  # monthly cycles
total_cycles <- ceiling(time_horizon_years * 12)

# 15. Mortality Hazard Ratios (relative to age-specific mortality)
hr.mortality.mci <- 1.61
hr.mortality.mild <- 2.23
hr.mortality.moderate <- 3.10
hr.mortality.severe <- 4.98


# Load mortality rate data
mortality_data <- read.csv("mortalityRates_US_2017.csv")

# Compute age-specific death probabilities for each disease severity state
mortality_data$prob_death_mci <- 1 - exp(-mortality_data$monthly_rate * hr.mortality.mci)
mortality_data$prob_death_mild <- 1 - exp(-mortality_data$monthly_rate * hr.mortality.mild)
mortality_data$prob_death_moderate <- 1 - exp(-mortality_data$monthly_rate * hr.mortality.moderate)
mortality_data$prob_death_severe <- 1 - exp(-mortality_data$monthly_rate * hr.mortality.severe)

# Final structured mortality table for use in transition matrix
mortality_table <- mortality_data[, c("age", "prob_death_mci", "prob_death_mild", "prob_death_moderate", "prob_death_severe")]


# Number of cycles
n_cycles <- total_cycles

# --- Standard care (no intervention) ---
# Transition probabilities: Standard care
p_mci_to_mild_std <- rep(p.mci_to_mild, n_cycles)
p_mild_to_moderate_std <- rep(p.mild_to_moderate, n_cycles)
p_moderate_to_severe_std <- rep(p.moderate_to_severe, n_cycles)

p_mci_stay_std <- 1 - p_mci_to_mild_std
p_mild_stay_std <- 1 - p_mild_to_moderate_std
p_moderate_stay_std <- 1 - p_moderate_to_severe_std
p_severe_stay_std <- rep(1, n_cycles)

# --- Aducanumab strategy ---
# Apply hazard ratio to progression probabilities
rate_mci <- -log(1 - p.mci_to_mild)
rate_mild <- -log(1 - p.mild_to_moderate)
rate_moderate <- -log(1 - p.moderate_to_severe)

adj_rate_mci_adu <- rate_mci * hr.aducanumab
adj_rate_mild_adu <- rate_mild * hr.aducanumab
adj_rate_moderate_adu <- rate_moderate * hr.aducanumab

p_mci_to_mild_adu <- rep(1 - exp(-adj_rate_mci_adu), n_cycles)
p_mild_to_moderate_adu <- rep(1 - exp(-adj_rate_mild_adu), n_cycles)
p_moderate_to_severe_adu <- rep(1 - exp(-adj_rate_moderate_adu), n_cycles)

p_mci_stay_adu <- 1 - p_mci_to_mild_adu
p_mild_stay_adu <- 1 - p_mild_to_moderate_adu
p_moderate_stay_adu <- 1 - p_moderate_to_severe_adu
p_severe_stay_adu <- rep(1, n_cycles)

# --- Donanemab strategy ---
adj_rate_mci_don <- rate_mci * hr.donanemab
adj_rate_mild_don <- rate_mild * hr.donanemab
adj_rate_moderate_don <- rate_moderate * hr.donanemab

p_mci_to_mild_don <- rep(1 - exp(-adj_rate_mci_don), n_cycles)
p_mild_to_moderate_don <- rep(1 - exp(-adj_rate_mild_don), n_cycles)
p_moderate_to_severe_don <- rep(1 - exp(-adj_rate_moderate_don), n_cycles)

p_mci_stay_don <- 1 - p_mci_to_mild_don
p_mild_stay_don <- 1 - p_mild_to_moderate_don
p_moderate_stay_don <- 1 - p_moderate_to_severe_don
p_severe_stay_don <- rep(1, n_cycles)


# Function to build transition probability matrix array
build_transition_matrix <- function(strat) {
  n_states <- length(state_names)
  tpm <- array(0, dim = c(n_states, n_states, n_cycles),
               dimnames = list(state_names, state_names, NULL))
  
  for (cycle in 1:n_cycles) {
    current_age <- initial_age + (cycle - 1) / 12
    age_index <- which.min(abs(mortality_table$age - current_age))
    
    # From "mci"
    tpm["mci", "death", cycle] <- mortality_table$prob_death_mci[age_index]
    
    # From "mild"
    tpm["mild", "death", cycle] <- mortality_table$prob_death_mild[age_index]
    
    # From "moderate"
    tpm["moderate", "death", cycle] <- mortality_table$prob_death_moderate[age_index]
    
    # From "severe"
    tpm["severe", "death", cycle] <- mortality_table$prob_death_severe[age_index]
    
    # From "death" (absorbing state)
    tpm["death", "death", cycle] <- 1
    
    # From "mci"
    if (strat == "no intervention") {
      p_prog <- p_mci_to_mild_std[cycle]
      p_stay <- p_mci_stay_std[cycle]
    } else if (strat == "aducanumab") {
      p_prog <- p_mci_to_mild_adu[cycle]
      p_stay <- p_mci_stay_adu[cycle]
    } else if (strat == "donanemab") {
      p_prog <- p_mci_to_mild_don[cycle]
      p_stay <- p_mci_stay_don[cycle]
    }
    death_prob <- tpm["mci", "death", cycle]
    tpm["mci", "mild", cycle] <- p_prog * (1 - death_prob)
    tpm["mci", "mci", cycle]  <- p_stay * (1 - death_prob)
    
    # From "mild"
    if (strat == "no intervention") {
      p_prog <- p_mild_to_moderate_std[cycle]
      p_stay <- p_mild_stay_std[cycle]
    } else if (strat == "aducanumab") {
      p_prog <- p_mild_to_moderate_adu[cycle]
      p_stay <- p_mild_stay_adu[cycle]
    } else if (strat == "donanemab") {
      p_prog <- p_mild_to_moderate_don[cycle]
      p_stay <- p_mild_stay_don[cycle]
    }
    death_prob <- tpm["mild", "death", cycle]
    tpm["mild", "moderate", cycle] <- p_prog * (1 - death_prob)
    tpm["mild", "mild", cycle]     <- p_stay * (1 - death_prob)
    
    # From "moderate"
    if (strat == "no intervention") {
      p_prog <- p_moderate_to_severe_std[cycle]
      p_stay <- p_moderate_stay_std[cycle]
    } else if (strat == "aducanumab") {
      p_prog <- p_moderate_to_severe_adu[cycle]
      p_stay <- p_moderate_stay_adu[cycle]
    } else if (strat == "donanemab") {
      p_prog <- p_moderate_to_severe_don[cycle]
      p_stay <- p_moderate_stay_don[cycle]
    }
    death_prob <- tpm["moderate", "death", cycle]
    tpm["moderate", "severe", cycle] <- p_prog * (1 - death_prob)
    tpm["moderate", "moderate", cycle] <- p_stay * (1 - death_prob)
    
    # From "severe"
    if (strat == "no intervention") {
      p_stay <- p_severe_stay_std[cycle]
    } else if (strat == "aducanumab") {
      p_stay <- p_severe_stay_adu[cycle]
    } else if (strat == "donanemab") {
      p_stay <- p_severe_stay_don[cycle]
    }
    death_prob <- tpm["severe", "death", cycle]
    tpm["severe", "severe", cycle] <- p_stay * (1 - death_prob)
    
    # Normalize rows (except for death row which is already valid)
    for (state in state_names[state_names != "death"]) {
      row_sum <- sum(tpm[state, , cycle])
      if (abs(row_sum - 1) > 1e-8) {
        tpm[state, , cycle] <- tpm[state, , cycle] / row_sum
      }
    }
  }
  
  return(tpm)
}


# Monthly discount rate
discount_monthly <- (1 + discount_rate)^(1 / 12) - 1

# Function to simulate Markov model
run_markov_model <- function(strategy) {
  tpm <- build_transition_matrix(strategy)
  trace <- matrix(0, nrow = n_cycles + 1, ncol = length(state_names),
                  dimnames = list(0:n_cycles, state_names))
  trace[1, ] <- cohort_size * initial_state_distribution
  
  qalys <- numeric(n_cycles)
  cost_healthcare <- numeric(n_cycles)
  cost_societal <- numeric(n_cycles)
  
  # Initialize treatment continuation vectors
  treat_adu <- rep(1, n_cycles)
  treat_adu[1:12] <- 1 - seq(0, 0.38, length.out = 12)
  treat_adu[13:n_cycles] <- 0.62
  
  treat_don <- rep(1, n_cycles)
  treat_don[1:6] <- 1 - 0.27
  treat_don[7:12] <- 1 - 0.55
  treat_don[13:n_cycles] <- 0.45
  
  for (cycle in 1:n_cycles) {
    trace[cycle + 1, ] <- trace[cycle, ] %*% tpm[ , , cycle]
    
    # Utility per state
    utilities <- c(mci = u.mci, mild = u.mild, moderate = u.moderate, 
                   severe = u.severe, death = u.death)
    qalys[cycle] <- sum(trace[cycle, ] * utilities) * (1 / (1 + discount_monthly)^cycle)
    
    # Healthcare cost
    hc_costs <- c(mci = cost.healthcare.mci, mild = cost.healthcare.mild,
                  moderate = cost.healthcare.moderate, severe = cost.healthcare.severe, death = 0)
    sc_costs <- c(mci = cost.societal.mci, mild = cost.societal.mild,
                  moderate = cost.societal.moderate, severe = cost.societal.severe, death = 0)
    
    # Treatment eligibility (no treatment in severe state)
    treated <- trace[cycle, c("mci", "mild", "moderate")]
    
    # Drug, infusion, and monitoring costs
    if (strategy == "aducanumab") {
      treat_flag <- treat_adu[cycle]
      drug_cost <- ifelse(cycle <= 12, cost.annual.aducanumab.year1, cost.annual.aducanumab.year2plus) / 12
      inf_cost <- ifelse(cycle <= 6, cost.infusion.aducanumab.m1_6,
                         ifelse(cycle <= 12, cost.infusion.aducanumab.m7_12,
                                cost.infusion.aducanumab.m13plus))
      drug_total <- sum(treated) * treat_flag * drug_cost
      infusion_total <- sum(treated) * treat_flag * inf_cost
    } else if (strategy == "donanemab") {
      treat_flag <- treat_don[cycle]
      drug_total <- sum(treated) * treat_flag * cost.annual.donanemab / 12
      inf_cost <- ifelse(cycle <= 6, cost.infusion.donanemab.m1_6,
                         ifelse(cycle <= 12, cost.infusion.donanemab.m7_12,
                                cost.infusion.donanemab.m13plus))
      infusion_total <- sum(treated) * inf_cost
    } else {
      drug_total <- 0
      infusion_total <- 0
    }
    
    # Screening cost only once
    screen_cost <- 0
    if (cycle == 1) {
      if (strategy == "aducanumab") {
        screen_cost <- cohort_size * cost.screening.aducanumab
      } else if (strategy == "donanemab") {
        screen_cost <- cohort_size * cost.screening.donanemab
      }
    }
    
    # ARIA symptomatic event cost (applies only in first 3 months)
    aria_cost <- 0
    if (cycle <= 3) {
      if (strategy == "aducanumab") {
        symp_cases <- sum(treated) * prob.aria.aducanumab * prob.symptomatic_aria.aducanumab
      } else if (strategy == "donanemab") {
        symp_cases <- sum(treated) * prob.aria.donanemab * prob.symptomatic_aria.donanemab
      } else {
        symp_cases <- 0
      }
      outpatient <- symp_cases * cost.aria_outpatient_visit
      imaging <- symp_cases * cost.aria_followup_imaging
      hospital <- symp_cases * 0.04 * cost.aria_hospitalization
      aria_cost <- outpatient + imaging + hospital
    }
    
    # QALY decrement due to ARIA
    if (cycle <= 3 && symp_cases > 0) {
      qalys[cycle] <- qalys[cycle] + symp_cases * aria_utility_decrement * cohort_size / (1 + discount_monthly)^cycle
    }
    
    # Aggregate costs
    cost_healthcare[cycle] <- (sum(trace[cycle, ] * hc_costs) +
                                 drug_total + infusion_total +
                                 screen_cost + aria_cost) / (1 + discount_monthly)^cycle
    cost_societal[cycle] <- (sum(trace[cycle, ] * (hc_costs + sc_costs)) +
                               drug_total + infusion_total +
                               screen_cost + aria_cost) / (1 + discount_monthly)^cycle
  }
  
  return(list(
    qalys = sum(qalys) / cohort_size,
    healthcare_costs = sum(cost_healthcare) / cohort_size,
    societal_costs = sum(cost_societal) / cohort_size
  ))
}

# Run model for each strategy
results <- list(
  no_intervention = run_markov_model("no intervention"),
  aducanumab = run_markov_model("aducanumab"),
  donanemab = run_markov_model("donanemab")
)
```
