## Supplementary material for "Replicating Health-Economic Simulation Models for Alzheimer’s Disease Using Artificial Intelligence": final R scripts: Ross_O6.R

```
# Define model input parameters based on Ross et al. (2022)

# State names
states <- c("mci", "mild", "moderate", "severe", "death")

# Strategy names
strategies <- c("standard_care", "aducanumab", "donanemab")

# Cohort characteristics
n_cohort <- 1000
initial_age <- 75.2
initial_mci_prop <- 0.65
initial_mild_prop <- 0.35

# Time settings
discount_rate <- 0.03
cycle_length <- 1 / 12  # Monthly cycles
time_horizon <- "lifetime"

# Transition probabilities (monthly)
p.mci_to_mild <- 0.007
p.mild_to_moderate <- 0.016
p.moderate_to_severe <- 0.026

# Hazard ratios for disease progression
hr.aducanumab <- 0.89
hr.donanemab <- 0.68

# Mortality hazard ratios (relative to general population)
hr.mci <- 1.61
hr.mild <- 2.23
hr.moderate <- 3.10
hr.severe <- 4.98

# Utilities (annual -> monthly)
u.mci <- 0.73 / 12
u.mild <- 0.69 / 12
u.moderate <- 0.53 / 12
u.severe <- 0.34 / 12
u.symptomatic_aria_decrement <- -0.065 / 12

# Background monthly healthcare sector costs ($)
cost.healthcare.mci <- 691
cost.healthcare.mild <- 1049
cost.healthcare.moderate <- 1274
cost.healthcare.severe <- 3764

# Background monthly societal costs (incremental)
cost.societal.mci <- 49
cost.societal.mild <- 1483
cost.societal.moderate <- 1932
cost.societal.severe <- 2153

# Drug costs (annual; convert to monthly)
cost.annual.aducanumab.year1 <- 20500
cost.annual.aducanumab.later <- 28200
cost.annual.donanemab <- 28200

# Monthly drug costs
cost.monthly.aducanumab.year1 <- cost.annual.aducanumab.year1 / 12
cost.monthly.aducanumab.later <- cost.annual.aducanumab.later / 12
cost.monthly.donanemab <- cost.annual.donanemab / 12

# Infusion and monitoring costs (monthly)
cost.infusion.aducanumab.m1_6 <- 112
cost.infusion.aducanumab.m7_12 <- 87
cost.infusion.aducanumab.later <- 80
cost.infusion.donanemab.m1_6 <- 599
cost.infusion.donanemab.m7_12 <- 302
cost.infusion.donanemab.later <- 287

# Screening costs (one-time)
cost.screening.aducanumab <- 6957
cost.screening.donanemab <- 17096

# ARIA-related costs
cost.aria.imaging <- 1060
cost.aria.hospitalization <- 35934
cost.aria.outpatient <- 128

# ARIA probabilities
p.aria.aducanumab <- 0.41
p.aria.donanemab <- 0.39
p.symptomatic_aria.aducanumab <- 0.41 * 0.24
p.symptomatic_aria.donanemab <- 0.39 * 0.16


# Load required data
mortality_data <- read.csv("mortalityRates_US_2017.csv")

# Calculate age-specific probabilities of death for each disease severity state
mortality_data$prob_death_mci <- 1 - exp(-mortality_data$monthly_rate * hr.mci)
mortality_data$prob_death_mild <- 1 - exp(-mortality_data$monthly_rate * hr.mild)
mortality_data$prob_death_moderate <- 1 - exp(-mortality_data$monthly_rate * hr.moderate)
mortality_data$prob_death_severe <- 1 - exp(-mortality_data$monthly_rate * hr.severe)

# Retain only the required columns
mortality_by_age <- mortality_data[, c("age", "prob_death_mci", "prob_death_mild", 
                                       "prob_death_moderate", "prob_death_severe")]


# Define number of cycles (e.g., 30 years with monthly cycles)
n_cycles <- 30 * 12

# --- Standard Care (no intervention) ---
p.mci_to_mild_std <- rep(p.mci_to_mild, n_cycles)
p.mci_stay_std <- 1 - p.mci_to_mild_std

p.mild_to_moderate_std <- rep(p.mild_to_moderate, n_cycles)
p.mild_stay_std <- 1 - p.mild_to_moderate_std

p.moderate_to_severe_std <- rep(p.moderate_to_severe, n_cycles)
p.moderate_stay_std <- 1 - p.moderate_to_severe_std

p.severe_stay_std <- rep(1, n_cycles)  # no forward progression beyond severe (death handled separately)

# --- Aducanumab ---
rate.mci_to_mild_adu <- -log(1 - p.mci_to_mild) * hr.aducanumab
p.mci_to_mild_adu <- rep(1 - exp(-rate.mci_to_mild_adu), n_cycles)
p.mci_stay_adu <- 1 - p.mci_to_mild_adu

rate.mild_to_moderate_adu <- -log(1 - p.mild_to_moderate) * hr.aducanumab
p.mild_to_moderate_adu <- rep(1 - exp(-rate.mild_to_moderate_adu), n_cycles)
p.mild_stay_adu <- 1 - p.mild_to_moderate_adu

rate.moderate_to_severe_adu <- -log(1 - p.moderate_to_severe) * hr.aducanumab
p.moderate_to_severe_adu <- rep(1 - exp(-rate.moderate_to_severe_adu), n_cycles)
p.moderate_stay_adu <- 1 - p.moderate_to_severe_adu

p.severe_stay_adu <- rep(1, n_cycles)

# --- Donanemab ---
rate.mci_to_mild_dona <- -log(1 - p.mci_to_mild) * hr.donanemab
p.mci_to_mild_dona <- rep(1 - exp(-rate.mci_to_mild_dona), n_cycles)
p.mci_stay_dona <- 1 - p.mci_to_mild_dona

rate.mild_to_moderate_dona <- -log(1 - p.mild_to_moderate) * hr.donanemab
p.mild_to_moderate_dona <- rep(1 - exp(-rate.mild_to_moderate_dona), n_cycles)
p.mild_stay_dona <- 1 - p.mild_to_moderate_dona

rate.moderate_to_severe_dona <- -log(1 - p.moderate_to_severe) * hr.donanemab
p.moderate_to_severe_dona <- rep(1 - exp(-rate.moderate_to_severe_dona), n_cycles)
p.moderate_stay_dona <- 1 - p.moderate_to_severe_dona

p.severe_stay_dona <- rep(1, n_cycles)


# Function to build the transition probability matrix
build_transition_matrix <- function(strat) {
  # Initialize transition array: [from_state, to_state, cycle]
  tp_matrix <- array(0, dim = c(length(states), length(states), n_cycles),
                     dimnames = list(from = states, to = states, cycle = NULL))
  
  for (t in 1:n_cycles) {
    # Age for current cycle
    current_age <- initial_age + (t - 1) / 12
    if (current_age > 99) current_age <- 99  # cap age at 99
    
    # Extract age-specific death probabilities
    age_row <- mortality_by_age[mortality_by_age$age == floor(current_age), ]
    death_mci <- age_row$prob_death_mci
    death_mild <- age_row$prob_death_mild
    death_moderate <- age_row$prob_death_moderate
    death_severe <- age_row$prob_death_severe
    
    # Assign probabilities to death state
    tp_matrix["mci", "death", t] <- death_mci
    tp_matrix["mild", "death", t] <- death_mild
    tp_matrix["moderate", "death", t] <- death_moderate
    tp_matrix["severe", "death", t] <- death_severe
    tp_matrix["death", "death", t] <- 1  # absorbing state
    
    # Transitions from MCI
    if (strat == "no intervention") {
      tp_matrix["mci", "mild", t] <- p.mci_to_mild_std[t] * (1 - death_mci)
      tp_matrix["mci", "mci", t]  <- p.mci_stay_std[t] * (1 - death_mci)
    } else if (strat == "aducanumab") {
      tp_matrix["mci", "mild", t] <- p.mci_to_mild_adu[t] * (1 - death_mci)
      tp_matrix["mci", "mci", t]  <- p.mci_stay_adu[t] * (1 - death_mci)
    } else if (strat == "donanemab") {
      tp_matrix["mci", "mild", t] <- p.mci_to_mild_dona[t] * (1 - death_mci)
      tp_matrix["mci", "mci", t]  <- p.mci_stay_dona[t] * (1 - death_mci)
    }
    
    # Transitions from Mild
    if (strat == "no intervention") {
      tp_matrix["mild", "moderate", t] <- p.mild_to_moderate_std[t] * (1 - death_mild)
      tp_matrix["mild", "mild", t]     <- p.mild_stay_std[t] * (1 - death_mild)
    } else if (strat == "aducanumab") {
      tp_matrix["mild", "moderate", t] <- p.mild_to_moderate_adu[t] * (1 - death_mild)
      tp_matrix["mild", "mild", t]     <- p.mild_stay_adu[t] * (1 - death_mild)
    } else if (strat == "donanemab") {
      tp_matrix["mild", "moderate", t] <- p.mild_to_moderate_dona[t] * (1 - death_mild)
      tp_matrix["mild", "mild", t]     <- p.mild_stay_dona[t] * (1 - death_mild)
    }
    
    # Transitions from Moderate
    if (strat == "no intervention") {
      tp_matrix["moderate", "severe", t] <- p.moderate_to_severe_std[t] * (1 - death_moderate)
      tp_matrix["moderate", "moderate", t] <- p.moderate_stay_std[t] * (1 - death_moderate)
    } else if (strat == "aducanumab") {
      tp_matrix["moderate", "severe", t] <- p.moderate_to_severe_adu[t] * (1 - death_moderate)
      tp_matrix["moderate", "moderate", t] <- p.moderate_stay_adu[t] * (1 - death_moderate)
    } else if (strat == "donanemab") {
      tp_matrix["moderate", "severe", t] <- p.moderate_to_severe_dona[t] * (1 - death_moderate)
      tp_matrix["moderate", "moderate", t] <- p.moderate_stay_dona[t] * (1 - death_moderate)
    }
    
    # Transitions from Severe (only staying in severe or dying)
    tp_matrix["severe", "severe", t] <- p.severe_stay_std[t] * (1 - death_severe)
    
    # Normalize each row if sum != 1
    for (s in states) {
      row_sum <- sum(tp_matrix[s, , t])
      if (row_sum > 0 && abs(row_sum - 1) > 1e-8) {
        tp_matrix[s, , t] <- tp_matrix[s, , t] / row_sum
      }
    }
  }
  
  return(tp_matrix)
}


# Monthly discount rate
discount_monthly <- (1 + discount_rate)^(1/12) - 1

# Initial state distribution
initial_state_dist <- c("mci" = initial_mci_prop, "mild" = initial_mild_prop,
                        "moderate" = 0, "severe" = 0, "death" = 0)

# Define utility and cost vectors
utilities <- c(mci = u.mci, mild = u.mild, moderate = u.moderate, severe = u.severe, death = 0)
healthcare_costs_vec <- c(mci = cost.healthcare.mci, mild = cost.healthcare.mild,
                          moderate = cost.healthcare.moderate, severe = cost.healthcare.severe, death = 0)
societal_costs_vec <- c(mci = cost.societal.mci, mild = cost.societal.mild,
                        moderate = cost.societal.moderate, severe = cost.societal.severe, death = 0)

# Define discontinuation functions
discontinue_adu <- function(cycle) {
  if (cycle <= 12) return(1 - 0.38 * (cycle / 12))
  return(1 - 0.38)
}

discontinue_dona <- function(cycle) {
  if (cycle == 6) return(1 - 0.27)
  else if (cycle == 12) return(1 - 0.55)
  else if (cycle > 12) return(0)
  return(1)
}

# Main function to run Markov model
run_markov <- function(strategy) {
  trace <- matrix(0, nrow = n_cycles + 1, ncol = length(states), dimnames = list(NULL, states))
  trace[1, ] <- initial_state_dist
  
  tp <- build_transition_matrix(strategy)
  
  qalys <- 0
  hc_costs <- 0
  soc_costs <- 0
  
  for (t in 1:n_cycles) {
    trace[t + 1, ] <- trace[t, ] %*% tp[ , , t]
    
    # Base QALYs and costs
    cycle_util <- sum(trace[t, ] * utilities)
    cycle_hc_cost <- sum(trace[t, ] * healthcare_costs_vec)
    cycle_soc_cost <- cycle_hc_cost + sum(trace[t, ] * societal_costs_vec)
    
    # Screening (one-time)
    if (t == 1) {
      if (strategy == "aducanumab") cycle_hc_cost <- cycle_hc_cost + cost.screening.aducanumab / n_cohort
      if (strategy == "donanemab") cycle_hc_cost <- cycle_hc_cost + cost.screening.donanemab / n_cohort
    }
    
    # Treatment eligibility (not in severe)
    treated <- sum(trace[t, c("mci", "mild", "moderate")])
    
    # Treatment continuation & costs
    if (strategy == "aducanumab") {
      cont <- discontinue_adu(t)
      drug_cost <- if (t <= 12) cost.monthly.aducanumab.year1 else cost.monthly.aducanumab.later
      inf_cost <- if (t <= 6) cost.infusion.aducanumab.m1_6 else if (t <= 12) cost.infusion.aducanumab.m7_12 else cost.infusion.aducanumab.later
      cycle_hc_cost <- cycle_hc_cost + cont * treated * (drug_cost + inf_cost) / n_cohort
    }
    
    if (strategy == "donanemab") {
      cont <- discontinue_dona(t)
      inf_cost <- if (t <= 6) cost.infusion.donanemab.m1_6 else if (t <= 12) cost.infusion.donanemab.m7_12 else cost.infusion.donanemab.later
      cycle_hc_cost <- cycle_hc_cost + (cont * treated * cost.monthly.donanemab +
                                          treated * inf_cost) / n_cohort
    }
    
    # ARIA-related costs and utility decrement
    if (t <= 3 && strategy %in% c("aducanumab", "donanemab")) {
      p_aria <- if (strategy == "aducanumab") p.symptomatic_aria.aducanumab else p.symptomatic_aria.donanemab
      cont <- if (strategy == "aducanumab") discontinue_adu(t) else discontinue_dona(t)
      aria_cases <- cont * treated * p_aria
      aria_cost <- aria_cases * (cost.aria.outpatient + cost.aria.imaging + 0.04 * cost.aria.hospitalization) / n_cohort
      cycle_hc_cost <- cycle_hc_cost + aria_cost
      cycle_util <- cycle_util + (aria_cases * u.symptomatic_aria_decrement) / n_cohort
    }
    
    # Apply discounting
    discount_factor <- 1 / ((1 + discount_monthly)^t)
    qalys <- qalys + cycle_util * discount_factor
    hc_costs <- hc_costs + cycle_hc_cost * discount_factor
    soc_costs <- soc_costs + cycle_soc_cost * discount_factor
  }
  
  return(list(qalys = qalys, healthcare_costs = hc_costs, societal_costs = soc_costs))
}

# Store results for each strategy
results <- list()
results$no_intervention <- run_markov("no intervention")
results$aducanumab <- run_markov("aducanumab")
results$donanemab <- run_markov("donanemab")
```
