## Supplementary material for "Replicating Health-Economic Simulation Models for Alzheimer’s Disease Using Artificial Intelligence": final R scripts: Ross_O7.R

```
# Input parameters for health economic Markov model from Ross et al. (2022)

# Define model states
states <- c("mci", "mild", "moderate", "severe", "death")

# Define treatment strategies
strategies <- c("standard_care", "aducanumab", "donanemab")

# Cohort and demographic settings
cohort_size <- 1000
initial_age <- 75.2

# Baseline state distribution
initial_state_dist <- c(mci = 0.65, mild = 0.35, moderate = 0, severe = 0, death = 0)

# Monthly progression probabilities (natural history)
p.mci_to_mild <- 0.007
p.mild_to_moderate <- 0.016
p.moderate_to_severe <- 0.026

# Disease progression hazard ratios (treatment effects)
hr.aducanumab <- 0.89
hr.donanemab <- 0.68

# Mortality hazard ratios by state (relative to age-specific background mortality)
hr.mci <- 1.61
hr.mild <- 2.23
hr.moderate <- 3.10
hr.severe <- 4.98

# Utilities (converted to monthly values)
u.mci <- 0.73 / 12
u.mild <- 0.69 / 12
u.moderate <- 0.53 / 12
u.severe <- 0.34 / 12

# Utility decrement due to symptomatic ARIA (applied for 3 months)
utility_decrement_aria <- -0.065

# Monthly background healthcare sector costs (USD)
c.hc.mci <- 691
c.hc.mild <- 1049
c.hc.moderate <- 1274
c.hc.severe <- 3764

# Monthly background societal costs (USD)
c.soc.mci <- 49
c.soc.mild <- 1483
c.soc.moderate <- 1932
c.soc.severe <- 2153

# Annual drug costs (USD)
c.drug.annual.aducanumab_year1 <- 20500
c.drug.annual.aducanumab_later <- 28200
c.drug.annual.donanemab <- 28200

# Monthly infusion and monitoring costs (USD)
c.infusion.aducanumab.m1_6 <- 112
c.infusion.aducanumab.m7_12 <- 87
c.infusion.aducanumab.later <- 80

c.infusion.donanemab.m1_6 <- 599
c.infusion.donanemab.m7_12 <- 302
c.infusion.donanemab.later <- 287

# Screening costs per eligible patient (USD)
c.screening.aducanumab <- 6957
c.screening.donanemab <- 17096

# ARIA-related costs (USD)
c.aria.followup_imaging <- 1060
c.aria.hospitalization <- 35934
c.aria.outpatient <- 128


# Load mortality rate data
mortality_data <- read.csv("mortalityRates_US_2017.csv")

# Compute age-specific death probabilities by AD state using HRs
mortality_data$prob_death_mci <- 1 - exp(-mortality_data$monthly_rate * hr.mci)
mortality_data$prob_death_mild <- 1 - exp(-mortality_data$monthly_rate * hr.mild)
mortality_data$prob_death_moderate <- 1 - exp(-mortality_data$monthly_rate * hr.moderate)
mortality_data$prob_death_severe <- 1 - exp(-mortality_data$monthly_rate * hr.severe)

# Final data frame with required columns
mortality_probs <- mortality_data[, c("age", "prob_death_mci", "prob_death_mild", "prob_death_moderate", "prob_death_severe")]


# Number of cycles for the model (e.g., 30 years * 12 months = 360 cycles)
n_cycles <- 360

# Transition probabilities for standard care (no intervention)
p_mci_to_mild_std <- rep(p.mci_to_mild, n_cycles)
p_mild_to_mod_std <- rep(p.mild_to_moderate, n_cycles)
p_mod_to_sev_std <- rep(p.moderate_to_severe, n_cycles)

p_mci_stay_std <- 1 - p_mci_to_mild_std
p_mild_stay_std <- 1 - p_mild_to_mod_std
p_mod_stay_std <- 1 - p_mod_to_sev_std
p_sev_stay_std <- rep(1, n_cycles)  # No progression beyond severe

# Aducanumab intervention
rate_mci_to_mild <- -log(1 - p.mci_to_mild)
rate_mild_to_mod <- -log(1 - p.mild_to_moderate)
rate_mod_to_sev <- -log(1 - p.moderate_to_severe)

adj_rate_mci_to_mild_adu <- rate_mci_to_mild * hr.aducanumab
adj_rate_mild_to_mod_adu <- rate_mild_to_mod * hr.aducanumab
adj_rate_mod_to_sev_adu <- rate_mod_to_sev * hr.aducanumab

p_mci_to_mild_adu <- rep(1 - exp(-adj_rate_mci_to_mild_adu), n_cycles)
p_mild_to_mod_adu <- rep(1 - exp(-adj_rate_mild_to_mod_adu), n_cycles)
p_mod_to_sev_adu <- rep(1 - exp(-adj_rate_mod_to_sev_adu), n_cycles)

p_mci_stay_adu <- 1 - p_mci_to_mild_adu
p_mild_stay_adu <- 1 - p_mild_to_mod_adu
p_mod_stay_adu <- 1 - p_mod_to_sev_adu
p_sev_stay_adu <- rep(1, n_cycles)

# Donanemab intervention
adj_rate_mci_to_mild_don <- rate_mci_to_mild * hr.donanemab
adj_rate_mild_to_mod_don <- rate_mild_to_mod * hr.donanemab
adj_rate_mod_to_sev_don <- rate_mod_to_sev * hr.donanemab

p_mci_to_mild_don <- rep(1 - exp(-adj_rate_mci_to_mild_don), n_cycles)
p_mild_to_mod_don <- rep(1 - exp(-adj_rate_mild_to_mod_don), n_cycles)
p_mod_to_sev_don <- rep(1 - exp(-adj_rate_mod_to_sev_don), n_cycles)

p_mci_stay_don <- 1 - p_mci_to_mild_don
p_mild_stay_don <- 1 - p_mild_to_mod_don
p_mod_stay_don <- 1 - p_mod_to_sev_don
p_sev_stay_don <- rep(1, n_cycles)


# Function to build transition probability array for a given strategy
build_transition_matrix <- function(strat) {
  
  # Initialize array: dimensions are [from_state, to_state, cycle]
  tmat <- array(0, dim = c(length(states), length(states), n_cycles),
                dimnames = list(from = states, to = states, cycle = NULL))
  
  for (cycle in 1:n_cycles) {
    
    # Determine current age (starting at 75.2 and increasing 1 year every 12 cycles)
    current_age <- min(99, floor(initial_age + (cycle - 1) / 12))
    
    # Get death probabilities for each state at this age
    p_death_mci <- mortality_probs$prob_death_mci[mortality_probs$age == current_age]
    p_death_mild <- mortality_probs$prob_death_mild[mortality_probs$age == current_age]
    p_death_moderate <- mortality_probs$prob_death_moderate[mortality_probs$age == current_age]
    p_death_severe <- mortality_probs$prob_death_severe[mortality_probs$age == current_age]
    
    # Assign death transition probabilities
    tmat["mci", "death", cycle] <- p_death_mci
    tmat["mild", "death", cycle] <- p_death_mild
    tmat["moderate", "death", cycle] <- p_death_moderate
    tmat["severe", "death", cycle] <- p_death_severe
    tmat["death", "death", cycle] <- 1  # Absorbing state
    
    # Transitions from MCI
    if (strat == "no intervention") {
      tmat["mci", "mild", cycle] <- p_mci_to_mild_std[cycle] * (1 - p_death_mci)
      tmat["mci", "mci", cycle]  <- p_mci_stay_std[cycle] * (1 - p_death_mci)
    } else if (strat == "aducanumab") {
      tmat["mci", "mild", cycle] <- p_mci_to_mild_adu[cycle] * (1 - p_death_mci)
      tmat["mci", "mci", cycle]  <- p_mci_stay_adu[cycle] * (1 - p_death_mci)
    } else if (strat == "donanemab") {
      tmat["mci", "mild", cycle] <- p_mci_to_mild_don[cycle] * (1 - p_death_mci)
      tmat["mci", "mci", cycle]  <- p_mci_stay_don[cycle] * (1 - p_death_mci)
    }
    
    # Transitions from Mild
    if (strat == "no intervention") {
      tmat["mild", "moderate", cycle] <- p_mild_to_mod_std[cycle] * (1 - p_death_mild)
      tmat["mild", "mild", cycle]     <- p_mild_stay_std[cycle] * (1 - p_death_mild)
    } else if (strat == "aducanumab") {
      tmat["mild", "moderate", cycle] <- p_mild_to_mod_adu[cycle] * (1 - p_death_mild)
      tmat["mild", "mild", cycle]     <- p_mild_stay_adu[cycle] * (1 - p_death_mild)
    } else if (strat == "donanemab") {
      tmat["mild", "moderate", cycle] <- p_mild_to_mod_don[cycle] * (1 - p_death_mild)
      tmat["mild", "mild", cycle]     <- p_mild_stay_don[cycle] * (1 - p_death_mild)
    }
    
    # Transitions from Moderate
    if (strat == "no intervention") {
      tmat["moderate", "severe", cycle] <- p_mod_to_sev_std[cycle] * (1 - p_death_moderate)
      tmat["moderate", "moderate", cycle] <- p_mod_stay_std[cycle] * (1 - p_death_moderate)
    } else if (strat == "aducanumab") {
      tmat["moderate", "severe", cycle] <- p_mod_to_sev_adu[cycle] * (1 - p_death_moderate)
      tmat["moderate", "moderate", cycle] <- p_mod_stay_adu[cycle] * (1 - p_death_moderate)
    } else if (strat == "donanemab") {
      tmat["moderate", "severe", cycle] <- p_mod_to_sev_don[cycle] * (1 - p_death_moderate)
      tmat["moderate", "moderate", cycle] <- p_mod_stay_don[cycle] * (1 - p_death_moderate)
    }
    
    # Transitions from Severe (only staying, besides death)
    if (strat == "no intervention") {
      tmat["severe", "severe", cycle] <- p_sev_stay_std[cycle] * (1 - p_death_severe)
    } else if (strat == "aducanumab") {
      tmat["severe", "severe", cycle] <- p_sev_stay_adu[cycle] * (1 - p_death_severe)
    } else if (strat == "donanemab") {
      tmat["severe", "severe", cycle] <- p_sev_stay_don[cycle] * (1 - p_death_severe)
    }
    
    # Normalize rows if row sum ≠ 1
    for (s in states) {
      row_sum <- sum(tmat[s, , cycle])
      if (row_sum > 0 && abs(row_sum - 1) > 1e-8) {
        tmat[s, , cycle] <- tmat[s, , cycle] / row_sum
      }
    }
  }
  
  return(tmat)
}


# Monthly discount rate for 3% annual
disc_rate <- 0.00246627
discount_factors <- 1 / ((1 + disc_rate) ^ (0:(n_cycles - 1)))

# Initialize results storage
results <- list()

# List of strategies
strategy_list <- c("no intervention", "aducanumab", "donanemab")

# State-specific utilities and costs (vectors)
utilities <- c(mci = u.mci, mild = u.mild, moderate = u.moderate, severe = u.severe, death = 0)
hc_costs <- c(mci = c.hc.mci, mild = c.hc.mild, moderate = c.hc.moderate, severe = c.hc.severe, death = 0)
soc_costs <- c(mci = c.soc.mci, mild = c.soc.mild, moderate = c.soc.moderate, severe = c.soc.severe, death = 0)

for (strategy in strategy_list) {
  
  strat_key <- gsub(" ", "_", strategy)
  tm <- build_transition_matrix(strategy)
  
  # Initialize trace matrix
  trace <- matrix(0, nrow = n_cycles + 1, ncol = length(states), dimnames = list(NULL, states))
  trace[1, ] <- cohort_size * initial_state_dist
  
  # Treatment status trackers
  on_treatment <- rep(TRUE, cohort_size)
  prop_treated <- rep(1, n_cycles + 1)
  
  if (strategy == "aducanumab") {
    for (i in 1:12) prop_treated[i + 1] <- 1 - (0.38 * i / 12)
    prop_treated[14:(n_cycles + 1)] <- prop_treated[13]
  } else if (strategy == "donanemab") {
    prop_treated[2:7] <- 1 - 0.27
    prop_treated[8:13] <- 1 - 0.55
    prop_treated[14:(n_cycles + 1)] <- 1 - 0.66
  }
  
  # Outcomes
  qalys <- numeric(n_cycles)
  hc_total <- numeric(n_cycles)
  soc_total <- numeric(n_cycles)
  
  for (cycle in 1:n_cycles) {
    
    trace[cycle + 1, ] <- trace[cycle, ] %*% tm[, , cycle]
    
    # QALYs with ARIA decrement in cycles 1-3
    qaly_cycle <- sum(trace[cycle, states] * utilities[states])
    if (strategy != "no intervention" && cycle <= 3) {
      aria_rate <- ifelse(strategy == "aducanumab", 0.41 * 0.24, 0.39 * 0.16)
      qaly_cycle <- qaly_cycle + (cohort_size * aria_rate * utility_decrement_aria)
    }
    qalys[cycle] <- qaly_cycle * discount_factors[cycle]
    
    # Background healthcare and societal costs
    hc_cycle <- sum(trace[cycle, states] * hc_costs[states])
    soc_cycle <- sum(trace[cycle, states] * (hc_costs[states] + soc_costs[states]))
    
    # Drug cost (annual → monthly); only treated & not in severe
    if (strategy == "aducanumab") {
      drug_cost <- ifelse(cycle <= 12, c.drug.annual.aducanumab_year1 / 12, c.drug.annual.aducanumab_later / 12)
    } else if (strategy == "donanemab") {
      drug_cost <- c.drug.annual.donanemab / 12
    } else {
      drug_cost <- 0
    }
    
    if (strategy != "no intervention") {
      treatable_states <- setdiff(states, c("severe", "death"))
      num_treated <- sum(trace[cycle, treatable_states]) * prop_treated[cycle]
      hc_cycle <- hc_cycle + num_treated * drug_cost
      
      # Infusion and monitoring costs
      if (strategy == "aducanumab") {
        if (cycle <= 6) mon_cost <- c.infusion.aducanumab.m1_6
        else if (cycle <= 12) mon_cost <- c.infusion.aducanumab.m7_12
        else mon_cost <- c.infusion.aducanumab.later
        hc_cycle <- hc_cycle + num_treated * mon_cost
      } else if (strategy == "donanemab") {
        if (cycle <= 6) mon_cost <- c.infusion.donanemab.m1_6
        else if (cycle <= 12) mon_cost <- c.infusion.donanemab.m7_12
        else mon_cost <- c.infusion.donanemab.later
        hc_cycle <- hc_cycle + num_treated * mon_cost
      }
      
      # ARIA costs in cycles 1–3
      if (cycle <= 3) {
        aria_prob <- ifelse(strategy == "aducanumab", 0.41 * 0.24, 0.39 * 0.16)
        n_aria <- cohort_size * aria_prob
        n_hosp <- n_aria * 0.04
        hc_cycle <- hc_cycle + n_aria * (c.aria.outpatient + c.aria.followup_imaging) + n_hosp * c.aria.hospitalization
      }
    }
    
    hc_total[cycle] <- hc_cycle * discount_factors[cycle]
    soc_total[cycle] <- soc_cycle * discount_factors[cycle]
  }
  
  # One-time screening costs
  screening_cost <- switch(strategy,
                           "no intervention" = 0,
                           "aducanumab" = c.screening.aducanumab,
                           "donanemab" = c.screening.donanemab)
  hc_total[1] <- hc_total[1] + screening_cost
  soc_total[1] <- soc_total[1] + screening_cost
  
  # Store per-person discounted totals
  results[[strat_key]] <- list(
    qalys = sum(qalys) / cohort_size,
    healthcare_costs = sum(hc_total) / cohort_size,
    societal_costs = sum(soc_total) / cohort_size
  )
}
```
