## Supplementary material for "Replicating Health-Economic Simulation Models for Alzheimer’s Disease Using Artificial Intelligence": final R scripts: Ross_O8.R

```
# Input Parameters for Health Economic Markov Model Based on Ross et al. (2022)

# Descriptive variables
states <- c("mci", "mild", "moderate", "severe", "death")
strategies <- c("standard_care", "aducanumab", "donanemab")
n.individuals <- 1000
starting.age <- 75.2

# Initial state distribution
initial_distribution <- c(mci = 0.65, mild = 0.35, moderate = 0, severe = 0, death = 0)

# Monthly disease progression probabilities (from untreated baseline)
p.mci_to_mild <- 0.007
p.mild_to_moderate <- 0.016
p.moderate_to_severe <- 0.026

# Treatment hazard ratios (progression)
hr.aducanumab <- 0.89
hr.donanemab <- 0.68

# Mortality hazard ratios by state (applied to age-specific mortality)
hr.mci <- 1.61
hr.mild <- 2.23
hr.moderate <- 3.10
hr.severe <- 4.98

# Monthly utility values (converted from annual values / 12)
u.mci <- 0.73 / 12
u.mild <- 0.69 / 12
u.moderate <- 0.53 / 12
u.severe <- 0.34 / 12

# Monthly background healthcare costs (USD)
c.healthcare.mci <- 691
c.healthcare.mild <- 1049
c.healthcare.moderate <- 1274
c.healthcare.severe <- 3764

# Monthly additional societal costs (USD)
c.societal.mci <- 49
c.societal.mild <- 1483
c.societal.moderate <- 1932
c.societal.severe <- 2153

# Annual drug costs (USD)
c.annual_drug.aducanumab.year1 <- 20500
c.annual_drug.aducanumab.later <- 28200
c.annual_drug.donanemab <- 28200

# Monthly infusion and monitoring costs (USD)
c.infusion_monitor.aducanumab.m1_6 <- 112
c.infusion_monitor.aducanumab.m7_12 <- 87
c.infusion_monitor.aducanumab.m13plus <- 80
c.infusion_monitor.donanemab.m1_6 <- 599
c.infusion_monitor.donanemab.m7_12 <- 302
c.infusion_monitor.donanemab.m13plus <- 287

# One-time screening costs (USD)
c.screening.aducanumab <- 6957
c.screening.donanemab <- 17096

# ARIA-related costs (applied per patient with ARIA)
c.aria_followup_imaging <- 1060
c.aria_hospitalization <- 35934
c.aria_outpatient_visits <- 128
u.aria_decrement <- -0.065  # for 3 months duration
p.aria.aducanumab <- 0.41
p.aria.donanemab <- 0.39

# Discount rate
discount.rate <- 0.03


# Load mortality data
mortality_data <- read.csv("mortalityRates_US_2017.csv")

# Compute age-specific death probabilities by AD severity state
mortality_data$prob_death_mci <- 1 - exp(-mortality_data$monthly_rate * hr.mci)
mortality_data$prob_death_mild <- 1 - exp(-mortality_data$monthly_rate * hr.mild)
mortality_data$prob_death_moderate <- 1 - exp(-mortality_data$monthly_rate * hr.moderate)
mortality_data$prob_death_severe <- 1 - exp(-mortality_data$monthly_rate * hr.severe)

# Resulting data frame with required columns
mortality_by_age <- mortality_data[, c("age", "prob_death_mci", "prob_death_mild", "prob_death_moderate", "prob_death_severe")]


# Number of model cycles (e.g., 12 cycles per year for lifetime simulation up to age 100)
n_cycles <- (100 - starting.age) * 12

# Transition probabilities for standard care
p.mci_to_mild_std <- rep(p.mci_to_mild, n_cycles)
p.mild_to_moderate_std <- rep(p.mild_to_moderate, n_cycles)
p.moderate_to_severe_std <- rep(p.moderate_to_severe, n_cycles)

p.mci_stay_std <- 1 - p.mci_to_mild_std
p.mild_stay_std <- 1 - p.mild_to_moderate_std
p.moderate_stay_std <- 1 - p.moderate_to_severe_std
p.severe_stay_std <- rep(1, n_cycles)  # no further progression from severe (death handled separately)

# Aducanumab: adjust progression probabilities
rate.mci_to_mild_adu <- -log(1 - p.mci_to_mild) * hr.aducanumab
rate.mild_to_moderate_adu <- -log(1 - p.mild_to_moderate) * hr.aducanumab
rate.moderate_to_severe_adu <- -log(1 - p.moderate_to_severe) * hr.aducanumab

p.mci_to_mild_adu <- rep(1 - exp(-rate.mci_to_mild_adu), n_cycles)
p.mild_to_moderate_adu <- rep(1 - exp(-rate.mild_to_moderate_adu), n_cycles)
p.moderate_to_severe_adu <- rep(1 - exp(-rate.moderate_to_severe_adu), n_cycles)

p.mci_stay_adu <- 1 - p.mci_to_mild_adu
p.mild_stay_adu <- 1 - p.mild_to_moderate_adu
p.moderate_stay_adu <- 1 - p.moderate_to_severe_adu
p.severe_stay_adu <- rep(1, n_cycles)

# Donanemab: adjust progression probabilities
rate.mci_to_mild_don <- -log(1 - p.mci_to_mild) * hr.donanemab
rate.mild_to_moderate_don <- -log(1 - p.mild_to_moderate) * hr.donanemab
rate.moderate_to_severe_don <- -log(1 - p.moderate_to_severe) * hr.donanemab

p.mci_to_mild_don <- rep(1 - exp(-rate.mci_to_mild_don), n_cycles)
p.mild_to_moderate_don <- rep(1 - exp(-rate.mild_to_moderate_don), n_cycles)
p.moderate_to_severe_don <- rep(1 - exp(-rate.moderate_to_severe_don), n_cycles)

p.mci_stay_don <- 1 - p.mci_to_mild_don
p.mild_stay_don <- 1 - p.mild_to_moderate_don
p.moderate_stay_don <- 1 - p.moderate_to_severe_don
p.severe_stay_don <- rep(1, n_cycles)


build_transition_matrix <- function(strat) {
  # Initialize 3D array: [from_state, to_state, cycle]
  tpm <- array(0, dim = c(length(states), length(states), n_cycles),
               dimnames = list(from = states, to = states, cycle = NULL))
  
  for (cycle in 1:n_cycles) {
    current_age <- floor(starting.age + (cycle - 1) / 12)
    
    # Skip if age is outside 60–99 range
    if (current_age < 60 || current_age > 99) next
    
    # Age-specific death probabilities
    age_row <- mortality_by_age[mortality_by_age$age == current_age, ]
    
    # From MCI
    tpm["mci", "death", cycle] <- age_row$prob_death_mci
    
    # From Mild
    tpm["mild", "death", cycle] <- age_row$prob_death_mild
    
    # From Moderate
    tpm["moderate", "death", cycle] <- age_row$prob_death_moderate
    
    # From Severe
    tpm["severe", "death", cycle] <- age_row$prob_death_severe
    
    # Death is an absorbing state
    tpm["death", "death", cycle] <- 1
    
    # MCI transitions
    if (strat == "no intervention") {
      tpm["mci", "mild", cycle] <- p.mci_to_mild_std[cycle] * (1 - tpm["mci", "death", cycle])
      tpm["mci", "mci", cycle]  <- p.mci_stay_std[cycle] * (1 - tpm["mci", "death", cycle])
    } else if (strat == "aducanumab") {
      tpm["mci", "mild", cycle] <- p.mci_to_mild_adu[cycle] * (1 - tpm["mci", "death", cycle])
      tpm["mci", "mci", cycle]  <- p.mci_stay_adu[cycle] * (1 - tpm["mci", "death", cycle])
    } else if (strat == "donanemab") {
      tpm["mci", "mild", cycle] <- p.mci_to_mild_don[cycle] * (1 - tpm["mci", "death", cycle])
      tpm["mci", "mci", cycle]  <- p.mci_stay_don[cycle] * (1 - tpm["mci", "death", cycle])
    }
    
    # Mild transitions
    if (strat == "no intervention") {
      tpm["mild", "moderate", cycle] <- p.mild_to_moderate_std[cycle] * (1 - tpm["mild", "death", cycle])
      tpm["mild", "mild", cycle]     <- p.mild_stay_std[cycle] * (1 - tpm["mild", "death", cycle])
    } else if (strat == "aducanumab") {
      tpm["mild", "moderate", cycle] <- p.mild_to_moderate_adu[cycle] * (1 - tpm["mild", "death", cycle])
      tpm["mild", "mild", cycle]     <- p.mild_stay_adu[cycle] * (1 - tpm["mild", "death", cycle])
    } else if (strat == "donanemab") {
      tpm["mild", "moderate", cycle] <- p.mild_to_moderate_don[cycle] * (1 - tpm["mild", "death", cycle])
      tpm["mild", "mild", cycle]     <- p.mild_stay_don[cycle] * (1 - tpm["mild", "death", cycle])
    }
    
    # Moderate transitions
    if (strat == "no intervention") {
      tpm["moderate", "severe", cycle] <- p.moderate_to_severe_std[cycle] * (1 - tpm["moderate", "death", cycle])
      tpm["moderate", "moderate", cycle] <- p.moderate_stay_std[cycle] * (1 - tpm["moderate", "death", cycle])
    } else if (strat == "aducanumab") {
      tpm["moderate", "severe", cycle] <- p.moderate_to_severe_adu[cycle] * (1 - tpm["moderate", "death", cycle])
      tpm["moderate", "moderate", cycle] <- p.moderate_stay_adu[cycle] * (1 - tpm["moderate", "death", cycle])
    } else if (strat == "donanemab") {
      tpm["moderate", "severe", cycle] <- p.moderate_to_severe_don[cycle] * (1 - tpm["moderate", "death", cycle])
      tpm["moderate", "moderate", cycle] <- p.moderate_stay_don[cycle] * (1 - tpm["moderate", "death", cycle])
    }
    
    # Severe transitions (only staying)
    if (strat == "no intervention") {
      tpm["severe", "severe", cycle] <- p.severe_stay_std[cycle] * (1 - tpm["severe", "death", cycle])
    } else if (strat == "aducanumab") {
      tpm["severe", "severe", cycle] <- p.severe_stay_adu[cycle] * (1 - tpm["severe", "death", cycle])
    } else if (strat == "donanemab") {
      tpm["severe", "severe", cycle] <- p.severe_stay_don[cycle] * (1 - tpm["severe", "death", cycle])
    }
    
    # Normalize rows (except death state)
    for (state in states[states != "death"]) {
      row_sum <- sum(tpm[state, , cycle])
      if (abs(row_sum - 1) > 1e-8 && row_sum > 0) {
        tpm[state, , cycle] <- tpm[state, , cycle] / row_sum
      }
    }
  }
  
  return(tpm)
}


# Monthly discount rate
discount_rate_monthly <- (1 + discount.rate)^(1 / 12) - 1

# Initialize results container
results <- list()

# Markov model function
run_markov_model <- function(strategy, tpm, utility_vec, hc_costs, soc_costs, screening_cost, annual_drug_cost, infusion_costs, aria_prob) {
  trace <- matrix(0, nrow = n_cycles + 1, ncol = length(states), dimnames = list(NULL, states))
  trace[1, ] <- initial_distribution * n.individuals
  
  qalys <- numeric(n_cycles)
  hc_total <- numeric(n_cycles)
  soc_total <- numeric(n_cycles)
  
  on_treatment <- rep(TRUE, n.individuals)
  discontinuation_vec <- rep(1, n_cycles)
  
  if (strategy == "aducanumab") {
    discontinuation_vec[1:12] <- 1 - seq(0, 0.38, length.out = 12)
    discontinuation_vec[13:n_cycles] <- 1 - 0.38
  } else if (strategy == "donanemab") {
    discontinuation_vec[1:6] <- 1 - 0.27
    discontinuation_vec[7:12] <- 1 - 0.55
    discontinuation_vec[13:n_cycles] <- 1 - 0.55
  }
  
  for (cycle in 1:n_cycles) {
    trace[cycle + 1, ] <- trace[cycle, ] %*% tpm[, , cycle]
    
    # Apply utility per state
    qaly_this_cycle <- sum(trace[cycle, states[1:4]] * unlist(utility_vec[states[1:4]]))
    
    # Discounted QALYs
    qalys[cycle] <- qaly_this_cycle / ((1 + discount_rate_monthly) ^ (cycle - 1))
    
    # Background healthcare and societal costs
    hc_cost <- sum(trace[cycle, states[1:4]] * unlist(hc_costs[states[1:4]]))
    soc_cost <- sum(trace[cycle, states[1:4]] * (unlist(hc_costs[states[1:4]]) + unlist(soc_costs[states[1:4]])))
    
    # Screening cost (only once)
    if (cycle == 1 && strategy != "no_intervention") {
      hc_cost <- hc_cost + screening_cost
      soc_cost <- soc_cost + screening_cost
    }
    
    # Annual drug costs and infusions
    if (strategy != "no_intervention") {
      alive <- sum(trace[cycle, states[1:4]])
      treated <- alive * discontinuation_vec[cycle]
      
      if (cycle <= 12) {
        drug_cost <- annual_drug_cost / 12
      } else {
        drug_cost <- annual_drug_cost / 12
      }
      
      inf_cost <- if (strategy == "aducanumab") {
        if (cycle <= 6) infusion_costs[1] else if (cycle <= 12) infusion_costs[2] else infusion_costs[3]
      } else {
        if (cycle <= 6) infusion_costs[1] else if (cycle <= 12) infusion_costs[2] else infusion_costs[3]
      }
      
      hc_cost <- hc_cost + treated * (drug_cost + inf_cost)
      soc_cost <- soc_cost + treated * (drug_cost + inf_cost)
    }
    
    # ARIA-related costs (only cycles 1-3)
    if (strategy != "no_intervention" && cycle <= 3) {
      symptomatic_aria <- sum(trace[cycle, states[1:2]]) * aria_prob * 0.24
      aria_cost <- symptomatic_aria * (c.aria_outpatient_visits + c.aria_followup_imaging + 0.04 * c.aria_hospitalization)
      
      hc_cost <- hc_cost + aria_cost
      soc_cost <- soc_cost + aria_cost
      
      # Apply utility decrement
      qaly_this_cycle <- qaly_this_cycle - symptomatic_aria * abs(u.aria_decrement)
      qalys[cycle] <- qaly_this_cycle / ((1 + discount_rate_monthly) ^ (cycle - 1))
    }
    
    hc_total[cycle] <- hc_cost / ((1 + discount_rate_monthly) ^ (cycle - 1))
    soc_total[cycle] <- soc_cost / ((1 + discount_rate_monthly) ^ (cycle - 1))
  }
  
  return(list(
    qalys = sum(qalys) / n.individuals,
    healthcare_costs = sum(hc_total) / n.individuals,
    societal_costs = sum(soc_total) / n.individuals
  ))
}

# Strategy: No intervention
tpm_no_int <- build_transition_matrix("no intervention")
results$no_intervention <- run_markov_model(
  strategy = "no_intervention",
  tpm = tpm_no_int,
  utility_vec = list(mci = u.mci, mild = u.mild, moderate = u.moderate, severe = u.severe),
  hc_costs = list(mci = c.healthcare.mci, mild = c.healthcare.mild, moderate = c.healthcare.moderate, severe = c.healthcare.severe),
  soc_costs = list(mci = c.societal.mci, mild = c.societal.mild, moderate = c.societal.moderate, severe = c.societal.severe),
  screening_cost = 0,
  annual_drug_cost = 0,
  infusion_costs = c(0, 0, 0),
  aria_prob = 0
)

# Strategy: Aducanumab
tpm_adu <- build_transition_matrix("aducanumab")
results$aducanumab <- run_markov_model(
  strategy = "aducanumab",
  tpm = tpm_adu,
  utility_vec = list(mci = u.mci, mild = u.mild, moderate = u.moderate, severe = u.severe),
  hc_costs = list(mci = c.healthcare.mci, mild = c.healthcare.mild, moderate = c.healthcare.moderate, severe = c.healthcare.severe),
  soc_costs = list(mci = c.societal.mci, mild = c.societal.mild, moderate = c.societal.moderate, severe = c.societal.severe),
  screening_cost = c.screening.aducanumab,
  annual_drug_cost = c.annual_drug.aducanumab.later,
  infusion_costs = c(c.infusion_monitor.aducanumab.m1_6, c.infusion_monitor.aducanumab.m7_12, c.infusion_monitor.aducanumab.m13plus),
  aria_prob = p.aria.aducanumab
)

# Strategy: Donanemab
tpm_don <- build_transition_matrix("donanemab")
results$donanemab <- run_markov_model(
  strategy = "donanemab",
  tpm = tpm_don,
  utility_vec = list(mci = u.mci, mild = u.mild, moderate = u.moderate, severe = u.severe),
  hc_costs = list(mci = c.healthcare.mci, mild = c.healthcare.mild, moderate = c.healthcare.moderate, severe = c.healthcare.severe),
  soc_costs = list(mci = c.societal.mci, mild = c.societal.mild, moderate = c.societal.moderate, severe = c.societal.severe),
  screening_cost = c.screening.donanemab,
  annual_drug_cost = c.annual_drug.donanemab,
  infusion_costs = c(c.infusion_monitor.donanemab.m1_6, c.infusion_monitor.donanemab.m7_12, c.infusion_monitor.donanemab.m13plus),
  aria_prob = p.aria.donanemab
)
```
