## Supplementary material for "Replicating Health-Economic Simulation Models for Alzheimer’s Disease Using Artificial Intelligence": final R scripts: Ross_O9.R

```
# Prompt 1: Input Parameters for Health Economic Markov Model Based on Ross et al. (2022)

# 1. Define State and Strategy Names
states <- c("mci", "mild", "moderate", "severe", "death")
strategies <- c("standard_care", "aducanumab", "donanemab")

# 2. Cohort Characteristics
n_individuals <- 1000
initial_age <- 75.2
initial_state_dist <- c(mci = 0.65, mild = 0.35, moderate = 0, severe = 0, death = 0)

# 3. Transition Probabilities (monthly)
p.mci_to_mild <- 0.007
p.mild_to_moderate <- 0.016
p.moderate_to_severe <- 0.026

# 4. Treatment Effects (HR on progression)
hr.aducanumab <- 0.89
hr.donanemab <- 0.68

# 5. Mortality Hazard Ratios by State
hr_mortality <- list(
  mci = 1.61,
  mild = 2.23,
  moderate = 3.10,
  severe = 4.98
)

# 6. Utility Weights (converted to monthly)
u.mci <- 0.73 / 12
u.mild <- 0.69 / 12
u.moderate <- 0.53 / 12
u.severe <- 0.34 / 12
u.aria_decrement <- -0.065 / 12

# 7. Costs (all in 2020 USD, monthly unless stated otherwise)
# Background healthcare costs (monthly)
c.healthcare <- list(
  mci = 691,
  mild = 1049,
  moderate = 1274,
  severe = 3764
)

# Additional societal costs (monthly)
c.societal <- list(
  mci = 49,
  mild = 1483,
  moderate = 1932,
  severe = 2153
)

# Annual drug costs
c.aducanumab_year1 <- 20500
c.aducanumab_year2plus <- 28200
c.donanemab <- 28200

# Monthly infusion and monitoring costs
c.infusion_monitoring <- list(
  aducanumab = c(m1_6 = 112, m7_12 = 87, m13plus = 80),
  donanemab = c(m1_6 = 599, m7_12 = 302, m13plus = 287)
)

# Screening costs (per eligible patient)
c.screening <- list(
  aducanumab = 6957,
  donanemab = 17096
)

# ARIA-related costs
c.aria <- list(
  outpatient_visit = 128,
  followup_imaging = 1060,
  hospitalization = 35934
)

# ARIA probabilities
p.aria <- list(
  aducanumab = 0.41,
  donanemab = 0.39
)
p.symptomatic_aria <- list(
  aducanumab = 0.24,
  donanemab = 0.16
)


# Prompt 2: Load Mortality Data and Compute State-Specific Death Probabilities

# Load mortality rates from CSV
mortality_df <- read.csv("mortalityRates_US_2017.csv")

# Compute death probabilities by disease severity using hazard ratios
mortality_df$prob_death_mci <- 1 - exp(-mortality_df$monthly_rate * hr_mortality$mci)
mortality_df$prob_death_mild <- 1 - exp(-mortality_df$monthly_rate * hr_mortality$mild)
mortality_df$prob_death_moderate <- 1 - exp(-mortality_df$monthly_rate * hr_mortality$moderate)
mortality_df$prob_death_severe <- 1 - exp(-mortality_df$monthly_rate * hr_mortality$severe)


# Prompt 3: Define Time-Dependent Transition Probability Vectors

# Define number of cycles (e.g., 40 years * 12 months = 480 months)
n_cycles <- 480

# --- Standard Care (no intervention) ---
# MCI
p_mci_to_mild_std <- rep(p.mci_to_mild, n_cycles)
p_mci_stay_std <- 1 - p_mci_to_mild_std

# Mild
p_mild_to_moderate_std <- rep(p.mild_to_moderate, n_cycles)
p_mild_stay_std <- 1 - p_mild_to_moderate_std

# Moderate
p_moderate_to_severe_std <- rep(p.moderate_to_severe, n_cycles)
p_moderate_stay_std <- 1 - p_moderate_to_severe_std

# Severe
p_severe_stay_std <- rep(1, n_cycles)  # no further progression from severe except death (handled separately)

# --- Aducanumab Intervention ---
# Apply HR to progression probabilities
# MCI
rate_mci <- -log(1 - p.mci_to_mild)
adj_rate_mci_adu <- rate_mci * hr.aducanumab
p_mci_to_mild_adu <- rep(1 - exp(-adj_rate_mci_adu), n_cycles)
p_mci_stay_adu <- 1 - p_mci_to_mild_adu

# Mild
rate_mild <- -log(1 - p.mild_to_moderate)
adj_rate_mild_adu <- rate_mild * hr.aducanumab
p_mild_to_moderate_adu <- rep(1 - exp(-adj_rate_mild_adu), n_cycles)
p_mild_stay_adu <- 1 - p_mild_to_moderate_adu

# Moderate
rate_moderate <- -log(1 - p.moderate_to_severe)
adj_rate_moderate_adu <- rate_moderate * hr.aducanumab
p_moderate_to_severe_adu <- rep(1 - exp(-adj_rate_moderate_adu), n_cycles)
p_moderate_stay_adu <- 1 - p_moderate_to_severe_adu

# Severe
p_severe_stay_adu <- rep(1, n_cycles)

# --- Donanemab Intervention ---
# MCI
adj_rate_mci_don <- rate_mci * hr.donanemab
p_mci_to_mild_don <- rep(1 - exp(-adj_rate_mci_don), n_cycles)
p_mci_stay_don <- 1 - p_mci_to_mild_don

# Mild
adj_rate_mild_don <- rate_mild * hr.donanemab
p_mild_to_moderate_don <- rep(1 - exp(-adj_rate_mild_don), n_cycles)
p_mild_stay_don <- 1 - p_mild_to_moderate_don

# Moderate
adj_rate_moderate_don <- rate_moderate * hr.donanemab
p_moderate_to_severe_don <- rep(1 - exp(-adj_rate_moderate_don), n_cycles)
p_moderate_stay_don <- 1 - p_moderate_to_severe_don

# Severe
p_severe_stay_don <- rep(1, n_cycles)


# Prompt 4A: Initialize Transition Probability Matrix Structure

build_transition_matrix <- function(strat) {
  n_states <- length(states)
  trans_mat <- array(0, dim = c(n_states, n_states, n_cycles),
                     dimnames = list(from = states, to = states, cycle = 1:n_cycles))
  
  for (t in 1:n_cycles) {
    # Get current age (assuming one year = 12 cycles, starting at 75.2)
    current_age <- floor(initial_age + (t - 1) / 12)
    if (current_age < 60) current_age <- 60
    if (current_age > 99) current_age <- 99
    
    # Extract mortality probabilities for current age
    age_row <- mortality_df[mortality_df$age == current_age, ]
    p_death_mci <- age_row$prob_death_mci
    p_death_mild <- age_row$prob_death_mild
    p_death_moderate <- age_row$prob_death_moderate
    p_death_severe <- age_row$prob_death_severe
    
    # Assign death transition probabilities
    trans_mat["mci", "death", t] <- p_death_mci
    trans_mat["mild", "death", t] <- p_death_mild
    trans_mat["moderate", "death", t] <- p_death_moderate
    trans_mat["severe", "death", t] <- p_death_severe
    trans_mat["death", "death", t] <- 1
    
    # [INSERT PROMPT 4B HERE] — define transitions to next disease stages (with or without treatment)
    
    if (strat == "no intervention") {
      trans_mat["mci", "mild", t]      <- p_mci_to_mild_std[t] * (1 - p_death_mci)
      trans_mat["mci", "mci", t]       <- p_mci_stay_std[t] * (1 - p_death_mci)
      
      trans_mat["mild", "moderate", t] <- p_mild_to_moderate_std[t] * (1 - p_death_mild)
      trans_mat["mild", "mild", t]     <- p_mild_stay_std[t] * (1 - p_death_mild)
      
      trans_mat["moderate", "severe", t] <- p_moderate_to_severe_std[t] * (1 - p_death_moderate)
      trans_mat["moderate", "moderate", t] <- p_moderate_stay_std[t] * (1 - p_death_moderate)
      
      trans_mat["severe", "severe", t] <- p_severe_stay_std[t] * (1 - p_death_severe)
      
    } else if (strat == "aducanumab") {
      trans_mat["mci", "mild", t]      <- p_mci_to_mild_adu[t] * (1 - p_death_mci)
      trans_mat["mci", "mci", t]       <- p_mci_stay_adu[t] * (1 - p_death_mci)
      
      trans_mat["mild", "moderate", t] <- p_mild_to_moderate_adu[t] * (1 - p_death_mild)
      trans_mat["mild", "mild", t]     <- p_mild_stay_adu[t] * (1 - p_death_mild)
      
      trans_mat["moderate", "severe", t] <- p_moderate_to_severe_adu[t] * (1 - p_death_moderate)
      trans_mat["moderate", "moderate", t] <- p_moderate_stay_adu[t] * (1 - p_death_moderate)
      
      trans_mat["severe", "severe", t] <- p_severe_stay_adu[t] * (1 - p_death_severe)
      
    } else if (strat == "donanemab") {
      trans_mat["mci", "mild", t]      <- p_mci_to_mild_don[t] * (1 - p_death_mci)
      trans_mat["mci", "mci", t]       <- p_mci_stay_don[t] * (1 - p_death_mci)
      
      trans_mat["mild", "moderate", t] <- p_mild_to_moderate_don[t] * (1 - p_death_mild)
      trans_mat["mild", "mild", t]     <- p_mild_stay_don[t] * (1 - p_death_mild)
      
      trans_mat["moderate", "severe", t] <- p_moderate_to_severe_don[t] * (1 - p_death_moderate)
      trans_mat["moderate", "moderate", t] <- p_moderate_stay_don[t] * (1 - p_death_moderate)
      
      trans_mat["severe", "severe", t] <- p_severe_stay_don[t] * (1 - p_death_severe)
    }
    
    # Normalize each row to ensure row sums equal 1
    for (s in states[states != "death"]) {
      row_sum <- sum(trans_mat[s, , t])
      if (row_sum > 0 && abs(row_sum - 1) > 1e-8) {
        trans_mat[s, , t] <- trans_mat[s, , t] / row_sum
      }
    }
  }
  
  return(trans_mat)
}


# Prompt 5: Run the Markov Model and Compute Discounted Outcomes

# Set monthly discount rate
discount_rate <- 0.0024662697723036864

# Initialize results list
results <- list()

# Utility vector
utilities <- c(mci = u.mci, mild = u.mild, moderate = u.moderate, severe = u.severe, death = 0)

# Cost vectors
cost_healthcare <- c(mci = c.healthcare$mci, mild = c.healthcare$mild, moderate = c.healthcare$moderate, severe = c.healthcare$severe, death = 0)
cost_societal <- c(mci = c.societal$mci, mild = c.societal$mild, moderate = c.societal$moderate, severe = c.societal$severe, death = 0)

# ARIA symptomatic proportions and costs
p_sympt_aria <- list(aducanumab = 0.24, donanemab = 0.16)
aria_cost <- c.aria$outpatient_visit + c.aria$followup_imaging + 0.04 * c.aria$hospitalization
aria_utility_loss <- -u.aria_decrement

# Treatment discontinuation functions
discontinuation_adu <- function(cycle) {
  if (cycle <= 12) return(1 - (0.38 * (cycle / 12)))
  return(1 - 0.38)
}
discontinuation_don <- function(cycle) {
  if (cycle <= 6) return(1 - 0.27)
  if (cycle <= 12) return(1 - 0.55)
  return(1 - 0.55)
}

# Run model for each strategy
for (strategy in c("no intervention", "aducanumab", "donanemab")) {
  strat_key <- gsub(" ", "_", strategy)
  trans <- build_transition_matrix(strategy)
  
  trace <- matrix(0, nrow = n_cycles + 1, ncol = length(states), dimnames = list(NULL, states))
  trace[1, ] <- initial_state_dist * n_individuals
  
  total_qalys <- 0
  total_healthcare <- 0
  total_societal <- 0
  on_treatment <- rep(1, n_individuals)
  
  for (t in 1:n_cycles) {
    trace[t + 1, ] <- trace[t, ] %*% trans[, , t]
    
    current <- trace[t, ]
    
    # Determine active treatment proportion
    if (strategy == "aducanumab") {
      treatment_prop <- discontinuation_adu(t)
    } else if (strategy == "donanemab") {
      treatment_prop <- discontinuation_don(t)
    } else {
      treatment_prop <- 0
    }
    
    # Determine patients on treatment (exclude "severe")
    treatable <- current["mci"] + current["mild"] + current["moderate"]
    on_tx <- treatable * treatment_prop
    
    # Utilities and QALYs
    cycle_qalys <- sum(current * utilities)
    
    # Apply ARIA disutility in cycles 1-3
    if (t <= 3 && strategy != "no intervention") {
      cycle_qalys <- cycle_qalys + (on_tx * p_sympt_aria[[strategy]] * aria_utility_loss)
    }
    
    # Apply discounting
    discount_factor <- (1 / (1 + discount_rate)) ^ t
    total_qalys <- total_qalys + (cycle_qalys * discount_factor)
    
    # Healthcare & societal costs
    cycle_hc_cost <- sum(current * cost_healthcare)
    cycle_soc_cost <- sum(current * (cost_healthcare + cost_societal))
    
    # Treatment-specific costs
    if (strategy == "aducanumab") {
      if (t <= 6) add_cost <- c.infusion_monitoring$aducanumab["m1_6"]
      else if (t <= 12) add_cost <- c.infusion_monitoring$aducanumab["m7_12"]
      else add_cost <- c.infusion_monitoring$aducanumab["m13plus"]
      drug_cost <- if (t <= 12) c.aducanumab_year1 / 12 else c.aducanumab_year2plus / 12
    } else if (strategy == "donanemab") {
      if (t <= 6) add_cost <- c.infusion_monitoring$donanemab["m1_6"]
      else if (t <= 12) add_cost <- c.infusion_monitoring$donanemab["m7_12"]
      else add_cost <- c.infusion_monitoring$donanemab["m13plus"]
      drug_cost <- c.donanemab / 12
    } else {
      add_cost <- 0
      drug_cost <- 0
    }
    
    cycle_hc_cost <- cycle_hc_cost + on_tx * (add_cost + drug_cost)
    cycle_soc_cost <- cycle_soc_cost + on_tx * (add_cost + drug_cost)
    
    # ARIA costs in cycles 1-3
    if (t <= 3 && strategy != "no intervention") {
      aria_events <- on_tx * p_sympt_aria[[strategy]]
      cycle_hc_cost <- cycle_hc_cost + aria_events * aria_cost
      cycle_soc_cost <- cycle_soc_cost + aria_events * aria_cost
    }
    
    # Apply discounting
    total_healthcare <- total_healthcare + (cycle_hc_cost * discount_factor)
    total_societal <- total_societal + (cycle_soc_cost * discount_factor)
  }
  
  # Add screening cost (only once at start)
  if (strategy == "aducanumab") {
    total_healthcare <- total_healthcare + c.screening$aducanumab
    total_societal <- total_societal + c.screening$aducanumab
  } else if (strategy == "donanemab") {
    total_healthcare <- total_healthcare + c.screening$donanemab
    total_societal <- total_societal + c.screening$donanemab
  }
  
  # Save results per person
  results[[strat_key]] <- list(
    qalys = total_qalys / n_individuals,
    healthcare_costs = total_healthcare / n_individuals,
    societal_costs = total_societal / n_individuals
  )
}
```
