## Supplementary material for "Replicating Health-Economic Simulation Models for Alzheimer’s Disease Using Artificial Intelligence": final R scripts: Ross_O10.R

```
# Health Economic Markov Model Inputs Based on Ross et al. (2022)

# Define state names
states <- c("mci", "mild", "moderate", "severe", "death")

# Define treatment strategies
strategies <- c("standard_care", "aducanumab", "donanemab")

# Simulation parameters
n_cohort <- 1000
start_age <- 75.2
cycle_length <- 1 / 12  # monthly cycles
annual_discount_rate <- 0.03

# Transition probabilities (monthly)
p.mci_to_mild <- 0.007
p.mild_to_moderate <- 0.016
p.moderate_to_severe <- 0.026

# Treatment hazard ratios for disease progression
hr_prog_aducanumab <- 0.89
hr_prog_donanemab <- 0.68

# Mortality hazard ratios by health state
hr_mort_mci <- 1.61
hr_mort_mild <- 2.23
hr_mort_moderate <- 3.10
hr_mort_severe <- 4.98

# Utilities (converted to monthly by dividing by 12)
u_mci <- 0.73 / 12
u_mild <- 0.69 / 12
u_moderate <- 0.53 / 12
u_severe <- 0.34 / 12
u_aria_decrement <- -0.065 / 12  # lasts 3 months

# Healthcare sector monthly costs per health state (USD)
c_hc_mci <- 691
c_hc_mild <- 1049
c_hc_moderate <- 1274
c_hc_severe <- 3764

# Societal additional monthly costs per health state (USD)
c_soc_mci <- 49
c_soc_mild <- 1483
c_soc_moderate <- 1932
c_soc_severe <- 2153

# Annual drug costs (USD)
drug_cost_aducanumab_year1 <- 20500
drug_cost_aducanumab_year2plus <- 28200
drug_cost_donanemab <- 28200

# Monthly infusion and monitoring costs (USD)
# Aducanumab
c_aduca_m1_6 <- 112
c_aduca_m7_12 <- 87
c_aduca_m13plus <- 80

# Donanemab
c_dona_m1_6 <- 599
c_dona_m7_12 <- 302
c_dona_m13plus <- 287

# One-time screening costs (USD)
screening_cost_aducanumab <- 6957
screening_cost_donanemab <- 17096

# ARIA-related costs
c_aria_imaging <- 1060
c_aria_hospitalization <- 35934
c_aria_outpatient <- 128

# ARIA probabilities (monthly proportions will be defined later if needed)
p_aria_aducanumab <- 0.41
p_aria_donanemab <- 0.39
p_aria_symptomatic_aducanumab <- 0.41 * 0.24
p_aria_symptomatic_donanemab <- 0.39 * 0.16


# Load mortality data
mortality_data <- read.csv("mortalityRates_US_2017.csv")

# Calculate age-specific death probabilities for each disease state
mortality_data$prob_death_mci <- 1 - exp(-mortality_data$monthly_rate * hr_mort_mci)
mortality_data$prob_death_mild <- 1 - exp(-mortality_data$monthly_rate * hr_mort_mild)
mortality_data$prob_death_moderate <- 1 - exp(-mortality_data$monthly_rate * hr_mort_moderate)
mortality_data$prob_death_severe <- 1 - exp(-mortality_data$monthly_rate * hr_mort_severe)


# Define number of cycles (assuming 12 cycles per year over a lifetime horizon, e.g., 30 years)
n_cycles <- 30 * 12

# --- Standard care (no intervention) ---
p_mci_to_mild_std <- rep(p.mci_to_mild, n_cycles)
p_mci_to_mci_std <- rep(1 - p.mci_to_mild, n_cycles)

p_mild_to_moderate_std <- rep(p.mild_to_moderate, n_cycles)
p_mild_to_mild_std <- rep(1 - p.mild_to_moderate, n_cycles)

p_moderate_to_severe_std <- rep(p.moderate_to_severe, n_cycles)
p_moderate_to_moderate_std <- rep(1 - p.moderate_to_severe, n_cycles)

p_severe_to_severe_std <- rep(1, n_cycles)  # No transition out of severe except death

# --- Aducanumab ---
# MCI to mild
rate_mci <- -log(1 - p.mci_to_mild)
adj_rate_mci_aducanumab <- rate_mci * hr_prog_aducanumab
p_mci_to_mild_adu <- rep(1 - exp(-adj_rate_mci_aducanumab), n_cycles)
p_mci_to_mci_adu <- rep(1 - p_mci_to_mild_adu[1], n_cycles)

# Mild to moderate
rate_mild <- -log(1 - p.mild_to_moderate)
adj_rate_mild_aducanumab <- rate_mild * hr_prog_aducanumab
p_mild_to_moderate_adu <- rep(1 - exp(-adj_rate_mild_aducanumab), n_cycles)
p_mild_to_mild_adu <- rep(1 - p_mild_to_moderate_adu[1], n_cycles)

# Moderate to severe
rate_moderate <- -log(1 - p.moderate_to_severe)
adj_rate_moderate_aducanumab <- rate_moderate * hr_prog_aducanumab
p_moderate_to_severe_adu <- rep(1 - exp(-adj_rate_moderate_aducanumab), n_cycles)
p_moderate_to_moderate_adu <- rep(1 - p_moderate_to_severe_adu[1], n_cycles)

# Severe state
p_severe_to_severe_adu <- rep(1, n_cycles)

# --- Donanemab ---
# MCI to mild
adj_rate_mci_donanemab <- rate_mci * hr_prog_donanemab
p_mci_to_mild_dona <- rep(1 - exp(-adj_rate_mci_donanemab), n_cycles)
p_mci_to_mci_dona <- rep(1 - p_mci_to_mild_dona[1], n_cycles)

# Mild to moderate
adj_rate_mild_donanemab <- rate_mild * hr_prog_donanemab
p_mild_to_moderate_dona <- rep(1 - exp(-adj_rate_mild_donanemab), n_cycles)
p_mild_to_mild_dona <- rep(1 - p_mild_to_moderate_dona[1], n_cycles)

# Moderate to severe
adj_rate_moderate_donanemab <- rate_moderate * hr_prog_donanemab
p_moderate_to_severe_dona <- rep(1 - exp(-adj_rate_moderate_donanemab), n_cycles)
p_moderate_to_moderate_dona <- rep(1 - p_moderate_to_severe_dona[1], n_cycles)

# Severe state
p_severe_to_severe_dona <- rep(1, n_cycles)


# Function to build the transition probability matrix
build_transition_matrix <- function(strat) {
  # Define number of states and initialize transition probability array
  n_states <- length(states)
  tpm <- array(0, dim = c(n_states, n_states, n_cycles),
               dimnames = list(states, states, NULL))
  
  # Loop over cycles
  for (t in 1:n_cycles) {
    # Get age for this cycle (starting age + months converted to years)
    age_t <- floor(start_age + (t - 1) / 12)
    
    # Skip if age is outside bounds of mortality data
    if (age_t < min(mortality_data$age) || age_t > max(mortality_data$age)) {
      next
    }
    
    # Subset mortality data for this age
    mort_row <- mortality_data[mortality_data$age == age_t, ]
    
    # Assign mortality transition probabilities per state
    tpm["mci", "death", t] <- mort_row$prob_death_mci
    tpm["mild", "death", t] <- mort_row$prob_death_mild
    tpm["moderate", "death", t] <- mort_row$prob_death_moderate
    tpm["severe", "death", t] <- mort_row$prob_death_severe
    
    # Absorbing state: probability of staying in death is 1
    tpm["death", "death", t] <- 1
    
    # Transitions for strategy-specific progression
    if (strat == "no intervention") {
      tpm["mci", "mild", t]     <- p_mci_to_mild_std[t] * (1 - tpm["mci", "death", t])
      tpm["mci", "mci", t]      <- p_mci_to_mci_std[t] * (1 - tpm["mci", "death", t])
      
      tpm["mild", "moderate", t] <- p_mild_to_moderate_std[t] * (1 - tpm["mild", "death", t])
      tpm["mild", "mild", t]     <- p_mild_to_mild_std[t] * (1 - tpm["mild", "death", t])
      
      tpm["moderate", "severe", t] <- p_moderate_to_severe_std[t] * (1 - tpm["moderate", "death", t])
      tpm["moderate", "moderate", t] <- p_moderate_to_moderate_std[t] * (1 - tpm["moderate", "death", t])
      
      tpm["severe", "severe", t] <- p_severe_to_severe_std[t] * (1 - tpm["severe", "death", t])
      
    } else if (strat == "aducanumab") {
      tpm["mci", "mild", t]     <- p_mci_to_mild_adu[t] * (1 - tpm["mci", "death", t])
      tpm["mci", "mci", t]      <- p_mci_to_mci_adu[t] * (1 - tpm["mci", "death", t])
      
      tpm["mild", "moderate", t] <- p_mild_to_moderate_adu[t] * (1 - tpm["mild", "death", t])
      tpm["mild", "mild", t]     <- p_mild_to_mild_adu[t] * (1 - tpm["mild", "death", t])
      
      tpm["moderate", "severe", t] <- p_moderate_to_severe_adu[t] * (1 - tpm["moderate", "death", t])
      tpm["moderate", "moderate", t] <- p_moderate_to_moderate_adu[t] * (1 - tpm["moderate", "death", t])
      
      tpm["severe", "severe", t] <- p_severe_to_severe_adu[t] * (1 - tpm["severe", "death", t])
      
    } else if (strat == "donanemab") {
      tpm["mci", "mild", t]     <- p_mci_to_mild_dona[t] * (1 - tpm["mci", "death", t])
      tpm["mci", "mci", t]      <- p_mci_to_mci_dona[t] * (1 - tpm["mci", "death", t])
      
      tpm["mild", "moderate", t] <- p_mild_to_moderate_dona[t] * (1 - tpm["mild", "death", t])
      tpm["mild", "mild", t]     <- p_mild_to_mild_dona[t] * (1 - tpm["mild", "death", t])
      
      tpm["moderate", "severe", t] <- p_moderate_to_severe_dona[t] * (1 - tpm["moderate", "death", t])
      tpm["moderate", "moderate", t] <- p_moderate_to_moderate_dona[t] * (1 - tpm["moderate", "death", t])
      
      tpm["severe", "severe", t] <- p_severe_to_severe_dona[t] * (1 - tpm["severe", "death", t])
    }
    
    # Normalize each row to ensure they sum to 1
    for (s in states) {
      row_sum <- sum(tpm[s, , t])
      if (row_sum > 0 && row_sum != 1) {
        tpm[s, , t] <- tpm[s, , t] / row_sum
      }
    }
  }
  
  return(tpm)
}

# Monthly discount factor
discount_monthly <- 1 / (1 + annual_discount_rate)^(1 / 12)

# Initialize results list
results <- list()

# Initial state distribution
init_dist <- c(mci = 0.65, mild = 0.35, moderate = 0, severe = 0, death = 0)

# Function to run Markov simulation
run_markov <- function(strat) {
  trace <- matrix(0, nrow = n_cycles + 1, ncol = length(states), dimnames = list(NULL, states))
  trace[1, ] <- init_dist
  
  tpm <- build_transition_matrix(strat)
  on_treatment <- rep(TRUE, n_cycles + 1)
  
  if (strat == "aducanumab") {
    on_treatment[1:13] <- 1 - seq(0, 0.38, length.out = 13)
    on_treatment[14:(n_cycles + 1)] <- on_treatment[13]
  } else if (strat == "donanemab") {
    on_treatment[1:7] <- 1 - 0.27
    on_treatment[8:13] <- 1 - 0.55
    on_treatment[14:(n_cycles + 1)] <- on_treatment[13]
  }
  
  qalys <- 0
  cost_healthcare <- 0
  cost_societal <- 0
  
  for (t in 1:n_cycles) {
    trace[t + 1, ] <- trace[t, ] %*% tpm[, , t]
    
    if (strat != "no intervention") {
      on_treatment[t + 1] <- on_treatment[t + 1] * (1 - trace[t + 1, "severe"])
    }
    
    q_t <- sum(trace[t, "mci"] * u_mci,
               trace[t, "mild"] * u_mild,
               trace[t, "moderate"] * u_moderate,
               trace[t, "severe"] * u_severe)
    
    if (t <= 3 && strat != "no intervention") {
      aria_decr <- ifelse(strat == "aducanumab", p_aria_symptomatic_aducanumab, p_aria_symptomatic_donanemab)
      q_t <- q_t + (sum(trace[t, c("mci", "mild", "moderate")]) * u_aria_decrement * aria_decr)
    }
    
    qalys <- qalys + q_t * discount_monthly^(t - 1)
    
    c_h_t <- sum(trace[t, "mci"] * c_hc_mci,
                 trace[t, "mild"] * c_hc_mild,
                 trace[t, "moderate"] * c_hc_moderate,
                 trace[t, "severe"] * c_hc_severe)
    
    c_soc_t <- sum(trace[t, "mci"] * c_soc_mci,
                   trace[t, "mild"] * c_soc_mild,
                   trace[t, "moderate"] * c_soc_moderate,
                   trace[t, "severe"] * c_soc_severe)
    
    drug_cost <- 0
    inf_mon_cost <- 0
    
    if (strat == "aducanumab" && on_treatment[t]) {
      drug_cost <- if (t <= 12) drug_cost_aducanumab_year1 else drug_cost_aducanumab_year2plus
      inf_mon_cost <- if (t <= 6) c_aduca_m1_6 else if (t <= 12) c_aduca_m7_12 else c_aduca_m13plus
    } else if (strat == "donanemab" && on_treatment[t]) {
      drug_cost <- drug_cost_donanemab
      inf_mon_cost <- if (t <= 6) c_dona_m1_6 else if (t <= 12) c_dona_m7_12 else c_dona_m13plus
    }
    
    n_treat <- if (strat == "no intervention") 0 else trace[t, "mci"] + trace[t, "mild"] + trace[t, "moderate"]
    n_treat <- n_treat * on_treatment[t]
    
    c_drug <- drug_cost / 12 * n_treat
    c_inf <- inf_mon_cost * n_treat
    
    if (t <= 3 && strat != "no intervention") {
      aria_prob <- if (strat == "aducanumab") p_aria_symptomatic_aducanumab else p_aria_symptomatic_donanemab
      n_aria <- sum(trace[t, c("mci", "mild", "moderate")]) * aria_prob
      c_aria <- n_aria * (c_aria_outpatient + c_aria_imaging + 0.04 * c_aria_hospitalization)
    } else {
      c_aria <- 0
    }
    
    cost_h <- (c_h_t + c_drug + c_inf + c_aria) * discount_monthly^(t - 1)
    cost_s <- (c_h_t + c_soc_t + c_drug + c_inf + c_aria) * discount_monthly^(t - 1)
    
    cost_healthcare <- cost_healthcare + cost_h
    cost_societal <- cost_societal + cost_s
  }
  
  screening_cost <- if (strat == "aducanumab") screening_cost_aducanumab else if (strat == "donanemab") screening_cost_donanemab else 0
  cost_healthcare <- cost_healthcare + screening_cost
  cost_societal <- cost_societal + screening_cost
  
  return(list(
    qalys = qalys,
    healthcare_costs = cost_healthcare,
    societal_costs = cost_societal
  ))
}

# Run model for each strategy
results$no_intervention <- run_markov("no intervention")
results$aducanumab <- run_markov("aducanumab")
results$donanemab <- run_markov("donanemab")
```
