## Supplementary material for "Replicating Health-Economic Simulation Models for Alzheimer’s Disease Using Artificial Intelligence": final R scripts: Ross_O11.R

```
# Define state and strategy names
states <- c("mci", "mild", "moderate", "severe", "death")
strategies <- c("standard_care", "aducanumab", "donanemab")

# Cohort characteristics
initial_age <- 75.2
cohort_size <- 1000
initial_distribution <- c(mci = 0.65, mild = 0.35, moderate = 0, severe = 0, death = 0)

# Disease progression probabilities (monthly)
p.mci_to_mild <- 0.007
p.mild_to_moderate <- 0.016
p.moderate_to_severe <- 0.026

# Hazard ratios for mortality by disease state
hr.mci <- 1.61
hr.mild <- 2.23
hr.moderate <- 3.10
hr.severe <- 4.98

# Treatment efficacy: hazard ratios for disease progression
hr_prog.aducanumab <- 0.89
hr_prog.donanemab <- 0.68

# Utilities (converted from annual to monthly)
util.mci <- 0.73 / 12
util.mild <- 0.69 / 12
util.moderate <- 0.53 / 12
util.severe <- 0.34 / 12
util.death <- 0

# Utility decrement for symptomatic ARIA (lasts 3 months)
util_aria_decrement <- -0.065 / 12

# Monthly healthcare sector costs (USD)
cost.healthcare.mci <- 691
cost.healthcare.mild <- 1049
cost.healthcare.moderate <- 1274
cost.healthcare.severe <- 3764
cost.healthcare.death <- 0

# Monthly additional societal costs (USD)
cost.societal.mci <- 49
cost.societal.mild <- 1483
cost.societal.moderate <- 1932
cost.societal.severe <- 2153
cost.societal.death <- 0

# Annual drug costs (USD)
drug_cost.aducanumab.year1 <- 20500
drug_cost.aducanumab.subsequent <- 28200
drug_cost.donanemab <- 28200

# Monthly infusion and monitoring costs (USD)
cost.infusion.aducanumab.months_1_6 <- 112
cost.infusion.aducanumab.months_7_12 <- 87
cost.infusion.aducanumab.after_12 <- 80
cost.infusion.donanemab.months_1_6 <- 599
cost.infusion.donanemab.months_7_12 <- 302
cost.infusion.donanemab.after_12 <- 287

# Screening costs (USD per eligible patient)
cost.screening.aducanumab <- 6957
cost.screening.donanemab <- 17096

# ARIA-related costs (USD)
cost.aria.followup_imaging <- 1060
cost.aria.hospitalization <- 35934
cost.aria.outpatient_visit <- 128

# ARIA incidence
p.aria.aducanumab <- 0.41
p.aria_symptomatic.aducanumab <- 0.24 * p.aria.aducanumab
p.aria.donanemab <- 0.39
p.aria_symptomatic.donanemab <- 0.16 * p.aria.donanemab


# Load required library
library(readr)

# Load mortality rate data
mortality_data <- read_csv("mortalityRates_US_2017.csv")

# Compute age-specific probability of death by disease severity
mortality_data <- mortality_data |>
  mutate(
    prob_death_mci = 1 - exp(-monthly_rate * hr.mci),
    prob_death_mild = 1 - exp(-monthly_rate * hr.mild),
    prob_death_moderate = 1 - exp(-monthly_rate * hr.moderate),
    prob_death_severe = 1 - exp(-monthly_rate * hr.severe)
  )

# Retain only relevant columns
mortality_probs <- mortality_data[, c("age", "prob_death_mci", "prob_death_mild", "prob_death_moderate", "prob_death_severe")]


# Number of cycles
n_cycles <- 360  # e.g., 30 years with monthly cycles

# Standard care (no intervention)
p_mci_to_mild_std <- rep(p.mci_to_mild, n_cycles)
p_mci_stay_std <- 1 - p_mci_to_mild_std

p_mild_to_moderate_std <- rep(p.mild_to_moderate, n_cycles)
p_mild_stay_std <- 1 - p_mild_to_moderate_std

p_moderate_to_severe_std <- rep(p.moderate_to_severe, n_cycles)
p_moderate_stay_std <- 1 - p_moderate_to_severe_std

p_severe_stay_std <- rep(1, n_cycles)  # no further progression defined beyond severe (death handled separately)

# Aducanumab intervention
rate_mci_to_mild <- -log(1 - p.mci_to_mild)
adj_rate_mci_to_mild_adu <- rate_mci_to_mild * hr_prog.aducanumab
p_mci_to_mild_adu <- rep(1 - exp(-adj_rate_mci_to_mild_adu), n_cycles)
p_mci_stay_adu <- 1 - p_mci_to_mild_adu

rate_mild_to_moderate <- -log(1 - p.mild_to_moderate)
adj_rate_mild_to_moderate_adu <- rate_mild_to_moderate * hr_prog.aducanumab
p_mild_to_moderate_adu <- rep(1 - exp(-adj_rate_mild_to_moderate_adu), n_cycles)
p_mild_stay_adu <- 1 - p_mild_to_moderate_adu

rate_moderate_to_severe <- -log(1 - p.moderate_to_severe)
adj_rate_moderate_to_severe_adu <- rate_moderate_to_severe * hr_prog.aducanumab
p_moderate_to_severe_adu <- rep(1 - exp(-adj_rate_moderate_to_severe_adu), n_cycles)
p_moderate_stay_adu <- 1 - p_moderate_to_severe_adu

p_severe_stay_adu <- rep(1, n_cycles)

# Donanemab intervention
adj_rate_mci_to_mild_don <- rate_mci_to_mild * hr_prog.donanemab
p_mci_to_mild_don <- rep(1 - exp(-adj_rate_mci_to_mild_don), n_cycles)
p_mci_stay_don <- 1 - p_mci_to_mild_don

adj_rate_mild_to_moderate_don <- rate_mild_to_moderate * hr_prog.donanemab
p_mild_to_moderate_don <- rep(1 - exp(-adj_rate_mild_to_moderate_don), n_cycles)
p_mild_stay_don <- 1 - p_mild_to_moderate_don

adj_rate_moderate_to_severe_don <- rate_moderate_to_severe * hr_prog.donanemab
p_moderate_to_severe_don <- rep(1 - exp(-adj_rate_moderate_to_severe_don), n_cycles)
p_moderate_stay_don <- 1 - p_moderate_to_severe_don

p_severe_stay_don <- rep(1, n_cycles)


# Function to build transition probability matrix
build_transition_matrix <- function(strat) {
  # Initialize transition array: from state × to state × cycle
  trans_mat <- array(0, dim = c(length(states), length(states), n_cycles),
                     dimnames = list(from = states, to = states, cycle = 1:n_cycles))
  
  for (t in 1:n_cycles) {
    age_t <- initial_age + (t - 1) / 12
    age_index <- which.min(abs(mortality_probs$age - age_t))
    
    # Get death probabilities from each state based on age
    p_death_mci <- mortality_probs$prob_death_mci[age_index]
    p_death_mild <- mortality_probs$prob_death_mild[age_index]
    p_death_moderate <- mortality_probs$prob_death_moderate[age_index]
    p_death_severe <- mortality_probs$prob_death_severe[age_index]
    
    # MCI row
    trans_mat["mci", "death", t] <- p_death_mci
    
    # Mild row
    trans_mat["mild", "death", t] <- p_death_mild
    
    # Moderate row
    trans_mat["moderate", "death", t] <- p_death_moderate
    
    # Severe row
    trans_mat["severe", "death", t] <- p_death_severe
    
    # Death is an absorbing state
    trans_mat["death", "death", t] <- 1
    
    if (strat == "no intervention") {
      trans_mat["mci", "mild", t]     <- p_mci_to_mild_std[t]     * (1 - p_death_mci)
      trans_mat["mci", "mci", t]      <- p_mci_stay_std[t]        * (1 - p_death_mci)
      
      trans_mat["mild", "moderate", t] <- p_mild_to_moderate_std[t] * (1 - p_death_mild)
      trans_mat["mild", "mild", t]     <- p_mild_stay_std[t]        * (1 - p_death_mild)
      
      trans_mat["moderate", "severe", t] <- p_moderate_to_severe_std[t] * (1 - p_death_moderate)
      trans_mat["moderate", "moderate", t] <- p_moderate_stay_std[t]    * (1 - p_death_moderate)
      
      trans_mat["severe", "severe", t] <- p_severe_stay_std[t] * (1 - p_death_severe)
      
    } else if (strat == "aducanumab") {
      trans_mat["mci", "mild", t]     <- p_mci_to_mild_adu[t]     * (1 - p_death_mci)
      trans_mat["mci", "mci", t]      <- p_mci_stay_adu[t]        * (1 - p_death_mci)
      
      trans_mat["mild", "moderate", t] <- p_mild_to_moderate_adu[t] * (1 - p_death_mild)
      trans_mat["mild", "mild", t]     <- p_mild_stay_adu[t]        * (1 - p_death_mild)
      
      trans_mat["moderate", "severe", t] <- p_moderate_to_severe_adu[t] * (1 - p_death_moderate)
      trans_mat["moderate", "moderate", t] <- p_moderate_stay_adu[t]    * (1 - p_death_moderate)
      
      trans_mat["severe", "severe", t] <- p_severe_stay_adu[t] * (1 - p_death_severe)
      
    } else if (strat == "donanemab") {
      trans_mat["mci", "mild", t]     <- p_mci_to_mild_don[t]     * (1 - p_death_mci)
      trans_mat["mci", "mci", t]      <- p_mci_stay_don[t]        * (1 - p_death_mci)
      
      trans_mat["mild", "moderate", t] <- p_mild_to_moderate_don[t] * (1 - p_death_mild)
      trans_mat["mild", "mild", t]     <- p_mild_stay_don[t]        * (1 - p_death_mild)
      
      trans_mat["moderate", "severe", t] <- p_moderate_to_severe_don[t] * (1 - p_death_moderate)
      trans_mat["moderate", "moderate", t] <- p_moderate_stay_don[t]    * (1 - p_death_moderate)
      
      trans_mat["severe", "severe", t] <- p_severe_stay_don[t] * (1 - p_death_severe)
    }
    
    # Normalize rows to ensure row sums equal 1
    for (state in states[-length(states)]) {  # exclude "death"
      row_sum <- sum(trans_mat[state, , t])
      if (abs(row_sum - 1) > 1e-8) {
        trans_mat[state, , t] <- trans_mat[state, , t] / row_sum
      }
    }
  }
  
  return(trans_mat)
}


# Load necessary library
library(dplyr)

# Discount factor per month
discount_rate_monthly <- (1 + 0.03)^(1 / 12) - 1
discount_factors <- 1 / ((1 + discount_rate_monthly)^(0:(n_cycles - 1)))

# Initialize result storage
results <- list()

# Function to run the Markov model for a given strategy
run_model <- function(strategy_name) {
  tm <- build_transition_matrix(strategy_name)
  trace <- matrix(0, nrow = n_cycles + 1, ncol = length(states),
                  dimnames = list(cycle = 0:n_cycles, state = states))
  trace[1, ] <- cohort_size * initial_distribution
  
  # Track treatment status
  treated <- rep(0, n_cycles + 1)
  if (strategy_name != "no intervention") {
    treated[1] <- sum(trace[1, c("mci", "mild")])
  }
  
  # Discontinuation logic
  if (strategy_name == "aducanumab") {
    discontinuation_rate <- 0.38 / 12
  } else if (strategy_name == "donanemab") {
    discont_schedule <- c(rep(0.27, 1), rep(0.28, 1), rep(0, n_cycles - 2))
  }
  
  # ARIA symptomatic proportion
  if (strategy_name == "aducanumab") {
    aria_sympt_prob <- p.aria_symptomatic.aducanumab
  } else if (strategy_name == "donanemab") {
    aria_sympt_prob <- p.aria_symptomatic.donanemab
  } else {
    aria_sympt_prob <- 0
  }
  
  # Trackers
  qalys <- rep(0, n_cycles)
  healthcare_costs <- rep(0, n_cycles)
  societal_costs <- rep(0, n_cycles)
  on_treatment <- treated[1]
  
  for (t in 1:n_cycles) {
    trace[t + 1, ] <- trace[t, ] %*% tm[ , , t]
    
    # Determine treatment population
    if (strategy_name == "aducanumab") {
      on_treatment <- on_treatment * (1 - discontinuation_rate)
    } else if (strategy_name == "donanemab") {
      on_treatment <- on_treatment * (1 - discont_schedule[t])
    }
    
    # Remove severe patients from treatment
    non_severe_treatment <- sum(trace[t + 1, c("mci", "mild", "moderate")])
    on_treatment <- min(on_treatment, non_severe_treatment)
    
    # QALYs
    u <- c(util.mci, util.mild, util.moderate, util.severe, util.death)
    qaly_t <- sum(trace[t + 1, ] * u)
    
    # ARIA disutility in cycles 1–3
    aria_decrement <- 0
    if (t <= 3 && strategy_name != "no intervention") {
      aria_decrement <- on_treatment * aria_sympt_prob * util_aria_decrement
    }
    
    qalys[t] <- (qaly_t + aria_decrement) * discount_factors[t]
    
    # Healthcare and societal costs
    hc_cost <- c(cost.healthcare.mci, cost.healthcare.mild, cost.healthcare.moderate,
                 cost.healthcare.severe, cost.healthcare.death)
    soc_cost <- c(cost.societal.mci, cost.societal.mild, cost.societal.moderate,
                  cost.societal.severe, cost.societal.death)
    
    healthcare_costs[t] <- sum(trace[t + 1, ] * hc_cost) * discount_factors[t]
    societal_costs[t] <- (sum(trace[t + 1, ] * (hc_cost + soc_cost))) * discount_factors[t]
    
    # Treatment-related costs
    if (strategy_name == "aducanumab") {
      if (t <= 6) {
        infusion_cost <- cost.infusion.aducanumab.months_1_6
      } else if (t <= 12) {
        infusion_cost <- cost.infusion.aducanumab.months_7_12
      } else {
        infusion_cost <- cost.infusion.aducanumab.after_12
      }
      
      if (t <= 12) {
        drug_cost <- drug_cost.aducanumab.year1 / 12
      } else {
        drug_cost <- drug_cost.aducanumab.subsequent / 12
      }
      
    } else if (strategy_name == "donanemab") {
      if (t <= 6) {
        infusion_cost <- cost.infusion.donanemab.months_1_6
      } else if (t <= 12) {
        infusion_cost <- cost.infusion.donanemab.months_7_12
      } else {
        infusion_cost <- cost.infusion.donanemab.after_12
      }
      
      drug_cost <- drug_cost.donanemab / 12
      
    } else {
      infusion_cost <- 0
      drug_cost <- 0
    }
    
    monitoring_cost <- infusion_cost
    total_treatment_cost <- on_treatment * (drug_cost + infusion_cost + monitoring_cost) * discount_factors[t]
    
    # ARIA-related costs
    if (strategy_name != "no intervention" && t <= 24) {
      aria_cases <- on_treatment * aria_sympt_prob
      outpatient <- cost.aria.outpatient_visit * aria_cases
      mri <- cost.aria.followup_imaging * aria_cases
      hospital <- cost.aria.hospitalization * 0.04 * aria_cases
      aria_costs <- outpatient + mri + hospital
    } else {
      aria_costs <- 0
    }
    
    healthcare_costs[t] <- healthcare_costs[t] + total_treatment_cost + aria_costs
    societal_costs[t] <- societal_costs[t] + total_treatment_cost + aria_costs
  }
  
  # Add screening costs once at start
  if (strategy_name == "aducanumab") {
    healthcare_costs[1] <- healthcare_costs[1] + cost.screening.aducanumab * discount_factors[1] * cohort_size
    societal_costs[1] <- societal_costs[1] + cost.screening.aducanumab * discount_factors[1] * cohort_size
  } else if (strategy_name == "donanemab") {
    healthcare_costs[1] <- healthcare_costs[1] + cost.screening.donanemab * discount_factors[1] * cohort_size
    societal_costs[1] <- societal_costs[1] + cost.screening.donanemab * discount_factors[1] * cohort_size
  }
  
  return(list(
    qalys = sum(qalys) / cohort_size,
    healthcare_costs = sum(healthcare_costs) / cohort_size,
    societal_costs = sum(societal_costs) / cohort_size
  ))
}

# Run for all strategies
results$no_intervention <- run_model("no intervention")
results$aducanumab <- run_model("aducanumab")
results$donanemab <- run_model("donanemab")
```
