## Supplementary material for "Replicating Health-Economic Simulation Models for Alzheimer’s Disease Using Artificial Intelligence": final R scripts: Ross_O12.R

```
# =============================
# Markov Model Input Parameters
# Ross et al. (2022)
# =============================

# -----------------------------
# Descriptive Parameters
# -----------------------------
state_names <- c("mci", "mild", "moderate", "severe", "death")
strategy_names <- c("standard_care", "aducanumab", "donanemab")
initial_age <- 75.2
cohort_size <- 1000

# -----------------------------
# Initial State Distribution
# -----------------------------
initial_state_dist <- c(mci = 0.65, mild = 0.35, moderate = 0, severe = 0, death = 0)

# -----------------------------
# Disease Progression Probabilities (Monthly)
# -----------------------------
p.mci_to_mild <- 0.007
p.mild_to_moderate <- 0.016
p.moderate_to_severe <- 0.026

# -----------------------------
# Treatment Effects (Hazard Ratios for Progression)
# -----------------------------
hr.aducanumab <- 0.89
hr.donanemab <- 0.68

# -----------------------------
# Utilities (Annual, converted to Monthly)
# -----------------------------
u.mci <- 0.73 / 12
u.mild <- 0.69 / 12
u.moderate <- 0.53 / 12
u.severe <- 0.34 / 12
u.symptomatic_aria <- -0.065 / 12  # Applied as decrement for 3 months

# -----------------------------
# Mortality Hazard Ratios by Health State
# -----------------------------
hr.mci_mortality <- 1.61
hr.mild_mortality <- 2.23
hr.moderate_mortality <- 3.10
hr.severe_mortality <- 4.98

# -----------------------------
# Costs (Monthly, 2020 USD)
# -----------------------------
# Background Healthcare Sector Costs
c.mci.healthcare <- 691
c.mild.healthcare <- 1049
c.moderate.healthcare <- 1274
c.severe.healthcare <- 3764

# Additional Societal Costs
c.mci.societal <- 49
c.mild.societal <- 1483
c.moderate.societal <- 1932
c.severe.societal <- 2153

# Drug Costs
c.aducanumab.year1 <- 20500
c.aducanumab.annual <- 28200
c.donanemab.annual <- 28200

# Monthly Infusion and Monitoring Costs
c.aducanumab.months1_6 <- 112
c.aducanumab.months7_12 <- 87
c.aducanumab.months13plus <- 80

c.donanemab.months1_6 <- 599
c.donanemab.months7_12 <- 302
c.donanemab.months13plus <- 287

# Screening Costs (one-time per eligible patient)
c.screening.aducanumab <- 6957
c.screening.donanemab <- 17096

# ARIA-related Costs
c.aria.followup_imaging <- 1060
c.aria.hospitalization <- 35934
c.aria.outpatient_visit <- 128

# ARIA-related Probabilities (Total and Symptomatic)
p.aria.aducanumab_total <- 0.41
p.aria.aducanumab_symptomatic <- 0.41 * 0.24
p.aria.donanemab_total <- 0.39
p.aria.donanemab_symptomatic <- 0.39 * 0.16


# =============================
# Load and Process Mortality Rates
# =============================

# Load the mortality life table
mortality_data <- read.csv("mortalityRates_US_2017.csv")

# Compute age-specific death probabilities by disease severity
mortality_data$prob_death_mci <- 1 - exp(-mortality_data$monthly_rate * hr.mci_mortality)
mortality_data$prob_death_mild <- 1 - exp(-mortality_data$monthly_rate * hr.mild_mortality)
mortality_data$prob_death_moderate <- 1 - exp(-mortality_data$monthly_rate * hr.moderate_mortality)
mortality_data$prob_death_severe <- 1 - exp(-mortality_data$monthly_rate * hr.severe_mortality)

# Retain only relevant columns
mortality_rates <- mortality_data[, c("age", "prob_death_mci", "prob_death_mild", "prob_death_moderate", "prob_death_severe")]


# =============================
# Time-Dependent Transition Probabilities
# =============================

# Number of model cycles (e.g., 40 years * 12 months = 480 months)
n_cycles <- 480

# ---- Standard Care ----
p.mci_to_mild_std <- rep(p.mci_to_mild, n_cycles)
p.mci_stay_std <- 1 - p.mci_to_mild_std

p.mild_to_moderate_std <- rep(p.mild_to_moderate, n_cycles)
p.mild_stay_std <- 1 - p.mild_to_moderate_std

p.moderate_to_severe_std <- rep(p.moderate_to_severe, n_cycles)
p.moderate_stay_std <- 1 - p.moderate_to_severe_std

p.severe_stay_std <- rep(1, n_cycles)  # No forward progression beyond severe in model

# ---- Aducanumab ----
rate.mci_to_mild_adu <- -log(1 - p.mci_to_mild) * hr.aducanumab
rate.mild_to_moderate_adu <- -log(1 - p.mild_to_moderate) * hr.aducanumab
rate.moderate_to_severe_adu <- -log(1 - p.moderate_to_severe) * hr.aducanumab

p.mci_to_mild_adu <- rep(1 - exp(-rate.mci_to_mild_adu), n_cycles)
p.mci_stay_adu <- 1 - p.mci_to_mild_adu

p.mild_to_moderate_adu <- rep(1 - exp(-rate.mild_to_moderate_adu), n_cycles)
p.mild_stay_adu <- 1 - p.mild_to_moderate_adu

p.moderate_to_severe_adu <- rep(1 - exp(-rate.moderate_to_severe_adu), n_cycles)
p.moderate_stay_adu <- 1 - p.moderate_to_severe_adu

p.severe_stay_adu <- rep(1, n_cycles)

# ---- Donanemab ----
rate.mci_to_mild_dona <- -log(1 - p.mci_to_mild) * hr.donanemab
rate.mild_to_moderate_dona <- -log(1 - p.mild_to_moderate) * hr.donanemab
rate.moderate_to_severe_dona <- -log(1 - p.moderate_to_severe) * hr.donanemab

p.mci_to_mild_dona <- rep(1 - exp(-rate.mci_to_mild_dona), n_cycles)
p.mci_stay_dona <- 1 - p.mci_to_mild_dona

p.mild_to_moderate_dona <- rep(1 - exp(-rate.mild_to_moderate_dona), n_cycles)
p.mild_stay_dona <- 1 - p.mild_to_moderate_dona

p.moderate_to_severe_dona <- rep(1 - exp(-rate.moderate_to_severe_dona), n_cycles)
p.moderate_stay_dona <- 1 - p.moderate_to_severe_dona

p.severe_stay_dona <- rep(1, n_cycles)


# =============================
# Transition Probability Matrix Builder
# =============================

build_transition_matrix <- function(strat) {
  # Initialize transition array: [from_state, to_state, cycle]
  trans_mat <- array(0, dim = c(length(state_names), length(state_names), n_cycles),
                     dimnames = list(from = state_names, to = state_names, cycle = NULL))
  
  for (t in 1:n_cycles) {
    # Current age (starting from 75.2, 1 year per 12 cycles)
    current_age <- floor(initial_age + (t - 1) / 12)
    current_age <- min(max(current_age, 60), 99)  # Ensure age within life table range
    
    # Get age-specific death probabilities
    p_death_mci <- mortality_rates$prob_death_mci[mortality_rates$age == current_age]
    p_death_mild <- mortality_rates$prob_death_mild[mortality_rates$age == current_age]
    p_death_moderate <- mortality_rates$prob_death_moderate[mortality_rates$age == current_age]
    p_death_severe <- mortality_rates$prob_death_severe[mortality_rates$age == current_age]
    
    # Set transitions to death (from each state)
    trans_mat["mci", "death", t] <- p_death_mci
    trans_mat["mild", "death", t] <- p_death_mild
    trans_mat["moderate", "death", t] <- p_death_moderate
    trans_mat["severe", "death", t] <- p_death_severe
    
    # Set death as absorbing state
    trans_mat["death", "death", t] <- 1
    
    # ---- Transitions from MCI ----
    if (strat == "no intervention") {
      trans_mat["mci", "mild", t] <- p.mci_to_mild_std[t] * (1 - p_death_mci)
      trans_mat["mci", "mci", t] <- p.mci_stay_std[t] * (1 - p_death_mci)
    } else if (strat == "aducanumab") {
      trans_mat["mci", "mild", t] <- p.mci_to_mild_adu[t] * (1 - p_death_mci)
      trans_mat["mci", "mci", t] <- p.mci_stay_adu[t] * (1 - p_death_mci)
    } else if (strat == "donanemab") {
      trans_mat["mci", "mild", t] <- p.mci_to_mild_dona[t] * (1 - p_death_mci)
      trans_mat["mci", "mci", t] <- p.mci_stay_dona[t] * (1 - p_death_mci)
    }
    
    # ---- Transitions from Mild ----
    if (strat == "no intervention") {
      trans_mat["mild", "moderate", t] <- p.mild_to_moderate_std[t] * (1 - p_death_mild)
      trans_mat["mild", "mild", t] <- p.mild_stay_std[t] * (1 - p_death_mild)
    } else if (strat == "aducanumab") {
      trans_mat["mild", "moderate", t] <- p.mild_to_moderate_adu[t] * (1 - p_death_mild)
      trans_mat["mild", "mild", t] <- p.mild_stay_adu[t] * (1 - p_death_mild)
    } else if (strat == "donanemab") {
      trans_mat["mild", "moderate", t] <- p.mild_to_moderate_dona[t] * (1 - p_death_mild)
      trans_mat["mild", "mild", t] <- p.mild_stay_dona[t] * (1 - p_death_mild)
    }
    
    # ---- Transitions from Moderate ----
    if (strat == "no intervention") {
      trans_mat["moderate", "severe", t] <- p.moderate_to_severe_std[t] * (1 - p_death_moderate)
      trans_mat["moderate", "moderate", t] <- p.moderate_stay_std[t] * (1 - p_death_moderate)
    } else if (strat == "aducanumab") {
      trans_mat["moderate", "severe", t] <- p.moderate_to_severe_adu[t] * (1 - p_death_moderate)
      trans_mat["moderate", "moderate", t] <- p.moderate_stay_adu[t] * (1 - p_death_moderate)
    } else if (strat == "donanemab") {
      trans_mat["moderate", "severe", t] <- p.moderate_to_severe_dona[t] * (1 - p_death_moderate)
      trans_mat["moderate", "moderate", t] <- p.moderate_stay_dona[t] * (1 - p_death_moderate)
    }
    
    # ---- Transitions from Severe ----
    if (strat == "no intervention") {
      trans_mat["severe", "severe", t] <- p.severe_stay_std[t] * (1 - p_death_severe)
    } else if (strat == "aducanumab") {
      trans_mat["severe", "severe", t] <- p.severe_stay_adu[t] * (1 - p_death_severe)
    } else if (strat == "donanemab") {
      trans_mat["severe", "severe", t] <- p.severe_stay_dona[t] * (1 - p_death_severe)
    }
    
    # Normalize each row to ensure row sums equal 1
    for (s in state_names) {
      row_sum <- sum(trans_mat[s, , t])
      if (row_sum > 0 && row_sum != 1) {
        trans_mat[s, , t] <- trans_mat[s, , t] / row_sum
      }
    }
  }
  
  return(trans_mat)
}


# =============================
# Run Markov Model and Compute Outcomes
# =============================

# Discount rate
discount_rate_annual <- 0.03
discount_rate_monthly <- (1 + discount_rate_annual)^(1 / 12) - 1
discount_weights <- 1 / ((1 + discount_rate_monthly) ^ (0:(n_cycles - 1)))

# Initialize results list
results <- list()

# Function to run model per strategy
run_markov_model <- function(strategy) {
  trans_mat <- build_transition_matrix(strategy)
  trace <- matrix(0, nrow = n_cycles + 1, ncol = length(state_names),
                  dimnames = list(NULL, state_names))
  trace[1, ] <- initial_state_dist * cohort_size
  
  qalys <- numeric(n_cycles)
  healthcare_costs <- numeric(n_cycles)
  societal_costs <- numeric(n_cycles)
  
  # Treatment tracking
  on_treatment <- rep(TRUE, n_cycles + 1)
  if (strategy == "aducanumab") {
    discontinuation_probs <- c(rep(38 / 12 / 100, 12), rep(0, n_cycles - 12))
    for (t in 2:(n_cycles + 1)) {
      on_treatment[t] <- on_treatment[t - 1] && runif(1) > discontinuation_probs[t - 1]
    }
  } else if (strategy == "donanemab") {
    on_treatment <- rep(TRUE, n_cycles + 1)
    on_treatment[7] <- FALSE  # 27% discontinue at month 6
    on_treatment[13] <- FALSE  # 55% discontinue at month 12
  }
  
  for (t in 1:n_cycles) {
    trace[t + 1, ] <- trace[t, ] %*% trans_mat[ , , t]
    
    # Utilities
    state_utils <- c(u.mci, u.mild, u.moderate, u.severe, 0)
    qalys[t] <- sum(trace[t, ] * state_utils)
    
    # Healthcare costs
    c_healthcare <- c(c.mci.healthcare, c.mild.healthcare, c.moderate.healthcare, c.severe.healthcare, 0)
    c_societal_extra <- c(c.mci.societal, c.mild.societal, c.moderate.societal, c.severe.societal, 0)
    
    # Drug costs
    drug_cost <- 0
    inf_mon_cost <- 0
    if (strategy == "aducanumab" && on_treatment[t]) {
      drug_cost <- ifelse(t <= 12, c.aducanumab.year1 / 12, c.aducanumab.annual / 12)
      inf_mon_cost <- ifelse(t <= 6, c.aducanumab.months1_6,
                             ifelse(t <= 12, c.aducanumab.months7_12, c.aducanumab.months13plus))
    } else if (strategy == "donanemab" && on_treatment[t]) {
      drug_cost <- c.donanemab.annual / 12
      inf_mon_cost <- ifelse(t <= 6, c.donanemab.months1_6,
                             ifelse(t <= 12, c.donanemab.months7_12, c.donanemab.months13plus))
    }
    
    n_treated <- sum(trace[t, c("mci", "mild", "moderate")])
    n_symptomatic_aria <- 0
    if (strategy == "aducanumab" && t <= 3) {
      n_symptomatic_aria <- n_treated * p.aria.aducanumab_symptomatic
    } else if (strategy == "donanemab" && t <= 3) {
      n_symptomatic_aria <- n_treated * p.aria.donanemab_symptomatic
    }
    
    # ARIA-related costs (for symptomatic cases)
    aria_cost <- n_symptomatic_aria * (c.aria.outpatient_visit +
                                         c.aria.followup_imaging +
                                         0.04 * c.aria.hospitalization)
    
    # Subtotal costs
    total_healthcare <- sum(trace[t, ] * c_healthcare) + n_treated * (drug_cost + inf_mon_cost) + aria_cost
    total_societal <- total_healthcare + sum(trace[t, ] * c_societal_extra)
    
    healthcare_costs[t] <- total_healthcare
    societal_costs[t] <- total_societal
  }
  
  # Apply screening cost at start
  screening_cost <- if (strategy == "aducanumab") c.screening.aducanumab else if (strategy == "donanemab") c.screening.donanemab else 0
  healthcare_costs[1] <- healthcare_costs[1] + screening_cost * cohort_size
  societal_costs[1] <- societal_costs[1] + screening_cost * cohort_size
  
  # Discounting
  total_qalys <- sum(qalys * discount_weights) / cohort_size
  total_healthcare <- sum(healthcare_costs * discount_weights) / cohort_size
  total_societal <- sum(societal_costs * discount_weights) / cohort_size
  
  return(list(qalys = total_qalys,
              healthcare_costs = total_healthcare,
              societal_costs = total_societal))
}

# Run model for all strategies
results$no_intervention <- run_markov_model("no intervention")
results$aducanumab <- run_markov_model("aducanumab")
results$donanemab <- run_markov_model("donanemab")
```
