## Supplementary material for "Replicating Health-Economic Simulation Models for Alzheimer’s Disease Using Artificial Intelligence": final R scripts: Ross_O13.R

```
# Health Economic Markov Model Inputs from Ross et al. (2022)

# Set state and strategy names
state_names <- c("mci", "mild", "moderate", "severe", "death")
strategy_names <- c("standard_care", "aducanumab", "donanemab")

# Simulation parameters
initial_age <- 75.2
cohort_size <- 1000
discount_rate <- 0.03

# Initial distribution
initial_dist <- c(mci = 0.65, mild = 0.35, moderate = 0, severe = 0, death = 0)

# Natural progression probabilities (monthly)
p.mci_to_mild <- 0.007
p.mild_to_moderate <- 0.016
p.moderate_to_severe <- 0.026

# Disease progression hazard ratios (HRs)
hr_progression_aducanumab <- 0.89
hr_progression_donanemab <- 0.68

# Mortality hazard ratios by state
hr_mortality_mci <- 1.61
hr_mortality_mild <- 2.23
hr_mortality_moderate <- 3.10
hr_mortality_severe <- 4.98

# Utilities (converted to monthly by dividing by 12)
u_mci <- 0.73 / 12
u_mild <- 0.69 / 12
u_moderate <- 0.53 / 12
u_severe <- 0.34 / 12
u_aria_decrement <- -0.065 / 12  # ARIA utility decrement (monthly impact assumed for 3 months)

# Monthly healthcare sector costs by disease state (USD)
cost_hc_mci <- 691
cost_hc_mild <- 1049
cost_hc_moderate <- 1274
cost_hc_severe <- 3764

# Monthly additional societal costs by disease state (USD)
cost_soc_mci <- 49
cost_soc_mild <- 1483
cost_soc_moderate <- 1932
cost_soc_severe <- 2153

# Annual drug costs (USD)
cost_drug_annual_aducanumab_yr1 <- 20500
cost_drug_annual_aducanumab_later <- 28200
cost_drug_annual_donanemab <- 28200

# Monthly infusion and monitoring costs (USD)
# Aducanumab
cost_infusion_monitor_aducanumab_1_6 <- 112
cost_infusion_monitor_aducanumab_7_12 <- 87
cost_infusion_monitor_aducanumab_13_plus <- 80
# Donanemab
cost_infusion_monitor_donanemab_1_6 <- 599
cost_infusion_monitor_donanemab_7_12 <- 302
cost_infusion_monitor_donanemab_13_plus <- 287

# Screening costs (one-time per eligible patient)
screening_cost_aducanumab <- 6957
screening_cost_donanemab <- 17096

# ARIA-related costs
aria_followup_imaging_cost <- 1060
aria_hospitalization_cost <- 35934
aria_outpatient_visit_cost <- 128

# ARIA event probabilities
p_aria_aducanumab <- 0.41
p_aria_donanemab <- 0.39


# Load age-specific mortality rates and compute disease-state-specific death probabilities

# Load CSV file
mortality_data <- read.csv("mortalityRates_US_2017.csv")

# Compute death probabilities for each disease state
mortality_data$prob_death_mci <- 1 - exp(-mortality_data$monthly_rate * hr_mortality_mci)
mortality_data$prob_death_mild <- 1 - exp(-mortality_data$monthly_rate * hr_mortality_mild)
mortality_data$prob_death_moderate <- 1 - exp(-mortality_data$monthly_rate * hr_mortality_moderate)
mortality_data$prob_death_severe <- 1 - exp(-mortality_data$monthly_rate * hr_mortality_severe)


# Define number of model cycles
n_cycles <- 480  # 40 years * 12 months

# --- Standard care (no intervention) ---
p_mci_to_mild_std <- rep(p.mci_to_mild, n_cycles)
p_mci_stay_std <- rep(1 - p.mci_to_mild, n_cycles)

p_mild_to_moderate_std <- rep(p.mild_to_moderate, n_cycles)
p_mild_stay_std <- rep(1 - p.mild_to_moderate, n_cycles)

p_moderate_to_severe_std <- rep(p.moderate_to_severe, n_cycles)
p_moderate_stay_std <- rep(1 - p.moderate_to_severe, n_cycles)

p_severe_stay_std <- rep(1, n_cycles)

# --- Aducanumab intervention ---
rate_mci_to_mild <- -log(1 - p.mci_to_mild)
rate_mild_to_moderate <- -log(1 - p.mild_to_moderate)
rate_moderate_to_severe <- -log(1 - p.moderate_to_severe)

adj_rate_mci_to_mild_adu <- rate_mci_to_mild * hr_progression_aducanumab
adj_rate_mild_to_moderate_adu <- rate_mild_to_moderate * hr_progression_aducanumab
adj_rate_moderate_to_severe_adu <- rate_moderate_to_severe * hr_progression_aducanumab

p_mci_to_mild_adu <- rep(1 - exp(-adj_rate_mci_to_mild_adu), n_cycles)
p_mci_stay_adu <- rep(1 - p_mci_to_mild_adu[1], n_cycles)

p_mild_to_moderate_adu <- rep(1 - exp(-adj_rate_mild_to_moderate_adu), n_cycles)
p_mild_stay_adu <- rep(1 - p_mild_to_moderate_adu[1], n_cycles)

p_moderate_to_severe_adu <- rep(1 - exp(-adj_rate_moderate_to_severe_adu), n_cycles)
p_moderate_stay_adu <- rep(1 - p_moderate_to_severe_adu[1], n_cycles)

p_severe_stay_adu <- rep(1, n_cycles)

# --- Donanemab intervention ---
adj_rate_mci_to_mild_dona <- rate_mci_to_mild * hr_progression_donanemab
adj_rate_mild_to_moderate_dona <- rate_mild_to_moderate * hr_progression_donanemab
adj_rate_moderate_to_severe_dona <- rate_moderate_to_severe * hr_progression_donanemab

p_mci_to_mild_dona <- rep(1 - exp(-adj_rate_mci_to_mild_dona), n_cycles)
p_mci_stay_dona <- rep(1 - p_mci_to_mild_dona[1], n_cycles)

p_mild_to_moderate_dona <- rep(1 - exp(-adj_rate_mild_to_moderate_dona), n_cycles)
p_mild_stay_dona <- rep(1 - p_mild_to_moderate_dona[1], n_cycles)

p_moderate_to_severe_dona <- rep(1 - exp(-adj_rate_moderate_to_severe_dona), n_cycles)
p_moderate_stay_dona <- rep(1 - p_moderate_to_severe_dona[1], n_cycles)

p_severe_stay_dona <- rep(1, n_cycles)


# Function to build the transition probability matrix for a given strategy
build_transition_matrix <- function(strat) {
  
  n_states <- length(state_names)
  trans_mat <- array(0, dim = c(n_states, n_states, n_cycles),
                     dimnames = list(state_names, state_names, NULL))
  
  for (t in 1:n_cycles) {
    
    # Current age (assuming 1 cycle = 1 month)
    age_t <- floor(initial_age + (t - 1) / 12)
    if (age_t > 99) age_t <- 99 
    
    # Find index for current age in mortality data
    idx <- which(mortality_data$age == age_t)
    
    # Assign age- and state-specific mortality probabilities
    trans_mat["mci", "death", t]      <- mortality_data$prob_death_mci[idx]
    trans_mat["mild", "death", t]     <- mortality_data$prob_death_mild[idx]
    trans_mat["moderate", "death", t] <- mortality_data$prob_death_moderate[idx]
    trans_mat["severe", "death", t]   <- mortality_data$prob_death_severe[idx]
    trans_mat["death", "death", t]    <- 1  # Absorbing state
    
    # [INSERT PROMPT 4B HERE] — define transitions to next disease stages (with or without treatment)
    
    if (strat == "no intervention") {
      trans_mat["mci", "mild", t]      <- p_mci_to_mild_std[t] * (1 - trans_mat["mci", "death", t])
      trans_mat["mci", "mci", t]       <- p_mci_stay_std[t] * (1 - trans_mat["mci", "death", t])
      
      trans_mat["mild", "moderate", t] <- p_mild_to_moderate_std[t] * (1 - trans_mat["mild", "death", t])
      trans_mat["mild", "mild", t]     <- p_mild_stay_std[t] * (1 - trans_mat["mild", "death", t])
      
      trans_mat["moderate", "severe", t] <- p_moderate_to_severe_std[t] * (1 - trans_mat["moderate", "death", t])
      trans_mat["moderate", "moderate", t] <- p_moderate_stay_std[t] * (1 - trans_mat["moderate", "death", t])
      
      trans_mat["severe", "severe", t] <- p_severe_stay_std[t] * (1 - trans_mat["severe", "death", t])
      
    } else if (strat == "aducanumab") {
      trans_mat["mci", "mild", t]      <- p_mci_to_mild_adu[t] * (1 - trans_mat["mci", "death", t])
      trans_mat["mci", "mci", t]       <- p_mci_stay_adu[t] * (1 - trans_mat["mci", "death", t])
      
      trans_mat["mild", "moderate", t] <- p_mild_to_moderate_adu[t] * (1 - trans_mat["mild", "death", t])
      trans_mat["mild", "mild", t]     <- p_mild_stay_adu[t] * (1 - trans_mat["mild", "death", t])
      
      trans_mat["moderate", "severe", t] <- p_moderate_to_severe_adu[t] * (1 - trans_mat["moderate", "death", t])
      trans_mat["moderate", "moderate", t] <- p_moderate_stay_adu[t] * (1 - trans_mat["moderate", "death", t])
      
      trans_mat["severe", "severe", t] <- p_severe_stay_adu[t] * (1 - trans_mat["severe", "death", t])
      
    } else if (strat == "donanemab") {
      trans_mat["mci", "mild", t]      <- p_mci_to_mild_dona[t] * (1 - trans_mat["mci", "death", t])
      trans_mat["mci", "mci", t]       <- p_mci_stay_dona[t] * (1 - trans_mat["mci", "death", t])
      
      trans_mat["mild", "moderate", t] <- p_mild_to_moderate_dona[t] * (1 - trans_mat["mild", "death", t])
      trans_mat["mild", "mild", t]     <- p_mild_stay_dona[t] * (1 - trans_mat["mild", "death", t])
      
      trans_mat["moderate", "severe", t] <- p_moderate_to_severe_dona[t] * (1 - trans_mat["moderate", "death", t])
      trans_mat["moderate", "moderate", t] <- p_moderate_stay_dona[t] * (1 - trans_mat["moderate", "death", t])
      
      trans_mat["severe", "severe", t] <- p_severe_stay_dona[t] * (1 - trans_mat["severe", "death", t])
    }
    
    # Normalize each row to sum to 1 (if not already)
    for (state in state_names) {
      row_sum <- sum(trans_mat[state, , t])
      if (row_sum > 0 && abs(row_sum - 1) > 1e-8) {
        trans_mat[state, , t] <- trans_mat[state, , t] / row_sum
      }
    }
  }
  
  return(trans_mat)
}


# Discount factor per month
disc_month <- (1 / (1 + discount_rate))^(0:(n_cycles - 1) / 12)

# Initialize result container
results <- list()

# Run model for each strategy
for (strategy in c("no intervention", "aducanumab", "donanemab")) {
  
  # Transition matrix
  tp_matrix <- build_transition_matrix(strategy)
  
  # Markov trace matrix [cycles x states]
  trace <- matrix(0, nrow = n_cycles + 1, ncol = length(state_names),
                  dimnames = list(NULL, state_names))
  trace[1, ] <- cohort_size * initial_dist
  
  # Treatment tracking (who remains on treatment)
  on_treatment <- rep(1, n_cycles + 1)
  if (strategy == "aducanumab") {
    # Linear discontinuation: 38% over 12 months
    discont_rate <- seq(0, 0.38, length.out = 12)
    on_treatment[2:13] <- 1 - discont_rate
    on_treatment[14:(n_cycles + 1)] <- 0.62
  } else if (strategy == "donanemab") {
    # Discontinuation: 27% at 6 months, 55% at 12 months
    on_treatment[2:7] <- 1
    on_treatment[8:13] <- 0.73
    on_treatment[14:(n_cycles + 1)] <- 0.45
  }
  
  # Initialize outcome accumulators
  qalys <- rep(0, n_cycles)
  hc_costs <- rep(0, n_cycles)
  soc_costs <- rep(0, n_cycles)
  
  for (t in 1:n_cycles) {
    
    # Update trace
    trace[t + 1, ] <- trace[t, ] %*% tp_matrix[, , t]
    
    # Apply utilities
    qalys[t] <- (
      trace[t, "mci"] * u_mci +
        trace[t, "mild"] * u_mild +
        trace[t, "moderate"] * u_moderate +
        trace[t, "severe"] * u_severe
    )
    
    # ARIA symptomatic effect (apply only in cycles 1-3, and to treated patients)
    if (strategy != "no intervention" && t <= 3) {
      if (strategy == "aducanumab") {
        aria_sympt_rate <- p_aria_aducanumab * 0.24
      } else {
        aria_sympt_rate <- p_aria_donanemab * 0.16
      }
      qalys[t] <- qalys[t] + trace[t, "mci"] * on_treatment[t] * aria_sympt_rate * u_aria_decrement
    }
    
    # Health sector costs
    hc_costs[t] <- (
      trace[t, "mci"] * cost_hc_mci +
        trace[t, "mild"] * cost_hc_mild +
        trace[t, "moderate"] * cost_hc_moderate +
        trace[t, "severe"] * cost_hc_severe
    )
    
    # Societal costs
    soc_costs[t] <- hc_costs[t] + (
      trace[t, "mci"] * cost_soc_mci +
        trace[t, "mild"] * cost_soc_mild +
        trace[t, "moderate"] * cost_soc_moderate +
        trace[t, "severe"] * cost_soc_severe
    )
    
    # Drug costs (only if not in severe, and still on treatment)
    if (strategy == "aducanumab" && trace[t, "severe"] < cohort_size) {
      if (t <= 12) {
        drug_cost <- cost_drug_annual_aducanumab_yr1 / 12
      } else {
        drug_cost <- cost_drug_annual_aducanumab_later / 12
      }
      patients_on_drug <- sum(trace[t, c("mci", "mild", "moderate")]) * on_treatment[t]
      hc_costs[t] <- hc_costs[t] + patients_on_drug * drug_cost
      soc_costs[t] <- soc_costs[t] + patients_on_drug * drug_cost
    } else if (strategy == "donanemab" && trace[t, "severe"] < cohort_size) {
      drug_cost <- cost_drug_annual_donanemab / 12
      patients_on_drug <- sum(trace[t, c("mci", "mild", "moderate")]) * on_treatment[t]
      hc_costs[t] <- hc_costs[t] + patients_on_drug * drug_cost
      soc_costs[t] <- soc_costs[t] + patients_on_drug * drug_cost
    }
    
    # Infusions and monitoring
    if (strategy == "aducanumab") {
      if (t <= 6) {
        infuse_cost <- cost_infusion_monitor_aducanumab_1_6
      } else if (t <= 12) {
        infuse_cost <- cost_infusion_monitor_aducanumab_7_12
      } else {
        infuse_cost <- cost_infusion_monitor_aducanumab_13_plus
      }
      hc_costs[t] <- hc_costs[t] + sum(trace[t, c("mci", "mild", "moderate")]) * on_treatment[t] * infuse_cost
      soc_costs[t] <- soc_costs[t] + sum(trace[t, c("mci", "mild", "moderate")]) * on_treatment[t] * infuse_cost
    } else if (strategy == "donanemab") {
      if (t <= 6) {
        infuse_cost <- cost_infusion_monitor_donanemab_1_6
      } else if (t <= 12) {
        infuse_cost <- cost_infusion_monitor_donanemab_7_12
      } else {
        infuse_cost <- cost_infusion_monitor_donanemab_13_plus
      }
      hc_costs[t] <- hc_costs[t] + sum(trace[t, c("mci", "mild", "moderate")]) * infuse_cost
      soc_costs[t] <- soc_costs[t] + sum(trace[t, c("mci", "mild", "moderate")]) * infuse_cost
    }
    
    # ARIA-related costs (apply to symptomatic cases only in cycles 1-3)
    if (strategy != "no intervention" && t <= 3) {
      if (strategy == "aducanumab") {
        p_aria_sympt <- p_aria_aducanumab * 0.24
      } else {
        p_aria_sympt <- p_aria_donanemab * 0.16
      }
      aria_cases <- trace[t, "mci"] * on_treatment[t] * p_aria_sympt
      aria_cost <- aria_cases * (
        aria_outpatient_visit_cost + aria_followup_imaging_cost +
          (0.04 * aria_hospitalization_cost)
      )
      hc_costs[t] <- hc_costs[t] + aria_cost
      soc_costs[t] <- soc_costs[t] + aria_cost
    }
  }
  
  # Apply discounting and compute totals per person
  total_qalys <- sum(qalys * disc_month) / cohort_size
  total_hc_costs <- sum(hc_costs * disc_month) / cohort_size
  total_soc_costs <- sum(soc_costs * disc_month) / cohort_size
  
  # Add one-time screening costs for treated groups
  if (strategy == "aducanumab") {
    total_hc_costs <- total_hc_costs + screening_cost_aducanumab / cohort_size
    total_soc_costs <- total_soc_costs + screening_cost_aducanumab / cohort_size
  } else if (strategy == "donanemab") {
    total_hc_costs <- total_hc_costs + screening_cost_donanemab / cohort_size
    total_soc_costs <- total_soc_costs + screening_cost_donanemab / cohort_size
  }
  
  # Save results
  strat_name <- gsub(" ", "_", strategy)
  results[[strat_name]] <- list(
    qalys = total_qalys,
    healthcare_costs = total_hc_costs,
    societal_costs = total_soc_costs
  )
}
```
