## Supplementary material for "Replicating Health-Economic Simulation Models for Alzheimer’s Disease Using Artificial Intelligence": final R scripts: Ross_O14.R

```
# ======================
# INPUT PARAMETERS: Ross et al. (2022) Model
# ======================

# State names
states <- c("mci", "mild", "moderate", "severe", "death")

# Strategy names
strategies <- c("standard_care", "aducanumab", "donanemab")

# Starting cohort
cohort_size <- 1000
starting_age <- 75.2

# Transition probabilities (monthly progression)
p.mci_to_mild <- 0.007
p.mild_to_moderate <- 0.016
p.moderate_to_severe <- 0.026

# Disease progression hazard ratios
hr_prog_aducanumab <- 0.89
hr_prog_donanemab <- 0.68

# Mortality hazard ratios by state
hr_mortality_mci <- 1.61
hr_mortality_mild <- 2.23
hr_mortality_moderate <- 3.10
hr_mortality_severe <- 4.98

# Utilities (converted to monthly by dividing by 12)
u.mci <- 0.73 / 12
u.mild <- 0.69 / 12
u.moderate <- 0.53 / 12
u.severe <- 0.34 / 12

# ARIA-related utility decrement (3-month impact modeled separately)
u_aria_decrement <- -0.065 / 12

# Healthcare sector monthly costs (2020 USD)
costs_healthcare_mci <- 691
costs_healthcare_mild <- 1049
costs_healthcare_moderate <- 1274
costs_healthcare_severe <- 3764

# Societal additional monthly costs (2020 USD)
costs_societal_mci <- 49
costs_societal_mild <- 1483
costs_societal_moderate <- 1932
costs_societal_severe <- 2153

# Annual drug costs
cost_aducanumab_year1 <- 20500
cost_aducanumab_annual <- 28200
cost_donanemab_annual <- 28200

# Monthly infusion and monitoring costs
cost_infusion_aducanumab_1_6 <- 112
cost_infusion_aducanumab_7_12 <- 87
cost_infusion_aducanumab_13plus <- 80
cost_infusion_donanemab_1_6 <- 599
cost_infusion_donanemab_7_12 <- 302
cost_infusion_donanemab_13plus <- 287

# Screening costs per eligible patient
cost_screening_aducanumab <- 6957
cost_screening_donanemab <- 17096

# ARIA-related costs
cost_aria_followup_imaging <- 1060
cost_aria_hospitalization <- 35934
cost_aria_outpatient <- 128

# ARIA event probabilities (monthly modeling required later)
prob_aria_aducanumab <- 0.41
prob_aria_donanemab <- 0.39
prob_symptomatic_aria_aducanumab <- 0.41 * 0.24
prob_symptomatic_aria_donanemab <- 0.39 * 0.16


# ======================
# LOAD AND PROCESS MORTALITY DATA
# ======================

# Load mortality rate CSV
mortality_data <- read.csv("mortalityRates_US_2017.csv")

# Compute age-specific death probabilities by severity
mortality_data$prob_death_mci <- 1 - exp(-mortality_data$monthly_rate * hr_mortality_mci)
mortality_data$prob_death_mild <- 1 - exp(-mortality_data$monthly_rate * hr_mortality_mild)
mortality_data$prob_death_moderate <- 1 - exp(-mortality_data$monthly_rate * hr_mortality_moderate)
mortality_data$prob_death_severe <- 1 - exp(-mortality_data$monthly_rate * hr_mortality_severe)

# Final output data frame
mortality_by_state <- mortality_data[, c("age", "prob_death_mci", "prob_death_mild", "prob_death_moderate", "prob_death_severe")]


# ======================
# TRANSITION PROBABILITY VECTORS (TIME-DEPENDENT)
# ======================

# Define number of cycles
n_cycles <- 480  # 40 years x 12 months

# ---- No Intervention (Standard Care) ----
p_mci_to_mild_std <- rep(p.mci_to_mild, n_cycles)
p_mild_to_moderate_std <- rep(p.mild_to_moderate, n_cycles)
p_moderate_to_severe_std <- rep(p.moderate_to_severe, n_cycles)

p_mci_stay_std <- rep(1 - p.mci_to_mild, n_cycles)
p_mild_stay_std <- rep(1 - p.mild_to_moderate, n_cycles)
p_moderate_stay_std <- rep(1 - p.moderate_to_severe, n_cycles)
p_severe_stay_std <- rep(1, n_cycles)

# ---- Aducanumab ----
rate_mci_to_mild_adu <- -log(1 - p.mci_to_mild) * hr_prog_aducanumab
rate_mild_to_moderate_adu <- -log(1 - p.mild_to_moderate) * hr_prog_aducanumab
rate_moderate_to_severe_adu <- -log(1 - p.moderate_to_severe) * hr_prog_aducanumab

p_mci_to_mild_adu <- rep(1 - exp(-rate_mci_to_mild_adu), n_cycles)
p_mild_to_moderate_adu <- rep(1 - exp(-rate_mild_to_moderate_adu), n_cycles)
p_moderate_to_severe_adu <- rep(1 - exp(-rate_moderate_to_severe_adu), n_cycles)

p_mci_stay_adu <- rep(1 - p_mci_to_mild_adu[1], n_cycles)
p_mild_stay_adu <- rep(1 - p_mild_to_moderate_adu[1], n_cycles)
p_moderate_stay_adu <- rep(1 - p_moderate_to_severe_adu[1], n_cycles)
p_severe_stay_adu <- rep(1, n_cycles)

# ---- Donanemab ----
rate_mci_to_mild_dona <- -log(1 - p.mci_to_mild) * hr_prog_donanemab
rate_mild_to_moderate_dona <- -log(1 - p.mild_to_moderate) * hr_prog_donanemab
rate_moderate_to_severe_dona <- -log(1 - p.moderate_to_severe) * hr_prog_donanemab

p_mci_to_mild_dona <- rep(1 - exp(-rate_mci_to_mild_dona), n_cycles)
p_mild_to_moderate_dona <- rep(1 - exp(-rate_mild_to_moderate_dona), n_cycles)
p_moderate_to_severe_dona <- rep(1 - exp(-rate_moderate_to_severe_dona), n_cycles)

p_mci_stay_dona <- rep(1 - p_mci_to_mild_dona[1], n_cycles)
p_mild_stay_dona <- rep(1 - p_mild_to_moderate_dona[1], n_cycles)
p_moderate_stay_dona <- rep(1 - p_moderate_to_severe_dona[1], n_cycles)
p_severe_stay_dona <- rep(1, n_cycles)


# ======================
# TRANSITION PROBABILITY MATRIX FUNCTION (STRUCTURE ONLY)
# ======================

build_transition_matrix <- function(strat) {
  
  # Number of states and cycles
  n_states <- length(states)
  trans_mat <- array(0, dim = c(n_states, n_states, n_cycles),
                     dimnames = list(from = states, to = states, cycle = 1:n_cycles))
  
  # Loop through each cycle to fill transition probabilities
  for (t in 1:n_cycles) {
    
    # Determine current age (assuming age increases every 12 cycles)
    current_age <- min(99, floor(starting_age + (t - 1) / 12))
    age_row <- mortality_by_state[mortality_by_state$age == current_age, ]
    
    # Assign probabilities of death from each health state
    trans_mat["mci", "death", t] <- age_row$prob_death_mci
    trans_mat["mild", "death", t] <- age_row$prob_death_mild
    trans_mat["moderate", "death", t] <- age_row$prob_death_moderate
    trans_mat["severe", "death", t] <- age_row$prob_death_severe
    trans_mat["death", "death", t] <- 1  # Absorbing state
    
    if (strat == "no intervention") {
      trans_mat["mci", "mild", t] <- p_mci_to_mild_std[t] * (1 - trans_mat["mci", "death", t])
      trans_mat["mci", "mci", t] <- p_mci_stay_std[t] * (1 - trans_mat["mci", "death", t])
      
      trans_mat["mild", "moderate", t] <- p_mild_to_moderate_std[t] * (1 - trans_mat["mild", "death", t])
      trans_mat["mild", "mild", t] <- p_mild_stay_std[t] * (1 - trans_mat["mild", "death", t])
      
      trans_mat["moderate", "severe", t] <- p_moderate_to_severe_std[t] * (1 - trans_mat["moderate", "death", t])
      trans_mat["moderate", "moderate", t] <- p_moderate_stay_std[t] * (1 - trans_mat["moderate", "death", t])
      
      trans_mat["severe", "severe", t] <- p_severe_stay_std[t] * (1 - trans_mat["severe", "death", t])
      
    } else if (strat == "aducanumab") {
      trans_mat["mci", "mild", t] <- p_mci_to_mild_adu[t] * (1 - trans_mat["mci", "death", t])
      trans_mat["mci", "mci", t] <- p_mci_stay_adu[t] * (1 - trans_mat["mci", "death", t])
      
      trans_mat["mild", "moderate", t] <- p_mild_to_moderate_adu[t] * (1 - trans_mat["mild", "death", t])
      trans_mat["mild", "mild", t] <- p_mild_stay_adu[t] * (1 - trans_mat["mild", "death", t])
      
      trans_mat["moderate", "severe", t] <- p_moderate_to_severe_adu[t] * (1 - trans_mat["moderate", "death", t])
      trans_mat["moderate", "moderate", t] <- p_moderate_stay_adu[t] * (1 - trans_mat["moderate", "death", t])
      
      trans_mat["severe", "severe", t] <- p_severe_stay_adu[t] * (1 - trans_mat["severe", "death", t])
      
    } else if (strat == "donanemab") {
      trans_mat["mci", "mild", t] <- p_mci_to_mild_dona[t] * (1 - trans_mat["mci", "death", t])
      trans_mat["mci", "mci", t] <- p_mci_stay_dona[t] * (1 - trans_mat["mci", "death", t])
      
      trans_mat["mild", "moderate", t] <- p_mild_to_moderate_dona[t] * (1 - trans_mat["mild", "death", t])
      trans_mat["mild", "mild", t] <- p_mild_stay_dona[t] * (1 - trans_mat["mild", "death", t])
      
      trans_mat["moderate", "severe", t] <- p_moderate_to_severe_dona[t] * (1 - trans_mat["moderate", "death", t])
      trans_mat["moderate", "moderate", t] <- p_moderate_stay_dona[t] * (1 - trans_mat["moderate", "death", t])
      
      trans_mat["severe", "severe", t] <- p_severe_stay_dona[t] * (1 - trans_mat["severe", "death", t])
    }
    
    # Normalize rows to ensure each row sums to 1
    for (state in states[1:4]) {  # exclude "death" since it's already set to 1
      row_sum <- sum(trans_mat[state, , t])
      if (abs(row_sum - 1) > 1e-8) {
        trans_mat[state, , t] <- trans_mat[state, , t] / row_sum
      }
    }
  }
  
  return(trans_mat)
}


# ======================
# MARKOV MODEL SIMULATION
# ======================

# Monthly discount rate from 3% annual
discount_rate <- (1 + 0.03)^(1/12) - 1
discount_factors <- 1 / (1 + discount_rate)^(0:(n_cycles - 1))

# Initial state distribution
init_dist <- numeric(length(states))
names(init_dist) <- states
init_dist["mci"] <- 0.65
init_dist["mild"] <- 0.35
init_dist["moderate"] <- 0
init_dist["severe"] <- 0
init_dist["death"] <- 0

# Treatment discontinuation probabilities
adu_treated <- rep(1, n_cycles)
adu_treated[1:12] <- 1 - seq(0, 0.38, length.out = 12)
adu_treated[13:n_cycles] <- 1 - 0.38

dona_treated <- rep(1, n_cycles)
dona_treated[1:6] <- 1 - 0.27
dona_treated[7:12] <- 1 - 0.55
dona_treated[13:n_cycles] <- 1 - 0.66

# Function to run model for one strategy
run_model <- function(strat) {
  
  trans_mat <- build_transition_matrix(strat)
  trace <- matrix(0, nrow = n_cycles + 1, ncol = length(states))
  colnames(trace) <- states
  trace[1, ] <- cohort_size * init_dist
  
  qalys <- numeric(n_cycles)
  cost_healthcare <- numeric(n_cycles)
  cost_societal <- numeric(n_cycles)
  
  for (t in 1:n_cycles) {
    trace[t + 1, ] <- trace[t, ] %*% trans_mat[, , t]
    
    current_state <- trace[t, ]
    
    # QALYs
    qaly_vec <- c(u.mci, u.mild, u.moderate, u.severe, 0)
    qaly_this_cycle <- sum(current_state * qaly_vec)
    
    # Apply ARIA QALY decrement in first 3 months
    if (t <= 3 && strat != "no intervention") {
      prob_sympt_aria <- if (strat == "aducanumab") prob_symptomatic_aria_aducanumab else prob_symptomatic_aria_donanemab
      qaly_this_cycle <- qaly_this_cycle + current_state["mci"] * prob_sympt_aria * u_aria_decrement
    }
    
    qalys[t] <- qaly_this_cycle
    
    # Base healthcare and societal costs
    cost_hc_vec <- c(costs_healthcare_mci, costs_healthcare_mild, costs_healthcare_moderate, costs_healthcare_severe, 0)
    cost_soc_vec <- c(costs_societal_mci, costs_societal_mild, costs_societal_moderate, costs_societal_severe, 0)
    cost_healthcare[t] <- sum(current_state * cost_hc_vec)
    cost_societal[t] <- sum(current_state * (cost_hc_vec + cost_soc_vec))
    
    # Add treatment costs
    if (strat == "aducanumab") {
      treat_pop <- current_state["mci"] + current_state["mild"] + current_state["moderate"]
      if (t <= 12) {
        drug_cost <- cost_aducanumab_year1 / 12
      } else {
        drug_cost <- cost_aducanumab_annual / 12
      }
      infusion_cost <- if (t <= 6) cost_infusion_aducanumab_1_6 else if (t <= 12) cost_infusion_aducanumab_7_12 else cost_infusion_aducanumab_13plus
      active_treatment <- treat_pop * adu_treated[t]
      cost_healthcare[t] <- cost_healthcare[t] + active_treatment * (drug_cost + infusion_cost)
      cost_societal[t] <- cost_societal[t] + active_treatment * (drug_cost + infusion_cost)
    }
    
    if (strat == "donanemab") {
      treat_pop <- current_state["mci"] + current_state["mild"] + current_state["moderate"]
      drug_cost <- cost_donanemab_annual / 12
      infusion_cost <- if (t <= 6) cost_infusion_donanemab_1_6 else if (t <= 12) cost_infusion_donanemab_7_12 else cost_infusion_donanemab_13plus
      active_treatment <- treat_pop * dona_treated[t]
      cost_healthcare[t] <- cost_healthcare[t] + active_treatment * (drug_cost + infusion_cost)
      cost_societal[t] <- cost_societal[t] + active_treatment * (drug_cost + infusion_cost)
    }
    
    # ARIA costs (first 3 months)
    if (t <= 3 && strat != "no intervention") {
      prob_sympt_aria <- if (strat == "aducanumab") prob_symptomatic_aria_aducanumab else prob_symptomatic_aria_donanemab
      aria_cases <- current_state["mci"] * prob_sympt_aria
      cost_aria <- aria_cases * (cost_aria_outpatient + cost_aria_followup_imaging + 0.04 * cost_aria_hospitalization)
      cost_healthcare[t] <- cost_healthcare[t] + cost_aria
      cost_societal[t] <- cost_societal[t] + cost_aria
    }
  }
  
  # One-time screening cost in cycle 1
  if (strat == "aducanumab") {
    cost_healthcare[1] <- cost_healthcare[1] + cohort_size * cost_screening_aducanumab
    cost_societal[1] <- cost_societal[1] + cohort_size * cost_screening_aducanumab
  }
  if (strat == "donanemab") {
    cost_healthcare[1] <- cost_healthcare[1] + cohort_size * cost_screening_donanemab
    cost_societal[1] <- cost_societal[1] + cohort_size * cost_screening_donanemab
  }
  
  # Discount
  total_qalys <- sum(qalys * discount_factors)
  total_hc_costs <- sum(cost_healthcare * discount_factors)
  total_soc_costs <- sum(cost_societal * discount_factors)
  
  # Return per-person results
  list(
    qalys = total_qalys / cohort_size,
    healthcare_costs = total_hc_costs / cohort_size,
    societal_costs = total_soc_costs / cohort_size
  )
}

# Run for all strategies
results <- list()
results$no_intervention <- run_model("no intervention")
results$aducanumab <- run_model("aducanumab")
results$donanemab <- run_model("donanemab")
```
