## Supplementary material for "Replicating Health-Economic Simulation Models for Alzheimer’s Disease Using Artificial Intelligence": final R scripts: Ross_O15.R

```
# Prompt 1: Define all input parameters for the Markov model from Ross et al. (2022)

# --- Descriptive Parameters ---
states <- c("mci", "mild", "moderate", "severe", "death")
strategies <- c("standard_care", "aducanumab", "donanemab")
cohort_size <- 1000
initial_age <- 75.2

# --- Initial Distribution ---
initial_state_dist <- c(mci = 0.65, mild = 0.35, moderate = 0, severe = 0, death = 0)

# --- Disease Progression Probabilities (monthly) ---
p.mci_to_mild <- 0.007
p.mild_to_moderate <- 0.016
p.moderate_to_severe <- 0.026
# No backward or same-stage transitions included as per model

# --- Treatment Effects (Hazard Ratios) ---
hr_progression_aducanumab <- 0.89
hr_progression_donanemab <- 0.68

# --- Mortality Hazard Ratios ---
hr_mortality_mci <- 1.61
hr_mortality_mild <- 2.23
hr_mortality_moderate <- 3.10
hr_mortality_severe <- 4.98

# --- Utilities (annual values converted to monthly) ---
u.mci <- 0.73 / 12
u.mild <- 0.69 / 12
u.moderate <- 0.53 / 12
u.severe <- 0.34 / 12
u.death <- 0

# Utility decrement for symptomatic ARIA
aria_utility_decrement <- -0.065 / 12  # Applied for 3 months

# --- Background Monthly Costs (Healthcare Sector) ---
c.hc.mci <- 691
c.hc.mild <- 1049
c.hc.moderate <- 1274
c.hc.severe <- 3764
c.hc.death <- 0

# --- Background Monthly Costs (Societal Increment) ---
c.soc.mci <- 49
c.soc.mild <- 1483
c.soc.moderate <- 1932
c.soc.severe <- 2153
c.soc.death <- 0

# --- Drug Costs ---
c.aducanumab_year1 <- 20500
c.aducanumab_year2plus <- 28200
c.donanemab_annual <- 28200

# --- Monthly Infusion and Monitoring Costs ---
# Aducanumab
c.aducanumab_months1_6 <- 112
c.aducanumab_months7_12 <- 87
c.aducanumab_months13plus <- 80
# Donanemab
c.donanemab_months1_6 <- 599
c.donanemab_months7_12 <- 302
c.donanemab_months13plus <- 287

# --- Screening Costs (One-Time) ---
c.screening_aducanumab <- 6957
c.screening_donanemab <- 17096

# --- ARIA-Related Costs ---
aria_probability_aducanumab <- 0.41
aria_probability_donanemab <- 0.39
c.aria_followup_imaging <- 1060
c.aria_hospitalization <- 35934
c.aria_outpatient_visit <- 128

# Note: ARIA assumed to occur in first 24 months, with most cases within the first 12
# Distribution of ARIA incidence by month (used later in model if needed):
aria_distribution <- c(m1_3 = 0.5, m4_12 = 0.4, m13_24 = 0.1)


# Prompt 2: Load and process mortality rates

# Load mortality data
mortality_data <- read.csv("mortalityRates_US_2017.csv")

# Compute age-specific death probabilities for each disease severity state
hr_mci <- 1.61
hr_mild <- 2.23
hr_moderate <- 3.10
hr_severe <- 4.98

mortality_processed <- data.frame(
  age = mortality_data$age,
  prob_death_mci = 1 - exp(-mortality_data$monthly_rate * hr_mci),
  prob_death_mild = 1 - exp(-mortality_data$monthly_rate * hr_mild),
  prob_death_moderate = 1 - exp(-mortality_data$monthly_rate * hr_moderate),
  prob_death_severe = 1 - exp(-mortality_data$monthly_rate * hr_severe)
)


# Prompt 3: Define time-dependent transition probability vectors

# Define number of model cycles (e.g., 360 for 30 years at monthly cycles)
n_cycles <- 360

# --- Base progression probabilities ---
p_mci_to_mild <- p.mci_to_mild
p_mild_to_moderate <- p.mild_to_moderate
p_moderate_to_severe <- p.moderate_to_severe

# --- Standard care (no intervention) ---
p_mci_to_mild_std <- rep(p_mci_to_mild, n_cycles)
p_mci_stay_std <- rep(1 - p_mci_to_mild, n_cycles)

p_mild_to_moderate_std <- rep(p_mild_to_moderate, n_cycles)
p_mild_stay_std <- rep(1 - p_mild_to_moderate, n_cycles)

p_moderate_to_severe_std <- rep(p_moderate_to_severe, n_cycles)
p_moderate_stay_std <- rep(1 - p_moderate_to_severe, n_cycles)

p_severe_stay_std <- rep(1, n_cycles)

# --- Aducanumab intervention ---
rate_mci_aducanumab <- -log(1 - p_mci_to_mild) * hr_progression_aducanumab
rate_mild_aducanumab <- -log(1 - p_mild_to_moderate) * hr_progression_aducanumab
rate_moderate_aducanumab <- -log(1 - p_moderate_to_severe) * hr_progression_aducanumab

p_mci_to_mild_adu <- rep(1 - exp(-rate_mci_aducanumab), n_cycles)
p_mci_stay_adu <- rep(1 - p_mci_to_mild_adu[1], n_cycles)

p_mild_to_moderate_adu <- rep(1 - exp(-rate_mild_aducanumab), n_cycles)
p_mild_stay_adu <- rep(1 - p_mild_to_moderate_adu[1], n_cycles)

p_moderate_to_severe_adu <- rep(1 - exp(-rate_moderate_aducanumab), n_cycles)
p_moderate_stay_adu <- rep(1 - p_moderate_to_severe_adu[1], n_cycles)

p_severe_stay_adu <- rep(1, n_cycles)

# --- Donanemab intervention ---
rate_mci_donanemab <- -log(1 - p_mci_to_mild) * hr_progression_donanemab
rate_mild_donanemab <- -log(1 - p_mild_to_moderate) * hr_progression_donanemab
rate_moderate_donanemab <- -log(1 - p_moderate_to_severe) * hr_progression_donanemab

p_mci_to_mild_dona <- rep(1 - exp(-rate_mci_donanemab), n_cycles)
p_mci_stay_dona <- rep(1 - p_mci_to_mild_dona[1], n_cycles)

p_mild_to_moderate_dona <- rep(1 - exp(-rate_mild_donanemab), n_cycles)
p_mild_stay_dona <- rep(1 - p_mild_to_moderate_dona[1], n_cycles)

p_moderate_to_severe_dona <- rep(1 - exp(-rate_moderate_donanemab), n_cycles)
p_moderate_stay_dona <- rep(1 - p_moderate_to_severe_dona[1], n_cycles)

p_severe_stay_dona <- rep(1, n_cycles)


# Prompt 4A: Initialize transition probability array with age-specific mortality and structural setup

build_transition_matrix <- function(strat) {
  # Initialize 3D transition probability array: states x states x cycles
  trans_mat <- array(0, dim = c(length(states), length(states), n_cycles),
                     dimnames = list(states, states, NULL))
  
  for (cycle in 1:n_cycles) {
    
    # Determine age at current cycle (assuming one cycle per month)
    age <- floor(initial_age + (cycle - 1) / 12)
    
    # Cap age at 99 if it exceeds bounds of mortality data
    age <- min(age, 99)
    
    # Extract mortality probabilities from processed data
    prob_death <- mortality_processed[mortality_processed$age == age, ]
    
    # Assign transition probabilities to death
    trans_mat["mci", "death", cycle] <- prob_death$prob_death_mci
    trans_mat["mild", "death", cycle] <- prob_death$prob_death_mild
    trans_mat["moderate", "death", cycle] <- prob_death$prob_death_moderate
    trans_mat["severe", "death", cycle] <- prob_death$prob_death_severe
    trans_mat["death", "death", cycle] <- 1
    
    # [INSERT PROMPT 4B HERE] — define transitions to next disease stages (with or without treatment)
    
    if (strat == "no intervention") {
      trans_mat["mci", "mild", cycle] <- p_mci_to_mild_std[cycle] * (1 - trans_mat["mci", "death", cycle])
      trans_mat["mci", "mci", cycle]  <- p_mci_stay_std[cycle] * (1 - trans_mat["mci", "death", cycle])
      
      trans_mat["mild", "moderate", cycle] <- p_mild_to_moderate_std[cycle] * (1 - trans_mat["mild", "death", cycle])
      trans_mat["mild", "mild", cycle]     <- p_mild_stay_std[cycle] * (1 - trans_mat["mild", "death", cycle])
      
      trans_mat["moderate", "severe", cycle] <- p_moderate_to_severe_std[cycle] * (1 - trans_mat["moderate", "death", cycle])
      trans_mat["moderate", "moderate", cycle] <- p_moderate_stay_std[cycle] * (1 - trans_mat["moderate", "death", cycle])
      
      trans_mat["severe", "severe", cycle] <- p_severe_stay_std[cycle] * (1 - trans_mat["severe", "death", cycle])
      
    } else if (strat == "aducanumab") {
      trans_mat["mci", "mild", cycle] <- p_mci_to_mild_adu[cycle] * (1 - trans_mat["mci", "death", cycle])
      trans_mat["mci", "mci", cycle]  <- p_mci_stay_adu[cycle] * (1 - trans_mat["mci", "death", cycle])
      
      trans_mat["mild", "moderate", cycle] <- p_mild_to_moderate_adu[cycle] * (1 - trans_mat["mild", "death", cycle])
      trans_mat["mild", "mild", cycle]     <- p_mild_stay_adu[cycle] * (1 - trans_mat["mild", "death", cycle])
      
      trans_mat["moderate", "severe", cycle] <- p_moderate_to_severe_adu[cycle] * (1 - trans_mat["moderate", "death", cycle])
      trans_mat["moderate", "moderate", cycle] <- p_moderate_stay_adu[cycle] * (1 - trans_mat["moderate", "death", cycle])
      
      trans_mat["severe", "severe", cycle] <- p_severe_stay_adu[cycle] * (1 - trans_mat["severe", "death", cycle])
      
    } else if (strat == "donanemab") {
      trans_mat["mci", "mild", cycle] <- p_mci_to_mild_dona[cycle] * (1 - trans_mat["mci", "death", cycle])
      trans_mat["mci", "mci", cycle]  <- p_mci_stay_dona[cycle] * (1 - trans_mat["mci", "death", cycle])
      
      trans_mat["mild", "moderate", cycle] <- p_mild_to_moderate_dona[cycle] * (1 - trans_mat["mild", "death", cycle])
      trans_mat["mild", "mild", cycle]     <- p_mild_stay_dona[cycle] * (1 - trans_mat["mild", "death", cycle])
      
      trans_mat["moderate", "severe", cycle] <- p_moderate_to_severe_dona[cycle] * (1 - trans_mat["moderate", "death", cycle])
      trans_mat["moderate", "moderate", cycle] <- p_moderate_stay_dona[cycle] * (1 - trans_mat["moderate", "death", cycle])
      
      trans_mat["severe", "severe", cycle] <- p_severe_stay_dona[cycle] * (1 - trans_mat["severe", "death", cycle])
    }
    
    # Normalize rows to ensure row sums = 1
    for (state in states) {
      row_sum <- sum(trans_mat[state, , cycle])
      if (row_sum > 0 && row_sum != 1) {
        trans_mat[state, , cycle] <- trans_mat[state, , cycle] / row_sum
      }
    }
  }
  
  return(trans_mat)
}


# Prompt 5: Run the Markov model and compute discounted outcomes

# Monthly discount factor for 3% annual rate
discount_factor <- 1 / ((1 + 0.03)^(1 / 12))

# Initialize list to store results
results <- list()

# Function to simulate model for a given strategy
run_model <- function(strategy) {
  trans_mat <- build_transition_matrix(strategy)
  trace <- matrix(0, nrow = n_cycles + 1, ncol = length(states), dimnames = list(NULL, states))
  trace[1, ] <- initial_state_dist * cohort_size
  
  qalys <- numeric(n_cycles)
  healthcare_costs <- numeric(n_cycles)
  societal_costs <- numeric(n_cycles)
  
  # Track treatment status for each state and cycle
  on_treatment <- matrix(TRUE, nrow = n_cycles + 1, ncol = length(states), dimnames = list(NULL, states))
  on_treatment[1, "moderate"] <- FALSE
  on_treatment[1, "severe"] <- FALSE
  on_treatment[1, "death"] <- FALSE
  
  # Treatment discontinuation logic
  discontinuation_aducanumab <- c(seq(0, 0.38, length.out = 12), rep(0.38, n_cycles - 12))
  remain_on_adu <- 1 - discontinuation_aducanumab
  switch_off_dona <- c(rep(0.27, 6), rep(0.55, 6), rep(1, n_cycles - 12))
  remain_on_dona <- 1 - switch_off_dona
  
  # One-time screening costs at cycle 1
  screening_cost <- switch(strategy,
                           "aducanumab" = c.screening_aducanumab,
                           "donanemab" = c.screening_donanemab,
                           "no intervention" = 0)
  
  for (cycle in 1:n_cycles) {
    trace[cycle + 1, ] <- trace[cycle, ] %*% trans_mat[, , cycle]
    
    # Determine on-treatment cohort
    if (strategy == "aducanumab") {
      treat_fraction <- remain_on_adu[cycle]
    } else if (strategy == "donanemab") {
      treat_fraction <- remain_on_dona[cycle]
    } else {
      treat_fraction <- 0
    }
    
    on_treatment[cycle + 1, "mci"] <- strategy != "no intervention" && treat_fraction > 0
    on_treatment[cycle + 1, "mild"] <- strategy != "no intervention" && treat_fraction > 0
    on_treatment[cycle + 1, "moderate"] <- FALSE
    on_treatment[cycle + 1, "severe"] <- FALSE
    on_treatment[cycle + 1, "death"] <- FALSE
    
    # QALYs
    utility_vector <- c(u.mci, u.mild, u.moderate, u.severe, u.death)
    qaly_cycle <- sum(trace[cycle, ] * utility_vector)
    
    # ARIA-related disutility in cycles 1–3 (symptomatic ARIA only)
    if (cycle <= 3 && strategy != "no intervention") {
      symptomatic_aria <- switch(strategy,
                                 "aducanumab" = aria_probability_aducanumab * 0.24,
                                 "donanemab" = aria_probability_donanemab * 0.16,
                                 0)
      qaly_cycle <- qaly_cycle + (trace[cycle, "mci"] + trace[cycle, "mild"]) * symptomatic_aria * aria_utility_decrement
    }
    
    # Discounted QALYs
    qalys[cycle] <- qaly_cycle * discount_factor^cycle
    
    # Healthcare sector costs (background + treatment)
    cost_hc_vector <- c(c.hc.mci, c.hc.mild, c.hc.moderate, c.hc.severe, c.hc.death)
    hc_cycle <- sum(trace[cycle, ] * cost_hc_vector)
    
    # Societal extra costs
    cost_soc_vector <- c(c.soc.mci, c.soc.mild, c.soc.moderate, c.soc.severe, c.soc.death)
    soc_cycle <- sum(trace[cycle, ] * cost_soc_vector)
    
    # Drug costs (annual, converted monthly), only if on treatment
    drug_cost_monthly <- switch(strategy,
                                "aducanumab" = ifelse(cycle <= 12, c.aducanumab_year1 / 12, c.aducanumab_year2plus / 12),
                                "donanemab" = c.donanemab_annual / 12,
                                0)
    
    drug_costs <- drug_cost_monthly * sum(trace[cycle, c("mci", "mild")] * treat_fraction)
    
    # Infusion and monitoring
    infusion_cost <- switch(strategy,
                            "aducanumab" = ifelse(cycle <= 6, c.aducanumab_months1_6,
                                                  ifelse(cycle <= 12, c.aducanumab_months7_12, c.aducanumab_months13plus)),
                            "donanemab" = ifelse(cycle <= 6, c.donanemab_months1_6,
                                                 ifelse(cycle <= 12, c.donanemab_months7_12, c.donanemab_months13plus)),
                            0)
    
    infusion_costs <- infusion_cost * sum(trace[cycle, c("mci", "mild")] * treat_fraction)
    
    # ARIA-related cost (outpatient + imaging + hospitalization for 4%)
    aria_cost_cycle <- 0
    if (cycle <= 3 && strategy != "no intervention") {
      symptomatic_cases <- switch(strategy,
                                  "aducanumab" = trace[cycle, c("mci", "mild")] * aria_probability_aducanumab * 0.24,
                                  "donanemab" = trace[cycle, c("mci", "mild")] * aria_probability_donanemab * 0.16)
      n_sympt_cases <- sum(symptomatic_cases)
      aria_cost_cycle <- n_sympt_cases * (c.aria_outpatient_visit + c.aria_followup_imaging + 0.04 * c.aria_hospitalization)
    }
    
    # Total discounted costs
    total_hc <- (hc_cycle + drug_costs + infusion_costs + aria_cost_cycle) * discount_factor^cycle
    total_soc <- (soc_cycle + drug_costs + infusion_costs + aria_cost_cycle) * discount_factor^cycle
    
    healthcare_costs[cycle] <- total_hc
    societal_costs[cycle] <- total_soc
  }
  
  return(list(
    qalys = sum(qalys) / cohort_size,
    healthcare_costs = sum(healthcare_costs) / cohort_size + screening_cost,
    societal_costs = sum(societal_costs) / cohort_size + screening_cost
  ))
}

# Run the model for each strategy
results$no_intervention <- run_model("no intervention")
results$aducanumab <- run_model("aducanumab")
results$donanemab <- run_model("donanemab")
```
