## Supplementary material for "Replicating Health-Economic Simulation Models for Alzheimer’s Disease Using Artificial Intelligence": final R scripts: validation15Trials_Ross.R

```
##### Code to compare the obtained final results to those of Wimo #####


# Clear console and environment
cat("\014") 
rm(list = ls()) 

# Load required libraries
library(knitr)
library(ggplot2)
library(tidyr)
library(dplyr)
library(extrafont)

# Define reference results from Ross et al. (2022)
reference_values <- list(
  no_intervention = list(
    qalys = 4.948,
    healthcare_costs = 118000,
    societal_costs = 205200
  ),
  aducanumab = list(
    qalys = 4.948 + 0.133,
    healthcare_costs = 118000 + 130100,
    societal_costs = 205200 + 127800
  ),
  donanemab = list(
    qalys = 4.948 + 0.408,
    healthcare_costs = 118000 + 78700,
    societal_costs = 205200 + 71600
  )
)

# Add total costs to each strategy
for (strategy in names(reference_values)) {
  reference_values[[strategy]]$total_costs <-
    reference_values[[strategy]]$healthcare_costs +
    reference_values[[strategy]]$societal_costs
}

# Prepare simulation names and metrics
sim_names <- paste0("Ross_O", 1:15)
metrics <- c("qalys", "healthcare_costs", "societal_costs", "total_costs")

# Function to compare each simulation against reference values
compare_simulation <- function(res, ref) {
  comparison <- c()
  for (strategy in c("no_intervention", "aducanumab", "donanemab")) {
    # Compute total costs in simulation results
    res[[strategy]]$total_costs <- res[[strategy]]$healthcare_costs + res[[strategy]]$societal_costs
    
    for (metric in metrics) {
      value <- res[[strategy]][[metric]]
      diff_abs <- value - ref[[strategy]][[metric]]
      diff_rel <- diff_abs / (ref[[strategy]][[metric]])
      comparison <- c(comparison, value, diff_abs, diff_rel)
    }
  }
  return(comparison)
}

# Run all simulations and store comparison results
results_matrix <- t(sapply(sim_names, function(sim_name) {
  env <- new.env()
  sys.source(paste0(sim_name, ".R"), envir = env)
  compare_simulation(env$results, reference_values)
}))

# Create column names
column_names <- c()
for (strategy in c("no_intervention", "aducanumab", "donanemab")) {
  for (metric in metrics) {
    column_names <- c(column_names,
                      paste0(strategy, "_", metric),
                      paste0(strategy, "_", metric, "_diff_abs"),
                      paste0(strategy, "_", metric, "_diff_rel"))
  }
}

# Construction du data frame final
comparison_df <- as.data.frame(results_matrix)
colnames(comparison_df) <- column_names
rownames(comparison_df) <- sim_names

# Affichage dans RStudio
View(comparison_df)


################ Bar plots for QALYs and Total Costs####################"

# Select relevant columns from comparison_df
Ross_data <- comparison_df %>%
  select(
    no_intervention_qalys,
    aducanumab_qalys,
    donanemab_qalys,
    no_intervention_total_costs,
    aducanumab_total_costs,
    donanemab_total_costs
  )

# Add simulation ID
ross_data$SimID <- rownames(ross_data)
ross_data <- ross_data[, c("SimID", setdiff(names(ross_data), "SimID"))]

# Create reference row
ref_row <- ross_data[1, , drop = FALSE]
ref_row[] <- NA
ref_row$SimID <- "Ross"
ref_row$no_intervention_qalys   <- reference_values$no_intervention$qalys
ref_row$aducanumab_qalys        <- reference_values$aducanumab$qalys
ref_row$donanemab_qalys         <- reference_values$donanemab$qalys
ref_row$no_intervention_total_costs <- reference_values$no_intervention$total_costs
ref_row$aducanumab_total_costs      <- reference_values$aducanumab$total_costs
ref_row$donanemab_total_costs       <- reference_values$donanemab$total_costs

# Add reference row on top
ross_data <- bind_rows(ref_row, ross_data)

# Rename and reorder factor levels
old_levels <- c("Ross", paste0("Ross_O", 1:15))
new_labels <- c("Ross", paste0("Sim_", 1:15))
ross_data$SimID <- factor(ross_data$SimID, levels = old_levels, labels = new_labels)

# Simplify column names
colnames(ross_data) <- gsub("no_intervention_", "no_", colnames(ross_data))
colnames(ross_data) <- gsub("aducanumab_", "aducanumab_", colnames(ross_data))
colnames(ross_data) <- gsub("donanemab_", "donanemab_", colnames(ross_data))
colnames(ross_data) <- gsub("final_costs", "costs", colnames(ross_data))

# Convert to long format for plotting
plot_data <- Ross_data %>%
  pivot_longer(
    -SimID,
    names_to = c("Strategy", "Metric"),
    names_pattern = "([^_]+)_(qalys|total_costs)",
    values_to = "Value"
  ) %>%
  mutate(
    Strategy = case_when(
      Strategy == "no" ~ "Standard of Care",
      Strategy == "aducanumab" ~ "Aducanumab",
      Strategy == "donanemab" ~ "Donanemab"
    ),
    is_ref = ifelse(SimID == "Ross", "ref", "sim"),
    fill_group = paste(Strategy, is_ref, sep = "_"),
    Metric = ifelse(Metric == "total_costs", "costs", Metric)  # renommage standardisé
  )

# Define custom colors
strategy_colors <- c(
  "Standard of Care_ref" = "#cc8400",
  "Standard of Care_sim" = "#ffbd59",
  "Aducanumab_ref" = "#6396a6",
  "Aducanumab_sim" = "#b9d8de",
  "Donanemab_ref" =  "#a6a6a6",
  "Donanemab_sim" = "#d9d9d9"
)

plot_data$fill_group <- as.character(plot_data$fill_group)
plot_data$fill_group <- trimws(plot_data$fill_group)
plot_data$fill_group <- factor(plot_data$fill_group, levels = names(strategy_colors))

# Mapping for reference metrics
ref_map <- list(qalys = "qalys", costs = "total_costs")
metric_labels <- c(qalys = "QALYs", costs = "Costs")

# Generate bar plots
for (m in names(m_labels)) {
  ref_soc <- reference_values$no_intervention[[ref_map[[m]]]]
  ref_adu <- reference_values$aducanumab[[ref_map[[m]]]]
  ref_dona <- reference_values$donanemab[[ref_map[[m]]]]
  
  p <- ggplot(filter(plot_data, Metric == m), aes(x = SimID, y = Value, fill = fill_group)) +
    geom_bar(stat = "identity", position = position_dodge(width = 0.8)) +
    scale_fill_manual(values = strategy_colors) +
    geom_hline(yintercept = ref_soc, linetype = "dashed", color = "#cc8400", linewidth = 0.6) +
    geom_hline(yintercept = ref_adu, linetype = "dashed", color = "#6396a6", linewidth = 0.6) +
    geom_hline(yintercept = ref_dona, linetype = "dashed", color = "#a6a6a6", linewidth = 0.6) +
    labs(
      title = paste("Ross –", m_labels[[m]], ": Original vs. AI-generated simulations"),
      x = "Simulation",
      y = m_labels[[m]]
          ) +
    theme_minimal() +
    theme(
      plot.title = element_text(hjust = 0.5, face = "bold", size = 14),
      axis.text.x = element_text(angle = 45, hjust = 1),
      text = element_text(family = "Verdana"),
      legend.position = "none"
      
    )
  print(p)
}
```
