## Supplementary figures and images for "Replicating Health-Economic Simulation Models for Alzheimer’s Disease Using Artificial Intelligence"

### SequenceOfPromptsSchema_Ross.png

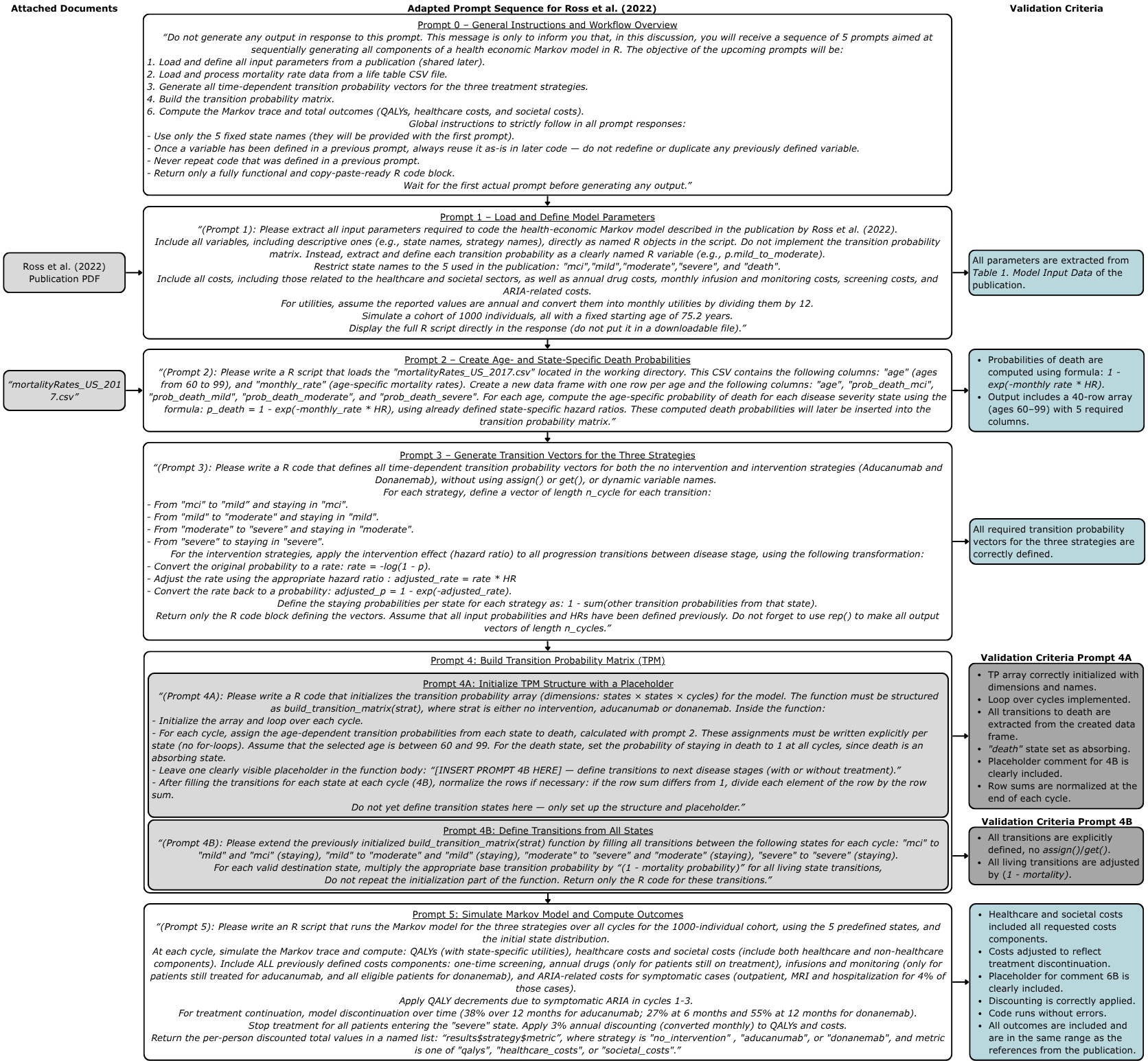

### Wimo_SequenceOfPromptsSchema.png

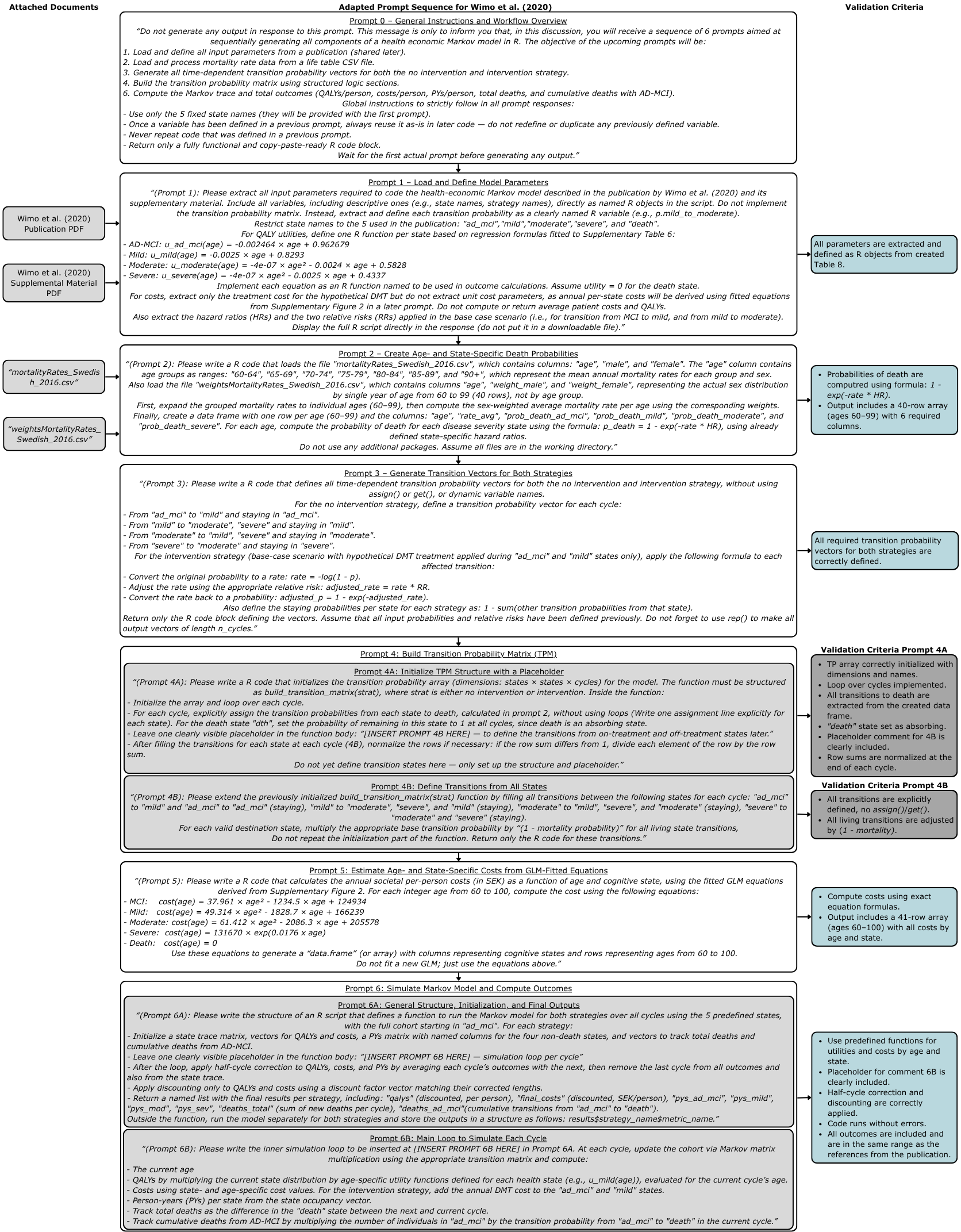
